# SARS-CoV-2 Genetic diversity and lineage dynamics of in Egypt

**DOI:** 10.1101/2022.01.05.22268646

**Authors:** Wael H. Roshdy, Mohamed K. khalifa, James Emmanuel San, Houriiyah Tegally, Eduan Wilkinson, Shymaa Showky, Daren Patrick Martin, Monika Moir, Amel Naguib, Nancy Elguindy, Mokhtar R. Gomaa, Manal Fahim, Hanaa Abu Elsood, Amira Mohsen, Ramy Galal, Mohamed Hassany, Richard J Lessells, Ahmed A. Al-Karmalawy, Rabeh EL-Shesheny, Ahmed M. Kandeil, Mohamed A. Ali, Tulio de Oliveira

## Abstract

COVID-19 was first diagnosed in Egypt on 14 February 2020. By the end of November 2021, over 333,840 cases and 18,832 deaths had been reported. As part of national genomic surveillance, 1,027 SARS-CoV-2 near whole-genomes had been generated and published by the end of May 2021. Here we describe the genomic epidemiology of SARS-CoV-2 in Egypt over this period using a subset of 976 high-quality Egyptian genomes analysed together with a representative set of global sequences within a phylogenetic framework. We show that a single lineage, C.36, introduced early in the pandemic was responsible for most cases in Egypt. Furthermore, we show that to remain dominant in the face of mounting immunity from previous infection and vaccination, this lineage evolved into various sub-lineages acquiring several mutations known to confer adaptive advantage and pathogenic properties. These results highlight the value of continuous genomic surveillance in regions where VOCs are not predominant and enforcement of public health measures to prevent expansion of existing lineages.

## Introduction

Over the first 21 months of the COVID-19 pandemic, several lineages of the SARS-CoV-2 characterized by diverse constellations of mutations have emerged. Some of these lineages have manifested phenotypes such as increased transmissibility, disease severity, and escape from neutralizing antibodies and have therefore been classified as variants of concern (VOC); e.g Alpha, Beta, Gamma, Delta and most recently, Omicron (Tao et al., 2021). Other lineages carrying mutational loads similar to those of VOCs and which therefore might in future present similar phenotypes to VOCs have been classified as variants of interest (VOI); e.g., Eta, Iota, Kappa, and Lambda. Recently, the World Health Organisation (WHO) and the European Center for Disease Prevention and Control (ECDCP) designated a new category of variant: variants under monitoring (VUM), that, although not yet confirmed as an immediate exceptional risk, might pose future threats to pharmaceutical and non-pharmaceutical interventions to control or end the pandemic (ECDC, 2021).

To date, studies to understand the impact of genomic structural variations on viral phenotypes have largely focused on VOCs and VOIs. Accordingly, stringent public health measures such as travel restrictions and lockdowns have been implemented at different times and in different regions of the world to control the spread of these variants. Considerably less attention has been given to understanding the emergence, local spread and persistence of non-VOC or VOI lineages in the parts of the world where they emerged. This has favored the continued evolution of some of these lineages in the regions where they persist: evolution that, independently of that ongoing elsewhere, has in some cases likely yielded variants with improved transmission and/or immune evasion phenotypes.

Here we describe the molecular epidemiology of SARS-CoV-2 in Egypt up to July 2021, focusing on the C.36 lineage that circulated within the country from early in the pandemic. This lineage evolved into sub-lineages C.36.1, C.36.3 and C.36.3.1, acquiring mutations known to confer fitness and pathogenic properties similar to those found in VOCs; with C.36.3 having since been classified as a VUM in June 2021. Our results highlight the importance of sustained surveillance and public health efforts to minimize the transmission, replication and evolution of variants within the regions where they arise.

## Results

### Genomic epidemiology of SARS-CoV-2 in Egypt

The first case of COVID-19 in Egypt was reported on 14 February 2020, in a foreign national of undisclosed nationality (AlJazeera, 2020; Fouchier et al., 2021). By 18 March 2021, at least 256 cases and 7 deaths had been reported. Since then, over 333,840 confirmed cases and 18,832 deaths (crude case-fatality rate 5.6 percent) have been reported (WHO, 2021), making Egypt, based on this metric at least, the most affected country in Africa after South Africa. At the onset of the pandemic, the Egyptian authorities devised a comprehensive response plan to control the spread of SARS-CoV-2, including various types of public health and social restriction measures to mitigate the effects of the virus (Medhat and El Kassas, 2020). On 19 March 2020, Egypt closed all airports and suspended air travel. Two days later, all social and religious gatherings were suspended, and curfew imposed until the end of March. Restrictions were followed by campaigns to increase awareness in the population (Saied et al., 2021). Unfortunately, these attempts and efforts were inadequate to curtail the spread of SARS-CoV-2 and, as a result, three waves of infection ensued (Fig. 1a). The surge in cases during the first wave, which lasted from April to August 2020 (Fig. 1a), was largely driven by religious and cultural celebrations in the holy month of Ramadan and the Fitr Islamic Holiday as well as a season of traditional wedding celebrations, and family gatherings characterized by little to no social distancing (Gomaa et al., 2021). This is in line with results from seroprevalence studies during this period which revealed high rates of transmission and seroprevalence within households and among large numbers of patients requiring admission. For example, in a study conducted on 290 households between April and October 2020, seroprevalence was estimated at 34.8% with a secondary attack rate of 89.8% (Fouchier et al., 2021) while a separate study in Cairo estimated a seroprevalence of 29.8% with middle aged men at higher risk (Girgis et al., 2021). An even higher rate of seroprevalence of 46.3% was reported among health care workers (El-Sokkary et al., 2021). The second wave coincided with the resumption of schools and commencement of the cold weather season (October 2020 to April 2021). Similar to the first wave, the third wave was also associated with the start of religious and cultural celebrations. Of the three waves, the first wave was associated with the most severe impact, characterized by the highest case counts and associated deaths. The attenuated effects of the second and third wave could be attributed to prevailing immunity from infections in the first wave and commencement of vaccination during the second wave. In December 2020, the Emirati government donated 50,000 Sinopharm vaccine doses to Egypt. This was followed by a donation of 300,000 Sinopharm doses from the Chinese government. Egypt also received 50,000 doses of Oxford’s AstraZeneca vaccine (ElSharkawy, 2021). Vaccination commenced in January 2021. By December 2021 30% of the population had been fully or partially vaccinated (https://ourworldindata.org/covid-vaccinations?country=OWID_WRL, 21 December 2021) (Mathieu et al., 2021).

**Fig 1.**
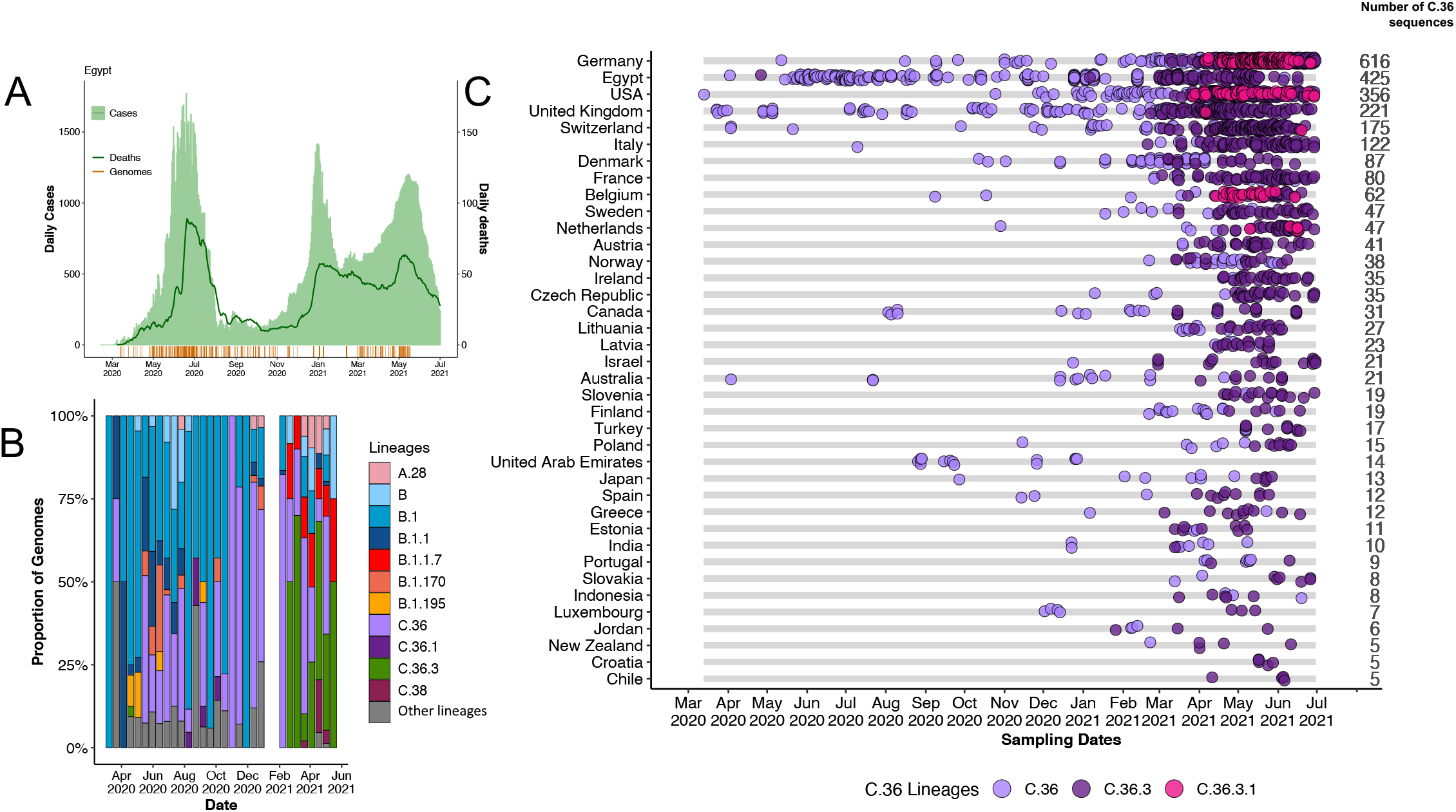
Epidemiological dynamics of the SARS-CoV-2 epidemic in Egypt. A) Histogram showing the number of daily COVID-19 cases overlayed with a line plot of deaths: in the bottom, timing of genomic sampling in Egypt B) Progressive distribution of the major SARS-CoV-2 lineages in Egypt. C) Temporal sampling of sequences of the C.36 SARS-CoV-2 lineages globally in countries with at least 5 sequences (ordered by total number of sequences).

### Phylogenetic analysis and Lineage Dynamics

On 13 March 2020, the first two SARS-CoV-2 genomes from Egypt were published (Kandeil et al., 2020). By June 2021, close to 1,027 SARS-CoV-2 near-full length genomes had been submitted to the GISAID (Shu and McCauley, 2017) database. Of these, 976 genomes were of good quality (genome coverage range 82.6 to 99.5%) (Table S1) and were associated with relevant metadata. To contextualize the epidemic in Egypt, we analyzed these sequences in a phylogenetic framework with a representative set of global sequences. The inferred phylogeny (Fig. 2A) enabled us to estimate the number of viral imports and exports between Egypt and the rest of the world. Our analysis suggests that there were at least 56 unique introductions of SARS-CoV-2 in Egypt (20 (34%) from Europe, 12 (20%) from Africa, 10 (17%) from Oceania, 9 (15%) from Asia and 7 (11%) from the Americas) (Figure S2A) with the earliest introduction from Australia. At country level, most of the viral introductions emanated from Australia (n = 9 (16%)) and United Kingdom (n = 9 (16%)). We also identified at least 364 export events mostly to Europe and USA (Fig 2B and C, Figure S2). Genomic analysis of the sequences revealed the circulation of at least 36 distinct SARS-CoV-2 lineages spread across the three waves which is in line with the number of introductions. The first wave, as is the case with most countries, was primarily dominated by lineage B viruses, i.e., the B.1 (n = 287, 32%) and B.1.1 (n = 57, 6%) PANGO lineages. Viruses belonging to these lineages carry the well-known D614G mutation that has been linked to increased transmissibility (Korber et al., 2020) but lack any of the other characteristic Spike mutations associated with VOCs or VOIs. Lineage A.28, the only A-lineage detected in Egypt was first sampled around August 2020. However, like in other parts of the continent, the A.28 lineage does not seem to have taken off and by May 2021, no sequences of the A.28 lineage have been sampled in Egypt.

**Fig 2.**
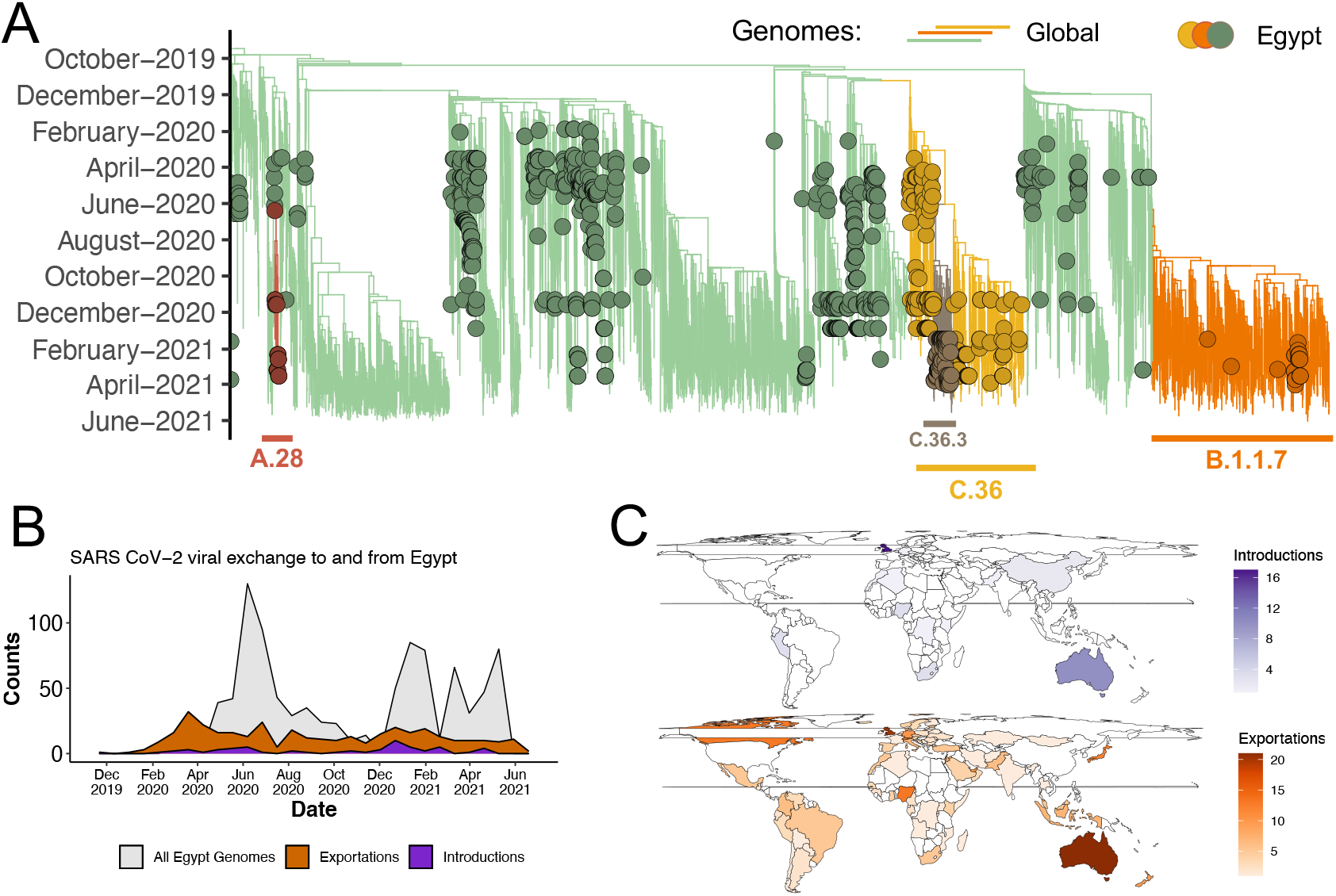
Phylogenetic reconstruction of SARS-CoV-2 epidemic in Egypt. A) Time-resolved maximum clade credibility phylogeny of major lineages circulating in Egypt. B) Area plot showing viral import-export as proportions of the sampled genomes. C) Inferred locations of imports into and exports of SARS-CoV-2 out of Egypt. Sampling for molecular and genomic analyses across Egypt were sporadic across 2020 and 2021. Although genomic sequencing only C.36/C.36.3 detected in Cairo (GISAID Metadata), qPCR assays were able to detect these lineages in various other locations (Fig 3B) at a high proportion of the total numbers of samples tested with qPCR assay (Fig 3A)

Although the Beta/B.1.351 variant which was first detected in South Africa (Tegally et al., 2021) dominated most of the second waves in Africa, at the time of writing it had not been detected in Egypt. This could be attributed to under-sampling, compounded by limited viral exchanges with the sub-Saharan countries, a known trend between countries in North Africa and those in sub-Saharan Africa (Wilkinson et al., 2021). The only VOC detected in Egypt was Alpha (Fig. 1b) with the first sequence being detected in March 2021. Interestingly, unlike in European countries reporting the rapid rise of Alpha throughout the first months of 2021, Egypt did not see similar growth in the number of cases attributed to Alpha following its introduction (Fig 1b). Rather, majority of the cases where travel-associated with minimal community transmission.

### Introduction of the C.36 lineage in Egypt

The C.36 lineage was detected very early on in the epidemic in Egypt (in March 2020) and has circulated within the country at varying frequencies since then (Fig 1B). Relative to B.1, the C.36 lineage is characterized by an additional mutation, Q677H in its Spike gene, upstream of the S1/S2 furin cleavage site. Notably, the Q677H mutation has also occurred independently in several other SARS-CoV-2 lineages and the VUM Eta/B.1.525; suggestive of a host adaptation advantage (Hodcroft et al., 2021). Our import export analysis suggests that the progenitor of this lineage it was likely introduced to Egypt from Australia (Fig.1 C). Despite the prevalence of C.36 lineage in Egypt and minimal viral export to neighboring countries such as Morocco, we did not find any evidence of regional spread to neighboring countries (Figure S1).

### Evolution and spread of C.36 sub-lineages

As of April 2020, the C.36 lineage begun to evolve into sub-lineages that include C.36.1, C.36.3, and C.36.3.1. The emergence of these sub-lineages is suggestive of a shift in the selective environment in which SARS-CoV-2 was evolving, i.e. from naïve susceptible to more resilient hosts likely due to mounting immunity from prior infections and later on the commencement of vaccination (Martin et al., 2021).

Lineage C.36.3 was particularly associated with a selective advantage (Fig 1B) and was responsible for the majority of the cases in Egypt during the third wave. As our genomic data was largely limited to Cairo, to characterize the spread of C.36.3 across Egypt, we utilized real-time polymerase chain reaction (RT-PCR) data from 1,076 nasopharyngeal swab samples with high quality demographic data. Of the 1,076 samples analysed, 462 samples (42.9%) belonged to the C.36.3 sub-lineage while only 35 samples (3.3%) belonged to the Alpha variant. Of the 462 samples, at least 33% where from Cairo, 13% from Sinai, and 11.8% from Giza, 9.9% from Sohage, 9.1% from Aswan, 8.6% from Alexandria, 4.9% Dakhlia, and 4.7% from Luxor; the remaining 8% was spread across the remaining governorates. Together these results show widespread community transmission of C.36.3 in all population centers of Egypt.

### Mutational and Selection analysis of the C.36.3

The C.36.3 lineage accumulated an additional six Spike mutations and eleven non-Spike mutations over the core set of mutations of the C.36 lineage (Figure 3 A and B). Among these, the Spike mutation L452R, also detected in other VOIs and VOCs (including Iota/B.1.526.1, Epsilon/B.1.427/B.1.429 and Delta/B.1.617.2 lineages) occurs in the receptor binding motif (RBM): the portion of the Spike that mediates contact with ACE2 (Shang et al., 2020). Occurrence of the L452R and R346S mutations in the RBD has been associated with reduction in neutralizing susceptibility to several RBM class II monoclonal antibodies (mAbs) as well as convalescent and vaccine sera (Amanat et al., 2021; Greaney et al., 2021; Li et al., 2020). The sub-lineage also acquired the ΔHV69-70 deletion originally detected in the Alpha variant which is associated with increased viral replication and infectivity (Kemp et al., 2021; Meng et al., 2021). Together, these mutations are likely to have conferred a significant adaptive advantage to the sub-lineage. Furthermore, it also harbors two additional mutations in the N-terminal domain (NTD), (S12F and W152R) that are potentially associated with an antigenic shift (England, 2021). Results from *in silico* experiments suggest the W152R mutation may weaken interactions between the Spike and neutralizing antibodies (Kubik et al., 2021).

**Fig 3.**
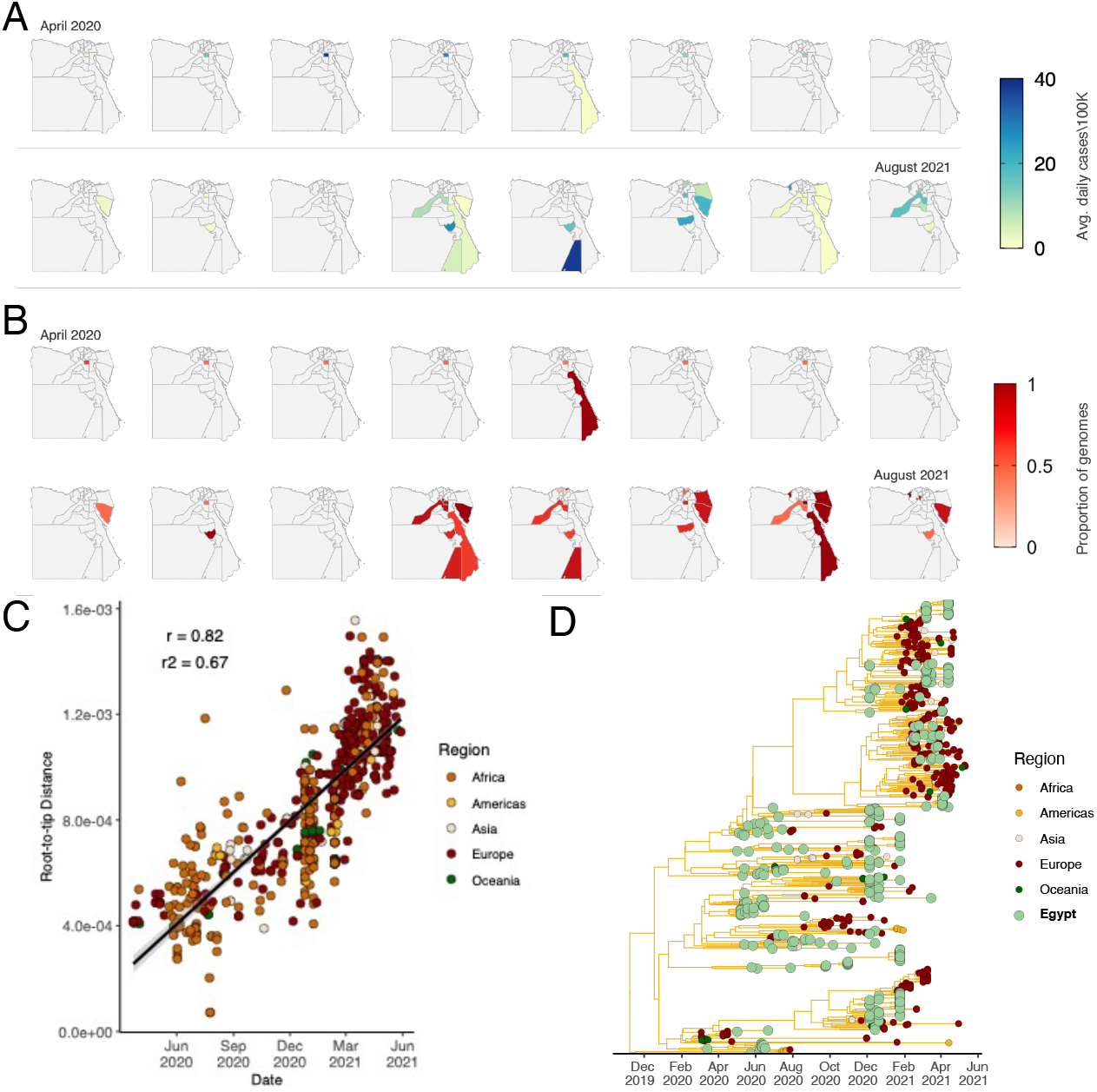
Sampling of the C.36 lineages in Egypt over time. A) Prevalence maps following the progression in the daily number of cases and, B) proportions of C.36.3 lineage samples per governorate in Egypt confirmed by RT-PCR from April 2020 to August 2021 C). Root-to-tip regression plot of the C.36 lineages. D) Maximum clade credibility phylogenetic tree including all the global sequences of C.36 lineages.

By July 2021, the C.36.3 lineage had evolved into the C.36.3.1 sublineage characterized by two additional Spike mutations: S477N and A845S (Figure 4C). The S477N mutation is known to increase the strength of ACE2 binding (Starr et al., 2020) while the biological consequence of the A845S mutation remains unknown.

**Fig 4.**
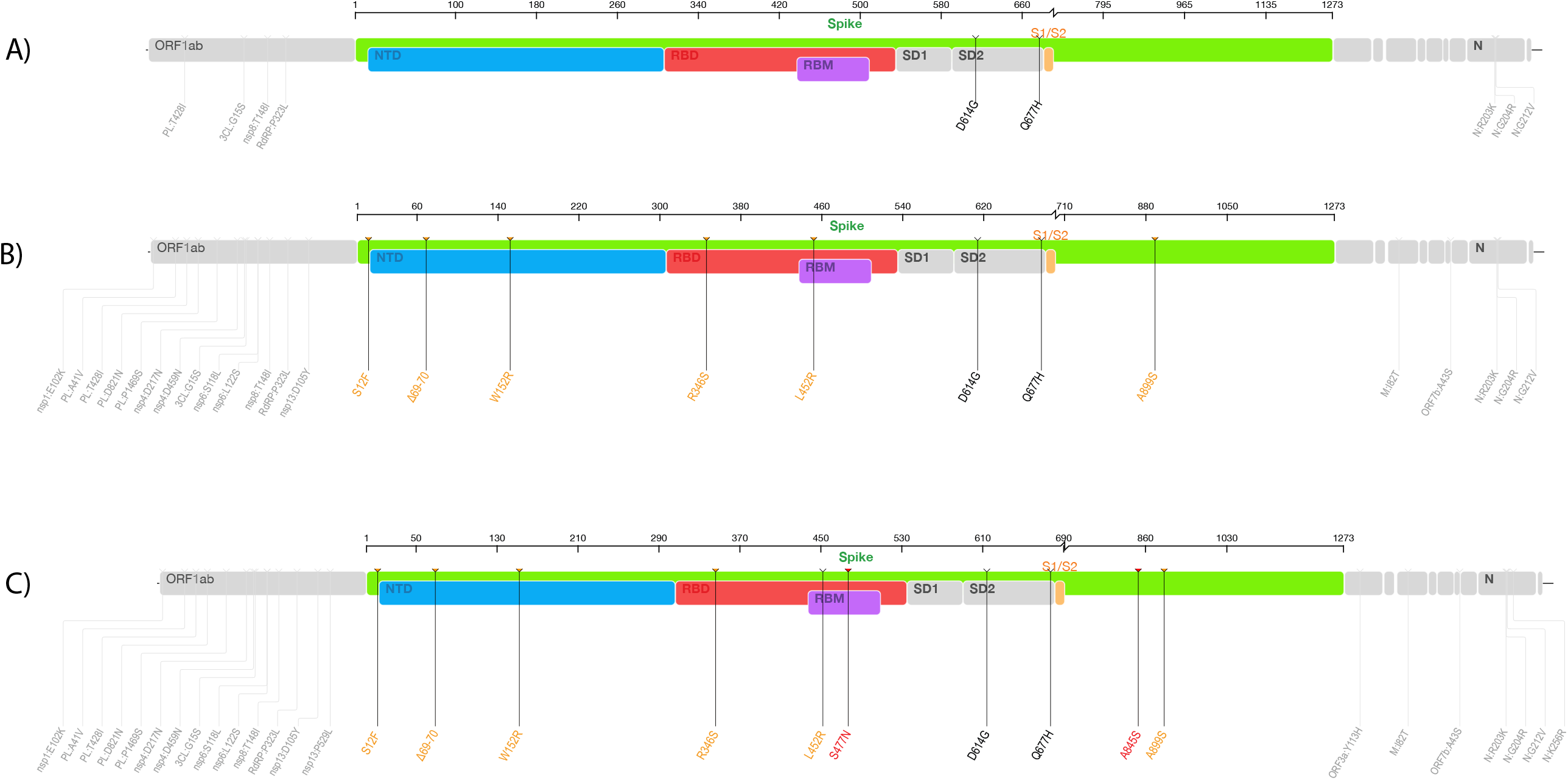
Genomic maps showing lineage defining mutations of the C.36 lineages. A) Lineage defining mutations of the C.36 lineage, B) lineage defining mutations of the C.36.3 sub-lineage, and C) lineage defining mutations in the C.36.3.1 sub-lineage. Additional Spike gene mutations in the C.36.3 lineage (n=6) are noted in orange and mutations unique to the C.36.3.1 (n=2) in red.

## Discussion

In this study, we have assessed the genomic epidemiology of SARS-CoV-2 in Egypt. We show that the virus was introduced into Egypt multiple times, resulting in the identification of at least 36 unique viral lineages detected in the population. Continued surveillance throughout 2020 and 2021 further illuminated the population dynamics of SARS-CoV-2 lineages circulating in Egypt. Specifically, we show that the first wave of the pandemic in Egypt was largely dominated by lineage B viruses (particularly B.1), while the second and third waves were dominated by C-like lineages (especially C.36). Genomic epidemiology allows for the identification of emerging variants and enhances our understanding of how the mutations carried by individual virus lineages contribute to the relative adaptive advantages they display in relation to other viruses with which the cocirculate. This can in turn inform the implementation of effective pandemic control measures and the development of vaccines and therapeutics.

The C.36 lineage was first introduced to Egypt around March 2020, potentially from Australia, and persisted through all three waves during which it accounted for the majority of the recorded cases. Its evolution and accumulation of several adaptive mutations that likely enabled it to escape natural and vaccine induced immunity highlights the need for sustained genomic surveillance to inform public health measures that are intended to control the pandemic. Like many other countries in Africa, there is a substantial burden of non-COVID19 disease in Egypt (El-Saadani et al., 2021; El-Sokkary et al., 2021): leading to large segments of the population having a reduced immunocompetence and might therefore favor the emergence of novel variants (Weigang et al., 2021). Prioritization of vaccine initiatives among the elderly and immunosuppressed groups is crucial to prevent continued evolution of variants that are presently circulating within the country.

The heterogeneous nature of the epidemic in Egypt also creates a favorable environment for recombination events that have the potential to produce viruses with enhanced adaptive and pathogenic properties. Such interlineage recombinants of SARS-CoV-2 with confirmed onward transmission have been reported in the UK, particularly from recombination with the Alpha variant that has also been detected in Egypt (Jackson et al., 2021). This demonstrates the potential of SARS-CoV-2 to evolve in similar settings and therefore the need for continued close monitoring of recombination events in Egypt. Additionally, public health measures including rapid response to confirmed cases, contact tracing and enforcement of self-isolation are crucial to prevent further evolution of the virus.

The evolution of the C.36 into various sub-lineages, some of interest reiterates a single important message: that no SARS-CoV-2 lineages should be ignored. This message is reinformed with the recent emergence of the Omicron variant suspected to have emerged from cryptic circulation of B.1.1. lineage viruses from late 2020 in Southern Africa (Viana et al., 2021). Furthermore, the continued epidemiological interaction of Egypt with the West as evidenced by the exportation events implied by our analyses (most of which exports were to Europe) highlights the need for a concerted united front against the pandemic between the rest of the world and Africa.

The main limitations of this study were (i) that most of the analysed genomic sequences where from a single Egyptian city (i.e. Cairo) and (ii) that the volume of sequencing was lower during the second and third waves that it was during the first wave. These two limitations greatly complicated the reconstruction of viral transmission dynamics within the country. There is a need for a more robust surveillance strategy in Egypt and this need to be met with sustainable funding to support genomic epidemiology initiatives to ensure that representative samples are collected across the country at regular time-intervals. A final important limitation of our study also is that the epidemiological data needed to contextualize much of the genomic sequencing data was absent: for example, patient travel details that might have been useful to corroborate the inferred importation and exportation events were not generally available. Epidemiological data of this sort is vital because the slow evolutionary rate of the virus at the onset of the pandemic meant that genetic linkages between sequences in different countries does not categorically prove that viral movements occurred between those countries.

In summary, by combining genomic and epidemiological data, we have been able to illuminate the viral dynamics behind the COVID19epidemic in Egypt. Particularly, we have shown that if not controlled, non-VOC lineages can evolve rapidly within a single country to acquire adaptive and pathogenic properties necessary to sustain community transmission and affect the prognosis of SARS-CoV-2 infections.

## Materials and methods

### Ethics statement

Ethical approval was obtained from the Ethics Committee of the National Research Centre, Giza, Egypt protocol number 14155, on 22 March 2020.

### Epidemiological data

Counts of daily cases and deaths analyzed were retrieved from the online data repository, Our World in Data (OWID) (Our World in Data, 2020). The effective reproductive number (R_e_) dataset was retrieved from the online repository, covid-19-Re/dailyRe-Data (Huisman, 2020; Huisman et al., 2021). Both epidemiological datasets analyzed were limited to data collected between 01 March 2020 and 15 July 2021 in line with genomic dataset.

### qPCR analyzed samples

The study examined 1,076 nasopharyngeal (NP) swab samples obtained from visa applicants, tourists, and patients in isolation throughout Egypt from 1 January 2020 to 1 August 2021. Briefly, samples were collected with flocked nasopharangeal swabs and immersed in viral transport medium for delivery to local governorate laboratories for testing with SARS-CoV2 PCR protocols. The chemagic 360 equipment (PerkinElmer Inc) was used to extract nucleic acid from the clinical samples. The local laboratories governorates performed the initial SARS-CoV-2 qPCR screening using the Viasure SARS-CoV-2 Real-Time PCR Detection Kit (Certest Biotec SL, Spain). The Applied Biosystems QuantStudio 7 Flex Real-Time PCR Detection equipment with QuantStudio Real-Time PCR software v.1.3 (Thermo Fisher Scientific, Waltham, MA) was used for RT-PCR. The samples were then sent to the central public health laboratory (CPHL) under optimal storage conditions. All accepted samples had a cycle threshold (CT) value less than 35. Positive SARS-CoV-2 samples were confirmed at CPHL using the Cobas 6800 system (Roche Holding AG).

### Mutation screening assays

The C36.3 lineage was confirmed by the presence of the amino acid deletion HV69-70 and substitution L452R in the Spike gene using a multiplex real-time RT-PCR assay and the VIASURE (Certest Biotec SL, Spain) SARS-CoV-2 detection detection kit on the Applied Biosystems QuantStudio 7 Flex Real-Time PCR detection equipment and QuantStudio Real-Time PCR software v.1.3 (Thermo Fisher Scientific, Waltham, MA). Additionally, Alpha variant mutations were confirmed using the SNPsig-Variplex SARS-CoV-2 SNP genotyping panel for the Alpha variant, Delta lineage mutations were identified using Thermo Fisher Scientific (Thermo Fisher Scientific, Waltham, MA) Applied Biosystems TaqMan SARS-CoV-2 Mutation Panel. Other SNP combinations such as 501Y, 484K, L452R and wild-types 501N, 484E, and 452L; deletion of amino acids H69 and V70, N501Y, E484K, K417N; k417T, P681R, P681H, L452R, K417N; and Q677H were identified with relevant kits following the manufacturer’s instructions.

### SARS-CoV-2 whole-genome sequencing

Ribosomal RNA was removed from previously extracted RNA using the ribo zero assay. This was followed by double stranded cDNA synthesis using the Truseq Stranded Total RNA (Illumina, San Diego, CA) per the manufacturer’s conditions and sequenced on the Illumina MiSeq sequencing platform. Strain typing assays (NGS) were cross-validated using a set of 250 SARS-CoV-2-positive samples and 50 SARS-CoV-2-negative samples.

### Genomic analysis and strain typing

Raw short read sequence data was assembled and analyzed using Dragen for SARS-CoV-2 variant detection (for the Truseq assay). Adequate sequencing required coverage of strain-distinguishing areas of the open reading frame 1a (ORF1a), S, N, and ORF8 genes to a depth of at least 100x. All but two sequences in this series yielded adequate strain-typeable sequences, and no sample contained a mixed population of viruses. Lineages and clades were assigned using the dynamic lineage classification tool Phylogenetic Assignment of Named Global Outbreak LINeages (Pangolin) (Rambaut et al., 2020) and NextStrain (Hadfield et al., 2018).

### Phylogenetic Analysis

All available Egyptian SARS-CoV-2 sequences on the GISAID database were retrieved (n=976; date of access: August 9, 2021) and analyzed against a globally representative dataset of SARS-CoV-2 isolates (n=6,561) to track and monitor the spatial temporal evolution of the virus across time and space. Global reference sequences were chosen based on their previous inclusion in the global and African phylogenetic builds (date of access: August 9, 2021) of the NextStrain platform (https://nextstrain.org/ncov/gisaid/global). Additionally, due to the importance of the C.36 lineage and sub-lineages in Egypt we included all globally sampled C.36 and sub-lineages on the GISAID database were included (date of access: August 9, 2021). This sampling strategy ensures that sufficient global viral diversity is included in the downstream analysis.

Briefly, sequences were aligned using NextAlign (Aksamentov and Neher, 2020) to obtain a good codon alignment of the sequences. A maximum likelihood tree topology was inferred from the resulting alignment in IQTREE 2 (Minh et al., 2020) using the General Time Reversible model of nucleotide substitution (Simon, 1986) and a total of 100 bootstrap replicates to infer support for branches in the resulting tree topology using Booster (Lemoine et al., 2018). Firstly, the topology was assessed in TemPest (Rambaut et al., 2016) to remove any potential outlier sequences. then, the tree topology was then transformed into a dated tree topology where the branches correspond to units of calendar time. This was achieved in TreeTime (Sagulenko et al., 2018) with the application of a strict molecular clock assumption at a rate of 8×10^−4^ mutations/site/year.

The dated SARS-CoV-2 phylogeny was then used along with the associated metadata that was obtained from GISAID to map discrete geographical locations to each of the tips and infer locations for internal nodes. This was done using the Mugration package extension of TreeTime. A custom python script was used to count the number of discrete changes occurring as we transcend the topology from the root towards the tips. In essence, this provides a crude estimate of the number of viral exchanges between Egypt and the rest of the world as well as when these exchanges occurred.

## Data Availability

All data produced are available online on GISAID

http://www.gisaid.org

## Acknowledgments

The authors acknowledge all members of Central Public Health Laboratories (CPHL) of the Ministry of Health and Population and all scientific researchers at the Center of Scientific Excellence for Influenza Virus; National Research Centre; Egypt.

## Author Contributions

Conceptualization: WHR, MKK, TDO, JES, EW, HT, SSA and A.M.K; Writing—original draft: WHR, MKK, JES, EW, HT, DPM, MM, SSA and AMK; Sampling: WHR, MKK, SSA; Sequencing: WHR, MKK, SSA and AMK; Writing—review and editing: WHR, TDO, DPM, AMK, JES, EW, HT; Visualization: HT, JES, EW, AMK, AAK; project administration: WHR, AN, NEG, MRG, RES, MF, HAE, AM, RG, MH, MAF, AE, MAA; funding acquisition: AN, NEG, MRG, RES, MF, HAE, AM, RG, MH, MAF, AE, MAA; All authors have read and agreed to the final version of the manuscript

## Figure Legends

Figure S1: Global spread of the C.36.3 lineage. The lineage has predominantly been circulating in Egypt

Figure S2. Import and export analysis of SARS-CoV-2 in Egypt. A) Viral introductions into Egypt from other continents. B) Viral exports from Egypt to other continents.

Table S1: Nextclade report of all Egypt genomes (n=976)

Table S2: GISAID acknowledgements

