## Supplementary Table 2 for "SARS-CoV-2 Genetic diversity and lineage dynamics of in Egypt"

We gratefully acknowledge the following Authors from the Originating laboratories responsible for obtaining the specimens, as well as the Submitting laboratories where the genome data were generated and shared via GISAID, on which this research is based.

All Submitters of data may be contacted directly via www.gisaid.org

Authors are sorted alphabetically.

| Accession ID | Originating Laboratory | Submitting Laboratory | Authors |
|---|---|---|---|
| EPI_ISL_1167045 | "Dr. Andrija Stampar" Teaching Institute of Public Health, Department of Clinical Microbiology | Istituto di Genomica Applicata | Davide Scaglione; Eleonora Paparelli; Fedrica Cattonaro; Gabriele Magris; Irena Jurman; Jasmina Vranes; Michele Morgante; Slobodanka Radovic; Vera Vendramin |
| EPI_ISL_993056, EPI_ISL_994661, EPI_ISL_996004 | "Dr. Andrija Stampar" Teaching Institute of Public Health, Department of Clinical Microbiology | Institute of Applied Genomics | Federica Cattonaro; Jasmina Vranes; Michele Morgante |
| EPI_ISL_2438647 | "National Center of Expertise" CSEC MH RK in Kostanay region | Reference laboratory for the control of viral infections | Aidar Usserbayev; Aknur Mutaliyeva; Andrey Komissarov; Artem Fadeev; Azamat Kenessov; Bekzhan Maikotov; Gaukhar Nussupbayeva; Madina Tleubergenova; Maria Pisareva; Nazym Tleumbetova |
| EPI_ISL_2438606 | "National Center of Expertise" CSEC MH RK in Kyzylorda Region | Reference laboratory for the control of viral infections | Aidar Usserbayev; Aknur Mutaliyeva; Andrey Komissarov; Artem Fadeev; Azamat Kenessov; Bekzhan Maikotov; Gaukhar Nussupbayeva; Madina Tleubergenova; Maria Pisareva; Nazym Tleumbetova |
| EPI_ISL_2438655 | "National Center of Expertise" CSEC MH RK in West Kazakhstan Region | Reference laboratory for the control of viral infections | Aidar Usserbayev; Aknur Mutaliyeva; Andrey Komissarov; Artem Fadeev; Azamat Kenessov; Bekzhan Maikotov; Gaukhar Nussupbayeva; Madina Tleubergenova; Maria Pisareva; Nazym Tleumbetova |
| EPI_ISL_2840043 | "Sangius" LLP | Reference laboratory for the control of viral infections | Aidar Usserbayev; Aknur Mutaliyeva; Andrey Komissarov; Artem Fadeev; Azamat Kenessov; Bekzhan Maikotov; Gaukhar Nussupbayeva; Madina Tleubergenova; Maria Pisareva; Nazym Tleumbetova |
| EPI_ISL_632310 | 1-Laboratory of Microbiology, National Reference Lab, Charles Nicolle Hospital; 2-University of Tunis ElManar, Faculty of Medicine of Tunis, LR99ES09, Tunis, Tunisia | 1-Clinical and Experimental Pharmacology Lab, LR16SP02, National Center of Pharmacovigilance, University of Tunis El Manar, Tunis, Tunisia. 2-Neurodegenerative diseases and psychiatric troubles, LR18SP03, Razi Hospital, University of Tunis El Manar, Tunis, Tunisia. 3- Ministry of Health, National Observatory of New and Emerging Diseases, 1006, Tunis, Tunisia | Alia Ben Kahla; Gaies Emna; Ilhem Boutiba-Ben Boubaker; Imen Kacem; Imen Mkada; Jalila Ben Khelil; Maher Kharrat; Mouna Ben Sassi; Mouna Safer; Nissaf Ben Alaya; Riadh Daghfous; Riadh Gouider.; Salma Abid; Sameh Trabelsi; Sana Ferjani; Soumaya Rammeh |
| EPI_ISL_635059, EPI_ISL_707698, EPI_ISL_733500, EPI_ISL_763065, EPI_ISL_794736 | 1-Laboratory of Microbiology, National Reference Lab, Charles Nicolle Hospital; 2-University of Tunis ElManar, Faculty of Medicine of Tunis, LR99ES09, Tunis, Tunisia | 1-Clinical and Experimental Pharmacology Lab, LR16SP02, National Center of Pharmacovigilance, University of Tunis El Manar, Tunis, Tunisia. 2-Neurodegenerative diseases and psychiatric troubles, LR18SP03, Razi Hospital, University of Tunis El Manar, Tunis, Tunisia. 3- Ministry of Health, National Observatory of New and Emerging Diseases, 1006, Tunis, Tunisia | Alia Ben Kahla; Alia BenKahla; Asma Ferjani; Awatef El Moussi; Gaies Emna; Guedi Ali Barreh; Habiba Ben Romdhane; Hanen El Jebari; Ilhem Boutiba-Ben Boubaker; Imen Kacem; Imen Mkada; Ines Mdini; Jalila Ben Khelil; Maher Kharrat; Mouna Ben Sassi; Mouna Safer; Nissaf Ben Alaya; Riadh Daghfous; Riadh Gouider.; Rouaa Ben Othman; Salma Abid; Sameh Trabelsi; Sana Ferjani; Sarra Chamman; Souissi Amira; Soumaya Rammeh; Zaineb Hamzaoui |
| EPI_ISL_492072 | 1. ViroGenetics - BSL3 Laboratory of Virology, Małopolska Centre of Biotechnology, Jagiellonian University; 2. II Department of Internal Medicine, Faculty of Medicine, Jagiellonian University Medical College; 3. Narodowy Instytut Zdrowia Publicznego – Państwowy Zakład Higieny (NIZP-PZH) | 1. ViroGenetics - BSL3 Laboratory of Virology, Małopolska Centre of Biotechnology, Jagiellonian University; 2. II Department of Internal Medicine, Faculty of Medicine, Jagiellonian University Medical College; 3. Narodowy Instytut Zdrowia Publicznego – Państwowy Zakład Higieny (NIZP-PZH). | Agnieszka Kołakowska-Kulesza; Aleksandra A. Zasada; Aleksandra Milewska; Ewelina Hallman-Szelińska; Katarzyna Owczarek; Katarzyna Pancer; Katarzyna Zacharczuk; Krzysztof Pyrć; Magdalena Rzeczkowska; Marek Sanak; Natalia Wolaniuk; Paweł P Łabaj; Tomasz Wołkowicz; Wojciech Branicki |
| EPI_ISL_877029 | 1.AO Universitaria 'S. Giovanni di Dio e Ruggi D'Aragona, Scuola Medica Salernitana' Hospital / 2.UOC di Virologia e Microbiologia, Università della Campania 'L. Vanvitelli' / 3.AO Universitaria 'Federico II' Napoli Hospital / 4.AORN 'San Giuseppe Moscati' Avellino Hospital / 5.AO 'San Pio - presidio G. Rummo' Benevento Hospital / 6.AO 'Sant'Anna e San Sebastiano' Caserta Hospital / 7.PO 'Maria Santissima Addolorata' Eboli Hospital / 8.Biogem Istituto di Ricerche Genetiche | 1. Genome Research Center for Health (CRGS) / 2. Laboratory of Molecular Medicine and Genomics(LMMGe) / 3. Center for Research in Pure and Applied Mathematics (CRMPA) | Alessandro Weisz (Corresponding Author); Alessia Cossu; Aniello Gentile; Annamaria Salvati; Antonello Saccomanno; Arnolfo Petruzziello; Assunta Sellitto; Carlo Ferravante; Domenico Memoli; Domenico Palumbo; Elena Alexandrova; Emilia Vaccaro; Francesca Marciano; Francesca Rizzo (Corresponding Author); Gianluigi Franci; Giorgio Giurato (Corresponding Author); Giovanni Nassa; Giovanni Pecoraro; Giuseppe Fenza; Giuseppe Portella; Gregorio Goffredi; Ilaria Terenzi; Jessica Lamberti; Maddalena Schioppa; Maria Grazia Foti; Maria Landi; Marianna Scrima; Mariarosaria Ingino; Massimiliano Galdiero; Maurizio Fumi; Michele Caraglia; Michele Cennamo; Oriana Strianese; Pasquale Pagliano; Rita Greco; Roberta Tarallo; Sonia Amabile; Teresa Rocco; Valeria Mirici Cappa; Vincenzo Rocco; Viola Melone; Vittoria Letizia; Ylenia D'Agostino |
| EPI_ISL_1361596, EPI_ISL_1547392 | 1.AO Universitaria 'S. Giovanni di Dio e Ruggi D'Aragona, Scuola Medica Salernitana' Hospital / 2.UOC di Virologia e Microbiologia, Università della Campania 'L. Vanvitelli' / 3.AO Universitaria 'Federico II' Napoli Hospital / 4.AORN 'San Giuseppe Moscati' Avellino Hospital / 5.AO 'San Pio - presidio G. Rummo' Benevento Hospital / 6.AO 'Sant'Anna e San Sebastiano' Caserta Hospital / 7.PO 'Maria Santissima Addolorata' Eboli Hospital / 8.Biogem Istituto di Ricerche Genetiche / 9. U.O.C. di Genetica Medica e di Laboratorio A.O.R.N., Azienda Ospedaliera di Rilievo Nazionale Antonio Cardarelli, Napoli / 10. Centro di riferimento Oncologico della Basilicata (IRCCS-CROB), Rionero in Vulture (PZ) / 11. Presidio Ospedaliero di Agropoli, Agropoli (SA). | 1. Genome Research Center for Health (CRGS) / 2. Laboratory of Molecular Medicine and Genomics(LMMGe) / 3. Center for Research in Pure and Applied Mathematics (CRMPA) | Alessandro Weisz; Alessandro Weisz (Corresponding Author); Alessia Cossu; Andreina Baj; Aniello Gentile; Annamaria Salvati; Antonello Saccomanno; Arnolfo Petruzziello; Assunta Sellitto; Caivano Antonella; Carlo Ferravante; D'Auria Fiorella; De Luca Luciana.; Domenico Memoli; Domenico Palumbo; Edmondo Adorisio; Elena Alexandrova; Emilia Vaccaro; Fausto Sessa; Francesca Marciano; Francesca Rizzo; Francesca Rizzo (Corresponding Author); Francesco Curcio; Gianluigi Franci; Giorgio Dirani; Giorgio Giurato; Giorgio Giurato (Corresponding Author); Giovanni Nassa; Giovanni Pecoraro; Giuseppe Fenza; Giuseppe Portella; Giusy Giordano; Gregorio Goffredi; Ilaria Terenzi; Jessica Lamberti; Maddalena Schioppa; Marcello Ametrano; Maria Grazia Foti; Maria Landi; Marianna Scrima; Mariano Fiorenza; Mariarosaria Ingino; Marina Tarsitano; Massimiliano Chetta; Massimiliano Galdiero; Maurizio Fumi; Michela Iacobellis; Michele Caraglia; Michele Cennamo; Morena D'Avenia; Oriana Strianese; Pasquale Pagliano; Rita Greco; Roberta Tarallo; Rosanna Piluscio; Silvia Zanoli; Simona Semprini; Sonia Amabile; Stefania Marzinotto; Teresa Rocco; Valeria Mirici Cappa; Vincenzo Rocco; Viola Melone; Vittoria Letizia; Vittorio Sambri; Ylenia D'Agostino |
| EPI_ISL_2140715 | 2 Military Hospital wc MAA | NHLS/UCT | Arash Iranzadeh; Bruna Galvao; Carolyn Williamson; Deelan Doolabh; Diana Hardie; Innocent Mudau; Kruger Marais; Lynn Tyers; Marvin Hsiao; Stephen Korsman |
| EPI_ISL_2547411 | 2 Military Hospital wc MAA | National Health Laboratory Service/University of Cape Town (NHLS/UCT) | Arash Iranzadeh; Bruna Galvao; Carolyn Williamson; Deelan Doolabh; Diana Hardie; Gert Marais; Innocent Mudau; Lynn Tyers; Marvin Hsiao; Stephen Korsman |
| EPI_ISL_2646244 | ACM Bio | Cerba lab | Aude Lessenne; Bénédicte Roquebert; Emmanuel Lecorche; Kader Merah; Laura Verdurme; Patrice Herisson; Sabine Trombert-Paolantoni; Stéphanie Haïm-Boukobza; Thierry Collin |
| EPI_ISL_2621213 | ADILAB | Universidad Nacional de Colombia - Laboratorio Genómico One Health | Andres F. Cardona-Rios; Carlos Franco-Muñoz; Carolina Muñoz-Arango; Celeny Ortiz; Daniel O. Maldonado-Perez; Diego A. Álvarez-Díaz; Hector Alejandro Ruiz-Moreno; Idabely Betancur Ortiz; Jorge E. Osorio; Juan P. Hernandez-Ortiz; Karl A Ciuoderis; Katherine Laiton-Donato; Laura Silvana Perez; Lina M. Hurtado; Marcela Mercado-Reyes; Maria Angélica Maya; Maria Stella López; Rita Almanza Payares; Sandra Ines Cano; Simón Villegas Velásquez |
| EPI_ISL_2617039, EPI_ISL_2617056 | AFRICA_CDC - Angola (Ship 12) | CERI, Centre for Epidemic Response and Innvoation, Stellenbosch University and KRISP, KZN Research Innovation and Sequencing Platform, UKZN. | Afonso P; David K; Emmanuel SJ; Freitas RH; Giandhari J; Inglês L; Lutucuta S; Miranda J; Morais J; Mufinda M; Naidoo Y; Neto Z; Paulo A Carralero RR Paixão JP; Pereira A; Pillay S; Tegally H; Wilkinson E; de Oliveira T |
| EPI_ISL_918437, EPI_ISL_918438 | AIID | Irish Coronavirus Sequencing Consortium-Teagasc Grange | Aljandro Abner Garcia Leon; Calum Walsh; Fiona Crispie; Gabriel Gonzalez; John Kenny; Matthew McCabe; Michael Carr; Patrick Mallon; Paul Cotter |
| EPI_ISL_2018254 | AK State Public Health Lab, State Health Department | Centers for Disease Control and Prevention Division of Viral Diseases, Pathogen Discovery | Alison Laufer Halpin; Ben L. Rambo-Martin; Clinton R. Paden; Dakota Howard; Darlene Wagner; Dave Wentworth; Dhwani Batra; Jasmine Padilla; Justin Lee; Katie Dillon; Krista Queen; Kristen Knipe; Kristine Lacek; Mark Burroughs; Matthew Schmerer; Mili Sheth; Peter Cook; Sam Shepard; Sarah Nobles; Shoshona Le; Suxiang Tong; Vivien Dugan; Yvette Unoarumhi |
| EPI_ISL_2608418 | AKUH Laboratories, Karachi, Pakistan | AKUH Laboratories, Karachi, Pakistan | Akbar Kanji; Asghar Nasir; Rumina Hasan; Safina A. Razzak; Sayed Ali Raza; Zahra Hasan |
| EPI_ISL_2383982 | AL Dept. of Public Health Bureau of Clinical Laboratories | Centers for Disease Control and Prevention Division of Viral Diseases, Pathogen Discovery | Alison Laufer Halpin; Ben L. Rambo-Martin; Clinton R. Paden; Dakota Howard; Darlene Wagner; Dave Wentworth; Dhwani Batra; Jasmine Padilla; Justin Lee; Katie Dillon; Krista Queen; Kristen Knipe; Kristine Lacek; Mark Burroughs; Matthew Schmerer; Mili Sheth; Peter Cook; Sam Shepard; Sarah Nobles; Shoshona Le; Suxiang Tong; Vivien Dugan; Yvette Unoarumhi |
| EPI_ISL_2868624 | ALBANY MEDICAL CENTER | Wadsworth Center, New York State Department of Health | Alexis Russell; Catharine Prussing; Daryl M. Lamson; Erasmus Schneider; Erica Lasek-Nesselquist; John Kelly; Jonathan Plitnick; Kirsten St. George; Matthew Shudt; Melissa A Leisner; Navjot Singh |
| EPI_ISL_2493041 | AMPATH | CERI, Centre for Epidemic Response and Innvoation, Stellenbosch University and KRISP, KZN Research Innovation and Sequencing Platform, UKZN. | Emmanuel SJ; Giandhari J; Khan S; Lessells R; Mdlalose K; Naidoo Y; Pillay S; Ramphal U; Tegally H; Wilkinson E; York D; de Oliveira T |
| EPI_ISL_2086992, EPI_ISL_2087012, EPI_ISL_2087018 | AMPATH LABORATORIES | National Institute for Communicable Diseases of the National Health Laboratory Service | Amoako DG; Bhiman JN; Ismail A; Mahlangu B; Mohale T; Ntuli N; Scheepers C |
| EPI_ISL_2658283 | AREA DE SALUD ALAJUELITA | Inciensa, Instituto Costarricense de Investigación y Enseñanza en Nutrición y Salud | Adriana Godínez; Claudio Soto-Garita; Estela Cordero; Francisco Duarte; Hebleen Porras; José Luis Vargas; Mariela Gutiérrez & Joselyn Prado; Melany Calderón |
| EPI_ISL_3037812 | AREA DE SALUD BAGACES | Inciensa, Instituto Costarricense de Investigación y Enseñanza en Nutrición y Salud | Adriana Godínez; Caterina Guzmán; Claudio Soto-Garita; Estela Cordero; Francisco Duarte; Hebleen Porras; José Luis Vargas; Mariela Gutiérrez Joselyn Prado; Melany Calderón; Nazareth Ruiz & Adriana Bermúdez |
| EPI_ISL_3037817 | AREA DE SALUD BELEN-FLORES - CLINICA DR. JORGE VOLIO | Inciensa, Instituto Costarricense de Investigación y Enseñanza en Nutrición y Salud | Adriana Godínez; Caterina Guzmán; Claudio Soto-Garita; Estela Cordero; Francisco Duarte; Hebleen Porras; José Luis Vargas; Mariela Gutiérrez Joselyn Prado; Melany Calderón; Nazareth Ruiz & Sandra Mora Leitón |
| EPI_ISL_2272981 | AREA DE SALUD CARRILLO | Inciensa, Instituto Costarricense de Investigación y Enseñanza en Nutrición y Salud | Adriana Godínez; Claudio Soto-Garita; Estela Cordero; Francisco Duarte; Hebleen Porras; Joselyn Prado & Adriana Bemúdez Espinoza; José Luis Vargas; Mariela Gutiérrez; Melany Calderón |
| EPI_ISL_2658271 | AREA DE SALUD CHOMES - MONTEVERDE | Inciensa, Instituto Costarricense de Investigación y Enseñanza en Nutrición y Salud | Adriana Godínez; Claudio Soto-Garita; Estela Cordero; Francisco Duarte; Hebleen Porras; Joselyn Prado & Javier Zárate-León; José Luis Vargas; Mariela Gutiérrez; Melany Calderón |

| | | | |
|---|---|---|---|
| EPI_ISL_3037821 | AREA DE SALUD COTO BRUS | Inciensa, Instituto Costarricense de Investigación y Enseñanza en Nutrición y Salud | Adriana Godínez; Caterina Guzmán; Claudio Soto-Garita; Estela Cordero; Francisco Duarte; Hebleen Porras; José Luis Vargas; Mariela Gutiérrez Joselyn Prado; Melany Calderón; Nazareth Ruiz & María Fernanda Matamoros |
| EPI_ISL_2502748 | AREA DE SALUD DESAMPARADOS 2 (COOPESALUD) | Inciensa, Instituto Costarricense de InvestigaciOn y Enseñanza en NutriciOn y Salud | Adriana Godínez; Claudio Soto-Garita; Estela Cordero; Francisco Duarte; Hebleen Porras; Jose Luis Vargas; Joselyn Prado & Paola Sánchez-Traña; Mariela Gutierrez; Melany CalderOn |
| EPI_ISL_3026033 | AREA DE SALUD Desamparados 1 - CLINICA DR. MARCIAL FALLAS [GRIFO ALTO/Desamparados/San Jose] | Inciensa, Instituto Costarricense de Investigación y Enseñanza en Nutrición y Salud | Adriana Godínez; Caterina Guzmán; Claudio Soto-Garita; Estela Cordero; Francisco Duarte; Hebleen Porras; Joselyn Prado; José Luis Vargas; Mariela Gutiérrez; Melany Calderón; Nazareth Ruíz |
| EPI_ISL_3026022 | AREA DE SALUD EL GUARCO [EL GUARCO/CARTAGO] | Inciensa, Instituto Costarricense de Investigación y Enseñanza en Nutrición y Salud | Adriana Godínez; Caterina Guzmán; Claudio Soto-Garita; Estela Cordero; Francisco Duarte; Hebleen Porras; Joselyn Prado; José Luis Vargas; Mariela Gutiérrez; Melany Calderón; Nazareth Ruíz & Mónica Charpentier |
| EPI_ISL_3026027 | AREA DE SALUD FORTUNA [San Carlos/ALAJUELA] | Inciensa, Instituto Costarricense de Investigación y Enseñanza en Nutrición y Salud | Adriana Godínez; Caterina Guzmán; Claudio Soto-Garita; Estela Cordero; Francisco Duarte; Hebleen Porras; Joselyn Prado; José Luis Vargas; Mariela Gutiérrez; Melany Calderón; Nazareth Ruíz & Carolina Arrieta |
| EPI_ISL_2828000 | AREA DE SALUD GARABITO | Inciensa, Instituto Costarricense de Investigación y Enseñanza en Nutrición y Salud | Adriana Godínez; Claudio Soto-Garita; Estela Cordero; Francisco Duarte; Hebleen Porras; Joselyn Prado & María José Gómez; José Luis Vargas; Mariela Gutiérrez; Melany Calderón |
| EPI_ISL_2502755 | AREA DE SALUD GOICOECHEA 2 - CLINICA DR. JIMENEZ NUÑEZ | Inciensa, Instituto Costarricense de InvestigaciOn y Enseñanza en NutriciOn y Salud | Adriana Godínez; Claudio Soto-Garita; Estela Cordero; Francisco Duarte; Hebleen Porras; Jose Luis Vargas; Joselyn Prado & Alberto Madrigal-Vega; Mariela Gutierrez; Melany CalderOn |
| EPI_ISL_2502736 | AREA DE SALUD GUATUSO | Inciensa, Instituto Costarricense de InvestigaciOn y Enseñanza en NutriciOn y Salud | Adriana Godínez; Claudio Soto-Garita; Estela Cordero; Francisco Duarte; Hebleen Porras; Jose Luis Vargas; Joselyn Prado & Francisco ChacOn-Valverde; Mariela Gutierrez; Melany CalderOn |
| EPI_ISL_3026031 | AREA DE SALUD Guatuso [Guatuso/ALAJUELA] | Inciensa, Instituto Costarricense de Investigación y Enseñanza en Nutrición y Salud | Adriana Godínez; Caterina Guzmán; Claudio Soto-Garita; Estela Cordero; Francisco Duarte; Hebleen Porras; Joselyn Prado; José Luis Vargas; Mariela Gutiérrez; Melany Calderón; Nazareth Ruíz & Francisco Chacón |
| EPI_ISL_2272966 | AREA DE SALUD HATILLO - CLINICA DR. SOLON NUÑEZ | Inciensa, Instituto Costarricense de Investigación y Enseñanza en Nutrición y Salud | Adriana Godínez; Claudio Soto-Garita; Estela Cordero; Francisco Duarte; Hebleen Porras; Joselyn Prado & Roberto Brilla-Ferrer; José Luis Vargas; Mariela Gutiérrez; Melany Calderón |
| EPI_ISL_2502739 | AREA DE SALUD MATA REDONDA -HOSPITAL -CLINICA DR. MORENO CAÑAS | Inciensa, Instituto Costarricense de InvestigaciOn y Enseñanza en NutriciOn y Salud | Adriana Godínez; Claudio Soto-Garita; Estela Cordero; Francisco Duarte; Hebleen Porras; Jose Luis Vargas; Joselyn Prado & Carolina Duarte; Mariela Gutierrez; Melany CalderOn |
| EPI_ISL_3037838 | AREA DE SALUD MATA REDONDA-HOSPITAL - CLINICA DR. MORENO CAÑAS | Inciensa, Instituto Costarricense de Investigación y Enseñanza en Nutrición y Salud | & Nazareth Ruiz; Adriana Godínez; Caterina Guzmán; Claudio Soto-Garita; Estela Cordero; Francisco Duarte; Hebleen Porras; José Luis Vargas; Mariela Gutiérrez Joselyn Prado; Melany Calderón |
| EPI_ISL_2502720 | AREA DE SALUD MORAVIA | Inciensa, Instituto Costarricense de InvestigaciOn y Enseñanza en NutriciOn y Salud | Adriana Godínez; Claudio Soto-Garita; Estela Cordero; Francisco Duarte; Hebleen Porras; Jose Luis Vargas; Joselyn Prado & Mariel LOpez; Mariela Gutierrez; Melany CalderOn |
| EPI_ISL_3037845 | AREA DE SALUD PAQUERA | Inciensa, Instituto Costarricense de Investigación y Enseñanza en Nutrición y Salud | Adriana Godínez; Caterina Guzmán; Claudio Soto-Garita; Estela Cordero; Francisco Duarte; Hebleen Porras; José Luis Vargas; Mariela Gutiérrez Joselyn Prado; Melany Calderón; Nazareth Ruiz & Andre Moreno Carvajal |
| EPI_ISL_2502757 | AREA DE SALUD PARAISO-CERVANTES | Inciensa, Instituto Costarricense de InvestigaciOn y Enseñanza en NutriciOn y Salud | Adriana Godínez; Claudio Soto-Garita; Estela Cordero; Francisco Duarte; Hebleen Porras; Jose Luis Vargas; Joselyn Prado & MOnica Charpentier; Mariela Gutierrez; Melany CalderOn |
| EPI_ISL_2502750 | AREA DE SALUD PEREZ ZELEDON | Inciensa, Instituto Costarricense de InvestigaciOn y Enseñanza en NutriciOn y Salud | Adriana Godínez; Claudio Soto-Garita; Estela Cordero; Francisco Duarte; Hebleen Porras; Jose Luis Vargas; Joselyn Prado & Fabricio Aguilar; Mariela Gutierrez; Melany CalderOn |
| EPI_ISL_3037819 | AREA DE SALUD PEREZ ZELEDON | Inciensa, Instituto Costarricense de Investigación y Enseñanza en Nutrición y Salud | Adriana Godínez; Caterina Guzmán; Claudio Soto-Garita; Estela Cordero; Francisco Duarte; Hebleen Porras; José Luis Vargas; Mariela Gutiérrez Joselyn Prado; Melany Calderón; Nazareth Ruiz & Yendry Ramírez |
| EPI_ISL_2658272, EPI_ISL_2658275 | AREA DE SALUD SAN RAFAEL | Inciensa, Instituto Costarricense de Investigación y Enseñanza en Nutrición y Salud | Adriana Godínez; Claudio Soto-Garita; Estela Cordero; Francisco Duarte; Hebleen Porras; Joselyn Prado & Javier Zárate-León; José Luis Vargas; Mariela Gutiérrez; Melany Calderón |
| EPI_ISL_2828008 | AREA DE SALUD SAN RAMON | Inciensa, Instituto Costarricense de Investigación y Enseñanza en Nutrición y Salud | Adriana Godínez; Claudio Soto-Garita; Estela Cordero; Francisco Duarte; Hebleen Porras; José Luis Vargas; Mariela Gutiérrez & Joselyn Prado; Melany Calderón |
| EPI_ISL_2272993 | AREA DE SALUD SANTO DOMINGO | Inciensa, Instituto Costarricense de Investigación y Enseñanza en Nutrición y Salud | Adriana Godínez; Claudio Soto-Garita; Estela Cordero; Francisco Duarte; Hebleen Porras; Joselyn Prado & Margarita Lee-Lui; José Luis Vargas; Mariela Gutiérrez; Melany Calderón |
| EPI_ISL_2502744 | AREA DE SALUD SIQUIRRES | Inciensa, Instituto Costarricense de InvestigaciOn y Enseñanza en NutriciOn y Salud | Adriana Godínez; Claudio Soto-Garita; Estela Cordero; Francisco Duarte; Hebleen Porras; Jose Luis Vargas; Joselyn Prado & Karla LOpez; Mariela Gutierrez; Melany CalderOn |
| EPI_ISL_2103384, EPI_ISL_2272958, EPI_ISL_2272977 | AREA DE SALUD SIQUIRRES | Inciensa, Instituto Costarricense de Investigación y Enseñanza en Nutrición y Salud | Adriana Godínez; Caterina Guzmán; Claudio Soto-Garita; Estela Cordero; Francisco Duarte; Hebleen Porras; Joselyn Prado; Joselyn Prado & Ileana Chaves-Peraza; José Luis Vargas; Mariela Gutiérrez; Melany Calderón; Nazareth Ruiz & Ileana Chaves-Peraza |
| EPI_ISL_3037825 | AREA DE SALUD TURRIALBA-JIMENEZ | Inciensa, Instituto Costarricense de Investigación y Enseñanza en Nutrición y Salud | Adriana Godínez; Caterina Guzmán; Claudio Soto-Garita; Estela Cordero; Francisco Duarte; Hebleen Porras; José Luis Vargas; Mariela Gutiérrez Joselyn Prado; Melany Calderón; Nazareth Ruiz & Mónica Charpentier |
| EPI_ISL_2502758 | AREA DE SALUD ZAPOTE-CATEDRAL - CLINICA DR. CARLOS DURAN | Inciensa, Instituto Costarricense de InvestigaciOn y Enseñanza en NutriciOn y Salud | Adriana Godínez; Claudio Soto-Garita; Estela Cordero; Francisco Duarte; Hebleen Porras; Jose Luis Vargas; Joselyn Prado & Marco Cháves-Otárola; Mariela Gutierrez; Melany CalderOn |
| EPI_ISL_1067612 | AREA DE SALUD ZAPOTE-CATEDRAL - CLINICA DR. CARLOS DURAN | Inciensa, Instituto Costarricense de InvestigaciOn y Enseñanza en NutriciOn y Salud | Adriana Godínez; Caterina Guzmán; Claudio Soto-Garita; Estela Cordero; Francisco Duarte; Hebleen Porras; Melany Calderón; Nazareth Ruiz & Melissa Carazo-Gutiérrez |
| EPI_ISL_2695444 | ARS Algarve - Laboratorio Laura Ayres | Instituto Nacional de Saude (INSA) | Borges et al |
| EPI_ISL_517616, EPI_ISL_518802 | Academic Hospital Paramaribo | Erasmus Medical Center | Bas Oude Munnink; Dion Gajadin; Ed Ijzerman; Emmanuelle Munger; Gary Gummels; Ingrid Krishnadath; Lycke Woittiez; Marion Koopmans; Mireille Van de Veer; Princes Wongsowidjojo; Radjesh Ori; Rohma Banwari; Stephen Vreden |
| EPI_ISL_2833781 | Adan Hospital | Kuwait Cancer Control Center | Mona Alateeqi; Shakir Bahzad |
| EPI_ISL_1445665, EPI_ISL_1650426, EPI_ISL_1689082, EPI_ISL_1702837, EPI_ISL_1737563, EPI_ISL_1835211, EPI_ISL_1837090, EPI_ISL_1837760, EPI_ISL_2040167, EPI_ISL_2040637, EPI_ISL_2040648, EPI_ISL_2040827, EPI_ISL_2041273, EPI_ISL_2042434, EPI_ISL_2090019, EPI_ISL_2090085, EPI_ISL_2146197, EPI_ISL_2148186, EPI_ISL_2148720, EPI_ISL_2148847, EPI_ISL_2150374, EPI_ISL_2158859, EPI_ISL_2159079, EPI_ISL_2159334, EPI_ISL_2180676, EPI_ISL_2187388, EPI_ISL_2201288, EPI_ISL_2202065, EPI_ISL_2203267, EPI_ISL_2243536, EPI_ISL_2280370, EPI_ISL_2368234, EPI_ISL_2370070, EPI_ISL_2421780, EPI_ISL_2489181, EPI_ISL_2489215, EPI_ISL_2528269, EPI_ISL_2600307, EPI_ISL_2687055, EPI_ISL_2687316, EPI_ISL_2687343, EPI_ISL_2709524 | | | |
| see above | Aegis Sciences Corporation | Centers for Disease Control and Prevention Division of Viral Diseases, Pathogen Discovery | Adrian Paskey; Alec Vest; Benjamin Rambo-Martin; Christopher Gulvick; Clinton R. Paden; Cyndi Clark; Dakota Howard; Darlene Wagner; Dhwani Batra; Dillon Nall; Duncan MacCannell; Ethan Sanders; Holly Houdeshell; Jason Caravas; Kara Moser; Matthew Hardison; Matthew Schmerer; Ola Kvalvaag; Patrick Campbell; Peter W. Cook; Rob Case; Scott Sammons; Shatavia Morrison; Shaun Westlund; Vikramsinha Ghorpade; Yvette Unoarumhi |
| EPI_ISL_2617050 | Africa_CDC - Malawi P4 S2 | CERI, Centre for Epidemic Response and Innvoation, Stellenbosch University and KRISP, KZN Research Innovation and Sequencing Platform, UKZN. | Auld A; Chilima B; Chiwaula M; Emmanuel SJ; Giandhari J; Kaba M; Kampira E; Kasambara W; Kim L; Lessells R; Maida A; Mvula B; Mwangomba W; Naidoo Y; Panja L; Pillay S; Tegally H; Wadonda N; Wilkinson E; de Oliveira T |
| EPI_ISL_2617090 | Africa_CDC - Malawi P6 S2 | CERI, Centre for Epidemic Response and Innvoation, Stellenbosch University and KRISP, KZN Research Innovation and Sequencing Platform, UKZN. | Auld A; Chilima B; Chiwaula M; Emmanuel SJ; Giandhari J; Kaba M; Kampira E; Kasambara W; Kim L; Lessells R; Maida A; Mvula B; Mwangomba W; Naidoo Y; Panja L; Pillay S; Tegally H; Wadonda N; Wilkinson E; de Oliveira T |
| EPI_ISL_2360250, EPI_ISL_2360251 | Afzalipoor Hospital | National Influenza Center | A Nejati; F Ajaminejad and T Mokhtari Azad; J Yavarian; K Sadeghi; N Ghavvami; NZ Shafiei Jandaghi; V Salimi |
| EPI_ISL_528935 | Agenzia di Tutela della Salute di Bergamo | Istitituto Zooprofilattico Sperimentale dell'Abruzzo e Molise "G.Caporale" | Ancora M; Cammà C; Curini V; Di Domenico M; Di Pasquale A; Lorusso A; Mangone I; Marcacci M; Puglia I; Rinaldi A; Savini G. |
| EPI_ISL_500783, EPI_ISL_590952, EPI_ISL_775387, EPI_ISL_1820053, EPI_ISL_2333261 | Akershus University Hospital, Department for Microbiology and Infectious Disease Control | Norwegian Institute of Public Health, Department of Virology | 'Kathrine Stene-Johansen; Atiya R Ali; Debech Nadia; Engebretsen Serina Beate; Garcia Llorente Ignacio; Hilde Elshaug; Hilde Vollan; Jon Bråte; Kamilla Heddeland Instefjord; Karoline Bragstad; Kathrine Stene-Johansen; Line Victoria Moen; Marie Paulsen Madsen; Olav Hungnes; Pedersen Benedikte Nevjen; Rasmus Riis Kopperud |
| EPI_ISL_625613 | Alameda County Public Health Lab | Chan-Zuckerberg Biohub | CZB Cliahub Consortium |
| EPI_ISL_1483588, EPI_ISL_2626698, EPI_ISL_2987166 | Alaska State Virology Laboratory | Alaska State Virology Laboratory | Elva House; Jack Chen; Jacob Zidek; Lisa Smith; Ph.D.; Stephanie DeRonde |
| EPI_ISL_2163060, EPI_ISL_2169909 | Alberta Precision Labs (APL) | Public Health Agency of Canada (PHAC) National Microbiology Laboratory | Buss; Croxen M; Deo A; Dieu P; E; Ferrato C; Gill K; Khan F; Koleva P; Li V; Lloyd C; Lynch T; Ma R; Murphy S; Pabbaraju K; Shokoples S; Thayer J; Tipples G; Whitehouse M; Wong A; Yu C; Zelyas N |
| EPI_ISL_2375924 | Albow Gardens Clinic wc ALB | NHLS/UCT | Arash Iranzadeh; Bruna Galvao; Carolyn Williamson; Deelan Doolabh; Diana Hardie; Innocent Mudau; Kruger Marais; Lynn Tyers; Marvin Hsiao; Stephen Korsman |
| EPI_ISL_1385811 | Alfa Diagnostica LLC | ONCOGENE LLC | ONCOGENE LLC |
| EPI_ISL_508174 | All india institute of Medical Sciences Rishikesh | National Institute of Biomedical Genomics | Amit Mangla; Arindam Maitra; Deepjyoti Kalita; Ravi Kant; Saumitra Das |
| EPI_ISL_2375992 | Alma CDC wc AHC | NHLS/UCT | Arash Iranzadeh; Bruna Galvao; Carolyn Williamson; Deelan Doolabh; Diana Hardie; Innocent Mudau; Kruger Marais; Lynn Tyers; Marvin Hsiao; Stephen Korsman |
| EPI_ISL_2854052 | Ampang Hospital | Institute for Medical Research, Infectious Disease Research Centre, National Institutes of Health, Ministry of Health Malaysia | Anasir MI; Azizan MA; Kamel K; Mohd Zawawi Z; Ramly N; Robert F; Suppiah J; Thayan R |
| EPI_ISL_2727238 | Ampath Greenacres Hospital | KRISP, KZn Research Innovation and Sequencing Platform | Giandhari Jennifer; Naidoo Yeshnee; Pillay Sureshnee; San James; Tegally Houriiyah; Tshabuila Derek; Wilkinson Eduan; Yajna Ramphal; de Oliveira Tulio |
| EPI_ISL_2178783, EPI_ISL_2178785, EPI_ISL_2178788, EPI_ISL_2178789, EPI_ISL_2178793, EPI_ISL_2178794, EPI_ISL_2193359 | | | |
| see above | Ampath Laboratories | National Institute for Communicable Diseases of the National | Amoako DG; Bhiman JN; Glass A; Gottberg A; Mahlangu B; Marshall T; Mohale T; Ntuli N; Scheepers C; Viana R |

| | | | |
|---|---|---|---|
| | | Health Laboratory Service | |
| EPI_ISL_1324593 | Anteja laboratorija (UAB Diagnostikos laboratorija) | Vilnius University Hospital Santaros Klinikos, Center of Laboratory Medicine | Daniel Naumovas; Dovile Ezerskyte; Gytis Dudas; Ingrida Olendraite; Laimonas Griskevicius; Ligita Raugaite; Mindaugas Stoskus; Monika Katenaite; Rimvydas Norvilas |
| EPI_ISL_1483700 | Area of Virology, Serology and Virology Division (SAVID), New South Wales Health Pathology Randwick | Area of Virology, Serology and Virology Division (SAVID), New South Wales Health Pathology Randwick | Au, J.; Bull, R.; Deveson, I.; Foster, C.; Jean, T.; Rawlinson, W.; Ruiz Silva, M.; Van Haal, S. |
| EPI_ISL_678312, EPI_ISL_678327, EPI_ISL_678328, EPI_ISL_707898, EPI_ISL_717702, EPI_ISL_1121974, EPI_ISL_1121975, EPI_ISL_1494718, EPI_ISL_1494722, EPI_ISL_1543926, EPI_ISL_1615594, EPI_ISL_1633353, EPI_ISL_2650011 | | | |
| see above | Area of Virology, Serology and Virology Division (SAViD), New South Wales Health Pathology Randwick | Virology Research Laboratory; Area of Virology, Serology and Virology Division (SAViD), New South Wales Health Pathology Randwick | Au, J.; Bull, R.; Deveson, I.; Foster, C.; Rawlinson, W.; Ruiz Silva, M.; Van Hal, S. |
| EPI_ISL_2931537, EPI_ISL_2931586 | Arkansas Public Health Laboratory, Arkansas Department of Health | University of Minnesota Genomics Center | Corbin Dirkx; Daryl M. Gohl; Jaquelyn Kuriger-Laber; John Garbe |
| EPI_ISL_1315065 | Armed Forces Medical Research Laboratories and Blood Bank, Egypt | Department of Microbial Biotechnology, Genetic Engineering Division, National Research Centre | Ahmed Elsayed; Ahmed Taha; Ayman Farghaly; Mohamed Khalifa; Mohamed Shemis; Reham Dawood |
| EPI_ISL_1315064 | Armed Forces Medical Research Laboratories and Blood Bank, Egypt | Department of Microbial Biotechnology, Genetic Engineering Division, National Research Centre, | Ahmed Elsayed; Ahmed Taha; Ayman Farghaly; Mohamed Khalifa; Mohamed Shemis; Reham Dawood |
| EPI_ISL_2849763 | Artic Network workflow-EPI2ME labs | King Khalid Bin Abdul Aziz street, South Surra, Kuwait City | Anfal Al-Adwani; Ebaa Al-Awadhi; Hussain Safar; Nada Madi |
| EPI_ISL_2423528 | Atlanta VA Medical Center | Genomics and Discovery, Respiratory Viruses Branch, Division of Viral Diseases, Centers for Disease Control and Prevention | Adam Retchless; Anna Kelleher; Anna Uehara; Brian Lynch; Clinton R. Paden; Dhwani Batra; Haibin Wang; Han Jia Justin Ng; Jasmine Padilla; Jing Zhang; Justin Lee; Krista Queen; Mark Burroughs; Mili Sheth; Morgan Davis; Peter Cook; Rachel Marine; Sarah Nobles; Suxiang Tong; Tara Coalter; Yan Li; Ying Tao |
| EPI_ISL_413490 | Auckland Hospital | Institute of Environmental Science and Research (ESR) | Erasmus Smit; Gary McAuliffe; Joep de Ligt; Lauren Jelly; Matt Storey; Matthew Blakiston; Sally Roberts; Xiaoyun Ren |
| EPI_ISL_853962, EPI_ISL_853964, EPI_ISL_1671058, EPI_ISL_1840870, EPI_ISL_1840872, EPI_ISL_1840952, EPI_ISL_1840962, EPI_ISL_2137339, EPI_ISL_2232813, EPI_ISL_2232857, EPI_ISL_2232886, EPI_ISL_2232895, EPI_ISL_2324297, EPI_ISL_2324298, EPI_ISL_2324300, EPI_ISL_2427162, EPI_ISL_2485020, EPI_ISL_2485057, EPI_ISL_2485157, EPI_ISL_2616224, EPI_ISL_2616312, EPI_ISL_2616365, EPI_ISL_2657417, EPI_ISL_2757990, EPI_ISL_2757992, EPI_ISL_2758026, EPI_ISL_2887420, EPI_ISL_2887619, EPI_ISL_2887630, EPI_ISL_2985444, EPI_ISL_3039920, EPI_ISL_3039925 | | | |
| see above | Austrian Agency for Health and Food Safety (AGES) | Bergthaler laboratory, CeMM Research Center for Molecular Medicine of the Austrian Academy of Sciences | Alexander Lercher; Alexandra Popa; Andreas Bergthaler; Anna Schedl; Bekir Erguner; Benedikt Agerer; Christoph Bock; Christoph Bock; Fabian Amman; Jakob-Wendelin Genger; Jan Laine; Lukas Endler; Maelle Le Moing; Martin Senekowitsch; Martin Senekowitsch; Matthew Thornton; Michael Schuster; Michael Schuster; Petr Triska; Thomas Penz |
| EPI_ISL_1670599, EPI_ISL_1919625 | Azienda Ospedaliera Terni | Istituto Zooprofilattico Sperimentale dell'Abruzzo e Molise "G. Caporale" | Ancora M; Calistri P; Cammà C; Caporale M; Curini V; Delli Compagni E; Di Domenico M; Di Lollo Valeria; Di Pasquale A; Lorusso A; Mangone I; Marcacci M; Palumbo M; Puglia I; Rinaldi A; Savini G; Scaccetti A; Scialabba S |
| EPI_ISL_1222791, EPI_ISL_1489770 | Azienda Sanitaria dell'Alto Adige Laboratorio Aziendale di Microbiologia e Virologia | Istituto di Genomica Applicata | Davide Scaglione; Eleonora Paparelli; Elisa Masi; Elisabetta Giacobazzi; Elisabetta Pagani; Gabriele Magris; Irena Jurman; Irene Bianconi; Michele Morgante; Stefanie Wieser; Vera Vendramin |
| EPI_ISL_467044 | B.J. Medical College and Civil hospital | Gujarat Biotechnology Research Centre | ; A M Kadri; Ankit Hinsu; Apurvasinh Puvar; Chaitanya Joshi; Dinesh Kumar; Harsh Bakshi; Janvi Raval; Kamlesh J Upadhyay; Komal Patel; Labdhi Pandya; Madhvi Joshi; Maharshi Pandya; Monika Gandhi; Nidhi Patel; Nirav Mungalpara; Nitin Savaliya; Pinal Trivedi; Pranay Shah; Pritesh Sabara; Priti Pandita; R D Dixit; Raghawendra Kumar; Snehal Bagatharia; Tejas Shah; Zarna Patel; Zuber Saiyed |
| EPI_ISL_2710311 | BARC / Lancet | KRISP, KZN Research Innovation and Sequencing Platform | Giandhari Jennifer; Naidoo Yeshnee; Pillay Sureshnee; San James; Sisonke; Tegally Houriiyah; Tshabuila Derek; Wilkinson Eduan; Yajna Ramphal; de Oliveira Tulio |
| EPI_ISL_2842230 | BBMP Urban | INSACOG-KA, NIMHANS | Ananthapadmanabha Kotambail; Anita S Desai; Anson Kunjumon George; Chetan G K; Chitra Pattabiraman; Darshan Sreenivas; Ellango Ramasamy; Gautham Arunachal Udupi; Mahesh Kumar.C.S; Sony Sharma; V Ravi |
| EPI_ISL_1169048 | BBTKLPP Banjarbaru, Kalimantan Selatan | National Institute of Health Research and Development | Arie Ardiansyah Nugraha; Hana Apsari Pawestri; Hartanti Dian Ikawati; Kartika Dewi Puspa; Nelly Puspandari; Subangkit; Vivi Setiawaty |
| EPI_ISL_415577, EPI_ISL_418828, EPI_ISL_968754, EPI_ISL_974610, EPI_ISL_974630, EPI_ISL_974656, EPI_ISL_974661, EPI_ISL_975034, EPI_ISL_2499685, EPI_ISL_2522213, EPI_ISL_2530954 | | | |
| see above | BCCDC Public Health Laboratory | BCCDC Public Health Laboratory | Ana Pacagnella; Choi; Corrinne Ng; Dan Fornika; Gilmour; Harrigan; Hoang; John Tyson; Kamelian; Kim Macdonald; Kimia Kamelian; Krajden; Lapointe; Lee; Levett; Li; Linda Hoang; Loman; Loretta Janz; Mel Krajden; Prystajecky; Prystajecky Natalie; Quick; Robert Azana; Robert Azana Terry Snutch; Sekirov; Shannon Russell; Snutch; Tyson |
| EPI_ISL_3032332, EPI_ISL_3032333 | BIOFAST | Instituto Butantan | Antonio Jorge Martins; Claudia Renata dos Santos Barros; David Schlesinger; Debora Botequio Moretti; Dimas Tadeu Covas; Elaine Cristina Marqueze; Elaine Vieira Santos; Evandra Strazza Rodrigues; Heidge Fukumasu; Jayme Augusto de Souza-Neto; José Salvatore Leister Patané; Luiz Alcantara; Luiz Lehmann Coutinho; Maria Carolina Elias; Maurício Lacerda Nogueira; Rafael dos Santos Bezerra; Raul Machado Neto; Rejane Maria Tommasini Grotto; Ricardo Haddad; Sandra Coccuzzo Sampaio Vessoni; Simone Kashima; Svetoslav Nanev Slavov; Vincent Louis Viala |
| EPI_ISL_2933674 | BIOMNIS LYON | CNR Virus des Infections Respiratoires - France SUD | Antonin Bal; Bruno Lina; Gregory Destras; Gwendolyne Burfin; Hadrien Regue; Laurence Josset; Martine Valette; Quentin Semanas |
| EPI_ISL_2141965 | BIOR | Latvian Biomedical Research and Study Centre | Daina Pule; Davids Fridmanis; Elina Dimina; Guntars Zarins; Irena Meistere; Ivars Silamikelis; Janis Klovins; Janis Pjalkovskis; Jurijs Perevoscikovs; Kaspars Megnis; Laila Silamikele; Lauma Freimane; Laura Ansone; Liga Birzniece; Monta Ustinova; Nikita Zrelovs; Uga Dumpis; Una Krumina; Vita Rovite |
| EPI_ISL_859567, EPI_ISL_859573, EPI_ISL_859607, EPI_ISL_859642, EPI_ISL_859646, EPI_ISL_859668, EPI_ISL_859695, EPI_ISL_859735, EPI_ISL_859896, EPI_ISL_859900, EPI_ISL_859910, EPI_ISL_859951, EPI_ISL_860061, EPI_ISL_860068, EPI_ISL_860081 | | | |
| see above | BTC, Khalifa University | BTC, Khalifa University | Al Safar et al |
| EPI_ISL_1263459 | BTKLPP Ambon | National Institute of Health Research and Development | Arie Ardiansyah Nugraha; Hana Apsari Pawestri; Hartanti Dian Ikawati; Kartika Dewi Puspa; Krisna Pangesti; Nelly Puspandari; Subangkit; Vivi Setiawaty |
| EPI_ISL_2970354 | Bacterial and Viral Diseases & Biotechnology and Bioinformatics, Armauer Hansen Research Institute | Bacterial and Viral Diseases & Biotechnology and Bioinformatics, Armauer Hansen Research Institute | A.A.; A.G.; Abdissa, A.; Abebe, M.; Adnew, B.; Alemayehu; Alemu, F.; Bayih; Beyene; D.A.; D.H.; G.T.; Gelanew, T.; Mihrete, A.; Mulu, A.; Negash; Seyoum, T.; Tefera |
| EPI_ISL_1159375 | Bahman Hospital | Laboratory of Molecular Biology and Cancer Immunology,Lebanese University Public Health England | Fadi Abdel Sater; Steven Pullan |
| EPI_ISL_2544801 | Balai Besar Penelitian dan Pengembangan dan Reservoir Penyakit, Salatiga | Genomik Solidaritas Indonesia Laboratorium | Alfin Mohammad Abdillah; Anindya Pradipta Susanto; Annisa Muthiah Sukirman; Anuraj Shankar; Ariel Pradipta; Budi Wiweko; Carissa Sintca Wijaya; Dhahlia Agustina Cahyono; Gracia Felias Enos Korompis; Joko Waluyo; Louisa Markus; Meutia Ayuputeri Kumaheri; Normastuti Adhini Tantyo; Pratiwi Sudarmono; Vania Gavrila Wikasa; Yudistira Aditya Rahmat |
| EPI_ISL_2343223 | Banteay Meanchey Rapid Response Team | Virology Unit, Institut Pasteur du Cambodge | Cecile Troupin; Chau Darapheak; Chin Savuth; Erik A Karlsson; Jurre Y Siegers; Kraing Sidonn; Leakhena Pum; Ly Sovann; Veasna Duong; Yi Sengdoeurn |
| EPI_ISL_766645 | Barnakuten | The Public Health Agency of Sweden | Department of Microbiology; The Public Health Agency of Sweden |
| EPI_ISL_833333 | Batangas City Health Office | Research Institute for Tropical Medicine | Catalino Demetria; Daria Manalo; Edelwisa Mercado; Francisco Gerardo Polotan; Hannah Leah Morito; Inez Andrea Medado; John Leonard Chan; Kirstyn Brunker; Ma Angelica Tujan; Othoniel Jan Onza |
| EPI_ISL_2406459, EPI_ISL_2406460 | Battambang Provincial Laboratory | Virology Unit, Institut Pasteur du Cambodge | Cecile Troupin; Chau Darapheak; Chin Savuth; Erik A Karlsson; Jurre Y Siegers; Kraing Sidonn; Leakhena Pum; Ly Sovann; Veasna Duong; Yi Sengdoeurn |
| EPI_ISL_1117377 | Beijing Center for Disease Prevention and Control | Beijing Center for Disease Prevention and Control | Bing Lyu; Daitao Zhang; Fu Li; Lijuan Chen; Quanyi Wang; Shujuan Cui; Yang Pan; Zhaomin Feng; Zhichao Liang |
| EPI_ISL_529216 | Beijing Institute of Microbiology and Epidemiology | Beijing Institute of Microbiology and Epidemiology | Cui, Y.; Fan; Guo, Y.; Hang; Hou, J.; Li, B.; Mi, Z.; Mu, J.; Qin, E.; Song; Teng; Wu, Y.; Xu, Z.; Yajun.; Yang, R.; Yong, Y.; Yue; Zhang, X. |
| EPI_ISL_509712 | Belize Ministry of Health | Pathogen Discovery, Respiratory Viruses Branch, Division of Viral Diseases, Centers for Disease Control and Prevention | Anna Uehara; Clinton Paden; Haibin Wang; Jing Zhang; Krista Queen; Suxiang Tong; Yan Li; Ying Tao |
| EPI_ISL_780383, EPI_ISL_780384, EPI_ISL_780398 | Bermuda Government Molecular Diagnostics Laboratory (MDL) | Respiratory Virus Unit, National Infection Service, Public Health England | Dr Ayoola Oyinloye (Bermuda); Dr Carika Weldon (Bermuda); PHE Covid Sequencing Team |
| EPI_ISL_2802860 | BioMoLab | Microbiologia Molecular, Instituto SELADIS, Universidad Mayor de San Andrés | Aneth Vasquez Michel; Carmen Delgado Barrera; Oscar M. Rollano-Peñaloza; Sandra Miranda Sardon |
| EPI_ISL_2802858, EPI_ISL_2802859, EPI_ISL_2802863 | BioMoLab | Molecular Genetics Laboratory, Instituto de Investigaciones Químicas, Universidad Mayor de San Andrés | Aneth Vasquez Michel; Carmen Delgado Barrera; Oscar M. Rollano-Peñaloza; Sandra Miranda Sardon |
| EPI_ISL_1502568, EPI_ISL_1502570, EPI_ISL_2107513, EPI_ISL_2107517 | Biochemistry and Molecular Biology Department-Faculty of Medicine, Al-Quds University | Biochemistry and Molecular Biology Department-Faculty of Medicine, Al-Quds University | Al-Jawabreh, A.; Dumaidi, K.; Ereqat, S.; Naserddin, A.; Nasereddin, A. |
| EPI_ISL_1707927, EPI_ISL_2878346, EPI_ISL_2878383 | Biogroup Bio Lam-LCD Saint-Denis | Department of Virology, Henri Mondor University Hospital, Assistance Publique Hôpitaux de Paris, Université Paris-Est Créteil, INSERM U955 | Alexandre Soulier; Christophe Rodriguez; Elisabeth Trawinski; Guillaume Gricourt; Jean-Michel Pawlotsky; Melissa N'Debi; Slim Fourati; Vanessa Demontant |
| EPI_ISL_429995, EPI_ISL_430011, EPI_ISL_635778, EPI_ISL_635781, EPI_ISL_730270, EPI_ISL_730479, EPI_ISL_730558, EPI_ISL_730559, EPI_ISL_730562, EPI_ISL_755242, EPI_ISL_878477 | | | |
| see above | Biolab Diagnostic Laboratories | Andersen lab at Scripps Research | Ahmad Tibi; Amid Abdelnour with SEARCH Alliance San Diego; Issa Abu-Dayyeh; Lama Hussein; Lina Mohammad; Zein Naber |
| EPI_ISL_977602, EPI_ISL_1406195, EPI_ISL_1406239, EPI_ISL_1822601, EPI_ISL_1823120, EPI_ISL_1823177, EPI_ISL_1823202, EPI_ISL_1824717, EPI_ISL_2105670, EPI_ISL_2105672, EPI_ISL_2105674, EPI_ISL_2617461, EPI_ISL_2617463, EPI_ISL_2658758, EPI_ISL_2658760, EPI_ISL_2658761, EPI_ISL_2658762, EPI_ISL_2693977, EPI_ISL_2693980, EPI_ISL_2868422 | | | |
| see above | Biolab Diagnostic Laboratories | Biolab Diagnostic Laboratories | Ahmad Tibi; Amid Abdelnour; Badia Saddedin; Eiad Atwa; Issa Abu-Dayyeh; Lama Hussein; Shaima Ali; Shayma Ali |
| EPI_ISL_527007 | Biological Prevention, Army | Biological Prevention, Army | A.E.; A.F.; A.M.; Ageez; B.E.; Elhoseiny; Gad; Harty; M.D.; M.F.; M.G.; Seadawy; Shabaan; Shamel |
| EPI_ISL_526975, EPI_ISL_526976, EPI_ISL_526977, EPI_ISL_526978, EPI_ISL_526979, EPI_ISL_526980, EPI_ISL_526981, EPI_ISL_526982, EPI_ISL_526983, EPI_ISL_526984, EPI_ISL_526985, EPI_ISL_526986, EPI_ISL_526987, EPI_ISL_526988, EPI_ISL_526989, EPI_ISL_526990, EPI_ISL_526991, EPI_ISL_526992, EPI_ISL_526993, EPI_ISL_526994, EPI_ISL_526995, EPI_ISL_526996, EPI_ISL_526997, EPI_ISL_526998, EPI_ISL_526999, EPI_ISL_527000, EPI_ISL_527001, EPI_ISL_527002, EPI_ISL_527003, EPI_ISL_527004, EPI_ISL_527005, EPI_ISL_527006 | | | |
| see above | Biological prevention, army | Biological prevention, army | A.E.; A.F.; A.M.; Ageez; B.E.; Elhoseiny; Elhosieny; Gad; Harty; M.D.; M.F.; M.G.; Seadawy; Shabaan; Shamel |
| EPI_ISL_510526, EPI_ISL_510532 | Biological prevention, army | Biological prevention, army | A.A.; A.F.; A.M. and Soliman; Ali; Amer; B.E.; B.S.; ELnabrawy; ElGohary; Elhoseiny; Elhoseny; Elnagdy; Elnakeeb; Gad; H.A.; Harty; Hassan; Kandeil; Karam; M.A.; M.D.; M.F.; M.G.; M.M. and Gad; Raouf; Seadawy; Shamel; T.A.; W.A.; Y.A.; k.E. |
| EPI_ISL_1524332, EPI_ISL_1524359, EPI_ISL_2931131, EPI_ISL_2931134, EPI_ISL_2931135, EPI_ISL_2931137, EPI_ISL_2931138 | | | |
| see above | Biology Department, College of Science, Al Muthanna University and Public Health Laboratory, Al-Muthanna Health | Department of Virology, Faculty of Medicine, University of Helsinki, Helsinki, Finland | Alaa Hameed; Ali Jasim; Batool Kadham Salman; Hussein Alburkat; Hussein Riadh Kitab; Murad Munahi; Nihad Al-Rashedi; Olli Vapalahti; Tarja Sironen; Teemu Smura |

| | | | |
|---|---|---|---|
| | Directorate | | |
| EPI_ISL_582030 | Biology Department, College of Science, Al-Muthanna University | International Centre for Genetic Engineering and Biotechnology (ICGEB) and ARGO Open Lab Platform | Alessandro Marcello; Danilo Licastro; Nihad Al-Rashedi; Simeone Dal Monego; Sreejith Rajasekharan |
| EPI_ISL_1015369 | Biology Lab, HIA PERCY | IRBA, 2MI | FOISSAUD V.; GORGE O.; NOLENT F.; SARILAR V. |
| EPI_ISL_907076, EPI_ISL_907077, EPI_ISL_907078, EPI_ISL_907079, EPI_ISL_907080, EPI_ISL_907081, EPI_ISL_907082, EPI_ISL_907083, EPI_ISL_907084, EPI_ISL_907085 | | | |
| see above | Biology, MCL | Biology, MCL | A.F.; A.N.; Elhoseiny; Gad; M.D.; M.F.; M.G.; Seadawy; Shamel; Zekri |
| EPI_ISL_794634 | Biology, MCL | Biology, MCL | A.F.; A.S.; B.S. and EL-Safty; EL-harty; EL-hosieny; Gad; M.D.; M.F.; M.G.; Seadawy; Shamel |
| EPI_ISL_1420697 | Biomedical Research Foundation of the Academy of Athens (BRFAA) | Greek Genome Center, Biomedical Research Foundation of the Academy of Athens (BRFAA) | Dimitrios Thanos; Emmanouil Athanasiadis; Ioannis Vatsellas; Katerina Zoi; Theodoros Loupis |
| EPI_ISL_2401515 | BioneXt Lab | Laboratoire national de sante, Microbiology, Microbial Genomics Platform | Anke Wienecke-Baldacchino; Catherine Ragimbeau; Fatu Djabi; Jessica Tapp; Lise Pignon; Raoul Salmon; Tamir Abdelrahman; Thibault Ferrandon |
| EPI_ISL_2878374, EPI_ISL_2984054 | Biopole Antilles | Department of Virology, Henri Mondor University Hospital, Assistance Publique Hôpitaux de Paris, Université Paris-Est Créteil, INSERM U955 | Alexandre Soulier; Christophe Rodriguez; Elisabeth Trawinski; Guillaume Gricourt; Jean-Michel Pawlotsky; Melissa N'Debi; Slim Fourati; Vanessa Demontant |
| EPI_ISL_1140703, EPI_ISL_1211107, EPI_ISL_1211289, EPI_ISL_1211353, EPI_ISL_1212796, EPI_ISL_1216676, EPI_ISL_1282001, EPI_ISL_1282133, EPI_ISL_1282295, EPI_ISL_1284692, EPI_ISL_1353663, EPI_ISL_1439255, EPI_ISL_1721811, EPI_ISL_2633951, EPI_ISL_2634081, EPI_ISL_2637135 | | | |
| see above | Bioscientia Labor Wermsdorf | Robert Koch Institute | |
| EPI_ISL_1284823, EPI_ISL_1432947, EPI_ISL_1721133, EPI_ISL_1848523, EPI_ISL_1848543, EPI_ISL_2109502, EPI_ISL_2121932, EPI_ISL_2122022, EPI_ISL_2125493, EPI_ISL_2125560, EPI_ISL_2126008, EPI_ISL_2260272, EPI_ISL_2265701, EPI_ISL_2265849, EPI_ISL_2265853, EPI_ISL_2313390, EPI_ISL_2313636, EPI_ISL_2632395, EPI_ISL_2632447, EPI_ISL_2632460, EPI_ISL_2632554 | | | |
| see above | Bioscientia MVZ Labor Karlsruhe GmbH | Robert Koch Institute | |
| EPI_ISL_1970307 | Biotechnology Division, NCDC Delhi | NCDC Delhi, Biotechnology Division | Hema Gogia; Hemlata Lall; Kalaiarasan Ponnusamy; Mahesh S Dhar; Manoj K Singh; Meena Datta; Partha Rakshit; Preeti Madan; Priyanka Singh; Radhakrishnan V. S; Robin Marwal; Sandhya Kabra; Sujeet K Singh; Uma Sharma |
| EPI_ISL_745169 | Bishop Lavis CDC wc BLP | National Health Laboratory Service (NHLS), Tygerberg | Bronwyn Kleinhans; Eduan Wilkindon; Gert van Zyl; Houriiyah Tegally; Kayla Delaney; Susan Engelbrecht; Tulio de Oliveira; Wolfgang Preiser |
| EPI_ISL_2380020, EPI_ISL_2608398 | Borneo Medical Centre | Institute of Health and Community Medicine | Chan Chia Jui; Chua Hock Hin; David Perera; Ooi Mong How; Tonnii Sia Loong Loong; Wong Jyn Shan; Wong Kiing Aik |
| EPI_ISL_871872, EPI_ISL_968214, EPI_ISL_1516831, EPI_ISL_1516846, EPI_ISL_1516847, EPI_ISL_2233458, EPI_ISL_2233463, EPI_ISL_2372269, EPI_ISL_2372270, EPI_ISL_2372271, EPI_ISL_2372273, EPI_ISL_2372275, EPI_ISL_2372276, EPI_ISL_2372277, EPI_ISL_2372280, EPI_ISL_2372283, EPI_ISL_2372286, EPI_ISL_2372287, EPI_ISL_2372294, EPI_ISL_2372297, EPI_ISL_2372299, EPI_ISL_2372305, EPI_ISL_2386153, EPI_ISL_2386154, EPI_ISL_2504067, EPI_ISL_2504068, EPI_ISL_2504069, EPI_ISL_2504072, EPI_ISL_2504074, EPI_ISL_2504075, EPI_ISL_2504077, EPI_ISL_2504080, EPI_ISL_2504082, EPI_ISL_2504084, EPI_ISL_2504087, EPI_ISL_2566206, EPI_ISL_2566207, EPI_ISL_2566208, EPI_ISL_2566209, EPI_ISL_2648202, EPI_ISL_2820337, EPI_ISL_2820338, EPI_ISL_2820339, EPI_ISL_2820340, EPI_ISL_2820341, EPI_ISL_2820342, EPI_ISL_2820343, EPI_ISL_2820345, EPI_ISL_2820347, EPI_ISL_2820348, EPI_ISL_2820349, EPI_ISL_2820350, EPI_ISL_2820352, EPI_ISL_2820353, EPI_ISL_2820354, EPI_ISL_2820355, EPI_ISL_2820357, EPI_ISL_2820358, EPI_ISL_2820359, EPI_ISL_2820360, EPI_ISL_2820361, EPI_ISL_2820362, EPI_ISL_2820364, EPI_ISL_2820365, EPI_ISL_2820366, EPI_ISL_2820367, EPI_ISL_2820368, EPI_ISL_2820370, EPI_ISL_2820371, EPI_ISL_2820373, EPI_ISL_2820374, EPI_ISL_2820418, EPI_ISL_2820419, EPI_ISL_2820427, EPI_ISL_2820429, EPI_ISL_2820435, EPI_ISL_2820436, EPI_ISL_2820438, EPI_ISL_2820439, EPI_ISL_2820440, EPI_ISL_2820442, EPI_ISL_2820444, EPI_ISL_2820445, EPI_ISL_2820453, EPI_ISL_2820455, EPI_ISL_2820460, EPI_ISL_2820461, EPI_ISL_2868330, EPI_ISL_2868335, EPI_ISL_2868338, EPI_ISL_2868339, EPI_ISL_2868348, EPI_ISL_2868351, EPI_ISL_2868359, EPI_ISL_2868361, EPI_ISL_2868362, EPI_ISL_2868363, EPI_ISL_2868373, EPI_ISL_2868378, EPI_ISL_2868387, EPI_ISL_2868388, EPI_ISL_2868397, EPI_ISL_2868398, EPI_ISL_2868402, EPI_ISL_2868406, EPI_ISL_2931798, EPI_ISL_2931799, EPI_ISL_2931800, EPI_ISL_2931801, EPI_ISL_2931803, EPI_ISL_2931805, EPI_ISL_2931807, EPI_ISL_2931808, EPI_ISL_2931809, EPI_ISL_2931810, EPI_ISL_2931811, EPI_ISL_2931812, EPI_ISL_2931813, EPI_ISL_2931814, EPI_ISL_2931815, EPI_ISL_2931816, EPI_ISL_2931817, EPI_ISL_2931819, EPI_ISL_2931820, EPI_ISL_2931822, EPI_ISL_2931823, EPI_ISL_2931824, EPI_ISL_2931825, EPI_ISL_2931826, EPI_ISL_2931830, EPI_ISL_2931834, EPI_ISL_2931835, EPI_ISL_2931836 | | | |
| see above | Botswana Harvard HIV Reference Laboratory | Botswana Harvard HIV Reference Laboratory | Boitumelo Zuze; Botshelo Radibe; David Lawrence; Dorcas Maruapula; Godfrey Simoonga; Irvin Kgetse; Joseph; Joseph Makhema; Kelebeletse K. Mokobela; Keoratile Ntshambiwa; Kereng Mphoyakgosi; Kgomotso Moruisi; Kwana Lechiile; Legodile Kooepile; Lindani Bochena; Madisa Mine; Makhema; Modisa Motswaledi; Mosepele Mosepele; Motlalepule L. Pone; Ontlametse T. Bareng; Pamela Smith-Lawrence; Roger Shapiro; Shahin Lockman; Sikhulile Moyo; Sikhulile Wonderful T. Choga; Simani Gaseitsiwe; Thongbotho Mphoyakgosi; Wonderful Choga; Wonderful T. Choga |
| EPI_ISL_1001001, EPI_ISL_1034758, EPI_ISL_1034760 | Bundeswehr Institute of Microbiology | Bundeswehr Institute of Microbiology | Alexandra Rehn; Enrico Georgi; Malena Bestehorn-Willmann; Markus Antwerpen; Mathias Walter; Mike Pillukat; Roman Wölfel; Sabine Zange |
| EPI_ISL_1834008 | Bundeswehrkrankenhaus Berlin | Bundeswehr Institute of Microbiology | Alexandra Rehn; Enrico Georgi; Malena Bestehorn-Willmann; Markus Antwerpen; Mathias Walter; Mike Pillukat; Roman Wölfel; Sabine Zange |
| EPI_ISL_2779327 | Bungoma County Referral Hospital | USAMRD-A, Basic Science Laboratory | Alan Lemtudo; Beth Mutai; Brian Andika; Carol Kifude; Clement Masakwe; Eric Muthanje; Esther Omuseni; Faith Sigei; Gathii Kimita; George Awinda; John Waitumbi; Josphat Nyataya; Rachel Githii; Rehema Liyai; Stephen Ochola |
| EPI_ISL_2779387, EPI_ISL_3031380 | Busia County Referral Hospital | USAMRD-A, Basic Science Laboratory | Alan Lemtudo; Beth Mutai; Brian Andika; Carol Kifude; Clement Masakwe; Eric Muthanje; Esther Omuseni; Faith Sigei; Gathii Kimita; George Awinda; John Waitumbi; Josphat Nyataya; Rachel Githii; Rehema Liyai; Stephen Ochola |
| EPI_ISL_2779289 | Busia border point | USAMRD-A, Basic Science Laboratory | Alan Lemtudo; Beth Mutai; Brian Andika; Carol Kifude; Clement Masakwe; Eric Muthanje; Esther Omuseni; Faith Sigei; Gathii Kimita; George Awinda; John Waitumbi; Josphat Nyataya; Rachel Githii; Rehema Liyai; Stephen Ochola |
| EPI_ISL_1371903, EPI_ISL_2289766 | C H DE LA POLYNESIE FRANCAISE | CNR Virus des Infections Respiratoires - France SUD | Antonin Bal; Bruno Lina; Bruno Simon; Gregory Destras; Gwendolyne Burfin; Hadrien Regue; Laurence Josset; Martine Valette; Quentin Semanas |
| EPI_ISL_2674322 | CARVAJAL LABORATORIOS | Instituto Nacional de Salud- Dirección de Investigación en Salud Pública | Carlos Franco-Muñoz; Carmen Osorio; Diana Malo; Diego A. Álvarez-Díaz; Diego Andrés Prada; Gerardo Santamaría; Hector Alejandro Ruiz-Moreno; Jhonnatan Reales-González; Jorge Rivera; Juan Camilo Martinez; Julian Naizaque; Katherine Laiton-Donato; Lisseth Pardo; Magdalena Wiesner; Marcela Mercado-Reyes; Maria T. Herrera-Sepúlveda; Marta Lopez Blanco; Martha Lucia Ospina Martinez; Paola Rojas; Sergio Gomez; Sheryll Corchuelo; Ángela Alarcon Cruz |
| EPI_ISL_2657865 | CARVAJALA LABORATORIO IPS | Instituto Nacional de Salud- Dirección de Investigación en Salud Pública | Carlos Franco-Muñoz; Carmen Osorio; Diana Malo; Diego A. Álvarez-Díaz; Diego Andrés Prada; Gerardo Santamaría; Hector Alejandro Ruiz-Moreno; Jhonnatan Reales-González; Jorge Rivera; Juan Camilo Martinez; Julian Naizaque; Katherine Laiton-Donato; Lisseth Pardo; Magdalena Wiesner; Marcela Mercado-Reyes; Maria T. Herrera-Sepúlveda; Marta Lopez Blanco; Martha Lucia Ospina Martinez; Paola Rojas; Sergio Gomez; Sheryll Corchuelo; Ángela Alarcon Cruz |
| EPI_ISL_2825738 | CDPH VBL | California Department of Public Health | CDPH-COVIDNet; UCLA Technology Center for Genomics & Bioinformatics |
| EPI_ISL_605780, EPI_ISL_605781, EPI_ISL_605782 | CEIRS Data Processing and Coordinating Center, St. Jude Center of Excellence for Influenza Research and Surveillance (CEIRS) | CEIRS Data Processing and Coordinating Center, St. Jude Center of Excellence for Influenza Research and Surveillance (CEIRS) | A.E.; Ali; El-Guindy; El-Sayes, M.; El-Shesheny, R.; El-Taweel, A.; Gomaa, M.; Kamel; Kandeil, A.; Kayali, G.; Kayed; Kutkat, O.; M.A.; M.N.; Mahmoud; Mahrous, N.; Moatasim, Y.; Mostafa, A.; N.M.; Naguib, A.; Roshdy; S.H.; Shehata, M.; Showky, S.; W.H.; Webby, R. |
| EPI_ISL_2471228, EPI_ISL_3043914 | CENTOGENE Frankfurt Laboratory: Niederlassung Industriepark Höchst | Robert Koch Institute | |
| EPI_ISL_1695254 | CENTRE HOSPITALIER DU HAUT BUGEY | CNR Virus des Infections Respiratoires - France SUD | Antonin Bal; Bruno Lina; Bruno Simon; Gregory Destras; Gwendolyne Burfin; Hadrien Regue; Laurence Josset; Martine Valette; Quentin Semanas |
| EPI_ISL_2373694 | CENTRE HOSPITALIER ST JOSEPH ST LUC | CNR Virus des Infections Respiratoires - France SUD | Antonin Bal; Bruno Lina; Bruno Simon; Gregory Destras; Gwendolyne Burfin; Hadrien Regue; Laurence Josset; Martine Valette; Quentin Semanas |
| EPI_ISL_644252 | CEPHR / Mater Hospital | Irish Coronavirus Sequencing Consortium - National Virus Reference Laboratory | Alejandro Abner Garcia Leon; Gabriel Gonzalez; Michael Carr; Patrick Mallon |
| EPI_ISL_2562031, EPI_ISL_2562045 | CH Princesse Grace | Cerba lab | Aude Lessenne; Bénédicte Roquebert; Emmanuel Lecorche; Kader Merah; Laura Verdurme; Patrice Herisson; Sabine Trombert-Paolantoni; Stéphanie Haïm-Boukobza; Thierry Collin |
| EPI_ISL_2933198 | CHR LA REUNION FELIX GUYON | CNR Virus des Infections Respiratoires - France SUD | Antonin Bal; Bruno Lina; Gregory Destras; Gwendolyne Burfin; Hadrien Regue; Laurence Josset; Martine Valette; Quentin Semanas |
| EPI_ISL_2694661, EPI_ISL_2694662, EPI_ISL_2709915, EPI_ISL_2709935 | CHRIS HANI BARAGWANATH | National Institute for Communicable Diseases of the National Health Laboratory Service | Amoako DG; Bhiman JN; Everatt J; Ismail A; Mahlangu B; Mnguni A; Mohale T; Ntuli N; Scheepers C |
| EPI_ISL_2273025, EPI_ISL_2273036, EPI_ISL_2273053, EPI_ISL_2273054, EPI_ISL_2382241, EPI_ISL_2382243, EPI_ISL_2382682, EPI_ISL_2454635, EPI_ISL_2454649, EPI_ISL_2662632, EPI_ISL_2662644, EPI_ISL_2662645, EPI_ISL_2662650 | | | |
| see above | CHRIS HANI BARAGWANATH LABORATORY | National Institute for Communicable Diseases of the National Health Laboratory Service | Amoako DG; Bhiman JN; Everatt J; Ismail A; Mahlangu B; Mnguni A; Mohale T; Ntuli N; Scheepers C |
| EPI_ISL_660356 | CHU Clermont-Ferrand | CNR Virus des Infections Respiratoires - France SUD | Amélie Brebion; Antonin Bal; Audrey Mirand; Bruno Lina; Christel Regagnon; Christine Archimbaud; Cécile Henquell; Gregory Destras; Gwendolyne Burfin; Hadrien Règue; Hélène Chabrolles; Laurence Josset; Martine Chambon; Martine Valette; Maxime Bisseux; Patricia Combes; Quentin Semanas |
| EPI_ISL_2403233, EPI_ISL_2403237, EPI_ISL_2403240, EPI_ISL_2403242, EPI_ISL_2709247, EPI_ISL_2709252, EPI_ISL_3039524, EPI_ISL_3039535 | | | |
| see above | CHU Pontchaillou | CHU Pontchaillou | DE TAYRAC Marie; DENOUAL Florent; ETCHEVERRY Amandine; FEBREAU Christine; GALIBERT Marie Dominique; GROLHIER Claire; JAGLINE Steven; PRONIER Charlotte; SASSI Mohamed; THIBAULT Vincent |
| EPI_ISL_593859, EPI_ISL_2351673 | CHU Purpan - Laboratoire de Virologie - Institut Fédératif de Biologie | CHU Purpan - Laboratoire de Virologie - Institut Fédératif de Biologie | Boyer P.; Bulach T.; Carcenac R.; Donnadieu C.; Dubois M.; Harter A.; Izopet J.; Latour J.; Milhes M.; Nicot F.; Ranger N.; Salin G.; Tremeaux P. |
| EPI_ISL_2509992 | CHUGA-IBP-laboratoire de Virologie | IBP-laboratoire de virologie | Alban Caporossi; Anne Signori -Schmuck; Anne-Karen Faure; Aurélie Truffot; Benjamin Nemoz; Hugo Jardin; Julien Andréani; Julien Lupo; Léa Ponderand; Pascal Poignard; Patrice Morand; Raphaële Germi; Sylvie Larrat |
| EPI_ISL_2249173 | CHULN - H Santa Maria | Instituto Nacional de Saude (INSA) and Centro de Investigaçao em Biodiversidade e Recursos Geneticos (CIBIO), Universidade do Porto | Borges et al |
| EPI_ISL_1545239, EPI_ISL_2367418 | CHUV | Laboratory of genomics and metagenomics | Claire Bertelli; Damien Jacot; Gilbert Greub; Sébastien Aeby; Trestan Pillonel |
| EPI_ISL_2683495, EPI_ISL_2683873 | CICM, Bamako | Malaria Research and Training Center-Bamako | Abdoul Karim Sangare; Abdoulaye Djimde; Amadou Daou; Antoine Dara; Bourema Kouriba |
| EPI_ISL_2672214 | CICM, Bamako | Malaria Research and Training Center-Parasito | Abdoul Karim Sangare; Abdoulaye Djimde; Amadou Daou; Antoine Dara; Bourema Kouriba |
| EPI_ISL_487463, EPI_ISL_487464 | CICM-Mali | Bundeswehr Institut of Microbiology | Antwerpen; Bestehorn-Willmann; Dürr; Heitzer; Kouriba; Maiga; Quedraogo; Rehn; Sangaré; Sogodogo; Traoré; Walter; Wölfel; Zimmermann |
| EPI_ISL_2502731 | CLINICA BIBLICA | Inciensa, Instituto Costarricense de InvestigaciOn y Enseñanza en NutriciOn y Salud | Adriana Godínez; Claudio Soto-Garita; Estela Cordero; Francisco Duarte; Hebleen Porras; Jose Luis Vargas; Joselyn Prado & Karla Gutierrez-González; Mariela Gutierrez; Melany CalderOn |
| EPI_ISL_1299497, EPI_ISL_1591100 | CMA Dano | Centre Muraz | Abdoul-Salam Ouedraogo; Ange Badjo; Armel Poda; Arsène Somé; Arsène Zongo; Essia Belarbi; Fabian Leendertz; Firmin Kaboré; Grit Schubert; Jasmin Schlotterbeck; Soumeya Ouangraoua; Thérèse Kagone; Yacouba Sawadogo |
| EPI_ISL_2438608 | COCR NEW TENGIZ Clinic | Reference laboratory for the control of viral infections | Aidar Usserbayev; Aknur Mutaliyeva; Andrey Komissarov; Artem Fadeev; Azamat Kenessov; Bekzhan Maikotov; Gaukhar Nussupbayeva; Madina Tleubergenova; Maria Pisareva; Nazym Tleumbetova |

| | | | |
|---|---|---|---|
| EPI_ISL_2484798 | COMMUNITY TESTING CENTRE | Hong Kong Department of Health | Alan K.L. Tsang; Dominic N.C. Tsang; Edman T.K. Lam; Gannon C.K. Mak; Ken H.L. Ng; Peter C.W. Yip; Peter K.C. Cheng; Rickjason C.W. Chan |
| EPI_ISL_2227272, EPI_ISL_2227273 | COVID-19 National Reference Laboratory, Pasteur Institute of Iran | Genetics Research Center, University of Social Welfare and Rehabilitation Sciences | Hossein Najmabadi; Hossein Najmabadi.; Kimia Kahrizi; Mahsa Tavakoli; Marzieh Mohseni; Mohammad Hassan Pouriayevali; Mostafa Salehi-Vaziri; Tahmineh Jalali; Zohreh Fattahi |
| EPI_ISL_1190767 | CREMER(Centre de Recherches sur les Maladies Emergentes et Ré-émergentes) | TransVIHMI(Recherches Translationnelles sur le VIH et les Maladies Infectieuses) | Ahidjo Ayouba; Celestin Godwe; Christelle Butel; Dowbiss Meta Djomsi; Eitel Mpoudi Ngole; Eric Delaporte; Esemu Livo; Laetitia Serrano; Marcel Tongo; Marie Amougou; Martin Maidadi Foudi; Martine Peeters; Nicole Vidal; Rodrigue Kamga |
| EPI_ISL_636980 | CS Xai Xai | KRISP, KZN Research Innovation and Sequencing Platform | Giandhari J; Ismael N; Nadia Sitoe; Nedio Mabunda; Paulo Arnaldo; Pillay S; Tegally H; Wilkinson E; de Oliveira T |
| EPI_ISL_910079 | CSIR-Centre for Cellular and Molecular Biology | CSIR-Centre for Cellular and Molecular Biology | Archana Bharadwaj Siva; B Himasri; Blessy B John; Divya Tej Sowpati; Karthik Bharadwaj Tallapaka; Lamuk Zaveri; Namami Gaur; Payel Mukherjee; Pratheusa Maccha; Priya Singh; Purushotham Vodnala; Rakesh K Mishra; Sofia Banu; Tulasi Nagabandi; Viswagithe S L |
| EPI_ISL_1380396, EPI_ISL_1838393, EPI_ISL_1838400, EPI_ISL_1838452, EPI_ISL_1838522, EPI_ISL_2775002 | CSIR-Centre for Cellular and Molecular Biology | CSIR-Centre for Cellular and Molecular Biology-INSACOG | Amareshwar Vodapalli; Ara Sreenivas; Archana Bharadwaj Siva; B Himasri; Blessy B John; Divya Tej Sowpati; Jandhyala Sai Krishna; Karthik Bharadwaj Tallapaka; Lamuk Zaveri; Namami Gaur; Onkar Kulkarni; Payel Mukherjee; Pratheusa Maccha; Priya Nurkurthy; Priya Singh; Purushotham Vodnala; Rakesh K Mishra; Sharath Chandra Thota; Shreekant Verma; Sofia Banu; Sumedha Avadhanula; Tulasi Nagabandi; Valli Nagalakshmi Undamatla; Vidhyadhari Methuku; Viswagithe S L |
| EPI_ISL_2754370 | CURE | Institut Pasteur de Montevideo | Andres Lizasoain; Belén González; Cecilia Alonso; Daiana Mir; Emiliano Pereira; Gonzalo Bello; Ighor Arantes; Juan Zanetti; Lucia Bilbao; Luciana Griffero; Lucía Spangenberg; Mailen Arleo; Mariana Brandes; María José Benítez-Galeano; Matías Castells; Matías Salvo; Matías Victoria; Mauricio Méndez; Melissa Duquía; Natalia Rego; Natalia Reyes; Odhille Chappos; Pablo Smircich; Pía Techera; Rodney Colina; Tamara Fernández-Calero; Tania Possi; Verónica Noya |
| EPI_ISL_935791 | Cadham Provincial laboratory | National Microbiology Laboratory (NML) | Anna Majer; Anneliese Landgraff; CanCOGeN's metadata curation team; Darian Hole; David Alexander; Elsie Grudeski; Gary Van Domselaar; Grace Seo; Jared Bullard; Jennifer Tanner; Kerry Dust; Kirsten Biggar; Madison Chapel; Morag Graham; Natalie Knox; Nathalie Bastien; Paul Van Caeseele; Philip Mabon; Public Health Agency of Canada CanCOGeN team; Rhiannon Huzarewich; Russell Mandes; Shari Tyson; Timothy Booth; Yan Li |
| EPI_ISL_413557 | California Department of Public Health | Chiu Laboratory, University of California, San Francisco | Chao-Yang Pan; Debra A. Wadford; Hugo Guevara; Scot Federman; Wei Gu; Xianding Deng; and Charles Y. Chiu |
| EPI_ISL_1817730 | Calvinia RSC Clinic | NHLS/UCT | Arash Iranzadeh; Bruna Galvao; Carolyn Williamson; Deelan Doolabh; Diana Hardie; Innocent Mudau; Kruger Marais; Lynn Tyers; Marvin Hsiao; Stephen Korsman |
| EPI_ISL_2547359, EPI_ISL_2547361 | Calvinia RSC Clinic | National Health Laboratory Service/University of Cape Town (NHLS/UCT) | Arash Iranzadeh; Bruna Galvao; Carolyn Williamson; Deelan Doolabh; Diana Hardie; Gert Marais; Innocent Mudau; Lynn Tyers; Marvin Hsiao; Stephen Korsman |
| EPI_ISL_1096139, EPI_ISL_1096140 | Cambodian National Public Health Laboratory, National Institute of Public Health | Virology Unit, Institut Pasteur du Cambodge | Chau Darapheak; Chin Savuth; Erik A Karlsson; Kraing Sidonn; Ly Sovann; Sokhoun Yann; Veasna Duong; Yi Sengdoeurn |
| EPI_ISL_576371, EPI_ISL_576372, EPI_ISL_576373, EPI_ISL_857315, EPI_ISL_857316, EPI_ISL_857317, EPI_ISL_857318, EPI_ISL_857319, EPI_ISL_857320, EPI_ISL_857321, EPI_ISL_857322, EPI_ISL_857323, EPI_ISL_857324, EPI_ISL_857325, EPI_ISL_857326, EPI_ISL_857327, EPI_ISL_857328, EPI_ISL_857329, EPI_ISL_857330, EPI_ISL_857331, EPI_ISL_857332, EPI_ISL_857333, EPI_ISL_857334, EPI_ISL_857335, EPI_ISL_857336, EPI_ISL_857337, EPI_ISL_857338, EPI_ISL_857339, EPI_ISL_857340, EPI_ISL_857341, EPI_ISL_857342, EPI_ISL_857343, EPI_ISL_857344, EPI_ISL_857345, EPI_ISL_857346, EPI_ISL_857347, EPI_ISL_862783, EPI_ISL_862784, EPI_ISL_862785, EPI_ISL_862786, EPI_ISL_862787, EPI_ISL_862788, EPI_ISL_862789, EPI_ISL_862790, EPI_ISL_862791, EPI_ISL_862792, EPI_ISL_862793, EPI_ISL_862794, EPI_ISL_862795, EPI_ISL_862796, EPI_ISL_862797, EPI_ISL_862798, EPI_ISL_862799, EPI_ISL_862800, EPI_ISL_862801, EPI_ISL_862802, EPI_ISL_862803, EPI_ISL_862804, EPI_ISL_862805, EPI_ISL_862806, EPI_ISL_862807, EPI_ISL_862808, EPI_ISL_862809, EPI_ISL_862810, EPI_ISL_862811, EPI_ISL_862812, EPI_ISL_862813, EPI_ISL_890195, EPI_ISL_890196, EPI_ISL_890197, EPI_ISL_890198, EPI_ISL_890199, EPI_ISL_890200, EPI_ISL_890201, EPI_ISL_890202, EPI_ISL_890203, EPI_ISL_890204, EPI_ISL_890205, EPI_ISL_890206, EPI_ISL_890207, EPI_ISL_890208, EPI_ISL_890209, EPI_ISL_890210, EPI_ISL_890211, EPI_ISL_890212, EPI_ISL_890213, EPI_ISL_890214, EPI_ISL_890215, EPI_ISL_890216, EPI_ISL_890217, EPI_ISL_890218, EPI_ISL_890219, EPI_ISL_890220, EPI_ISL_890221, EPI_ISL_890222, EPI_ISL_890223, EPI_ISL_890224, EPI_ISL_890225, EPI_ISL_890226, EPI_ISL_890227, EPI_ISL_890228, EPI_ISL_890229, EPI_ISL_890230, EPI_ISL_890231, EPI_ISL_907086, EPI_ISL_907087, EPI_ISL_907088, EPI_ISL_907089, EPI_ISL_907090, EPI_ISL_907091, EPI_ISL_907092, EPI_ISL_907093, EPI_ISL_907094, EPI_ISL_907095, EPI_ISL_907096, EPI_ISL_907097, EPI_ISL_907098, EPI_ISL_907099, EPI_ISL_907100, EPI_ISL_907101, EPI_ISL_907102, EPI_ISL_907103, EPI_ISL_907104, EPI_ISL_907105, EPI_ISL_907106, EPI_ISL_907107, EPI_ISL_907108, EPI_ISL_907109, EPI_ISL_907110, EPI_ISL_907111, EPI_ISL_907112, EPI_ISL_907113, EPI_ISL_907114, EPI_ISL_907115, EPI_ISL_907116, EPI_ISL_907117, EPI_ISL_907118 | | | |
| see above | Cancer Biology Department, National Cancer Institute | Cancer Biology Department, National Cancer Institute | A.A.; A.E.; A.N.; Abouelhoda, M.; Ahmed; Bahnassy; Elhosieny; F.W.; Gad; H.K.; Hafez; Hamdy; M.G.; M.M.; M.S.; O.S.; Sedawy; Soliman; Soliman, L.; Zekri |
| EPI_ISL_1040916 | Cancer Biology Department, National Cancer Institute | Cancer Biology Department, National Cancer Institute | A.A.; A.E.; A.N.; Ahmed; Bahnassy; Elhosieny; F.W.; Gad; H.K.; Hafez; Hamdy; L. and Abouelhoda, M.; M.G.; M.M.; M.S.; O.S.; Sedawy; Soliman; Zekri |
| EPI_ISL_3021345, EPI_ISL_3021415 | Cantacuzino National Military-Medical Institute, Viral Respiratory Infections Laboratory | Cantacuzino Institute Virology | Carmen Cherciu; Luiza Ustea; Mihaela Lazar; Mihaela Oprea; Nicoleta Paraschiv; Sorin Dinu |
| EPI_ISL_548138, EPI_ISL_548140, EPI_ISL_579063, EPI_ISL_579066, EPI_ISL_579070, EPI_ISL_579086, EPI_ISL_579090, EPI_ISL_579426, EPI_ISL_579463, EPI_ISL_579496, EPI_ISL_622788, EPI_ISL_622789, EPI_ISL_622795, EPI_ISL_622799, EPI_ISL_622800, EPI_ISL_622810, EPI_ISL_622827, EPI_ISL_661260, EPI_ISL_682296, EPI_ISL_755638, EPI_ISL_843198, EPI_ISL_1016881, EPI_ISL_1082255, EPI_ISL_1904855, EPI_ISL_1967890, EPI_ISL_2406488, EPI_ISL_2811947, EPI_ISL_2811950, EPI_ISL_2811951, EPI_ISL_2811952, EPI_ISL_2811953, EPI_ISL_2811958 | | | |
| see above | Canterbury Health Laboratories | Institute of Environmental Science and Research (ESR) | Anja Werno; Antje van der Linden; Arlo Upton; Chris Mansell; David Hammer; Dragana Drinkovic; Erasmus Smit; Gary McAuliffe; Hana Sofia Andersson; Hermes Perez; James Ussher; Jill Sherwood; Jing Wang; Joep de Ligt; Josh Freeman; Julia Howard; Juliet Elvy; Lauren Jelly; Mary DeAlmeida; Matt Blakiston; Matt Storey; Matthew Rogers; Max Bloomfield; Michael Addidle; Michelle Balm; Muhammad Faisal; Nikki Freed; Olin Silander; Olivia Stroeven; Paula scholes; Rachel Boyle; Sally Roberts; SallyAnn Harbison; Sarah Jefferies; Sharmini Muttaiyah; Susan Lin; Susan Morpeth; Susan Taylor; Timothy Blackmore; Vani Sathyendran; Veronica Playle; Virginia Hope; Xiaoyun Ren |
| EPI_ISL_977538, EPI_ISL_977583 | Caribbean Public Health Agency | Carrington Lab, Department of PreClinical Sciences | Adesh Ramsubhag; Arianne Brown-Jordan; Avery Hinds; Chinna Chinnadurai; Christine V. F. Carrington; Christopher Oura; Dr. Sharon Belmar-George; Dr. Sharon Belmar-George Avery Hinds; Gabriel Escobar; Jaya Jayaraman; Jerome Foster; Karla Georges; Marsha Ivey; Naresh Nandram; Nikita S. D. Sahadeo; Nuno Faria; Oliver Pybus; Rahul Naidu; Rajini Haraksingh; Risha Singh; Sarah Hill; Stanley Giddings; SueMin Nathaniel; Vernie Ramkissoon |
| EPI_ISL_977659 | Caribbean Public Health Agency | Carrington Lab, Department of PreClinical Sciences, Building 36, First Floor Biochemistry Unit, Faculty of Medical Sciences, The University of the West Indies | Adesh Ramsubhag; Arianne Brown-Jordan; Avery Hinds; Chinna Chinnadurai; Christine V. F. Carrington; Christopher Oura; Gabriel Escobar; Jaya Jayaraman; Jerome Foster; Karla Georges; Kenneth George; Marsha Ivey; Naresh Nandram; Nikita S. D. Sahadeo; Nuno Faria; Oliver Pybus; Rahul Naidu; Rajini Haraksingh; Risha Singh; Sarah Hill; Stanley Giddings; SueMin Nathaniel; Vernie Ramkissoon |
| EPI_ISL_872194, EPI_ISL_1591250, EPI_ISL_1591351, EPI_ISL_1591518, EPI_ISL_1591718 | Caribbean Public Health Agency | Carrington Lab, Department of PreClinical Sciences, Faculty of Medical Sciences, The University of the West Indies | Adesh Ramsubhag; Arianne Brown-Jordan; Avery Hinds; Chinna Chinnadurai; Christine V. F. Carrington; Christopher Oura; Gabriel Escobar; Jaya Jayaraman; Jerome Foster; Karla Georges; Marsha Ivey; Naresh Nandram; Nikita S. D. Sahadeo; Nuno Faria; Oliver Pybus; Rahul Naidu; Rajini Haraksingh; Risha Singh; Roshan Parasram; Sarah Hill; Stanley Giddings; SueMin Nathaniel; Vernie Ramkissoon |
| EPI_ISL_872193 | Caribbean Public Health Agency | Carrington Lab, Department ofBuilding 36, First Floor Biochemistry Unit, Faculty of Medical Sciences, The University of the West Indies | Adesh Ramsubhag; Arianne Brown-Jordan; Avery Hinds; Chinna Chinnadurai; Christine V. F. Carrington; Christopher Oura; Gabriel Escobar; Jaya Jayaraman; Jerome Foster; Karla Georges; Marsha Ivey; Naresh Nandram; Nikita S. D. Sahadeo; Nuno Faria; Oliver Pybus; Rahul Naidu; Rajini Haraksingh; Risha Singh; Sarah Hill; Stanley Giddings; SueMin Nathaniel; Vernie Ramkissoon |
| EPI_ISL_459858 | Center for Genome Regulation (CRG) | Center for Mathematical Modeling and Center for Genome Regulation. Santiago, Chile | Allende ML; Gaete A; González M.; Maass A; Palma R; Travisany D; Urra C; Varas M |
| EPI_ISL_1594295, EPI_ISL_1594306 | Center for Laboratory Medicine St. Gallen | Center for Laboratory Medicine St. Gallen | Yannick Gerth |
| EPI_ISL_2835623, EPI_ISL_2835624, EPI_ISL_2835634, EPI_ISL_2835635, EPI_ISL_2835636, EPI_ISL_2835637, EPI_ISL_2835640 | | | |
| see above | Center for Medical and Sanitary Research (CERMES) | CDC Atlanta | Adamou Lagare; Dhwani Batra; Justin Lee |
| EPI_ISL_853839 | Center for Virology, Medical University of Vienna | Bergthaler laboratory, CeMM Research Center for Molecular Medicine of the Austrian Academy of Sciences | Alexander Lercher; Alexandra Popa; Andreas Bergthaler; Anna Schedl; Benedikt Agerer; Christoph Bock; Jakob-Wendelin Genger; Jan Laine; Lukas Endler; Martin Senekowitsch; Michael Schuster; Thomas Penz |
| EPI_ISL_2385405 | Center of Hygiene and Epidemiology in Belgorod Region | WHO National Influenza Centre Russian Federation | Alexey Masharsky; Andrey Komissarov; Artem Fadeev; Daria Danilenko; Dmitry Lioznov; Elena Nabieva; Georgii Bazykin; Kirill Varchenko; Ksenia Safina; Kseniya Komissarova; Maria Baturova; Maria Pisareva; Maria Timofeeva; Mikhail Bakaev; Nikita Yolshin; Oula Masour; Tamila Musaeva; Veronika Eder |
| EPI_ISL_1346779 | Center of Medical Microbiology, Virology, and Hospital Hygiene, University of Duesseldorf | Center of Medical Microbiology, Virology, and Hospital Hygiene, University of Duesseldorf | Alexander Dilthey; Andreas Walker; Daniel Strelow; Jessica Nicolai; Jörg Timm; Klaus Pfeffer; Lisanna Hülse; Malte Kohns Vasconcelos; Maximilian Damagnez; Nadine Lübke; Tobias Wienemann; Torsten Houwaart |
| EPI_ISL_2227328, EPI_ISL_2227329, EPI_ISL_2227330, EPI_ISL_2227331, EPI_ISL_2227332, EPI_ISL_2227333, EPI_ISL_2227334, EPI_ISL_2227335, EPI_ISL_2227336, EPI_ISL_2227337, EPI_ISL_2227338, EPI_ISL_2227339, EPI_ISL_2227340, EPI_ISL_2227341, EPI_ISL_2227342, EPI_ISL_2227343, EPI_ISL_2227344, EPI_ISL_2227345, EPI_ISL_2227346, EPI_ISL_2227347, EPI_ISL_2227348, EPI_ISL_2227349, EPI_ISL_2227350, EPI_ISL_2232252, EPI_ISL_2232253, EPI_ISL_2232254, EPI_ISL_2232255, EPI_ISL_2232256, EPI_ISL_2232257, EPI_ISL_2232258, EPI_ISL_2232259, EPI_ISL_2232260, EPI_ISL_2232261, EPI_ISL_2232262, EPI_ISL_2232263, EPI_ISL_2232264, EPI_ISL_2232265, EPI_ISL_2232266, EPI_ISL_2232267, EPI_ISL_2232268, EPI_ISL_2232269, EPI_ISL_2232270, EPI_ISL_2232271, EPI_ISL_2232272, EPI_ISL_2232273, EPI_ISL_2232274, EPI_ISL_2232275, EPI_ISL_2232276, EPI_ISL_2232277, EPI_ISL_2232278, EPI_ISL_2232279, EPI_ISL_2232280, EPI_ISL_2232281, EPI_ISL_2232282, EPI_ISL_2232283, EPI_ISL_2232284, EPI_ISL_2232285, EPI_ISL_2232286, EPI_ISL_2232287, EPI_ISL_2232288, EPI_ISL_2232289, EPI_ISL_2232290, EPI_ISL_2232291, EPI_ISL_2232292, EPI_ISL_2232293, EPI_ISL_2232294, EPI_ISL_2232295, EPI_ISL_2232296, EPI_ISL_2232297, EPI_ISL_2232298, EPI_ISL_2232299, EPI_ISL_2232300, EPI_ISL_2232301, EPI_ISL_2232302, EPI_ISL_2232303, EPI_ISL_2232304, EPI_ISL_2232305, EPI_ISL_2232306, EPI_ISL_2232307, EPI_ISL_2232308, EPI_ISL_2232309, EPI_ISL_2232310, EPI_ISL_2232311, EPI_ISL_2232312, EPI_ISL_2232313, EPI_ISL_2232314, EPI_ISL_2232315, EPI_ISL_2232316, EPI_ISL_2232317, EPI_ISL_2232318, EPI_ISL_2232319, EPI_ISL_2232320, EPI_ISL_2232321, EPI_ISL_2232322, EPI_ISL_2232323, EPI_ISL_2232324, EPI_ISL_2232325, EPI_ISL_2232326, EPI_ISL_2232327, EPI_ISL_2232328, EPI_ISL_2232329, EPI_ISL_2232330, EPI_ISL_2232331, EPI_ISL_2232332, EPI_ISL_2232333, EPI_ISL_2232334, EPI_ISL_2232335, EPI_ISL_2232336, EPI_ISL_2232337, EPI_ISL_2232338, EPI_ISL_2232339, EPI_ISL_2232340, EPI_ISL_2232341, EPI_ISL_2232342, EPI_ISL_2232343, EPI_ISL_2232344, EPI_ISL_2232345, EPI_ISL_2232346, EPI_ISL_2232347, EPI_ISL_2232348, EPI_ISL_2232349, EPI_ISL_2232350, EPI_ISL_2232351, EPI_ISL_2232352, EPI_ISL_2232353, EPI_ISL_2232354, EPI_ISL_2232355, EPI_ISL_2232356, EPI_ISL_2232357, EPI_ISL_2232358, EPI_ISL_2232359, EPI_ISL_2232360, EPI_ISL_2232361, EPI_ISL_2232362, EPI_ISL_2232363, EPI_ISL_2232364, EPI_ISL_2232365, EPI_ISL_2232366, EPI_ISL_2232367, EPI_ISL_2232368, EPI_ISL_2232369, EPI_ISL_2232370, EPI_ISL_2232371, EPI_ISL_2232372, EPI_ISL_2232373, EPI_ISL_2232374, EPI_ISL_2232375, EPI_ISL_2232376, EPI_ISL_2232377, EPI_ISL_2232378, EPI_ISL_2232379, EPI_ISL_2232380, EPI_ISL_2232381, EPI_ISL_2232382, EPI_ISL_2232383, EPI_ISL_2232384, EPI_ISL_2232385, EPI_ISL_2232386, EPI_ISL_2232387, EPI_ISL_2232388, EPI_ISL_2232389, EPI_ISL_2232390, EPI_ISL_2232391, EPI_ISL_2232392, EPI_ISL_2232393, EPI_ISL_2232394, EPI_ISL_2232395, EPI_ISL_2232396, EPI_ISL_2232397, EPI_ISL_2232398, EPI_ISL_2232399, EPI_ISL_2232400, EPI_ISL_2232401, EPI_ISL_2232402, EPI_ISL_2232403, EPI_ISL_2232404, EPI_ISL_2232405, EPI_ISL_2232406, EPI_ISL_2232407, EPI_ISL_2380050, EPI_ISL_2380051, EPI_ISL_2380052, EPI_ISL_2380053, EPI_ISL_2380054, EPI_ISL_2380055, EPI_ISL_2380056, EPI_ISL_2380057, EPI_ISL_2380058, EPI_ISL_2380059, EPI_ISL_2380060, EPI_ISL_2380061, EPI_ISL_2380062, EPI_ISL_2380063, EPI_ISL_2380064, EPI_ISL_2380065, EPI_ISL_2380066, EPI_ISL_2380067, EPI_ISL_2380068, EPI_ISL_2380069, EPI_ISL_2380070, EPI_ISL_2380071, EPI_ISL_2380072, EPI_ISL_2380073, EPI_ISL_2380075, EPI_ISL_2380076, EPI_ISL_2380077, EPI_ISL_2380078, EPI_ISL_2380079, EPI_ISL_2380080, EPI_ISL_2380081, EPI_ISL_2380082, EPI_ISL_2380083, EPI_ISL_2380084, EPI_ISL_2380085, EPI_ISL_2380086, EPI_ISL_2380087, EPI_ISL_2380088, EPI_ISL_2380089, EPI_ISL_2380090, EPI_ISL_2380091, EPI_ISL_2380092, EPI_ISL_2380093, EPI_ISL_2380094, EPI_ISL_2380095, EPI_ISL_2380096, EPI_ISL_2380097, EPI_ISL_2380098 | | | |
| see above | Center of Scientific Excellence for Influenza Viruses (CSEIV), National Research Centre | Center of Scientific Excellence for Influenza Viruses (CSEIV), National Research Centre | Ahmed E Kayed; Ahmed El-Taweel; Ahmed Kandeil; Ahmed Mostafa; Amal Naguib; Ghazi Kayali; M K Khalifa; Mahmoud Shehata; Mina Kamel; Mohamed Ahmed Ali; Mohamed El Sayes; Mohamed Hassany; Mokhtar Gomaa; Nancy M. El Guindy; Noura M Abo Shama; Omnia Kutkat; Rabeh El-Shesheny; Richard Webby; Sara Mahmoud; Shymaa S Ahmed; Wael Roshdy; Yassmin Moatasim |
| EPI_ISL_430820 | Center of Scientific Excellence for Influenza Viruses, National Research Centre (NRC), Egypt. | Center of Scientific Excellence for Influenza Viruses, National Research Centre (NRC), Egypt. | Abo Shama; Ahmed E Kayed; Ahmed El-Taweel; Ahmed Kandeil; Ahmed Mostafa; Amal Naguib; M Noura; Mahmoud Shehata; Mina Kamel; Mohamed Ahmed Ali; Mohamed El Sayes; Mokhtar Gomaa; Nancy M. El Guindy; Omnia Kutkat; Rabeh El-Shesheny; Sara Mahmoud; Shymaa Showky Ahmed; Wael Roshdy; Yassmin Moatasim |
| EPI_ISL_430819 | Center of Scientific Excellence for Influenza Viruses,National Research Centre (NRC), Egypt. | Center of Scientific Excellence for Influenza Viruses,National Research Centre (NRC), Egypt. | Abo Shama; Ahmed E Kayed; Ahmed El-Taweel; Ahmed Kandeil; Ahmed Mostafa; Amal Naguib; M Noura; Mahmoud Shehata; Mina Kamel; Mohamed Ahmed Ali; Mohamed El Sayes; Mokhtar Gomaa; Nancy M. El Guindy; Omnia Kutkat; Rabeh El-Shesheny; Sara Mahmoud; Shymaa Showky Ahmed; Wael Roshdy; Yassmin Moatasim |
| EPI_ISL_406031 | Centers for Disease Control, R.O.C. (Taiwan) | Centers for Disease Control, R.O.C. (Taiwan) | Ji-Rong Yang; Jung-Jung Mu; Ming-Tsan Liu; Shu-Ying Li; Yu-Chi Lin |
| EPI_ISL_815257, EPI_ISL_815368 | Centogene | Centogene | Krishna Kumar Kandaswamy; Peter Bauer; Vivi Hue-Trang Lieu |
| EPI_ISL_2123097 | Centogene; Dr. Bauer Laboratoriums GmbH | Robert Koch Institute | |
| EPI_ISL_2834910, EPI_ISL_2834911, EPI_ISL_2834914, EPI_ISL_2834915 | Central Health Laboratory | Central Health Laboratory ,Victoria Hospital, Candos,Ministry of Health and Wellness, Mauritius | Bahadoor BS; Jannoo N; Manraj SS; Mathur H; Ramuth M; Sonoo J; Sujeewon C |
| EPI_ISL_2834925, EPI_ISL_2834926, EPI_ISL_2834927, EPI_ISL_2834928 | Central Health Laboratory/Airport Health Laboratory | Central Health Laboratory ,Victoria Hospital, Candos,Ministry of Health and Wellness, Mauritius | Bahadoor BS; Jannoo N; Manraj SS; Mathur H; Patoo M; Ramuth M; Sonoo J; Sujeewon C |

| | | | |
|---|---|---|---|
| EPI_ISL_794593, EPI_ISL_794594, EPI_ISL_794595, EPI_ISL_794596, EPI_ISL_794597, EPI_ISL_794598, EPI_ISL_794599, EPI_ISL_794600, EPI_ISL_794601, EPI_ISL_794602, EPI_ISL_794603, EPI_ISL_794604, EPI_ISL_2313046, EPI_ISL_2313047, EPI_ISL_2313048, EPI_ISL_2313049, EPI_ISL_2313050, EPI_ISL_2313051, EPI_ISL_2313052, EPI_ISL_2313053, EPI_ISL_2313054, EPI_ISL_2313055, EPI_ISL_2313056, EPI_ISL_2313057, EPI_ISL_2313058, EPI_ISL_2313059, EPI_ISL_2313060, EPI_ISL_2313061, EPI_ISL_2313062, EPI_ISL_2313063, EPI_ISL_2313064, EPI_ISL_2313065, EPI_ISL_2313066, EPI_ISL_2313067, EPI_ISL_2313068, EPI_ISL_2313069 | | | |
| see above | Central Laboratories, Egyptian Ministry of Health and Population | Central Laboratories, Egyptian Ministry of Health and Population | A.E.; Ali; El Guindy; El Sayes; El Taweel, A.; El-Shesheny; El-Shesheny, R.; Gomaa, M.; Hassany, M.; Kamel; Kandeil, A.; Kayali, G.; Kayed; Khalifa; Kutkat, O.; M.A.; M.K.; M.N.; Mahmoud; Mahrous, N.; Moatasim, Y.; Mostafa, A.; N.M.; Naguib, A.; R.E.; Roshdy; S.H.; Saleh, M.; Shawky, S.; Shehata, M.; W.H. |
| EPI_ISL_2502381, EPI_ISL_2502387, EPI_ISL_2502393, EPI_ISL_2502402, EPI_ISL_2502426, EPI_ISL_2502427, EPI_ISL_2502430, EPI_ISL_2502435, EPI_ISL_2502438, EPI_ISL_2502441 | | | |
| see above | Central Laboratory, Bureau of Public Health (BOG) and Academic Hospital Paramaribo | Erasmus Medical Center | Bas B Oude Munnink; Cherise Beek; Consuella Partowidjojo; Dion Gajadin; Ed PF IJzerman; Emmanuelle Munger; Gary Gummels; Ingrid SK Krishnadath; Lycke Woittiez; Marion PG Koopmans; Mireille Van de Veer; Phyllis Pinas; Princes Wongsowidjojo; Radjesh Ori; Ranisha Doerbalie; Rohma Banwari; Soeradj Harkisoen; Stephen Vreden; Tilotmadebie Ramlal; Verne Nanhoe |
| EPI_ISL_1822763, EPI_ISL_1822784 | Central Medical Laboratory | Texas Children's Microbiome Center | Adrianna Maliga; Francis Morey; Gerhaldine Morazan; Jennifer K. Spinler; Kristy O. Murray; Melissa Diaz-Musa; Miguel Saldana; Russell I Manzanero; Ruth Ann Luna; Sarah M. Gunter; Shannon E. Ronca |
| EPI_ISL_529032 | Central Molecular Microbiology Laboratory and Next Generation Sequencing Reference Laboratory, Clinical and Chemical Pathology Department, Faculty of Medicine, CAIRO UNIVERSITY | Next Generation Sequencing Reference Laboratory, Faculty of Medicine, CAIRO UNIVERSITY and The Center for Genome and Microbiome Research, Faculty of Pharmacy, CAIRO UNIVERSITY | May Abdelfattah; May Sherif Soliman; Ramy Karam Aziz |
| EPI_ISL_529031 | Central Molecular Microbiology Laboratory, Clinical and Chemical Pathology Department, Faculty of Medicine, CAIRO UNIVERSITY | Next Generation Sequencing Reference Laboratory, Faculty of Medicine, Cairo University and The Center for Genome and Microbiome Research, Faculty of Pharmacy, CAIRO UNIVERSITY | May Abdelfattah; May Sherif Soliman; Ramy Karam Aziz |
| EPI_ISL_2464001 | Central Public Health Lab, National Public Health Organization | Central Public Health Lab, National Public Health Organization | Gregory Spanakos et al; Kyriaki Tryfinopoulou |
| EPI_ISL_794605, EPI_ISL_794606, EPI_ISL_794607, EPI_ISL_794608 | Central Public Health Laboratories, Egyptian Ministry of Health and Population | Central Public Health Laboratories, Egyptian Ministry of Health and Population | A.E.; Ali; El Guindy; El Sayes; El Taweel, A.; El-Shesheny, R.; Gomaa, M.; Kamel; Kandeil, A.; Kayali, G.; Kayed; Khalifa; Kutkat, O.; M.A.; M.K.; M.N.; Mahmoud; Mahrous, N.; Moatasim, Y.; Mostafa, A.; N.M.; Naguib, A.; Roshdy; S.H.; Saleh, M.; Shawky, S.; Shehata, M.; W.H. |
| EPI_ISL_693471, EPI_ISL_693473, EPI_ISL_693474, EPI_ISL_693477, EPI_ISL_693478, EPI_ISL_693481, EPI_ISL_693482, EPI_ISL_693483, EPI_ISL_1307643, EPI_ISL_1307660, EPI_ISL_1322329, EPI_ISL_1322330 | | | |
| see above | Central Public Health Laboratory | National Public Health Laboratory, National Centre for Infectious Diseases | Esorom Daoni; Lin Cui; Raymond Tzer Pin Lin; Sophie Octavia; Theresa Palou; Tze Minn Mak; Zhenyang Zhou |
| EPI_ISL_978512 | Central Public Health Laboratory - LACEN -Bahia, Salvador, Brazil | Central Public Health Laboratory - LACEN -Bahia, Salvador, Brazil | Arabela Leal; Breno Dominguez; Felicidade Pereira; Jaqueline Gomes; Luciana Oliveira; Luiz Alcantara; Marcela Gómez; Marta Giovanetti; Patrícia Cajado; Stephane Tosta; Vagner Fonseca; Vanessa Nardy |
| EPI_ISL_3045139, EPI_ISL_3045144, EPI_ISL_3045167, EPI_ISL_3045179, EPI_ISL_3045180, EPI_ISL_3045220 | Central Public Health Laboratory, National Public Health Organization | Central Public Health Laboratory, National Public Health Organization | A.Katsoulidou et al; G.Spanakos; Kyriaki Tryfinopoulou |
| EPI_ISL_2232157 | Central Public Health Laboratory, National Public Health Organization | Greek Genome Center, Biomedical Research Foundation of the Academy of Athens (BRFAA) | Dimitrios Thanos; Emmanouil Athanasiadis; Giannis Vatsellas; Katerina Zoi; Kyriaki Tryfinopoulou; Theodoros Loupis |
| EPI_ISL_2786923 | Central Virology Laboratory, Ministry of Health | Central Virology Laboratory, Ministry of Health | Amos Adler; Efrat Bucris; Ella Mendelson; Michal Mandelboim; Moran Shwartswort-Cohen; Neta S. Zuckerman; Noam Protter; Oran Erster; Orna Mor; Saar Burstein |
| EPI_ISL_2141877 | Centrala Laboratorija | Latvian Biomedical Research and Study Centre | Davids Fridmanis; Elina Dimina; Guntars Zarins; Ivars Silamikelis; Jana Osite; Janis Klovins; Janis Pjalkovskis; Jurijs Perevoscikovs; Kaspars Megnis; Laila Silamikele; Lauma Freimane; Laura Ansone; Liga Birzniece; Marta Priedite; Monta Ustinova; Nikita Zrelovs; Uga Dumpis; Una Krumina; Vita Rovite |
| EPI_ISL_961838, EPI_ISL_1312520 | Centrala laboratorija | Latvian Biomedical Research and Study Centre | Davids Fridmanis; Davids Fridmanis; Guntars Zarins; Ivars Silamikelis; Ivars Silamikelis; Jana Osite; Janis Klovins; Janis Pjalkovskis; Janis Klovins; Jurijs Perevoscikovs; Kaspars Megnis; Kaspars Megnis; Laila Silamikele; Laila Silamikele; Lauma Freimane; Lauma Freimane; Laura Ansone; Laura Ansone; Liga Birzniece; Liga Birzniece; Marta Priedite; Monta Ustinova; Monta Ustinova; Nikita Zrelovs; Nikita Zrelovs; Uga Dumpis; Uga Dumpis; Una Krumina; Vita Rovite; Vita Rovite |
| EPI_ISL_1357603 | Centre For Biotechnology Research And Development | THE AFRICA GENOMICS CENTRE AND CONSULTANCY | Adede Hawi; Cecilia Waruhiu; Damaris Matoke-Muhia; George Michuki; John Njuguna; Lilian Kanjau; Ravena Mubichi |
| EPI_ISL_414625 | Centre Hospitalier Régional Universitaire de Nantes Laboratoire de Virologie | National Reference Center for Viruses of Respiratory Infections, Institut Pasteur, Paris | Angela Brisebarre; Flora Donati Vincent Enouf; Marianne Coste-Burel; Marion Barbet; Maud Vanpeene; Méline Bizard; Mélnie Albert; Sylvie Behillil; Sylvie van der Werf |
| EPI_ISL_1001044, EPI_ISL_1001045, EPI_ISL_1001050, EPI_ISL_1001051, EPI_ISL_1001053, EPI_ISL_1001059, EPI_ISL_1001062, EPI_ISL_1001064, EPI_ISL_1001068, EPI_ISL_1790120, EPI_ISL_1790131 | | | |
| see above | Centre Pasteur du Cameroun | Institut Pasteur de Dakar | Carniel Elisabeth; Dia Ndongo; Diagne Moussa Moïse; Diallo Amadou; Diop Mamadou; Faye Ousmane; Loucoubar Cheikh; Ndiaye Ndack; Njouom Richard; Sall Amadou Alpha; Sankhe Safietou |
| EPI_ISL_788936, EPI_ISL_788938 | Centre de Recherche et de Formation en Infectiologie Guinée | TransVIHMI, IRD/INSERM/Monpellier University | Abdoul Karim SOUMAH; Abdoulaye TOURE; Ahidjo AYOUBA; Alimou CAMARA; Alpha Kabinet KEITA; Bouna Yatassaye; Christelle BUTEL; Eric DELAPORTE; Jean-louis MONEMOU; Joel KOIVOGUI; Kaba KOUROUMA; Laetitia SERRANO; Mamadou Bhoye KEITA; Mamadou Saliou BAH; Mamadou Saliou SOW; Mandiou DIAKITE; Martine PEETERS; Moriba POVOGUI; Penda Malhado DIALLO; Sakoba KEITA |
| EPI_ISL_1615655, EPI_ISL_1913075, EPI_ISL_1913078, EPI_ISL_1913080, EPI_ISL_2097218, EPI_ISL_2097219, EPI_ISL_2097220, EPI_ISL_2097221, EPI_ISL_2097223, EPI_ISL_2097228, EPI_ISL_2097229, EPI_ISL_2097230, EPI_ISL_2097231, EPI_ISL_2156795, EPI_ISL_2156822, EPI_ISL_2424136, EPI_ISL_2424137, EPI_ISL_2424157, EPI_ISL_2424158, EPI_ISL_2424159, EPI_ISL_2424160, EPI_ISL_2424161, EPI_ISL_2424162, EPI_ISL_2424163, EPI_ISL_2434970, EPI_ISL_2434971, EPI_ISL_2434974, EPI_ISL_2434975, EPI_ISL_2434979, EPI_ISL_2434980, EPI_ISL_2442203, EPI_ISL_2442276, EPI_ISL_2442277, EPI_ISL_2442279, EPI_ISL_2442338, EPI_ISL_2442363, EPI_ISL_2442383, EPI_ISL_2442384 | | | |
| see above | Centre de Recherches Médicales de Lambaréné (CERMEL) | Centre de Recherches Médicales de Lambaréné (CERMEL) | Anicet Mouity Matoumba; Armel MINTSA NDONG; Bertrand Lell and Ayola Akim Adegnika; Bénédicte NDEBOKO; Georgelin NGUEMA ONDO; Georgelin Nguema Ondo; Gédéon Prince Manouana; Jean Bernard LEKANA-DOUKI; Jean Bernard Lekana-Douki; Joël-Fleury DJOBA SIAWAYA; Joël-Fleury Djoba Siawaya; Michel Ngonga Dikongo; Moustapha Nzamba Maloum; Rodrigue BIKANGUI; Rodrigue Bikangui; Sam O'neilla Oye Bingono; Samira ZOA ASSOUMOU; Samira Zoa Assoumou; Srinivas reddy Pallerla; Steffen Borrmann; Thirumalaisamy P. Velavan |
| EPI_ISL_978545, EPI_ISL_978548, EPI_ISL_2968046, EPI_ISL_2968064, EPI_ISL_2968065, EPI_ISL_2968069 | Centre de Virologie des Maladies infectueuses Tropicales | Functional Genomic Platform UATRS-biology, CNRST | ; Abdelilah LARAQUI; Abderrazzak Rfaki; Ahmed REGGAD; Charifa DRISSI TOUZANI; Elmostafa BENAISSA; Elmostafa EL FAHIME; Farida HILALI; Hemlali Mouhssine; Hicham EL ANNAZ; Idriss-Amine LAHLOU; Ikram BNOUYAHIA; Khalid ENNIBI; Marouane MELLOUL; Mly Abdelaziz ELALAOUI; Mostafa ELOUENNASS; Nadia EL MRIMAR; Nadia TOUIL; Nadia Touil; Rachid ABI; Rida TAGAJDID; Safae ELKOCHRI; Sanaa ALAOUI-Amine; Tahar BAJJOU; Yassine SEKHSOKH; Youssef AKHOUAD; Zohour KASMY |
| EPI_ISL_471456 | Centre de Virologie des Maladies Tropicales | Functional Genomic Platform/Service Analyses Biologique/UATRS/ Centre National Pour la Recherche Scientifique Et Technique (CNRST) | Abdelillah LARAQUI; Abdelkader LAATIRIS; Ahmed REGGAD; Elmostafa EL FAHIME; Farida HILALI; Hicham EL ANNAZ; Idriss-Amine LAHLOU; Khalid ENNIBI; Marouane MELLOUL; Mly Abdelaziz ELALAOUI; Mostafa ELOUENNASS; Nadia TOUIL; Rachid ABI; Rida TAGAJDID; Safae ELKOCHRI; Sanaa ALAOUI-Amine; Tahar BAJJOU; Yassine SEKHSOKH; Youssef AKHOUAD; Zohour KASMY |
| EPI_ISL_792550, EPI_ISL_1233067, EPI_ISL_1717082, EPI_ISL_1717092, EPI_ISL_1717093, EPI_ISL_1970356, EPI_ISL_1970358, EPI_ISL_1970371, EPI_ISL_1970374, EPI_ISL_1970399, EPI_ISL_1970410, EPI_ISL_1970413, EPI_ISL_2481412, EPI_ISL_2481418, EPI_ISL_2481425, EPI_ISL_2803170, EPI_ISL_2803176, EPI_ISL_2803179, EPI_ISL_2803193, EPI_ISL_2803199, EPI_ISL_2803251, EPI_ISL_2803298 | | | |
| see above | Centre for Dengue Research and AICBU, Department of Immunology and Molecular Medicine | Centre for Dengue Research and AICBU, Department of Immunology and Molecular Medicine | Chandima Jeewandara; Deshan Madhusanka; Deshni Jayathilaka; Dinuka Ariyaratne; Diyanath Ranasinghe; Gathsaurie Neelika Malavige; Laksiri Gomes; Tibutius Thanesh Pramanayagam |
| EPI_ISL_668448, EPI_ISL_668449, EPI_ISL_668452, EPI_ISL_668455 | Centre for Dengue Research, Department of Immunology and Molecular Medicine | Centre for Dengue Research, Department of Immunology and Molecular Medicine | Chandima Jeewandara; Deshni Jayathilaka; Dinuka Ariyaratne; Diyanath Ranasinghe; Gathsaurie Neelika Malavige; Laksiri Gomes |
| EPI_ISL_602565 | Centre for Dengue Research, Department of Immunology and Molecular Medicine | Centre for Dengue Research, Department of Immunology and Molecular Medicine, | Chandima Jeewandara; Deshni Jayathilaka; Dinuka Ariyaratne; Dinuka Guruge; Diyanath Ranasinghe; Gathsaurie Neelika Malavige; Laksiri Gomes; Ruwan Wiajayamuni |
| EPI_ISL_679977, EPI_ISL_763605, EPI_ISL_841936 | Centre for Enzyme Innovation, University of Portsmouth / Translational Research Laboratory, Portsmouth Hospitals NHS Trust | COVID-19 Genomics UK (COG-UK) Consortium | Allyson Lloyd; Angela Beckett; Anoop Chauhan; Ethan Butcher; Garry Scarlett; Kelly Bicknell; Robert Impey; Samuel Robson; Sarah Wyllie; Scott Elliott; Sharon Glaysher; Yann Bourgeois |
| EPI_ISL_654794 | Centre for Human Virology & Genomics, Nigerian Institute of Medical Research | Centre for Human Virology & Genomics, Nigerian Institute of Medical Research | Shaibu, J. |
| EPI_ISL_2399460, EPI_ISL_2399462, EPI_ISL_2502019 | Centre for Human Virology and Genomics, Microbiology Department, Nigerian Institute of Medical Research | Central Research Laboratory, Nigerian Institute of Medical Research | Ayorinde Babatunde James; Azuka Patrick Okwuraiwe; Babatunde Lawal Salako; Bamidele Iwalokun; Chika Kingsley Onwuamah; Grace Oni; Joseph Ojonugwa Shaibu; Josiah Ayoola Isong; Joy Ayoola; Muinah Adenike Fowora; Ngozi Mirabel Otuonye; Nyam Itse Yusuf; Olufemi Samuel Amoo; Phasha-Muchemenye Mmatshepho; Rahaman A. Ahmed; Rosemary Ajuma Audu; Sharon Abimbola; Sola Ajibaye; Uyi Emokpae; Yusuf Jimoh |
| EPI_ISL_413550 | Centre for Human and Zoonotic Virology (CHAZVY), College of Medicine University of Lagos/Lagos University Teaching Hospital (LUTH), part of the Laboratory Network of the Nigeria Centre for Disease Control (NCDC) | African Centre of Excellence for Genomics of Infectious Diseases (ACEGID), Redeemer's University, Ede, Osun State, Nigeria | Ajogbasile F.V.; Folarin O.A.; Ihekweazu C. Happi C.T.; Kayode A.; Oguzie J.; Oluniyi P.E. |
| EPI_ISL_407893, EPI_ISL_413594 | Centre for Infectious Diseases and Microbiology Laboratory Services | NSW Health Pathology - Institute of Clinical Pathology and Medical Research; Westmead Hospital; University of Sydney | ; A; Alicia; Carter I; Chen SC; Eden J-S; Gall; Gray K; Holmes EC; Kok J and Dwyer DE for the 2019-nCoV Study Group; Kok J and Dwyer DE for the 2019-nCoV Study Group*; Lam C; M; Maddocks S; O'Sullivan MV; Rahman H; Rockett R; Sintchenko V; Timms; V |
| EPI_ISL_2842788 | Centro Medico ABC | Instituto Nacional de Medicina Genomica | Cedro-Tanda A; Cesar Lara; Daniel Aguirre; Escobar-Arrazola; Gonzalez-Barrera D; Herrera-Montalvo LA.; Hidalgo-Miranda A; Jacomine Reyes-Carrasco; M; Mendoza-Vargas A; Munguia-Garza P; Ramirez-Vega O; Rangel-DeLeon D; Reyes-Grajeda JP; Roxana Trejo-González |
| EPI_ISL_837557, EPI_ISL_837559, EPI_ISL_837564, EPI_ISL_837565, EPI_ISL_837569, EPI_ISL_837572, EPI_ISL_837573, EPI_ISL_837577, EPI_ISL_837578 | | | |
| see above | Centro Nacional de Enfermedades Tropicales (CENETROP) | Laboratory of Respiratory Viruses and Measles, Oswaldo Cruz Institute, FIOCRUZ | Ana Carolina Mendonca; Anna Carolina Paixao; Cinthia Avila; Fernando Motta; Luciana Appolinario; Marilda Siqueira on behalf of the Fiocruz COVID-19 Genomic Surveillance Network; Paola Resende; Roxana Loayza |
| EPI_ISL_2274031, EPI_ISL_2274037, EPI_ISL_2274038 | Centro Nacional de Enfermidades Tropicales (CENETROP) | Laboratory of Respiratory Viruses and Measles, Oswaldo Cruz Institute, FIOCRUZ | Alice Sampaio Rocha; Ana Carolina Mendonca; Anna Carolina Paixao; Cinthia Avila; Elisa Cavalcante Pereira; Fernando Motta; Luciana Appolinario; Marilda Siqueira on behalf of the Fiocruz COVID-19 Genomic Surveillance Network; Paola Resende; Renata Serrano Lopes; Roxana Loayza; Taina Venas |

| | | | |
|---|---|---|---|
| EPI_ISL_1531863, EPI_ISL_2231524 | Centro de Diagnostico COVID-19 UABC Tijuana | Andersen lab at Scripps Research | German Ibarra; Jonathan Vincent Baena; Jorge Luis Jimenez Niebla; Manuel Sanchez Alavez; Oscar Efren Zazueta Fierro; SEARCH Alliance San Diego with Idanya Rubi Serafin Higuera |
| EPI_ISL_635531 | Centro de Diagn√≥stico COVID-19 UABC Tijuana | Andersen lab at Scripps Research | Germán Ibarra; Jonathan Vincent Baena; Jorge Luis Jiménez Niebla; Manuel Sánchez Alavez; Oscar Efrén Zazueta Fierro; SEARCH Alliance San Diego with Idanya Rubí Serafín Higuera |
| EPI_ISL_2339876, EPI_ISL_2339903, EPI_ISL_2339907 | Centro de Estudio de Enfermedades Autoinmunes (CREA), Universidad del Rosario, Bogota, Colombia | Centro de Investigaciones en Microbiología y Biotecnología-UR (CIMBIUR), Facultad de Ciencias Naturales, Universidad del Rosario, Bogotá, Colombia | Carolina Ramírez-Santana; Gustavo Salguero; Juan David Ramírez; Juan Esteban Gallo; Juan-Manuel Anaya; Luz H. Patiño; Marina Muñoz; Nathalia Ballesteros; Sergio Castañeda |
| EPI_ISL_2031726 | Centro de Innovación en Vigilancia Epidemiológica (CiVE), Institut Pasteur Montevideo, Uruguay | Centro de Innovación en Vigilancia Epidemiológica (CiVE), Institut Pasteur Montevideo, Uruguay | Alicia Costábile; Alvaro Fajardo; Andrés Lizosaín; Belén González; Bernardina Rivera; Cecilia Alonso; Cecilia Salazar; Gonzalo Moratorio; Gregorio Iraola; Henry Albornoz; Ignacio Ferrés; Inés Bellini; Juan Zanetti; Julio Medina; Lucia Bilbao; Luciana Griffero; Lucía Spangenberg; Ma Noel Bentancor; Ma Pía Techera; Mailen Arleo; Martina Alonso; María José Benítez; Matías Maidana; Mauricio Méndez; Melissa Duquía; Mercedes Paz; Natalia Rego; Natalia Reyes; Odhille Chappos; Paula Perbolianachis; Pilar Moreno; Rodney Colina; Rodrigo Arce; Tamara Fernández; Tania Possi |
| EPI_ISL_941116, EPI_ISL_941123, EPI_ISL_941139 | Centro de Investigaciones en Microbiología y Biotecnología-UR (CIMBIUR), Facultad de Ciencias Naturales, Universidad del Rosario, Bogotá, Colombia | Centro de Investigaciones en Microbiología y Biotecnología-UR (CIMBIUR), Facultad de Ciencias Naturales, Universidad del Rosario, Bogotá, Colombia Icahn School of Medicine at Mount Sinai, New York, USA | Adriana van de Guchte; Alberto Paniz-Mondolfi; Alejandro Feged-Rivadeneira; Ana S. Gonzalez-Reiche; Andrés Angel; Carolina Flórez; Carolina Hernández; Emilia Mia Sordillo; Felipe González-Casabianca; Hala Alejel Alshammary; Harm van Bakel; Iván Carroll; Jaime Cascante; Jayeeta Dutta; Juan David Ramírez; Luz Helena Patiño; Marina Muñoz; Matthew M. Hernandez; Mauricio Santos-Vega; Mónica Palma-Cuero; Nathalia Ballesteros; Sergio Gomez; Viviana Simon; Zenab Khan |
| EPI_ISL_941982 | Centro de Investigaciones en Microbiología y Biotecnología-UR (CIMBIUR), Facultad de Ciencias Naturales, Universidad del Rosario, Bogotá, Colombia | Centro de Investigaciones en Microbiología y Biotecnología-UR (CIMBIUR), Facultad de Ciencias Naturales, Universidad del Rosario, Bogotá, Colombia Instituto Nacional de Salud, Bogotá, Colombia Icahn School of Medicine at Mount Sinai, New York, USA | Adriana van de Guchte; Alberto Paniz-Mondolfi; Ana S. Gonzalez-Reiche; Carolina Flórez; Carolina Hernández; Emilia Mia Sordillo; Hala Alejel Alshammary; Harm van Bakel; Jayeeta Dutta; Juan David Ramírez; Luz Helena Patiño; Marina Muñoz; Matthew M. Hernandez; Nathalia Ballesteros; Sergio Gomez; Viviana Simon; Zenab Khan |
| EPI_ISL_491941 | Centro de Investigaciones, Universidad de Especialidades Espíritu Santo | Institute of Microbiology, Universidad San Francisco de Quito | Belén Prado-Vivar; Bernardo Gutiérrez; Derly Andrade; Edith Lopez; Fernando Espinoza; Gabriel Morey; Gabriel Trueba; Jose Pedro Barberan; Juan Carlos Fernandez; Juan José Guadalupe; Michelle Grunauer; Monica Becerra-Wong; Patricio Rojas-Silva; Paúl Cárdenas; Ruben Armas; Sully Márquez; Verónica Barragán |
| EPI_ISL_2442098, EPI_ISL_2442113 | Centro de Investigação em Saúde de Manhiça (CISM) & ISGlobal, Institut de Salut Global de Barcelona | Instituto de Biomedicina de València - CSIC | Alfredo Mayor; Arsenia Massinga; Inacio Mandomando; Irving Cancino; Iñaki Comas; Manoli Torres Puente; Santiago Jiménez-Serrano |
| EPI_ISL_450523 | Centrālā Laboratorija | Latvian Biomedical Research and Study Centre | Ivars Silamiķelis; Jana Osīte; Jānis Kloviņš; Kaspars Megnis; Marta Priedīte; Monta Ustinova; Stella Lapiņa; Uga Dumpis; Vita Rovīte; Ņikita Zrelovs |
| EPI_ISL_2798876, EPI_ISL_2798895, EPI_ISL_2798922, EPI_ISL_2798996, EPI_ISL_2799000, EPI_ISL_2799099, EPI_ISL_2799215, EPI_ISL_2799803 | | | |
| see above | Centrālā Laboratorija, SIA | Riga East University Hospital, National Microbiology Reference Laboratory; Eurofins Genomics Europe Sequencing GmbH | Arzu Algulieva; Diāna Dušacka; Dārta Pūpola; Ilva Pole; Jana Osīte; Jevgenijs Bodrenko; Jūlija Čevere; Reinis Vangravs; Reinis Zeltmatis; Sergejs Ņikišins; Stella Lapiņa; Ģirts Šķenders |
| EPI_ISL_486412, EPI_ISL_492988 | Centrālā laboratorija | Latvian Biomedical Research and Study Centre | Ivars Silamiķelis; Jana Osīte; Jānis Kloviņš; Kaspars Megnis; Marta Priedīte; Monta Ustinova; Stella Lapiņa; Uga Dumpis; Vita Rovīte; Ņikita Zrelovs |
| EPI_ISL_861489 | Cerballiance Côte d'Azur | CERBA LAB | Adjtoutah Z; Mangiarotti T; Prots L. |
| EPI_ISL_2646262 | Cerballiance-IDF | Cerba lab | Aude Lessenne; Bénédicte Roquebert; Emmanuel Lecorche; Kader Merah; Laura Verdurme; Patrice Herisson; Sabine Trombert-Paolantoni; Stéphanie Haïm-Boukobza; Thierry Collin |
| EPI_ISL_2504131, EPI_ISL_2504133, EPI_ISL_2504138, EPI_ISL_2504139, EPI_ISL_2504141, EPI_ISL_2504142 | Chantal Biya International Reference Centre (CIRCB) | International Centre for Genetic Engineering and Biotechnology (ICGEB) and ARGO Open Lab for Genome Sequencing | Alessandro Marcello; Alexis Ndjolo; Celine Nkenfou; Collins Chenwi; Danilo Licastro; Desire Takou; Emanuele Orsini; Grace Angong Belournou; Joseph Fokam; Ndine Fainguem; Simeone Dal Monego |
| EPI_ISL_753955, EPI_ISL_754011 | Charité Universitätsmedizin Berlin, Institut für Virologie/Labor Berlin | Charité Universitätsmedizin Berlin, Institut für Virologie | Barbara Mühlemann; Christian Drosten; Julia Schneider; Jörn Beheim-Schwarzbach; Talitha Veith; Terry Jones; Victor M Corman |
| EPI_ISL_2987446 | Chattogram Maa-O-Shishu Hospital / Disease Biology & Molecular Epidemiology Research Lab, University of Chittagong | Virology Laboratory, International Centre for Diarrhoeal Disease Research, Bangladesh (ICDDR,B) | Adnan Mannan; Fahim Reza; H.M. Hamidullah Mehedi; Md. Abdur Rob; Md. Minhazul Hoque; Mohammad Enayet Hossain; Mojnu Miah; Mustafizur Rahman; Nahid Sultana; Rajdeep Biswas; Rashedul Hasan; Sanjoy Biswas |
| EPI_ISL_735497 | Chattogram Veterinary and Animal Sciences University | Central Biological Research Laboratory and Department of Biochemistry and Molecular Biology | H. M. Abdullah Al Masud; Imam Hossen; Md. Arif Hossain; Md. Imranul Hoq; Md. Khondakar Raziur Rahman; Md. Omer Faruq; Mohammad Omar Faruque; Robiul Hasan Bhuiyan; Sajib Rudra; Shanta Paul |
| EPI_ISL_1211989 | Chemisches und Veterinäruntersuchungsamt Ostwestfalen-Lippe | Robert Koch Institute | |
| EPI_ISL_419297, EPI_ISL_419298, EPI_ISL_419309 | Chiba Prefectural Institute of Public Health | Pathogen Genomics Center, National Institute of Infectious Diseases | Hajime Kamiya; Kentaro Itokawa; Makoto Kuroda; Masakatsu Taira; Masanori Hashino; Motoi Suzuki; Rina Tanaka; Tsuyoshi Sekizuka; Yushi Hachisu |
| EPI_ISL_700332, EPI_ISL_906087, EPI_ISL_1360422, EPI_ISL_1634454, EPI_ISL_1634455, EPI_ISL_2446012, EPI_ISL_2692860, EPI_ISL_2692865, EPI_ISL_2692876 | | | |
| see above | Child Health Research Foundation | Child Health Research Foundation | Afroza Akter Tanni; Arif Mohammad Tanmoy; CHRF Bangladesh Genomics Team; Md Hafizur Rahman; Roly Malaker; Samir K Saha; Senjuti Saha; Sharmistha Goswami; Syed Muktadir Al Sium |
| EPI_ISL_2445988 | Chittagong General Hospital | Child Health Research Foundation | Adnan Mannan; CHRF Bangladesh Genomics Team; H. M. Hamidullah Mehedi; Md. Abdur Rob; Md. Minhazul Hoque; Rajdeep Biswas |
| EPI_ISL_1970348 | Chongqing International Travel Health Care Center | Chongqing Municipal Center for Disease Control and Prevention | Dong Wang; Haiyan Wen; Hua Ling; Lan Zhou; Mingyue Wang; Rong Rong; Sheng Ye; Shuang Chen; Wenge Tang; Yun Tang; Zhangping Tan; Zhen Yu |
| EPI_ISL_1970347 | Chongqing Yuzhong District Center For Disease Control And Prevention | Chongqing Municipal Center for Disease Control and Prevention | Hua Ling; Jiaqi Li; Mingyue Wang; Rong Rong; Sheng Ye; Shuang Chen; Wenge Tang; Yingbing Zhou; Yun Tang; Yuyue Luo; Zhangping Tan; Zhen Yu |
| EPI_ISL_3038908 | Chulabhorn Hospital | National Institute of Health, Department of Medical Sciences, Ministry of Public Health, Thailand | ; Natchaya Khiadsang; Nuttida Thongpramul; Pakorn Piromtong; Pilailuk Okada; Ratana Tacharoenmuang; Siripaporn Phuygun; Sittiporn Parnmen; Sunthareeya Waicharoen; Thanutsapa Thanadachakul; Warawan Wongboot; sirikanda wimol |
| EPI_ISL_2248847, EPI_ISL_2248852, EPI_ISL_2248853, EPI_ISL_2445744 | Cintramedica | Instituto Nacional de Saude (INSA) | Borges et al |
| EPI_ISL_1722998, EPI_ISL_1844396 | Clemenshospital - Alexianer Clemenshospital MÃ¼nster | Robert Koch Institute | |
| EPI_ISL_1363787 | Clin & Gen Lab | Microbiologia Molecular, Instituto SELADIS, Universidad Mayor de San Andrés | Aneth Vasquez Michel; Oscar M. Rollano-Peñaloza |
| EPI_ISL_1250842 | Clin & Gen Lab | Molecular Genetics Laboratory, Instituto de Investigaciones Químicas, Universidad Mayor de San Andrés | Aneth Vasquez Michel; Oscar M. Rollano-Peñaloza |
| EPI_ISL_2987572 | Clinic of infective diseases | Laboratory of virology and molecular diagnostics, Institute of Public Health | Boshevska G; Janchevska E.; Kuzmanovska M |
| EPI_ISL_462450, EPI_ISL_462464 | Clinical Center, University of Sarajevo | Charite Universitatsmedizin Berlin, Institute of Virology | Almedina Hadzihasanovic-Moro; Amela Dedeic-Ljubovic; Barbara Muehlemann; Christian Drosten; Irma Salimovic-Besic; Jorn Beheim-Schwarzbach; Julia Schneider; Selma Mutevelic; Suzana Arapcic; Talitha Veith; Terry Jones; Victor M Corman |
| EPI_ISL_1300651, EPI_ISL_2938102, EPI_ISL_2938104, EPI_ISL_2938106, EPI_ISL_2938118 | Clinical Center, University of Sarajevo; Unit for Clinical Microbiology | Clinical Center, University of Sarajevo; Unit for Clinical Microbiology | Amela Dedeić-Ljubović; Edina Zahirović; Golubinka Boshevska; Irma Salimović-Bešić; Maja Kuzmanovska; Sandra Vegar-Zubović; Sebija Izetbegović; Suzana Arapčić |
| EPI_ISL_2652206, EPI_ISL_2652209, EPI_ISL_3021385, EPI_ISL_3021390 | Clinical Emergency County Hospital, Craiova | Cantacuzino Institute Virology | Carmen Cherciu; Luiza Ustea; Mihaela Lazar; Mihaela Oprea; Nicoleta Paraschiv; Sorin Dinu |
| EPI_ISL_516431, EPI_ISL_678259 | Clinical Hospital - Shtip | Research Center for Genetic Engineering and Biotechnology "Georgi D. Efremov" , Macedonian Academy of Sciences and Arts | RCGEB - MASA |
| EPI_ISL_3020158 | Clinical Hospital of Infectious Diseases „Sfanta Parascheva" Iasi | National Institute of Infectious Diseases-Prof. Dr. Matei Bals Molecular Diagnostics Laboratory | Corina Casangiu; Dan Otelea; Leontina Banica; Marius Surleac; Petre Milu; Robert Hohan; Simona Paraschiv |
| EPI_ISL_447454 | Clinical Microbiology Laboratory, Sheba Medical Center | Stern Lab | Stern Lab |
| EPI_ISL_968146, EPI_ISL_1334559 | Clinical Molecular Microbiology Laboratory, UNC Hospital | Dirk Dittmer | Andrea McDaniel-Harper; Angelica Juarez; Ashwini Roy-Chaudhury; Billy Fisher; Blossom Damania.; Brent A. Eason; Cameroon Grant; Carolina Caro-Vegas; Cecilia Thompson; Danielle Chappell; Dirk Dittmer; Evelyn Hoffman; Jason Smedberg; Jason Wong; Justin T. Landis; Linda Pluta; Melissa B. Miller; Patricio Cano; Razia Moorad; Ryan McNamara; Shawn Hawken |
| EPI_ISL_2535703 | Clinical Research Centre Hospital Sibu | Institute of Health and Community Medicine | Chan Chia Jui; Chua Hock Hin; David Perera; Ooi Mong How; Tonnii Sia Loong Loong; Wong Jyn Shan; Wong Kiing Aik |
| EPI_ISL_2608412 | Clinical Research Centre, Sarawak General Hospital | Institute of Health and Community Medicine | Chan Chia Jui; Chua Hock Hin; David Perera; Ooi Mong How; Tonnii Sia Loong Loong; Wong Jyn Shan; Wong Kiing Aik |
| EPI_ISL_581842, EPI_ISL_581843, EPI_ISL_830804, EPI_ISL_930860, EPI_ISL_2102098, EPI_ISL_2270056, EPI_ISL_2768083 | | | |

| | | | |
|---|---|---|---|
| see above | Clinical Virology | Clinical Bacteriology | Adrian Egli; Alexander Gensch; Alfredo Mari; Christian Nickel; Fanny Wegner; Hans Hirsch; Hans Pargger; Helena MB Seth-Smith; Julia Bielicki; Karoline Leuzinger; Kirstine K. Soegaard; Madlen Stange; Manuel Battegay; Martin Siegemund; Michael Osthoff; Michael Schweitzer; Myrta Brunner; Rita Schneider-SliFemalea; Rita Schneider-Sliwa; Roland Bingisser; Sarah Tschudin-Sutter; Simon Fuchs; Stefano Bassetti; Tim Roloff |
| EPI_ISL_2035560 | Clinical Virology | Clinical Virology | Anissa Chouikha; Henda Triki; Kais Ghedira; Mariem Gdoura; Sondos Haddad; Wasfi Fares |
| EPI_ISL_605804 | Clinical Virology Laboratory, Institute of Liver and Biliary Sciences | ILBS - IGIB | Abhishek Padhi; Ekta Gupta; Jaswinder Singh Maras; Reshu Agarwal; Sheetalnath Rooge; Shridhar Sivasubbu; Shvetank Sharma; Vinod Scaria |
| EPI_ISL_2370909 | Clinical and Basic Functional Sciences, Department of Microbiology | Greek Genome Center, Biomedical Research Foundation of the Academy of Athens (BRFAA) | Dimitrios Thanos; Emmanouil Athanasiadis; Giannis Vatsellas; Katerina Zoi; Konstantina Gartzonika; Theodoros Loupis |
| EPI_ISL_1397826 | Columbia University Irving Medical Center | Wadsworth Center, New York State Department of Health | Alexis Russel; Daryl M. Lamson; Erasmus Schneider; Erica Lasek-Nesselquist; John Kelly; Jonathan Plitnick; Kirsten St. George; Matthew Shudt; Melissa A Leisner; Navjot Singh |
| EPI_ISL_2383896 | Commonwealth Healthcare Center | Centers for Disease Control and Prevention Division of Viral Diseases, Pathogen Discovery | Alison Laufer Halpin; Ben L. Rambo-Martin; Clinton R. Paden; Dakota Howard; Darlene Wagner; Dave Wentworth; Dhwani Batra; Jasmine Padilla; Justin Lee; Katie Dillon; Krista Queen; Kristen Knipe; Kristine Lacek; Mark Burroughs; Matthew Schmerer; Mili Sheth; Peter Cook; Sam Shepard; Sarah Nobles; Shoshona Le; Suxiang Tong; Vivien Dugan; Yvette Unoarumhi |
| EPI_ISL_632271, EPI_ISL_632285, EPI_ISL_682300, EPI_ISL_1660433, EPI_ISL_1660444, EPI_ISL_1660449, EPI_ISL_1660464, EPI_ISL_1660470, EPI_ISL_2466680, EPI_ISL_2622084, EPI_ISL_2663322, EPI_ISL_2663323, EPI_ISL_2715285, EPI_ISL_2715288, EPI_ISL_2715289, EPI_ISL_2812541, EPI_ISL_2932623, EPI_ISL_2932641, EPI_ISL_2932642, EPI_ISL_2932655, EPI_ISL_2932663, EPI_ISL_2932666, EPI_ISL_2932669, EPI_ISL_2932673, EPI_ISL_2932678, EPI_ISL_2992091 | | | |
| see above | Communicable Disease Laboratory, Public Health Directorate | Communicable Disease Laboratory, Public Health Directorate | AlAbbas, Z.; AlHujairi, Z.; AlTaif, Z.; AlWasti, H.; Almoamen, G.; Altaif, Z.; Alwasti, H.; Marhoon, A. |
| EPI_ISL_2375921 | Concordia Clinic | NHLS/UCT | Arash Iranzadeh; Bruna Galvao; Carolyn Williamson; Deelan Doolabh; Diana Hardie; Innocent Mudau; Kruger Marais; Lynn Tyers; Marvin Hsiao; Stephen Korsman |
| EPI_ISL_454110 | ControlVet | Instituto Nacional de Saude (INSA) | Borges et al |
| EPI_ISL_2636156 | Corona-Testzentrum ifp Institut für Produktqualität GmbH | Robert Koch Institute | |
| EPI_ISL_3014093, EPI_ISL_3014108, EPI_ISL_3014126 | Cotugno | TIGEM | Antonio Grimaldi Patrizia Annunziata Francesco Panariello Biancamaria Pierri Claudia Tiberio Teresa Giuliano Valentina Bouche Chiara Colantuono Maria Concetta Cuomo Denise Di Concilio Lucio Di Filippo Anna Manfredi Marcello Salvi Antonio Limone Luigi Atripaldi Pellegrino Cerino Andrea Ballabio Davide Cacchiarelli |
| EPI_ISL_1084757 | Croatian Institute of Public Health | Croatian Institute of Public Health | Irena Tabain; Ivana Ferenčak |
| EPI_ISL_1695973 | DC Public Health Lab/ Dept. of Forensic Sciences | Centers for Disease Control and Prevention Division of Viral Diseases, Pathogen Discovery | Alison Laufer Halpin; Ben L. Rambo-Martin; Clinton R. Paden; Dakota Howard; Darlene Wagner; Dave Wentworth; Dhwani Batra; Jasmine Padilla; Justin Lee; Katie Dillon; Krista Queen; Kristen Knipe; Kristine Lacek; Mark Burroughs; Matthew Schmerer; Mili Sheth; Peter Cook; Sam Shepard; Sarah Nobles; Shoshona Le; Suxiang Tong; Vivien Dugan; Yvette Unoarumhi |
| EPI_ISL_1094341 | DC Public Health Lab/ Dept. of Forensic Sciences | Respiratory Viruses Branch, Division of Viral Diseases, Centers for Disease Control and Prevention | Anna Montmayeur; Anna Uehara; Ben L. Rambo-Martin; Clinton R. Paden; Dhwani Batra; Haibin Wang; Jasmine Padilla; Jing Zhang; Justin Lee; Krista Queen; Lori Rowe; Mark Burroughs; Mili Sheth; Peter W. Cook; Rachel Marine; Sarah Nobles; Suxiang Tong; Yan Li; Ying Tao |
| EPI_ISL_2779433 | DFMH | USAMRD-A, Basic Science Laboratory | Alan Lemtudo; Beth Mutai; Brian Andika; Carol Kifude; Clement Masakwe; Eric Muthanje; Esther Omuseni; Faith Sigei; Gathii Kimita; George Awinda; John Waitumbi; Josphat Nyataya; Rachel Githii; Rehema Liyai; Stephen Ochola |
| EPI_ISL_2342561, EPI_ISL_2342562, EPI_ISL_2342563 | DNA Laboratories Sdn Bhd | Malaysia Genome Institute | Azrin Ahmad; Enizza Kasim; Irni Suhayu Sapian; Mohd Faizal Abu Bakar; Mohd Noor Mat Isa; Nor Azfa Johari.; Nurhezreen Md Iqbal; Shamsidar Sopie; Siti Noraini Othman; Wong Yong Wee; Yusuf Muhammad Noor |
| EPI_ISL_2983222, EPI_ISL_2983224, EPI_ISL_2987211, EPI_ISL_2987214 | DNA Labs | National Reference Laboratory, Nigeria Centre for Disease Control | Anthony Ahumibe; Catherine Okoi; Chimaobi Chukwu; Dr Chikwe Ihekweazu; Dr Ndodo Nnaemeka; Dr Omoare Adesuyi; Nwando Mba; Olusola Anuoluwapo Akanbi |
| EPI_ISL_683829 | DOHMH Central Harlem | New York City Public Health Laboratory | Jade Wang; et al. |
| EPI_ISL_823949 | DOHMH Jamaica | New York City Public Health Laboratory | Jade Wang; et al. |
| EPI_ISL_857252 | DOHMH Morrisania | New York City Public Health Laboratory | Jade Wang; et al. |
| EPI_ISL_745438 | DOHMH PHL | New York City Public Health Laboratory | Jade Wang; et al. |
| EPI_ISL_1181670, EPI_ISL_1511145 | DPHL | Delaware Public Health Lab | Gregory Hovan; Rebecca Savage |
| EPI_ISL_2593764, EPI_ISL_2595726, EPI_ISL_2595860, EPI_ISL_2596341, EPI_ISL_2596803, EPI_ISL_2597228, EPI_ISL_2597229, EPI_ISL_2597312, EPI_ISL_2612300 | | | |
| see above | DSMRC | DSMRC | Khine Zaw Oo; Ko Ko Lwin; Ko Ko Win; Kyee Myint; Nay Myo Aung; Pai Khant Kyaw; Phyo Kyaw Aung; Thet Wai Oo; Zaw Win Htun |
| EPI_ISL_2289557 | DYOMEDEA NEOLAB | CNR Virus des Infections Respiratoires - France SUD | Antonin Bal; Bruno Lina; Bruno Simon; Gregory Destras; Gwendolyne Burfin; Hadrien Regue; Laurence Josset; Martine Valette; Quentin Semanas |
| EPI_ISL_2894008 | DYOMEDEA-LABORATOIRE DE LA SAUVEGARDE | CNR Virus des Infections Respiratoires - France SUD | Antonin Bal; Bruno Lina; Gregory Destras; Gwendolyne Burfin; Hadrien Regue; Laurence Josset; Martine Valette; Quentin Semanas |
| EPI_ISL_416542, EPI_ISL_416543 | Dasman Diabetes Institute | Dasman Diabetes Institute | Ebaa AlOzairi; Fahd Al-Mulla; Motasem Melhem; Qais Al-Duwairi; Rasheeba Iqbal; Sara Al-Qabandi; Sumi John |
| EPI_ISL_416541 | Dasman Diabetes Institute and Virology Laboratory Ministry of Health | Dasman Diabetes Institute | Ebaa AlOzairi; Fahd Al-Mulla; Motasem Melhem; Qais Al-Duwairi; Rasheeba Iqbal; Sara Al-Qabandi; Sumi John |
| EPI_ISL_2140727 | De Rust (Blommenek) Clinic wc DRC | NHLS/UCT | Arash Iranzadeh; Bruna Galvao; Carolyn Williamson; Deelan Doolabh; Diana Hardie; Innocent Mudau; Kruger Marais; Lynn Tyers; Marvin Hsiao; Stephen Korsman |
| EPI_ISL_2372406, EPI_ISL_2648206, EPI_ISL_2648207, EPI_ISL_2648208, EPI_ISL_2648209, EPI_ISL_2648211, EPI_ISL_2648213, EPI_ISL_2648214, EPI_ISL_2648215, EPI_ISL_2648217, EPI_ISL_2648218, EPI_ISL_2648219, EPI_ISL_2648220 | | | |
| see above | Debswana Orapa Mine Hospital Laboratory | Botswana Harvard HIV Reference Laboratory | Boitumelo Zuze; Botshelo Radibe; Dorcas Maruapula; Joseph Makhema; Keoratile Ntshambiwa; Kgomotso Moruisi; Koketso Maotwe; Legodile Kooepile; Lesedi Magama; Madisa Mine; Mosepele Mosepele; Mpo Molapisi; Ontlametse T. Bareng; Pamela Smith-Lawrence; Roger Shapiro; Shahin Lockman; Sikhulile Moyo; Simani Gaseitsiwe; Thongbotho Mphoyakgosi; Wonderful T. Choga |
| EPI_ISL_812293 | Delaware Public Health Lab | Delaware Public Health Lab | Gregory Hovan |
| EPI_ISL_693691, EPI_ISL_693730 | Delaware Public Health Laboratory | Delaware Public Health Laboratory | Gregory Hovan |
| EPI_ISL_2432518, EPI_ISL_2432922, EPI_ISL_2432953 | Demy Health | National Reference Laboratory, Nigeria Centre for Disease Control | Anthony Ahumibe; Catherine Okoi; Chimaobi Chukwu; Dr Chikwe Ihekweazu; Dr Ndodo Nnaemeka; Dr Omoare Adesuyi; Grace Esebanmen; Naidoo Dhamari; Nwando Mba; Olusola Anuoluwapo Akanbi |
| EPI_ISL_430796 | Departamento de Biología y genética molecular, IACA Laboratorios. | Área de Secuenciación del Laboratorio de Virología del Hospital de Niños Dr. Ricardo Gutierrez on behalf of 'Proyecto Argentino Interinstitucional de genomica de SARS-CoV-2' (PAIS Consortium) | A; AS; E; Goya; LE; Lusso; MI; MS; Masciovecchio MV; Mistchenko; Nabaes Jodar; Natale; S; Streitenberger ER; Suárez; Tittarelli; Valinotto; Viegas, M. |
| EPI_ISL_949238 | Departamento de Microbiología, CDB, Hospital Clínic, Barcelona | SeqCOVID-SPAIN consortium/IBV(CSIC) | Aida Peiró and SeqCOVID-SPAIN consortium; Andrea Vergara; Elisa Rubio; Jéssica Navero; Mikel Martínez |
| EPI_ISL_1340753, EPI_ISL_1340755, EPI_ISL_1340756, EPI_ISL_1340758 | Departamento de Virologia, Laboratorio Central de Salud Pública, Avenida Venezuela y Teniente Escurra, Asunción, Paraguay | Laboratory of Respiratory Viruses and Measles, Oswaldo Cruz Institute, FIOCRUZ | Alice Sampaio Rocha; Ana Carolina Mendonca; Ana Carolina Paixao; Cynthia Vazquez; Fernando Motta; Luciana Appolinario; Marilda Siqueira on behalf of the Fiocruz COVID-19 Genomic Surveillance Network; Paola Resende; Renata Serrano Lopes |
| EPI_ISL_516923 | Department for Molecular Diagnostics, Centre for Medical Microbiology, Institute of Public Health of Montenegro | Charité Universitätsmedizin Berlin, Institut für Virologie | Barbara Muehlemann; Christian Drosten; Julia Schneider; Jörn Beheim-Schwarzbach; Marija Govedarica and Danijela Vujošević; Talitha Veith; Terry Jones; Victor M Corman |
| EPI_ISL_471529, EPI_ISL_477169, EPI_ISL_754180, EPI_ISL_1914665, EPI_ISL_1914938, EPI_ISL_1921859, EPI_ISL_3204236, EPI_ISL_3204239, EPI_ISL_3204245 | | | |
| see above | Department for Virology, Molecular Biology and Genome Research, R. G. Lugar Center for Public Health Research, National Center for Disease Control and Public Health (NCDC) of Georgia. | Department for Virology, Molecular Biology and Genome Research, R. G. Lugar Center for Public Health Research, National Center for Disease Control and Public Health (NCDC) of Georgia. | Adam Kotorashvili; Amiran Gamkrelidze.; Ana Papkiauri; Ann Machablishvili; Anna Kasradze; Davit Tsaguria; Ekaterine Khmaladze; Ekaterine Zangaladze; Ekaterine Zhghenti; Giorgi Gogoladze; Giorgi Tomashvili; Gvantsa Brachveli; Gvantsa Chanturia; Irma Burjanadze; Ketevan Sidamonidze; Khatuna Zakhashvili; Lela Sabadze; Lela Urushadze; Magda Dgebuadze; Maia Alkhazashvili; Mari Gavashelidze; Mariam Zakalashvili; Marine Murtskhvaladze; Meri Pantsulaia; Nato Kotaria; Nino Berishvili; Paata Imnadze; Roena Sukhiashvili; Salome Javashvili; Tamar Jashiashvili; Tata Imnadze; Tea Tevdoradze |
| EPI_ISL_1642768 | Department fÃ¼r Labormedizin Abteilung III Bereich Molekulare Diagnostik UniversitÃ¤tsklinikum Halle(Saale) | Robert Koch Institute | |
| EPI_ISL_2723562, EPI_ISL_2896229 | Department of Acute Infectious Diseases Control and Prevention, Yunnan Provincial Center for Disease Control and Prevention | Department of Acute Infectious Diseases Control and Prevention, Yunnan Provincial Center for Disease Control and Prevention | Jienan Zhou; Meiling Zhang; Senquan Jia; Xiaoqing Fu; Zhaosheng Liu |
| EPI_ISL_2429651, EPI_ISL_2665812, EPI_ISL_2666405, EPI_ISL_2667413, EPI_ISL_2669906, EPI_ISL_2670493, EPI_ISL_2670876, EPI_ISL_2671330, EPI_ISL_2680019, EPI_ISL_2684126, EPI_ISL_2819459, EPI_ISL_2854368, EPI_ISL_2854578, EPI_ISL_2862744, EPI_ISL_2971522, EPI_ISL_2992466, EPI_ISL_2992648, EPI_ISL_3029549 | | | |
| see above | Department of Bacteria, Parasites and Fungi, Statens Serum Institut, Copenhagen, Denmark | Statens Serum Institut Bioinformatics and Microbial Genomics | Danish Covid-19 Genome Consortium |
| EPI_ISL_515082 | Department of Biochemistry, Cell and Molecular Biology | WACCBIP, University of Ghana | A.K.; Adu, B.; Amenga-Etego; Ampofo, W.; Amuzu; Anang; Arjarquah, A.; Asante, I.; Awandare; Bediako, Y.; Boatemaa, L.; Bonney, E.; Bonney, K.; C.M.; D.S.; Eshun, M.; G.A.; G.B.; J.K.; J.M.; Kotey, E.; Kumordjie, S.; Kyei; L.N.; Magnussen, V.; Morang'a; Mutungi; Ngoi; Quashie, P.; Tei-Maya, F. |
| EPI_ISL_884839, EPI_ISL_884841, EPI_ISL_884856 | Department of Biochemistry, Cell and Molecular Biology, West African Centre for Cell Biology of Infectious Pathogens (WACCBIP), University of Ghana | Department of Biochemistry, Cell and Molecular Biology, West African Centre for Cell Biology of Infectious Pathogens (WACCBIP), University of Ghana | A.-K.; A.B.; Abass; Akoriyea; Amenga-Etego; Amoako, E.; Amuzu; Awandare; Bediako, Y.; Boakye; C.M.; D.S.; Diallo; G.A.; J.M.; Kibinge, N.; Kumi-Ansah, F.; L.N.; Magnussen, V.; Mohammed, A.; Morang'a; Ngoi; O.D.; Odoom, T.; Quashie, P.; S.K.; Tapela, K.; Tei-Maya, F. |
| EPI_ISL_421195, EPI_ISL_540509, EPI_ISL_678471, EPI_ISL_737311, EPI_ISL_1708595, EPI_ISL_1826581, EPI_ISL_1826750 | | | |
| see above | Department of Clinical Microbiology | GIGA Medical Genomics | Axelle Chaslain; Bouchra Boujemla; Cécile Meex; Céline Fombellida-Lopez; Keith Durkin; Maria Artesi; Marie-Pierre Hayette; Nathalie Renotte; Pierrette Melin; Raphaël Boreux; Sébastien Bontems; Vincent Bours; Vincent Bours. |

| | | | |
|---|---|---|---|
| EPI_ISL_419216 | Department of Clinical Pathology, Pamela Youde Nethersole Eastern Hospital | Department of Health Technology and Informatics, Faculty of Health and Social Science, The Hong Kong Polytechnic University | Alan Ka-Lun WU; Alex Yat-Man HO; Barry Kin-Chung WONG; David Ho-Keung SHUM; Eugene Yuk-Keung TSO; Gilman Kit-Hang SIU; Hiu-Yin LAO; Kam-Tong YIP; Kenneth Siu-Sing LEUNG; Kingsley King-Gee TAM; Kit-Man SIN; Kitty Sau-Chun FUNG; Kwok-Cheung LUNG; Lam-Kwong LEE; Man-Chun CHAN; Ming-Pan CHOI; Miranda Chong-Yee YAU; Raymond Wai-To LIU; Sandy Ka-Yee CHAU; Shea Ping YIP; Tak-Lun QUE; Timothy Ting-Leung NG; Wai-Shing LEUNG; Wing Cheong YAM; Wing-Kin TO; Yuk-Yung NG |
| EPI_ISL_1731082 | Department of Genetics and Department of Microbiology, University Hospital Motol | Department of Microbiology, University Hospital Motol | Ales Briksi; Katerina Chuda; Katerina Pehlikova; Malgorzata Libik; Milan Macek; Ondrej Cinek; Pavel Drevinek; Petr Broz; Petr Hubacek; Zdenek Kepka |
| EPI_ISL_1280115 | Department of Genetics, Medirex | Laboratory of Genomics and Bioinformatics, Comenius University Science Park | Anna Gičová; Diana Rusňáková; Gabriel Minárik; Jaroslav Budiš; Miroslav Böhmer; Renáta Lukačková; Tatiana Sedláčková; Tomáš Szemes; Werner Krampl |
| EPI_ISL_1121891 | Department of Health & Mental Hygiene Crown Heights | New York City Public Health Laboratory | Jade Wang; et al. |
| EPI_ISL_1018120, EPI_ISL_1018293, EPI_ISL_1018326, EPI_ISL_2710858, EPI_ISL_2713166, EPI_ISL_2713181, EPI_ISL_2713213, EPI_ISL_2713247, EPI_ISL_2713256, EPI_ISL_2713258, EPI_ISL_2713309, EPI_ISL_2723759, EPI_ISL_2723767, EPI_ISL_2723768, EPI_ISL_2860793, EPI_ISL_3039610 | | | |
| see above | Department of Health Technology and Informatics, The Hong Kong Polytechnic University | Department of Health Technology and Informatics, The Hong Kong Polytechnic University | Alan Ka-Lun Wu; Alex Yat-Man Ho; Barry Kin-Chung Wong; Chloe Toi-Mei Chan; David Ho-Keung Shum; Denise Sze-Hang Wong; Gilman Kit-Hang Siu; Hiu-Yin Lao; Hoi-Ching Jim; Ivan Tak-Fai Wong; Jake Siu-Lun Leung; Kam-Tong Yip; Kenneth Siu-Sing Leung; Kingsley King-Gee Tam; Kitty Sau-Chun Fung; Kristine Luk; Lam-Kwong Lee; Miranda Chong-Yee Yau; Sandy Ka-Yee Chau; Shea Ping Yip; Tak-Lun Que; Timothy Ting-Leung Ng; Wing Cheong Yam; Wing-Hei Lo; Wing-Kin To; Yvette Wai-Man Lai |
| EPI_ISL_408430 | Department of Infectious and Tropical Diseases, Bichat Claude Bernard Hospital, Paris | National Reference Center for Viruses of Respiratory Infections, Institut Pasteur, Paris | Angela Brisebarre; Flora Donati; Marion Barbet; Maud Vanpeene; Mélanie Albert; Méline Bizard; Sylvie Behillil; Sylvie van der Werf; Vincent Enouf; Xavier Lescure; Yazdan Yazdanpanah |
| EPI_ISL_413019 | Department of Internal Medicine, Triemli Hospital | Institute of Medical Virology, University of Zurich | Alexandra Trkola; Andrea Zbinden; Fiona Steiner; Gerhard Eich; Jon Huder; Jürg Böni; Maryam Zaheri; Michael Huber; Patrick Redli; Riccarda Capaul; Stefan Schmutz; Verena Kufner |
| EPI_ISL_1965488, EPI_ISL_2137132 | Department of Laboratory Medicine, Division of Clinical Virology, University of Medicine, Vienna | Bergthaler laboratory, CeMM Research Center for Molecular Medicine of the Austrian Academy of Sciences | Andreas Bergthaler; Anna Schedl; Bekir Erguner; Benedikt Agerer; Christoph Bock; Fabian Amman; Jan Laine; Lukas Endler; Maelle Le Moing; Martin Senekowitsch; Michael Schuster; Petr Triska; Thomas Penz |
| EPI_ISL_422411, EPI_ISL_422417, EPI_ISL_447614, EPI_ISL_1667474 | Department of Laboratory Medicine, National Taiwan University Hospital | Microbial Genomics Core Lab, National Taiwan University Centers of Genomic and Precision Medicine | Chiao-Ling Li; Pei-Jer Chen; Shan-Chwen Chang; Shiou-Hwei Yeh; Sui-Yuan Chang; Ya-Yun Lai; You-Yu Lin |
| EPI_ISL_476795, EPI_ISL_538436, EPI_ISL_538458, EPI_ISL_648689, EPI_ISL_648761 | Department of Laboratory Medicine, Tan Tock Seng Hospital | Department of Laboratory Medicine, Tan Tock Seng Hospital | Barkham TMS; Chen YYC; Li C; Lim JX; Maurer-Stroh S; Nagarajan N; Sessions OM; Tang WY; Zair X |
| EPI_ISL_1599170, EPI_ISL_1820321, EPI_ISL_2138809 | Department of Medical Microbiology - section Molde, Molde Hospital | Norwegian Institute of Public Health, Department of Virology | Atiya R Ali; Debech Nadia; Engebretsen Serina Beate; Garcia Llorente Ignacio; Hilde Elshaug; Hilde Vollan; Jon Bråte; Kamilla Heddeland Instefjord; Karoline Bragstad; Kathrine Stene-Johansen; Marie Paulsen Madsen; Olav Hungnes; Pedersen Benedikte Nevjen; Rasmus Riis Kopperud |
| EPI_ISL_861719 | Department of Medical Microbiology, Hospital Pengajar Universiti Putra Malaysia | Malaysia Genome Institute | Avisha Richards; Azrin Ahmad; Enizza Kasim; Hui-Yee Chee; Irni Suhayu Sapian; Mohd Faizal Abu Bakar; Mohd Noor Mat Isa; Muhammad MI; Narcisse Joseph; Nor Azfa Johari; Nor Zahrin Hasran; Nurhezreen Md Iqbal; Shamsidar Sopie; Siti Noraini Othman; Syafinaz Amin-Nordin; Yusuf Muhammad Noor |
| EPI_ISL_1192188, EPI_ISL_1820301, EPI_ISL_1919387, EPI_ISL_1989268, EPI_ISL_2142789, EPI_ISL_2225820, EPI_ISL_2225844, EPI_ISL_2225890, EPI_ISL_2225891, EPI_ISL_2225993, EPI_ISL_2333513, EPI_ISL_2333514, EPI_ISL_2549126, EPI_ISL_2612273, EPI_ISL_2927874, EPI_ISL_3152014, EPI_ISL_3152024 | | | |
| see above | Department of Medical Microbiology, St. Olavs hospital | Norwegian Institute of Public Health, Department of Virology | 'Kathrine Stene-Johansen; Atiya R Ali; Debech Nadia; Engebretsen Serina Beate; Garcia Llorente Ignacio; Hilde Elshaug; Hilde Vollan; Jon Bråte; Kamilla Heddeland Instefjord; Karoline Bragstad; Kathrine Stene-Johansen; Line Victoria Moen; Marie Paulsen Madsen; Olav Hungnes; Pedersen Benedikte Nevjen; Rasmus Riis Kopperud |
| EPI_ISL_506999 | Department of Medical Microbiology, University Malaya Medical Centre | Department of Medical Microbiology, Faculty of Medicine, University of Malaya | I-Ching SAM; Jennifer Chong; University Malaya Medical Centre COVID Team; Yoke Fun CHAN; Yoong Min CHONG |
| EPI_ISL_512844 | Department of Medical Research | DMR_Myanmar | Aung Kyaw Kyaw; Aung Zaw Latt; Hlaing Myat Thu; Hnin Ohnmar Soe; Htin Lin; Kay Thi Aye; Lai Lai San; Myat Htut Nyunt; Nan Aye Thida Oo; Ni Ni Zaw; Phyu Win Ei; Su Mon Win; Theingi Win Myat; Wah Wah Aung; Yi Yi Kyaw; Zaw Than Htun |
| EPI_ISL_2547429 | Department of Medical Virology, School of Medicine Ahvaz Jundishapur University of Medical sciences | Genetics Research Center, University of Social Welfare and Rehabilitation Sciences | Ahmad Tavakoli; Azarakhsh Azaran; Farid yousefi; Hossein Najmabadi.; Kimia Kahrizi; Maryam Beheshtian; Marzieh Mohseni; Mohammad Farahmand; Mohsen Savaie; Seyed Mohammad Jazayeri; Zohreh Fattahi |
| EPI_ISL_437442 | Department of MicroBiology, Government Medical College, Surat | Gujarat Biotechnology Research Centre | Akanksha Verma; Amit Kanani; Ankit Hinsu; Apurvasinh Puvar; Bhavesh Modi; Binita Aring; Chaitanya Joshi; Dinesh Kumar; Dipa Kinariwala; Dipeshwari Shewale; Disha Patel; Gaurishankar Shrimali; Geeta Vaghela; Janvi Raval; Kairavi Joshi; Kamlesh J Upadhyay; Madhvi Joshi; Maharshi Pandya; Monika Gandhi; Neeta Khandelwal; Nidhi Sood; Nitin Savaliya; Pinal Trivedi; Pranay Shah; Pritesh Sabara; R D Dixit; Raghawendra Kumar; Ramesh Pandit; Snehal Bagatharia; Sonia Barve; Tejas Shah; Zuber Saiyed |
| EPI_ISL_1273103 | Department of Microbial Biotechnology, Genetic Engineering Division, National Research Centre | Department of Microbial Biotechnology, Genetic Engineering Division, National Research Centre | A.E. and Shemis, M.; A.G.; Dawood; Farghaly, A.; Fikry; Khalifa; M.K.; R.M.; Taha |
| EPI_ISL_1745697 | Department of Microbiology, Faculty of Medicine and Dentistry, Palacky University and University Hospital Olomouc | Institute of Molecular and Translational Medicine / Laboratory of Experimental Medicine, Faculty of Medicine and Dentistry, Palacky University | Barbora Blumová; Hana Jaworek; Marián Hajdúch; Pavel Sauer; Rastislav Slavkovský; Tomáš Pospíšíl; Vladimíra Koudeláková |
| EPI_ISL_2833646, EPI_ISL_2833648, EPI_ISL_2833649, EPI_ISL_2833650, EPI_ISL_2833651, EPI_ISL_2833652, EPI_ISL_2833653, EPI_ISL_2833654, EPI_ISL_2833655, EPI_ISL_2833657, EPI_ISL_2833658, EPI_ISL_2833659, EPI_ISL_2833661, EPI_ISL_2833663, EPI_ISL_2833664, EPI_ISL_2833665, EPI_ISL_2833666 | | | |
| see above | Department of Microbiology, National Institute for Public Health of Kosova | Charité Universitätsmedizin Berlin, Institut für Virologie | Aferdita Hyseni; Barbara Mühlemann; Blendi Jerliu; Christian Drosten; Donjeta Hajdari; Julia Schneider; Jörn Beheim-Schwarzbach; Nazmi Mehmeti; Pranvera Abazi; Talitha Veith; Terry Jones; Victor M Corman; Xhevat Jakupi; Zana Deva |
| EPI_ISL_1027643, EPI_ISL_1027652, EPI_ISL_1027653 | Department of Microbiology, National Institute for Public Health of Kosova | Charité Universitätsmedizin Berlin, Institut für Virologie | Barbara Mühlemann; Christian Drosten; Donjeta Hajdari; Julia Schneider; Jörn Beheim-Schwarzbach; Talitha Veith; Terry Jones; Victor M Corman; Xhevat Jakupi; Zana Deva |
| EPI_ISL_1034592, EPI_ISL_1034598, EPI_ISL_1034716, EPI_ISL_2423558 | Department of Microbiology, The University of Hong Kong | Department of Microbiology, The University of Hong Kong | Kelvin K.W. To; Kwok-Yung Yuen |
| EPI_ISL_1117809, EPI_ISL_1840981 | Department of Microbiology, University Innsbruck | Bergthaler laboratory, CeMM Research Center for Molecular Medicine of the Austrian Academy of Sciences | Andreas Bergthaler; Anna Schedl; Bekir Erguner; Benedikt Agerer; Christoph Bock; Fabian Amman; Jan Laine; Lukas Endler; Maelle Le Moing; Martin Senekowitsch; Michael Schuster; Petr Triska; Thomas Penz |
| EPI_ISL_463744, EPI_ISL_463745, EPI_ISL_1164626, EPI_ISL_1164627, EPI_ISL_1164628, EPI_ISL_1164649, EPI_ISL_1164662, EPI_ISL_1164664, EPI_ISL_1164671 | | | |
| see above | Department of Molecular Virology, Cyprus Institute of Neurology and Genetics | Department of Molecular Virology, Cyprus Institute of Neurology and Genetics | Anastasis Oulas; Andreas Hadjisavvas; Christina Christodoulou; Christina Tryfonos; Dana Koptides; Denise Alexandrou; George Krashias; George Spyrou; Jan Richter; Maria Loizidou; Mihalis Panayiotidis; Olga Kalakouta; Pavlos Fanis; Stavros Bashiardes |
| EPI_ISL_2692906 | Department of Pathology, School of Medicine, Imam Hospital Complex, Tehran University of Medical Sciences, Tehran, Iran | Genetics Research Center, University of Social Welfare and Rehabilitation Sciences | Ali Jafarpour; Alireza Abdollahi; Azam Ghaziasadi; Azar Hadadi; Hossein Najmabadi.; Khadijeh Jalalvand; Kimia Kahrizi; Marzieh Mohseni; Reza Najafipour; Saber Soltani; Seyed Mohammad Jazayeri; Seyedeh elham Mortazavi; Zohreh Fattahi |
| EPI_ISL_448062 | Department of Pathology, University of Cambridge | COVID-19 Genomics UK (COG-UK) Consortium | Aminu S. Jahun; Anna Yakovleva; Charlotte J. Houldcroft; Fahad A Khokhar; Grant Hall; Ian Goodfellow; Laura G Caller; Luke W Meredith; M. Estée Török; Martin D. Curran; Myra Hosmillo; Sarah L. Caddy; Theresa Feltwell; William L. Hamilton |
| EPI_ISL_439557, EPI_ISL_439647, EPI_ISL_459182, EPI_ISL_459193, EPI_ISL_459217, EPI_ISL_459427, EPI_ISL_459489, EPI_ISL_459495 | | | |
| see above | Department of Pathology, University of Cambridge | Wellcome Sanger Institute for the COVID-19 Genomics UK (COG-UK) consortium | Alex Alderton; Aminu S. Jahun; Anna Yakovleva; Charlotte J. Houldcroft; Cordelia Langford; David K. Jackson; Dominic Kwiatkowski; Ewan Harrison; Fahad A Khokhar; Grant Hall; Ian Goodfellow; Ian Johnston; John Sillitoe on behalf of the Wellcome Sanger Institute COVID-19 Surveillance Team (http://www.sanger.ac.uk/covid-team); Laura G Caller; Luke W Meredith; M. Estée Török; Martin D. Curran; Myra Hosmillo; Roberto Amato; Sarah L. Caddy; Sonia Goncalves; Theresa Feltwell; William L. Hamilton; and Alex Alderton |
| EPI_ISL_1657104, EPI_ISL_1657117, EPI_ISL_1969986, EPI_ISL_1969990, EPI_ISL_2099878, EPI_ISL_2099884, EPI_ISL_2190107 | | | |
| see above | Department of Public Health Bucharest | National Institute of Infectious Diseases-Prof. Dr. Matei Bals Molecular Diagnostics Laboratory | Andreea Tudor; Corina Casangiu; Dan Otelea; Leontina Banica; Marius Surleac; Ovidiu Vlaicu; Simona Paraschiv |
| EPI_ISL_2687986 | Department of Public Health Buzau | National Institute of Infectious Diseases-Prof. Dr. Matei Bals Molecular Diagnostics Laboratory | Corina Casangiu; Dan Otelea; Leontina Banica; Marius Surleac; Petre Milu; Robert Hohan; Simona Paraschiv |
| EPI_ISL_2687992, EPI_ISL_3045846 | Department of Public Health Constanta | National Institute of Infectious Diseases-Prof. Dr. Matei Bals Molecular Diagnostics Laboratory | Corina Casangiu; Dan Otelea; Leontina Banica; Marius Surleac; Petre Milu; Robert Hohan; Simona Paraschiv |
| EPI_ISL_3045850, EPI_ISL_3045852, EPI_ISL_3045853, EPI_ISL_3045867, EPI_ISL_3045876 | Department of Public Health Iasi | National Institute of Infectious Diseases-Prof. Dr. Matei Bals Molecular Diagnostics Laboratory | Corina Casangiu; Dan Otelea; Leontina Banica; Marius Surleac; Petre Milu; Robert Hohan; Simona Paraschiv |
| EPI_ISL_1677895, EPI_ISL_1795690, EPI_ISL_1795878 | Department of Public Health Microbiology Ljubljana, National Laboratory for Health, Environment and Food | Department for Public Health Microbiology Ljubljana, National Laboratory for Health, Environment and Food | José Gonçalves; Katarina Prosenc; Martin Bosilj; Metka Paragi; Natasa Berginc; Tom Koritnik |
| EPI_ISL_582513, EPI_ISL_979973 | Department of Respiratory and other Viral Infections of L.V.Gromashevsky Institute of Epidemiology & Infectious Diseases NAMS of Ukraine | Department of Respiratory and other Viral Infections of L.V.Gromashevsky Institute of Epidemiology & Infectious Diseases NAMS of Ukraine, JSC "Farmak" | Alla Mironenko; Andriy Goy; Ihor Kravchuk; Larysa Radchenko; Ludmyla Bolotova; Nataliia Teteriuk |
| EPI_ISL_1385784, EPI_ISL_1385786, EPI_ISL_1385791, EPI_ISL_1385794, EPI_ISL_1398039, EPI_ISL_1406394, EPI_ISL_1710506, EPI_ISL_1711673, EPI_ISL_1732172, EPI_ISL_1969995, EPI_ISL_2313112, EPI_ISL_2434781, EPI_ISL_2434982, EPI_ISL_2438547, EPI_ISL_2757735, EPI_ISL_2757736, EPI_ISL_2757738, EPI_ISL_2757739, EPI_ISL_2757740, EPI_ISL_2757743, EPI_ISL_2757756, EPI_ISL_2757758, EPI_ISL_2757759, EPI_ISL_2757761, EPI_ISL_2757762, EPI_ISL_2894974, EPI_ISL_2894982 | | | |
| see above | Department of Virology | Department of Virology | Aamer Ikram; Abdul Ahad; Adnan Haider; Ammar Amjad; Austin Leach; Joel Montgomery; John Klena; Ketan Patel; Massab Umair; Melissa Mobley; Muhammad Ammar; Muhammad Salman; Nazish Badar; Qasim Ali; Sana Tamim; Shannon Whitmer; Syed Adnan Haider; Zaira Rehman |
| EPI_ISL_732649, EPI_ISL_738136, EPI_ISL_759798, EPI_ISL_759830, EPI_ISL_1218648, EPI_ISL_1595768, EPI_ISL_2604473, EPI_ISL_2608455, EPI_ISL_2608457, EPI_ISL_2608770, EPI_ISL_2608977, EPI_ISL_2609443, EPI_ISL_3014476, EPI_ISL_3014729, EPI_ISL_3014888, EPI_ISL_3015049 | | | |
| see above | Department of Virology and Immunology, University of | Department of Virology, Faculty of Medicine, University of | Essi Korhonen; Fathiah Zakham; Hanna Jarva; Hanna Liimatainen; Hannimari Kallio-Kokko; Harri Kangas; Hussein Alburkat; Jenni Virtanen; Maija Lappalainen; Maija Suvanto; Mert Erdin; Olli Vapalahti; Pekka Ellonen; Phuoc Truong; Ravi Kant; Sari Hannula; Satu |

| | | | |
|---|---|---|---|
| | Helsinki and Helsinki University Hospital, Huslab Finland | Helsinki, Helsinki, Finland | Kurkela; Teemu Smura; fathiah Zakham |
| EPI_ISL_2896259 | Department of Virology, Molecular Biology and Genome Research | Department of Virology, Molecular Biology and Genome Research | Kotaria, N.; Kotorashvili, A.; Murtskhvaladze, M. |
| EPI_ISL_855567, EPI_ISL_855570 | Department of Virology, Principal Military Hospital of Instruction of Tunis | Bundeswehr Institute of Microbiology | Habiba Naija; Kilian Stoecker; Malena Bestehorn-Willmann; Markus H. Antwerpen; Mathias C. Walter; Roman Wölfel & Mohamed Ben Moussa; Simone Eckstein; Susann Handrick |
| EPI_ISL_757286 | Department of Virology, Public Health Laboratories Division | Department of Virology, Public Health Laboratories Division | Aamer Ikram; Massab Umair; Muhammad Salman |
| EPI_ISL_2254131 | Department of Virology, School of Medicine, Shiraz University of Medical Sciences, Shiraz, Iran | Genetics Research Center, University of Social Welfare and Rehabilitation Sciences | Hossein Najmabadi; Kimia Kahrizi; Marzieh Mohseni; Moattari A; Moghadami M; Zohreh Fattahi |
| EPI_ISL_926067, EPI_ISL_927136, EPI_ISL_928108, EPI_ISL_928152, EPI_ISL_930199, EPI_ISL_1022501, EPI_ISL_1022644, EPI_ISL_1022682, EPI_ISL_1022684, EPI_ISL_1065948, EPI_ISL_1125881, EPI_ISL_1126064, EPI_ISL_1126279, EPI_ISL_1858552, EPI_ISL_1859527, EPI_ISL_1859734, EPI_ISL_1860060, EPI_ISL_1860341, EPI_ISL_1862355, EPI_ISL_1862402, EPI_ISL_1862888, EPI_ISL_1863717, EPI_ISL_1864758, EPI_ISL_1865226, EPI_ISL_1865320, EPI_ISL_1866574, EPI_ISL_1867960, EPI_ISL_1868550, EPI_ISL_1868837, EPI_ISL_1869258, EPI_ISL_1871039, EPI_ISL_1871224, EPI_ISL_1871274, EPI_ISL_1871949, EPI_ISL_1872826, EPI_ISL_1872870, EPI_ISL_1873858, EPI_ISL_1874487, EPI_ISL_1874779, EPI_ISL_1875598, EPI_ISL_1875866, EPI_ISL_1876066, EPI_ISL_1876442, EPI_ISL_1876721, EPI_ISL_1877285, EPI_ISL_1877995, EPI_ISL_1878462, EPI_ISL_1879001, EPI_ISL_1881008, EPI_ISL_1881989, EPI_ISL_1882010, EPI_ISL_1882472, EPI_ISL_1882982, EPI_ISL_1883563, EPI_ISL_1883815, EPI_ISL_1884113, EPI_ISL_1886538, EPI_ISL_1887618, EPI_ISL_1888054, EPI_ISL_1888124, EPI_ISL_1890115, EPI_ISL_1891575, EPI_ISL_1891764, EPI_ISL_1893809, EPI_ISL_1894309, EPI_ISL_1895070, EPI_ISL_1896329, EPI_ISL_2023813, EPI_ISL_2025423, EPI_ISL_2028832, EPI_ISL_2175777, EPI_ISL_2176487, EPI_ISL_2300915 | | | |
| see above | Department of Virus and Microbiological Special Diagnostics, Statens Serum Institut, Copenhagen, Denmark | Aalborg University | Danish Covid-19 Genome Consortium |
| EPI_ISL_669126, EPI_ISL_669629, EPI_ISL_670863, EPI_ISL_757623, EPI_ISL_792954 | Department of Virus and Microbiological Special Diagnostics, Statens Serum Institut, Copenhagen, Denmark | Albertsen Lab, Department of Chemistry and Bioscience, Aalborg University, Denmark | Danish Covid-19 Genome Consortium |
| EPI_ISL_437631 | Department of Virus and Microbiological Special Diagnostics, Statens Serum Institut, Copenhagen, Denmark, Artillerivej 5, 2300 Copenahgen S | Albertsen lab, Department of Chemistry and Bioscience, Aalborg University, Denmark | Rasmus Kirkegaard |
| EPI_ISL_614409, EPI_ISL_616443, EPI_ISL_616724, EPI_ISL_617431, EPI_ISL_618152, EPI_ISL_618309, EPI_ISL_618562, EPI_ISL_619869, EPI_ISL_620017, EPI_ISL_621663 | | | |
| see above | Department of Virus and Microbiological Special Diagnostics, Statens Serum Institut, Denmark | Albertsen lab, Department of Chemistry and Bioscience, Aalborg University, Denmark | Danish Covid-19 Genome Consortia |
| EPI_ISL_1919409, EPI_ISL_2400054, EPI_ISL_2491413 | Dept. of Medical Microbiology, Stavanger University Hospital, Helse Stavanger HF | Norwegian Institute of Public Health, Department of Virology | Atiya R Ali; Debech Nadia; Engebretsen Serina Beate; Garcia Llorente Ignacio; Hilde Elshaug; Hilde Vollan; Jon Bråte; Kamilla Heddeland Instefjord; Karoline Bragstad; Kathrine Stene-Johansen; Line Victoria Moen; Marie Paulsen Madsen; Olav Hungnes; Pedersen Benedikte Nevjen; Rasmus Riis Kopperud |
| EPI_ISL_1118120, EPI_ISL_1118121 | Dept. of Microbiology and Infection Control, Akershus University Hospital HF | Dept. of Microbiology and Infection Control, Akershus University Hospital HF | Alexander Hesselberg Løvestad; Hege Vangstein Aamot |
| EPI_ISL_410532 | Dept. of Pathology, National Institute of Infectious Diseases | Pathogen Genomics Center, National Institute of Infectious Diseases | Harutaka Katano; Hideki Hasegawa; Kazuya Shirato; Makoto Kuroda; Makoto Takeda; Motoi Suzuki; Naganori Nao; Shutoku Matsuyama; Tadaki Suzuki; Takaji Wakita; Tsuyoshi Sekizuka |
| EPI_ISL_774872 | Designated Reference Institute for Chemical Measurements (DRICM) | DNA SOLUTION LTD. | Abdul Khaleque; Abu Sufian; Hasan Ul Haider; Jannatun Naima; Kazi Nadim Hasan; MSM Chowdhury; Mala Khan; Mamudul Hasan Razu; Md. Imran Khan; Mizanur Rahman; Mohammad Fazle Alam Rabbi |
| EPI_ISL_2171041, EPI_ISL_2171267 | Dhulikhel Hospital, Kathmandu University Hospital | Molecular and Genomics Research Lab, Dhulikhel Hospital, Kathmandu University Hospital | Dipesh Tamrakar; Manu Vanaerschot; Meghnath Dhimal; Navin Adhikari; Nishan Katuwal; Pradip Gyanwali; Rajeev Shrestha; Saroj Bhattarai; Surendra Kumar Madhup |
| EPI_ISL_2101016, EPI_ISL_2101017 | Diagnostic Laboratory-Microbiology | Biomedical Laboratory-2 | ; A. Dharshan De Silva; Dilini Nakawita; Dilini Nakkawita; Dumitha Govindapala; Harshi Abeygoonawardena; Himali S. Jayasinghearachchi; Pawani Gamage; Priyamali Jayasekera; Thamarasi Senaratne; Upeksha Kulasekera |
| EPI_ISL_1447336, EPI_ISL_1533051, EPI_ISL_1533073 | Diagnostic and Research Center of Infectious Diseases, Medical Faculty, Andalas University | Diagnostic and Research Center of Infectious Diseases, Medical Faculty, Andalas University | Andani Eka Putra; Ayu Novita Trisnawati; Dede Rahman Agustian; Desmawati; Dessy Arisanty; Fauzul Azhim; Gestina Aliska; Ikhwan R. Sudji; Juane Plantika Menra; Linosefa; Mutia Lailani; Nia Ayuni Putri; Nita Afriani; SM Rezvi; Sekar Asri Tresnaningtyas; Siskalil Fahma; Syafrizayanti; Syandrez Prima Putra; Yolani Syaputri |
| EPI_ISL_2636693, EPI_ISL_2762335, EPI_ISL_2762339, EPI_ISL_2957236, EPI_ISL_2957275 | Diagnosticum - Labor Neukirchen | Robert Koch Institute | |
| EPI_ISL_2657185 | Diagnostyka | 1. Tricity SARS-CoV-2 sequencing consortium: University of Gdansk, Medical University of Gdansk, Vaxican Ltd., Invicta Ltd. 2. National Institute of Public Health - National Institute of Hygiene, Warsaw, Poland | Celina Cybulska; Karolina Gackowska; Katarzyna Groth; Katarzyna Zacharczuk; Krystyna Bienkowska Szewczyk; Lukasz Rabalski; Maciej Grzybek; Maciej Kosinski; Magdalena Nowakowska; Marcin Lubocki; Małgorzata Sadkowska-Todys; Tomasz Wołkowicz |
| EPI_ISL_2319055, EPI_ISL_2937950, EPI_ISL_2937967, EPI_ISL_3133633 | Diagnostyka. Laboratoria Medyczne. | 1. ViroGenetics - BSL3 Laboratory of Virology, Małopolska Centre of Biotechnology, Jagiellonian University; 2. genXone SA, Research & Development Laboratory | Aleksandra Gidlewicz; Anna Brylak; Brylak A; Drweska-Matelska N; Gidlewicz A; Grabowski J; Gromowski, T.; Grzegorz Nowicki; Jakub Grabowski; Januszczak S; Karol Szeszko; Kaszuba M; Kowalski, M.; Krych L; Labaj; Maciej Sykulski; Mazur-Panasiuk, N.; Michał Kaszuba; Natalia Drweska-Matelska; Nowicki G; P.P.; Pyrc, K.; Sykulski M; Sylwia Januszczak; Szeszko K; Szulc, P.; Wydmanski W.; Wydmanski, W.; Łukasz Krych |
| EPI_ISL_1752833 | Diagnostyka. Laboratoria Medyczne. | 1. ViroGenetics - BSL3 Laboratory of Virology, Małopolska Centre of Biotechnology, Jagiellonian University; 2. genXone SA, Research & Development Laboratory | Brylak A; Drweska-Matelska N; Gidlewicz A; Grabowski J; Gromowski, T.; Januszczak S; Kaszuba M; Kowalski, M.; Krych L; Labaj; Mazur-Panasiuk, N.; Nowicki G; P.P.; Pyrc, K.; Sykulski M; Szeszko K; Szulc, P.; Wydmanski W. |
| EPI_ISL_2597974 | Dianalabs SA | Genesupport | Geraldine Jost; Katia Jaton; Nadia Liassine; Tanguy ARAUD |
| EPI_ISL_1855939 | Dinas Kesehatan Kota Bogor | Indonesian Research Center for Veterinary Science / BBalitvet | Diana Nurjanah; Fadjry Djufry; Harimurti Nuradji; Ilham Chaidir; Ni Luh Putu Indi Dharmayanti; Risa Indriani; Sri Nowo Retno |
| EPI_ISL_747337, EPI_ISL_747419, EPI_ISL_760064, EPI_ISL_1063655, EPI_ISL_1063656, EPI_ISL_2161069, EPI_ISL_2161113, EPI_ISL_2161137, EPI_ISL_2332342, EPI_ISL_2361092, EPI_ISL_2361095, EPI_ISL_2361097, EPI_ISL_2361167, EPI_ISL_2464346, EPI_ISL_2464347, EPI_ISL_2710485, EPI_ISL_2710499, EPI_ISL_2754407, EPI_ISL_2754473, EPI_ISL_2754619, EPI_ISL_2967308, EPI_ISL_2967332, EPI_ISL_2967375, EPI_ISL_2967572, EPI_ISL_3026379, EPI_ISL_3026450, EPI_ISL_3026451, EPI_ISL_3026459, EPI_ISL_3026473, EPI_ISL_3026476, EPI_ISL_3026508, EPI_ISL_3026560, EPI_ISL_3026673, EPI_ISL_3026704, EPI_ISL_3026742, EPI_ISL_3026834 | | | |
| see above | Division of Emerging Infectious Diseases, Bureau of Infectious Diseases Diagnosis Control, Korea Disease Control and Prevention Agency | Division of Emerging Infectious Diseases, Bureau of Infectious Diseases Diagnosis Control, Korea Disease Control and Prevention Agency | Ae Kyung Park; Chae Young Lee; Chaeyoung Lee; Eun-Jin Kim; Heui Man Kim; Il-Hwan Kim; Jeong-Ah Kim; Jeong-Min Kim; Jin Sun No; Namjoo Lee; Sang Hee Woo |
| EPI_ISL_2695860, EPI_ISL_2695862, EPI_ISL_2695874, EPI_ISL_2876313, EPI_ISL_2876320, EPI_ISL_2876336, EPI_ISL_2876382, EPI_ISL_3032493, EPI_ISL_3032496, EPI_ISL_3032506, EPI_ISL_3032513, EPI_ISL_3032515, EPI_ISL_3032516 | | | |
| see above | Division of Medical Virology, National Health Laboratory Service (NHLS), Tygerberg Hospital / Stellenbosch University | Division of Medical Virology, National Health Laboratory Service (NHLS), Tygerberg Hospital / Stellenbosch University | Bronwyn Kleinhans; Gert van Zyl; Kamela Mahlakwane; San Emmanual James; Susan Engelbrecht; Tania Stander; Tongai Maponga; Tulio de Oliveira; Wolfgang Preiser |
| EPI_ISL_1591415, EPI_ISL_1591427 | Division of Medical Virology, National Health Laboratory Service (NHLS), Tygerberg Hospital / Stellenbosch University | Division of Medical Virology, Stellenbosch University and NHLS Tygerberg Hospital | Bronwyn Kleinhans; Gert van Zyl; Susan Engelbrecht; Wolfgang Preiser |
| EPI_ISL_412869, EPI_ISL_426173, EPI_ISL_497962, EPI_ISL_506987, EPI_ISL_510627 | Division of Viral Diseases, Center for Laboratory Control of Infectious Diseases, Korea Centers for Diseases Control and Prevention | Division of Viral Diseases, Center for Laboratory Control of Infectious Diseases, Korea Centers for Diseases Control and Prevention | Daesang Lee; Dong Hyun Song; Heui Man Kim; Hye-Jun Jo; Jeong-Min Kim; Jun-Sub Kim; Junhyeong Jang; Mi-Seon Kim; Myung Guk Han; Namjoo Lee; Sang Hee Woo; Sehee Park; Seong Tae Jeong; Yoon-Seok Chung |
| EPI_ISL_2521795 | Dr S raju, Director of public heath and preventive medicine | inStem NCBS - INSACOG | Uma Ramakrishnan Dasaradhi Palakodeti Aswin SaiNarain |
| EPI_ISL_1055411 | Dr. Leonard A. Miller Centre for Health Services | National Microbiology Laboratory (NML) | Adel Malek; Anna Majer; Anneliese Landgraff; CanCOGeN's metadata curation team; Darian Hole; Elsie Grudeski; Gary Van Domselaar; George Zahariadis; Grace Seo; Jennifer Tanner; Kerri Smith; Kirsten Biggar; Laura Gilbert; Madison Chapel; Morag Graham; Natalie Knox; Nathalie Bastien; Philip Mabon; Public Health Agency of Canada CanCOGeN team; Rhiannon Huzarewich; Robert Needle; Russell Mandes; Shari Tyson; Timothy Booth; Yan Li; Yang Yu |
| EPI_ISL_461385, EPI_ISL_632354, EPI_ISL_801407, EPI_ISL_2483471, EPI_ISL_2860624 | Dutch COVID-19 response team | Erasmus Medical Center | Anne van der Linden; Annemiek van der Eijk; Aura Timen; Bas Oude Munnink; Claudia Schapendonk; Corien Swaan; Corine GeurtsvanKessel; David Nieuwenhuijse; Emmanuelle Munger; Irina Chestakova; Jeroen van Kampen; Jolanda Voermans; Madelief Mollers; Manon Haverkate; Marion Koopmans; Marjan Boter; Mark Pronk; Mart Stein; Pascal Lexmond; Reina Sikkema; Richard Molenkamp; Sandra Kengne Kamga Mobou; Stefan van Nieuwkoop; Theo Bestebroer; on behalf of the Dutch national COVID-19 respo; on behalf of the Dutch national COVID-19 response team. |
| EPI_ISL_547447, EPI_ISL_547449, EPI_ISL_547451, EPI_ISL_636492, EPI_ISL_636519, EPI_ISL_824273, EPI_ISL_1014558, EPI_ISL_1014573, EPI_ISL_1014646, EPI_ISL_1035245, EPI_ISL_1232248, EPI_ISL_1232305, EPI_ISL_1232602, EPI_ISL_1288137, EPI_ISL_1371227, EPI_ISL_1457590, EPI_ISL_1521321, EPI_ISL_1521326, EPI_ISL_1522167, EPI_ISL_1522202, EPI_ISL_1522223, EPI_ISL_1596268, EPI_ISL_1597320, EPI_ISL_1597411, EPI_ISL_1703535, EPI_ISL_1704912, EPI_ISL_1704914, EPI_ISL_1704921, EPI_ISL_1704945, EPI_ISL_1704948, EPI_ISL_1704954, EPI_ISL_1704959, EPI_ISL_1704990, EPI_ISL_1705155, EPI_ISL_1705178, EPI_ISL_1705190, EPI_ISL_1705242, EPI_ISL_1705251, EPI_ISL_1705268, EPI_ISL_1705285, EPI_ISL_1705293, EPI_ISL_1705301, EPI_ISL_1705568, EPI_ISL_1705864, EPI_ISL_1705904, EPI_ISL_1705929, EPI_ISL_1792360, EPI_ISL_1792363, EPI_ISL_1792862, EPI_ISL_1792863, EPI_ISL_1792864, EPI_ISL_1792865, EPI_ISL_1792868, EPI_ISL_1961869, EPI_ISL_1961897, EPI_ISL_1961912, EPI_ISL_1961920, EPI_ISL_1962066, EPI_ISL_1962160, EPI_ISL_1962166, EPI_ISL_1962181, EPI_ISL_1962712, EPI_ISL_1962940, EPI_ISL_1962944, EPI_ISL_1962971, EPI_ISL_2093518, EPI_ISL_2093534, EPI_ISL_2093545, EPI_ISL_2093608, EPI_ISL_2093659, EPI_ISL_2093759, EPI_ISL_2093798, EPI_ISL_2093885, EPI_ISL_2094043, EPI_ISL_2094340, EPI_ISL_2094396, EPI_ISL_2218611, EPI_ISL_2218731, EPI_ISL_2219699, EPI_ISL_2220430, EPI_ISL_2220456, EPI_ISL_2220523, EPI_ISL_2220549, EPI_ISL_2302309, EPI_ISL_2302360, EPI_ISL_2302965, EPI_ISL_2302971, EPI_ISL_2303552, EPI_ISL_2303931, EPI_ISL_2303978, EPI_ISL_2405457, EPI_ISL_2405458, EPI_ISL_2405505, EPI_ISL_2405516, EPI_ISL_2405518, EPI_ISL_2405526, EPI_ISL_2405538, EPI_ISL_2405550, EPI_ISL_2405561, EPI_ISL_2405570, EPI_ISL_2405573, EPI_ISL_2405584, EPI_ISL_2405585, EPI_ISL_2405594, EPI_ISL_2405596, EPI_ISL_2405597, EPI_ISL_2405608, EPI_ISL_2405609, EPI_ISL_2405620, EPI_ISL_2406008, EPI_ISL_2406319, EPI_ISL_2406352, EPI_ISL_2474249, EPI_ISL_2475174, EPI_ISL_2475191, EPI_ISL_2475363, EPI_ISL_2475518, EPI_ISL_2475682, EPI_ISL_2475716, EPI_ISL_2475722, EPI_ISL_2475732, EPI_ISL_2476242, EPI_ISL_2476276, EPI_ISL_2476311, EPI_ISL_2476312, EPI_ISL_2476313, EPI_ISL_2609665, EPI_ISL_2610116, EPI_ISL_2610186, EPI_ISL_2610234, EPI_ISL_2610471, EPI_ISL_2610546, EPI_ISL_2610658, EPI_ISL_2610674, EPI_ISL_2610690, EPI_ISL_2610721, EPI_ISL_2610722, EPI_ISL_2610775, EPI_ISL_2672255, EPI_ISL_2672409, EPI_ISL_2672546, EPI_ISL_2673234, EPI_ISL_2673239, EPI_ISL_2673240, EPI_ISL_2673274, EPI_ISL_2673295, EPI_ISL_2787916, EPI_ISL_2787926, EPI_ISL_2788186, EPI_ISL_2788368, EPI_ISL_2862759, EPI_ISL_2862935, EPI_ISL_2862937, EPI_ISL_2862941, EPI_ISL_2981152, EPI_ISL_2981385, EPI_ISL_2981482, EPI_ISL_2981711, EPI_ISL_2981868, EPI_ISL_2981896, EPI_ISL_2981901 | | | |
| see above | Dutch COVID-19 response team | National Institute for Public Health and the Environment (RIVM) | Adam Meijer; AnneMarie van den Brandt; Annelies Kroneman; Bas van der Veer; Chantal Reusken; Dennis Schmitz; Dirk Eggink; Euníce Then; Florian Zwagemaker; Harry Vennema; James Groot; Jeroen Cremer; Jolienke Hardeman; Karim Hajji; Kim Freriks; Linda van de Nes; Lisa Wijsman; Lynn Aarts; Melissa van Tuil; Robert Kohl; Ryanne Jaarsma; Sanne Bos; Sharon van den Brink; Sjoerd Kuiling; on behalf of the national COVID-19 response team |
| EPI_ISL_2140728 | Dysselsdorp Clinic wc DDC | NHLS/UCT | Arash Iranzadeh; Bruna Galvao; Carolyn Williamson; Deelan Doolabh; Diana Hardie; Innocent Mudau; Kruger Marais; Lynn Tyers; Marvin Hsiao; Stephen Korsman |
| EPI_ISL_2246957 | Dział Diagnostyki Laboratoryjnej Wojewódzki Szpital | 1. Academic Center for Pathomorphological and Genetic- | Anetta Sulewska; Jacek Nikliński; Janusz Dzięcioł; Joanna Kiśluk; Katarzyna Zacharczuk; Konrad Raczkowski; Magdalena Nowakowska; Małgorzata Sadkowska-Todys; Piotr Karabowicz; Piotr Majewski; Przemysław Biecek. Joanna Reszeć; Radosław Charkiewicz; |

| | | | |
|---|---|---|---|
| | Specjalistyczny im.J.Gromkowskiego | Molecular Diagnostics ltd, Bialystok, Poland 2. National Institute of Public Health - National Institute of Hygiene, Warsaw, Poland | Tomasz Wołkowicz |
| EPI_ISL_421656 | E. Gulbja Laboratorija | Latvian Biomedical Research and Study Centre | Dmitrijs Perminovs; Ivars Silamiķelis; Jānis Kloviņš; Kaspars Megnis; Mikus Gavars; Monta Ustinova; Uga Dumpis; Vita Rovīte; Ņikita Zrelovs |
| EPI_ISL_1312417, EPI_ISL_1312435, EPI_ISL_1312683, EPI_ISL_1312689, EPI_ISL_1321930 | E. Gulbja laboratorija | Latvian Biomedical Research and Study Centre | Davids Fridmanis; Dmitrijs Perminovs; Elina Dimina; Guntars Zarins; Ivars Silamikelis; Janis Klovins; Janis Pjalkovskis; Jurijs Perevoscikovs; Kaspars Megnis; Laila Silamikele; Lauma Freimane; Laura Ansone; Liga Birzniece; Mikus Gavars; Monta Ustinova; Nikita Zrelovs; Uga Dumpis; Una Krumina; Vita Rovite |
| EPI_ISL_1582980 | E.S.E. HOSPITAL SAN JOSE DE MAICAO | Instituto Nacional de Salud- Dirección de Investigación en Salud Pública | Carlos Franco-Muñoz; Carmen Osorio; Diana Malo; Diego A. Álvarez-Díaz; Diego Andrés Prada; Gerardo Santamaría; Hector Alejandro Ruiz-Moreno; Jhonnatan Reales-González; Juan Camilo Martinez; Julian Naizaque; Katherine Laiton-Donato; Lisseth Pardo; Magdalena Wiesner; Marcela Mercado-Reyes; Maria T. Herrera-Sepúlveda; Marta Lopez Blanco; Martha Lucia Ospina Martinez; Paola Rojas; Sergio Gomez; Sheryll Corchuelo; Ángela Alarcon Cruz |
| EPI_ISL_1915793, EPI_ISL_1972358 | EHA Clinics | National Reference Laboratory, Nigeria Centre for Disease Control | Anthony Ahumibe; Catherine Okoi; Chimaobi Chukwu; Dr Chikwe Ihekweazu; Dr Ndodo Nnaemeka; Dr Omoare Adesuyi; Grace Esebanmen; Naidoo Dhamari; Nwando Mba; Olusola Anuoluwapo Akanbi |
| EPI_ISL_2987317, EPI_ISL_2987318, EPI_ISL_2987335 | ERMELO LABORATORY | National Institute for Communicable Diseases of the National Health Laboratory Service | Amoako DG; Bhiman JN; Everatt J; Ismail A; Mahlangu B; Mnguni A; Mohale T; Ntuli N; Scheepers C |
| EPI_ISL_2370457, EPI_ISL_2370853 | ETHNIKO KENTRO AIMODOSIAS E.KE.A. | Greek Genome Center, Biomedical Research Foundation of the Academy of Athens (BRFAA) | Dimitrios Thanos; Efthimia Petinaki; Emmanouil Athanasiadis; Giannis Vatsellas; Katerina Zoi; Kostas Stamoulis; Theodoros Loupis |
| EPI_ISL_1811773, EPI_ISL_1811814 | EXCITE Lab | Andersen lab at Scripps Research | Alexandre Bolze; Alice Summerfield; Celena Andrade; Charlotte Rivera-Garcia; David Becker; Efren Sandoval; Elizabeth Cirulli; Francisco Tanudjaja; Geraint Levan; James Lu + SEARCH; Jason Nguyen; Jimmy Ramirez; Kelly Schiabor Barrett; Magnus Isaksson; Marc Laurent; Nicole L Washington; Ryan Cho; Sherry Wang; Simon White; Tyler Cassens; William Lee |
| EPI_ISL_593513 | Eastern Ontario Regional Laboratory Association | McMaster University | Ahmed Draia; Andrew G. McArthur; Emily Panousis; Hooman Derakhshani; Jalees Nasir; Leanne Mortimer; Robert Slinger |
| EPI_ISL_2477989, EPI_ISL_2478875, EPI_ISL_2479322, EPI_ISL_2939133, EPI_ISL_2940521, EPI_ISL_2940651, EPI_ISL_2940877, EPI_ISL_2940897 | | | |
| see above | Edmonton Provincial Lab | Public Health Agency of Canada (PHAC) National Microbiology Laboratory | Buss; Croxen M; Deo A; Dieu P; E; Ferrato C; Gill K; Khan F; Koleva P; Li V; Lloyd C; Lynch T; Ma R; Murphy S; Pabbaraju K; Shokoples S; Thayer J; Tipples G; Whitehouse M; Wong A; Yu C; Zelyas N |
| EPI_ISL_468044, EPI_ISL_468045, EPI_ISL_468046, EPI_ISL_468047, EPI_ISL_468048, EPI_ISL_468049, EPI_ISL_468050, EPI_ISL_468051, EPI_ISL_468052, EPI_ISL_468053, EPI_ISL_468054, EPI_ISL_468055, EPI_ISL_468056, EPI_ISL_468057, EPI_ISL_468058, EPI_ISL_468059, EPI_ISL_468060, EPI_ISL_468061, EPI_ISL_468062, EPI_ISL_475722, EPI_ISL_475723, EPI_ISL_475724, EPI_ISL_477161, EPI_ISL_478672, EPI_ISL_479686, EPI_ISL_479687, EPI_ISL_479688, EPI_ISL_479689, EPI_ISL_479690, EPI_ISL_479691, EPI_ISL_479692, EPI_ISL_479693, EPI_ISL_479694, EPI_ISL_479695, EPI_ISL_479696, EPI_ISL_479697, EPI_ISL_479698, EPI_ISL_479699, EPI_ISL_479700, EPI_ISL_479701, EPI_ISL_479702, EPI_ISL_479703, EPI_ISL_479704, EPI_ISL_479705, EPI_ISL_479706, EPI_ISL_479707, EPI_ISL_479708, EPI_ISL_479709, EPI_ISL_479710, EPI_ISL_479711, EPI_ISL_479712, EPI_ISL_479713, EPI_ISL_479714, EPI_ISL_479715, EPI_ISL_479716, EPI_ISL_479717, EPI_ISL_479718, EPI_ISL_479719, EPI_ISL_479720, EPI_ISL_479721, EPI_ISL_479722, EPI_ISL_479723, EPI_ISL_479724, EPI_ISL_479725, EPI_ISL_479726, EPI_ISL_479727, EPI_ISL_479728, EPI_ISL_479729, EPI_ISL_479730, EPI_ISL_479731, EPI_ISL_479732, EPI_ISL_479733, EPI_ISL_479734, EPI_ISL_479735, EPI_ISL_524426, EPI_ISL_524427, EPI_ISL_529141, EPI_ISL_529142, EPI_ISL_529143, EPI_ISL_529144, EPI_ISL_529145 | | | |
| see above | Egyptian National Cancer Institute (ENCI) | Egyptian National Cancer Institute (ENCI) | A.A.; A.N.; Abdel Rahman N; Abdel Rahman N.; Abdelhamid, W.; Abouelhoda; Abouelhoda, M.; Ahmed; Ali; Ali, M.; Amer; Bahnassy; Elkhateeb; Elsissy; Ezzelarab; Gad, A.; H.K.; Hafez; Hamdy; Hassan; Hassan, W.; K.E.; Khattab; M.A.; M.H.; M.M.; M.S.; Mahmoud; Mohamed; O.S.; Raouf, A.; S.M.; Samir, M.; Soliman; W.A.; Zekri |
| EPI_ISL_469275 | Egyptian National Cancer Institute (ENCI) | Human Genome Center | A.A.; Abdel Rahman N; Abdelhamid, W.; Abouelhoda; Ahmed; Ali, M.; Amer; Bahnassy; Elkhateeb; Elsissy; Ezzelarab; Gad, A.; H.K.; Hafez; Hamdy; Hassan, W.; K.E.; M.H.; M.M.; M.S.; Mohamed; O.S.; Raouf, A.; S.M.; Samir, M.; Soliman; Zekri |
| EPI_ISL_2029260 | Ekstralab Tuzla | Alea Genetic Centre | Adis Kandic; Dino Pecar; Enis Kandic; Lana Salihefendic; Nusret Butkovic; Rijad Konjhodzic |
| EPI_ISL_1969960, EPI_ISL_1969963 | Emergency County Hospital Targu Mures | National Institute of Infectious Diseases-Prof. Dr. Matei Bals Molecular Diagnostics Laboratory | Andreea Tudor; Corina Casangiu; Dan Otelea; Leontina Banica; Marius Surleac; Ovidiu Vlaicu; Simona Paraschiv |
| EPI_ISL_754751 | Emory Molecular Diagnostics Laboratory, Emory Healthcare | Piantadosi Lab, Emory Department of Pathology | Ahmed Babiker; Anne Piantadosi |
| EPI_ISL_2321508 | Emory University | Centers for Disease Control and Prevention Division of Viral Diseases, Pathogen Discovery | Alison Laufer Halpin; Ben L. Rambo-Martin; Clinton R. Paden; Dakota Howard; Darlene Wagner; Dave Wentworth; Dhwani Batra; Jasmine Padilla; Justin Lee; Katie Dillon; Krista Queen; Kristen Knipe; Kristine Lacek; Mark Burroughs; Matthew Schmerer; Mili Sheth; Peter Cook; Sam Shepard; Sarah Nobles; Shoshona Le; Suxiang Tong; Vivien Dugan; Yvette Unoarumhi |
| EPI_ISL_2438624 | Enbekshikazakh multidisciplinary interdistrict hospital | Reference laboratory for the control of viral infections | Aidar Usserbayev; Aknur Mutaliyeva; Andrey Komissarov; Artem Fadeev; Azamat Kenessov; Bekzhan Maikotov; Gaukhar Nussupbayeva; Madina Tleubergenova; Maria Pisareva; Nazym Tleumbetova |
| EPI_ISL_2086267 | Enthabeni Lancet Laboratories | KRISP, KZN Research Innovation and Sequencing Platform | Emmanuel SJ; Giandhari J; Khan S; Lessells R; Mdlalose K; Naidoo Y; Pillay S; Ramphal U; Tegally H; Wilkinson E; York D; de Oliveira T |
| EPI_ISL_2112529 | Epidemiology of Microbial Diseases, Yale School of Public Health | Epidemiology of Microbial Diseases, Yale School of Public Health | A.E.; A.F.; Alpert, T.; Breban; Brito; C.B.; C.C.; Fauver; Grubaugh; I.M.; Iwasaki, A.; J.E.; J.R.; Kalinich; Landry; Lucas, C.; M.E.; M.I.; M.L.; N.D.; Ott; Petrone; Rothman; Vogels; Watkins |
| EPI_ISL_1898554, EPI_ISL_2241494, EPI_ISL_2241623 | Ethiopian Biotechnology Institute (EBTI) | International Centre for Genetic Engineering and Biotechnology (ICGEB) and ARGO Open Lab for Genome Sequencing | Alessandro Marcello; Danilo Licastro; Emanuele Orsini; Getnet Hailu; Hailu Dadi; Kassahun Tesfaye; Keyru Tuki; Kominist Asmamaw; Molalegne Bitew; Simeone Dal Monego; Yakob Gebregziabher Tsegay |
| EPI_ISL_877391 | Eurofins Diatherix | Hudsonalpha Genome Sequencing Center | Ada Stewart; Jane Grimwood; Jenell Webber; Jennifer Cart; John Lovell; Joshua Stough; Leslie Malone; Lori H. Handley; Melissa Williams; Stefan Brzezinski; Teresa Jones; and Jeremy Schmutz |
| EPI_ISL_1570090, EPI_ISL_1571260, EPI_ISL_1571269, EPI_ISL_1846037, EPI_ISL_1846048, EPI_ISL_1846115, EPI_ISL_1846176, EPI_ISL_1846201, EPI_ISL_2259578 | | | |
| see above | Eurofins LifeCodexx GmbH | Robert Koch Institute | |
| EPI_ISL_1571477, EPI_ISL_2110761, EPI_ISL_2110762, EPI_ISL_2316402, EPI_ISL_2316415, EPI_ISL_2845449 | Eurofins MVZ Labor Gelsenkirchen | Robert Koch Institute | |
| EPI_ISL_2803756 | Everight Diagnostic Laboratory | National Reference Laboratory, Nigeria Centre for Disease Control | Anthony Ahumibe; Catherine Okoi; Chimaobi Chukwu; Dr Chikwe Ihekweazu; Dr Ndodo Nnaemeka; Dr Omoare Adesuyi; Grace Esebanmen; Nwando Mba; Olusola Anuoluwapo Akanbi |
| EPI_ISL_2803754 | Everight Diagnostic Laboratory | National Reference Laboratory, Nigeria Centre for Disease Control Durumi III, Gaduwa, Abuja, Nigeria | Anthony Ahumibe; Catherine Okoi; Chimaobi Chukwu; Dr Chikwe Ihekweazu; Dr Ndodo Nnaemeka; Dr Omoare Adesuyi; Grace Esebanmen; Nwando Mba; Olusola Anuoluwapo Akanbi |
| EPI_ISL_1712303 | FSBI «NATIONAL MEDICAL RESEARCH CENTER FOR OBSTETRICS, GYNECOLOGY AND PERINATOLOGY NAMED AFTER ACADEMICIAN V.I.KULAKOV» MINISTRY OF HEALTHCARE OF THE RUSSIAN FEDERATION | Center for Precision Genome Editing and Genetic Technologies for Biomedicine, Pirogov Medical University, Moscow, Russian Federation | Anastasia Shut; Andrey Krivoy; Dmitriy Korostin; Margarita Korzhanova; Vera Belova; Yegor Botsmanov |
| EPI_ISL_1491576, EPI_ISL_1538420, EPI_ISL_1538429 | Faculty of Medicine Vajira Hospital | COVID-19 Network Investigations (CONI) Alliance | Ampan Vimonvattana; Anan Manomaipiboon; Angkana Huang; Anthony R. Jones; Arporn Wangwiwatsin; Bhakbhoom Panthan; Chayanit Phutthanu; Chonticha Klungtong; Duangkamon Loesbanluechai; Ekawat Pasomsub; Elizabeth Batty; Insee Sensorn; Jakravoot Maneerit; Janjira Thaipadungpanit; Khajohn Joonlasak; Kingkan Rakmanee; Krittikorn Kumpornsin; Namfon Kotanan; Stefan Fernandez; Sunisa Dongphooyao; Thanat Chookajorn; Theerarat Kochakarn; Tonsan Hansirisathit; Treewat Watthanachockchai; Uraporn Phumisantiphong; Wasun Chantratita; Wipawee Thongsopa; Wudtichai Manasatienkij |
| EPI_ISL_1273102 | Faculty of Medicine, Al-Quds University | Faculty of Medicine, Al-Quds University | Al-Jawabreh, A.; Dumaidi, K.; Ereqat, S.; Nasereddin, A. |
| EPI_ISL_451966 | Federal Budget Institution of Science, State Research Center for Applied Microbiology & Biotechnology | Federal Budget Institution of Science, State Research Center for Applied Microbiology & Biotechnology | Abaimova A; Bakhteeva I; Blagodatskikh S; Bogun A; Borzilov A; Chekan L; Chernysh S; Denisenko E; Dentovskaya S; Detushev K; Detusheva E; Dyatlov I; Firstova V; Frolov V; Fursov M; Fursova N; Galkina E; Gapelchenkova T; Goncharova J; Gorbatov A; Hlyntseva A; Ivanov S; Kalmantayev T; Kalmantayeva O; Kanashenko M; Kartsev N; Kartseva A; Khomyakov A; Khramov M; Kislichkina A; Kolchanova A; Koroleva-Ushakova A; Kosilova I; Krasilnikova E; Kuzin V; Kuzina E; Makarova M; Marin M; Novikova T; Platonov M; Podkopaev Y; Ryabko A; Shaikhutdinova R; Shemyakin I; Shishkina L; Silkina M; Sizova A; Skryabin Y; Slukin P; Slukina N; Solomentsev V; Solovieva A; Teymurazov M; Timofeev V; Titareva G; Trunyakova A; Tyurin E; Vagayskaya A; Zeninskaya N; Zhumakaev R |
| EPI_ISL_2507666, EPI_ISL_2507677, EPI_ISL_2508274 | Fimlab Laboratoriot Oy Tampere | Expert Microbiology, National Institute for Health and Welfare | Carita Savolainen-Kopra; Erika Lindh; Haider al-Hello; Jani Halkilahti; Kirsi Liitsola; Niina Ikonen; Olli Vapalahti; Pekka Ellonen; Phuoc Truong; Päivi Laurila; Ravi Kant; Sari Hannula; Soile Blomqvist; Teemu Smura |
| EPI_ISL_509726, EPI_ISL_1030086, EPI_ISL_2008721, EPI_ISL_2008761 | Florida Bureau of Public Health Laboratories | Florida Bureau of Public Health Laboratories | Jason Blanton; Sarah Schmedes |
| EPI_ISL_590899, EPI_ISL_708077 | Foerde Hospital, Department of Microbiology | Norwegian Institute of Public Health, Department of Virology | Hilde Elshaug; Hilde Vollan; Kamilla Heddeland Instefjord; Karoline Bragstad; Kathrine Stene-Johansen; Marie Paulsen Madsen; Olav Hungnes; Rasmus Riis Kopperud |
| EPI_ISL_476139 | Folkhalsomyndigheten | The Public Health Agency of Sweden | Anna Risberg; Anna-Malin Linde; Karin Tegmark-Wisell; Maria Lind Karlberg; Mattias Haukland; Olov Svartstrom; Oskar Karlsson Lindsjo; Petra Edquist; Reza Advani; Sandra Broddesson; Shamam Muradrasoli |
| EPI_ISL_2889833, EPI_ISL_2889839, EPI_ISL_2889841, EPI_ISL_2889844, EPI_ISL_2889846, EPI_ISL_2889847, EPI_ISL_2889848, EPI_ISL_2889849, EPI_ISL_2889850, EPI_ISL_2889854 | | | |
| see above | Fondation Congolaise pour la recherche medicale (FCRM) | Fondation Congolaise pour la Recherche Médicale | Abel Lissom; Batchi-Bouyou Armel Landry; Francine Ntoumi; Jean Claude Djontu; Mfoutou Mapanguy Claujens Chastel; Thirumalaisamy P. Velavan |
| EPI_ISL_581455, EPI_ISL_581491, EPI_ISL_581492 | Fondation Congolaise pour la recherche medicale (FCRM) | NGS Competence Center Tübingen, Institut für Medizinische Mikrobiologie und Hygiene, Universitätsklinikum Tübingen | Angel Angelov |
| EPI_ISL_2087052 | Fondation Congolaise pour la recherche medicale (FCRM), Francine Ntoumi | Centre de Recherche sur les Maladies Infectieuses | Mfoutou Mapanguy Claujeans Chastel and Batchi-Bouyou Armel Landry; Prof. Dr. Thirumalaisamy P. Velavan; Prof. Francine Ntoumi |
| EPI_ISL_2964949, EPI_ISL_2964950, EPI_ISL_2964955, EPI_ISL_3040125, EPI_ISL_3040129, EPI_ISL_3040130, EPI_ISL_3040133, EPI_ISL_3040134, EPI_ISL_3040135, EPI_ISL_3040137 | | | |
| see above | Fondation Congolaise pour la recherche medicale (FCRM), Francine Ntoumi | Fondation Congolaise pour la Recherche Médicale | Batchi-Bouyou Armel Landry; Dr. Abel Lissom; Dr. Jean Claude Djontu; Mfoutou Mapanguy Claujens Chastel; Prof. Dr. Thirumalaisamy P. Velavan; Prof. Francine Ntoumi |
| EPI_ISL_1671925, EPI_ISL_1840863, EPI_ISL_1854769, | Fondation Congolaise pour la recherche medicale (FCRM), Francine Ntoumi | Institute of Tropical Medicine | Mfoutou Mapanguy Claujeans Chastel and Batchi-Bouyou Armel Landry; Prof. Dr. Thirumalaisamy P. Velavan; Prof. Dr. Thirumalaisamy P. Velavan and Prof. Francine Ntoumi; Prof. Francine Ntoumi |

| | | | |
|---|---|---|---|
| EPI_ISL_1854789, EPI_ISL_1854794, EPI_ISL_1857281 | | | |
| EPI_ISL_912358, EPI_ISL_912371, EPI_ISL_912394 | Fondation Congolaise pour la recherche medicale (FCRM), Francine Ntoumi | NGS Competence Center Tuebingen, Institut für Medizinische Mikrobiologie und Hygiene, Universitaetsklinikum Tübingen | Angel Angelov |
| EPI_ISL_1666171, EPI_ISL_1992782, EPI_ISL_2038596, EPI_ISL_2525602, EPI_ISL_2785604, EPI_ISL_2785740, EPI_ISL_2928732 | | | |
| see above | Fulgent Genetics | Centers for Disease Control and Prevention Division of Viral Diseases, Pathogen Discovery | Adrian Paskey; Becky Tsai; Benafsh Sapra; Benjamin Rambo-Martin; Christopher Gulvick; Clinton R. Paden; Dakota Howard; Darlene Wagner; Dhwani Batra; Doreen Ng; Duncan MacCannell; Harry Gao; James Xie; Jason Caravas; John Gao; Joseph Fierro; Kara Moser; Matthew Schmerer; Mickey Li; Peter W. Cook; Scott Sammons; Shatavia Morrison; Yan Meng; Yvette Unoarumhi |
| EPI_ISL_1026982, EPI_ISL_1027144, EPI_ISL_1068425 | Fulgent Genetics | Fulgent Genetics | Becky Tsai; Benafsh Sapra; Doreen Ng; Harry Gao; James Xie; John Gao; Joseph Fierro; Mickey Li; Yan Meng |
| EPI_ISL_1182612 | Fundação Ezequiel Dias (FUNED) | Coordenação Geral de Laboratórios de Saúde Pública (CGLAB/DAEVS/SVS/MS) | Vagner Fonseca; et al. |
| EPI_ISL_493362, EPI_ISL_2788586, EPI_ISL_2925776, EPI_ISL_3008324 | Furst Medical Laboratory | Norwegian Institute of Public Health, Department of Virology | Atiya R Ali; Debech Nadia; Engebretsen Serina Beate; Garcia Llorente Ignacio; Hilde Elshaug; Hilde Vollan; Jon Bråte; Kamilla Heddeland Instefjord; Karoline Bragstad; Kathrine Stene-Johansen; Line Victoria Moen; Marie Paulsen Madsen; Olav Hungnes; Pedersen Benedikte Nevjen; Rasmus Riis Kopperud |
| EPI_ISL_1318590 | GA Department of Public Health Laboratory | Centers for Disease Control and Prevention Division of Viral Diseases, Pathogen Discovery | Anna Montmayeur; Anna Uehara; Ben L. Rambo-Martin; Clinton R. Paden; Dhwani Batra; Haibin Wang; Jasmine Padilla; Jing Zhang; Justin Lee; Katie Dillon; Krista Queen; Kristen Knipe; Kristine Lacek; Lori Rowe; Mark Burroughs; Matthew Schmerer; Mili Sheth; Peter W. Cook; Rachel Marine; Sam Shepard; Sarah Nobles; Shoshona Le; Suxiang Tong; Yan Li; Ying Tao |
| EPI_ISL_1672509, EPI_ISL_2333929 | GH A.CHENEVIER-H.MONDOR | Department of Virology, Henri Mondor University Hospital, Assistance Publique Hôpitaux de Paris, Université Paris-Est Créteil, INSERM U955 | Alexandre Soulier; Christophe Rodriguez; Elisabeth Trawinski; Guillaume Gricourt; Jean-Michel Pawlotsky; Melissa N'Debi; Slim Fourati; Vanessa Demontant |
| EPI_ISL_2779388 | GK Prison Kiambu | USAMRD-A, Basic Science Laboratory | Alan Lemtudo; Beth Mutai; Brian Andika; Carol Kifude; Clement Masakwe; Eric Muthanje; Esther Omuseni; Faith Sigei; Gathii Kimita; George Awinda; John Waitumbi; Josphat Nyataya; Rachel Githii; Rehema Liyai; Stephen Ochola |
| EPI_ISL_2779313, EPI_ISL_2779316 | GK Prisons Dispensary Kapsabet | USAMRD-A, Basic Science Laboratory | Alan Lemtudo; Beth Mutai; Brian Andika; Carol Kifude; Clement Masakwe; Eric Muthanje; Esther Omuseni; Faith Sigei; Gathii Kimita; George Awinda; John Waitumbi; Josphat Nyataya; Rachel Githii; Rehema Liyai; Stephen Ochola |
| EPI_ISL_2864606 | Ga East Hospital | UHAS COVID-19 Lab | John O. Gyapong and the UHAS COVID-19 Lab Team; Jones Gyamfi; Kwabena O. Duedu; Reuben Ayivor-Djanie |
| EPI_ISL_831977 | Gavle klinisk mikrobiologi | The Public Health Agency of Sweden | Department of Microbiology; The Public Health Agency of Sweden |
| EPI_ISL_730577, EPI_ISL_984749 | Gazi University Faculty of Medicine, Medical Virology Laboratory | Gazi University Faculty of Medicine, Medical Virology Laboratory | Erdem Şahin; Gülendam Bozdayı; Hager Muftah; Işıl Fidan; Kayhan Çağlar; Murat Dizbay; Selin Yiğit; Shaknoza Sarzhanova; Özlem Güzel Tunçcan |
| EPI_ISL_3026015, EPI_ISL_3026016, EPI_ISL_3026019, EPI_ISL_3026020 | Geelong Centre for Emerging Infectious Diseases, Deakin University | Geelong Centre for Emerging Infectious Diseases, Deakin University | Alexandersen, S.; Bhatta; Chamings, A.; T.R. |
| EPI_ISL_2681409, EPI_ISL_2681439 | General Hospital "8th of September" | Laboratory of virology and molecular diagnostics, Institute of Public Health | Boshevska G; Janchevska E.; Kuzmanovska M |
| EPI_ISL_933566, EPI_ISL_934314 | General Hospital - Bitola | Research Center for Genetic Engineering and Biotechnology "Georgi D. Efremov" , Macedonian Academy of Sciences and Arts | RCGEB - MASA |
| EPI_ISL_933775 | General Hospital - Ohrid | Research Center for Genetic Engineering and Biotechnology "Georgi D. Efremov" , Macedonian Academy of Sciences and Arts | RCGEB - MASA |
| EPI_ISL_677674 | General Hospital - Prilep | Research Center for Genetic Engineering and Biotechnology "Georgi D. Efremov" , Macedonian Academy of Sciences and Arts | RCGEB - MASA |
| EPI_ISL_933602, EPI_ISL_934413 | General Hospital - Shtip | Research Center for Genetic Engineering and Biotechnology "Georgi D. Efremov" , Macedonian Academy of Sciences and Arts | RCGEB - MASA |
| EPI_ISL_3020343 | General Hospital Tetovo | Laboratory of virology and molecular diagnostics, Institute of Public Health | Boshevska G; Janchevska E.; Kuzmanovska M |
| EPI_ISL_406798 | General Hospital of Central Theater Command of People's Liberation Army of China | BGI & Institute of Microbiology, Chinese Academy of Sciences & Shandong First Medical University & Shandong Academy of Medical Sciences & General Hospital of Central Theater Command of People's Liberation Army of China | Weifeng Shi and Zhenhong Hu; Weijun Chen; Yuhai Bi |
| EPI_ISL_746523, EPI_ISL_1167697, EPI_ISL_1167721, EPI_ISL_1167794, EPI_ISL_1167814, EPI_ISL_1167878, EPI_ISL_1167922, EPI_ISL_1167925, EPI_ISL_1300477, EPI_ISL_1321468, EPI_ISL_1321493, EPI_ISL_1321508, EPI_ISL_1534569, EPI_ISL_1534583, EPI_ISL_1541011, EPI_ISL_1541012, EPI_ISL_1541035, EPI_ISL_1633372, EPI_ISL_1633383, EPI_ISL_1633418, EPI_ISL_1633500, EPI_ISL_2009203, EPI_ISL_2009256, EPI_ISL_2009293, EPI_ISL_2009637, EPI_ISL_2391042, EPI_ISL_2391174, EPI_ISL_2391176, EPI_ISL_2391269, EPI_ISL_2391355, EPI_ISL_2391412, EPI_ISL_2391442, EPI_ISL_2543475, EPI_ISL_2543539, EPI_ISL_2543664, EPI_ISL_2557275, EPI_ISL_2597292, EPI_ISL_2659168, EPI_ISL_2659197, EPI_ISL_2756220, EPI_ISL_2756329, EPI_ISL_2756330, EPI_ISL_2756336, EPI_ISL_2756337, EPI_ISL_2892690, EPI_ISL_2892741, EPI_ISL_2892797, EPI_ISL_2894889, EPI_ISL_2896177 | | | |
| see above | Genetica Molecular and Subdepartamento de Virologia ISP Chile | Instituto de Salud Publica de Chile | Andres Castillo; Barbara Parra; Constanza Campano; Gisselle Barra; Jaime Lagos; Javier Tognarelli; Jorge Fernandez; Karen Orostica; Loredana Arata; Patricia Bustos; Rodrigo Fasce; Soledad Ulloa |
| EPI_ISL_480245, EPI_ISL_735371 | Genomic Laboratory (GLAB) (Conjoint lab of Health Directorate of Istanbul and Istanbul Technical University) | Genomic Laboratory (GLAB), Istanbul Technical University | Arzu Irvem; Ayse Serra Ozel; Betsi Kose; Betsi Köse; Bugra Agaoglu; Elifnaz Çelik; Gizem Alkurt; Gizem Dinler Doganay; Ilker Karacan; Jale Yildiz; Levent Doganay; Mehtap Aydin; Mehtap Aydın; Nihat Bugra Agaoglu; Nilsun Altunal; Nisan Denizce Can; Ozlem Akgun Dogan; Payam Zolfagharian; Tugba Kizilboga Akgun; Yasemin Kendir Demirkol |
| EPI_ISL_1673680 | Genomic Research Lab, BCSIR | Genomic Research Lab, BCSIR | Abu Sayeed Mohammad Mahmud; Barna Goswami; Eshrar Osman; Iffat Jahan; Md. Ahasan Habib; Md. Murshed Hasan Sarkar; Md. Saddam Hossain; Md. Salim Khan; Mohammad Mohi Uddin; Mohammad Samir Uzzaman; Shahina Akter; Tanjina Akhter Banu |
| EPI_ISL_2862015 | Genomic Research Laboratory, Bangabandhu Sheikh Mujib Medical University (BSMMU) Shahbag, Dhaka_1000 | Genomic Research Laboratory, BSMMU | Arifa Akram; Laila Anjuman Banu; Md.Saydur Rahman; Md.Sharfuddin Ahmed; Sharadindu Kanti Sinha |
| EPI_ISL_632908 | Genomic Sciences, Rehman Medical Institute | Genomic Sciences, Rehman Medical Institute | Afridi; Ali, J.; H. and Jehanzeb, V.; Haider; Jan; S.A.; Sabiha, B.; U.K. |
| EPI_ISL_812783, EPI_ISL_812784, EPI_ISL_812785, EPI_ISL_812786, EPI_ISL_812787, EPI_ISL_812788, EPI_ISL_812789, EPI_ISL_812790, EPI_ISL_812791, EPI_ISL_812792, EPI_ISL_812793, EPI_ISL_812794, EPI_ISL_812795, EPI_ISL_812796, EPI_ISL_812797, EPI_ISL_812798, EPI_ISL_812799, EPI_ISL_812800, EPI_ISL_812801, EPI_ISL_812802, EPI_ISL_812803, EPI_ISL_812804, EPI_ISL_812805, EPI_ISL_812806, EPI_ISL_812807, EPI_ISL_812808, EPI_ISL_812809, EPI_ISL_812810, EPI_ISL_812811, EPI_ISL_812812, EPI_ISL_812813, EPI_ISL_812814, EPI_ISL_812815, EPI_ISL_812816, EPI_ISL_812817, EPI_ISL_812818, EPI_ISL_812819, EPI_ISL_812820, EPI_ISL_812821, EPI_ISL_812822, EPI_ISL_812823, EPI_ISL_812824, EPI_ISL_812825, EPI_ISL_812826, EPI_ISL_812827, EPI_ISL_812828, EPI_ISL_812829, EPI_ISL_812830, EPI_ISL_812831, EPI_ISL_812832, EPI_ISL_812833, EPI_ISL_812834, EPI_ISL_812835, EPI_ISL_812836, EPI_ISL_812837, EPI_ISL_812838, EPI_ISL_812839, EPI_ISL_812840, EPI_ISL_812841, EPI_ISL_812842, EPI_ISL_812843, EPI_ISL_812844, EPI_ISL_812845, EPI_ISL_812846, EPI_ISL_812847, EPI_ISL_812848, EPI_ISL_812849, EPI_ISL_812850, EPI_ISL_812851, EPI_ISL_812852, EPI_ISL_812853, EPI_ISL_812854, EPI_ISL_812855, EPI_ISL_812856, EPI_ISL_812857, EPI_ISL_812858, EPI_ISL_812859, EPI_ISL_812860, EPI_ISL_812861, EPI_ISL_812862, EPI_ISL_812863, EPI_ISL_812864, EPI_ISL_812865, EPI_ISL_812866, EPI_ISL_812867, EPI_ISL_812868, EPI_ISL_812869, EPI_ISL_812870, EPI_ISL_812871, EPI_ISL_812872, EPI_ISL_2566467, EPI_ISL_2566468, EPI_ISL_2566469, EPI_ISL_2566470, EPI_ISL_2566471, EPI_ISL_2566472, EPI_ISL_2566473, EPI_ISL_2566474, EPI_ISL_2566475, EPI_ISL_2566476, EPI_ISL_2566477, EPI_ISL_2566478, EPI_ISL_2566479, EPI_ISL_2566480, EPI_ISL_2566481, EPI_ISL_2566482, EPI_ISL_2566483, EPI_ISL_2566484, EPI_ISL_2566485, EPI_ISL_2566486, EPI_ISL_2566487, EPI_ISL_2566488, EPI_ISL_2566489, EPI_ISL_2566490, EPI_ISL_2566491, EPI_ISL_2566492, EPI_ISL_2566493, EPI_ISL_2566494, EPI_ISL_2566495, EPI_ISL_2566496, EPI_ISL_2566497, EPI_ISL_2566498, EPI_ISL_2566499, EPI_ISL_2566500, EPI_ISL_2566501, EPI_ISL_2566502, EPI_ISL_2566503, EPI_ISL_2566504, EPI_ISL_2566505, EPI_ISL_2566506, EPI_ISL_2566507, EPI_ISL_2566508, EPI_ISL_2566509, EPI_ISL_2566510, EPI_ISL_2566511, EPI_ISL_2566512, EPI_ISL_2566513, EPI_ISL_2566514, EPI_ISL_2566515, EPI_ISL_2566516, EPI_ISL_2566517, EPI_ISL_2566518, EPI_ISL_2566519, EPI_ISL_2566520, EPI_ISL_2566521, EPI_ISL_2566522, EPI_ISL_2566523, EPI_ISL_2566524, EPI_ISL_2566525, EPI_ISL_2566526, EPI_ISL_2566527, EPI_ISL_2566528, EPI_ISL_2566529, EPI_ISL_2566530 | | | |
| see above | Genomics Program, Children Cancer Hospital | Genomics Program, Children Cancer Hospital | Abdelaziz, H.; Abdo, I.; Abouelnaga, S.; Almeldin, A.; Amer, K.; Bakry, U.; Diab, A.; El-Shaqnqery, H.; El-Zayat, M.; ElHaddad, A.; ElHalafawy, A.; Elnaqeeb, M.; Farawela, H.; Farawyla, H.; Gomaa, C.; Hadad, A.; Halafawy, A.; Hammad, M.; Hassan, R.; Hassan, W.; Hatem, A.; Hossam, M.; Hussein, S.; Ismail, J.; Jalal, D.; Magdeldin, S.; Mansour, T.; Monuir, G.; Saaid, M.; Said, D.; Salah, H.; Samir, O.; Sayed, A.; Shalaby, L.; Soliman, M.; Soliman, S.; Yahia, A.; shalaby, L. |
| EPI_ISL_2510688 | Genomics Program, Children Cancer Hospital | Genomics Program, Children Cancer Hospital | Abdelaziz, H.; Abdo, I.; Abouelnaga, S.; Almeldin, A.; Amer, K.; Diab, A.; El-Shaqnqery, H.; El-Zayat, M.; ElHaddad, A.; ElHalafawy, A.; Elnaqeeb, M.; Farawela, H.; Hammad, M.; Hassan, R.; Hassan, W.; Hatem, A.; Hossam, M.; Hussein, S.; Ismail, J.; Jalal, D.; Mansour, T.; O. and Sayed, A.; Saaid, M.; Said, D.; Salah, H.; Samir; Shalaby, L.; Soliman, S.; Yahia, A. |
| EPI_ISL_2434978 | Genomik Solidaritas Indonesia Laboratorium | Genomik Solidaritas Indonesia Laboratorium | Alfin Mohammad Abdillah; Anindya Pradipta Susanto; Annisa Muthiah Sukirman; Ariel Pradipta; Carissa Sintca Wijaya; Dhahlia Agustina Cahyono; Gracia Felias Enos Korompis; Meutia Ayuputeri Kumaheri; Normastuti Adhini Tantyo; Vania Gavrila Wikasa |
| EPI_ISL_2140696, EPI_ISL_2802110 | George Hospital wc GRH | NHLS/UCT | Arash Iranzadeh; Bruna Galvao; Carolyn Williamson; Deelan Doolabh; Diana Hardie; Gert Marais; Innocent Mudau; Kruger Marais; Lynn Tyers; Marvin Hsiao; Stephen Korsman |
| EPI_ISL_1744345, EPI_ISL_1744346, EPI_ISL_2248892, EPI_ISL_2340163, EPI_ISL_2340181 | Germano de Sousa | Instituto Nacional de Saude (INSA) | Borges et al |
| EPI_ISL_2340334, EPI_ISL_2340355 | Germano de Sousa | Instituto Nacional de Saude (INSA) and BioSystems & Integrative Sciences Institute (BioISI) Genomics Unit, FCUL | Borges et al |
| EPI_ISL_1853560 | Germano de Sousa | Instituto Nacional de Saude (INSA) and Instituto Gulbenkian de Ciencia (IGC) | Borges et al |
| EPI_ISL_960672 | Germano de sousa | Instituto Gulbenkian de Ciencia | Cathy Paulino; João Costa; João Sobral; Maria Costa; Ricardo Leite; Susana Ladeiro |
| EPI_ISL_933769 | Gerontology Institute - Skopje | Research Center for Genetic Engineering and Biotechnology "Georgi D. Efremov" , Macedonian Academy of Sciences and Arts | RCGEB - MASA |

| | | | |
|---|---|---|---|
| EPI_ISL_890188 | Gonoshasthaya-RNA Research Center, Gonoshasthaya-RNA Molecular Diagnostics and Research Center | Gonoshasthaya-RNA Research Center, Gonoshasthaya-RNA Molecular Diagnostics and Research Center | Adnan, N.; Ahmed; Akter, S.; Ali, T.; Azmuda, N.; Chaity, M.; Haq; Jahan, N.; Jamiruddin; Jamiruddin, M.; Khandker; Khondoker; Liza; M.A.; M.F.; M.R.; M.U.; Marzan, M.; Mou; Nahar, S.; Oishee, M.; S.M.; S.S.; Sharif, N.; Sharmin, S.; T.J. |
| EPI_ISL_774976 | Gonoshasthya-RNA Molecular Diagnostic and Research Center | Gonoshasthya-RNA Molecular Diagnostic and Research Center | Firoz Ahmed; Maha Jamiruddin; Mahfuza Marzan; Md. Ahsanul Haq; Mohd. Raeed Jamiruddin; Mohib Ullah Khondoker; Mousumi Chaity; Mumtarin Jannat Oishee; Nafisa Azmuda; Nihad Adnan; Nowshin Jahan; Salma Akter; Sayeda Moriam Liza; Shahad Saif Khandker; Shahana Sharmin; Tamanna Ali; Taslin Jahan Mou |
| EPI_ISL_496860, EPI_ISL_1225352, EPI_ISL_1225354, EPI_ISL_1225513, EPI_ISL_1225516, EPI_ISL_1225521, EPI_ISL_1225563, EPI_ISL_1225574, EPI_ISL_1225577, EPI_ISL_1502846, EPI_ISL_1502915, EPI_ISL_1502937, EPI_ISL_1502983, EPI_ISL_1502997, EPI_ISL_1503020, EPI_ISL_1503114, EPI_ISL_1503116 | | | |
| see above | Gorgas Memorial Laboratory of Health Studies | Gorgas Memorial Laboratory of Health Studies | Adriana Weeden; Alejandra Valoy; Alexander A Martinez; Alexander Martinez; Ambar Moreno; Anyuri Ortiz; Brechla Moreno; Castillo Jorge; Claudia Gonzalez; Claudia Gonzalez Sandra Lopez-Verges; Daniel Castillo; Danilo Franco; Davis Beltran; Dimelza Arauz; Elimelec Valdespino; Franco Danilo; Gonzalez Claudia; Gretel Vasquez; Ilka Guerra; Isela Guerrero; Jessica Gondola; Jim Chang; Juan Miguel Pascale; Layda Abrego; Leyda Abrego; Lisseth Saenz; Lopez-Verges Sandra; Mabel Martinez-Montero; Maria Chen-German; Marlene Castillo; Marlenne Castillo; Martinez Alexander; Melissa Gaitan; Moreno Ambar; Moreno Brechla; Oris Chavarria; Ortiz Alma; Rita Corrales; Rita Rodriguez; Sandra Lopez-Verges; Yamilka Diaz; Yaneth Pitti; Zumara Chaverra |
| EPI_ISL_1302710 | Gravity Diagnostics | Kentucky State Public Health Lab | Joshua Tobias; Karim George; Matthew Johnson; Rachel Zinner; Rhonda Lucas; Stephanie Lunn; Vaneet Arora; William Grooms |
| EPI_ISL_2602113, EPI_ISL_2614003, EPI_ISL_2985763 | Gravity Diagnostics, LLC | Gravity Diagnostics, LLC | Gravity Diagnostics |
| EPI_ISL_2301649, EPI_ISL_2301783, EPI_ISL_2369069, EPI_ISL_2371278 | Greek Genome Center, Biomedical Research Foundation of the Academy of Athens (BRFAA) | Greek Genome Center, Biomedical Research Foundation of the Academy of Athens (BRFAA) | Dimitrios Thanos; Emmanouil Athanasiadis; Giannis Vatsellas; Katerina Zoi; Theodoros Loupis |
| EPI_ISL_1040654, EPI_ISL_1706573, EPI_ISL_2140691, EPI_ISL_2140704, EPI_ISL_2140712, EPI_ISL_2375973, EPI_ISL_2621054, EPI_ISL_2876955 | | | |
| see above | Groote Schuur Hospital wc GSH | NHLS/UCT | Arash Iranzadeh; Bruna Galvao; Carolyn Williamson; Deelan Doolabh; Diana Hardie; Emmanuel SJ; Gert Marais; Innocent Mudau; Kruger Marais; Lynn Tyers; Marvin Hsiao; Stephen Korsman; Tegally H; de Oliveira T |
| EPI_ISL_2547418 | Groote Schuur Hospital wc GSH | National Health Laboratory Service/University of Cape Town (NHLS/UCT) | Arash Iranzadeh; Bruna Galvao; Carolyn Williamson; Deelan Doolabh; Diana Hardie; Gert Marais; Innocent Mudau; Lynn Tyers; Marvin Hsiao; Stephen Korsman |
| EPI_ISL_699059 | Group 42 (G42) Healthcare, Abu Dhabi, United Arab Emirates; Department of Health, The United Arab Emirates | G42 Healthcare | Ashish Koshy; Budoor Alqarni; Denghui Liu; Fang Chen; Hanif Khalak; Huanming Yang; Javier Quilez; Jian Wang; Junhua Li; Ke Liang; Long Lin; Mohammed Saifuddin Fasihuddin; Nan Qiao; Nawal Ahmed Mohamed Al Kaabi; Pauline Ogrodzki; Pei Wu; Peng Xiao; Pengjuan Liu; Rong Liu; Sally Mahmoud; Siyang Liu; Stephen S. Francis; Tao Ma; Vinay Kusuma; Walid Abbas Zaher; Weibin Liu; Wenjun He; Xavier Anton; Xin Jin; Xin Meng; Xinyu Huang; Xun Xu; Zhaorong Yuan |
| EPI_ISL_3033081 | Grupo de Genómica y Bioinformática del Instituto de Investigación de la Cadena Láctea CONICET-INTA on behalf of 'Proyecto Argentino Interinstitucional de genomica de SARS-CoV-2' (PAIS Consortium) | Grupo de Genómica y Bioinformática del Instituto de Investigación de la Cadena Láctea CONICET-INTA on behalf of 'Proyecto Argentino Interinstitucional de genomica de SARS-CoV-2' (PAIS Consortium) | AF; Amadio; Eberhardt; Irazoqui; JM; MF |
| EPI_ISL_1824180, EPI_ISL_2339776, EPI_ISL_2339777, EPI_ISL_2339778, EPI_ISL_2339779, EPI_ISL_2339780, EPI_ISL_2339781, EPI_ISL_2339782, EPI_ISL_2339783, EPI_ISL_2339784, EPI_ISL_2339785, EPI_ISL_2339786, EPI_ISL_2339787, EPI_ISL_2339788, EPI_ISL_2339789, EPI_ISL_2339790, EPI_ISL_2339791, EPI_ISL_2339792, EPI_ISL_2339793, EPI_ISL_2339794, EPI_ISL_2603146 | | | |
| see above | Grupo de Investigación en Enfermedades Tropicales del Ejército (GINETEJ), Laboratorio de Referencia e Investigación, Dirección de Sanidad Ejército, Bogotá, Colombia | Centro de Investigaciones en Microbiología y Biotecnología-UR (CIMBIUR), Facultad de Ciencias Naturales, Universidad del Rosario, Bogotá, Colombia | Camilo A. Correa-Cárdenas; Carolina Oliveros; Claudia Méndez; Elizabeth K. Márquez; Frank de los Santos Ortiz; Juan David Ramírez; Julie Pérez; Lorena Albarracín; Luz H. Patiño; Maria Clara Duque; Marina Muñoz; María Teresa Alvarado; Nathalia Ballesteros; Sergio Castañeda; Sergio Gutíerrez-Riveros; Yanira Romero; Zulma Cucunubá |
| EPI_ISL_1273073, EPI_ISL_1823585, EPI_ISL_2451659, EPI_ISL_2787627, EPI_ISL_2787642 | Guam Public Health Laboratory | Centers for Disease Control and Prevention Division of Viral Diseases, Pathogen Discovery | Alison Laufer Halpin; Anna Montmayeur; Anna Uehara; Ben L. Rambo-Martin; Clinton R. Paden; Dakota Howard; Darlene Wagner; Dave Wentworth; Dhwani Batra; Haibin Wang; Jasmine Padilla; Jing Zhang; Justin Lee; Katie Dillon; Krista Queen; Kristen Knipe; Kristine Lacek; Lori Rowe; Mark Burroughs; Matthew Schmerer; Mili Sheth; Peter Cook; Peter W. Cook; Rachel Marine; Sam Shepard; Sarah Nobles; Shoshona Le; Suxiang Tong; Vivien Dugan; Yan Li; Ying Tao; Yvette Unoarumhi |
| EPI_ISL_509701 | Guatemala Ministry of Public Health | Pathogen Discovery, Respiratory Viruses Branch, Division of Viral Diseases, Centers for Disease Control and Prevention | Anna Uehara; Clinton Paden; Haibin Wang; Jing Zhang; Krista Queen; Suxiang Tong; Yan Li; Ying Tao |
| EPI_ISL_547739, EPI_ISL_942722 | Gundersen Molecular Diagnostics Laboratory | Kabara Cancer Research Institute | Craig S. Richmond; Paraic A. Kenny |
| EPI_ISL_1853561 | H Fernando Fonseca | Instituto Nacional de Saude (INSA) and Institute of Biomedicine (iBiMed), Universidade de Aveiro | Borges et al |
| EPI_ISL_693535 | H Vila Franca Xira | Instituto Nacional de Saude (INSA) | Borges et al |
| EPI_ISL_2695606, EPI_ISL_2709863 | HELEN JOSEPH | National Institute for Communicable Diseases of the National Health Laboratory Service | Amoako DG; Bhiman JN; Everatt J; Ismail A; Mahlangu B; Mnguni A; Mohale T; Ntuli N; Scheepers C |
| EPI_ISL_2984814, EPI_ISL_2984893, EPI_ISL_3024595 | HELEN JOSEPH LABORATORY | National Institute for Communicable Diseases of the National Health Laboratory Service | Amoako DG; Bhiman JN; Everatt J; Ismail A; Mahlangu B; Mnguni A; Mohale T; Ntuli N; Scheepers C |
| EPI_ISL_2385290, EPI_ISL_2523862, EPI_ISL_2899790 | HELIX LLC | WHO National Influenza Centre Russian Federation | Andrey Komissarov; Artem Fadeev; Daria Danilenko; Dmitry Lioznov; Elena Nabieva; Georgii Bazykin; Kirill Varchenko; Ksenia Safina; Kseniya Komissarova; Maria Pisareva; Maria Timofeeva; Mikhail Bakaev; Nikita Yolshin; Oula Mansour; Oula Masour; Tamila Musaeva; Veronika Eder |
| EPI_ISL_1180745, EPI_ISL_1965453 | HG Pharma GmbH | Bergthaler laboratory, CeMM Research Center for Molecular Medicine of the Austrian Academy of Sciences | Andreas Bergthaler; Anna Schedl; Bekir Erguner; Benedikt Agerer; Christoph Bock; Fabian Amman; Jan Laine; Lukas Endler; Maelle Le Moing; Martin Senekowitsch; Michael Schuster; Petr Triska; Thomas Penz |
| EPI_ISL_955236 | HGR 1 VICENTE GUERRERO | BIOBANCO / COCTI | Arias C; Borja-Aburto VH; Grajales-Muñiz C; Grande R; Isa P; López S; Muñoz-Medina JE; Ochoa Carrera LA; Rojas-Mendoza T; Santacruz Tinoco CE; Sánchez A; Taboada B |
| EPI_ISL_1805649 | HIC | Instituto Nacional de Salud- Dirección de Investigación en Salud Pública | Carlos Franco-Muñoz; Carmen Osorio; Diana Malo; Diego A. Álvarez-Díaz; Diego Andrés Prada; Gerardo Santamaría; Hector Alejandro Ruiz-Moreno; Jhonnatan Reales-González; Jorge Rivera; Juan Camilo Martinez; Julian Naizaque; Katherine Laiton-Donato; Lisseth Pardo; Magdalena Wiesner; Marcela Mercado-Reyes; Maria T. Herrera-Sepúlveda; Marta Lopez Blanco; Martha Lucia Ospina Martinez; Paola Rojas; Sergio Gomez; Sheryll Corchuelo; Ángela Alarcon Cruz |
| EPI_ISL_2484763, EPI_ISL_2484784, EPI_ISL_3019487, EPI_ISL_3019492 | HOME QUARANTINE TASKFORCE | Hong Kong Department of Health | Alan K.L. Tsang; Dominic N.C. Tsang; Edman T.K. Lam; Gannon C.K. Mak; Ken H.L. Ng; Peter C.W. Yip; Peter K.C. Cheng; Rickjason C.W. Chan |
| EPI_ISL_1940037 | HOSAHALLI | inStem NCBS - INSACOG | Uma Ramakrishnan Dasaradhi Palakodeti Aswin SaiNarain |
| EPI_ISL_2209295 | HOSP MUN DE MOGI DAS CRUZES PREF WALDEMAR COSTA FILHO | Instituto Butantan / Mendelics | Antonio Jorge Martins; Claudia Renata dos Santos Barros; David Schlesinger; Debora Botequio Moretti; Dimas Tadeu Covas; Elaine Cristina Marqueze; Elaine Vieira Santos; Evandra Strazza Rodrigues; Heidge Fukumasu; Jayme Augusto de Souza-Neto; José Salvatore Leister Patané; Luiz Alcantara; Luiz Lehmann Coutinho; Maria Carolina Elias; Maurício Lacerda Nogueira; Rafael dos Santos Bezerra; Raul Machado Neto; Rejane Maria Tommasini Grotto; Ricardo Haddad; Sandra Coccuzzo Sampaio Vessoni; Simone Kashima; Svetoslav Nanev Slavov; Vincent Louis Viala |
| EPI_ISL_2272985 | HOSPITAL CIUDAD NEILY | Inciensa, Instituto Costarricense de Investigación y Enseñanza en Nutrición y Salud | Adriana Godínez; Claudio Soto-Garita; Estela Cordero; Francisco Duarte; Hebleen Porras; Joselyn Prado & Raúl Zeledón-Mayorga; José Luis Vargas; Mariela Gutiérrez; Melany Calderón |
| EPI_ISL_2103379, EPI_ISL_2103412, EPI_ISL_2272972 | HOSPITAL DR. FERNANDO ESCALANTE PRADILLA | Inciensa, Instituto Costarricense de Investigación y Enseñanza en Nutrición y Salud | Adriana Godínez; Caterina Guzmán; Claudio Soto-Garita; Estela Cordero; Francisco Duarte; Hebleen Porras; Joselyn Prado & María Fernanda Matamoros Ceciliano; José Luis Vargas; Mariela Gutiérrez; Mariela Gutiérrez. Joselyn Prado; Melany Calderón; Nazareth Ruiz & Joselyn Prado; Nazareth Ruiz & Lizzeth Blanco |
| EPI_ISL_2103402 | HOSPITAL DR. MAX TERAN VALLS | Inciensa, Instituto Costarricense de Investigación y Enseñanza en Nutrición y Salud | Adriana Godínez; Caterina Guzmán; Claudio Soto-Garita; Estela Cordero; Francisco Duarte; Hebleen Porras; Joselyn Prado; José Luis Vargas; Mariela Gutiérrez; Melany Calderón; Nazareth Ruiz & María José Gómez-Umaña |
| EPI_ISL_3037832 | HOSPITAL GUAPILES [POCOCI/LIMON] | Inciensa, Instituto Costarricense de Investigación y Enseñanza en Nutrición y Salud | Adriana Godínez; Caterina Guzmán; Claudio Soto-Garita; Estela Cordero; Francisco Duarte; Hebleen Porras; José Luis Vargas; Mariela Gutiérrez Joselyn Prado; Melany Calderón; Nazareth Ruiz & Cristian Pérez |
| EPI_ISL_2502729 | HOSPITAL LA ANEXION | Inciensa, Instituto Costarricense de InvestigaciOn y Enseñanza en NutriciOn y Salud | Adriana Godínez; Claudio Soto-Garita; Estela Cordero; Francisco Duarte; Hebleen Porras; Jose Luis Vargas; Joselyn Prado & Ivanna Krize-Morún; Mariela Gutierrez; Melany CalderOn |
| EPI_ISL_3037842 | HOSPITAL LA ANEXION [NICOYA/GUANACASTE] | Inciensa, Instituto Costarricense de Investigación y Enseñanza en Nutrición y Salud | Adriana Godínez; Caterina Guzmán; Claudio Soto-Garita; Estela Cordero; Francisco Duarte; Hebleen Porras; José Luis Vargas; Mariela Gutiérrez Joselyn Prado; Melany Calderón; Nazareth Ruiz & Arnoldo Zamora |
| EPI_ISL_2657682, EPI_ISL_3037844 | HOSPITAL METROPOLITANO | Inciensa, Instituto Costarricense de Investigación y Enseñanza en Nutrición y Salud | Adriana Godínez; Caterina Guzmán; Claudio Soto-Garita; Estela Cordero; Francisco Duarte; Hebleen Porras; Joselyn Prado; José Luis Vargas; Mariela Gutiérrez; Mariela Gutiérrez Joselyn Prado; Melany Calderón; Nazareth Ruiz & Margarita Lee; Nazareth Ruiz & Margarita Lee-Lui |
| EPI_ISL_2502746 | HOSPITAL MEXICO | Inciensa, Instituto Costarricense de InvestigaciOn y Enseñanza en NutriciOn y Salud | Adriana Godínez; Claudio Soto-Garita; Estela Cordero; Francisco Duarte; Hebleen Porras; Jose Luis Vargas; Joselyn Prado & Juan Vilallobos; Mariela Gutierrez; Melany CalderOn |
| EPI_ISL_1827515, EPI_ISL_2658274 | HOSPITAL MONSEÑOR SANABRIA | Inciensa, Instituto Costarricense de Investigación y Enseñanza en Nutrición y Salud | Adriana Godínez; Claudio Soto-Garita; Estela Cordero; Francisco Duarte; Hebleen Porras; Joselyn Prado & Andrea Moreno-Carvajal; Joselyn Prado & Javier Zárate-León; José Luis Vargas; Mariela Gutiérrez; Melany Calderón |
| EPI_ISL_1261540 | HOSPITAL RAFAEL MENDEZ | Instituto de Salud Carlos III | A. Monzón; E; F. Casas; I. Jiménez; I.Cascales; M. Sandonís; P. Zaballos; S. Cuesta; S. Iglesias-Caballero; S. Pozo; S. Varona; V. Camarero; Vázquez-Morón |
| EPI_ISL_2674346 | HOSPITAL REGIONAL DE LA ORINOQUIA | Instituto Nacional de Salud- Dirección de Investigación en Salud Pública | Carlos Franco-Muñoz; Carmen Osorio; Diana Malo; Diego A. Álvarez-Díaz; Diego Andrés Prada; Gerardo Santamaría; Hector Alejandro Ruiz-Moreno; Jhonnatan Reales-González; Jorge Rivera; Juan Camilo Martinez; Julian Naizaque; Katherine Laiton-Donato; Lisseth Pardo; Magdalena Wiesner; Marcela Mercado-Reyes; Maria T. Herrera-Sepúlveda; Marta Lopez Blanco; Martha Lucia Ospina Martinez; Paola Rojas; Sergio Gomez; Sheryll Corchuelo; Ángela Alarcon Cruz |
| EPI_ISL_2502741 | HOSPITAL SAN FRANCISCO DE ASIS | Inciensa, Instituto Costarricense de InvestigaciOn y Enseñanza en NutriciOn y Salud | Adriana Godínez; Claudio Soto-Garita; Estela Cordero; Francisco Duarte; Hebleen Porras; Jose Luis Vargas; Joselyn Prado & Adrián Fallas-Mora; Mariela Gutierrez; Melany CalderOn |
| EPI_ISL_2103406, EPI_ISL_2658278 | HOSPITAL SAN FRANCISCO DE ASIS | Inciensa, Instituto Costarricense de Investigación y Enseñanza en Nutrición y Salud | Adriana Godínez; Caterina Guzmán; Claudio Soto-Garita; Estela Cordero; Francisco Duarte; Hebleen Porras; Joselyn Prado & Juan Carlos Villalobos-Ugalde; José Luis Vargas; Mariela Gutiérrez; Melany Calderón; Nazareth Ruiz & Joselyn Prado |
| EPI_ISL_2502753, EPI_ISL_2502760, EPI_ISL_2502761, | HOSPITAL SAN JUAN DE DIOS | Inciensa, Instituto Costarricense de InvestigaciOn y Enseñanza en NutriciOn y Salud | Adriana Godínez; Claudio Soto-Garita; Estela Cordero; Francisco Duarte; Hebleen Porras; Jose Luis Vargas; Joselyn Prado & Marco Cháves-Otárola; Mariela Gutierrez; Melany CalderOn |

| | | | |
|---|---|---|---|
| EPI_ISL_2502762 | | | |
| EPI_ISL_682245, EPI_ISL_2657687, EPI_ISL_2827974 | HOSPITAL SAN JUAN DE DIOS | Inciensa, Instituto Costarricense de Investigación y Enseñanza en Nutrición y Salud | Adriana Godinez & Melany Calderon; Adriana Godínez; Claudio Soto-Garita; Estela Cordero; Francisco Duarte; Hebleen Porras; Joselyn Prado & Marco Cháves-Otárola; Joselyn Prado & Paola Sánchez; José Luis Vargas; Mariela Gutiérrez; Melany Calderón |
| EPI_ISL_1510697, EPI_ISL_2331700 | HOSPITAL UNIVERSITARIO INSULAR DE GRAN CANARIA | Instituto de Salud Carlos III | A. Monzón; ARACELI; CRISTOBAL; F. Casas; I. Jiménez; I.DEL ROSARIO QUINTANA; I.HERNANDEZ BETANCOR; M. Sandonís; P. Zaballos; S. Cuesta; S. Iglesias-Caballero; S. Pozo; S. Varona; V. Camarero; Vázquez-Morón |
| EPI_ISL_682265 | HOSPITAL UPALA | Inciensa, Instituto Costarricense de Investigación y Enseñanza en Nutrición y Salud | Adriana Godinez & Melany Calderon; Claudio Soto-Garita; Estela Cordero; Francisco Duarte; Hebleen Porras |
| EPI_ISL_636975 | HP Pemba | KRISP, KZN Research Innovation and Sequencing Platform | Giandhari J; Ismael N; Nadia Sitoe; Nedio Mabunda; Paulo Arnaldo; Pillay S; Tegally H; Wilkinson E; de Oliveira T |
| EPI_ISL_412971 | HUS Diagnostiikkakeskus, Hallinto | Department of Virology Faculty of Medicine, Medicum University of Helsinki | Hannimari Kallio-Kokko; Olli Vapalahti; Suvi Kuivanen; Teemu Smura |
| EPI_ISL_2193710 | Hackensack Medical Center | New York Genome Center | Andre Corvelo; Barry Kreiswirth; David Perlin; Dayna M. Oschwald; Jose Mediavilla; Kaelea Composto; Kar Chow; Liang Chen; Marcus Cunningham; Michael Zody; Samantha Fennessey; Tom Maniatis |
| EPI_ISL_2096776, EPI_ISL_2096885 | Hadassah Medical Center Clinical Virology Laboratory, Hadassah Ein Kerem | Hadassah Hebrew University Viral Sequencing Group, Hadassah Hebrew University Medical Center | Dana G. Wolf; Esther Oiknine-Djian; Hadar Golan Berman; Mila Rivkin; Sheera Adar |
| EPI_ISL_615105 | Halmstad klinisk mikrobiologi | The Public Health Agency of Sweden | Anna Risberg; Anna-Malin Linde; Karin Tegmark-Wisell; Maria Lind Karlberg; Mattias Haukland; Mia Brytting; Olov Svartstrom; Oskar Karlsson Lindsjo; Petra Edquist; Reza Advani; Sandra Broddesson |
| EPI_ISL_2333091 | Haugesund Hospital, laboratory for Medical Microbiology | Norwegian Institute of Public Health, Department of Virology | Atiya R Ali; Debech Nadia; Engebretsen Serina Beate; Garcia Llorente Ignacio; Hilde Elshaug; Hilde Vollan; Jon Bråte; Kamilla Heddeland Instefjord; Karoline Bragstad; Kathrine Stene-Johansen; Line Victoria Moen; Marie Paulsen Madsen; Olav Hungnes; Pedersen Benedikte Nevjen; Rasmus Riis Kopperud |
| EPI_ISL_1318065, EPI_ISL_2333443 | Haukeland University Hospital, Dept. of Microbiology | Norwegian Institute of Public Health, Department of Virology | 'Kathrine Stene-Johansen; Atiya R Ali; Debech Nadia; Engebretsen Serina Beate; Garcia Llorente Ignacio; Hilde Elshaug; Hilde Vollan; Jon Bråte; Kamilla Heddeland Instefjord; Karoline Bragstad; Kathrine Stene-Johansen; Line Victoria Moen; Marie Paulsen Madsen; Olav Hungnes; Pedersen Benedikte Nevjen; Rasmus Riis Kopperud |
| EPI_ISL_2938119 | Health Center Novi Grad Sarajevo | Clinical Center, University of Sarajevo; Unit for Clinical Microbiology | Amela Dedeić-Ljubović; Edina Zahirović; Irma Salimović-Bešić; Sandra Vegar-Zubović; Sebija Izetbegović |
| EPI_ISL_2938097 | Health Center Velika Kladuša | Clinical Center, University of Sarajevo; Unit for Clinical Microbiology | Amela Dedeić-Ljubović; Edina Zahirović; Irma Salimović-Bešić; Sandra Vegar-Zubović; Sebija Izetbegović |
| EPI_ISL_2080427, EPI_ISL_2469286 | Health Services Laboratories | Wellcome Sanger Institute for the COVID-19 Genomics UK (COG-UK) Consortium | Cordelia Langford; David K. Jackson; Dominic Kwiatkowski; Ewan Harrison; Health Services Laboratories and Alex Alderton; Ian Johnston; Jeffrey Barrett; John Sillitoe on behalf of the Wellcome Sanger Institute COVID-19 Surveillance Team; Roberto Amato; Sonia Goncalves |
| EPI_ISL_2004109, EPI_ISL_2004111, EPI_ISL_2689834, EPI_ISL_2712906 | Health sciences Department, Universidad Técnica Particular de Loja | Institute of Microbiology, Universidad San Francisco de Quito | Belén Prado-Vivar; Bernardo Gutiérrez; Danilo Sosa; Fernanda Zurita; Fernando Serrano; Gabriel Trueba; Juan José Guadalupe; Katty Ojeda; Michelle Grunauer; Monica Becerra-Wong; Paola Dalgo; Patricio Rojas-Silva; Paúl Cárdenas; Rita Rodriguez; Rommel Guevara; Sully Márquez; Verónica Barragán |
| EPI_ISL_796015 | Hebei Provincial Center for Disease Control and Prevention, Shijiazhuang, Hebei Province; National Institute for Viral Disease Control and Prevention, China CDC | Hebei Provincial Center for Disease Control and Prevention, Shijiazhuang, Hebei Province; National Institute for Viral Disease Control and Prevention, China CDC | George F. Gao; Nankun Liu; Qi Li; Shunxiang Qi; Wenbo Xu; Xiang Zhao; Yang Song |
| EPI_ISL_640018 | Heidelberg Clinic wc HBC | NHLS/UCT | Arash Iranzadeh; Bruna Galvao; Carolyn Williamson; Deelan Doolabh; Diana Hardie; Innocent Mudau; Kruger Marais; Lynn Tyers; Marvin Hsiao; Stephen Korsman |
| EPI_ISL_700598, EPI_ISL_2877001 | Heideveld Emergency Centre | NHLS/UCT | Arash Iranzadeh; Bruna Galvao; Carolyn Williamson; Deelan Doolabh; Diana Hardie; Gert Marais; Innocent Mudau; Kruger Marais; Lynn Tyers; Marvin Hsiao; Stephen Korsman |
| EPI_ISL_1088823 | Helix / Illumina | Respiratory Viruses Branch, Division of Viral Diseases, Centers for Disease Control and Prevention | Alexandre Bolze; Ary Ascencio; Ben L. Rambo-Martin; Brad Sickler; Charlotte Rivera-Garcia; Christine Tran; Clinton R. Paden; Dakota Howard; David Becker; Dhwani Batra; Duncan MacCannell; Efren Sandoval; Eileen de Feo; Elizabeth Cirulli; Eric Allen; Geraint Levan; James Lu; Jan Antico; Jason Nguyen; Jimmy Ramirez; Jingtao Liu; Kelly Schiabor Barrett; Kim Gietzen; Magnus Isaksson; Marc Laurent; Matthew Tolentino; Nicole L. Washington; Peter W. Cook; Phil Febbo; Ryan Cho; Shannon Wickline; Sherry Wang; Simon White; Summer Galloway; Suxiang Tong; Tyler Cassens; William Lee |
| EPI_ISL_1269377, EPI_ISL_1270167, EPI_ISL_1443915, EPI_ISL_1924266, EPI_ISL_2159579, EPI_ISL_2268352, EPI_ISL_2269936, EPI_ISL_2269998, EPI_ISL_2320596, EPI_ISL_2598842 | | | |
| see above | Helix/Illumina | Centers for Disease Control and Prevention Division of Viral Diseases, Pathogen Discovery | Adrian Paskey; Alexandre Bolze; Ary Ascencio; Ben L. Rambo-Martin; Benjamin Rambo-Martin; Brad Sickler; Charlotte Rivera-Garcia; Christine Tran; Christopher Gulvick; Clinton R. Paden; Dakota Howard; Darlene Wagner; David Becker; Dhwani Batra; Duncan MacCannell; Efren Sandoval; Eileen de Feo; Elizabeth Cirulli; Eric Allen; Geraint Levan; James Lu; Jan Antico; Jason Caravas; Jason Nguyen; Jimmy Ramirez; Jingtao Liu; Kara Moser; Kelly Schiabor Barrett; Kim Gietzen; Magnus Isaksson; Marc Laurent; Matthew Schmerer; Matthew Tolentino; Nicole L. Washington; Peter W. Cook; Phil Febbo; Ryan Cho; Scott Sammons; Shannon Wickline; Shatavia Morrison; Sherry Wang; Simon White; Summer Galloway; Suxiang Tong; Tyler Cassens; William Lee; Yvette Unoarumhi |
| EPI_ISL_1036688, EPI_ISL_1036697 | Helix/Illumina | Respiratory Viruses Branch, Division of Viral Diseases, Centers for Disease Control and Prevention | Alexandre Bolze; Ary Ascencio; Ben L. Rambo-Martin; Brad Sickler; Charlotte Rivera-Garcia; Christine Tran; Clinton R. Paden; Dakota Howard; David Becker; Dhwani Batra; Duncan MacCannell; Efren Sandoval; Eileen de Feo; Elizabeth Cirulli; Eric Allen; Geraint Levan; James Lu; Jan Antico; Jason Nguyen; Jimmy Ramirez; Jingtao Liu; Kelly Schiabor Barrett; Kim Gietzen; Magnus Isaksson; Marc Laurent; Matthew Tolentino; Nicole L. Washington; Peter W. Cook; Phil Febbo; Ryan Cho; Shannon Wickline; Sherry Wang; Simon White; Summer Galloway; Suxiang Tong; Tyler Cassens; William Lee |
| EPI_ISL_2367588 | Hellenic National Blood Transfusion Center - EKEA | Greek Genome Center, Biomedical Research Foundation of the Academy of Athens (BRFAA) | Dimitrios Thanos; Efthimia Petinaki; Emmanouil Athanasiadis; Giannis Vatsellas; Katerina Zoi; Kostas Stamoulis; Theodoros Loupis |
| EPI_ISL_2617410 | Hellenic National Blood Transfusion Center - EKEA | Greek Genome Center, Biomedical Research Foundation of the Academy of Athens (BRFAA) | Dimitrios Thanos; Efthimia Petinaki; Emmanouil Athanasiadis; Giannis Vatsellas; Katerina Zoi; Kostas Stamoulis; Theodoros Loupis |
| EPI_ISL_501235 | Hellenic Pasteur Institute, National Influenza Reference laboratory of Southern Greece & Unit of Bioinformatics and Applied Genomics | Hellenic Pasteur Institute, National Influenza Reference laboratory of Southern Greece & Unit of Bioinformatics and Applied Genomics | Andreas Mentis; Androniki Voulgari-Kokota; Antonios Kalliaropoulos; Aspasia Kontou; Athanasios Kossyvakis; Evangelidou Maria; Horefti Elina; Timokratis Karamitros; Vasiliki Pogka |
| EPI_ISL_430469 | Hellenic Pasteur Institute, Public Health Laboratories | Hellenic Pasteur Institute, Public Health Laboratories, Unit of Bioinformatics and Applied Genomics | Andreas Mentis; Androniki Voulgari-Kokota; Antonios Kalliaropoulos; Aspasia Kontou; Athanasios Kossyvakis; Evangelidou Maria; Horefti Elina; Timokratis Karamitros; Vasiliki Pogka |
| EPI_ISL_450529 | Hematology Laboratory, Section of Molecular Diagnostics, University Clinical Centre, Medical University of Gdansk | Department of Virology, Faculty of Medicine, University of Helsinki, Helsinki, Finland | Aneta Szulc; Maciej Grzybek; Marlena Robakowska; Olli Vapalahti; Teemu Smura |
| EPI_ISL_451654, EPI_ISL_906752 | Hematology Laboratory, Section of Molecular Diagnostics, University Clinical Centre, Medical University of Gdansk | Laboratory of Recombinant Vaccines | Adam Sodol; Aneta Szulc; Boguslaw Szewczyk; Ewa Milosz; Krystyna Bienkowska-Szewczyk; Krzysztof Lewandowski; Lukasz Rabalski; Maciej Grzybek; Maciej Kosinski; Marlena Robakowska |
| EPI_ISL_700262 | Hematopathology Laboratory, ACTREC, TMC | Hematopathology Laboratory, ACTREC, TMC | ACTREC; Hematopathology Laboratory |
| EPI_ISL_2140721 | Hillside Clinic wc BFW | NHLS/UCT | Arash Iranzadeh; Bruna Galvao; Carolyn Williamson; Deelan Doolabh; Diana Hardie; Innocent Mudau; Kruger Marais; Lynn Tyers; Marvin Hsiao; Stephen Korsman |
| EPI_ISL_2812851, EPI_ISL_2828084, EPI_ISL_3015123 | Histopath | NSW Health Pathology - Institute of Clinical Pathology and Medical Research; Westmead Hospital; University of Sydney | CIDM-PH et al. |
| EPI_ISL_2779330, EPI_ISL_2779346 | Homabay County Referral Hospital | USAMRD-A, Basic Science Laboratory | Alan Lemtudo; Beth Mutai; Brian Andika; Carol Kifude; Clement Masakwe; Eric Muthanje; Esther Omuseni; Faith Sigei; Gathii Kimita; George Awinda; John Waitumbi; Josphat Nyataya; Rachel Githii; Rehema Liyai; Stephen Ochola |
| EPI_ISL_1257882 | Hopital Général de Douala | Institut Pasteur de Dakar | Dia Ndongo; Diagne Moussa Moïse; Diallo Amadou; Diop Mamadou; Elisabeth Carniel; Faye Ousmane; Loucoubar Cheikh; Njouom Richard; Sall Amadou Alpha; Sankhe Safietou |
| EPI_ISL_1257892 | Hopital Laquintinie de Douala | Institut Pasteur de Dakar | Dia Ndongo; Diagne Moussa Moïse; Diallo Amadou; Diop Mamadou; Elisabeth Carniel; Faye Ousmane; Loucoubar Cheikh; Njouom Richard; Sall Amadou Alpha; Sankhe Safietou |
| EPI_ISL_1821607, EPI_ISL_2029952, EPI_ISL_2029977, EPI_ISL_2188319, EPI_ISL_2188320, EPI_ISL_2293517, EPI_ISL_2374409, EPI_ISL_2462905, EPI_ISL_2462927, EPI_ISL_2464242, EPI_ISL_2628279, EPI_ISL_2757663, EPI_ISL_2757673, EPI_ISL_2757678, EPI_ISL_2790847, EPI_ISL_2835879, EPI_ISL_2835906, EPI_ISL_2835907, EPI_ISL_3031652, EPI_ISL_3031654, EPI_ISL_3031655 | | | |
| see above | Hospital | National Reference Center for Viruses of Respiratory Infections, Institut Pasteur, Paris | Alexandra Ducancelle; Angela Brisebarre; Antoine Talarmin; Camille Capel; Ch De Cornouaille; Christophe Malabat; CléMence Guillaume; Corinne Maufrais; Damien Mornico; Didier Mattera; Emmanuelle Permal; Etienne Simon-Lorière; Frédéric Lemoine; Hub de Bioinformatique et Biostatistique; JéRôMe Durivault; L. Courdavault; Louise Lefrançois; LéA Pilorge; Marianne Coste-Burel; Marianne Burgard; Marion Barbet; Maud Vanpeene; Méline Bizard; Patricia Barbut; Sylvaine Bastian; Sylvie Behillil; Sylvie Van der Werf; Victoire Baillet; Vincent Enouf |
| EPI_ISL_1917304 | Hospital Center Emile Mayrisch | Laboratoire national de sante, Microbiology, Microbial Genomics Platform | Anke Wienecke-Baldacchino; Catherine Ragimbeau; Cynthia Oxacelay; Fatu Djabi; Jessica Tapp; Lise Pignon; Raoul Salmon; Tamir Abdelrahman |
| EPI_ISL_770028 | Hospital Dr. Raul Blanco Cervantes | Inciensa, Instituto Costarricense de Investigación y Enseñanza en Nutrición y Salud | Adriana Godínez; Claudio Soto-Garita; Estela Cordero; Francisco Duarte; Hebleen Porras; Melany Calderón & Mariel López |
| EPI_ISL_2965560, EPI_ISL_2965569, EPI_ISL_2965583, EPI_ISL_2965584 | Hospital Español | Centro de Innovación en Vigilancia Epidemiológica (CiVE), Institut Pasteur Montevideo, Uruguay | Alicia Costábile; Alvaro Fajardo; Ana Moller; Andrés Lizasoain; Belén González; Bernardina Rivera; Cecilia Alonso; Cecilia Salazar; Gonzalo Bello; Gonzalo Moratorio; Gregorio Iraola; Henry Albornoz; Ignacio Ferrés; Javier Hurtado; Juan Zanetti; Julio Medina; Luciana Griffero; Lucía Spangenberg; Ma Noel Bentancor; Ma Pía Techera; Mailen Arleo; Martina Alonso; Matías Maidana; Mauricio Méndez; Melissa Duquía; Mercedes Paz; Natalia Rego; Natalia Reyes; Nicolas Nin; Odhille Chappos; Paula Perbolianachis; Pilar Moreno; Rodney Colina; Rodrigo Arce; Tamara Fernández-Calero; Tania Possi; Veronica Noya; Viviana Bortagaray |
| EPI_ISL_1647957, EPI_ISL_2483973 | Hospital General Universitario Gregorio Marañón | Hospital General Universitario Gregorio Marañón | Cristina Rodriguez-Grande; Darío García de Viedma; Julia Suárez; Laura Pérez-Lago; Marta Herranz Martin; Patricia Muñoz; Pedro Sola Campoy; Pilar Catalán; Sergio Buenestado Serrano; Victor Manuel de la Cueva |
| EPI_ISL_467234, EPI_ISL_541903, EPI_ISL_780044, EPI_ISL_780047 | Hospital General Universitario Gregorio Marañón | SeqCOVID-SPAIN consortium/IBV(CSIC) | Darío García de Viedma; Darío García de Viedma and SeqCOVID-SPAIN consortium; Jon Sicilia; Julia Suárez; Laura Pérez-Lago; Marta Herranz; Patricia Muñoz; Patricia Muñoz and SeqCOVID-SPAIN consortium; Pilar Catalán |
| EPI_ISL_2014355 | Hospital Herrera Llerandi | Asociación de Salud Integral/Clínica Familiar Luis Ángel García | Ana S. Gonzalez-Reiche; Claudia Rangel; Danicela Mercado; Eduardo Arathoon; Hilda Ruiz; Luis Aguirre; Luis Rivas; Narda Medina; Oscar Bonilla; Osmar Gamboa. |
| EPI_ISL_769992 | Hospital Metropolitano | Inciensa, Instituto Costarricense de Investigación y Enseñanza en Nutrición y Salud | Adriana Godínez; Claudio Soto-Garita; Estela Cordero; Francisco Duarte; Hebleen Porras; Melany Calderón & Margarita Lee Lui |
| EPI_ISL_480327 | Hospital Nacional de Niños | Charité Virology-University of Costa Rica | Andrei Montero Bonilla; Andres Moreira-Soto; Cristian Pérez Corrales; Eugenia Corrales-Aguilar; Ignacio Postigo-Hidalgo; Jan Felix Drexler |

| | | | |
|---|---|---|---|
| EPI_ISL_471270 | Hospital Oncológico Solca Núcleo de Quito | Institute of Microbiology, Universidad San Francisco de Quito | Belén Prado-Vivar; Bernardo Gutiérrez; Gabriel Trueba; Grace Salazar; Juan José Guadalupe; Marcos Di Stefano; Michelle Grunauer; Patricio Rojas-Silva; Paúl Cárdenas; Sully Márquez; Verónica Barragán |
| EPI_ISL_524463 | Hospital Regional de Cotia | Instituto Adolfo Lutz, Interdiciplinary Procedures Center, Strategic Laboratory | Claudia Regina Gonçalves; Claudio Tavares Sacchi; Erica Valessa Ramos Gomes |
| EPI_ISL_491452 | Hospital San Rafael de Alajuela | Inciensa, Instituto Costarricense de Investigación y Enseñanza en Nutrición y Salud | Adriana Godinez & Melany Calderon; Claudio Soto-Garita; Estela Cordero; Francisco Duarte; Hebleen Brenes |
| EPI_ISL_2982423 | Hospital Universitari Arnau de Vilanova | Hospital Universitari Vall d'Hebron - Vall d'Hebron Institut de Recerca | Alejandra González-Sánchez; Andrés Antón; Ariadna Rando; Carla Castillo; Cristina Andrés; Damir Garcia-Cehic; Josep Quer; Juliana Esperalba; Karen García; Maria Carmen Martin; Maria Gema Codina; Maria Piñana; Rodrigo Vásquez; Tomàs Pumarola |
| EPI_ISL_2516574 | Hospital Universitari Bellvitge | Microbiology Department | Aida Gonzalez-Diaz; Carmen Ardanuy; Jordi Camara; Jordi Niubó; Laura Calatayud; M Angeles Domínguez; Miguel Fernandez-Huerta; Sara Marti |
| EPI_ISL_3009987 | Hospital Universitari Vall d'Hebron - Vall d'Hebron Institut de Recerca | Hospital Universitari Vall d'Hebron - Vall d'Hebron Institut de Recerca | Alejandra González-Sánchez; Andrés Antón; Ariadna Rando; Carla Castillo; Cristina Andrés; Damir Garcia-Cehic; Josep Quer; Juliana Esperalba; Karen García; Maria Carmen Martin; Maria Gema Codina; Maria Piñana; Rodrigo Vásquez; Tomàs Pumarola |
| EPI_ISL_691687 | Hospital Universitario de Ceuta | Instituto de Salud Carlos III | A. Monzón; F. Casas; I. Hijano, S.; I. Jiménez; Iglesias-Caballero; M. Camarero; M. Cuesta; M. González-Esguevillas; M. Pozo; M. Zaballos; P. Jiménez; S. Juliá; S. Molinero Calamita; S. Varona |
| EPI_ISL_1470312 | Hospital Universitario de Gran Canaria Dr. Negrín | SeqCOVID-SPAIN consortium/IBV(CSIC) | Ana Bordes Benítez and SeqCOVID-SPAIN consortium; Francisco J. Chamizo López; M. Carmen Pérez González |
| EPI_ISL_455314 | Hospital Virgen del Rocio | Instituto de Salud Carlos III | A. Monzón; F. Casas; I; I. Jiménez; Iglesias-Caballero; J. Lepe; M. Camarero; M. Cuesta; M. González-Esguevillas; M. Molinero Calamita; M. Zaballos; P. Jiménez; S. Juliá; S. Pozo; S. Varona |
| EPI_ISL_671815 | Hospital de la Santa Creu i Sant Pau. Servicio de Microbiología | SeqCOVID-SPAIN consortium/IBV(CSIC) | Elisenda Miró and SeqCOVID-SPAIN consortium; Ferran Navarro; Núria Rabella |
| EPI_ISL_458028 | Hospital for Tropical Diseases | COVID-19 Network Investigations (CONI) Alliance | Akanitt Jittmittraphap; Angkana Huang; Anthony R. Jones; Arporn Wangwiwatsin; Bhakbhoom Panthan; Chonticha Klungtong; Ekawat Pasomsub; Elizabeth Batty; Insee Sensorn; Janjira Thaipadungpanit; Khajohn Joonsalak; Kingkan Rakmanee; Krittikorn Kumpornsin; Namfon Kotanan; Nantarat Chantawat; Pornsawan Leaungwutiwong; Stefan Fernandez; Thanat Chookajorn; Theerarat Kochakarn; Treewat Watthanachockchai; Wasun Chantratita; Weena Janwitthayanan; Wudtichai Manasatienkij |
| EPI_ISL_2788566, EPI_ISL_2788595 | Hospital of Southern Norway - Kristiansand, Department of Medical Microbiology | Norwegian Institute of Public Health, Department of Virology | Atiya R Ali; Debech Nadia; Engebretsen Serina Beate; Garcia Llorente Ignacio; Hilde Elshaug; Hilde Vollan; Jon Bråte; Kamilla Heddeland Instefjord; Karoline Bragstad; Kathrine Stene-Johansen; Line Victoria Moen; Marie Paulsen Madsen; Olav Hungnes; Pedersen Benedikte Nevjen; Rasmus Riis Kopperud |
| EPI_ISL_1077280, EPI_ISL_2219818 | Houston Methodist Hospital | Houston Methodist Hospital | Ilya J. Finkelstein; James J. Davis; Jessica Cambric; Jimmy Gollihar; Kristina Reppond; Layne Pruitt; Madison N. Shyer; Marcus Nguyen; Matthew Ojeda Saavedra; Paul A. Christensen; Prasanti Yerramilli; Randall J. Olsen; Robert Olson; Ryan Gadd; S. Wesley Long; Sishir Subedi; and James M. Musser |
| EPI_ISL_475525 | Huddinge VC | The Public Health Agency of Sweden | Anna Risberg; Anna-Malin Linde; Karin Tegmark-Wisell; Maria Lind Karlberg; Mattias Haukland; Mia Brytting; Olov Svartstrom; Oskar Karlsson Lindsjo; Reza Advani; Sandra Broddesson |
| EPI_ISL_2466657 | Human Genetic Research Center, Kawsar Biotech Company | Kawsar Biotech Company | Abbasalipour, M.; Azadmanesh, K.; Chamran Hospital, C.; Khosravi; M.A.; National Institute for Medical Research Development, N.; Zeinali, R.; Zeinali, S. |
| EPI_ISL_645082 | Human Genome Variation Research Group, Malopolska Centre of Biotechnology | Human Genome Variation Research Group, Malopolska Centre of Biotechnology | Botwina, P.; Branicki, W.; Dabrowska, A.; Foremny, J.; Gromowski, T.; Klajmon, A.; Kopera, K.; Kowalski, M.; Labaj; Marszalek, K.; Owczarek, K.; P.P.; Pisarek, A.; Pospiech, E.; Pyrc, K.; Sanak, M.; Swadzba, J.; Szczepanski, A. |
| EPI_ISL_526224, EPI_ISL_526225, EPI_ISL_1041201 | Hungarian Defence Forces Military Medical Centre | National Laboratory of Virology, Szentágothai Research Centre | Balázs Somogyi; Bálint Eszenyi; Csaba Pereszlényi; Endre Gábor Tóth; Ferenc Jakab; Gábor Kemenesi; Ágnes Balázs-Nagy |
| EPI_ISL_1296394 | Hôpital Bichat Claude Bernard, Laboratoire de Virologie | IAME UMR1137 Inserm, Université de Paris, Hôpital Bichat | Alexandre Storto; Amélie Recoing; Antoine Bridier-Nahmias; Benoit Visseaux; Charlotte Charpentier; Diane Descamps; Gilles Collin; Lena Daniel; Mélanie Bertine; Nadhira Houhou-Fidouh; Quentin Le Hingrat; Siham Hamri |
| EPI_ISL_2637514 | Hôpital Cochin | Department of Virology, Henri Mondor University Hospital, Assistance Publique Hôpitaux de Paris, Université Paris-Est Créteil, INSERM U955 | Alexandre Soulier; Christophe Rodriguez; Elisabeth Trawinski; Guillaume Gricourt; Jean-Michel Pawlotsky; Melissa N'Debi; Slim Fourati; Vanessa Demontant |
| EPI_ISL_710532 | Hôpital Fattouma-Bourguiba de Monastir | Laboratoire des Procédés de Criblage Moléculaire et Cellulaire-Centre de Biotechnologie de Sfax | A. and Masmoudi, S.; Abdelmoulah, F.; Abid, N.; Ajili, F.; Aouni, M.; Ben Ayed, I.; Bensaid, M.; Chtourou, A.; Elargoubi, A.; Fki-berrajah, L.; Gaaloul, I.; Gargouri, S.; Hammami, A.; Kamoun, S.; Karray Hakim, H.; Kharat, N.; Mastouri, M.; Mhalla, S.; Nabli, A.; Rebai; Smeti, I.; Souissi, A.; Stambouli, N.; Turki, M. |
| EPI_ISL_961579 | Hôpital Georges L. Dumont | National Microbiology Laboratory (NML) | Anna Majer; Anneliese Landgraff; CanCOGeN's metadata curation team; Darian Hole; Elsie Grudeski; Gary Van Domselaar; Grace Seo; Guillaume Desnoyers; Jennifer Tanner; Kirsten Biggar; Madison Chapel; Morag Graham; Natalie Knox; Nathalie Bastien; Philip Mabon; Public Health Agency of Canada CanCOGeN team; Rhiannon Huzarewich; Richard Garceau; Russell Mandes; Shari Tyson; Timothy Booth; Yan Li |
| EPI_ISL_535716 | Hôpital de Verdun | Laboratoire de santé publique du Québec | Guillaume Bourque; Ioannis Ragoussis; Jesse Shapiro; Mark Lathrop and Michel Roger on behalf of the CoVSeQ research group; Sandrine Moreira |
| EPI_ISL_2384050, EPI_ISL_2384052 | IA State Hygienic Laboratory | Centers for Disease Control and Prevention Division of Viral Diseases, Pathogen Discovery | Alison Laufer Halpin; Ben L. Rambo-Martin; Clinton R. Paden; Dakota Howard; Darlene Wagner; Dave Wentworth; Dhwani Batra; Jasmine Padilla; Justin Lee; Katie Dillon; Krista Queen; Kristen Knipe; Kristine Lacek; Mark Burroughs; Matthew Schmerer; Mili Sheth; Peter Cook; Sam Shepard; Sarah Nobles; Shoshona Le; Suxiang Tong; Vivien Dugan; Yvette Unoarumhi |
| EPI_ISL_2840309 | ID Bureau of Laboratories | Centers for Disease Control and Prevention Division of Viral Diseases, Pathogen Discovery | Alison Laufer Halpin; Ben L. Rambo-Martin; Clinton R. Paden; Dakota Howard; Darlene Wagner; Dave Wentworth; Dhwani Batra; Jasmine Padilla; Justin Lee; Katie Dillon; Krista Queen; Kristen Knipe; Kristine Lacek; Mark Burroughs; Matthew Schmerer; Mili Sheth; Peter Cook; Sam Shepard; Sarah Nobles; Shoshona Le; Suxiang Tong; Vivien Dugan; Yvette Unoarumhi |
| EPI_ISL_903753 | ID Bureau of Laboratories | Genomics and Discovery, Respiratory Viruses Branch, Division of Viral Diseases, Centers for Disease Control and Prevention | Anna Montmayeur; Anna Uehara; Ben L. Rambo-Martin; Clinton R. Paden; Dhwani Batra; Haibin Wang; Jasmine Padilla; Jing Zhang; Justin Lee; Krista Queen; Lori Rowe; Mark Burroughs; Mili Sheth; Peter W. Cook; Rachel Marine; Sarah Nobles; Suxiang Tong; Yan Li; Ying Tao |
| EPI_ISL_2234876, EPI_ISL_2444806, EPI_ISL_2444813, EPI_ISL_2444814, EPI_ISL_2444815, EPI_ISL_2444817, EPI_ISL_2444819, EPI_ISL_2444820, EPI_ISL_2444821, EPI_ISL_2444822, EPI_ISL_2444823, EPI_ISL_2444825, EPI_ISL_2444827, EPI_ISL_2444829, EPI_ISL_2444830, EPI_ISL_2444834, EPI_ISL_2444836, EPI_ISL_2444838, EPI_ISL_2444839 | | | |
| see above | IICS-UNA | IICS-UNA | Adriana Valenzuela; Alejandra Rojas; Chyntia Diaz; Eva Nara; Fatima Cardozo; Florencia del Puerto; Joel Ortiz; Jonas Fernandez; Laura Franco; Laura Mendoza; Leticia Rojas; Magaly Martinez; Maria Eugenia Galeano. |
| EPI_ISL_2758180 | ILV Kärnten | Bergthaler laboratory, CeMM Research Center for Molecular Medicine of the Austrian Academy of Sciences | Andreas Bergthaler; Anna Schedl; Bekir Erguner; Benedikt Agerer; Christoph Bock; Fabian Amman; Jan Laine; Lukas Endler; Maelle Le Moing; Martin Senekowitsch; Matthew Thornton; Michael Schuster; Petr Triska; Thomas Penz |
| EPI_ISL_2827770, EPI_ISL_2827774, EPI_ISL_2827780, EPI_ISL_2827784 | IMAT S.A.S. | Instituto Nacional de Salud- Dirección de Investigación en Salud Pública | Carlos Franco-Muñoz; Carmen Osorio; Diana Malo; Diego A. Álvarez-Díaz; Diego Andrés Prada; Gerardo Santamaría; Hector Alejandro Ruiz-Moreno; Jhonnatan Reales-González; Jorge Rivera; Juan Camilo Martinez; Julian Naizaque; Katherine Laiton-Donato; Lisseth Pardo; Magdalena Wiesner; Marcela Mercado-Reyes; Maria T. Herrera-Sepúlveda; Marta Lopez Blanco; Martha Lucia Ospina Martinez; Paola Rojas; Sergio Gomez; Sheryll Corchuelo; Ángela Alarcon Cruz |
| EPI_ISL_1439879, EPI_ISL_2845684 | IMD - Labor Oderland | Robert Koch Institute | |
| EPI_ISL_1722245 | IMD - Medizinisches Labor Rostock | Robert Koch Institute | |
| EPI_ISL_1721083 | IMD Labor Frankfurt | Robert Koch Institute | |
| EPI_ISL_2657686 | INCIENSA | Inciensa, Instituto Costarricense de Investigación y Enseñanza en Nutrición y Salud | Adriana Godínez; Claudio Soto-Garita; Estela Cordero; Francisco Duarte; Hebleen Porras; Joselyn Prado & Grettel Chanto-Chacón; José Luis Vargas; Mariela Gutiérrez; Melany Calderón |
| EPI_ISL_2319343 | INDRE | Instituto Nacional de Medicina Genomica | Cedro-Tanda A; Garcia-Cardenas FJ; Gisela Barrera-Badillo; Gonzalez-Barrera D; Gonzalez-Woge MA; Herrera-Montalvo LA.; Hidalgo-Miranda A; Irma Lopez-Martinez; Jose Ernesto Ramirez González; Mendoza-Vargas A; Munguia-Garza P; Rangel-DeLeon D; Reyes-Grajeda JP |
| EPI_ISL_1577752, EPI_ISL_1577815, EPI_ISL_2628208, EPI_ISL_2628297, EPI_ISL_2628307 | INHRR | Laboratorio de Virología Molecular | Aguilar M; Alarcon V; Carmen L Loureiro; CoViVen Group; D Angelo P; Delgado M; Domingo J Garzaro; Esmeralda Vizzi; Flor H Pujol; Garzaro D; Héctor R Rangel; Jaspe RC; José Luis Zambrano; Lieska Rodríguez; Loureiro CL; Mariana Hidalgo; Pierina D´Angelo; Pujol FH; Rangel HR; Rodriguez L; Rossana C Jaspe; Víctor Alarcón; Yoneira Sulbaran; Zambrano JL; Zoila Moros |
| EPI_ISL_2878585, EPI_ISL_2881318 | INSACOG Surveillance | INSACOG at CSIR Institute of Genomics and Integrative Biology | INSACOG |
| EPI_ISL_2812628 | INSACOG-MANIPUR | National Institute of Biomedical Genomics – INSACOG | Arindam Maitra; Khumanthem Ranjana; Nanaocha Sharma; Nidhan Kumar Biswas; R K Manglem Singh; Saumitra Das; Sreedhar Chinnaswamy |
| EPI_ISL_3019053 | INSACOG-Sikkim | National Institute of Biomedical Genomics – INSACOG | Arindam Maitra; Kaden Zangmu Bhutia; Nidhan Kumar Biswas; Saumitra Das; Sreedhar Chinnaswamy Tshering Ongmu Bhutia; Tara D Sharma |
| EPI_ISL_2503286, EPI_ISL_2503354, EPI_ISL_2548404, EPI_ISL_2676269, EPI_ISL_3019399 | INSACOG-WB | National Institute of Biomedical Genomics – INSACOG | Ajay Chakraborti; Arindam Maitra; Bhaswati Bandyopadhyay; Nidhan Kumar Biswas; Saumitra Das; Sreedhar Chinnaswamy; Tamal Ghosh |
| EPI_ISL_2672210, EPI_ISL_2672216, EPI_ISL_2672330, EPI_ISL_2672993 | INSP, Bamako | Malaria Research and Training Center-Parasito | Abdoulaye Djimde; Amadou Daou; Antoine Dara; Demba Koita; Ibrehima Guindo |
| EPI_ISL_2484167 | INSP-Bamako | Malaria Research and Training Center | Abdoulaye Djimde; Amadou Daou; Antoine Dara; Demba Koita; Ibrehima Guindo |
| EPI_ISL_826816, EPI_ISL_826825, EPI_ISL_826830, EPI_ISL_826834 | INSPI-CRN DE INFLUENZA Y OTROS VIRUS RESPIRATORIOS | Instituto de Salud Publica de Chile | Alfredo Bruno; Andres Castillo; Barbara Parra; Domenica de Mora; Gisselle Barra; Jaime Lagos; Javier Tognarelli; Jimmy Garcez; Jorge Fernandez; Loredana Arata; Manuel Gonzalez; Martiza Olmedo; Michelle Paez; Patricia Bustos; Rodrigo Fasce; Solon Narvaez |
| EPI_ISL_2488721, EPI_ISL_2488741, EPI_ISL_2488743, EPI_ISL_2757586, EPI_ISL_2757591, EPI_ISL_2757595, EPI_ISL_2757601, EPI_ISL_2757646, EPI_ISL_2893553, EPI_ISL_2893554, EPI_ISL_2893592, EPI_ISL_2893605, EPI_ISL_2893608, EPI_ISL_2893620, EPI_ISL_2893664, EPI_ISL_2897542, EPI_ISL_2897546, EPI_ISL_2988508, EPI_ISL_2988528, EPI_ISL_2988546, EPI_ISL_2988550, EPI_ISL_2988562, EPI_ISL_3010424, EPI_ISL_3010431, EPI_ISL_3010445 | | | |
| see above | INSPI-CRN DE INFLUENZA Y OTROS VIRUS RESPIRATORIOS | NIC-INSPI | Alfredo Bruno; Domenica de Mora.; Jimmy Garcés; Johanna Laines; Lizbeth Patiño; Manuel Gonzalez; Maritza Olmedo; Michelle Páez |
| EPI_ISL_940775 | INSPI-CRN de Influenza y otros virus respiratorios | INSPI-Centro de Investigación Multidisciplinaria de la DTIDI | Alberto Orlando.; Alfredo Bruno; Andrés Carrazco-Montalvo; Doménica de Mora; Leandro Patiño; Manuel González; Maritza Olmedo; Mary Regato-Arrata; Melissa Zambrano; Orson Mestanza |
| EPI_ISL_1630269 | IRESSEF | Abbott | Adbou Padane; Ambroise Ahouidi; Aminata Dia; Aminata Mboup; Ana Olivo; Anna julienne selbe Ndiaye; Barbara Harris; Cyrille Diedhiou; Gavin Cloherty; Mary Rodgers; Moustapha Mbow; Nafissatou Leye; Ndeye Diabou Diagne; Papa Alassane Diaw; Souleymane Mboup; Todd Meyer |
| EPI_ISL_2015097, | IU-Cerrahpasa, Cerrahpasa School of Medicine, COVID-19 Lab | IU-Cerrahpasa, Cerrahpasa School of Medicine, COVID-19 Lab | Kenan Midilli; Mert Kuskucu; Yesim Tuyji Tok |

| | | | |
|---|---|---|---|
| EPI_ISL_2015111 | | | |
| EPI_ISL_2628209, EPI_ISL_2628302, EPI_ISL_2628303, EPI_ISL_2628304, EPI_ISL_2628305, EPI_ISL_2628306 | IVIC | Laboratorio de Virología Molecular | Carmen L Loureiro; CoViVen Group; Domingo J Garzaro; Esmeralda Vizzi; Flor H Pujol; Héctor R Rangel; José Luis Zambrano; Lieska Rodríguez; Mariana Hidalgo; Pierina D´Angelo; Rossana C Jaspe; Víctor Alarcón; Yoneira Sulbaran; Zoila Moros |
| EPI_ISL_738121 | IZSM-U.O.C. Virologia | Istituto Zooprofilattico Sperimentale del Mezzogiorno | Giovanna Fusco; Lorena Cardillo; Maurizio Viscardi |
| EPI_ISL_2350569 | Ibaraki Prefectural Institute of Public Health | Ibaraki Prefectural Institute of Public Health | Keiko Goto; Kentaro Itokawa; Makoto Kuroda; Masanori Hashino; Rina Tanaka; Tsuyoshi Sekizuka |
| EPI_ISL_2834665, EPI_ISL_2934986 | Idaho Bureau of Laboratories | Center for Global Health, University of New Mexico Health Sciences Center | Christopher Ball; Darrell Dinwiddie; Daryl Domman; Kurt Schwalm; Matthew Burns; Robert Voermans; Valerie Morley |
| EPI_ISL_1323469 | Illinois Department of Public Health | Gagnon Lab, Southern Illinois University | Keith Gagnon |
| EPI_ISL_1018095, EPI_ISL_2001061, EPI_ISL_2001063, EPI_ISL_2001093, EPI_ISL_2508380 | Immunology, Noguchi Memorial Institute for Medical Research | Immunology, Noguchi Memorial Institute for Medical Research | A.W.; Adu, B.; Adusei-Poku; Agbenyo; Agbodzi, B.; Ampofo; Appiah-Kubi, J.; Asare; Assane, H.; Attiku; Awunyo, S.; Bonney; Dagnran, A.; Dorkenoo, A.; Egyir, B.; Halatoko; Issa, Z.; J.K.; K.M.; K.O.; Kossi, K.; Kumordjie, S.; Lamboni, L.; Layibo, Y.; M.A.; Maman, I.; Mohktar, Q.; Odoom; Opoku Frempong, H.; Oteng, F.; Owusu-Nyantakyi, C.; P.O.; S.B.; Sadji; Salah, D.; Salou, M.; Uche; W.K.; Y.A.; Yeboah, C. |
| EPI_ISL_2881744 | InDRE | Instituto Nacional de Medicina Genomica | Cedro-Tanda A; Garcia-Cardenas FJ; Gisela Barrera-Badillo; Gonzalez-Barrera D; Gonzalez-Woge MA; Herrera-Montalvo LA.; Hidalgo-Miranda A; Irma Lopez-Martinez; Jose Ernesto Ramirez González; Mendoza-Vargas A; Munguia-Garza P; Rangel-DeLeon D; Reyes-Grajeda JP |
| EPI_ISL_1517392, EPI_ISL_1517399, EPI_ISL_1517404, EPI_ISL_1517416 | Inciensa, Instituto Costarricense de Investigación y Enseñanza en Nutrición y Salud | Inciensa, Instituto Costarricense de Investigación y Enseñanza en Nutrición y Salud | Cristian Peréz-Corrales; Cristian Peréz-Corrales & Valeria Peralta-Barquero; Valeria Peralta-Barquero & César Cerdas Quesada; Valeria Peralta-Barquero & Mauricio Bolaños; Valeria Peralta-Barquero & Patricia Salas |
| EPI_ISL_476704 | Incubadora Venezolana de Ciencia, Venezuela | Incubadora Venezolana de Ciencia, Venezuela / Instituto Nacional de Salud, Bogotá, Colombia / Grupo de Investigaciones Microbiológicas-UR (GIMUR), Departamento de Biología, Facultad de Ciencias Naturales, Universidad del Rosario, Bogotá, Colombia / Icahn School of Medicine at Mount Sinai, New York, USA | Alberto Paniz-Mondolfi; Ana S. Gonzalez-Reiche; Angelica Rico; Aníbal A. Teherán; Carolina Florez; Carolina Hernández; Emilia Mia Sordillo; Esther C. Barros; Harm van Bakel; Jesús E. Jaimes; Juan David Ramírez; Lisseth Pardo; Lourdes Delgado; Luis Perez-Garcia; Marina Muñoz; Matthew M. Hernandez; Sergio Gomez; Viviana Simon |
| EPI_ISL_3000158 | Infection Prevention & Control Unit | Central Virology laboratory, Ministry of Health | Asaf Biber; Danis Sofer; Efrat Bucris; Ella Mendelson; Gili Regev-Yochay; Ilana Tal; Michal Mandelboim; Miki Goldenfeld; Neta Zuckerman; Orna Mor; Sabrina Hason; Sharon Amit; Shiraz Gefen-Halevi |
| EPI_ISL_2693906 | Infectious Disease and Tropical Medicine Research Center, Resistant Tuberculosis Institute, Zahedan University of Medical Sciences, Zahedan, Iran | Genetics Research Center, University of Social Welfare and Rehabilitation Sciences | Ali Jafarpour; Azam Ghaziasadi; Ebrahim Kord; Hossein Najmabadi.; Khadijeh Jalalvand; Kimia Kahrizi; Marzieh Mohseni; Seyed Mohammad Hashemi-Shahri; Seyed Mohammad Jazayeri; Seyedeh elham Mortazavi; Zohreh Fattahi |
| EPI_ISL_1969944 | Infectious Diseases Hospital IASI | National Institute of Infectious Diseases-Prof. Dr. Matei Bals Molecular Diagnostics Laboratory | Andreea Tudor; Corina Casangiu; Dan Otelea; Leontina Banica; Marius Surleac; Ovidiu Vlaicu; Simona Paraschiv |
| EPI_ISL_856714, EPI_ISL_856715 | Infectious Diseases Unit, Department of Internal Medicine, Azienda Ospedaliera-Universitaria di Padova | Laboratory of Infectious Diseases, Department of Biomedical and Clinical Sciences L. Sacco, University of Milan | Alessia Lai; Anna Maria Cattelan; Annalisa Bergna; Carla Della Ventura; Claudia Balotta; Davide Leoni; Gianguglielmo Zehender on behalf of SARS-CoV-2 ITALIAN RESEARCH ENTERPRISE-(SCIRE) Collaborative Group; Lolita Sasset; Massimo Galli |
| EPI_ISL_2107507, EPI_ISL_2151337 | Infectious Diseases, King Faisal Hospital Research Center | Infectious Diseases, King Faisal Hospital Research Center | Al-Qahtani A, A.; Alahideb B, M.; Alhamlan F, S.; Almaghrabi R, S.; Alsanea M; Alsanea M, S.; Althawadi S, I.; Balavenkatesh Mani; Balavenkatesh Mani, M.; M. and UdayaRaja G, K.; Mutabagani M, S.; S. and Alahideb B, M.; UdayaRaja G, K. |
| EPI_ISL_2013699, EPI_ISL_2014214, EPI_ISL_2089063, EPI_ISL_2282745, EPI_ISL_2282747 | Infinity Biologix | Centers for Disease Control and Prevention Division of Viral Diseases, Pathogen Discovery | Adrian Paskey; Benjamin Rambo-Martin; Chirayu Goswami; Christian Bixby; Christopher Gulvick; Clinton R. Paden; Dakota Howard; Darlene Wagner; Dhwani Batra; Duncan MacCannell; Jason Caravas; Jonathan Schultz; Kara Moser; Matthew Schmerer; Peter W. Cook; Robin Grimwood; Russ Hager; Scott Sammons; Shatavia Morrison; Yihe Wang; Yvette Unoarumhi |
| EPI_ISL_1989269 | Innlandet Hospital Trust, Division Lillehammer, Department for Medical Microbiology | Norwegian Institute of Public Health, Department of Virology | Atiya R Ali; Debech Nadia; Engebretsen Serina Beate; Garcia Llorente Ignacio; Hilde Elshaug; Hilde Vollan; Jon Bråte; Kamilla Heddeland Instefjord; Karoline Bragstad; Kathrine Stene-Johansen; Marie Paulsen Madsen; Olav Hungnes; Pedersen Benedikte Nevjen; Rasmus Riis Kopperud |
| EPI_ISL_2281228, EPI_ISL_2281229, EPI_ISL_2281230, EPI_ISL_2281237, EPI_ISL_2281238, EPI_ISL_2281245, EPI_ISL_2281256 | | | |
| see above | Insitut Pasteur Bangui | Institut Pasteur de Dakar | Amadou Alpha Sall; Amadou Diallo; Benjamin Selekon; Cheikh Loucoubar; Christian Malaka; Mamadou Diop; Marie-Astrid Vernet; Moussa Moïse Diagne; Ndack Ndiaye; Ndongo Dia; Ousmane Faye; Rivalyn Nakoune Yandoko; Safietou Sankhe; Sandra Garba Ouangole |
| EPI_ISL_2828655 | Instituo Adolfo Lutz Central | Instituto Adolfo Lutz, Interdiciplinary Procedures Center, Strategic Laboratory | Caio Vinicius Dias Lopes; Claudia Regina Gonçalves; Claudio Tavares Sacchi; Erica Valessa Ramos Gomes; Karoline Rodrigues Campos |
| EPI_ISL_1907961, EPI_ISL_1908147, EPI_ISL_1908148, EPI_ISL_1908155, EPI_ISL_1908158, EPI_ISL_1908735, EPI_ISL_1908805, EPI_ISL_1908896, EPI_ISL_1908967, EPI_ISL_1909099, EPI_ISL_1909211, EPI_ISL_1909241, EPI_ISL_1909242, EPI_ISL_1909243, EPI_ISL_1909244, EPI_ISL_1909247, EPI_ISL_1909250, EPI_ISL_1909252, EPI_ISL_1913009, EPI_ISL_1913018, EPI_ISL_1913019, EPI_ISL_1913061, EPI_ISL_1913077, EPI_ISL_1913081, EPI_ISL_1913083 | | | |
| see above | Institut National d'Hygiène | Laboratoire de Biotechnologie | Abdelomunim Essabbar; Fatima El Falaki; Fatima ElFalaki; Hicham Oumzil; Lahcen Belyamani and Azeddine Ibrahimi; Mohamde Rhajaoui; Mohamed Rhajaoui; Mouna Ouadghiri; Saaïd Amzazi; Tarik Aanniz |
| EPI_ISL_1433132, EPI_ISL_1435763, EPI_ISL_1436817 | Institut National d'Hygiène | Unité Mixte Internationale TransVIHMI (UMI 233 IRD – U1175 INSERM - Université de Montpellier) IRD (Institut de recherche pour le développement) | Abla A. KONOU; Adodo SADJI; Ahidjo AYOUBA; Akoélé SILIADIN; Alassane OURO-MEDELI; Amivi EHLAN; Améyo DORKENOO; Anoumou DAGNRA; Christelle BUTEL; Déléma MABA; Eric DELAPORTE; Issaka Maman; Kokou TEGUENI; Laetitia SERRANO; Martine PEETERS; Messanh DOUFFAN; Mireille PRINCE-DAVID; Mounerou SALOU; Sidonie A.M.KAGNISSODE; Sika DOSSIM; Wembo A. HALATOKO |
| EPI_ISL_2111158 | Institut National d'Hygiène Consortium National de Veille Génomique - Maroc | Laboratoire de Biotechnologie | Abdelomunim Essabbar; Fatima El Falaki; Hicham Oumzil; Lahcen Belyamani and Azeddine Ibrahimi; Mohamed Rhajaoui; Mouna Ouadghiri; Saaïd Amzazi; Tarik Aanniz |
| EPI_ISL_1415425, EPI_ISL_1416192, EPI_ISL_1443003 | Institut National d'hygiène | "Unité Mixte Internationale TransVIHMI (UMI 233 IRD – U1175 INSERM - Université de Montpellier) IRD (Institut de recherche pour le développement)" | Abla A. KONOU; Adodo SADJI; Ahidjo AYOUBA; Akoélé SILIADIN; Alassane OURO-MEDELI; Amivi EHLAN; Améyo DORKENOO; Anoumou DAGNRA; Christelle BUTEL; Déléma MABA; Eric DELAPORTE; Issaka Maman; Kokou TEGUENI; Laetitia SERRANO; Martine PEETERS; Messanh DOUFFAN; Mireille PRINCE-DAVID; Mounerou SALOU; Sidonie A.M.KAGNISSODE; Sika DOSSIM; Wembo A. HALATOKO |
| EPI_ISL_1508958, EPI_ISL_1508960, EPI_ISL_1509101 | Institut National d'hygiène | Unité Mixte Internationale TransVIHMI (UMI 233 IRD – U1175 INSERM - Université de Montpellier) IRD (Institut de recherche pour le développement) | Abla A. KONOU; Adodo SADJI; Ahidjo AYOUBA; Akoélé SILIADIN; Alassane OURO-MEDELI; Amivi EHLAN; Améyo DORKENOO; Anoumou DAGNRA; Christelle BUTEL; Déléma MABA; Eric DELAPORTE; Issaka Maman; Kokou TEGUENI; Koku AGBODEKA; Laetitia SERRANO; Martine PEETERS; Messanh DOUFFAN; Mireille PRINCE-DAVID; Mounerou SALOU; Sidonie A.M.KAGNISSODE; Sika DOSSIM; Syntyche DEVATCHAGNI; Wembo A. HALATOKO |
| EPI_ISL_1321604, EPI_ISL_1406144, EPI_ISL_2278010 | Institut National d’Hygiène (INH) | Unité Mixte Internationale TransVIHMI (UMI 233 IRD – U1175 INSERM - Université de Montpellier)IRD (Institut de recherche pour le développement) | Abla A. KONOU; Adodo SADJI; Ahidjo AYOUBA; Akoélé SILIADIN; Alassane OURO-MEDELI; Amivi EHLAN; Améyo DORKENOO; Anoumou DAGNRA; Christelle BUTEL; Déléma MABA; Eric DELAPORTE; Issaka Maman; Kokou TEGUENI; Laetitia SERRANO; Martine PEETERS; Messanh DOUFFAN; Mireille PRINCE-DAVID; Mounerou SALOU; Sidonie A.M.KAGNISSODE; Sika DOSSIM; Wembo A. HALATOKO |
| EPI_ISL_418206, EPI_ISL_418210, EPI_ISL_418212, EPI_ISL_476562, EPI_ISL_480783, EPI_ISL_480789, EPI_ISL_485708, EPI_ISL_486869 | | | |
| see above | Institut Pasteur Dakar | Institut Pasteur de Dakar | Amadou Alpha Sall; Amadou Alpha Sall.; Amadou Alpha sall; Mamadou Diop; Mamadou Malado Jallow; Mamadou diop; Marie Henriette Dior Ndione; Moussa Moise Diagne; Ndongo Dia; Ousmane Faye; Safietou Sanke |
| EPI_ISL_498239 | Institut Pasteur de Dakar | Institut Pasteur de Dakar | Amadou Alpha Sall.; Mamadou Diop; Mamadou Malado Jallow; Marie Henriette Dior Ndione; Moussa Moise Diagne; Ndongo Dia; Ousmane Faye; Safietou Sankhe Mbengue |
| EPI_ISL_2245787, EPI_ISL_2245795, EPI_ISL_2245816, EPI_ISL_2245819, EPI_ISL_2245821, EPI_ISL_2245840, EPI_ISL_2245855, EPI_ISL_2245859, EPI_ISL_2245862, EPI_ISL_2245864, EPI_ISL_2245865, EPI_ISL_2245882, EPI_ISL_2245892, EPI_ISL_2245894, EPI_ISL_2245897, EPI_ISL_2245910, EPI_ISL_2245931, EPI_ISL_2245957, EPI_ISL_2245958, EPI_ISL_2245960, EPI_ISL_2245964, EPI_ISL_2245966, EPI_ISL_2245977 | | | |
| see above | Institut Pasteur de Guinée | Institut Pateur de Dakar | Dia Ndongo; Diagne Moussa Moïse; Diallo Amadou; Diop Mamadou; Faye Ousmane; Grayo Solene; Loucoubar Cheikh; Maimouna Mbanne; Mbengue Safietou Sankhe; Mohamed Kane; Ndiaye Ndack; Sall Amadou Alpha; Tordo Noel |
| EPI_ISL_613421, EPI_ISL_613438, EPI_ISL_613441, EPI_ISL_613446, EPI_ISL_613455 | Institut Pasteur de la Guadeloupe | Institut Pasteur de la Guadeloupe | Angela Brisebarre; Antoine Talarmin; Camille Capel; Cherina Fleming; Etienne Simon-Lorière; Marion Barbet; Maud Vanpeene; Méline Bizard; Radjin Steingrover; Stéphanie Guyomard; Sylvie Behillil; Sylvie van der Werf; Sébastien Breurec; Vincent Enouf |
| EPI_ISL_459971 | Institut Pasteur du Maroc | Institut Pasteur du Maroc | Abdellah Faouzi; Anass Abbad; Anderrahmane Maaroufi; Angela Brisebarre; Camille Capel; Etienne Simon-Lorière; Jalal Nourlil; Latifa Anga; Marion Barbet; Maud Vanpeene; Mjid Eloualid; Méline Bizard; Sylvie Behillil; Sylvie van der Werf; Vincent Enouf |
| EPI_ISL_475820, EPI_ISL_1008302 | Institut für Virologie am Department für Hygiene, Mikrobiologie und Public Health | Bergthaler laboratory, CeMM Research Center for Molecular Medicine of the Austrian Academy of Sciences | Alexander Lercher; Alexandra Popa; Andreas Bergthaler; Anna Schedl; Bekir Erguner; Benedikt Agerer; Christoph Bock; Daniela Schmid; Dorothee von Laer; Elisabeth Puchhammer-Stoeckl; Franz Allerberger; Gregor Hörmann; Guenter Weiss; Henrique Colaco; Jakob-Wendelin Genger; Jan Laine; Judith Aberle; Kinga Rigler-Hohenwarter; Lukas Endler; Maelle Le Moing; Manfred Nairz; Mark Smyth; Martin Senekowitsch; Michael Schuster; Peter Hufnagl; Rainer Gattringer; Stephan Aberle; Thomas Penz; Wegene Borena |
| EPI_ISL_955168 | Institute for Biocides and Medical Ecology | Institute of microbiology and Immunology, Faculty of Medicine, University of Belgrade | Jankovic, M.; Jovanovic, T.; Knezevic, A.; Milicevic, O.; Sekler, M.; Tesovic, B.; Vidanovic, D. |
| EPI_ISL_1657085, EPI_ISL_1915108 | Institute for Developing Science and Health Initiatives (ideSHi) | Institute for Developing Science and Health Initiatives (ideSHi) | Fidausi Qadri; Hassan Afrad; Sadia Rahman; Tahmina Shirin |
| EPI_ISL_1822604, EPI_ISL_1827861, EPI_ISL_1827862, EPI_ISL_1827865, EPI_ISL_1827890, EPI_ISL_1827915, EPI_ISL_1827932, EPI_ISL_1827945 | | | |
| see above | Institute for Health Research, Epidemiological Surveillance and Training (IRESSEF) | Abbott Laboratories | Adbou Padane; Ambroise Ahouidi; Aminata Dia; Aminata Mboup; Ana Olivo; Anna julienne selbe Ndiaye; Barbara Harris; Cyrille Diedhiou; Gavin Cloherty; Mary Rodgers; Moustapha Mbow; Nafissatou Leye; Ndeye Diabou Diagne; Papa Alassane Diaw; Souleymane Mboup; Todd Meyer |
| EPI_ISL_574790 | Institute for Infectious Diseases, University of Bern | Institute for Infectious Diseases, University of Bern | Alban Ramette; Christian Baumann; Cora Sägesser; Franziska Suter-Riniker; Michel C Koch; Miguel A Terrazos Miani; Peter Keller; Stephen L Leib |
| EPI_ISL_2758186 | Institute for Laboratory Diagnostics and Microbiology, Klinikum Klagenfurt am Wörthersee | Bergthaler laboratory, CeMM Research Center for Molecular Medicine of the Austrian Academy of Sciences | Andreas Bergthaler; Anna Schedl; Bekir Erguner; Benedikt Agerer; Christoph Bock; Fabian Amman; Jan Laine; Lukas Endler; Maelle Le Moing; Martin Senekowitsch; Matthew Thornton; Michael Schuster; Petr Triska; Thomas Penz |

| | | | |
|---|---|---|---|
| EPI_ISL_490090, EPI_ISL_490096, EPI_ISL_490101, EPI_ISL_490103, EPI_ISL_644672, EPI_ISL_718289, EPI_ISL_718301, EPI_ISL_944102, EPI_ISL_1114744, EPI_ISL_1263458, EPI_ISL_1406244, EPI_ISL_1673342, EPI_ISL_1696746, EPI_ISL_1696754, EPI_ISL_1787317, EPI_ISL_2091000, EPI_ISL_2625633, EPI_ISL_2685838 | | | |
| see above | Institute for Medical Research, Infectious Disease Research Centre, National Institutes of Health, Ministry of Health Malaysia | Institute for Medical Research, Infectious Disease Research Centre, National Institutes of Health, Ministry of Health Malaysia | Azizan MA; Kalyanasundram J; Kamel K; Mohd Zawawi Z; Mohd-Zawawi Z; Ramly N; Robert F; Suppiah J; Thayan R |
| EPI_ISL_2324270 | Institute for Medical and Chemical Laboratory Diagnostics, Kepler Universitätsklinikum | Bergthaler laboratory, CeMM Research Center for Molecular Medicine of the Austrian Academy of Sciences | Andreas Bergthaler; Anna Schedl; Bekir Erguner; Benedikt Agerer; Christoph Bock; Fabian Amman; Jan Laine; Lukas Endler; Maelle Le Moing; Martin Senekowitsch; Michael Schuster; Petr Triska; Thomas Penz |
| EPI_ISL_812955, EPI_ISL_1491539, EPI_ISL_1511131 | Institute for Urban Disease Control and Prevention | COVID-19 Network Investigations (CONI) Alliance | Amornmas Kongklieng; Anek Mungaomklang; Angkana Huang; Anthony R. Jones; Arporn Wangwiwatsin; Bhakbhoom Panthan; Chonticha Klungtong; Duangkamon Loesbanluechai; Ekawat Pasomsub; Elizabeth Batty; Insee Sensorn; Janjira Thaipadungpanit; Kamolthip Atsawawaranunt; Khajohn Joonlasak; Khajohn Joonsalak; Kingkan Rakmanee; Krittikorn Kumpornsin; Namfon Kotanan; Prayuth Kaewmalang; Pukkaporn Parnwijitkul; Stefan Fernandez; Thanat Chookajorn; Theerarat Kochakarn; Treewat Watthanachockchai; Vichan Pawun; Wasun Chantratita; Wudtichai Manasatienkij |
| EPI_ISL_626228 | Institute for Virology, University Hospital Essen | Center of Medical Microbiology, Virology, and Hospital Hygiene, University of Duesseldorf | Alexander Dilthey; Andreas Walker; Daniel Strelow; Jessica Nicolai; Jörg Timm; Klaus Pfeffer; Lisanna Hülse; Malte Kohns Vasconcelos; Maximilian Damagnez; Nadine Lübke; Olympia E. Anastasiou; Tobias Wienemann; Torsten Houwaart; Ulf Dittmer |
| EPI_ISL_2137158 | Institute for Water Quality and Resource Management, Technical University Vienna | Bergthaler laboratory, CeMM Research Center for Molecular Medicine of the Austrian Academy of Sciences | Andreas Bergthaler; Anna Schedl; Bekir Erguner; Benedikt Agerer; Christoph Bock; Fabian Amman; Jan Laine; Lukas Endler; Maelle Le Moing; Martin Senekowitsch; Michael Schuster; Petr Triska; Thomas Penz |
| EPI_ISL_1057041, EPI_ISL_1057098 | Institute of Biocides and Medical Ecology, Belgarde, Serbia | Virology Department Institute of Microbiology and Immunology Faculty of Medicine University of Belgrade | Abazovic Dzihan; Banko Ana; Despot Dragana; Loncar Ana; Milicevic Ognjen; Miljanovic Danijela |
| EPI_ISL_891266 | Institute of Biocides and Medical Ecology, Belgrade, Serbia | Virology department Institute of microbiology and immunology Faculty of Medicine University of Belgrade | Abazovic Dzihan; Banko Ana; Despot Dragana; Loncar Ana; Milicevic Ognjen; Miljanovic Danijela |
| EPI_ISL_2180456, EPI_ISL_2284894, EPI_ISL_2600368, EPI_ISL_2600369, EPI_ISL_2774039, EPI_ISL_2774043, EPI_ISL_2774074 | | | |
| see above | Institute of Epidemiology, Disease Control and Research (IEDCR) | IEDCR-ideSHi-icddr,b | Firdausi Qadri; Hassan Afrad; Manjur Hossain Khan; Sadia Rahman; Tahmina Shirin |
| EPI_ISL_1938476 | Institute of Epidemiology, Disease Control and Research (IEDCR) | Institute for Developing Science and Health Initiatives (ideSHi) | Fidausi Qadri; Hassan Afrad; Sadia Rahman; Tahmina Shirin |
| EPI_ISL_864573 | Institute of Medical Microbiology and Hospital Hygiene | Institute of Medical Microbiology and Hospital Hygiene | Aljoscha Tersteegen; Prof. Dr. Achim Kaasch |
| EPI_ISL_1511107, EPI_ISL_1668835, EPI_ISL_1964492, EPI_ISL_2492324, EPI_ISL_2492327, EPI_ISL_2644269, EPI_ISL_2932442, EPI_ISL_2955269, EPI_ISL_2966919, EPI_ISL_3039361 | | | |
| see above | Institute of Microbiology and Immunology, Faculty of Medicine, University of Ljubljana | Institute of Microbiology and Immunology, Faculty of Medicine, University of Ljubljana | Alen Suljič; Andraž Celar; Dominika Šturm; Doroteja Vlaj; Mario Poljak; Matic Brvar; Miša Korva; Patricija Pozvek; Samo Zakotnik; Tatjana Avšič – Županc; Tomaž Mark Zorec; Špela Pleh |
| EPI_ISL_417482, EPI_ISL_477016, EPI_ISL_491936, EPI_ISL_516650, EPI_ISL_516651, EPI_ISL_527813, EPI_ISL_539789, EPI_ISL_660530, EPI_ISL_697798, EPI_ISL_728202, EPI_ISL_824285, EPI_ISL_877557, EPI_ISL_1805654, EPI_ISL_1805657, EPI_ISL_1805711, EPI_ISL_1896678, EPI_ISL_1896691, EPI_ISL_1896695, EPI_ISL_2004102, EPI_ISL_2004105, EPI_ISL_2086711, EPI_ISL_2100425, EPI_ISL_2100438, EPI_ISL_2134873, EPI_ISL_2228102, EPI_ISL_2228105, EPI_ISL_2361460, EPI_ISL_2361462, EPI_ISL_2361472, EPI_ISL_2361476, EPI_ISL_2484966, EPI_ISL_2534327, EPI_ISL_2689843, EPI_ISL_2774510, EPI_ISL_2861153 | | | |
| see above | Institute of Microbiology, Universidad San Francisco de Quito | Institute of Microbiology, Universidad San Francisco de Quito | ; Alberto Velez; Andrea Macías; Belén Prado-Vivar; Bernardo Darquea; Bernardo Gutiérrez; Carla Torres; Carlos Guerrero; Carlos Mena; Carolina Pacheco; Christian Zambrano; DamarisSandoya; David Zuñiga; Diana Zambrano; Edison Chavez; Erika Muñoz; Ernesto Pazos; Eulalia Pazmiño; Fernada Zurita; Fernanda Zurita; Fernando Serrano; Francisco Ríos; Fredy Loor; Gabriel Trueba; Gabriela Zambrano; Guzmán Bernabéu Lorenzo; Hermelinda Paguay; Hugo Hernandéz; Hugo Hernández; Jorge Luis Velez; Jorge Montaño; Juan Carlos Zuñiga; Juan José Guadalupe; Juan Pablo Haro; Juan Zuñiga; Katalina Pacheco; Katherine Ojeda; Khurram Mahbbob; Ligia Briceño; Luis Flores; Luis Morales; Manuel Jaramillo; Manuel Jibaja; Mayra Beltrán; Melissa Ortega; Melva Morales; Michelle Grunauer; Milton Tobar; Miriam Ruiz; Monica Becerra-Wong; Monica Becerra-Wong Juan José Guadalupe; Nabih Dahik; Oscar Mena; Paola Dalgo; Patricio Reyes; Patricio Rojas-Silva; Patricio Vega-Mariño; Paúl Cárdenas; Prado-Vivar; Rafael Parra; Raiza Briceño; Ramiro Echeverría Tapia; Rommel Guevara; Sully Márquez; Tania Guayasamin; Tania Lluma; Tanya Guayasamin; Verónica Barragán; Victor Vinicio Román |
| EPI_ISL_1745684, EPI_ISL_1745694, EPI_ISL_1745705, EPI_ISL_2346373, EPI_ISL_2727514, EPI_ISL_3006762 | Institute of Molecular and Translational Medicine / Laboratory of Experimental Medicine, Faculty of Medicine and Dentistry, Palacky University and University Hospital Olomouc | Institute of Molecular and Translational Medicine / Laboratory of Experimental Medicine, Faculty of Medicine and Dentistry, Palacky University | Barbora Blumová; Hana Dřímalová; Hana Jaworek; Marián Hajdúch; Rastislav Slavkovský; Tomáš Pospíšíl; Vladimíra Koudeláková |
| EPI_ISL_402123 | Institute of Pathogen Biology, Chinese Academy of Medical Sciences & Peking Union Medical College | Institute of Pathogen Biology, Chinese Academy of Medical Sciences & Peking Union Medical College | Chao Wu; Jianwei Wang; Lili Ren; Qi Jin; Yiwei Liu; Zhiqiang Wu; Zichun Xiang |
| EPI_ISL_2932530, EPI_ISL_2932531, EPI_ISL_2986987 | Institute of Public Health Kraljevo | Veterinary Specialized Institute Kraljevo | Afonso, C.; Banovic Djeri, B.; Djukic V, .; Jankovic, M.; Jovanovic, T.; Knezevic, A.; Petrovic, T.; Sekler, M.; Tesovic, B.; Vidanovic, D.; Volkening, J. |
| EPI_ISL_2657582 | Institute of Public Health Šibenik-Knin County | Croatian Institute of Public Health | Irena Tabain; Ivana Ferenčak |
| EPI_ISL_1712568 | Institute of Virology Department of Hygiene, Microbiology and Public Health at Innsbruck Medical University | Institute of Virology Department of Hygiene, Microbiology and Public Health at Innsbruck Medical University | Andre Volland; Barbara Falkensammer; David Bante; Dorothee von Laer; Heribert Stoiber; Janine Kimpel; Lisa Pipperger; Lydia Riepler; Wegene Borena |
| EPI_ISL_577740, EPI_ISL_577742, EPI_ISL_904001, EPI_ISL_1994596, EPI_ISL_2351249, EPI_ISL_2894457 | Institute of Virology, Biomedical Research Center of the Slovak Academy of Sciences, Bratislava | Faculty of Natural Sciences, Comenius University, Bratislava | Boris Klempa; Brona Brejova; Broňa Brejová; Dominika Fričová; Edita Staroňová; Elena Tichá; Jozef Nosek; Juraj Kopacek; Juraj Kopáček; Kristina Borsova; Kristína Boršová; Lubomira Lukacikova; Martina Lickova; Martina Ličková; Martina Nebohacova; Martina Neboháčová; Monika Slavikova; Monika Sláviková; Sabina Fumacova Havlikova; Sabina Fumačová Havlíková; Tomas Vinar; Tomáš Vinař; Viktoria Cabanova; Viktoria Hodorova; Viktória Hodorová; Viktória Čabanová; Ľubomíra Lukáčiková |
| EPI_ISL_1654810, EPI_ISL_1654830 | Institute of Virology, Vaccines and Sera "Torlak" | Institute of microbiology and Immunology, Faculty of Medicine, University of Belgrade | Jankovic, M.; Jovanovic, T.; Knezevic, A.; Milicevic, O.; Sekler, M.; Tesovic, B.; Vidanovic, D. |
| EPI_ISL_1017684, EPI_ISL_1017686, EPI_ISL_1017688 | Institute of Virology, Vaccines and Sera ''Torlak'' | Institute of microbiology and Immunology, Faculty of Medicine, University of Belgrade | Jankovic, M.; Jovanovic, T.; Knezevic, A.; Milicevic, O.; Sekler, M.; Tesovic, B.; Vidanovic, D. |
| EPI_ISL_833163 | Instituto Adolfo Lutz - Regional de Marilia | Instituto Adolfo Lutz, Interdiciplinary Procedures Center, Strategic Laboratory | Claudia Regina Gonçalves; Claudio Tavares Sacchi; Erica Valessa Ramos Gomes; Karoline Rodrigues Campos |
| EPI_ISL_2614622 | Instituto Adolfo Lutz - Regional de Presidente Prudente | Instituto Adolfo Lutz, Interdiciplinary Procedures Center, Strategic Laboratory | Caio Vinicius Dias Lopes; Claudia Regina Gonçalves; Claudio Tavares Sacchi; Erica Valessa Ramos Gomes; Karoline Rodrigues Campos; Leonardo Jose Tadeu de Araujo |
| EPI_ISL_984246 | Instituto Adolfo Lutz Central | Instituto Adolfo Lutz, Interdiciplinary Procedures Center, Strategic Laboratory | Claudia Regina Gonçalves; Claudio Tavares Sacchi; Erica Valessa Ramos Gomes; Karoline Rodrigues Campos |
| EPI_ISL_2344311, EPI_ISL_2344420 | Instituto Butantan | Instituto de Medicina Tropical de Sao Paulo | Brazil-UK Centre for Arbovirus Discovery Diagnosis Genomics and Epidemiology (CADDE) Genomic Network - Instituto de Medicina Tropical |
| EPI_ISL_491196, EPI_ISL_491207, EPI_ISL_491212 | Instituto Gulbenkian de Ciência | Instituto Gulbenkian de Ciência | Cathy Paulino; Joao Sobral; João Costa; Ricardo Leite; Susana Ladeiro |
| EPI_ISL_2348751, EPI_ISL_2348766, EPI_ISL_2348777 | Instituto Nacional De Investigación En Salud Pública-Crn De Influenza Y Otros Virus Respiratorios | NIC-Instituto Nacional de Investigación en Salud Pública | Alfredo Bruno; Domenica de Mora.; Jimmy Garcés; Johanna Laines; Lizbeth Patiño; Manuel Gonzalez; Maritza Olmedo; Michelle Páez |
| EPI_ISL_837720 | Instituto Nacional de Enfermedades Respiratorias (INER) | Instituto Nacional de Enfermedades Respiratorias (INER) | Alejandra Hernández-Terán; Alma Rincón-Rubio; Celia Boukadida; Edgar Sevilla-Reyes; Eduardo Becerril-Vargas; Fidencio Mejía-Nepomuceno; Hector Esteban Paz-Juárez; Joel Armando Vázquez-Pérez; Jorge Salas-Hernández; José Arturo Martínez-Orozco; Margarita Matías-Florentino; Mario Mújica-Sánchez; Olivia Briceño; Santiago Ávila-Ríos |
| EPI_ISL_574294 | Instituto Nacional de Investigacion en Salud Pública | Instituto Nacional de Investigación en Salud Pública | Alberto Orlando Narvaez; Alfredo Bruno; Andrés Carrasco; Denisse Portugal; Doménica de Mora Coloma; Leandro Patino Patino; Manuel Gonzalez; Orson Mestanza |
| EPI_ISL_491950 | Instituto Nacional de Investigación en Salud Pública - INSPI | INSPI - Charité | Alberto Orlando; Alexandra Usiña; Alfredo Bruno Caicedo; Andres Moreira-Soto; Anna-Lena Sander; Denisses Portugal; Domenica de Mora Coloma; Jan Felix Drexler; Juan Carlos Zeballos; Manuel Gonzalez; Maritza Olmedo; Nina Krause; Silvia Salgado |
| EPI_ISL_2492549, EPI_ISL_2492604, EPI_ISL_2492663, EPI_ISL_2492696, EPI_ISL_2492728, EPI_ISL_2492780, EPI_ISL_2492786, EPI_ISL_2492787, EPI_ISL_2492788, EPI_ISL_2492789, EPI_ISL_2492799, EPI_ISL_2492803, EPI_ISL_2492818, EPI_ISL_2492822, EPI_ISL_2492846, EPI_ISL_2492847, EPI_ISL_2492882, EPI_ISL_2492915, EPI_ISL_2493087, EPI_ISL_2617133, EPI_ISL_2617181, EPI_ISL_2617182, EPI_ISL_2685951, EPI_ISL_2685957, EPI_ISL_2685960, EPI_ISL_2685973, EPI_ISL_2841653, EPI_ISL_2841654 | | | |
| see above | Instituto Nacional de Investigação em Saúde | CERI, Centre for Epidemic Response and Innvoation, Stellenbosch University and KRISP, KZN Research Innovation and Sequencing Platform, UKZN. | Afonso P; David K; Emmanuel SJ; Freitas RH; Giandhari J; Inglês L; Lutucuta S; Miranda J; Morais J; Mufinda M; Naidoo Y; Neto Z; Paulo A Carralero RR Paixão JP; Pereira A; Pillay S; Tegally H; Wilkinson E; de Oliveira T |
| EPI_ISL_1347897, EPI_ISL_1545274, EPI_ISL_1545278, EPI_ISL_1545383, EPI_ISL_2494880, EPI_ISL_2494884, EPI_ISL_2494900, EPI_ISL_2609486, EPI_ISL_2609493, EPI_ISL_2609531, EPI_ISL_2609540, EPI_ISL_2609541, EPI_ISL_2609561 | | | |
| see above | Instituto Nacional de Investigação em Saúde | KRISP, KZN Research Innovation and Sequencing Platform | Afonso P; David K; Emmanuel SJ; Freitas RH; Giandhari J; Inglês L; Lutucuta S; Miranda J; Morais J; Mufinda M; Naidoo Y; Neto Z; Paulo A Carralero RR Paixão JP; Pereira A; Pillay S; Tegally H; Wilkinson E; de Oliveira T |
| EPI_ISL_933521, EPI_ISL_1591655, EPI_ISL_1711328, EPI_ISL_2230812, EPI_ISL_2894314 | Instituto Nacional de Medicina Genomica | Instituto Nacional de Medicina Genomica | Alcaraz N; Canseco Mendez JC; Cedro-Tanda A; Cisneros- Villanueva M; Cisneros-Villanueva M; Escobar-Arrazola; Garcia-Cardenas FJ; Gonzalez-Barrera D; Gonzalez-Woge MA; Herrera-Montalvo LA; Herrera-Montalvo LA.; Hidalgo-Miranda A; M; Mendoza-Vargas A; Miranda-Ortiz H; Munguia-Garza P; Peñaloza-Figueroa F; Ramirez-Vega O; Rangel-DeLeon D; Reyes-Grajeda JP; Rosas-Escobar P |
| EPI_ISL_2500953, EPI_ISL_2500971 | Instituto Nacional de Salud | Instituto Nacional de Salud- Dirección de Investigación en Salud Pública | Carlos Franco-Muñoz; Carmen Osorio; Diana Malo; Diego A. Álvarez-Díaz; Diego Andrés Prada; Gerardo Santamaría; Hector Alejandro Ruiz-Moreno; Jhonnatan Reales-González; Jorge Rivera; Juan Camilo Martinez; Julian Naizaque; Katherine Laiton-Donato; Lisseth Pardo; Magdalena Wiesner; Marcela Mercado-Reyes; Maria T. Herrera-Sepúlveda; Marta Lopez Blanco; Martha Lucia Ospina Martinez; Paola Rojas; Sergio Gomez; Sheryll Corchuelo; Ángela Alarcon Cruz |
| EPI_ISL_791087, EPI_ISL_791096 | Instituto Nacional de Salud - Unidad de Secuenciación y Análisis Genómico | Instituto Nacional de Salud - Dirección de Investigación en Salud Pública | Carlos Franco-Muñoz; Diego A. Álvarez-Díaz; Diego Andrés Prada; Gerardo Santamaría; Jonathan Reales; Julian Naizaque; Katherine Laiton-Donato; Magdalena Wiesner; Marcela Mercado-Reyes; Maria T. Herrera; Martha Lucia Ospina Martinez; Mauricio Pacheco-Montealegre; Paola Muñoz-Laiton; Sheryll Corchuelo |

| | | | |
|---|---|---|---|
| EPI_ISL_941955, EPI_ISL_941986 | Instituto Nacional de Salud, Bogotá, Colombia | Centro de Investigaciones en Microbiología y Biotecnología-UR (CIMBIUR), Facultad de Ciencias Naturales, Universidad del Rosario, Bogotá, Colombia Instituto Nacional de Salud, Bogotá, Colombia Icahn School of Medicine at Mount Sinai, New York, USA | Adriana van de Guchte; Alberto Paniz-Mondolfi; Ana S. Gonzalez-Reiche; Carolina Flórez; Carolina Hernández; Emilia Mia Sordillo; Hala Alejel Alshammary; Harm van Bakel; Jayeeta Dutta; Juan David Ramírez; Luz Helena Patiño; Marina Muñoz; Matthew M. Hernandez; Nathalia Ballesteros; Sergio Gomez; Viviana Simon; Zenab Khan |
| EPI_ISL_447804 | Instituto Nacional de Salud, Bogotá, Colombia | Grupo de Investigaciones Microbiológicas-UR (GIMUR), Departamento de Biología, Facultad de Ciencias Naturales, Universidad del Rosario, Bogotá, Colombia Instituto Nacional de Salud, Bogotá, Colombia Icahn School of Medicine at Mount Sinai, New York, USA | Adriana Castillo; Alberto Paniz-Mondolfi; Ana S. Gonzalez-Reiche; Angelica Rico; Aníbal A. Teherán; Carolina Florez; Carolina Hernandez; David Martínez; Emilia Mia Sordillo; Esther C. Barros; Harm van Bakel; Jesús E. Jaimes; Juan David Ramírez; Laura Vega; Lisseth Pardo; Marina Muñoz; Martha L. Ospina; Matthew M. Hernandez; Nathalia Ballesteros; Sergio Castañeda; Sergio Gomez; Viviana Simon |
| EPI_ISL_526967 | Instituto Nacional de Salud, Bogotá, Colombia | Instituto Nacional de Salud, Bogotá, Colombia | Astrid C. Flórez; Carlos Andrés Durán; Carlos Franco-Muñoz; Carolina Ferro; Christian Julian Villabona-Arenas; Diana Marcela Walteros-Acero; Diego A. Álvarez-Díaz; Diego Andrés Prada; Franklin Prieto; Jonathan Reales; Jose A. Usme-Ciro; Katherine Laiton-Donato; Liz Villabona-Arenas; Marcela Mercado-Reyes; Martha Lucia Ospina Martinez; Mauricio Pacheco-Montealegre; Sussy Echeverria; Zulma M. Cucunubá |
| EPI_ISL_2617100, EPI_ISL_2617116, EPI_ISL_2688576, EPI_ISL_2688577, EPI_ISL_2688578 | Instituto Nacional de Saude (INS), Mozambique | CERI, Centre for Epidemic Response and Innvoation, Stellenbosch University and KRISP, KZN Research Innovation and Sequencing Platform, UKZN. | Emmanuel S; Giandhari J; Nadia Sitoe; Naidoo Yeshnee; Nalia Ismael; Nedio Mabunda; Paulo Arnaldo; Pillay S; Tegally H; Tshabuila Derek; Wilkinson E; Yajna Ramphal; de Oliveira T |
| EPI_ISL_887419, EPI_ISL_887429, EPI_ISL_887433, EPI_ISL_887501, EPI_ISL_964947, EPI_ISL_1132836, EPI_ISL_1132840, EPI_ISL_2396914, EPI_ISL_2396917, EPI_ISL_2396929, EPI_ISL_2396934, EPI_ISL_2396950, EPI_ISL_2396994 | | | |
| see above | Instituto Nacional de Saude (INS), Mozambique | KRISP, KZN Research Innovation and Sequencing Platform | Emmanuel S; Giandhari J; Nadia Sitoe; Nalia Ismael; Nedio Mabunda; Paulo Arnaldo; Pillay S; Tegally H; Wilkinson E; de Oliveira T |
| EPI_ISL_2843966 | Instituto de Biotecnologia - UNESP-Botucatu-SP | Instituto de Biotecnologia - UNESP-Botucatu-SP | Cecília Artico Banho; Cíntia Bittar; Fábio Sossai Possebon; Guilherme Campos; Helena Lage Ferreira; Jorge A. Petroli Marchesi; João Pessoa Araújo Jr.; Leila Sabrina Ullmann; Lívia Sacchetto; Maisa C. Pereira Parra; Marília Moraes; Maurício L. Nogueira; Paula Rahal; Paulo Inacio da Costa |
| EPI_ISL_516625, EPI_ISL_576266, EPI_ISL_576274, EPI_ISL_658893 | Instituto de Diagnostico y Referencia Epidemiologicos (INDRE) | Instituto de Diagnostico y Referencia Epidemiologicos (INDRE) | Abril Rodriguez-Maldonado; Adnan Araiza-Rodriguez; Claudia Wong-Arambula; Dayanira Arellano-Suarez; Edgar Mendieta-Condado; Ernesto Ramirez-Gonzalez; Ernesto Ramirez-Gonzalez.; Fabiola Garces-Ayala; Gisela Barrera-Badillo; Gisela Barrera-Badillo.; Irma Lopez-Martinez; Lucia Hernandez-Rivas; Natividad Cruz-Ortiz; Tatiana Nunez-Garcia |
| EPI_ISL_452141 | Instituto de Diagnostico y Referencia Epidemiologicos (INDRE) | Instituto de diagnóstico y Referencia Epidemiologicos (INDRE) | Araiza-Rodriguez Adnan; Barrera-Badillo Gisela; Garces-Ayala Fabiola; Hernandez-Rivas Lucia; Lopez-Martinez Irma.; Mendieta-Condado Edgar; Ramirez-Gonzalez Ernesto; Rodriguez-Maldonado Abril; Wong-Arambula Claudia |
| EPI_ISL_1060682, EPI_ISL_1060743, EPI_ISL_1060756, EPI_ISL_1060760, EPI_ISL_1060762 | Instituto de Diagnostico y Referencia Epidemiologicos (INDRE)_RNLSP | Instituto de Diagnostico y Referencia Epidemiologicos (INDRE) | Abril Rodriguez-Maldonado; Adnan Araiza-Rodriguez; Claudia Wong-Arambula; David Fragoso-Fonseca; Ernesto Ramirez-Gonzalez.; Fabiola Garces-Ayala; Gisela Barrera-Badillo; Irma Lopez-Martinez; Lucia Hernandez-Rivas; Mayra Jimenez-Morales; Nancy Munoz-Hernandez; Natividad Cruz-Ortiz; Sergio Rangel-Guerrero; Tatiana Nunez-Garcia |
| EPI_ISL_913933, EPI_ISL_913944, EPI_ISL_913959, EPI_ISL_1054977, EPI_ISL_1168563 | Instituto de Diagnostico y Referencia Epidemiologicos INDRE_RNLSP | Instituto de Diagnostico y Referencia Epidemiologicos (INDRE) | Abril Rodriguez-Maldonado; Adnan Araiza-Rodriguez; Ariadna Medina-Benitez; Claudia Wong-Arambula; David Fragoso-Fonseca; Ernesto Ramirez-Gonzalez.; Fabiola Garces-Ayala; Gisela Barrera-Badillo; Irma Lopez-Martinez; Joaquin Quiroz-Mercado; Lucia Hernandez-Rivas; Mayra Jimenez-Morales; Nancy Munoz-Hernandez; Natividad Cruz-Ortiz; Sergio Rangel-Guerrero; Tatiana Nunez-Garcia; Vanessa Rivero-Arredondo |
| EPI_ISL_1301527, EPI_ISL_1301559, EPI_ISL_1301712 | Instituto de Diagnostico y Referencia Epidemiologicos InDRE_RNLSP | Instituto de Biotecnología de la UNAM | Alejandra Hernández-Terán; Alejandro Sanchez-Flores; Alma Rincón-Rubio; Andrea Santos Coy-Arechavaleta; Authors from IBT; Blanca Taboada; Celia Boukadida; Clara Esperanza Santacruz-Tinoco; Edgar Mendieta-Condado; Eduardo Becerril-Vargas; Fidencio Mejía-Nepomuceno; Francisco Pulido; Gisela Barrera-Badillo; Gloria Vazquez; Hector Esteban Paz-Juárez; IMSS; InDRE and INER (in alphabetical order): Carlos F. Arias; Irma Lopez-Martinez; Jerome Jean Verleyen; Joel Armando Vázquez-Pérez; Jorge Salas-Hernández; José Arturo Martínez-Orozco; José Ernesto Ramírez-González; José Esteban Muñoz-Medina; Larissa Fernandes-Matano; Lucia Hernandez-Rivas; Luis Alberto Ochoa-Carrera; Margarita Matías-Florentino; Mario Mújica-Sánchez; Natividad Cruz-Ortiz; Pavel Isa; Ricardo Grande; Santiago Ávila-Ríos; Tatiana Nunez-Garcia; Teresita Rojas-Mendoza |
| EPI_ISL_2776172, EPI_ISL_2776174, EPI_ISL_2776175, EPI_ISL_2776177, EPI_ISL_2776178, EPI_ISL_2776179, EPI_ISL_2776182, EPI_ISL_2776197 | | | |
| see above | Instituto de Medicina Tropical & Salud Global (IMTSAG) | Grubaugh Lab - Yale School of Public Health | Alejandaro Vallejo Degaudenzi; Anderson Brito; Annie Watkins; Chaney Kalinich; Chantal Vogels; Elisa Contreras; Esperanza Mendoza; Isabel Ott; Jessica Rothman; Joseph Fauver; Kendall Billig; Mallery Breban; Mary Petrone; Nathan Grubaugh; Robert Paulino-Ramirez; Tara Alpert; Tobias Koch; Victor Virgilio Calderon |
| EPI_ISL_3020245, EPI_ISL_3020248, EPI_ISL_3020257, EPI_ISL_3020269 | Instituto de Medicina Tropical Alexander Von Humboldt, Sede Cusco | Laboratorio de Genómica Microbiana, Universidad Peruana Cayetano Heredia | Alejandra Dávila-Barclay; Diego Cuicapuza; Guillermo Salvatierra; Janet Huancachoque; Luis González; Pablo Tsukayama; Pedro E. Romero; Pool Marcos |
| EPI_ISL_729916 | Instituto de Medicina Tropical, Universidad Nacional Toribio Rodríguez de Mendoza de Amazonas | Laboratorio de Genómica Microbiana, Universidad Peruana Cayetano Heredia | Alejandra Dávila-Barclay; Brenda Ayzanoa; Carla Montenegro; Cecilia Pajuelo; Janet Huancachoque; Luis González; Pablo Tsukayama; Pedro E. Romero; Pool Marcos; Rafael Tapia; Stella Chenet |
| EPI_ISL_1627995 | Instytut Onkologii | 1. National Institute of Public Health - National Institute of Hygiene; 2. Eurofins Genomics Europe Sequencing GmbH | ECDC COVID-19 WGS support team; Eurofins Genomics Europe Sequencing Team; Gierczyński Rafał; Sadkowska-Todys Małgorzata; Wołkowicz Tomasz; Zacharczuk Katarzyna |
| EPI_ISL_1652050, EPI_ISL_1652051, EPI_ISL_1652052, EPI_ISL_1652053, EPI_ISL_1652054, EPI_ISL_1652055 | Integrated Biorepository of H3Africa Uganda – IBRH3AU | Molecular Biology Laboratory | Ashaba Fred Katabazi; Bernard Ssentalo Bagaya; David Patrick Kateete; Edgar Kigozi; Emmanuel Nasinghe; Eric Katagirya; Gerald Mboowa; Lwanga Newton; Misaki Wayengera; Moses Joloba; Moses Luutu; Nsubuga Gideon; Rogers Kamulegeya; Samuel Kirimunda; Sarah Stanley; Savannah Mwesigwa |
| EPI_ISL_1919645, EPI_ISL_1919646 | Ipoh Public Health Laboratory (MKAI), Ministry of Health Malaysia | Institute for Medical Research, Infectious Disease Research Centre, National Institutes of Health, Ministry | Kamel K; Mohd Zawawi Z; Suppiah J; Thayan R |
| EPI_ISL_1919858, EPI_ISL_2650007, EPI_ISL_2815329, EPI_ISL_2815330 | Ipoh Public Health Laboratory (MKAI), Ministry of Health Malaysia | Institute for Medical Research, Infectious Disease Research Centre, National Institutes of Health, Ministry of Health Malaysia | Azizan MA; Kamel K; Mohd Zawawi Z; Ramly N; Robert F; Suppiah J; Thayan R |
| EPI_ISL_2795660, EPI_ISL_3038900 | Iran University of Medical Sciences, Tehran, Iran | Genetics Research Center, University of Social Welfare and Rehabilitation Sciences | Farah Bokharaei-Salim; Hossein Najmabadi.; Kimia Kahrizi; Marzieh Mohseni; Seyed Jalal Kiani; Tahereh Donyavi; Zohreh Fattahi |
| EPI_ISL_1697387 | Iressef / LBV | Iressef Genomics lab | Abdou PADANE; Adjiratou Aissatou BA; Alioune TINE; Ambroise AHOUIDI; Aminata DIA; Aminata MBOUP; Anna Julienne Selbe NDIAYE; Assane DIENG; Astou Gaye GAYE; Barada CISSE; Birahim Piere NDIAYE; Gora LO; Khadim GUEYE; Makhtar Camara; Mengue FALL; Moustapha MBOW; Nafisatou LEYE; Ndeye Coumba Toure KANE; OUMY DIOP; Papa Alassane DIAW; Pauline SENE; SADA DIALLO; Serigne Mbaye Lo NDIAYE; Souleymane MBOUP; Yacine DIA |
| EPI_ISL_2620888, EPI_ISL_2620889, EPI_ISL_2873822, EPI_ISL_2873823, EPI_ISL_2873828, EPI_ISL_2873829, EPI_ISL_2873833, EPI_ISL_2873834, EPI_ISL_2873835, EPI_ISL_2887849, EPI_ISL_2887851, EPI_ISL_2887852, EPI_ISL_2887857 | | | |
| see above | Iressef Genomics lab | IRESSEF | Abdou PADANE; Ambroise AHOUIDI; Aminata DIA; Aminata MBOUP; Astou Gaye GAYE; Barada CISSE; Birahim Piere NDIAYE; Diabou Diagne; Gora LO; Khadim GUEYE; Moustapha MBOW; Nafisatou LEYE; Ndeye Coumba Toure KANE; Papa Alassane DIAW; Samba Ndiour; Seni Ndiaye; Souleymane MBOUP; Yacine DIA |
| EPI_ISL_1167146 | Iressef Genomics lab | L'institut de Recherche en Santé, de Surveillance Épidémiologique et de Formation (IRESSEF) | Abdou PADANE; Abdoulie KANTEH; Abdul Karim SESAY; Ambroise AHOUIDI; Aminata DIA; Aminata MBOUP; Astou Gaye GAYE; Barada CISSE; Birahim Piere NDIAYE; Gora LO; Khadim GUEYE; Moustapha MBOW; Nafisatou LEYE; Ndeye Coumba Toure KANE; Papa Alassane DIAW; Souleymane MBOUP; Yacine DIA |
| EPI_ISL_2779373 | Isebania Sub County Hospital | USAMRD-A, Basic Science Laboratory | Alan Lemtudo; Beth Mutai; Brian Andika; Carol Kifude; Clement Masakwe; Eric Muthanje; Esther Omuseni; Faith Sigei; Gathii Kimita; George Awinda; John Waitumbi; Josphat Nyataya; Rachel Githii; Rehema Liyai; Stephen Ochola |
| EPI_ISL_419299, EPI_ISL_419300 | Ishikawa Prefectural Institute of Public Health and Environmental Science | Pathogen Genomics Center, National Institute of Infectious Diseases | Eri Nariai; Hajime Kamiya; Kentaro Itokawa; Makoto Kuroda; Masanori Hashino; Motoi Suzuki; Rina Tanaka; Sanae Kuramoto; Tsuyoshi Sekizuka |
| EPI_ISL_2505234, EPI_ISL_2508151, EPI_ISL_2508154 | Islab, Pohjois-Savon aluelaboratorio | Expert Microbiology, National Institute for Health and Welfare | Carita Savolainen-Kopra; Erika Lindh; Haider al-Hello; Jani Halkilahti; Kirsi Liitsola; Niina Ikonen; Olli Vapalahti; Pekka Ellonen; Phuoc Truong; Päivi Laurila; Ravi Kant; Sari Hannula; Soile Blomqvist; Teemu Smura |
| EPI_ISL_1240650 | Israel Central Virology Laboratory | Israel National Consortium for SARS-CoV-2 sequencing | Assaf Rokney; Dana Bar-Ilan; David A. Zeevi; Efrat Dahan Bucris; Efrat Glick-Saar; Efrat Rorman; Ella Mendelson; Ephraim Fass; Eva Nachum; Gal Zizelski Valenci; Gideon Rechavi; Israel Nissan; Joseph Jaffe; Maya Davidovich Cohen; Michal Mandelboim; Mor Rubinstein; Neta Zuckerman; Omer Murik; Omri Nayshool; Oran Erster; Orna Mor; Tzvia Mann |
| EPI_ISL_474989, EPI_ISL_516889, EPI_ISL_516900, EPI_ISL_575334, EPI_ISL_649094, EPI_ISL_776862 | Israel Central Virology laboratory | Israel Central Virology laboratory | Efrat Dahan Bucris; Ella Mendelson; Michal Mandelboim; Neta Zuckerman; Oran Erster; Orna Mor |
| EPI_ISL_944436, EPI_ISL_1073463, EPI_ISL_1209484, EPI_ISL_1209811, EPI_ISL_2084501, EPI_ISL_2085005, EPI_ISL_2085242, EPI_ISL_2085776, EPI_ISL_2086077, EPI_ISL_2182466, EPI_ISL_2183616, EPI_ISL_2183687, EPI_ISL_2636241 | | | |
| see above | Israel Central Virology laboratory | Israel National Consortium for SARS-CoV-2 sequencing | Assaf Rokney; Dana Bar-Ilan; David A. Zeevi; Efrat Dahan Bucris; Efrat Glick-Saar; Efrat Rorman; Ella Mendelson; Ephraim Fass; Eva Nachum; Gal Zizelski Valenci; Gideon Rechavi; Israel Nissan; Joseph Jaffe; Maya Davidovich Cohen; Michal Mandelboim; Miranda Geva; Mor Rubinstein; Neta Zuckerman; Netanel Abu; Omer Murik; Omri Nayshool; Oran Erster; Orna Mor; Tzvia Mann |
| EPI_ISL_1670660 | Istituto Zooprofilattico Sperimentale Umbria e Marche "Togo Rosati" | Istituto Zooprofilattico Sperimentale dell'Abruzzo e Molise "G. Caporale" | Ancora M; Biagetti M; Calistri P; Cammà C; Caporale M; Curini V; Delli Compagni E; Di Domenico M; Di Lollo Valeria; Di Pasquale A; Giammarioli M; Lorusso A; Mangone I; Marcacci M; Puglia I; Rinaldi A; Savini G; Scialabba S |
| EPI_ISL_837247, EPI_ISL_1039312, EPI_ISL_1759528, EPI_ISL_2227133, | Istituto Zooprofilattico Sperimentale del Mezzogiorno | TIGEM | Andrea Ballabio; Anna Manfredi; Antonio Grimaldi; Antonio Grimaldi Patrizia Annunziata Francesco Panariello Biancamaria Pierri Claudia Tiberio Teresa Giuliano Valentina Bouche Chiara Colantuono Maria Concetta Cuomo Denise Di Concilio Lucio Di Filippo Anna Manfredi Marcello Salvi Antonio Limone Luigi Atripaldi Pellegrino Cerino Andrea Ballabio Davide Cacchiarelli; Antonio Limone; Biancamaria Pierri; Chiara Colantuono; Davide Cacchiarelli.; Denise Di Concilio; Francesco Panariello; Lucio Di Filippo; Marcello Salvi; Maria Concetta Cuomo; Patrizia Annunziata; Pellegrino Cerino; Valentina Bouche |

| | | | |
|---|---|---|---|
| EPI_ISL_2455775 | | | |
| EPI_ISL_1632906, EPI_ISL_2143715, EPI_ISL_2144052 | Istituto Zooprofilattico Sperimentale del Mezzogiorno | Telethon Institute of Genetics and Medicine (TIGEM) | Antonio Grimaldi Patrizia Annunziata Francesco Panariello Biancamaria Pierri Claudia Tiberio Teresa Giuliano Valentina Bouche Chiara Colantuono Maria Concetta Cuomo Denise Di Concilio Lucio Di Filippo Anna Manfredi Marcello Salvi Antonio Limone Luigi Atripaldi Pellegrino Cerino Andrea Ballabio Davide Cacchiarelli |
| EPI_ISL_1390714 | Istituto Zooprofilattico Sperimentale del Mezzogiorno - Azienda Ospedaliera "Pugliese Ciaccio" di Catanzaro | TIGEM | Andrea Ballabio; Anna Manfredi; Antonio Grimaldi; Antonio Limone Pasquale Minchella Luigi Atripaldi Pellegrino Cerino; Biancamaria Pierri Claudia Tiberio Valentina Bouche; Chiara Colantuono; Davide Cacchiarelli; Denise Di Concilio; Francesco Panariello; Lucio Di Filippo; Marcello Salvi; Maria Concetta Cuomo; Patrizia Annunziata |
| EPI_ISL_1298481 | Ivano-Frankivsk CCH#1 | The Institute of Molecular Biology and Genetics of NASU | M.Tukalo et al. |
| EPI_ISL_2272918 | JIPMER Puducherry | inStem NCBS - INSACOG | Uma Ramakrishnan Dasaradhi Palakodeti Aswin SaiNarain |
| EPI_ISL_2844844 | Jaber Quarantine | Virology Lab, Jaber Al Ahmad Al Sabah Hospital | Adil Afridi; Dr. Ebaa Al-Awadhi; Dr. Zahrah Buhamad; Haroon Masih |
| EPI_ISL_2984226, EPI_ISL_2984232, EPI_ISL_2984244 | Jahra Hospital | Kuwait Cancer Control Center | Mona Alateeqi; Shakir Bahzad |
| EPI_ISL_450793 | Jamaica Ministry of Health and Wellness | Pathogen Discovery, Respiratory Viruses Branch, Division of Viral Diseases, Centers for Disease Control and Prevention | Anna Montmayeur; Anna Uehara; Bettina Bankamp; Clinton R. Paden; Haibin Wang; Jing Zhang; Krista Queen; Rachel Marine; Suxiang Tong; Yan Li; Ying Tao; Zachary Weiner |
| EPI_ISL_779255, EPI_ISL_779261, EPI_ISL_779263, EPI_ISL_779281, EPI_ISL_779283, EPI_ISL_779284, EPI_ISL_2151345, EPI_ISL_2151346, EPI_ISL_2151365 | | | |
| see above | Jamil-ur-Rahman Center for Genome Research, Dr. Panjwani Center for Molecular Medicine and Drug Research | Jamil-ur-Rahman Center for Genome Research, Dr. Panjwani Center for Molecular Medicine and Drug Research | A.H.; Ain, N.; Ansari, S.; Hashmi; I.A.; Irfan, M.; Khan; Khan, I.; Nisa, Z.; Rashid, M.; Shakeel, M.; Siddiqui, S. |
| EPI_ISL_2662827 | Jamil-ur-Rahman Center for Genome Research, Dr. Panjwani Center for Molecular Medicine and Drug Research, International Center for Chemical and Biological Sciences, University of Karachi | Jamil-ur-Rahman Center for Genome Research, Dr. Panjwani Center for Molecular Medicine and Drug Research, International Center for Chemical and Biological Sciences, University of Karachi | Ain, Nu.; B.T.; I.A.; Iqbal, W.; Irfan, M.; Jahan, S.; Javed; Kakar, N.; Khan, S.; Nisa, Zu.; Rehman, Z.; S. and Khan; Sarwar, B.; Shakeel, M.; Siddiqi; Zehra, M. |
| EPI_ISL_2779319, EPI_ISL_2779320, EPI_ISL_3031392, EPI_ISL_3031395, EPI_ISL_3031423, EPI_ISL_3031426, EPI_ISL_3031427, EPI_ISL_3031429, EPI_ISL_3031432, EPI_ISL_3031433 | | | |
| see above | Jaramogi Oginga Odinga Teaching and Referral Hospital | USAMRD-A, Basic Science Laboratory | Alan Lemtudo; Beth Mutai; Brian Andika; Carol Kifude; Clement Masakwe; Eric Muthanje; Esther Omuseni; Faith Sigei; Gathii Kimita; George Awinda; John Waitumbi; Josphat Nyataya; Rachel Githii; Rehema Liyai; Stephen Ochola |
| EPI_ISL_3039592 | Jessa | Jessa | Berden et al. on behalf of the Jessa_cmdLab |
| EPI_ISL_2248928 | Joaquim Chaves Saude | Instituto Nacional de Saude (INSA) | Borges et al |
| EPI_ISL_2885164 | Johns Hopkins Hospital Department of Pathology | Johns Hopkins Hospital Department of Pathology | Adannaya Amadi; C. Paul Morris; Chun Huai Luo; Heba H. Mostafa; Matthew Schwartz; Nicholas Gallagher |
| EPI_ISL_2932611, EPI_ISL_2932615, EPI_ISL_2932618 | Jordan University of Science Technology/ Irbid filed Hospital/ MOH | Princess Haya Biotechnology Center/ Jordan University of Science Technology | Areej Alquran; Emad Abu Yaqeen; Hazem Haddad; Maha Karam; Saied Jaradat; Shereen Issa; Suha Hasan |
| EPI_ISL_2271955 | Joubert Clinic | National Institute for Communicable Diseases of the National Health Laboratory Service | Amoako DG; Bhiman JN; Ismail A; Mahlangu B; Mohale T; Ntuli N; Scheepers C |
| EPI_ISL_2779335 | KCRH KEMRI-DEID Surveillance Site | USAMRD-A, Basic Science Laboratory | Alan Lemtudo; Beth Mutai; Brian Andika; Carol Kifude; Clement Masakwe; Eric Muthanje; Esther Omuseni; Faith Sigei; Gathii Kimita; George Awinda; John Waitumbi; Josphat Nyataya; Rachel Githii; Rehema Liyai; Stephen Ochola |
| EPI_ISL_2779337, EPI_ISL_2779455, EPI_ISL_2779456, EPI_ISL_2779547, EPI_ISL_3031383, EPI_ISL_3031385, EPI_ISL_3031407, EPI_ISL_3031413, EPI_ISL_3031417 | | | |
| see above | KCRH KEMRI/DEID Surveillance Site | USAMRD-A, Basic Science Laboratory | Alan Lemtudo; Beth Mutai; Brian Andika; Carol Kifude; Clement Masakwe; Eric Muthanje; Esther Omuseni; Faith Sigei; Gathii Kimita; George Awinda; John Waitumbi; Josphat Nyataya; Rachel Githii; Rehema Liyai; Stephen Ochola |
| EPI_ISL_2348630 | KEMRI | KEMRI & ICGEB | Alessandro Marcello; Emanuele Orsini; Eric A Lelo; Janet Majanja; Samwel L Symekher; Silvanos M Opanda; Wadegu Meshack; Wallace D Bulimo |
| EPI_ISL_2602986, EPI_ISL_2602988, EPI_ISL_2602989, EPI_ISL_2602990, EPI_ISL_2602991, EPI_ISL_2602992 | KEMRI Center for Biotechnology Research and Development | KEMRI-Wellcome Trust Research Programme,Kilifi | ; Githinji G.; Matoke D.; Mburu M.W.; Mohamed K.S.; Onyango C.; de Laurent Z. |
| EPI_ISL_2602781, EPI_ISL_2602794, EPI_ISL_2602797, EPI_ISL_2602799, EPI_ISL_2602807, EPI_ISL_2602814, EPI_ISL_2602816, EPI_ISL_2602817, EPI_ISL_2602822, EPI_ISL_2602838, EPI_ISL_2602856, EPI_ISL_2602877, EPI_ISL_2602891, EPI_ISL_2602918, EPI_ISL_2602919, EPI_ISL_2602920, EPI_ISL_2602923, EPI_ISL_2602925, EPI_ISL_2602927, EPI_ISL_2602930, EPI_ISL_2602932, EPI_ISL_2602933, EPI_ISL_2602938, EPI_ISL_2602944, EPI_ISL_2602945, EPI_ISL_2602946, EPI_ISL_2602947, EPI_ISL_2602949, EPI_ISL_2602951 | | | |
| see above | KEMRI-CGHR | KEMRI-Wellcome Trust Research Programme,Kilifi | ; Githinji G.; Matoke D.; Mburu M.W.; Mohamed K.S.; Onyango C.; de Laurent Z. |
| EPI_ISL_457866, EPI_ISL_457876, EPI_ISL_457906 | KEMRI-CGMR-C | KEMRI-Wellcome Trust Research Programme/KEMRI-CGMR-C Kilifi | Githinji G. et al 2020 |
| EPI_ISL_1440093, EPI_ISL_1440096, EPI_ISL_1440106, EPI_ISL_1440115, EPI_ISL_1440118, EPI_ISL_1440122, EPI_ISL_1440257, EPI_ISL_1440319, EPI_ISL_1610444, EPI_ISL_1610516, EPI_ISL_1610567, EPI_ISL_2602477, EPI_ISL_2602480, EPI_ISL_2602508, EPI_ISL_2602519, EPI_ISL_2602520, EPI_ISL_2602521, EPI_ISL_2602522, EPI_ISL_2602527, EPI_ISL_2602528, EPI_ISL_2602529, EPI_ISL_2602531, EPI_ISL_2602532, EPI_ISL_2602534, EPI_ISL_2602549, EPI_ISL_2602553, EPI_ISL_2602557, EPI_ISL_2602559, EPI_ISL_2602560, EPI_ISL_2602566, EPI_ISL_2602569, EPI_ISL_2602570, EPI_ISL_2602571, EPI_ISL_2602572, EPI_ISL_2602583, EPI_ISL_2602594, EPI_ISL_2602596, EPI_ISL_2602597, EPI_ISL_2602598, EPI_ISL_2602602, EPI_ISL_2602605, EPI_ISL_2602607, EPI_ISL_2602610, EPI_ISL_2602612, EPI_ISL_2602620, EPI_ISL_2602625, EPI_ISL_2602635, EPI_ISL_2602637, EPI_ISL_2602643, EPI_ISL_2602644, EPI_ISL_2602647, EPI_ISL_2602650, EPI_ISL_2602657, EPI_ISL_2602659, EPI_ISL_2602660, EPI_ISL_2602661, EPI_ISL_2602662, EPI_ISL_2602663, EPI_ISL_2602667, EPI_ISL_2602674, EPI_ISL_2602681, EPI_ISL_2602683, EPI_ISL_2602684, EPI_ISL_2602685, EPI_ISL_2602686, EPI_ISL_2602687, EPI_ISL_2602688, EPI_ISL_2602689, EPI_ISL_2602695, EPI_ISL_2602696, EPI_ISL_2602697, EPI_ISL_2602698, EPI_ISL_2602699, EPI_ISL_2602700, EPI_ISL_2602701, EPI_ISL_2602703, EPI_ISL_2602706, EPI_ISL_2602713, EPI_ISL_2602714, EPI_ISL_2602716, EPI_ISL_2602723, EPI_ISL_2602730, EPI_ISL_2602745, EPI_ISL_2602748, EPI_ISL_2602749, EPI_ISL_2602750, EPI_ISL_2602751, EPI_ISL_2602753, EPI_ISL_2602754, EPI_ISL_2602762, EPI_ISL_2602763, EPI_ISL_2602996, EPI_ISL_2603012, EPI_ISL_2603021, EPI_ISL_2603036, EPI_ISL_2603038, EPI_ISL_2603039, EPI_ISL_2603040, EPI_ISL_2603043, EPI_ISL_2603044, EPI_ISL_2603045, EPI_ISL_2603047, EPI_ISL_2603048, EPI_ISL_2603049, EPI_ISL_2603050, EPI_ISL_2603052, EPI_ISL_2603053, EPI_ISL_2603054, EPI_ISL_2603055, EPI_ISL_2603056, EPI_ISL_2603057, EPI_ISL_2603058, EPI_ISL_2603059, EPI_ISL_2603061, EPI_ISL_2603063, EPI_ISL_2603065, EPI_ISL_2603066, EPI_ISL_2603067, EPI_ISL_2603068, EPI_ISL_2603069, EPI_ISL_2603070, EPI_ISL_2603071, EPI_ISL_2603072, EPI_ISL_2603074, EPI_ISL_2603075, EPI_ISL_2603078, EPI_ISL_2603079, EPI_ISL_2603082, EPI_ISL_2603085, EPI_ISL_2603086, EPI_ISL_2603088, EPI_ISL_2603089, EPI_ISL_2603091, EPI_ISL_2603093, EPI_ISL_2603095, EPI_ISL_2603096, EPI_ISL_2603097, EPI_ISL_2603099, EPI_ISL_2603103, EPI_ISL_2603104, EPI_ISL_2603105, EPI_ISL_2603106, EPI_ISL_2603108, EPI_ISL_2603111, EPI_ISL_2603113, EPI_ISL_2603115, EPI_ISL_2603118, EPI_ISL_2603119, EPI_ISL_2603120, EPI_ISL_2603121 | | | |
| see above | KEMRI-Wellcome Trust Research Programme,Kilifi | KEMRI-Wellcome Trust Research Programme,Kilifi | ; Githinji G.; Matoke D.; Mburu M.W.; Mohamed K.S.; Onyango C.; Thiongo K.; de Laurent Z.; deLaurent Z. |
| EPI_ISL_568715, EPI_ISL_568863, EPI_ISL_806550, EPI_ISL_806561, EPI_ISL_806574, EPI_ISL_806617, EPI_ISL_806638, EPI_ISL_806643, EPI_ISL_806653, EPI_ISL_806667, EPI_ISL_806704, EPI_ISL_806705, EPI_ISL_855536, EPI_ISL_855537, EPI_ISL_855544, EPI_ISL_855548, EPI_ISL_968817, EPI_ISL_968822, EPI_ISL_968850, EPI_ISL_968922, EPI_ISL_968961, EPI_ISL_968992, EPI_ISL_968993, EPI_ISL_969014, EPI_ISL_969074, EPI_ISL_1039226, EPI_ISL_1039228 | | | |
| see above | KEMRI-Wellcome Trust Research Programme/KEMRI-CGMR-C Kilifi | KEMRI-Wellcome Trust Research Programme/KEMRI-CGMR-C Kilifi | Githinji et al; Githinji et al 2020 |
| EPI_ISL_2779322, EPI_ISL_2779323 | KEMRI/DEID Surveillance Site (Isebani Sub-County Hospital) | USAMRD-A, Basic Science Laboratory | Alan Lemtudo; Beth Mutai; Brian Andika; Carol Kifude; Clement Masakwe; Eric Muthanje; Esther Omuseni; Faith Sigei; Gathii Kimita; George Awinda; John Waitumbi; Josphat Nyataya; Rachel Githii; Rehema Liyai; Stephen Ochola |
| EPI_ISL_3031404, EPI_ISL_3031405 | KEMRI/DEID Surveillance Site (Kombewa Sub County Hospital) | USAMRD-A, Basic Science Laboratory | Alan Lemtudo; Beth Mutai; Brian Andika; Carol Kifude; Clement Masakwe; Eric Muthanje; Esther Omuseni; Faith Sigei; Gathii Kimita; George Awinda; John Waitumbi; Josphat Nyataya; Rachel Githii; Rehema Liyai; Stephen Ochola |
| EPI_ISL_2779325, EPI_ISL_2779348 | KEMRI/DEID Surveillance Site (Lamu County Hospital) | USAMRD-A, Basic Science Laboratory | Alan Lemtudo; Beth Mutai; Brian Andika; Carol Kifude; Clement Masakwe; Eric Muthanje; Esther Omuseni; Faith Sigei; Gathii Kimita; George Awinda; John Waitumbi; Josphat Nyataya; Rachel Githii; Rehema Liyai; Stephen Ochola |
| EPI_ISL_2779362, EPI_ISL_2779376 | KEMRI/DEID Surveillance Site (Mtongwe Navy Base Hospital) | USAMRD-A, Basic Science Laboratory | Alan Lemtudo; Beth Mutai; Brian Andika; Carol Kifude; Clement Masakwe; Eric Muthanje; Esther Omuseni; Faith Sigei; Gathii Kimita; George Awinda; John Waitumbi; Josphat Nyataya; Rachel Githii; Rehema Liyai; Stephen Ochola |
| EPI_ISL_2382194, EPI_ISL_2382195, EPI_ISL_2382197, EPI_ISL_2382198, EPI_ISL_2382200, EPI_ISL_2382201, EPI_ISL_2382202, EPI_ISL_2382203, EPI_ISL_2382204, EPI_ISL_2382205, EPI_ISL_2382209, EPI_ISL_2382210, EPI_ISL_2382214, EPI_ISL_2382217, EPI_ISL_2382702, EPI_ISL_2382710, EPI_ISL_2382715, EPI_ISL_2382720, EPI_ISL_2382725, EPI_ISL_2582845, EPI_ISL_2582848, EPI_ISL_2582868, EPI_ISL_2582870, EPI_ISL_2582872, EPI_ISL_2582885, EPI_ISL_2582963, EPI_ISL_2582977, EPI_ISL_2662665, EPI_ISL_2662678, EPI_ISL_2662685, EPI_ISL_2662699, EPI_ISL_2662700, EPI_ISL_2984802, EPI_ISL_2984803, EPI_ISL_2984804, EPI_ISL_2984805, EPI_ISL_2984806, EPI_ISL_2984807, EPI_ISL_2984808, EPI_ISL_2984809 | | | |
| see above | KIMBERLEY LABORATORY | National Institute for Communicable Diseases of the National Health Laboratory Service | Amoako DG; Bhiman JN; Everatt J; Ismail A; Mahlangu B; Mnguni A; Mohale T; Ntuli N; Scheepers C |
| EPI_ISL_436684, EPI_ISL_455631 | KRISP, KZN Research Innovation and Sequencing Platform | KRISP, KZN Research Innovation and Sequencing Platform | Chimukangara B; Deforche K; Giandhari J; Lessells R; Pillay S; Tegally H; Wilkinson E; de Oliveira T |
| EPI_ISL_2384202, EPI_ISL_2690054 | KS Health and Environmental Laboratories | Centers for Disease Control and Prevention Division of Viral Diseases, Pathogen Discovery | Alison Laufer Halpin; Ben L. Rambo-Martin; Clinton R. Paden; Dakota Howard; Darlene Wagner; Dave Wentworth; Dhwani Batra; Jasmine Padilla; Justin Lee; Katie Dillon; Krista Queen; Kristen Knipe; Kristine Lacek; Mark Burroughs; Matthew Schmerer; Mili Sheth; Peter Cook; Sam Shepard; Sarah Nobles; Shoshona Le; Suxiang Tong; Vivien Dugan; Yvette Unoarumhi |
| EPI_ISL_1048849, EPI_ISL_2424511, EPI_ISL_2425042, EPI_ISL_2425316, EPI_ISL_2833359 | KU Leuven, Rega Institute, Clinical and Epidemiological Virology | KU Leuven, Rega Institute, Clinical and Epidemiological Virology | Bert Vanmechelen; Joan Marti-Carerras; Piet Maes; Tony Wawina-Bokalanga |
| EPI_ISL_1446361 | KY State Public Health Lab | Centers for Disease Control and Prevention Division of Viral Diseases, Pathogen Discovery | Alison Laufer Halpin; Ben L. Rambo-Martin; Clinton R. Paden; Dakota Howard; Darlene Wagner; Dave Wentworth; Dhwani Batra; Jasmine Padilla; Justin Lee; Katie Dillon; Krista Queen; Kristen Knipe; Kristine Lacek; Mark Burroughs; Matthew Schmerer; Mili Sheth; Peter Cook; Sam Shepard; Sarah Nobles; Shoshona Le; Suxiang Tong; Vivien Dugan; Yvette Unoarumhi |
| EPI_ISL_495421 | Kafkas University, Faculty of Medicine, Department of Medical Microbiology | Kafkas University, Faculty of Medicine, Department of Medical Microbiology | Didem Ozgur; E. Ediz Tutuncu; Murat Karamese |
| EPI_ISL_1412522, EPI_ISL_2153683, EPI_ISL_2627909, EPI_ISL_2688192, EPI_ISL_2833876, EPI_ISL_2842088, EPI_ISL_3030691 | | | |
| see above | Kansas Health and Environmental Lab | Kansas Health and Environmental Lab | Ben Olsen; Jonathan Barnell; Katherine Wiggins; Mike Grose; and Phil Adam |
| EPI_ISL_2438618 | Karasai Central District Hospital | Reference laboratory for the control of viral infections | Aidar Usserbayev; Aknur Mutaliyeva; Andrey Komissarov; Artem Fadeev; Azamat Kenessov; Bekzhan Maikotov; Gaukhar Nussupbayeva; Madina Tleubergenova; Maria Pisareva; Nazym Tleumbetova |
| EPI_ISL_2365354, EPI_ISL_2365857, EPI_ISL_2835993 | Kariminejad-Najmabadi Pathology & Genetics Center, Tehran, Iran | Genetics Research Center, University of Social Welfare and Rehabilitation Sciences | Hossein Najmabadi.; Kimia Kahrizi; Mahdieh Koshki; Maryam Azad; Marzieh Mohseni; Siavash Ghaderi; Zohreh Fattahi |

| | | | |
|---|---|---|---|
| EPI_ISL_475544, EPI_ISL_510835 | Karolinska Universitetslaboratoriet | The Public Health Agency of Sweden | Anna Risberg; Anna-Malin Linde; Karin Tegmark-Wisell; Maria Lind Karlberg; Mattias Haukland; Mia Brytting; Olov Svartstrom; Oskar Karlsson Lindsjo; Petra Edquist; Reza Advani; Sandra Broddesson; Shaman Muradrasoli |
| EPI_ISL_2648222, EPI_ISL_2648231, EPI_ISL_2648235, EPI_ISL_2648238, EPI_ISL_2820291, EPI_ISL_2820294, EPI_ISL_2820302, EPI_ISL_2820303, EPI_ISL_2820309, EPI_ISL_2820310, EPI_ISL_2820316, EPI_ISL_2820322, EPI_ISL_2820329, EPI_ISL_2820330, EPI_ISL_2820381, EPI_ISL_2820385, EPI_ISL_2820396, EPI_ISL_2820402, EPI_ISL_2820411, EPI_ISL_2820412 | | | |
| see above | Kasane Primary Hospital Laboratory | Botswana Harvard HIV Reference Laboratory | Agnes Karutwaeng; Boitumelo Zuze; Botshelo Radibe; Dorcas Maruapula; Joseph Makhema; Keoratile Ntshambiwa; Legodile Kooepile; Madisa Mine; Mosepele Mosepele; Ontlametse T. Bareng; Roger Shapiro; Rose Munyere; Shahin Lockman; Sikhulile Moyo; Simani Gaseitsiwe; Thongbotho Mphoyakgosi; Wonderful T. Choga |
| EPI_ISL_754063 | Kathmandu University | Nepal Health Research Council | Meghnath Dhimal; Pradip Gyanwali |
| EPI_ISL_1859725 | Kazan Federal University, Academic Consortium | FEDERAL RESEARCH AND CLINICAL CENTER OF PHYSICAL-CHEMICAL MEDICINE OF FEDERAL MEDICAL BIOLOGICAL AGENCY, Academic Consortium | Alexander Manolov; Alexander Pavlenko; Alexander Rumyantsev; Alexander Samoilov; Alexander Troitsky; Alexander Tulichev; Alexander Vasiliev; Alexey Volnukhin; Alla Petrova; Andrey Koritsky; Anna Blagonravova; Anna Varizhuk; Daria Boldyreva; Daria Kharlampieva; Daria Matyushkina; Dmitry Fedorov; Dmitry Pushkar; Ekaterina Moskaleva; Elena Galova; Elena Ilina; Elizaveta Orlova; Galina Trifonova; Gaukhar Yusubalieva; Ilshat Gafurov; Irina Shirokova; Konstantin Ivanov; Konstantin Kolontarev; Ksenia Klimina; Larisa Balykova; Lyubov Rychkova; Nadezhda Kolyshkina; Natalia Pozhenko; Oleg Ogarkov; Pavel Bashkirov; Polina Khromova; Polina Tikhonova; Ramiz Bakhtyev; Raushaniya Gaifullina; Sergey Rumyantsev; Svetlana Solovieva; Vadim Govorun; Valentin Manuvera; Varvara Musarova; Vasily Lazarev; Vladimir Baklaushev; Vladimir Rumyantsev; Vladimir Veselovsky; Vladislav Babenko; Vyacheslav Kuropatkin; Vyacheslav Sinkov; Yulia Galeeva; Yuri Udalov |
| EPI_ISL_1184116 | Kentucky State Public Health Lab | Kentucky State Public Health Lab | Joshua Tobias; Karim George; Matthew Johnson; Rachel Zinner; Rhonda Lucas; Stephanie Lunn; Vaneet Arora; William Grooms |
| EPI_ISL_1239232, EPI_ISL_1239238, EPI_ISL_1239247 | Kimberley Hospital, National Health Laboratory Services, Northern Cape, South Africa | National Institute for Communicable Diseases of the National Health Laboratory Service | Amoako DG; Bhiman JN; Ismail A; Mahlangu B; Mohale T; Ntuli N; Scheepers C |
| EPI_ISL_3031398 | King'ong'o Dispensary | USAMRD-A, Basic Science Laboratory | Alan Lemtudo; Beth Mutai; Brian Andika; Carol Kifude; Clement Masakwe; Eric Muthanje; Esther Omuseni; Faith Sigei; Gathii Kimita; George Awinda; John Waitumbi; Josphat Nyataya; Rachel Githii; Rehema Liyai; Stephen Ochola |
| EPI_ISL_483550, EPI_ISL_483551 | Kingdom of Bahrein Ministry of Health | Erasmus Medical Center | Amjad Ghanem Mohamed; Anne van der Linden; Bas Oude Munnink; Claudia Schapendonk; David Nieuwenhuijse; Ebrahim Shehad; Fatema; Hashmeya Al Wasti; Irina Chestakova; Marion Koopmans; Mark Pronk; Pascal Lexmond; Reina Sikkema; Richard Molenkamp; Stefan van Nieuwkoop; Theo Bestebroer; on behalf of the Dutch national COVID-19 response team. |
| EPI_ISL_1510532 | Klaipeda University Hospital | Vilnius University Hospital Santaros Klinikos, Center of Laboratory Medicine | Daniel Naumovas; Dovile Ezerskyte; Gytis Dudas; Ingrida Olendraite; Laimonas Griskevicius; Ligita Raugaite; Mindaugas Stoskus; Monika Katenaite; Rimvydas Norvilas |
| EPI_ISL_2693464 | Klinika za infektivne bolesti "Dr. Fran Mihaljević" | Hrvatski zavod za javno zdravstvo | Irena Tabain; Ivana Ferenčak |
| EPI_ISL_654892 | Klinisk mikrobiologi | The Public Health Agency of Sweden | Anna Risberg; Anna-Malin Linde; Karin Tegmark-Wisell; Maria Lind Karlberg; Mattias Haukland; Mia Brytting; Olov Svartstrom; Oskar Karlsson Lindsjo; Petra Edquist; Reza Advani; Sandra Broddesson |
| EPI_ISL_548962 | Klinisk mikrobiologi, Region Västerbotten | Unit for Biological Agents, Department for CBRN Defence and Security, Swedish Defence Research Agency | FOI Bioinformatics team |
| EPI_ISL_419296 | Kochi Prefectural Institute of Public Health | Pathogen Genomics Center, National Institute of Infectious Diseases | Akihiko Tokaji; Hajime Kamiya; Kentaro Itokawa; Makoto Kuroda; Masanori Hashino; Motoi Suzuki; Rina Tanaka; Tsuyoshi Sekizuka |
| EPI_ISL_2815331 | Kuala Lumpur General Hospital | Institute for Medical Research, Infectious Disease Research Centre, National Institutes of Health, Ministry of Health Malaysia | Azizan MA; Kamel K; Mohd Zawawi Z; Ramly N; Robert F; Suppiah J; Thayan R |
| EPI_ISL_2649991 | Kudat Hospital | Institute for Medical Research, Infectious Disease Research Centre, National Institutes of Health, Ministry of Health Malaysia | Azizan MA; Kamel K; Mohd Zawawi Z; Ramly N; Robert F; Suppiah J; Thayan R |
| EPI_ISL_2689954, EPI_ISL_2689960 | LA Office of Public Health Laboratories | Centers for Disease Control and Prevention Division of Viral Diseases, Pathogen Discovery | Alison Laufer Halpin; Ben L. Rambo-Martin; Clinton R. Paden; Dakota Howard; Darlene Wagner; Dave Wentworth; Dhwani Batra; Jasmine Padilla; Justin Lee; Katie Dillon; Krista Queen; Kristen Knipe; Kristine Lacek; Mark Burroughs; Matthew Schmerer; Mili Sheth; Peter Cook; Sam Shepard; Sarah Nobles; Shoshona Le; Suxiang Tong; Vivien Dugan; Yvette Unoarumhi |
| EPI_ISL_2779525 | LAB-BMC | USAMRD-A, Basic Science Laboratory | Alan Lemtudo; Beth Mutai; Brian Andika; Carol Kifude; Clement Masakwe; Eric Muthanje; Esther Omuseni; Faith Sigei; Gathii Kimita; George Awinda; John Waitumbi; Josphat Nyataya; Rachel Githii; Rehema Liyai; Stephen Ochola |
| EPI_ISL_2893835 | LABORATOIRE FORTE BIO ST PAUL LES DAX | CNR Virus des Infections Respiratoires - France SUD | Antonin Bal; Bruno Lina; Gregory Destras; Gwendolyne Burfin; Hadrien Regue; Laurence Josset; Martine Valette; Quentin Semanas |
| EPI_ISL_2278210 | LABORATOIRE GCS INTER HOSPITALIER | CHU Purpan - Laboratoire de Virologie - Institut Fédératif de Biologie | Bulach T.; Donnadieu C.; Izopet J.; Latour J.; Milhes M.; Nicot F.; Ranger N.; Salin G.; Tremeaux P. |
| EPI_ISL_2676632 | LABORATOIRE d'ANALYSES de BIOLOGIE MEDICALES | CNR Virus des Infections Respiratoires - France SUD | Antonin Bal; Bruno Lina; Gregory Destras; Gwendolyne Burfin; Hadrien Regue; Laurence Josset; Martine Valette; Quentin Semanas |
| EPI_ISL_2657871 | LABORATORIO CLINICA IMBANACO | Instituto Nacional de Salud- Dirección de Investigación en Salud Pública | Carlos Franco-Muñoz; Carmen Osorio; Diana Malo; Diego A. Álvarez-Díaz; Diego Andrés Prada; Gerardo Santamaría; Hector Alejandro Ruiz-Moreno; Jhonnatan Reales-González; Jorge Rivera; Juan Camilo Martinez; Julian Naizaque; Katherine Laiton-Donato; Lisseth Pardo; Magdalena Wiesner; Marcela Mercado-Reyes; Maria T. Herrera-Sepúlveda; Marta Lopez Blanco; Martha Lucia Ospina Martinez; Paola Rojas; Sergio Gomez; Sheryll Corchuelo; Ángela Alarcon Cruz |
| EPI_ISL_2834727 | LABORATORIO CLINICO HIGUERA ESCALANTE | Instituto Nacional de Salud- Dirección de Investigación en Salud Pública | Carlos Franco-Muñoz; Carmen Osorio; Diana Malo; Diego A. Álvarez-Díaz; Diego Andrés Prada; Gerardo Santamaría; Hector Alejandro Ruiz-Moreno; Jhonnatan Reales-González; Jorge Rivera; Juan Camilo Martinez; Julian Naizaque; Katherine Laiton-Donato; Lisseth Pardo; Magdalena Wiesner; Marcela Mercado-Reyes; Maria T. Herrera-Sepúlveda; Marta Lopez Blanco; Martha Lucia Ospina Martinez; Paola Rojas; Sergio Gomez; Sheryll Corchuelo; Ángela Alarcon Cruz |
| EPI_ISL_682275 | LABORATORIOS LABIN | Inciensa, Instituto Costarricense de Investigación y Enseñanza en Nutrición y Salud | Adriana Godinez; Claudio Soto-Garita; Estela Cordero; Francisco Duarte; Hebleen Porras; Melany Calderon & Pei Ling Chan |
| EPI_ISL_2893755 | LABOSUD | GenBio | Gwladys SEVRIN |
| EPI_ISL_717909 | LACEN RJ - Noel Nutels | Bioinformatics Laboratory / LNCC | Alexandra L Gerber; Amilcar Tanuri; Ana Paula de C Guimarães; Ana Tereza R de Vasconcelos; Andréa Cony Cavalcanti; Carolina M Voloch; Claudia dos Santos Rodrigues; Cynthia C Cardoso; Diana Mariani; Luiz G P de Almeida; Otavio Bustrolini; Ronaldo da Silva F Jr; Terezinha M P P Castiñeira |
| EPI_ISL_2821315, EPI_ISL_2821319, EPI_ISL_3046170, EPI_ISL_3046275, EPI_ISL_3046313, EPI_ISL_3046320, EPI_ISL_3046326, EPI_ISL_3046345, EPI_ISL_3046346 | | | |
| see above | LACEN/PE | WallauLab on behalf of Fiocruz COVID-19 Genomic Surveillance Network | Alexandre Freitas da Silva; Antonio Mauro Rezende; Cassia Docena; Constância Flávia Junqueira Ayres; Cássia Docena; Duschinka Ribeiro Duarte Guedes; Elisama Helvecio; Filipe Zimmer Dezordi; Gabriel Luz Wallau; Gustavo Barbosa de Lima; Lais Ceschini Machado; Larissa Krokovsky; Laís Ceschini Machado; Lilian Caroliny Amorim Silva; Marcelo Henrique dos Santos Paiva; Matheus Filgueira Bezerra; Sinval Pinto Brandão Filho |
| EPI_ISL_2463301, EPI_ISL_2463303 | LANCET LABORATORY | National Institute for Communicable Diseases of the National Health Laboratory Service | Amoako DG; Bhiman JN; Everatt J; Glass A; Sisonke Team; Viana R |
| EPI_ISL_2433867, EPI_ISL_2433906 | LANCET, LABORATORIES | National Institute for Communicable Diseases of the National Health Laboratory Service | Amoako DG; Bhiman JN; Ismail A; Mahlangu B; Mohale T; Ntuli N; Scheepers C |
| EPI_ISL_1935347 | LBM CLUSES | CNR Virus des Infections Respiratoires - France SUD | Antonin Bal; Bruno Lina; Bruno Simon; Gregory Destras; Gwendolyne Burfin; Hadrien Regue; Laurence Josset; Martine Valette; Quentin Semanas |
| EPI_ISL_1788463 | LBM DE LA SAUVEGARDE | CNR Virus des Infections Respiratoires - France SUD | Antonin Bal; Bruno Lina; Bruno Simon; Gregory Destras; Gwendolyne Burfin; Hadrien Regue; Laurence Josset; Martine Valette; Quentin Semanas |
| EPI_ISL_2827783, EPI_ISL_2827787 | LDSP - CAQUETA HOSPITAL MALVINAS | Instituto Nacional de Salud- Dirección de Investigación en Salud Pública | Carlos Franco-Muñoz; Carmen Osorio; Diana Malo; Diego A. Álvarez-Díaz; Diego Andrés Prada; Gerardo Santamaría; Hector Alejandro Ruiz-Moreno; Jhonnatan Reales-González; Jorge Rivera; Juan Camilo Martinez; Julian Naizaque; Katherine Laiton-Donato; Lisseth Pardo; Magdalena Wiesner; Marcela Mercado-Reyes; Maria T. Herrera-Sepúlveda; Marta Lopez Blanco; Martha Lucia Ospina Martinez; Paola Rojas; Sergio Gomez; Sheryll Corchuelo; Ángela Alarcon Cruz |
| EPI_ISL_2674311 | LDSP NARIÑO | Instituto Nacional de Salud- Dirección de Investigación en Salud Pública | Carlos Franco-Muñoz; Carmen Osorio; Diana Malo; Diego A. Álvarez-Díaz; Diego Andrés Prada; Gerardo Santamaría; Hector Alejandro Ruiz-Moreno; Jhonnatan Reales-González; Jorge Rivera; Juan Camilo Martinez; Julian Naizaque; Katherine Laiton-Donato; Lisseth Pardo; Magdalena Wiesner; Marcela Mercado-Reyes; Maria T. Herrera-Sepúlveda; Marta Lopez Blanco; Martha Lucia Ospina Martinez; Paola Rojas; Sergio Gomez; Sheryll Corchuelo; Ángela Alarcon Cruz |
| EPI_ISL_2894632 | LDSP QUINDIO | Instituto Nacional de Salud- Dirección de Investigación en Salud Pública | Carlos Franco-Muñoz; Carmen Osorio; Diana Malo; Diego A. Álvarez-Díaz; Diego Andrés Prada; Gerardo Santamaría; Hector Alejandro Ruiz-Moreno; Jhonnatan Reales-González; Jorge Rivera; Juan Camilo Martinez; Julian Naizaque; Katherine Laiton-Donato; Lisseth Pardo; Magdalena Wiesner; Marcela Mercado-Reyes; Maria T. Herrera-Sepúlveda; Marta Lopez Blanco; Martha Lucia Ospina Martinez; Paola Rojas; Sergio Gomez; Sheryll Corchuelo; Ángela Alarcon Cruz |
| EPI_ISL_3007486 | LDSP VILLAVICENCIO - CLINICA PRIMAVERA DE VILLAVICENCIO | Instituto Nacional de Salud- Dirección de Investigación en Salud Pública | Carlos Franco-Muñoz; Carmen Osorio; Diana Malo; Diego A. Álvarez-Díaz; Diego Andrés Prada; Gerardo Santamaría; Hector Alejandro Ruiz-Moreno; Jhonnatan Reales-González; Jorge Rivera; Juan Camilo Martinez; Julian Naizaque; Katherine Laiton-Donato; Lisseth Pardo; Magdalena Wiesner; Marcela Mercado-Reyes; Maria T. Herrera-Sepúlveda; Marta Lopez Blanco; Martha Lucia Ospina Martinez; Paola Rojas; Sergio Gomez; Sheryll Corchuelo; Ángela Alarcon Cruz |
| EPI_ISL_3024517, EPI_ISL_3024525 | LEBOWAKGOMO LABORATORY | National Institute for Communicable Diseases of the National Health Laboratory Service | Amoako DG; Bhiman JN; Everatt J; Ismail A; Mahlangu B; Mnguni A; Mohale T; Ntuli N; Scheepers C |
| EPI_ISL_2495924, EPI_ISL_2937905 | LESP Aguascalientes | Instituto de Diagnostico y Referencia Epidemiologicos (INDRE) | Abril Rodriguez-Maldonado; Ariadna Medina-Benitez; Claudia Wong-Arambula; Ernesto Ramirez-Gonzalez.; Gisela Barrera-Badillo; Irma Lopez-Martinez; Joaquin Quiroz-Mercado; Lucia Hernandez-Rivas; Maribel Gonzalez-Villa; Natividad Cruz-Ortiz; Sergio Rangel-Guerrero; Tatiana Nunez-Garcia; Vanessa Rivero-Arredondo |
| EPI_ISL_2296064, EPI_ISL_3033354 | LESP Baja California | Instituto de Diagnostico y Referencia Epidemiologicos (INDRE) | Abril Rodriguez-Maldonado; Ariadna Medina-Benitez; Claudia Wong-Arambula; Ernesto Ramirez-Gonzalez.; Gisela Barrera-Badillo; Irma Lopez-Martinez; Joaquin Quiroz-Mercado; Lucia Hernandez-Rivas; Maribel Gonzalez-Villa; Natividad Cruz-Ortiz; Sergio Rangel-Guerrero; Tatiana Nunez-Garcia; Vanessa Rivero-Arredondo |
| EPI_ISL_1651893, EPI_ISL_1821182, EPI_ISL_2533818 | LESP Baja California Sur | Instituto de Diagnostico y Referencia Epidemiologicos (INDRE) | Abril Rodriguez-Maldonado; Ariadna Medina-Benitez; Claudia Wong-Arambula; Ernesto Ramirez-Gonzalez.; Gisela Barrera-Badillo; Irma Lopez-Martinez; Joaquin Quiroz-Mercado; Lucia Hernandez-Rivas; Natividad Cruz-Ortiz; Sergio Rangel-Guerrero; Tatiana Nunez-Garcia; Vanessa Rivero-Arredondo |
| EPI_ISL_2094509, EPI_ISL_2094511, EPI_ISL_2533816, EPI_ISL_2559361 | LESP Campeche | Instituto de Diagnostico y Referencia Epidemiologicos (INDRE) | Abril Rodriguez-Maldonado; Ariadna Medina-Benitez; Claudia Wong-Arambula; Ernesto Ramirez-Gonzalez.; Gisela Barrera-Badillo; Irma Lopez-Martinez; Joaquin Quiroz-Mercado; Lucia Hernandez-Rivas; Natividad Cruz-Ortiz; Sergio Rangel-Guerrero; Tatiana Nunez-Garcia; Vanessa Rivero-Arredondo |
| EPI_ISL_2492494 | LESP Chiapas | Instituto de Diagnostico y Referencia Epidemiologicos (INDRE) | Abril Rodriguez-Maldonado; Ariadna Medina-Benitez; Claudia Wong-Arambula; Ernesto Ramirez-Gonzalez.; Gisela Barrera-Badillo; Irma Lopez-Martinez; Joaquin Quiroz-Mercado; Lucia Hernandez-Rivas; Natividad Cruz-Ortiz; Sergio Rangel-Guerrero; Tatiana Nunez-Garcia; Vanessa Rivero-Arredondo |
| EPI_ISL_2858979 | LESP Chihuahua | Instituto de Diagnostico y Referencia Epidemiologicos (INDRE) | Abril Rodriguez-Maldonado; Ariadna Medina-Benitez; Claudia Wong-Arambula; Ernesto Ramirez-Gonzalez.; Gisela Barrera-Badillo; Irma Lopez-Martinez; Joaquin Quiroz-Mercado; Lucia Hernandez-Rivas; Maribel Gonzalez-Villa; Natividad Cruz-Ortiz; Sergio Rangel-Guerrero; Tatiana Nunez-Garcia; Vanessa Rivero-Arredondo |
| EPI_ISL_1651889, EPI_ISL_2674689, EPI_ISL_2688985 | LESP Coahuila | Instituto de Diagnostico y Referencia Epidemiologicos (INDRE) | Abril Rodriguez-Maldonado; Ariadna Medina-Benitez; Claudia Wong-Arambula; Ernesto Ramirez-Gonzalez.; Gisela Barrera-Badillo; Irma Lopez-Martinez; Joaquin Quiroz-Mercado; Lucia Hernandez-Rivas; Natividad Cruz-Ortiz; Sergio Rangel-Guerrero; Tatiana Nunez-Garcia; Vanessa Rivero-Arredondo |
| EPI_ISL_2920686 | LESP Colima | Instituto de Diagnostico y Referencia Epidemiologicos (INDRE) | Abril Rodriguez-Maldonado; Ariadna Medina-Benitez; Claudia Wong-Arambula; Ernesto Ramirez-Gonzalez.; Gisela Barrera-Badillo; Irma Lopez-Martinez; Joaquin Quiroz-Mercado; Lucia Hernandez-Rivas; Natividad Cruz-Ortiz; Sergio Rangel-Guerrero; Tatiana Nunez-Garcia; Vanessa Rivero-Arredondo |
| EPI_ISL_1661512, EPI_ISL_2296101, EPI_ISL_2476421 | LESP Guanajuato | Instituto de Diagnostico y Referencia Epidemiologicos (INDRE) | Abril Rodriguez-Maldonado; Ariadna Medina-Benitez; Claudia Wong-Arambula; Ernesto Ramirez-Gonzalez.; Gisela Barrera-Badillo; Irma Lopez-Martinez; Joaquin Quiroz-Mercado; Lucia Hernandez-Rivas; Natividad Cruz-Ortiz; Sergio Rangel-Guerrero; Tatiana Nunez-Garcia; Vanessa Rivero-Arredondo |

| | | | |
|---|---|---|---|
| EPI_ISL_2663360, EPI_ISL_3033348 | LESP Hidalgo | Instituto de Diagnostico y Referencia Epidemiologicos (INDRE) | Abril Rodriguez-Maldonado; Ariadna Medina-Benitez; Claudia Wong-Arambula; Ernesto Ramirez-Gonzalez.; Gisela Barrera-Badillo; Irma Lopez-Martinez; Joaquin Quiroz-Mercado; Lucia Hernandez-Rivas; Maribel Gonzalez-Villa; Natividad Cruz-Ortiz; Sergio Rangel-Guerrero; Tatiana Nunez-Garcia; Vanessa Rivero-Arredondo |
| EPI_ISL_1821200, EPI_ISL_2094505, EPI_ISL_2533767 | LESP Jalisco | Instituto de Diagnostico y Referencia Epidemiologicos (INDRE) | Abril Rodriguez-Maldonado; Ariadna Medina-Benitez; Claudia Wong-Arambula; Ernesto Ramirez-Gonzalez.; Gisela Barrera-Badillo; Irma Lopez-Martinez; Joaquin Quiroz-Mercado; Lucia Hernandez-Rivas; Natividad Cruz-Ortiz; Sergio Rangel-Guerrero; Tatiana Nunez-Garcia; Vanessa Rivero-Arredondo |
| EPI_ISL_2246830, EPI_ISL_2340892, EPI_ISL_2340899, EPI_ISL_2340901, EPI_ISL_3033443, EPI_ISL_3033447, EPI_ISL_3046115, EPI_ISL_3046118 | | | |
| see above | LESP Michoacan | Instituto de Diagnostico y Referencia Epidemiologicos (INDRE) | Abril Rodriguez-Maldonado; Ariadna Medina-Benitez; Claudia Wong-Arambula; Ernesto Ramirez-Gonzalez.; Gisela Barrera-Badillo; Irma Lopez-Martinez; Joaquin Quiroz-Mercado; Lucia Hernandez-Rivas; Maribel Gonzalez-Villa; Natividad Cruz-Ortiz; Sergio Rangel-Guerrero; Tatiana Nunez-Garcia; Vanessa Rivero-Arredondo |
| EPI_ISL_1651907, EPI_ISL_1821146, EPI_ISL_1857248, EPI_ISL_2495935 | LESP Morelos | Instituto de Diagnostico y Referencia Epidemiologicos (INDRE) | Abril Rodriguez-Maldonado; Ariadna Medina-Benitez; Claudia Wong-Arambula; Ernesto Ramirez-Gonzalez.; Gisela Barrera-Badillo; Irma Lopez-Martinez; Joaquin Quiroz-Mercado; Lucia Hernandez-Rivas; Natividad Cruz-Ortiz; Sergio Rangel-Guerrero; Tatiana Nunez-Garcia; Vanessa Rivero-Arredondo |
| EPI_ISL_2158204 | LESP Nayarit | Instituto de Diagnostico y Referencia Epidemiologicos (INDRE) | Abril Rodriguez-Maldonado; Ariadna Medina-Benitez; Claudia Wong-Arambula; Ernesto Ramirez-Gonzalez.; Gisela Barrera-Badillo; Irma Lopez-Martinez; Joaquin Quiroz-Mercado; Lucia Hernandez-Rivas; Natividad Cruz-Ortiz; Sergio Rangel-Guerrero; Tatiana Nunez-Garcia; Vanessa Rivero-Arredondo |
| EPI_ISL_1805485, EPI_ISL_2296059 | LESP Oaxaca | Instituto de Diagnostico y Referencia Epidemiologicos (INDRE) | Abril Rodriguez-Maldonado; Ariadna Medina-Benitez; Claudia Wong-Arambula; Ernesto Ramirez-Gonzalez.; Gisela Barrera-Badillo; Irma Lopez-Martinez; Joaquin Quiroz-Mercado; Lucia Hernandez-Rivas; Natividad Cruz-Ortiz; Sergio Rangel-Guerrero; Tatiana Nunez-Garcia; Vanessa Rivero-Arredondo |
| EPI_ISL_2455897, EPI_ISL_2455901, EPI_ISL_2495928, EPI_ISL_2495953, EPI_ISL_2559289, EPI_ISL_2559309, EPI_ISL_2689824, EPI_ISL_3046095 | | | |
| see above | LESP Queretaro | Instituto de Diagnostico y Referencia Epidemiologicos (INDRE) | Abril Rodriguez-Maldonado; Ariadna Medina-Benitez; Claudia Wong-Arambula; Ernesto Ramirez-Gonzalez.; Gisela Barrera-Badillo; Irma Lopez-Martinez; Joaquin Quiroz-Mercado; Lucia Hernandez-Rivas; Maribel Gonzalez-Villa; Natividad Cruz-Ortiz; Sergio Rangel-Guerrero; Tatiana Nunez-Garcia; Vanessa Rivero-Arredondo |
| EPI_ISL_2296087, EPI_ISL_2492495 | LESP Quintana Roo | Instituto de Diagnostico y Referencia Epidemiologicos (INDRE) | Abril Rodriguez-Maldonado; Ariadna Medina-Benitez; Claudia Wong-Arambula; Ernesto Ramirez-Gonzalez.; Gisela Barrera-Badillo; Irma Lopez-Martinez; Joaquin Quiroz-Mercado; Lucia Hernandez-Rivas; Natividad Cruz-Ortiz; Sergio Rangel-Guerrero; Tatiana Nunez-Garcia; Vanessa Rivero-Arredondo |
| EPI_ISL_1821128, EPI_ISL_2663403, EPI_ISL_2689794 | LESP Sonora | Instituto de Diagnostico y Referencia Epidemiologicos (INDRE) | Abril Rodriguez-Maldonado; Ariadna Medina-Benitez; Claudia Wong-Arambula; Ernesto Ramirez-Gonzalez.; Gisela Barrera-Badillo; Irma Lopez-Martinez; Joaquin Quiroz-Mercado; Lucia Hernandez-Rivas; Natividad Cruz-Ortiz; Sergio Rangel-Guerrero; Tatiana Nunez-Garcia; Vanessa Rivero-Arredondo |
| EPI_ISL_2920762, EPI_ISL_3033552 | LESP Tabasco | Instituto de Diagnostico y Referencia Epidemiologicos (INDRE) | Abril Rodriguez-Maldonado; Ariadna Medina-Benitez; Claudia Wong-Arambula; Ernesto Ramirez-Gonzalez.; Gisela Barrera-Badillo; Irma Lopez-Martinez; Joaquin Quiroz-Mercado; Lucia Hernandez-Rivas; Maribel Gonzalez-Villa; Natividad Cruz-Ortiz; Sergio Rangel-Guerrero; Tatiana Nunez-Garcia; Vanessa Rivero-Arredondo |
| EPI_ISL_1821158, EPI_ISL_2246840, EPI_ISL_2455939, EPI_ISL_2455950 | LESP Tamaulipas | Instituto de Diagnostico y Referencia Epidemiologicos (INDRE) | Abril Rodriguez-Maldonado; Ariadna Medina-Benitez; Claudia Wong-Arambula; Ernesto Ramirez-Gonzalez.; Gisela Barrera-Badillo; Irma Lopez-Martinez; Joaquin Quiroz-Mercado; Lucia Hernandez-Rivas; Natividad Cruz-Ortiz; Sergio Rangel-Guerrero; Tatiana Nunez-Garcia; Vanessa Rivero-Arredondo |
| EPI_ISL_2937845 | LESP Tlaxcala | Instituto de Diagnostico y Referencia Epidemiologicos (INDRE) | Abril Rodriguez-Maldonado; Ariadna Medina-Benitez; Claudia Wong-Arambula; Ernesto Ramirez-Gonzalez.; Gisela Barrera-Badillo; Irma Lopez-Martinez; Joaquin Quiroz-Mercado; Lucia Hernandez-Rivas; Maribel Gonzalez-Villa; Natividad Cruz-Ortiz; Sergio Rangel-Guerrero; Tatiana Nunez-Garcia; Vanessa Rivero-Arredondo |
| EPI_ISL_1821163, EPI_ISL_2559279 | LESP Veracruz | Instituto de Diagnostico y Referencia Epidemiologicos (INDRE) | Abril Rodriguez-Maldonado; Ariadna Medina-Benitez; Claudia Wong-Arambula; Ernesto Ramirez-Gonzalez.; Gisela Barrera-Badillo; Irma Lopez-Martinez; Joaquin Quiroz-Mercado; Lucia Hernandez-Rivas; Natividad Cruz-Ortiz; Sergio Rangel-Guerrero; Tatiana Nunez-Garcia; Vanessa Rivero-Arredondo |
| EPI_ISL_2533779, EPI_ISL_2545762, EPI_ISL_3033498 | LESP Yucatan | Instituto de Diagnostico y Referencia Epidemiologicos (INDRE) | Abril Rodriguez-Maldonado; Ariadna Medina-Benitez; Claudia Wong-Arambula; Ernesto Ramirez-Gonzalez.; Gisela Barrera-Badillo; Irma Lopez-Martinez; Joaquin Quiroz-Mercado; Lucia Hernandez-Rivas; Maribel Gonzalez-Villa; Natividad Cruz-Ortiz; Sergio Rangel-Guerrero; Tatiana Nunez-Garcia; Vanessa Rivero-Arredondo |
| EPI_ISL_1752510 | LHUB-ULB | LHUB-ULB | Charlotte Michel; Marie Hallin; Olivier Vandenberg |
| EPI_ISL_1524979, EPI_ISL_2943031 | LHUB-ULB | Labo Klinische Biologie, UZA | Basil Britto Xavier; Christine Lammens; Herman Goossens; Ines Verbesselt; Jasmine Coppens; Kathleen Holemans; Marie Le Mercier; Veerle Matheeussen |
| EPI_ISL_1312503, EPI_ISL_1613434 | LIC | Latvian Biomedical Research and Study Centre | Davids Fridmanis; Diana Dusacka; Elina Dimina; Guntars Zarins; Ivars Silamikelis; Janis Klovins; Janis Pjalkovskis; Jurijs Perevoscikovs; Kaspars Megnis; Laila Silamikele; Lauma Freimane; Laura Ansone; Liga Birzniece; Monta Ustinova; Nikita Zrelovs; Reinis Zeltmatis; Uga Dumpis; Una Krumina; Vita Rovite |
| EPI_ISL_482737, EPI_ISL_482740 | LNR National Reference Laboratory, Mohammed VI University of Health Sciences | Medical Biotechnology Laboratory, Rabat Medical and Pharmacy School, Mohammed The Vth University in Rabat | Chakib NEJJARI; Houda BENRAHMA; Idrissa DIAWARA; Imane SMYEJ; Jalil El ATAR; Jalila RAHOUI; Lahcen BELYAMANI and Azeddine IBRAHIMI; Laila SBABOU; Loubna ALLAM; M.W. CHEMAO-ELFIHRI; Meriem LAAMARTI; Mouna OUADGHIRI; Rachid EL JAOUDI; Rachid MENTAG; Rokia LAAMARTI; Saaid AMZAZI; Souad KARTTI |
| EPI_ISL_889543, EPI_ISL_1913263 | LSUHS Emerging Viral Threat Laboratory | Microbial Genome Sequencing Center | Alexander Mijalis; Andrew D. Yurochko; Camille F. Abshire; Christopher G. Kevil; Daniel J. Snyder; Gregory L. Ware; Jennifer L. Carroll; Jeremy P. Kamil; John A. Vanchiere; Maarten Van Diest; Martin J. Sapp; Mohammed N.A. Siddiquey; Rona S. Scott; Vaughn S. Cooper |
| EPI_ISL_475082, EPI_ISL_717592 | Lab voor klinische biologie | Onderzoeksgroep Virologie | Bruno Verhasselt; Hans Nauwynck; Laurens Lambrechts; Linos Vandekerckhove; Marthe Pauwels; Nick Vereecke; Sebastiaan Theuns |
| EPI_ISL_775596 | Lab. Biologi Molekular FKIK Unwar, Bali | National Institute of Health Research and Development | AA; Dewi; HA; HD; Ikawati; KD; KNA; N; Nugraha; Pangesti; Pawestri; Puspa; Puspandari; SR; Setiawaty; Soekarso; Subangkit; T; V |
| EPI_ISL_1239665 | Lab. Biologii molekularnej USK | 1. National Institute of Public Health - National Institute of Hygiene; 2. Eurofins Genomics Europe Sequencing GmbH | ECDC COVID-19 WGS support team; Eurofins Genomics Europe Sequencing Team; Gierczyńki Rafał; Sadkowska-Todys Małgorzata; Wołkowicz Tomasz; Zacharczuk Katarzyna |
| EPI_ISL_2844508 | Lab. Microbiologia e Virologia Cotugno A.O. dei Colli - Istituto Zooprofilattico Sperimentale del Mezzogiorno | TIGEM | Antonio Grimaldi Patrizia Annunziata Francesco Panariello Biancamaria Pierri Claudia Tiberio Teresa Giuliano Valentina Bouche Chiara Colantuono Maria Concetta Cuomo Denise Di Concilio Lucio Di Filippo Anna Manfredi Marcello Salvi Antonio Limone Luigi Atripaldi Pellegrino Cerino Andrea Ballabio Davide Cacchiarelli |
| EPI_ISL_1154601 | LabKom - Labor Augsburg MVZ GmbH | Robert Koch Institute | |
| EPI_ISL_1642661, EPI_ISL_2260869, EPI_ISL_2314139 | LabKom - Labor Hannover MVZ GmbH | Robert Koch Institute | |
| EPI_ISL_2260098 | LabKom - Labor Mainz MVZ GmbH | Robert Koch Institute | |
| EPI_ISL_456211, EPI_ISL_456393, EPI_ISL_579106, EPI_ISL_579425, EPI_ISL_637092, EPI_ISL_661258, EPI_ISL_661261, EPI_ISL_661262, EPI_ISL_755631, EPI_ISL_794619, EPI_ISL_877212, EPI_ISL_1016858, EPI_ISL_1016867, EPI_ISL_1082265, EPI_ISL_1172031, EPI_ISL_1315312, EPI_ISL_1469088, EPI_ISL_1469093, EPI_ISL_1469097, EPI_ISL_1469107, EPI_ISL_1621303, EPI_ISL_1621304, EPI_ISL_1621309, EPI_ISL_1621314, EPI_ISL_1621316, EPI_ISL_1621319, EPI_ISL_1904850, EPI_ISL_1904853, EPI_ISL_1967901, EPI_ISL_1967902, EPI_ISL_2103202, EPI_ISL_2406489, EPI_ISL_2406490, EPI_ISL_2406495, EPI_ISL_2650017, EPI_ISL_2650021, EPI_ISL_2650022, EPI_ISL_2964924, EPI_ISL_2964927, EPI_ISL_2964930, EPI_ISL_2964932, EPI_ISL_2964935 | | | |
| see above | LabPLUS | Institute of Environmental Science and Research (ESR) | Anja Werno; Antje van der Linden; Arlo Upton; Chris Mansell; David Hammer; Dragana Drinkovic; Erasmus Smit; Gary McAuliffe; Hana Sofia Andersson; Hermes Perez; James Ussher; Jill Sherwood; Jing Wang; Joep de Ligt; Josh Freeman; Julia Howard; Juliet Elvy; Lauren Jelly; Mary DeAlmeida; Matt Blakiston; Matt Storey; Matthew Rogers; Max Bloomfield; Michael Addidle; Michelle Balm; Muhammad Faisal; Nikki Freed; Olin Silander; Olivia Stroeven; Rachel Boyle; Sally Roberts; SallyAnn Harbison; Sarah Jefferies; Sharmini Muttaiyah; Susan Morpeth; Susan Taylor; Timothy Blackmore; Vani Sathyendran; Veronica Playle; Virginia Hope; Xiaoyun Ren |
| EPI_ISL_548013 | LabTests | Institute of Environmental Science and Research (ESR) | Anja Werno; Antje van der Linden; Arlo Upton; Chris Mansell; David Hammer; Dragana Drinkovic; Erasmus Smit; Gary McAuliffe; Hana Sofia Andersson; Hermes Perez; James Ussher; Jill Sherwood; Jing Wang; Joep de Ligt; Josh Freeman; Julia Howard; Juliet Elvy; Lauren Jelly; Mary DeAlmeida; Matt Blakiston; Matt Storey; Matthew Rogers; Max Bloomfield; Michael Addidle; Michelle Balm; Muhammad Faisal; Nikki Freed; Olin Silander; Sally Roberts; Sarah Jefferies; Sharmini Muttaiyah; Susan Morpeth; Susan Taylor; Timothy Blackmore; Vani Sathyendran; Veronica Playle; Virginia Hope; Xiaoyun Ren |
| EPI_ISL_912603, EPI_ISL_1219949, EPI_ISL_1821587, EPI_ISL_2029615, EPI_ISL_2029621, EPI_ISL_2029888, EPI_ISL_2029936, EPI_ISL_2030060, EPI_ISL_2188487, EPI_ISL_2709261 | | | |
| see above | Labo Analyses Med | National Reference Center for Viruses of Respiratory Infections, Institut Pasteur, Paris | Adamou Lagare; Amaury Vaysse; Angela Brisebarre; Brieuc Gestin; Camille Capel; Christophe Malabat; Corinne Maufrais; Durivault JéRôMe; Emmanuelle Permal; Etienne Simon-Lorière; Frédéric Lemoine; Gael Millot; Louise Lefrançois; Marion Barbet; Maud Vanpeene; Méline Bizard; Patricia Tamby; Sylvaine Bastian; Sylvie Behillil; Sylvie Van der Werf; Sylvie van der Werf; Vincent Enouf |
| EPI_ISL_2003054 | Labolatorium Biologi Molekularnej USK | 1. Academic Center for Pathomorphological and Genetic-Molecular Diagnostics ltd, Bialystok, Poland 2. National Institute of Public Health - National Institute of Hygiene, Warsaw, Poland | Anetta Sulewska; Jacek Nikliński; Janusz Dzięcioł; Joanna Kiśluk; Katarzyna Zacharczuk; Konrad Raczkowski; Magdalena Nowakowska; Małgorzata Sadkowska-Todys; Piotr Karabowicz; Piotr Majewski; Przemysław Biecek. Joanna Reszeć; Radosław Charkiewicz; Tomasz Wołkowicz |
| EPI_ISL_1746802 | Labolatorium Biologi Molekularnej USK | 1. National Institute of Public Health - National Institute of Hygiene; 2. Eurofins Genomics Europe Sequencing GmbH | ECDC COVID-19 WGS support team; Eurofins Genomics Europe Sequencing Team; Gierczyńki Rafał; Sadkowska-Todys Małgorzata; Wołkowicz Tomasz; Zacharczuk Katarzyna |
| EPI_ISL_2897150 | Labolatorium Biologi Molekularnej USK | 1. Tricity SARS-CoV-2 sequencing consortium: University of Gdansk, Medical University of Gdansk, Vaxican Ltd., Invicta Ltd. 2. National Institute of Public Health - National Institute of Hygiene, Warsaw, Poland | Celina Cybulska; Karolina Gackowska; Katarzyna Groth; Katarzyna Zacharczuk; Krystyna Bienkowska Szewczyk; Lukasz Rabalski; Maciej Grzybek; Maciej Kosinski; Magdalena Nowakowska; Marcin Lubocki; Małgorzata Sadkowska-Todys; Tomasz Wołkowicz |
| EPI_ISL_2761448 | Labor 28 MVZ GmbH | Robert Koch Institute | |
| EPI_ISL_2845726 | Labor Becker & Kollegen (Standort München) | Robert Koch Institute | |
| EPI_ISL_2390079 | Labor Blackholm MVZ | Robert Koch Institute | |
| EPI_ISL_1463910, EPI_ISL_1716024, EPI_ISL_1862119, EPI_ISL_1862322, EPI_ISL_2384709 | Labor Dr. Fenner und Kollegen | Heinrich Pette Institute, Leibniz Institute for Experimental Virology | Adam Grundhoff; Alexis Robitaille; Johannes Knobloch; Martin Aepfelbacher; Nicole Fischer; Thomas Günther |
| EPI_ISL_2129685 | Labor Dr. Heidrich & Kollegen MVZ GmbH Hamburg | Robert Koch Institute | |
| EPI_ISL_2125015 | Labor Dr. Wisplinghoff - Koln | Robert Koch Institute | |
| EPI_ISL_1850332 | Labor Dr. Wisplinghoff - KÃ¶ln | Robert Koch Institute | |
| EPI_ISL_1215851, | Labor Dr. Wisplinghoff - Köln | Robert Koch Institute | |

| | | | |
|---|---|---|---|
| EPI_ISL_2266929 | | | |
| EPI_ISL_803955 | Labor Dr. Wisplinghoff - Köln | Robert Koch Institute, Influenza and respiratory viruses FG17 & Bioinformatics MF1, Berlin, Germany | Aleksandar Radonic; Dr. R. Grosser; Marianne Wedde; Oliver Drechsel; Ralf Duerrwald; Rene Kmiecinski; Stefan Kroeger; Stephan Fuchs; Thorsten Wolff |
| EPI_ISL_3044981 | Labor Mönchengladbach MVZ Dr. Stein + Kollegen GbR | Robert Koch Institute | |
| EPI_ISL_1140227, EPI_ISL_1140343, EPI_ISL_1140360, EPI_ISL_1285209 | Labor ZOTZ\|KLIMAS; MVZ Düsseldorf-Centrum | Robert Koch Institute | |
| EPI_ISL_775220 | Laboratoire Biolife | Laboratoire de Biotechnologie | Afaf Alaoui; Amina Benouda; Bouchra Belfquih; Hanae Dakka; Lahcen belyamani; Mohamed Chenaoui; Mohammed Walid Chemao Elfihri; Mouna Ouadghiri; Otmane Touzani; Saaid Amzazi and Azeddine Ibrahimi; Tarik Aanniz |
| EPI_ISL_1508956, EPI_ISL_1575436, EPI_ISL_1575441, EPI_ISL_1675141 | Laboratoire Biolim/FSS/UL | Unité Mixte Internationale TransVIHMI (UMI 233 IRD – U1175 INSERM - Université de Montpellier) IRD (Institut de recherche pour le développement) | Abla A. KONOU; Adodo SADJI; Ahidjo AYOUBA; Akoélé SILIADIN; Alassane OURO-MEDELI; Amivi EHLAN; Améyo DORKENOO; Anoumou DAGNRA; Christelle BUTEL; Déléma MABA; Eric DELAPORTE; Issaka Maman; Kokou TEGUENI; Koku AGBODEKA; Laetitia SERRANO; Martine PEETERS; Messanh DOUFFAN; Mireille PRINCE-DAVID; Mounerou SALOU; Sidonie A.M.KAGNISSODE; Sika DOSSIM; Syntyche DEVATCHAGNI; Wembo A. HALATOKO |
| EPI_ISL_2835003 | Laboratoire Central Analyse Médicale mohammedia | Functional Genomic Platform_UATRS_CNRST_Rabat | Amina Barkat; Belayachi Lamiae; Elmostafa El fahime; Hajjaji Mohammed; Hasnae Benkirane; Hassan Aguenaou; Jaafar Heikel; Marouane Melloul; Mouhcine Gardoul; Mouhssine Hemlali; Nadia Touil; Oumaima Ait Si Mohammed; Sanaa Alaoui-Amine; Taha Chouati Omar Askander |
| EPI_ISL_1905079, EPI_ISL_2318037 | Laboratoire Central de Virologie | Laboratoire de Biotechnologie | Abdelomunim Essabbar; Amal Zouaki; Ghizlane EL Amin; Hakima Kabbaj; Lahcen Belyamani and Azeddine Ibrahimi; Mouna Ouadghiri; Myriam Seffar; Naima El Hafidi; Saaïd Amzazi; Tarik Aanniz |
| EPI_ISL_2978186, EPI_ISL_2978187, EPI_ISL_2978188, EPI_ISL_2978191, EPI_ISL_2978195, EPI_ISL_2978196, EPI_ISL_2978200, EPI_ISL_2978201, EPI_ISL_2978203, EPI_ISL_2978206, EPI_ISL_2978211, EPI_ISL_2978223, EPI_ISL_2978224, EPI_ISL_2978225, EPI_ISL_2978226, EPI_ISL_2978236 | | | |
| see above | Laboratoire National de Santé Publique, Brazzaville, République du Congo | Pathogen Genomics Lab, National Institute for Biomedical Research (INRB) | Amuri Aziza; Andrew Rambaut; Catherine Pratt; Eddy Kinganda-Lusamaki; Edith Nkwembe; Emmanuel Lokilo Lofiko; Fabien Roch Niama; Francisca Muyembe Mawete; Gabriel Kabamba; Ian Goodfellow; Jean Claude Makangara; Jean-Jacques Muyembe Tamfum; Josh Quick; Matthias Pauthner; Michael Wiley; Nick Loman; Placide Mbala-Kingebeni; Raphael Lumembe; Steve Ahuka-Mundeke; Trevor Bedford |
| EPI_ISL_459898 | Laboratoire National de Sante, Microbiology, Virology | Laboratoire National de Sante, Microbiology, Epidemiology and Microbial Genomics | Anke Wienecke-Baldacchino; Catherine Ragimbeau; Guillaume Fournier; Jessica Tapp; Tamir Abdelrahman; Trung Nguyen Nguyen |
| EPI_ISL_1972306 | Laboratoire National de Santé Publique du Cameroun | Pathogen Genomics Lab, National Institute for Biomedical Research (INRB) | Amuri Aziza; Andrew Rambaut; Catherine Pratt; Eddy Kinganda-Lusamaki; Edith Nkwembe; Emmanuel Lokilo Lofiko; Francisca Muyembe Mawete; Gabriel Kabamba; Ian Goodfellow; Jean Claude Makangara; Jean-Jacques Muyembe Tamfum; Josh Quick; Marie Claire Okomo; Matthias Pauthner; Michael Wiley; Nick Loman; Placide Mbala-Kingebeni; Raphael Lumembe; Steve Ahuka-Mundeke; Trevor Bedford |
| EPI_ISL_2157560, EPI_ISL_2492505, EPI_ISL_2492517, EPI_ISL_2492525 | Laboratoire National de Santé Publique – LNSP (HAITI - LNSP) | Laboratory of Respiratory Viruses and Measles, Oswaldo Cruz Institute, FIOCRUZ | Alice Sampaio Rocha; Ana Carolina Mendonca; Anna Carolina Paixao; Elisa Cavalcante Pereira; Fernando Motta; Ito Journel; Jaques Boncy; Luciana Appolinario; Marilda Siqueira on behalf of the Fiocruz COVID-19 Genomic Surveillance Network; Paola Resende; Patrick Delly; Renata Serrano Lopes; Taina Venas |
| EPI_ISL_1760554 | Laboratoire Professeur Daniel GAHOUMA (LPDG) | Centre de recherches médicales de Lambaréné (CERMEL) | Ayola A. Adegnika; Ayong Moure; Bertrand Lell; Bénédicte Ndeboko; Emilio Skarwan; Georgelin Nguema Ondo; Gédéon P. Manouana; Haruka Abe; Jiro Yasuda; Joel Fleury Djoba Siawaya; Rodrigue Bikangui; Rotimi Myrabelle Avome Houechenou; Samira Zoa-Assoumou; Yuri Ushijima |
| EPI_ISL_2373998 | Laboratoire Virologie Saint Louis APHP | Laboratoire Virologie Saint Louis APHP | Constance Delauguerre; Jérome Le Goff; Linda Feghoul; Marie Laure Chaix; Marie Laure Néré; Maud Salmona; Severine Mercier Delarue; Sophia Achaibou |
| EPI_ISL_1103576, EPI_ISL_1810947, EPI_ISL_1896699, EPI_ISL_1904886 | Laboratoire central de Virologie | Laboratoire de Biotechnologie | Abdelomunim Essabbar; Amal Zouaki; Ghizlane EL Amin; Hakima Kabbaj; Lahcen Belyamani and Azeddine Ibrahimi; Mouna Ouadghiri; Myriam Seffar; Saaïd Amzazi; Tarik Aanniz |
| EPI_ISL_2227202, EPI_ISL_2227203 | Laboratoire d'analyse biomédicale du camp Ouezzin Coulibaly | Centre Muraz | Abdoul-Salam Ouedraogo; Amariane Koné; Armel Poda; Arsène Zongo; Essia Belarbi; Fabian Leendertz; Grit Schubert; Halidou Tinto; Soumeya Ouangraoua; Thérèse Kagoné; Yacouba Sawadogo; Zekiba Tarnagda |
| EPI_ISL_2820514 | Laboratoire de Diagnostic, Centre de Soins 1, Caisse Nationale de Sécurité Sociale (CNSS) de Djibouti | IHU Méditerranée Infection | Anthony Levasseur; Christian A. Devaux.; Didier Raoult; Idil Salah abdillahi; Ikram Omar Osman; Iman Abdillahi Hassan; Jeremy Delerce; Linda Houhamdi; Ludivine Brechar; Mohamed Houmed Aboubaker; Philippe Colson; Pierre-Edouard Fournier; Zeinab Ali Waberi |
| EPI_ISL_2820520 | Laboratoire de Diagnostic, Centre de Soins 1, Caisse Nationale de Sécurité Sociale (CNSS) de Djibouti, | IHU Méditerranée Infection. | Anthony Levasseur; Christian A. Devaux.; Didier Raoult; Idil Salah abdillahi; Ikram Omar Osman; Iman Abdillahi Hassan; Jeremy Delerce; Linda Houhamdi; Ludivine Brechar; Mohamed Houmed Aboubaker; Philippe Colson; Pierre-Edouard Fournier; Zeinab Ali Waberi |
| EPI_ISL_2820626 | Laboratoire de Diagnostic, Centre de Soins 1, Caisse Nationale de Sécurité Sociale (CNSS) de Djibouti. | IHU Méditerranée Infection. | Anthony Levasseur; Christian A. Devaux.; Didier Raoult; Idil Salah abdillahi; Ikram Omar Osman; Iman Abdillahi Hassan; Jeremy Delerce; Linda Houhamdi; Ludivine Brechar; Mohamed Houmed Aboubaker; Philippe Colson; Pierre-Edouard Fournier; Zeinab Ali Waberi |
| EPI_ISL_660448, EPI_ISL_660452, EPI_ISL_660471, EPI_ISL_660477, EPI_ISL_660478, EPI_ISL_660479, EPI_ISL_660492, EPI_ISL_660499, EPI_ISL_660515, EPI_ISL_2142708, EPI_ISL_2142721, EPI_ISL_2142743 | | | |
| see above | Laboratoire de Microbiologie CHU Souro Sanou | Centre Muraz | Abdoul-Salam Ouedraogo; Amariane Koné; Armel Poda; Arsène Zongo; Essia Belarbi; Fabian Leendertz; Grit Schubert; Halidou Tinto; Lassana Sangaré; Soumeya Ouangraoua; Thérèse Kagoné; Yacouba Sawadogo; Zekiba Tarnagda |
| EPI_ISL_2289108 | Laboratoire de Microbiologie CHU Souro Sanou | Laboratoire bacteriologie virologie CHUSS | Abdoul-Salam Ouedraogo; Abdoulie Kanteh; Abdul Sesay; Annette Erhart; Armel Poda; François Kiemdé; Halidou Tinto; Mariama Kujabi; Yacouba Sawadogo |
| EPI_ISL_1116464, EPI_ISL_1116467, EPI_ISL_1116468, EPI_ISL_1118923 | Laboratoire de Microbiologie- CHU Habib Bourguiba – Sfax | Laboratoire des Procédés de Criblage Moléculaire et Cellulaire-Centre de Biotechnologie de Sfax | A. and Masmoudi, S.; Abdelmoulah, F.; Abid, N.; Ajili, F.; Aouni, M.; Ben Ayed, I.; Bensaid, M.; Chtourou, A.; Elargoubi, A.; Fki-berrajah, L.; Gaaloul, I.; Gargouri, S.; Hammami, A.; Kamoun, S.; Karray Hakim, H.; Kharat, N.; Mastouri, M.; Mhalla, S.; Nabli, A.; Rebai; Smeti, I.; Souissi, A.; Stambouli, N.; Turki, M. |
| EPI_ISL_414600 | Laboratoire de Virologie Institut de Virologie - INSERM U 1109 Hôpitaux Universitaires de Strasbourg | National Reference Center for Viruses of Respiratory Infections, Institut Pasteur, Paris | Angela Brisebarre; Flora Donati Vincent Enouf; Marion Barbet; Maud Vanpeene; Méline Bizard; Mélnie Albert; Samira Fafi-Kremer; Sylvie Behillil; Sylvie van der Werf |
| EPI_ISL_825711, EPI_ISL_890133, EPI_ISL_890145, EPI_ISL_890158, EPI_ISL_1366018, EPI_ISL_1379080, EPI_ISL_1400646, EPI_ISL_1400702, EPI_ISL_1400733, EPI_ISL_1403232, EPI_ISL_1403325, EPI_ISL_1403327, EPI_ISL_1523424, EPI_ISL_1534163, EPI_ISL_1696760, EPI_ISL_1696761, EPI_ISL_1696762, EPI_ISL_1696763, EPI_ISL_1696764, EPI_ISL_1696765, EPI_ISL_1696818, EPI_ISL_1696894, EPI_ISL_1696913, EPI_ISL_1697201, EPI_ISL_1697202, EPI_ISL_1697203, EPI_ISL_1697205, EPI_ISL_2007707, EPI_ISL_2007710, EPI_ISL_2293764, EPI_ISL_2293840, EPI_ISL_2293901, EPI_ISL_2388272, EPI_ISL_2390825, EPI_ISL_2390950, EPI_ISL_2390959, EPI_ISL_2821389, EPI_ISL_2821439, EPI_ISL_2821747, EPI_ISL_2822078, EPI_ISL_2822086, EPI_ISL_2822095, EPI_ISL_2822121, EPI_ISL_2822220, EPI_ISL_2822230, EPI_ISL_2822242, EPI_ISL_2822257, EPI_ISL_2822259, EPI_ISL_2824272, EPI_ISL_2824348, EPI_ISL_2824702, EPI_ISL_2824716, EPI_ISL_2824921, EPI_ISL_2824929, EPI_ISL_2970215, EPI_ISL_2970217, EPI_ISL_2970307, EPI_ISL_2970418, EPI_ISL_2970502, EPI_ISL_2970657, EPI_ISL_2975903, EPI_ISL_2976528, EPI_ISL_2976529, EPI_ISL_2976530, EPI_ISL_2976531, EPI_ISL_3144438 | | | |
| see above | Laboratoire de santé publique du Québec | Laboratoire de santé publique du Québec | Guillaume Bourque; Ioannis Ragoussis; Jesse Shapiro; Mark Lathrop and Michel Roger on behalf of the CoVSeQ research group; Mark Lathrop and Michel Roger on behalf of the CoVSeQ research group (http://covseq.ca/researchgroup); Sandrine Moreira |
| EPI_ISL_476822, EPI_ISL_476824, EPI_ISL_476826, EPI_ISL_476829, EPI_ISL_476830, EPI_ISL_476831 | Laboratoire des Fièvres Hémorragiques Virales du Benin | Charité-Universitätsmedizin Berlin | Anges; Drexler; Jan Felix; Moreira-Soto Andres; Sander Anna-Lena; Yadouleton |
| EPI_ISL_910672, EPI_ISL_910845, EPI_ISL_1383353 | Laboratoire national de sante, Microbiology, Virology | Laboratoire national de sante, Microbiology, Microbial Genomics Platform | Anke Wienecke-Baldacchino; Catherine Ragimbeau; Fatu Djabi; Jessica Tapp; Lise Pignon; Raoul Salmon; Tamir Abdelrahman; Trung Nguyen Nguyen |
| EPI_ISL_739880, EPI_ISL_740129 | Laboratoire national de santé, Microbiology, Virology | Laboratoire national de santé, Microbiology, Epidemiology and Microbial Genomics | Anke Wienecke-Baldacchino; Catherine Ragimbeau; Fatu Djabi; Jessica Tapp; Tamir Abdelrahman; Trung Nguyen Nguyen |
| EPI_ISL_739901, EPI_ISL_739947, EPI_ISL_740028, EPI_ISL_744139, EPI_ISL_744561, EPI_ISL_744958, EPI_ISL_771034 | | | |
| see above | Laboratoire national de santé, Microbiology, Virology | Laboratoire national de santé, Microbiology, Microbial Genomics Platform | Anke Wienecke-Baldacchino; Catherine Ragimbeau; Fatu Djabi; Jessica Tapp; Lise Pignon; Raoul Salmon; Tamir Abdelrahman |
| EPI_ISL_1384323 | Laboratoires d'analyses medicales - Ketterthill | Laboratoire national de sante, Microbiology, Microbial Genomics Platform | Anke Wienecke-Baldacchino; Caroline Scheiber; Catherine Ragimbeau; Fatu Djabi; Jessica Tapp; Lise Pignon; Raoul Salmon; Serge Vedy; Tamir Abdelrahman |
| EPI_ISL_2645438, EPI_ISL_2863801, EPI_ISL_2863816 | Laboratorio Central de Saude Publica do Estado do Para (LACEN/PA) | Laboratory of Respiratory Viruses and Measles, Oswaldo Cruz Institute, FIOCRUZ | Agatha Cristinne Prudencio Soares; Alice Sampaio Rocha; Ana Carolina Mendonca; Anna Carolina Paixao; Elisa Cavalcante Pereira; Fernando Motta; Ighor Leonardo Arantes Gomes; Luciana Appolinario; Marilda Siqueira on behalf of the Fiocruz COVID-19 Genomic Surveillance Network; Paola Resende; Renata Serrano Lopes; Taina Venas; Valnete Andrade |
| EPI_ISL_981041 | Laboratorio Central Sección Virología. Hospital Regional Artemides Zatti (Viedma, Rio Negro); Laboratorio MICROBIOM (General Roca, Río Negro); Hospital Zonal Dr. Ramón Carrillo (San Carlos De Bariloche, Río Negro) | Laboratorio Central Mg. Luis Alfredo Pianciola on behalf of 'Proyecto Argentino Interinstitucional de genomica de SARS-CoV-2' (PAIS Consortium) | Antonela De Fino; C Pintos; C Ziehm; Darío Fabián di Prátula; J Ousset; L Pianciola.; Liliana Fonseca; M Fernandez; M Mazzeo; M Nabaes; Marcela Nóbile; María Laura Álvarez; Patricia Valeria Blanco; Silvana Cecchi; Yesica Espasandin |
| EPI_ISL_1288281, EPI_ISL_1811413, EPI_ISL_1811433, EPI_ISL_1811479 | Laboratorio Central de Epidemiología (LCE) | Instituto de Biotecnología de la UNAM | Alejandro Sanchez-Flores; Alfredo Herrera-Estrella; Alicia Ocaña-Mondragón; Angel Gustavo Salas-Lais; Bernardo Martínez-Miguel; Blanca Taboada; Brenda Irasema Maldonado-Meza; Carla Ivón Herrera-Najera; Carlos F. Arias; Celia Boukadida; Clara Esperanza Santacruz-Tinoco; Concepción Grajales-Muñiz; Consorcio Mexicano de Vigilancia Genómica (CoViGen-Mex). Authors (in alphabetical order): Julio Elias Alvarado-Yaah; Célida Duque-Molina; Fernando Fontove-Herrera; Francisco Pulido; Gloria Elena Espinosa-Ayala; Gloria Elena Espinoza-Ayala; Gloria María Molina-Salinas; Gloria Vazquez; Hector Esteban Paz-Juárez; Hector Montoya-Fuentes; Helen Haydee Fernanda Ramírez-Plascencia; Jorge Ivan Salinal-Nevarez; José Antonio Enciso-Moreno; José Esteban Muñoz-Medina; José de Jesús Nuñez-Contreras; Juan Bautista Chale-Dzul; Julio Elias Alvarado-Yaah; Luis Alberto Ochoa-Carrera; Margarita Matías-Florentino; María Guadalupe Santiago-Mauricio; María Guadalupe de Jesús Mireles-Rivera; Nelly Sélem-Mojica; Pavel Isa; Ricardo Grande; Santiago Ávila-Ríos; Victor Eduardo Garcia-Arias; Víctor Hugo Borja-Aburto |
| EPI_ISL_1301498, EPI_ISL_1302158 | Laboratorio Central de Epidemiología IMSS | Instituto de Biotecnología de la UNAM | Alejandra Hernández-Terán; Alejandro Sanchez-Flores; Alma Rincón-Rubio; Andrea Santos Coy-Arechavaleta; Authors from IBT; Blanca Taboada; Celia Boukadida; Clara Esperanza Santacruz-Tinoco; Edgar Mendieta-Condado; Eduardo Becerril-Vargas; Fidencio Mejía-Nepomuceno; Francisco Pulido; Gisela Barrera-Badillo; Gloria Vazquez; Hector Esteban Paz-Juárez; IMSS; InDRE and INER (in alphabetical order): Carlos F. Arias; Irma Lopez-Martinez; Jerome Jean Verleyen; Joel Armando Vázquez-Pérez; Jorge Salas-Hernández; José Arturo Martínez-Orozco; José Ernesto Ramírez-González; José Esteban Muñoz-Medina; Larissa Fernandes-Matano; Lucia Hernandez-Rivas; Luis Alberto Ochoa-Carrera; Margarita Matías-Florentino; Mario Mújica-Sánchez; Natividad Cruz-Ortiz; Pavel Isa; Ricardo Grande; Santiago Ávila-Ríos; Tatiana Nunez-Garcia; Teresita Rojas-Mendoza |
| EPI_ISL_1492641, EPI_ISL_1492656, EPI_ISL_1492658, EPI_ISL_1492661, EPI_ISL_1492666, EPI_ISL_1492672, EPI_ISL_1492675, EPI_ISL_1492679 | | | |
| see above | Laboratorio Central de Salud Publica de Paraguay | Laboratorio Central de Salud Publica de Paraguay | Andrea Gómez de la Fuente; Cynthia Vázquez; Flavia Aburjaile; Juan Torales; Luiz Carlos Junior Alcantara; Marta Giovanetti; María José Ortega; María Liz Gamarra; Shirley Villalba; Talita Adelino; Vagner Fonseca |
| EPI_ISL_2645411 | Laboratorio Central de Saude Publica do Estado Maranhao (LACEN-MA) | Laboratory of Respiratory Viruses and Measles, Oswaldo Cruz Institute, FIOCRUZ | Alice Sampaio Rocha; Ana Carolina Mendonca; Anna Carolina Paixao; Elisa Cavalcante Pereira; Fernando Motta; Lidio Gonçalves Lima Neto; Luciana Appolinario; Marilda Siqueira on behalf of the Fiocruz COVID-19 Genomic Surveillance Network; Paola Resende; Renata Serrano Lopes; Taina Venas |

| | | | |
|---|---|---|---|
| EPI_ISL_2645396, EPI_ISL_2645667 | Laboratorio Central de Saude Publica do Estado de Alagoas (LACEN/AL) | Laboratory of Respiratory Viruses and Measles, Oswaldo Cruz Institute, FIOCRUZ | Alice Sampaio Rocha; Ana Carolina Mendonca; Anderson Brandao Leite; Anna Carolina Paixao; Elisa Cavalcante Pereira; Fernando Motta; Luciana Appolinario; Marilda Siqueira on behalf of the Fiocruz COVID-19 Genomic Surveillance Network; Paola Resende; Renata Serrano Lopes; Taina Venas |
| EPI_ISL_2863619 | Laboratorio Central de Saude Publica do Estado de Sergipe (LACEN/SE) | Laboratory of Respiratory Viruses and Measles, Oswaldo Cruz Institute, FIOCRUZ | Alice Sampaio Rocha; Ana Carolina Mendonca; Anna Carolina Paixao; Cliomar Alves dos Santos; Elisa Cavalcante Pereira; Fernando Motta; Luciana Appolinario; Marilda Siqueira on behalf of the Fiocruz COVID-19 Genomic Surveillance Network; Paola Resende; Renata Serrano Lopes; Tainá Moreira Martins Venas |
| EPI_ISL_3045555 | Laboratorio Central de Saude Publica do Estado do Espirito Santo (LACEN/ES) | Laboratory of Respiratory Viruses and Measles, Oswaldo Cruz Institute, FIOCRUZ | Alice Sampaio Rocha; Ana Carolina Mendonca; Anna Carolina Paixao; Eilisa Cavalcante Pereira; Fernando Motta; Luciana Appolinario; Marilda Siqueira on behalf of the Fiocruz COVID-19 Genomic Surveillance Network; Paola Resende; Renata Serrano Lopes; Rodrigo Ribeiro Rodrigues; Taina Venas |
| EPI_ISL_2775494 | Laboratorio Central de Saude Publica do Estado do Parana (Instituto de Biologia Molecular do Paraná (LAC)EN-PR) | Instituto Carlos Chagas - Fiocruz | Alessandra De Melo Aguiar; Andreia Akemi Suzukawa; Andréa Rodrigues Ávila; Bruno Dallagiovanna; Dalila Zanette; Eduardo Balsanelli; Emanuel Maltempi de Souza; Fabio Passetti; Fabricio Klerynton Marchini; Fábio de Oliveira Pedrosa; Guilherme Becker; Helisson Faoro; Hellen Geremias dos Santos; Irina Nastassja Riediger; Letusa Albrecht; Lucas Blanes; Luis Gustavo Morello; Lysangela Ronalte Alves; Maria do Carmo Debur; Mauro de Medeiros Oliveira; Michelle Orane Schemberger; Paola Cristina Resende; Sheila Cristina Nardeli; Tiago Gräf; Valter Antônio de Baura |
| EPI_ISL_2645467, EPI_ISL_2731458 | Laboratorio Central de Saude Publica do Estado do Parana (LACEN/PR) | Laboratory of Respiratory Viruses and Measles, Oswaldo Cruz Institute, FIOCRUZ | Alice Sampaio Rocha; Ana Carolina Mendonca; Anna Carolina Paixao; Elisa Cavalcante Pereira; Fernando Motta; Irina Riediger; Luciana Appolinario; Marilda Siqueira on behalf of the Fiocruz COVID-19 Genomic Surveillance Network; Paola Resende; Renata Serrano Lopes; Taina Venas |
| EPI_ISL_792519 | Laboratorio Central de la Ciudad de Santa Fe | Grupo de Genómica y Bioinformática del Instituto de Investigación de la Cadena Láctea CONICET-INTA on behalf of 'Proyecto Argentino Interinstitucional de genomica de SARS-CoV-2' (PAIS Consortium) | AF; Amadio; C; Eberhardt; G; Irazoqui; JM; MF; Mugna; Ojeda; Pastor; Rompato; V |
| EPI_ISL_2271696 | Laboratorio Central, Ministerio de Salud Cordoba | Instituto de Patologia Vegetal (CIAP-INTA) on behalf of 'Proyecto Argentino Interinstitucional de genomica de SARS-CoV-2' (PAIS Consortium) | Barbas, G.; Castro, G.; Debat, HJ.; FD; Fernandez; M; M.B.; Marquez, N.; Pisano; Re, V. |
| EPI_ISL_792539, EPI_ISL_2940267, EPI_ISL_2940286 | Laboratorio Central, Ministerio de Salud Córdoba | Instituto de Patologia Vegetal (CIAP-INTA) on behalf of 'Proyecto Argentino Interinstitucional de genomica de SARS-CoV-2' (PAIS Consortium) | Barbas, G.; Castro, G.; Debat, HJ.; FD; Fernández; M; M.B.; MB; Marquez, N.; Pisano; Re; Re, V.; V |
| EPI_ISL_424667 | Laboratorio Estatal de Salud Publica del Estado de México | Instituto de Diagnóstico y Referencia Epidemiológicos | Adnan Araiza Rodríguez; Alejandro Sánchez; Alfredo Ponce de León Garduño; Blanca Taboada; Carlos F. Arias.; Carolina González Torres; Celia Boukadida; Cesar Raúl González Bonilla; Concepción Grajales Muñiz; Edgar Mendieta Condado; Eduardo Becerril Vargas; Fabiola Garcés Ayala; Fernando Ledesma Barrientos; Francisco Javier Gaytán Cervantes; Francisco Pulido; Gisela Barrera Badillo; Gloria Vázquez; Guillermo M. Ruiz-Palacios; Irma López Martínez; Joel Armando Vázquez Pérez; José Arturo Martínez Orozco; José Ernesto Ramírez González; José Esteban Muñoz Medina; Lucia Hernández Rivas; Luis Alberto García Andrade; Mario Mújica Sánchez; Pavel Isa; Pilar Ramos Cervantes; Ricardo Grande; Santiago Avila Ríos; Victor Hugo Borja Aburto; Violeta Ibarra Gonzalez |
| EPI_ISL_779189, EPI_ISL_779196 | Laboratorio Estatal de Salud Pública de Nuevo León | Laboratorio de Infectología Molecular, Departamento de Bioquímica y Medicina Molecular,Facultad de Medicina - Universidad Autónoma de Nuevo León | Ana M. Rivas-Estilla; Consuelo Treviño-Garza; Daniel Arellanos-Soto; Else del Carmen García-García; Gloria A. Jasso-de-la-Peña; Kame A. Galán-Huerta; Manuel E. de-la-O-Cavazos; María F. Herrera-Saldivar; Natalia Martínez-Acuña; Roberto Montes-de-Oca; Samuel Buentello-Wong; Sonia A. Lozano-Sepúlveda |
| EPI_ISL_2825105, EPI_ISL_2825117, EPI_ISL_2825135, EPI_ISL_2826967, EPI_ISL_2826996, EPI_ISL_2827807, EPI_ISL_2827824, EPI_ISL_2827828, EPI_ISL_2827835, EPI_ISL_2827848 | | | |
| see above | Laboratorio Nacional de Salud | Laboratorio Nacional de Salud | Gabriela García; Linda Mendoza |
| EPI_ISL_837551, EPI_ISL_837579, EPI_ISL_837589 | Laboratorio Nacional de Salud | Laboratory of Respiratory Viruses and Measles, Oswaldo Cruz Institute, FIOCRUZ | Ana Carolina Mendonca; Anna Carolina Paixao; Cesar Roberto Conde Pereira; Claudia Estrada; Fernando Motta; Luciana Appolinario; Marilda Siqueira on behalf of the Fiocruz COVID-19 Genomic Surveillance Network; Paola Resende |
| EPI_ISL_2274010, EPI_ISL_3045384, EPI_ISL_3045410, EPI_ISL_3045422 | Laboratorio Nacional de Salud Pública Dr. Defilló - LNSPDD | Laboratory of Respiratory Viruses and Measles, Oswaldo Cruz Institute, FIOCRUZ | Alice Sampaio Rocha; Ana Carolina Mendonca; Anna Carolina Paixao; Elisa Cavalcante Pereira; Fernando Motta; Grey Benoit Vasquez; Isaac Miguel Sanchez; Ivonne Imbert; Lucia de la Cruz; Luciana Appolinario; Marilda Siqueira on behalf of the Fiocruz COVID-19 Genomic Surveillance Network; Nurys de Castro; Paola Resende; Renata Serrano Lopes; Ronald Skewes; Taina Venas |
| EPI_ISL_2648244, EPI_ISL_2648250, EPI_ISL_2648251, EPI_ISL_2648252, EPI_ISL_2648256, EPI_ISL_2648260, EPI_ISL_2650523, EPI_ISL_2650524, EPI_ISL_2650529, EPI_ISL_2650533, EPI_ISL_2650534, EPI_ISL_2650535, EPI_ISL_2650537 | | | |
| see above | Laboratorio Nacional de Vigilancia de la Salud – Sección de Virologia | Genomics and Proteomics Departament, Gorgas Memorial Institute For Health Studies | Alexander Martinez; Ambar Moreno; Claudia Díaz; Claudia Gonzalez; Elda Martínez; Jessica Gondola; Leyda Abrego; Marlene Castillo; Mitzi Castro; Nelly Amador; Oris Chavarria; Sandra Paola Paz; Sofia Carolina Alvarado |
| EPI_ISL_1091264 | Laboratorio PGM | Laboratorio de Infectología Molecular, Departamento de Bioquímica y Medicina Molecular,Facultad de Medicina - Universidad Autónoma de Nuevo León | Ana M. Rivas-Estilla; Daniel Arellanos-Soto; Eduardo Garza-de-la-Peña; Gabriela Elizondo; Javier Ramos-Jimenez; Kame A. Galán-Huerta; María F. Herrera-Saldivar; Natalia Martínez-Acuña; Sonia A. Lozano-Sepúlveda |
| EPI_ISL_1396389 | Laboratorio Salud Publica Mendoza | Nodo de Secuenciación Tierra del Fuego - Hospital Regional Ushuaia - Centro Austral De Investigaciones Científicas - Universidad Nacional De Tierra Del Fuego on behalf of 'Proyecto Argentino Interinstitucional de genomica de SARS-CoV-2' (PAIS Consortium) | Aguirre Carolina; Alberto Carena; Alejandro Ezequiel Rojas; Andrea Falaschi; Belén Ortiz; Bosio Lia; Carlos Espul; Cristian Garay; Cristina Fernanda Nardi; Fernando Gallego; Fernando Giuliani; Ivan Dario Gramundi; Luciana Martinez; Maria Belen Peralta Roca; Patricia Robledo; Santiago Guillermo Ceballos; Silvia Zerrer; Silvina Denita |
| EPI_ISL_2462063 | Laboratorio de Biologia Molecular, Hospital San Pedro Claver | Microbiologia Molecular, Instituto SELADIS, Universidad Mayor de San Andrés | Aneth Vasquez Michel; Carmen Delgado Barrera; Oscar M. Rollano-Peñaloza; Sandra Miranda Sardon |
| EPI_ISL_2600378 | Laboratorio de Biologia Molecular, Hospital San Pedro Claver | Molecular Genetics Laboratory, Instituto de Investigaciones Químicas, Universidad Mayor de San Andrés | Aneth Vasquez Michel; Carmen Delgado Barrera; Oscar M. Rollano-Peñaloza; Sandra Miranda Sardon |
| EPI_ISL_1278284 | Laboratorio de Biologia Molecular, SEDES-Potosi | Molecular Genetics Laboratory, Instituto de Investigaciones Químicas, Universidad Mayor de San Andrés | Aneth Vasquez Michel; Oscar M. Rollano-Peñaloza |
| EPI_ISL_457941, EPI_ISL_457956, EPI_ISL_457965 | Laboratorio de Biología Molecular Asociación Española Primera en Salud | Departments of Pathology and Medicine, New York University School of Medicine | Adriana Heguy; Christian Marier; Gael Westby; Gonzalo Manrique; Maria Noel Zubillaga; Maria Victoria Elizondo; Matthew T Maurano; Paul Zappile |
| EPI_ISL_2427504, EPI_ISL_2427581, EPI_ISL_2427601, EPI_ISL_2427614, EPI_ISL_2427669, EPI_ISL_2427683, EPI_ISL_2427694, EPI_ISL_2427702, EPI_ISL_2427705, EPI_ISL_2427711, EPI_ISL_2427714, EPI_ISL_2427724, EPI_ISL_2427733, EPI_ISL_2427751, EPI_ISL_2427752, EPI_ISL_2427768, EPI_ISL_2427769, EPI_ISL_2427770, EPI_ISL_2427772, EPI_ISL_2427774, EPI_ISL_2427775, EPI_ISL_2427777, EPI_ISL_2427779 | | | |
| see above | Laboratorio de Biología Molecular Médica Uruguaya | Departments of Pathology and Medicine, New York University School of Medicine | Adriana Heguy; Cecilia Sorhouet; Christian Marier; Dacia Dimartino; Gonzalo Manrique; Maria Cristina Mogdasy; Maria Noel Zubillaga; Maria Victoria Elizondo; Paul Zappile |
| EPI_ISL_626549 | Laboratorio de Biología Molecular, Facultad de Medicina, Universidad de Atacama, Copiapo, Chile/ FONDAP CRG, Universidad Andrés Bello, Santiago, Chile | Center for Mathematical Modeling and Center for Genome Regulation. Santiago, Chile | Allende ML; Bastias M; Castro E; Echeverría C; González M; M; Maass A; Manríquez R; Meneses C.; Montecino; Orellana A; Sanhueza D; Travisany D |
| EPI_ISL_1068097 | Laboratorio de Ecologia de Doencas Transmissiveis na Amazonia, Instituto Leonidas e Maria Deane - Fiocruz Amazonia | Laboratorio de Ecologia de Doencas Transmissiveis na Amazonia, Instituto Leonidas e Maria Deane - Fiocruz Amazonia | André Corado; Debora Duarte; Felipe Naveca on behalf of the Fiocruz COVID-19 Genomic Surveillance Network; Fernanda Nascimento; George Silva; Karina Pessoa; Luciana Gonçalves; Maria Júlia Brandão; Matilde Mejía; Michele Jesus; Valdinete Nascimento; Victor Souza; Ágatha Costa |
| EPI_ISL_1629797 | Laboratorio de Genómica Microbiana, Universidad Peruana Cayetano Heredia | Laboratorio de Genómica Microbiana, Universidad Peruana Cayetano Heredia | Alejandra Dávila-Barclay; Diego Cuicapuza; Guillermo Salvatierra; Janet Huancachoque; Lenin Maturrano; Luis González; Pablo Tsukayama; Pedro E. Romero; Pool Marcos |
| EPI_ISL_779182, EPI_ISL_961785 | Laboratorio de Infectología, Servicio de Infectología, Hospital Universitario Dr. José Eleuterio González - Universidad Autónoma de Nuevo León | Laboratorio de Infectología Molecular, Departamento de Bioquímica y Medicina Molecular,Facultad de Medicina - Universidad Autónoma de Nuevo León | Adrian Camacho-Ortiz; Ana M. Rivas-Estilla; Daniel Arellanos-Soto; Eduardo Perez-Alba; Elvira Garza-González; Kame A. Galán-Huerta; Laura Nuzzolo-Shihadeh; María F. Herrera-Saldivar; Natalia Martínez-Acuña; Paola Bocanegra-Ibarias; Samantha M. Flores-Treviño; Sonia A. Lozano-Sepúlveda |
| EPI_ISL_953404 | Laboratorio de Investigaciones de Baney | "Swiss Tropical and Public Health Institute" | "Carlos Cortes; Bonifacio Manguire Nlavo; Claudia Daubenberger; Diosdado Odjama Nseng Ada; Elizabeth Nyakarungu; Guillermo Garcia; Maximilian Mpina; Mitoha Ondo O Ayekaba; Philip Wonder Phiri"; Salome Hosch; Tobias Schindler |
| EPI_ISL_1672556, EPI_ISL_2002672, EPI_ISL_2002686 | Laboratorio de Investigaciones de Baney | Swiss Tropical and Public Health Institute | Bonifacio Manguire Nlavo; Carlos Cortes; Claudia Daubenberger; Diosdado Odjama Nseng Ada; Elizabeth Nyakarungu; Guillermo Garcia; Maximilian Mpina; Mitoha Ondo O Ayekaba; Philip Wonder Phiri; Philipp Wagner; Salome Hosch; Tobias Schindler; Yahya Maidane |
| EPI_ISL_648307, EPI_ISL_648318, EPI_ISL_648341, EPI_ISL_648350, EPI_ISL_648356, EPI_ISL_648372, EPI_ISL_648379, EPI_ISL_649171, EPI_ISL_649172 | | | |
| see above | Laboratorio de Investigaciones de Baney | University Hospital Basel, Clinical Bacteriology | Adrian Egli; Alfredo Mari; Bonifacio Manguire Nlavo; Carlos Cortes; Claudia Daubenberger; Diosdado Odjama Nseng Ada; Elizabeth Nyakarungu; Guillermo Garcia; Helena Seth-Smith; Madlen Stange; Maximilian Mpina; Mitoha Ondo O Ayekaba; Philip Wonder Phiri; Salome Hosch; Tim Roloff; Tobias Schindler |
| EPI_ISL_2854117, EPI_ISL_2854145 | Laboratorio de Medicina Molecular, Universidad de Magallanes | Centro Asistencial Docente y de Investigacion, Universidad de Magallanes | Constanza Ceroni; Diego Alvarez; Jacqueline Aldridge; Jorge González; Marcelo Navarrete; Roberto Uribe-Paredes |
| EPI_ISL_2462066 | Laboratorio de Referencia Departamental en Inmunología, Sedes-Pando | Molecular Genetics Laboratory, Instituto de Investigaciones Químicas, Universidad Mayor de San Andrés | Aneth Vasquez Michel; Carmen Delgado Barrera; Oscar M. Rollano-Peñaloza; Sandra Miranda Sardon |
| EPI_ISL_524473 | Laboratorio de Referencia Nacional de Virus Respiratorio. Centro Nacional de Salud Publica. Instituto Nacional de Salud Peru. | Laboratorio de Referencia Nacional de Biotecnología y Biología Molecular. Centro Nacional de Salud Publica. Instituto Nacional de Salud Peru. | Carlos Padilla Rojas; Henri Bailon Calderon; Johanna Balbuena Torres; Karolyn Vega Chozo; Marco Galarza Perez; Maribel Huaringa Nuñez; Nancy Rojas Serrano.; Omar Caceres Rey; Priscila Lope Pari |
| EPI_ISL_1575438 | Laboratorio de Referencia Nacional de Virus Respiratorio. Instituto Nacional de Salud Peru | Laboratorio de Referencia Nacional de Biotecnología y Biología Molecular. Instituto Nacional de Salud Peru | Carlos Padilla Rojas; Henri Bailon Calderon; Johanna Balbuena Torrez; Karolyn Vega Chozo; Luis Barcena; Marco Galarza Perez; Maribel Huaringa Nuñez; Nancy Rojas Serrano; Omar Caceres Rey; Priscila Lope Pari |
| EPI_ISL_491429, EPI_ISL_1532220 | Laboratorio de Referencia Nacional de Virus Respiratorio. Instituto Nacional de Salud Perú | Laboratorio de Referencia Nacional de Biotecnología y Biología Molecular. Instituto Nacional de Salud Perú | Carlos Padilla Rojas; Henri Bailon Calderon; Johanna Balbuena Torrez; Karolyn Vega Chozo; Luis Barcena; Marco Galarza Perez; Maribel Huaringa Nuñez; Nancy Rojas Serrano; Omar Caceres Rey; Priscila Lope Pari |
| EPI_ISL_1111143, EPI_ISL_1111223, EPI_ISL_1111249, EPI_ISL_1111480, | Laboratorio de Referencia Nacional de Virus Respiratorio. Instituto Nacional de Salud Perú | Laboratorio de Referencia Nacional de Enteropatógenos. Instituto Nacional de Salud del Perú | Fiorella Orellana Peralta; Iris Silva Molina; Junior Caro Castro; Ronnie Gavilan Chavez; Veronica Hurtado Vela; Willi Quino Sifuentes |

| | | | |
|---|---|---|---|
| EPI_ISL_1111501 | | | |
| EPI_ISL_729872, EPI_ISL_729879, EPI_ISL_812456 | Laboratorio de Referencia Nacional de Virus Respiratorios, Instituto Nacional de Salud Peru | Laboratorio de Genómica Microbiana, Universidad Peruana Cayetano Heredia | Alejandra Dávila-Barclay; Brenda Ayzanoa; Camila Castillo-Vilcahuamán; Guillermo Salvatierra; Janet Huancachoque; Luis González; Marco Galarza; Nancy Rojas; Oscar Escalante; Pablo Tsukayama; Pedro E. Romero; Pool Marcos; Priscila Lope |
| EPI_ISL_2536706, EPI_ISL_2536739, EPI_ISL_2536758, EPI_ISL_2536790, EPI_ISL_2536791, EPI_ISL_2536794, EPI_ISL_2536840, EPI_ISL_2536845, EPI_ISL_2921339, EPI_ISL_2921345, EPI_ISL_2921359, EPI_ISL_2921372, EPI_ISL_2921373, EPI_ISL_2921385, EPI_ISL_2921427, EPI_ISL_2921474, EPI_ISL_3023412, EPI_ISL_3023495, EPI_ISL_3023554, EPI_ISL_3023587, EPI_ISL_3023618 | | | |
| see above | Laboratorio de Referencial Nacional de Virus Respiratorios | Laboratorio de Referencial Nacional de Virus Respiratorios | Carlos Padilla Rojas; Henri Bailon Calderon; Iris Silva Molina; Joseph Huayra Niquen; Lely Solari Zerpa; Luis Barcena Flores; Marco Galarza Perez; Nancy Rojas Serrano; Omar Caceres Rey; Orson Mestanza Millones; Priscila Lope Pari; Sandra Morales Ruiz; Steve Acedo Lazo; Veronica Hurtado Vela |
| EPI_ISL_1738785, EPI_ISL_1738789 | Laboratorio de Salud Pública Bogota | Gencore - Universidad de los Andes | Alejandro Gomez; Ana Maria Palacio; David Gonzalez; Gabriela Delgado; Johana Hernandez; Luisa Sacristan; Marcela Guevara; Silvia Restrepo |
| EPI_ISL_717827 | Laboratorio de Virologia Molecular / UFRJ | Bioinformatics Laboratory / LNCC | Alexandra L Gerber; Amilcar Tanuri; Ana Paula de C Guimarães; Ana Tereza R de Vasconcelos; Andréa Cony Cavalcanti; Carolina M Voloch; Claudia dos Santos Rodrigues; Cynthia C Cardoso; Diana Mariani; Luiz G P de Almeida; Otavio Bustrolini; Ronaldo da Silva F Jr; Terezinha M P P Castiñeira |
| EPI_ISL_792399 | Laboratorio de Virología del Hospital de Niños Dr. Ricardo Gutierrez | Biocódices SA. on behalf of 'Proyecto Argentino Interinstitucional de genomica de SARS-CoV-2' (PAIS Consortium) | Acevedo; Alexay; Alvarez Lopez; Berros JM; C; Dopazo H; E; Gravis; Jacques; ME; Mistchenko; O; S; Zubrzycki J |
| EPI_ISL_1396198, EPI_ISL_1396199, EPI_ISL_2007487 | Laboratorio de Virología del Hospital de Niños Dr. Ricardo Gutierrez | Área de Secuenciación del Laboratorio de Virología del Hospital de Niños Dr. Ricardo Gutierrez on behalf of 'Proyecto Argentino Interinstitucional de genomica de SARS-CoV-2' (PAIS Consortium) | A; Acevedo; Acuña; Alexay; Alvarez Lopez; Barreda Frank; C; D; E; G; Goya; Grandis; Jacques; LE; Labarta; Lusso; M; ME; MI; Medina; Mistchenko; N; Nabaes Jodar; Natale; O; S; Streitenberger; Thomas; Valinotto; Viegas, M.; Villegas |
| EPI_ISL_1760549 | Laboratorio di Microbiologia | Laboratorio di Microbiologia | Martinetti Lucchini Gladys; Valeria Spina |
| EPI_ISL_1239601 | Laboratorium Badawczo-Rozwojowe Vitagenum | 1. National Institute of Public Health - National Institute of Hygiene; 2. Eurofins Genomics Europe Sequencing GmbH | ECDC COVID-19 WGS support team; Eurofins Genomics Europe Sequencing Team; Gierczyński Rafał; Sadkowska-Todys Małgorzata; Wołkowicz Tomasz; Zacharczuk Katarzyna |
| EPI_ISL_882960 | Laboratorium Diagnostyi Mikrobiologicznej z Pracownią Prątka Gruźlicy SPSzW im. Jana Bożego w Lublinie | National Institute of Public Health - National Institute of Hygiene | Gawor Jan; Wołkowicz Tomasz; Zacharczuk Katarzyna |
| EPI_ISL_1891882 | Laboratorium Mikrobiologiczne Dolnośląskie Centrum Transplantacji Komórkowych z Krajowym Bankiem Dawców Szpiku | 1. Academic Center for Pathomorphological and Genetic-Molecular Diagnostics ltd, Bialystok, Poland 2. National Institute of Public Health - National Institute of Hygiene, Warsaw, Poland | Anetta Sulewska; Jacek Nikliński; Janusz Dzięcioł; Joanna Kiśluk; Katarzyna Zacharczuk; Konrad Raczkowski; Magdalena Nowakowska; Małgorzata Sadkowska-Todys; Piotr Karabowicz; Piotr Majewski; Przemysław Biecek. Joanna Reszeć; Radosław Charkiewicz; Tomasz Wołkowicz |
| EPI_ISL_1612196, EPI_ISL_1681431, EPI_ISL_1681649, EPI_ISL_1681687, EPI_ISL_1685076, EPI_ISL_1799239, EPI_ISL_1800114, EPI_ISL_1802363, EPI_ISL_1802595, EPI_ISL_1926483, EPI_ISL_1928869, EPI_ISL_1931457, EPI_ISL_1931706, EPI_ISL_2045663, EPI_ISL_2184581, EPI_ISL_2305197, EPI_ISL_2305215, EPI_ISL_2305316, EPI_ISL_2305435, EPI_ISL_2305757, EPI_ISL_2306269, EPI_ISL_2398872, EPI_ISL_2398939, EPI_ISL_2480633, EPI_ISL_2481359, EPI_ISL_2481449, EPI_ISL_2481857, EPI_ISL_2611208, EPI_ISL_2611644, EPI_ISL_2872882, EPI_ISL_2872931, EPI_ISL_2927056, EPI_ISL_3017270 | | | |
| see above | Laboratory Corporation of America | Centers for Disease Control and Prevention Division of Viral Diseases, Pathogen Discovery | Adrian Paskey; Amanda Douglas; Amanda Suchanek; Andrea Throop; Ayla Burns; Benjamin Rambo-Martin; Bobbi Croy; Brian Krueger; Brian Norvell; Christopher Gulvick; Christos Petropoulos; Clinton R. Paden; Craig Lukasik; Dakota Howard; Darlene Wagner; Debbie Boles; Dhwani Batra; Duncan MacCannell; Eyad Almasri; Goran Stevovic; Howard Engler; Hrushikesh Deshmukh; Jake Humphrey; Jana Schroth; Jason Caravas; Joe Voshell; John Pruitt; Jonathan Meltzer; Jonathan Williams; Kara Moser; Kimberly Wagner; Lax Iyer; Lisa Pfefferle; Lyndon Tilson; Manoj Jain; Marcia Eisenberg; Mary Ann Cristobal; Mary Williamson; Matthew Robinson; Matthew Schmerer; Michael Levandoski; Mike Sapeta; Mindy Nye; Minoo Agarwal; Mohan Kolli; Nuthawin Charoensri; Oren Cohen; Peter W. Cook; Prashant Gupta; Qian Zeng; Rama Ghatti; Scott Parker; Scott Ryan; Scott Sammons; Shatavia Morrison; Stanley Letovsky; Steven Ragan; Suresh Babu Selvaraju; Susan Countryman; Susan Hicks; Suzanne Dale; Thomas Urban; Tim Kuphal; Tricia Zwiefelhofer; Vincent Drouillon; Yvette Unoarumhi |
| EPI_ISL_1021332, EPI_ISL_1026027, EPI_ISL_1029613 | Laboratory Corporation of America | Respiratory Viruses Branch, Division of Viral Diseases, Centers for Disease Control and Prevention | Ben L. Rambo-Martin; Clinton R. Paden; Dakota Howard; Dhwani Batra; Duncan MacCannell; Peter W. Cook; Suxiang Tong |
| EPI_ISL_582530, EPI_ISL_3102050 | Laboratory Diagnostic, Veterinary Specialized Institute Kraljevo | Laboratory Diagnostic, Veterinary Specialized Institute Kraljevo | Afonso, C.; Banovic Djeri, B.; Djukic, V.; Jankovic, M.; Jovanovic, T.; Knezevic, A.; Petrovic, T.; Sekler, M.; Tesovic, B.; Vidanovic, D.; Volkening, J. |
| EPI_ISL_754904, EPI_ISL_754906 | Laboratory Diagnostics and Clinical Immunology of Developmental Age, Medical University of Warsaw | genXone SA, Research & Development Laboratory; The Faculty of Mathematics, Informatics and Mechanics of the University of Warsaw | Anna Gambin; Grzegorz Nowicki; Jakub Grabowski; Maciej Sykulski; Michał Kaszuba; Monika Mańkowska-Woźniak; Natalia Drwęska-Matelska; Urszula Demkow; Łukasz Krych |
| EPI_ISL_667809 | Laboratory Medicine | Department of Laboratory Medicine, Lin-Kou Chang Gung Memorial Hospital, Taoyuan, Taiwan | Cheng-Hsun Chiu; Cheng-Ta Yang; Chung-Guei Huang; Guang-Wu Chen; Kuo-Chien Tsao; Kuo-Ming Lee; Mei-Jen Hsiao; Peng-Nien Huang; Po-Wei Huang; Shin-Ru Shih; Shu-Li Yang; Yi-Chun Liu; Yu-Nong Gong |
| EPI_ISL_2467938 | Laboratory for HIV and opportunistic infections diagnosis The Republican Research and Practical Center for Epidemiology and Microbiology (RRPCEM) | Laboratory for HIV and opportunistic infections diagnosis The Republican Research and Practical Center for Epidemiology and Microbiology (RRPCEM | Alina Drozd; Artur Akhremchuk; Elena Gasich; Hanna Gudel; Katsiaryna Belyakova; Kirill Bulda; Leonid Valentovich; Nastassia Kabankova |
| EPI_ISL_933533, EPI_ISL_1138899, EPI_ISL_1653933, EPI_ISL_2080973 | Laboratory for HIV and opportunistic infections diagnosis The Republican Research and Practical Center for Epidemiology and Microbiology (RRPCEM) | Laboratory for HIV and opportunistic infections diagnosis The Republican Research and Practical Center for Epidemiology and Microbiology (RRPCEM) | Alena Mikhalenka; Alexander Kilchevcky; Alina Drozd; Anatoly Krasko; Anna Gudel; Artur Akhremchuk; Elena Gasich; Katsiaryna Belyakova; Kirill Bulda; Leonid Valentovich; Leonid Valentovich; Nastassia Kabankova; Vladimir Gorbunov; Yauhen Sysaliatsin |
| EPI_ISL_471421 | Laboratory for Respiratory Viruses, National Influenza Centre, Cantacuzino National Military-Medical Institute for Research and Development | Cantacuzino Institute | Luiza Ustea; Mihaela Lazar; Nicoleta Paraschiv; Tim Durfee |
| EPI_ISL_435045 | Laboratory of Applied Genetics | RSE "National Center for Biotechnology" | Alexandr Shevtsov; Asylulan Amirgazin; Ilyas Akhmetollayev; Ruslan Kalendar; Viktoriya Lutsay; Yerlan Ramanculov |
| EPI_ISL_2832105, EPI_ISL_2832106 | Laboratory of Genetics and Personalized Medicine, Zan Mitrev Clinic | Laboratory of Genetics and Personalized Medicine, Zan Mitrev Clinic | Kungulovski G. et al. |
| EPI_ISL_2349110 | Laboratory of Immunohematology, Division of Hematology | Greek Genome Center, Biomedical Research Foundation of the Academy of Athens (BRFAA) | Athanasia Mouzaki; Dimitrios Thanos; Emmanouil Athanasiadis; Giannis Vatsellas; Katerina Zoi; Theodoros Loupis |
| EPI_ISL_2312569, EPI_ISL_2344050, EPI_ISL_2344165 | Laboratory of Immunohematology, Division of Hematology | Greek Genome Center, Biomedical Research Foundation of the Academy of Athens (BRFAA) | Athanasia Mouzaki; Dimitrios Thanos; Emmanouil Athanasiadis; Giannis Vatsellas; Katerina Zoi; Theodoros Loupis |
| EPI_ISL_434456, EPI_ISL_437895 | Laboratory of Microbiology, Medical School, National and Kapodistrian University of Athens | Laboratory of Biology, Department of Medicine, Democritus University of Thrace | Bampali, M.; Dovrolis, N.; Froukala, E.; Gatzidou, E.; Kassela K.; N. and Karakasiliotis, I.; Spanakis; Stavropoulou, A.; Tsakris, A.; Veletza, S. |
| EPI_ISL_654016, EPI_ISL_654017, EPI_ISL_2896981 | Laboratory of Microbiology, National Reference Lab, Charles Nicolle Hospital; 2-University of Tunis ElManar, Faculty of Medicine of Tunis, LR99ES09, Tunis, Tunisia | 1-Clinical and Experimental Pharmacology Lab, LR16SP02, National Center of Pharmacovigilance, University of Tunis El Manar, Tunis, Tunisia. 2-Neurodegenerative diseases and psychiatric troubles, LR18SP03, Razi Hospital, University of Tunis El Manar, Tunis, Tunisia. 3- Ministry of Health, National Observatory of New and Emerging Diseases, 1006, Tunis, Tunisia | Alia Ben Kahla; Alia Benkahla; Emna Gaies; Gaies Emna; Ilhem Boutiba-Ben Boubaker; Ilhem Boutiba-Ben Boubaker.; Imen Kacem; Imen Mkada; Ines Mdini; Jalila Ben Khelil; Maher Kharrat; Mouna Ben Sassi; Mouna Safer; Nissaf Ben Alaya; Riadh Daghfous; Riadh Gouider; Riadh Gouider.; Roua Ben Othman; Salma Abid; Sameh Trabelsi; Sana Ferjani; Sarra Chammam; Soumaya Rammeh |
| EPI_ISL_803119, EPI_ISL_2153433, EPI_ISL_2154331 | Laboratory of Microbiology, National Reference Lab, Charles Nicolle Hospital; 2-University of Tunis ElManar, Faculty of Medicine of Tunis, LR99ES09, Tunis, Tunisia | Clinical and Experimental Pharmacology Lab, LR16SP02, National Center of Pharmacovigilance, University of Tunis El Manar, Tunis, Tunisia. 2-Neurodegenerative diseases and psychiatric troubles, LR18SP03, Razi Hospital, University of Tunis El Manar, Tunis, Tunisia. 3- Ministry of Health, National Observatory of New and Emerging Diseases, 1006, Tunis, Tunisia | Ahmed Fakhfakh; Alia BenKahla; Gaies Emna; Habiba Ben Romdhane; Hanen El Jebari; Ilhem Boutiba-Ben Boubaker; Ilhem Boutiba-Ben Boubaker.; Imen Kacem; Imen Mdini; Jalila Ben Khelil; Maher Kharrat; Manel Ben Sassi; Mouna Ben Sassi; Mouna Safer; Nissaf Ben Alaya; Raja Mahfoudh; Riadh Daghfous; Riadh Gouider; Riadh Gouider.; Roua Ben Othman; Salma Abid; Sameh Trabelsi; Sana Ferjani; Sarra Chamman; Souissi Amira; Taha Maatoug; Zaineb Hamzaoui |
| EPI_ISL_2893208, EPI_ISL_2893213, EPI_ISL_2893214, EPI_ISL_2893217, EPI_ISL_2893218, EPI_ISL_2893219, EPI_ISL_2893220 | | | |
| see above | Laboratory of Molecular Biology and Cancer Immunology, Faculty of Sciences, Lebanese University | Microbial Pathogenomics Lab - LAU | Bassam Badran; Fadi Abdel Sater; Georgi Merhi; Hamad Hassan; Jad Koweyes; Rawan Makki; Sima tokajian |
| EPI_ISL_450746 | Laboratory of Molecular Biology, Diagnostyka sp. z o.o. | Laboratory of Recombinant Vaccines | Anna Piotrowska-Mietelska; Boguslaw Szewczyk; Krystyna Bienkowska-Szewczyk; Lukasz Rabalski; Maciej Kosinski |
| EPI_ISL_2301773, EPI_ISL_2301776, EPI_ISL_2312615 | Laboratory of Molecular Biology, Mamatsio General Hospital of Kozani | Greek Genome Center, Biomedical Research Foundation of the Academy of Athens (BRFAA) | Dimitrios Thanos; Emmanouil Athanasiadis; Giannis Vatsellas; Katerina Zoi; Konstantina Gartzonika; Theodoros Loupis |
| EPI_ISL_801607, EPI_ISL_801609 | Laboratory of Molecular Virology, Pontificia Universidad Católica de Chile | MSHS Pathogen Surveillance Program | Adolfo Garcia-Sastre; Adriana van De Guchte; Ajay Obla; Ana Maria Contreras; Ana S. Gonzalez-Reiche; Bremy Alburquerque; Carlos Palma; Constanza Maldonado; Edward C. Holmes; Eileen Serrano; Erick Salinas; Hala Alshammary; Harm van Bakel; Jayeeta Dutta; Jorge Levican; Juan Soto; Leonardo I. Almonacid; M. Belen Leyton; Marcela Ferres; Matthew M. Hernandez; Melissa Smith; Rafael A. Medina.; Robert Sebra; Shwetha Hara Sridhar; Tamara García-Salum; Viviana Simon; Ying-Chih Wang; Zenab Khan |
| EPI_ISL_2557397 | Laboratory of Respiratory Viruses and Measles, Oswaldo Cruz Institute, FIOCRUZ | Laboratory of Respiratory Viruses and Measles, Oswaldo Cruz Institute, FIOCRUZ | Alice Sampaio Rocha; Ana Carolina Mendonca; Anna Carolina Paixao; Elisa Cavalcante Pereira; Fernando Motta; Luciana Appolinario; Marilda Siqueira on behalf of the Fiocruz COVID-19 Genomic Surveillance Network; Paola Resende; Renata Serrano Lopes; Taina Venas |
| EPI_ISL_1082301, EPI_ISL_1082303 | Laboratory of Virology of Federal Budget Health Care Institution "Center of Hygiene and Epidemiology in Arkhangelsk region" | Group of Genomics and Postgenomic Technologies of Central Research Institute of Epidemiology | Akimkin VG; Bulanenko VP; Cherkashina AS; Golubeva AG; Kaptelova VV; Kondrasheva LV; Korneenko EV; Ponomareva YA; Saenko SS; Samoilov AE; Semyina LV; Shishko LA; Speranskaya AS; Tivanova EV; Valdokhina AV |
| EPI_ISL_1341503, EPI_ISL_1489929 | Laboratory of Virology, National center of expertise | RSE "National Center of Expertise" and RSE "National center for Biotechnology" | Abdaliyev Askar; Amirgazin Asylulan; Balykbaev Kanat; Kamalova Dinara; Ramankulov Erlan; Sharipova Saule; Shevtsov Alexandr; Tungushbayev Talgat |

| | | | |
|---|---|---|---|
| EPI_ISL_1191740 | Laboratory of virology and molecular diagnostics | Laboratory of virology and molecular diagnostics | Boshevska Golubinka; Janchevska Elizabeta; Kuzmanovska Maja |
| EPI_ISL_2392161, EPI_ISL_2681400, EPI_ISL_2681403, EPI_ISL_2681404, EPI_ISL_2681431, EPI_ISL_2681436, EPI_ISL_2681438, EPI_ISL_2987553, EPI_ISL_2987557, EPI_ISL_2987571, EPI_ISL_2987574 | | | |
| see above | Laboratory of virology and molecular diagnostics, Institute of Public Health | Laboratory of virology and molecular diagnostics, Institute of Public Health | Boshevska G; Janchevska E.; Kuzmanovska M |
| EPI_ISL_454585 | Laboratory of virology, National Center of Expertise | Laboratory of molecular-genetic research, National Center for Expertise, Kazakhstan National Center for Biotechnology, Kazakhstan | ; Abdaliyev Askar; Akhmetollayev Ilyas; Amirgazin Asylulan; Aushakhmetova Zabira; Kalendar Ruslan; Lutsay Viktoriya; Rakhmetova Akbota; Ramankulov Yerlan; Shevtsov Alexandr |
| EPI_ISL_1364615, EPI_ISL_1448018 | Laboratory of virology, National center of expertise | RSE "National Center for Biotechnology" and RSE "National Center of Expertise" | Abdaliyev Askar; Amirgazin Asylulan; Balykbaev Kanat; Kamalova Dinara; Ramankulov Yerlan; Sharipova Saule; Shevtsov Alexandr; Tungushbayev Talgat |
| EPI_ISL_2696437, EPI_ISL_2696689 | Laboratory, Gravity Diagnostics | Laboratory, Gravity Diagnostics | Diagnostics, G. |
| EPI_ISL_576119 | Laboratory, The Bio Arte Limited | Laboratory, The Bio Arte Limited | Biazzo, M.; Madeddu, S.; Pinzauti, D.; Santoro, F. |
| EPI_ISL_2157506 | Laboratório Central de Saude Publica do Estado do Paraná (LACEN/PR) | Laboratory of Respiratory Viruses and Measles, Oswaldo Cruz Institute, FIOCRUZ | Alice Sampaio Rocha; Ana Carolina Mendonca; Anna Carolina Paixao; Elisa Cavalcante Pereira; Fernando Motta; Irina Nastassja Riediger; Luciana Appolinario; Maria do Carmo Debur; Marilda Siqueira on behalf of the Fiocruz COVID-19 Genomic Surveillance Network; Paola Resende; Renata Serrano Lopes; Taina Venas |
| EPI_ISL_2777330, EPI_ISL_2777341, EPI_ISL_2777778, EPI_ISL_2777954 | Laboratório Central de Saúde Pública do Amazonas - LACEN-AM | Laboratorio de Ecologia de Doencas Transmissiveis na Amazonia, Instituto Leonidas e Maria Deane - Fiocruz Amazonia | André Corado; Debora Duarte; Felipe Naveca; Fernanda Nascimento; George Silva; Karina Pessoa; Luciana Gonçalves; Maria Júlia Brandão; Matilde Mejía; Michele Jesus; Valdinete Nascimento; Victor Souza; Ágatha Costa |
| EPI_ISL_541387 | Laboratório Central de Saúde Pública do Estado de Sergipe (LACEN-SE) | Laboratory of Respiratory Viruses and Measles, Oswaldo Cruz Institute, FIOCRUZ | Ana Carolina Mendonça; Anna Carolina Paixão; Clioma Santos; Fernando Motta; Jonathan Lopes; Luciana Appolinario; Marilda Siqueira on behalf of the Fiocruz COVID-19 Genomic Surveillance Network; Paola Resende |
| EPI_ISL_415128 | Laboratório Central de Saúde Pública do Estado do Espírito Santo (LACEN-ES) | Laboratory of Respiratory Viruses and Measles, Oswaldo Cruz Institute, FIOCRUZ | Aline Mattos; Allison Fabri; Braulia Caetano; Cristiana Garcia; Fernando Motta; Joilson Xavier; Jonathan Lopes; Luciana Appolinario; Maria Nóbrega; Maria Ogrzewalska; Marilda Siqueira on behalf of the Fiocruz COVID-19 Genomic Surveillance Network; Milene Miranda; Paola Resende; Sunando Roy |
| EPI_ISL_729832 | Laboratório Central de Saúde Pública do Estado do Rio Grande do Sul (LACEN-RS) | Laboratory of Respiratory Viruses and Measles, Oswaldo Cruz Institute, FIOCRUZ | Ana Carolina Mendonça; Anna Carolina Paixão; Fernando Motta; Luciana Appolinario; Marilda Siqueira on behalf of the Fiocruz COVID-19 Genomic Surveillance Network; Marilda Tereza Mar da Rosa; Paola Resende; Tatiana Schaffer Gregianini |
| EPI_ISL_2150977, EPI_ISL_2158597, EPI_ISL_2158598, EPI_ISL_2158601 | Laboratório de Biologia Molecular Jean Piaget | MRCG at LSHTM, Genomics lab | Abdoulie Kanteh; Abdul Karim Sesay; Adul Candé; Aicha Balde; Aladje Balde; Bakary Sanyang; Bubacar Delgado Pinto Embaló; Dabiri Damilari; Edmira Maria da Costa; Erica Luis Maria Magalhães; Faatu Cassama; Mariama Kujabi; Milanca Agostinho Cá; Paulina Joãozinho da Costa Jarra Manneh; Rei José Pereira; Rui Inndi; Sainabou Laye Ndure; Simão Tchuda Bióté |
| EPI_ISL_2835169 | Laboratório de Biotecnologia Aplicada (LBA) - Laboratório de Biologia Molecular - Hospital das Clínicas, Faculdade de Medicina de Botucatu, Departamento de Bioprocessos e Biotecnologia - Faculdade de Ciências Agronômicas, UNESP – Botucatu/SP | Laboratory of Respiratory Viruses and Measles, Oswaldo Cruz Institute, FIOCRUZ | Alice Sampaio Rocha; Ana Carolina Mendonca; Anna Carolina Paixao; Elisa Cavalcante Pereira; Felipe Allan da Silva Costa; Fernando Motta; Jayme Augusto de Souza Neto; Leonardo Nazario de Moraes; Luciana Appolinario; Marilda Siqueira on behalf of the Fiocruz COVID-19 Genomic Surveillance Network; Paola Resende; Patricia Akemi Assato; Rejane Maria Tommasini; Renata Serrano Lopes; Taina Venas |
| EPI_ISL_832012 | Laboratório de Microbiologia Molecular - Universidade FEEVALE | Universidade Federal de Ciências da Saúde de Porto Alegre | Amanda de Menezes Mayer; Carla Andretta Moreira Neves; Claudia Elizabeth Thompson; Fernando Rosado Spilki; Gabriel Dickin Caldana; Gabriela Bettella Cybis; Lívia Kmetzsch; Patrícia Aline Gröhs Ferrareze; Ricardo Ariel Zimerman; Vinícius Bonetti Franceschi |
| EPI_ISL_1216912 | Labormedizin Darmstadt | Robert Koch Institute | |
| EPI_ISL_1233630, EPI_ISL_1233663, EPI_ISL_1299921, EPI_ISL_1914638, EPI_ISL_1973556, EPI_ISL_2274472, EPI_ISL_2484522, EPI_ISL_2768045, EPI_ISL_3030090 | | | |
| see above | Labormedizinisches Zentrum Dr Risch | Clinical Bacteriology | Adrian Egli; Alfredo Mari; Fanny Wegner; Hans Hirsch; Helena MB Seth-Smith; Julia Bielicki; Karoline Leuzinger; Lorenz Risch; Madlen Stange; Manuel Battegay; Martin Risch; Nadia Wohlwend; Tim Roloff |
| EPI_ISL_2677281, EPI_ISL_2677286 | Labortorio Central de Saude Publica do Estado de Santa Catarina (LACEN/SC) | Laboratory of Respiratory Viruses and Measles, Oswaldo Cruz Institute, FIOCRUZ | Alice Sampaio Rocha; Ana Carolina Mendonca; Anna Carolina Paixao; Darcita Buerger Rovaris; Elisa Cavalcante Pereira; Fernando Motta; Luciana Appolinario; Marilda Siqueira on behalf of the Fiocruz COVID-19 Genomic Surveillance Network; Paola Resende; Renata Serrano Lopes; Sandra Bianchini Fernandes; Taina Venas |
| EPI_ISL_660133 | Lancet | National Health Laboratory Service (NHLS), Tygerberg | Bronwyn Kleinhans; Davis M-A; Draper C; Eduan Wilkindon; Gert van Zyl; Houriiyah Tegally; Hsiao M; Kayla Delaney; Siegfried N; Susan Engelbrecht; Tulio de Oliveira; Williamson C; Wolfgang Preiser |
| EPI_ISL_2013052 | Lancet Laboratories | National Institute for Communicable Diseases of the National Health Laboratory Service | Amoako DG; Bhiman JN; Glass A; Mahlangu B; Mohale T; Ntuli N; Scheepers C; Viana R |
| EPI_ISL_738336, EPI_ISL_812275, EPI_ISL_812281 | Landstuhl Regional Medical Center | United States Air Force School of Aerospace Medicine | Amanda Javorina; Anthony Fries; Clarise Starr; Cole Anderson; Elizabeth Macias; Fritz Castillo; Jennifer Meyer; Sarah Purves; William Gruner |
| EPI_ISL_407079 | Lapland Central Hospital | Department of Virology, University of Helsinki and Helsinki University Hospital, Helsinki, Finland | Hannimari Kallio-Kokko; Olli Vapalahti; Suvi Kuivanen; Teemu Smura |
| EPI_ISL_2811989 | Laverty Pathology | NSW Health Pathology - Institute of Clinical Pathology and Medical Research; Westmead Hospital; University of Sydney | CIDM-PH et al. |
| EPI_ISL_2863684 | Lboratorio Central de Saude Publica do Estado do Parana (LACEN/PR) | Laboratory of Respiratory Viruses and Measles, Oswaldo Cruz Institute, FIOCRUZ | Agatha Cristinne Prudencio; Alice Sampaio Rocha; Ana Carolina Mendonca; Anna Carolina Paixao; Elisa Cavalcante Pereira; Fernando Motta; Ighor Leonardo Arantes Gomes; Irina Riediger; Luciana Appolinario; Marilda Siqueira on behalf of the Fiocruz COVID-19 Genomic Surveillance Network; Paola Resende; Renata Serrano Lopes; Taina Venas |
| EPI_ISL_2779329 | Life Care Hospital (Bungoma) | USAMRD-A, Basic Science Laboratory | Alan Lemtudo; Beth Mutai; Brian Andika; Carol Kifude; Clement Masakwe; Eric Muthanje; Esther Omuseni; Faith Sigei; Gathii Kimita; George Awinda; John Waitumbi; Josphat Nyataya; Rachel Githii; Rehema Liyai; Stephen Ochola |
| EPI_ISL_2136815 | Life Sciences Center, Vilnius University | Institute of Biotechnology, Life Sciences Center, Vilnius University | Albertas Timinskas; Alma Gedvilaite; Danguole Ziogiene; Emilija Vasiliunaite; Milda Norkiene |
| EPI_ISL_1510207 | Life Sciences Center, Vilnius University | Vilnius University Hospital Santaros Klinikos, Center of Laboratory Medicine | Daniel Naumovas; Dovile Ezerskyte; Gytis Dudas; Ingrida Olendraite; Laimonas Griskevicius; Ligita Raugaite; Mindaugas Stoskus; Monika Katenaite; Rimvydas Norvilas |
| EPI_ISL_701677, EPI_ISL_701777, EPI_ISL_703603, EPI_ISL_709416, EPI_ISL_720734, EPI_ISL_720848, EPI_ISL_720858, EPI_ISL_731623, EPI_ISL_800014, EPI_ISL_835513, EPI_ISL_1256078, EPI_ISL_1414488, EPI_ISL_1415461, EPI_ISL_1415755, EPI_ISL_1415960, EPI_ISL_1594716, EPI_ISL_1594784, EPI_ISL_1653511, EPI_ISL_1699106, EPI_ISL_2005299, EPI_ISL_2468924, EPI_ISL_2553223, EPI_ISL_2995319 | | | |
| see above | Lighthouse Lab in Alderley Park | Wellcome Sanger Institute for the COVID-19 Genomics UK (COG-UK) Consortium | Cordelia Langford; David K. Jackson; Dominic Kwiatkowski; Ewan Harrison; Ian Johnston; Jacquelyn Wynn; Jeffrey Barrett; John Sillitoe on behalf of the Wellcome Sanger Institute COVID-19 Surveillance Team; Mairead Hyland; Roberto Amato; Sonia Goncalves; The Lighthouse Lab in Alderley Park and Alex Alderton |
| EPI_ISL_597618, EPI_ISL_597757, EPI_ISL_606128, EPI_ISL_611066, EPI_ISL_625220 | Lighthouse Lab in Alderley Park | Wellcome Sanger Institute for the COVID-19 Genomics UK (COG-UK) consortium | Cordelia Langford; David K. Jackson; Dominic Kwiatkowski; Ewan Harrison; Ian Johnston; Jacquelyn Wynn; John Sillitoe on behalf of the Wellcome Sanger Institute COVID-19 Surveillance Team; John Sillitoe on behalf of the Wellcome Sanger Institute COVID-19 Surveillance Team (http://www.sanger.ac.uk/covid-team); Mairead Hyland; Roberto Amato; Sonia Goncalves; The Lighthouse Lab in Alderley Park and Alex Alderton |
| EPI_ISL_1263915, EPI_ISL_1275626, EPI_ISL_1275723, EPI_ISL_1275862, EPI_ISL_1276977, EPI_ISL_1316141, EPI_ISL_1316216, EPI_ISL_1316218, EPI_ISL_1327314, EPI_ISL_1327532, EPI_ISL_1327570, EPI_ISL_1329240, EPI_ISL_1332662, EPI_ISL_1332697, EPI_ISL_1409592, EPI_ISL_1409671, EPI_ISL_1467652, EPI_ISL_1473620, EPI_ISL_1538088, EPI_ISL_1584041, EPI_ISL_1652995, EPI_ISL_1719191, EPI_ISL_1831454 | | | |
| see above | Lighthouse Lab in Cambridge | Wellcome Sanger Institute for the COVID-19 Genomics UK (COG-UK) Consortium | Cordelia Langford; David K. Jackson; Dominic Kwiatkowski; Ewan Harrison; Ian Johnston; Jeffrey Barrett; John Sillitoe on behalf of the Wellcome Sanger Institute COVID-19 Surveillance Team; Rob Howes; Roberto Amato; Sonia Goncalves; The Lighthouse Lab in Cambridge and Alex Alderton |
| EPI_ISL_608558 | Lighthouse Lab in Cambridge | Wellcome Sanger Institute for the COVID-19 Genomics UK (COG-UK) consortium | Cordelia Langford; David K. Jackson; Dominic Kwiatkowski; Ewan Harrison; Ian Johnston; John Sillitoe on behalf of the Wellcome Sanger Institute COVID-19 Surveillance Team; Rob Howes; Roberto Amato; Sonia Goncalves; The Lighthouse Lab in Cambridge and Alex Alderton |
| EPI_ISL_720767, EPI_ISL_760686, EPI_ISL_760985, EPI_ISL_859475, EPI_ISL_989853, EPI_ISL_1084473, EPI_ISL_1333020, EPI_ISL_1333066, EPI_ISL_1333267, EPI_ISL_1333902, EPI_ISL_1413324, EPI_ISL_2960675 | | | |
| see above | Lighthouse Lab in Glasgow | Wellcome Sanger Institute for the COVID-19 Genomics UK (COG-UK) Consortium | Anna Dominiczak and Alex Alderton; Carol Clugston; Cordelia Langford; David Gray; David K. Jackson; Dominic Kwiatkowski; Ewan Harrison; Harper VanSteenhouse; Ian Johnston; Jeffrey Barrett; John Sillitoe on behalf of the Wellcome Sanger Institute COVID-19 Surveillance Team; John Sillitoe on behalf of the Wellcome Sanger Institute COVID-19 Surveillance Team (http://www.sanger.ac.uk/covid-team); Roberto Amato; Sonia Goncalves; Yumi Kasai |
| EPI_ISL_530530, EPI_ISL_530548, EPI_ISL_532638, EPI_ISL_532883, EPI_ISL_533001, EPI_ISL_533038, EPI_ISL_533069 | | | |
| see above | Lighthouse Lab in Glasgow | Wellcome Sanger Institute for the COVID-19 Genomics UK (COG-UK) consortium | Anna Dominiczak and Alex Alderton; Carol Clugston; Cordelia Langford; David Gray; David K. Jackson; Dominic Kwiatkowski; Ewan Harrison; Harper VanSteenhouse; Ian Johnston; John Sillitoe; Roberto Amato; Sonia Goncalves; Yumi Kasai |
| EPI_ISL_878751, EPI_ISL_1072347, EPI_ISL_1332568, EPI_ISL_1631640, EPI_ISL_1698027, EPI_ISL_1698076, EPI_ISL_2317665 | | | |
| see above | Lighthouse Lab in Milton Keynes | Wellcome Sanger Institute for the COVID-19 Genomics UK (COG-UK) Consortium | Cordelia Langford; David K. Jackson; Dominic Kwiatkowski; Ewan Harrison; Ian Johnston; Jeffrey Barrett; John Sillitoe on behalf of the Wellcome Sanger Institute COVID-19 Surveillance Team; Roberto Amato; Sonia Goncalves; The Lighthouse Lab in Milton Keynes and Alex Alderton |
| EPI_ISL_552471, EPI_ISL_553021, EPI_ISL_553243, EPI_ISL_557188, EPI_ISL_606702, EPI_ISL_606838 | Lighthouse Lab in Milton Keynes | Wellcome Sanger Institute for the COVID-19 Genomics UK (COG-UK) consortium | Cordelia Langford; David K. Jackson; Dominic Kwiatkowski; Ewan Harrison; Ian Johnston; John Sillitoe on behalf of the Wellcome Sanger Institute COVID-19 Surveillance Team; John Sillitoe on behalf of the Wellcome Sanger Institute COVID-19 Surveillance Team (http://www.sanger.ac.uk/covid-team); Roberto Amato; Sonia Goncalves; The Lighthouse Lab in Milton Keynes and Alex Alderton |
| EPI_ISL_2635692 | Limbach - MVZ Humangenetik Ulm | Robert Koch Institute | |
| EPI_ISL_603088, EPI_ISL_603119, EPI_ISL_636840, EPI_ISL_636842, EPI_ISL_636870, EPI_ISL_636880, EPI_ISL_636881, EPI_ISL_636888 | | | |
| see above | Lithuanian University of Health Sciences Hospital, Department of Laboratory Medicine | Lithuanian University of Health Sciences, Molecular cardiology lab. | Arnoldas Pautienius; Astra Vitkauskiene; Dovydas Gecys; Ingrida Olendraitė; Kamile Tamusauskaite; Laura Pareckaite; Lukas Zemaitis; Vaiva Lesauskaite |
| EPI_ISL_1502008 | Lurie Children's Hospital of Chicago | Northwestern University - Ozer Lab | Egon A. Ozer; Judd F. Hultquist; Lacy M. Simons; Larry K. Kociolek; Michael G. Ison; Ramon Lorenzo-Redondo; Taylor J. Dean; William J. Muller; Xiaotian; Zheng |

| | | | |
|---|---|---|---|
| EPI_ISL_2827933, EPI_ISL_2827939, EPI_ISL_2984851, EPI_ISL_2984857, EPI_ISL_3024551, EPI_ISL_3024553, EPI_ISL_3024567, EPI_ISL_3024568 | | | |
| see above | MANKWENG LABORATORY | National Institute for Communicable Diseases of the National Health Laboratory Service | Amoako DG; Bhiman JN; Everatt J; Ismail A; Mahlangu B; Mnguni A; Mohale T; Ntuli N; Scheepers C |
| EPI_ISL_1258302, EPI_ISL_1594077, EPI_ISL_2495486, EPI_ISL_2495574, EPI_ISL_2495647, EPI_ISL_2495669 | MB-Cadham Provincial laboratory | National Microbiology Laboratory (NML) | Anna Majer; Anneliese Landgraff; CanCOGeN's metadata curation team; Darian Hole; David Alexander; Elsie Grudeski; Gary Van Domselaar; Grace Seo; Jared Bullard; Jennifer Tanner; Kerry Dust; Kirsten Biggar; Madison Chapel; Morag Graham; Natalie Knox; Nathalie Bastien; Paul Van Caeseele; Philip Mabon; Public Health Agency of Canada CanCOGeN team; Rhiannon Huzarewich; Russell Mandes; Shari Tyson; Timothy Booth; Yan Li |
| EPI_ISL_1152607, EPI_ISL_1435428 | MDI Limbach Berlin GmbH | Robert Koch Institute | |
| EPI_ISL_2120501, EPI_ISL_2472756, EPI_ISL_2632870, EPI_ISL_2678707, EPI_ISL_2678719, EPI_ISL_2761877 | MDI Limbach Berlin GmbH; MVZ Labor Berlin | Robert Koch Institute | |
| EPI_ISL_568978, EPI_ISL_569028, EPI_ISL_569395, EPI_ISL_569496, EPI_ISL_900072, EPI_ISL_900176, EPI_ISL_1020363, EPI_ISL_1694792 | | | |
| see above | MEPHI, Aix Marseille University | MEPHI, Aix Marseille University | Anthony LEVASSEUR |
| EPI_ISL_2895751, EPI_ISL_2895753, EPI_ISL_2895755, EPI_ISL_2895756, EPI_ISL_2895757, EPI_ISL_2895759, EPI_ISL_2895760, EPI_ISL_2895763, EPI_ISL_2895764, EPI_ISL_2895765, EPI_ISL_2942282 | | | |
| see above | MIDDELBURG LABORATORY | National Institute for Communicable Diseases of the National Health Laboratory Service | Amoako DG; Bhiman JN; Everatt J; Ismail A; Mahlangu B; Mnguni A; Mohale T; Ntuli N; Scheepers C |
| EPI_ISL_2566568 | MKK Mühlenkreiskliniken, Johannes Wesling Klinikum Minden | Hannover Medical School, Institute of Virology | Lars Steinbrück |
| EPI_ISL_1226380 | MO State Public Health Laboratory | Genomics and Discovery, Respiratory Viruses Branch, Division of Viral Diseases, Centers for Disease Control and Prevention | Anna Montmayeur; Anna Uehara; Ben L. Rambo-Martin; Clinton R. Paden; Dhwani Batra; Haibin Wang; Jasmine Padilla; Jing Zhang; Justin Lee; Katie Dillon; Krista Queen; Kristen Knipe; Kristine Lacek; Lori Rowe; Mark Burroughs; Matthew Schmerer; Mili Sheth; Peter W. Cook; Rachel Marine; Sam Shepard; Sarah Nobles; Shoshona Le; Suxiang Tong; Yan Li; Ying Tao |
| EPI_ISL_2556117, EPI_ISL_2557251 | MOH - Jaber Al-Ahmad Hospital (Innovation Research Laboratory) | MOH - Jaber Al-Ahmad Hospital (Innovation Research Laboratory) | Mohammad Alghounaim; Salman Al-Sabah |
| EPI_ISL_2779435 | MOH-K | USAMRD-A, Basic Science Laboratory | Alan Lemtudo; Beth Mutai; Brian Andika; Carol Kifude; Clement Masakwe; Eric Muthanje; Esther Omuseni; Faith Sigei; Gathii Kimita; George Awinda; John Waitumbi; Josphat Nyataya; Rachel Githii; Rehema Liyai; Stephen Ochola |
| EPI_ISL_2779526, EPI_ISL_2779528, EPI_ISL_2779534 | MOH-Q | USAMRD-A, Basic Science Laboratory | Alan Lemtudo; Beth Mutai; Brian Andika; Carol Kifude; Clement Masakwe; Eric Muthanje; Esther Omuseni; Faith Sigei; Gathii Kimita; George Awinda; John Waitumbi; Josphat Nyataya; Rachel Githii; Rehema Liyai; Stephen Ochola |
| EPI_ISL_2346387, EPI_ISL_2346398, EPI_ISL_2346404, EPI_ISL_2346424, EPI_ISL_2346425, EPI_ISL_2346426, EPI_ISL_2346428, EPI_ISL_2346429, EPI_ISL_2346430, EPI_ISL_2346432, EPI_ISL_2346434, EPI_ISL_2346435 | | | |
| see above | MRC/UVRI & LSHTM Uganda Research Unit, Central Public Health Laboratories | MRC/UVRI & LSHTM Uganda Research Unit, Central Public Health Laboratories | Dan Lule Bugembe; Isaac Sseeewanyana; Matthew Cotten; My V.T. Phan; Patrick Semanda; Pontiano Kaleebu; Susan Nabadda |
| EPI_ISL_1970565, EPI_ISL_1970566, EPI_ISL_1970567, EPI_ISL_1970568 | MRC/UVRI & LSHTM Uganda Research Unit | MRC/UVRI & LSHTM Uganda Research Unit | Dan Lule Bugembe; Isaac Sseeewanyana; Matthew Cotten; My V.T. Phan; Patrick Semanda; Pontiano Kaleebu; Susan Nabadda |
| EPI_ISL_954236, EPI_ISL_954239, EPI_ISL_1469319, EPI_ISL_1469324, EPI_ISL_1469337, EPI_ISL_1469381, EPI_ISL_1469382, EPI_ISL_1469385 | | | |
| see above | MRC/UVRI & LSHTM Uganda Research Unit | Where sequence data have been generated and submitted to GISAID | Dan Lule Bugembe; Isaac Sseeewanyana; Matthew Cotten; My V.T. Phan; Patrick Semanda; Pontiano Kaleebu; Susan Nabadda |
| EPI_ISL_2690446, EPI_ISL_2690447, EPI_ISL_2690448, EPI_ISL_2690449, EPI_ISL_2690450, EPI_ISL_2690451, EPI_ISL_2690452, EPI_ISL_2690456, EPI_ISL_2690457, EPI_ISL_2690458, EPI_ISL_2690459, EPI_ISL_2690460, EPI_ISL_2690461, EPI_ISL_2690462, EPI_ISL_2690463, EPI_ISL_2690485, EPI_ISL_2690496, EPI_ISL_2690500, EPI_ISL_2690513 | | | |
| see above | MRC/UVRI & LSHTM Uganda Research Unit, Central Public Health Laboratories, Rakai Health Sciences Program | MRC/UVRI & LSHTM Uganda Research Unit, Central Public Health Laboratories | Dan Lule Bugembe; Hellen Nansumba; Isaac Sseeewanyana; Matthew Cotten; My V.T. Phan; Patrick Semanda; Pontiano Kaleebu; Susan Nabadda |
| EPI_ISL_2690464, EPI_ISL_2690465, EPI_ISL_2690466, EPI_ISL_2690467, EPI_ISL_2690468, EPI_ISL_2690469, EPI_ISL_2690470, EPI_ISL_2690471, EPI_ISL_2690472, EPI_ISL_2690473, EPI_ISL_2690474, EPI_ISL_2690475, EPI_ISL_2690476, EPI_ISL_2690477, EPI_ISL_2690478 | | | |
| see above | MRC/UVRI & LSHTM Uganda Research Unit, Central Public Health Laboratories, Rakai Health Sciences Program | MRC/UVRI & LSHTM Uganda Research Unit, Rakai Health Sciences Program | Charles Ssuuna; Dan Lule Bugembe; Matthew Cotten; My V.T. Phan; Pontiano Kaleebu; Ronald Moses Galiwango; Steven J Reynolds |
| EPI_ISL_428857, EPI_ISL_471158, EPI_ISL_471160, EPI_ISL_471166, EPI_ISL_561027, EPI_ISL_561085, EPI_ISL_561218, EPI_ISL_1216072, EPI_ISL_1216077, EPI_ISL_1216118, EPI_ISL_1234526, EPI_ISL_1234531, EPI_ISL_1731549, EPI_ISL_1731557, EPI_ISL_2142749, EPI_ISL_2820697, EPI_ISL_2941560, EPI_ISL_2958626, EPI_ISL_2958629, EPI_ISL_2958631, EPI_ISL_2958632, EPI_ISL_2958636, EPI_ISL_2958637, EPI_ISL_2958638, EPI_ISL_2958639, EPI_ISL_2958644, EPI_ISL_2958645, EPI_ISL_2958646, EPI_ISL_2958647 | | | |
| see above | MRCG at LSHTM Genomics lab | MRCG at LSHTM Genomics lab | Abdoulie Kanteh; Abdul Karim sesay; Bakary Sanyang; Dabiri Damilari; Damiri Damilari; Jarra Manneh; Mariama Kujabi; Sainabou laye Ndure; Sesay et al |
| EPI_ISL_903937 | MS Public Health Laboratory | Genomics and Discovery, Respiratory Viruses Branch, Division of Viral Diseases, Centers for Disease Control and Prevention | Anna Montmayeur; Anna Uehara; Ben L. Rambo-Martin; Clinton R. Paden; Dhwani Batra; Haibin Wang; Jasmine Padilla; Jing Zhang; Justin Lee; Krista Queen; Lori Rowe; Mark Burroughs; Mili Sheth; Peter W. Cook; Rachel Marine; Sarah Nobles; Suxiang Tong; Yan Li; Ying Tao |
| EPI_ISL_2224388, EPI_ISL_2920904 | MS Public Health Laboratory | University of Mississippi Medical Center, Molecular and Genomics Core Facility | Ashley C. Johnson; D. Ashley Robinson; Ithiel J. Frame; Krishna K. Ayyalasomayajula; Michael R. Garrett; Wenjie Wu |
| EPI_ISL_802148, EPI_ISL_1300812, EPI_ISL_1300813, EPI_ISL_1301126 | MSHS Clinical Microbiology Laboratories | MSHS Pathogen Surveillance Program | Adolfo García-Sastre; Adriana van de Guchte; Ajay Obla; Alberto Paniz-Mondolfi; Ana S. Gonzalez-Reiche; Andrew Kasarskis; Angela Amoako; Ashley S. Salimbangon; Ashley Salimbangon; Betsaida Salom Melo; Bremy Alburquerque; Brianne Ciferri; Charles Gleason; Daniel Floda; Deena R. Altman; Denise Jurczyszak; Elena Hirsch; Emilia Mia Sordillo; Emily Ferreri; Gintaras Deikus; Giulio Kleiner; Gopi Patel; Hala Alshammary; Harm van Bakel; Irina Oussenko; Jayeeta Dutta; Juan Soto; Julia Matthews; Katherine Beach; Kathryn Twyman; Kayla Russo; Komal Srivastava; Levy Sominsky; Mahmoud Awawda; Marta Luksza; Matthew M. Hernandez; Melissa Gitman; Michael D. Nowak; Mitchell J. Sullivan; Nancy Francoeur; Rachel Chernet; Robert Sebra; Sarah Schaefer; Shelcie Fabre; Shwetha Hara Sridhar; Viviana Simon; Ying-Chih Wang; Zenab Khan |
| EPI_ISL_2690428 | MSK Microbiology Lab | MSK Microbiology Lab | Esther Babady; Krupa Jani; Tracy McMillen |
| EPI_ISL_1722072, EPI_ISL_1722136 | MVZ Labor Dr. Fenner und Kollegen (Standort Hamburg) | Robert Koch Institute | |
| EPI_ISL_2261223 | MVZ Labor Dr. Quade & Kollegen GmbH | Robert Koch Institute | |
| EPI_ISL_2636786, EPI_ISL_3044511 | MVZ Labor Krone GbR | Robert Koch Institute | |
| EPI_ISL_2314763 | MVZ Medizinisches Labor Hannover GmbH | Robert Koch Institute | |
| EPI_ISL_1936102, EPI_ISL_1936103, EPI_ISL_1936104, EPI_ISL_1936105, EPI_ISL_1936106, EPI_ISL_1936107, EPI_ISL_1936108, EPI_ISL_1936109, EPI_ISL_1936110, EPI_ISL_1936111, EPI_ISL_1936112, EPI_ISL_1936113, EPI_ISL_1936114, EPI_ISL_1936115, EPI_ISL_1936116, EPI_ISL_1936117, EPI_ISL_1936118, EPI_ISL_1936119, EPI_ISL_1936120, EPI_ISL_1936121, EPI_ISL_1936122, EPI_ISL_1936123, EPI_ISL_1936124, EPI_ISL_1936125, EPI_ISL_1936126, EPI_ISL_1936127, EPI_ISL_1936128, EPI_ISL_1936129, EPI_ISL_1936130, EPI_ISL_1936131, EPI_ISL_1936132, EPI_ISL_1936133, EPI_ISL_1936134, EPI_ISL_1936135, EPI_ISL_1936136, EPI_ISL_1936137, EPI_ISL_1936138, EPI_ISL_1936139, EPI_ISL_1936140, EPI_ISL_1936141, EPI_ISL_1936142, EPI_ISL_1936143, EPI_ISL_1936144, EPI_ISL_1936145, EPI_ISL_1936194, EPI_ISL_1936195, EPI_ISL_1936196, EPI_ISL_1936197, EPI_ISL_1936198, EPI_ISL_1936199, EPI_ISL_1936200, EPI_ISL_1936201, EPI_ISL_1936202, EPI_ISL_1936203, EPI_ISL_1936204, EPI_ISL_1936205, EPI_ISL_1936206, EPI_ISL_1936207, EPI_ISL_1936208, EPI_ISL_1936209, EPI_ISL_1936210, EPI_ISL_1936211, EPI_ISL_1936212, EPI_ISL_1936213, EPI_ISL_1936214, EPI_ISL_1936215, EPI_ISL_1936216, EPI_ISL_1936217, EPI_ISL_1936218, EPI_ISL_1936219, EPI_ISL_1936220, EPI_ISL_1936221, EPI_ISL_1936222, EPI_ISL_1936223, EPI_ISL_1936224, EPI_ISL_1936225, EPI_ISL_1936226, EPI_ISL_1936227, EPI_ISL_1936228, EPI_ISL_1936229, EPI_ISL_1936230, EPI_ISL_1936231, EPI_ISL_1936232, EPI_ISL_1936233, EPI_ISL_1936234, EPI_ISL_1936235, EPI_ISL_1936236, EPI_ISL_1936237, EPI_ISL_1936240, EPI_ISL_1936241, EPI_ISL_1936242, EPI_ISL_1936243, EPI_ISL_1936244, EPI_ISL_1936245, EPI_ISL_1936246, EPI_ISL_1936247, EPI_ISL_1936248, EPI_ISL_1936249, EPI_ISL_1936250, EPI_ISL_1936251, EPI_ISL_1936252, EPI_ISL_1936253, EPI_ISL_1936254, EPI_ISL_1936255, EPI_ISL_1936256, EPI_ISL_1936257, EPI_ISL_1936258, EPI_ISL_1936259, EPI_ISL_1936260, EPI_ISL_1936261, EPI_ISL_1936262, EPI_ISL_1936263, EPI_ISL_1936264, EPI_ISL_1936265, EPI_ISL_1936266, EPI_ISL_1936267, EPI_ISL_1936268, EPI_ISL_1936269, EPI_ISL_1936270, EPI_ISL_1936271, EPI_ISL_1936272, EPI_ISL_1936273, EPI_ISL_1936274, EPI_ISL_1936275, EPI_ISL_1936276, EPI_ISL_1936277, EPI_ISL_1936278, EPI_ISL_1936279, EPI_ISL_1936280, EPI_ISL_1936281, EPI_ISL_1936282, EPI_ISL_1936283, EPI_ISL_1936284, EPI_ISL_1936285, EPI_ISL_1936286, EPI_ISL_1936287, EPI_ISL_1936288, EPI_ISL_1936289, EPI_ISL_1936290, EPI_ISL_1936291, EPI_ISL_1936292, EPI_ISL_1936293, EPI_ISL_1936294, EPI_ISL_1936296, EPI_ISL_1936297, EPI_ISL_1936298, EPI_ISL_1936299, EPI_ISL_1936300, EPI_ISL_1936301, EPI_ISL_1936302, EPI_ISL_1936303, EPI_ISL_1936304, EPI_ISL_1936305, EPI_ISL_1936306, EPI_ISL_1936307, EPI_ISL_1936308, EPI_ISL_1936309, EPI_ISL_1936310, EPI_ISL_1936311, EPI_ISL_1936312, EPI_ISL_1936313, EPI_ISL_1936314, EPI_ISL_1936315, EPI_ISL_1936316, EPI_ISL_1936317, EPI_ISL_1936318, EPI_ISL_1936319, EPI_ISL_1936320, EPI_ISL_1936321, EPI_ISL_1936322, EPI_ISL_1936323, EPI_ISL_1936324, EPI_ISL_1936325, EPI_ISL_1936326, EPI_ISL_1936327, EPI_ISL_1936328, EPI_ISL_1936329, EPI_ISL_1936330, EPI_ISL_1936331, EPI_ISL_1936332, EPI_ISL_1936333, EPI_ISL_1936334, EPI_ISL_1936335, EPI_ISL_1936336, EPI_ISL_1936337, EPI_ISL_1936338, EPI_ISL_1936339, EPI_ISL_1936340, EPI_ISL_1936341, EPI_ISL_1936342, EPI_ISL_1936343, EPI_ISL_1936344, EPI_ISL_1936345, EPI_ISL_1936346, EPI_ISL_1936347, EPI_ISL_1936348, EPI_ISL_1936349, EPI_ISL_1936350, EPI_ISL_1936351, EPI_ISL_1936352, EPI_ISL_1936353, EPI_ISL_1936354, EPI_ISL_1936355, EPI_ISL_1936356, EPI_ISL_1936357, EPI_ISL_1936358, EPI_ISL_1936359, EPI_ISL_1936360, EPI_ISL_1936361, EPI_ISL_1936362, EPI_ISL_1936363, EPI_ISL_1936364, EPI_ISL_1936365, EPI_ISL_1936366, EPI_ISL_1936367, EPI_ISL_1936368, EPI_ISL_1936369, EPI_ISL_1936370, EPI_ISL_1936371, EPI_ISL_1936372, EPI_ISL_1936373, EPI_ISL_1936374, EPI_ISL_1969078, EPI_ISL_1969079, EPI_ISL_1969080, EPI_ISL_1969081 | | | |
| see above | Main Chemical Laboratories Egypt Army | Main Chemical Laboratories Egypt Army | Abdullah Salama; AbedElrahman Zekri; Ahmed Gad; Bassem Elharty; Laila Elsawy; Mervat Hassan; Mohamed Abdel-Monem; Mohamed El-Esawi; Mohamed Seadawy; Mohamed Shamel; Mostfa Elhoseiny; Mostfa Yakout; Ola Elroby; Sabah Ahmed; Sherine Helmy |
| EPI_ISL_2647352, EPI_ISL_2875445 | Mako Medical | Centers for Disease Control and Prevention Division of Viral Diseases, Pathogen Discovery | Adrian Paskey; Benjamin Rambo-Martin; Christopher Gulvick; Clinton R. Paden; Dakota Howard; Darlene Wagner; Dhwani Batra; Duncan MacCannell; Jason Caravas; Kara Moser; Lauren Moon; Matthew Schmerer; Matthew Tugwell; Peter W. Cook; Scott Sammons; Shatavia Morrison; Yvette Unoarumhi |
| EPI_ISL_582124 | Malaysia Genome Institute | Malaysia Genome Institute | Azrin Ahmad; Enizza Kasim; Irni Suhayu Sapian; Mohd Faizal Abu Bakar; Mohd Noor Mat Isa; Nor Azfa Johari; Nurhezreen Md Iqbal; Shahrul Hisham Zainal Ariffin; Shamsidar Sopie; Siti Noraini Othman; Yusuf Muhammad Noor |
| EPI_ISL_1827700 | Manzana | National Institute for Communicable Diseases of the National Health Laboratory Service | Amoako DG; Bhiman JN; Ismail A; Mahlangu B; Maphalala GP; Mohale T; Ntuli N; Scheepers C |
| EPI_ISL_2373638 | Mapmygenome | CSIR-Centre for Cellular and Molecular Biology - INSACOG | Amareshwar Vodapalli; Ara Sreenivas; B Himasri; Divya Tej Sowpati; Karthik Bharadwaj Tallapaka; Lamuk Zaveri; Onkar Kulkarni; Rakesh K Mishra; Sharath Chandra Thota; Shreekant Verma; Sofia Banu; Viswagithe S L |
| EPI_ISL_1910606, EPI_ISL_1910607, EPI_ISL_1910617, EPI_ISL_1913025 | Mashrek Medical Diagnostic Center | Microbial Pathogenomics Lab - LAU | Georgi Merhi; Jad Koweyes; Sima Tokajian; Tamara Salloum |
| EPI_ISL_692915 | Massachusetts State Public Health Laboratory | Massachusetts State Public Health Laboratory | Andrew Lang; Glen Gallagher; Sandra Smole; Timelia Fink |

| | | | |
|---|---|---|---|
| EPI_ISL_2547344 | Matjieskloof Clinic | National Health Laboratory Service/University of Cape Town (NHLS/UCT) | Arash Iranzadeh; Bruna Galvao; Carolyn Williamson; Deelan Doolabh; Diana Hardie; Gert Marais; Innocent Mudau; Lynn Tyers; Marvin Hsiao; Stephen Korsman |
| EPI_ISL_1196004 | Mbabane PHU | National Institute for Communicable Diseases of the National Health Laboratory Service | Amoako DG; Bhiman JN; Ismail A; Mahlangu B; Maphalala GP; Mohale T; Ntuli N; Scheepers C |
| EPI_ISL_1722079, EPI_ISL_2110393, EPI_ISL_2260594 | Med. Labor Prof. Schenk Dr. Ansorge & Kollegen | Robert Koch Institute | |
| EPI_ISL_456378 | MedLab Central Ltd | Institute of Environmental Science and Research (ESR) | Anja Werno; Antje van der Linden; Arlo Upton; Chris Mansell; David Hammer; Dragana Drinkovic; Erasmus Smit; Gary McAuliffe; Hana Sofia Andersson; James Ussher; Jill Sherwood; Joep de Ligt; Josh Freeman; Julia Howard; Juliet Elvy; Lauren Jelly; Mary DeAlmeida; Matt Blakiston; Matt Storey; Matthew Rogers; Max Bloomfield; Michael Addidle; Michelle Balm; Sally Roberts; Sarah Jefferies; Sharmini Muttaiyah; Susan Morpeth; Susan Taylor; Timothy Blackmore; Vani Sathyendran; Veronica Playle; Virginia Hope; Xiaoyun Ren |
| EPI_ISL_475745, EPI_ISL_475746, EPI_ISL_475747, EPI_ISL_475748, EPI_ISL_475749, EPI_ISL_475750, EPI_ISL_475751, EPI_ISL_475752, EPI_ISL_475753, EPI_ISL_482759, EPI_ISL_482760, EPI_ISL_482761, EPI_ISL_482762, EPI_ISL_482763, EPI_ISL_482764, EPI_ISL_482765, EPI_ISL_482766, EPI_ISL_482767, EPI_ISL_482768, EPI_ISL_482769, EPI_ISL_482770, EPI_ISL_482771, EPI_ISL_482772, EPI_ISL_482773, EPI_ISL_482774, EPI_ISL_482775, EPI_ISL_483035, EPI_ISL_483036, EPI_ISL_483037, EPI_ISL_483038, EPI_ISL_1098839, EPI_ISL_1109484, EPI_ISL_1109485, EPI_ISL_1109486, EPI_ISL_1109624, EPI_ISL_1109625, EPI_ISL_1109626, EPI_ISL_1109627, EPI_ISL_1109628, EPI_ISL_1109629, EPI_ISL_1109630, EPI_ISL_1141525, EPI_ISL_1165078, EPI_ISL_1165079, EPI_ISL_1165080, EPI_ISL_1165082, EPI_ISL_1165083, EPI_ISL_1165087, EPI_ISL_1167189, EPI_ISL_1167193, EPI_ISL_1586895 | | | |
| see above | Medical Ain Shams Research Institute (MASRI), Ain Shams University | Medical Ain Shams Research Institute (MASRI), Ain Shams University | Ahmad Moustafa; Ashraf Omar; Aya Mohamed; Fatma Ebied; Ghada Ismael; Hagar Elshora; Hala Hafez; Hesham Elghazaly; Hoda Abd Elsatar; Hoda Ezz Elarab; Iman Foda; Mahmoud Elmeitini; Mahmoud Elmeteini; Manal Hamdy Elsaid; Mohamed Elhadidi; Osama Mansour; Osama Mansour.; Reham Kassab; Reham Mamdouh; Samia Abdo; Samia Abdou Girgis; Sara Elnakeep; Sara Hassan Agwa; Shaimaa Moustafa; Shimaa Moustafa |
| EPI_ISL_873164, EPI_ISL_2107508, EPI_ISL_2107509, EPI_ISL_2107510, EPI_ISL_2107511, EPI_ISL_2107512, EPI_ISL_2107524 | | | |
| see above | Medical Laboratory Sciences, Arab American University | Medical Laboratory Sciences, Arab American University | Al-Jawabreh, A.; Al-Jawabreh, H.; Dumaidi, K.; Dumaidi, k.; Ereqat, S.; Nasereddin, A. |
| EPI_ISL_882646 | Medical Research Center, Faculty of Medicine, Syarif Hidayatullah State Islamic University Jakarta | Medical Research Center, Faculty of Medicine, Syarif Hidayatullah State Islamic University Jakarta | Chris Adhiyanto; Endah Wulandari; Fika Ekayanti; Flori Ratna Sari; Hari Hendarto; Laifa Hendarmin; Zeti Harriyati |
| EPI_ISL_581457 | Medizinische Klinik Innere Medizin I, Universitätsklinikum Tübingen | NGS Competence Center Tübingen, Institut für Medizinische Mikrobiologie und Hygiene, Universitätsklinikum Tübingen | Angel Angelov |
| EPI_ISL_1726791 | Medizinisches Labor Wahl LÃ¼denscheid | Robert Koch Institute | |
| EPI_ISL_1722630 | Medizinisches Zentrallabor Altenburg GmbH & Co.KG | Robert Koch Institute | |
| EPI_ISL_1158449 | Michigan Department of Health and Human Services, Bureau of Laboratories | Michigan Department of Health and Human Services, Bureau of Laboratories | Blankenship HM; Riner D; Soehnlen MK |
| EPI_ISL_426477 | Microbial Genomics Laboratory, Institut Pasteur Montevideo | Microbial Genomics Laboratory, Institut Pasteur Montevideo, Uruguay | Cecilia Salazar; Florencia Díaz-Viraqué; Gonzalo Moratorio; Gregorio Iraola; Marianoel Pereira; Pilar Moreno |
| EPI_ISL_480346 | Microbial Genomics Laboratory, Institut Pasteur de Montevideo | Microbial Genomics Laboratory, Institut Pasteur de Montevideo | Cecilia Salazar; Gonzalo Moratorio; Gregorio Iraola; Ignacio Ferrés; Marianoel Pereira; Pilar Moreno |
| EPI_ISL_519427 | Microbiological Diagnostic Unit - Public Health Laboratory (MDU-PHL) | MDU-PHL | Sait, M.; Schultz M.; Seemann T.; Sherry, N. |
| EPI_ISL_427149, EPI_ISL_430668 | Microbiological Diagnostic Unit Public Health Laboratory | Microbiological Diagnostic Unit Public Health Laboratory | Sait, M.; Schultz M.; Seemann T.; Sherry, N. |
| EPI_ISL_513925 | Microbiology & Bioinformatics and Biostatistics, Kohat University of Science and Technology (Pakistan) & Shanghai Jiao Tong University (China) | Microbiology & Bioinformatics and Biostatistics, Kohat University of Science and Technology (Pakistan) & Shanghai Jiao Tong University (China) | A.S.; Ali, S.; D.Q.; Khan; M.T.; Muhammad; N. and Wei; T.A. |
| EPI_ISL_2136858 | Microbiology Department, Laboratori Clínic Metropolitana Nord. Hospital Universitari Germans Trias i Pujol | Can Ruti SARS-CoV-2 Sequencing Hub (HUGTiP/IrsiCaixa/IGTP) | Alba Sánchez; Anna Not; Antoni E Bordoy; Bonaventura Clotet; Cristina Casañ; Cristina Esteban; Francesc Catala-Moll; Gemma Clara; Ignacio Blanco; Marc Noguera-Julian; Maria Casadellà; Mariona Parera; Mercedes Guerrero; Montserrat Giménez; Pere-Joan Cardona; Pilar Armengol; Roger Paredes; Verónica Saludes; and Elisa Martró on behalf of the Can Ruti SARS-CoV-2 Sequencing Hub. |
| EPI_ISL_2931852 | Microbiology Department. Complexo Hospitalario Universitario de Vigo | Microbiology Department. Complexo Hospitalario Universitario de Vigo | Alfaya N; Alonso I; Alvarez M; Cabrera JJ; Carballo R; Cores O; Cortizo S; Davina C; Martinez L; Mediero G; Perez S; Potel C; Regueiro B; Rey S; Vasallo FJ; del-Campo V |
| EPI_ISL_1303252 | Microbiology Division, South Carolina Department of Health and Environmental Control | Microbiology Division, South Carolina Department of Health and Environmental Control | Flores, H.; Freeman, J. |
| EPI_ISL_2688489 | Microbiology Laboratory, Attikon University Hospital, Athens | Central National Laboratory ,Public Health Organization | N.Siafakas; S.Pournaras et al |
| EPI_ISL_1416330 | Microbiology and Virology Laboratory, "Policlinico Riuniti, Azienda Ospedaliero Universitaria, Foggia" | Microbiology and Virology Laboratory, "Policlinico Riuniti, Azienda Ospedaliero Universitaria, Foggia" | Annamaria D'Aprile; Daniela Pisanelli; Donatella Cedola; Fabio Arena; Loris Micelli; Maria Pia Patrizio; Maria Rosaria Lipsi; Maurizio Margaglione and Rosella De Nittis.; Settimia Altamura; Tiziana Rollo; Valeria Delli Carri |
| EPI_ISL_1064063 | Microbiology and Virology Unit, Azienda Ospedale Padova, Padova, Italy | Department of Molecular Medicine, Computational Medicine Group, Univeresity of Padova, Padova, Italy | Andrea Crisanti; Andrea Spitaleri; Claudia Del Vecchio; Daniela Maria Cirillo; Dejan Lazarevic; Elisa Franchin; Enrico Lavezzo; Fabio Simeoni; Federico Bianca; Francesca Saluzzo; Francesco Onelia; Giovanni Lorenzin; Giovanni Tonon; Laura Manuto; Marco Grazioli; Stefano Toppo |
| EPI_ISL_3136161 | Microbiology laboratory, National Center for Global Health and Medicine Hospital | Pathogen Genomics Center, National Institute of Infectious Diseases | Harutaka Katano; Kentaro Itokawa; Makoto Kuroda; Masanori Hashino; Norio Ohmagari; Rina Tanaka; Shinji Saito; Shun Iida; Tadaki Suzuki; Tsuyoshi Sekizuka; Yuichiro Hirata |
| EPI_ISL_2348487 | Middle East Institute of Health University Hospital | Microbial Pathogenomics Lab - LAU | Edmond Abboud; Georgi Merhi; Jad Koweyes; Sima Tokajian |
| EPI_ISL_548032, EPI_ISL_548103, EPI_ISL_548145, EPI_ISL_579102, EPI_ISL_794624, EPI_ISL_1016845, EPI_ISL_1082258, EPI_ISL_1469089, EPI_ISL_1621293, EPI_ISL_1621326, EPI_ISL_1967893, EPI_ISL_1967898, EPI_ISL_1967900, EPI_ISL_2406492, EPI_ISL_2650019, EPI_ISL_2650024, EPI_ISL_2811945, EPI_ISL_2811946, EPI_ISL_2811954, EPI_ISL_2811956, EPI_ISL_2964937 | | | |
| see above | Middlemore Hospital | Institute of Environmental Science and Research (ESR) | Anja Werno; Antje van der Linden; Arlo Upton; Chris Mansell; David Hammer; Dragana Drinkovic; Erasmus Smit; Gary McAuliffe; Hana Sofia Andersson; Hermes Perez; James Ussher; Jill Sherwood; Jing Wang; Joep de Ligt; Josh Freeman; Julia Howard; Juliet Elvy; Lauren Jelly; Mary DeAlmeida; Matt Blakiston; Matt Storey; Matthew Rogers; Max Bloomfield; Michael Addidle; Michelle Balm; Muhammad Faisal; Nikki Freed; Olin Silander; Olivia Stroeven; Paula scholes; Rachel Boyle; Sally Roberts; SallyAnn Harbison; Sarah Jefferies; Sharmini Muttaiyah; Susan Lin; Susan Morpeth; Susan Taylor; Timothy Blackmore; Vani Sathyendran; Veronica Playle; Virginia Hope; Xiaoyun Ren |
| EPI_ISL_718146, EPI_ISL_2367475, EPI_ISL_2535605, EPI_ISL_2535657, EPI_ISL_2535659 | Ministry of Health Hospitals | Institute of Health and Community Medicine | Chan Chia Jui; Chua Hock Hin; David Perera; Ooi Mong How; Tonnii Sia Loong Loong; Wong Jyn Shan; Wong Kiing Aik |
| EPI_ISL_428713, EPI_ISL_814079, EPI_ISL_1020467, EPI_ISL_1117613, EPI_ISL_2158034, EPI_ISL_2158057, EPI_ISL_2158066, EPI_ISL_2158074, EPI_ISL_2232628, EPI_ISL_2232646, EPI_ISL_2232672, EPI_ISL_2232673, EPI_ISL_2232674, EPI_ISL_2232675, EPI_ISL_2232676, EPI_ISL_2232695, EPI_ISL_2363590, EPI_ISL_2363596, EPI_ISL_2363632, EPI_ISL_2363736, EPI_ISL_2403280, EPI_ISL_2958500, EPI_ISL_2984650, EPI_ISL_2984684, EPI_ISL_2984763, EPI_ISL_2987762, EPI_ISL_2987829, EPI_ISL_2987841, EPI_ISL_2987847, EPI_ISL_2987930, EPI_ISL_2987933, EPI_ISL_2988072, EPI_ISL_3032772, EPI_ISL_3032778, EPI_ISL_3063381, EPI_ISL_3063382, EPI_ISL_3066742, EPI_ISL_3066746 | | | |
| see above | Ministry of Health Turkey | Ministry of Health Turkey | Ayşe Başak Altaş; Fatma Bayrakdar; Gulay Korukluoglu; Gülay Korukluoğlu; Selçuk Kılıç; Suleyman Yalcin; Süleyman Yalcin; Yasemin Cosgun; Yasemin Coşgun |
| EPI_ISL_2846023 | Ministry of Health, Jaber Al-Ahmad Hospital | Virology Unit, Department of Microbiology, Faculty of Medicine, Kuwait University | Anfal Al-Adwani; Ebaa Al-Awadhi; Hussain Safar; Nada Madi |
| EPI_ISL_1712690, EPI_ISL_1712705, EPI_ISL_1713194, EPI_ISL_1713231, EPI_ISL_1713276, EPI_ISL_1713546, EPI_ISL_1713637, EPI_ISL_2274279, EPI_ISL_2274319, EPI_ISL_2274351, EPI_ISL_2274354, EPI_ISL_2274358, EPI_ISL_2274390, EPI_ISL_2408295, EPI_ISL_2408310, EPI_ISL_2408353, EPI_ISL_2408365, EPI_ISL_2842881, EPI_ISL_2842914, EPI_ISL_2843042, EPI_ISL_2843050, EPI_ISL_2843081, EPI_ISL_2843089, EPI_ISL_2843116, EPI_ISL_2843135, EPI_ISL_2843139, EPI_ISL_2843146, EPI_ISL_2843149, EPI_ISL_2843237, EPI_ISL_2843241, EPI_ISL_2843246, EPI_ISL_2843259, EPI_ISL_2843273, EPI_ISL_2843280, EPI_ISL_2843284, EPI_ISL_2843285 | | | |
| see above | Ministry of Public Health / Hamad Medical Corporation | Biomedical Research Center (BRC), Qatar University / Qatar Genome Project (QGP) | Asmaa A. Al-Thani. MOPH and HMC: Abdullatif Al-Khal; Asmaa A. Al-Thani. MOPH and HMC: Abdullatif Al-Khal; BRC: Fatiha M. Benslimane; Chadi Saad; Dana Al-Batesh; Dina Elgakhlab QGP: Fatima H. Al-Kuwari; Dina Elgakhlab QGP: Fatima H. Al-Kuwari; Einas A. E. Al-Kuwari; Hadi M. Yassine; Hamad E. Al-Romaihi; Hamda Alromaihi; Heba A. Al-Khatib; Mashael A. Al-Bader; Mohammed Al-Thani; Muna A. S. Al-Maslamani; Oal Al-Jamal; Peter V. Coyle; Reham A. El-Kahlout. QBB: Tasneem Al-Hamad; Reham A. El-Kahlout. QBB: Tasneem Al-Hamad; Roberto Bertollini; Salih Al-Marri |
| EPI_ISL_1714165, EPI_ISL_1714185, EPI_ISL_1714328, EPI_ISL_1714331, EPI_ISL_1714468, EPI_ISL_1714531, EPI_ISL_1714557 | | | |
| see above | Ministry of Public Health / Hamad Medical Corporation | Weill Cornell Medical College - Qatar (WCM-Q), Genomics Core Laboratory / Qatar Genome Project (QGP) | Chadi Saad MOPH and HMC: Abdullatif Al-Khal; Dina Elgakhlab; Einas A. E. Al-Kuwari; Hamad E. Al-Romaihi; Hamda Alromaihi; Joel A Malek. QGP: Fatima H. Al-Kuwari; Laith Abu-Raddad; Mashael A. Al-Bader; Meryem Bensaad; Mohammed Al-Thani; Muna A. S. Al-Maslamani; Peter V. Coyle; Reham A. El-Kahlout. QBB: Tasneem Al-Hamad; Roberto Bertollini; Salih Al-Marri; Shameem Younuskunju; WCMQ: Ayeda A. Ahmed; Yasmin Mohamoud |
| EPI_ISL_708485, EPI_ISL_1760680, EPI_ISL_2015217, EPI_ISL_2443802, EPI_ISL_2536893 | Minnesota Department of Health, Public Health Laboratory | Minnesota Department of Health, Public Health Laboratory | Alexandra Lorentz; Jacob Garfin; Matt Plumb; and Xiong Wang |
| EPI_ISL_1362768, EPI_ISL_1392816, EPI_ISL_2035597 | Missouri State Public Health Laboratory | Missouri State Public Health Laboratory | Ashley New; Joshua Barry; Matthew Sinn |
| EPI_ISL_2140698 | Mitchells Plain Hospital wc MPH | NHLS/UCT | Arash Iranzadeh; Bruna Galvao; Carolyn Williamson; Deelan Doolabh; Diana Hardie; Innocent Mudau; Kruger Marais; Lynn Tyers; Marvin Hsiao; Stephen Korsman |
| EPI_ISL_2861095, EPI_ISL_2928345 | Mobile County Health Department | Synergy Laboratories | Megan Cornwell |
| EPI_ISL_435127, EPI_ISL_463740, EPI_ISL_520723 | Mohammed Bin Rashid University of Medicine and Health Sciences | Al Jalila Genomics Center | Abdulmajeed Alkhaja; Abiola Catherine Senok; Ahmad Abou Tayoun; Alawi Alsheikh-Ali; Divinlal Harilal; Hamda Khansaheb; Hanan Al Suwaidi; Mohammed Uddin; Norbert Nowotny; Qutayba Hamid; Rabih Halwani; Rifat Hamoudi; Rupa Murthy Varghese; Sathishkumar Ramaswamy; Tom Loney; Zulfa Omar Deesi |
| EPI_ISL_2101058, EPI_ISL_2101204 | Molecular Biology Laboratory, Faculty Medicine and Health Sciences, Warmadewa University | Eijkman Institute for Molecular Biology, National Agency for Research and Innovation; Molecular Biology Laboratory, Faculty Medicine and Health Sciences, Warmadewa University | Amin Soebandrio; Edison Johar; Erly Sintya; Frilasita A Yudhaputri; Hidayat Trimarsanto; Iskandar Adnan; Khin Saw Myint; Lidwina Priliani; Lydia V. Panggalo; Muhammad Rezki Rasyak; Safarina G Malik; Sri Masyeni; Sukma Oktavianthi; Willy Agustine |
| EPI_ISL_2095819 | Molecular Diagnostics Mater Dei Hospital Pathology Department Malta | Molecular Diagnostics Mater Dei Hospital Pathology Department Malta | C Cilia; G Zahra; L Grech; R Borg |

| | | | |
|---|---|---|---|
| EPI_ISL_2006673, EPI_ISL_2101345, EPI_ISL_2101350, EPI_ISL_2101358, EPI_ISL_2102068, EPI_ISL_2102070, EPI_ISL_2105875, EPI_ISL_2105881, EPI_ISL_2105939, EPI_ISL_2113984, EPI_ISL_2114905, EPI_ISL_2399438, EPI_ISL_2399457, EPI_ISL_2399611, EPI_ISL_2716425, EPI_ISL_2716474, EPI_ISL_2718538, EPI_ISL_2854661, EPI_ISL_2970370, EPI_ISL_2970563, EPI_ISL_2970965, EPI_ISL_2973300, EPI_ISL_2988654, EPI_ISL_2988658 | | | |
| see above | Molecular Diagnostics Pathology Department Mater Dei Hospital Malta | Molecular Diagnostics Pathology Department Mater Dei Hospital Malta | C Cilia; G Zahra; L Grech; M Briffa; R Borg |
| EPI_ISL_681690, EPI_ISL_681700 | Molecular Medicine Laboratory, University of Magallanes | Centro Asistencial Docente y de Investigacion, Universidad de Magallanes | Diego Alvarez; Hermy Alvarez; Jacqueline Aldridge; Jorge González; Marcelo Navarrete; Marco Montes de Oca; Roberto Uribe-Paredes |
| EPI_ISL_895793, EPI_ISL_895834 | Molecular biology division, Institute of Clinical Biochemistry and Diagnostics, Charles University, Faculty of Medicine in Hradec Králové and University Hospital Hradec Králové | Molecular biology division, Institute of Clinical Biochemistry and Diagnostics, Charles University, Faculty of Medicine in Hradec Králové and University Hospital Hradec Králové | Helena Kovaříková; Ivana Baranová; Jitka Novotná; Kateřina Hrochová; Kateřina Pehlíková; Petr Brož; Tereza Baťková; Vladimír Palička. Cooperation project with BioVendor-R&D and bioinformatics company BIOXSYS s r.o. |
| EPI_ISL_2031957 | Molecular diagnostic laboratory of Federal Budget Institution of Science "Central Research Institute of Epidemiology" of The Federal Service on Customers' Rights Protection and Human Well-being Surveillance | Group of Genomics and Postgenomic Technologies of Central Research Institute of Epidemilology | Akimkin V.G.; Kaptelova V.V.; Kondrasheva L.Y.; Korneenko E.V.; Nadtoka M.I.; Saenko S.S.; Samojlov A.E.; Shipulina O.Y.; Sinicyn S.O.; Smirnova Y.S.; Speranskaya A.S.; Tivanova E.V. |
| EPI_ISL_2626244 | Molecular diagnostic laboratory of Federal Budget Institution of Science "Central Research Institute of Epidemiology" of The Federal Service on Customers' Rights Protection and Human Well-being Surveillance | Group of Genomics and Postgenomic Technologies of Central Research Institute of Epidemilology | Akimkin V.G.; Cherkashina A.S.; Kaptelova V.V.; Kondrasheva L.Y.; Korneenko E.V.; Nadtoka M.I.; Saenko S.S.; Samoilov A.E.; Sinitsyn S.O.; Smirnova Y.S.; Speranskaya A.S.; Tivanova E.V. |
| EPI_ISL_1365029, EPI_ISL_1365031, EPI_ISL_1367691, EPI_ISL_1663669, EPI_ISL_1663671 | Molecular diagnostic unit for viral haemorrhagic fevers and emerging viruses, Bouaké CHU Laboratory | Molecular diagnostic unit for viral haemorrhagic fevers and emerging viruses, Bouaké CHU Laboratory | Adjaratou Traoré; Bamba Fatoumata Touré; Chantal Akoua-Koffi; Coulibaly Mbegnan; Diané Bamourou; Essia Belarbi; Etilé Anoh; Fabian Leendertz; Grit Schubert; Kra Ouffoué; Monemo Pacome; Oby Wayoro; Safiatou Karidioula; Soundélé Maïté |
| EPI_ISL_614348, EPI_ISL_614364, EPI_ISL_614382, EPI_ISL_614391, EPI_ISL_681834, EPI_ISL_682058, EPI_ISL_1662579, EPI_ISL_1662582, EPI_ISL_1662584 | | | |
| see above | Molecular diagnostic unit for viral haemorrhagic fevers and emerging viruses, Bouaké CHU Laboratory | Project group Epidemology of Highly Pathogenic Microorganisms, Robert Koch-Institute | Adjaratou Traoré; Bamba Fatoumata Touré; Chantal Akoua-Koffi; Coulibaly Mbegnan; Diané Bamourou; Essia Belarbi; Etilé Anoh; Fabian Leendertz; Grit Schubert; Kra Ouffoué; Monemo Pacome; Safiatou Karidioula; Soundélé Maïté |
| EPI_ISL_1881945 | Molekylær Medicinsk Afdeling, Aarhus University Hospital, Aarhus, Denmark | Aalborg University | Danish Covid-19 Genome Consortium |
| EPI_ISL_2350967, EPI_ISL_2351003 | Mongkutwattana General Hospital | Division of Genomic Medicine and Innovation support,Department of Medical Sciences, Ministry of Public Health, Thailand | Archawin Rojanawiwat; Jirapha Pakdee; Natthakul Bunneang; Nuanjun Wichukchinda; Penpitcha Thawong; Pilailuk Akkapaiboon Okada; Pundharika Piboonsiri; Surakameth Mahasirimongkol; Waritta Sawaengdee |
| EPI_ISL_889366 | Motol University Hospital | Institute of Applied Biotechnologies a.s. | Kateřina Kvapilová; Martin Kašný; Ondřej Brzoň; Pavel Dřevínek; Petr Klempt; Petr Kvapil |
| EPI_ISL_3031397 | Mount Kenya Hospital | USAMRD-A, Basic Science Laboratory | Alan Lemtudo; Beth Mutai; Brian Andika; Carol Kifude; Clement Masakwe; Eric Muthanje; Esther Omuseni; Faith Sigei; Gathii Kimita; George Awinda; John Waitumbi; Josphat Nyataya; Rachel Githii; Rehema Liyai; Stephen Ochola |
| EPI_ISL_1534267 | Mowbray Maternity Hospital wc MMH | NHLS/UCT | Arash Iranzadeh; Bruna Galvao; Carolyn Williamson; Deelan Doolabh; Diana Hardie; Innocent Mudau; Kruger Marais; Lynn Tyers; Marvin Hsiao; Stephen Korsman |
| EPI_ISL_2429125, EPI_ISL_2429128 | Mubarak Al-Kabeer Hospital | Virology Unit, Department of Microbiology, Faculty of Medicine, Kuwait University | Anfal Al-Adwani; Hussain Safar; Nada Madi |
| EPI_ISL_2899875, EPI_ISL_2964432, EPI_ISL_2964451 | N.F. Gamaleya Research Center for Epidemiology and Microbiology | WHO National Influenza Centre Russian Federation | Alexander Gintsburg; Alexey Shchetinin; Alina Odintsova; Andrei Botikov; Andrei Pochtovyi; Andrei Siniavin; Andrey Komissarov; Anna Kovyrshina; Artem Fadeev; Artem Tkachuk; Daria Danilenko; Denis Kleymenov; Denis Logunov; Dmitry Lioznov; Dmitry Shcheblyakov; Elena Mazunina; Elena Nabieva; Elena Shidlovskaya; Elizaveta Divisenko; Evgeniia Bykonia; Georgii Bazykin; Inna Dolzhikova; Kirill Varchenko; Ksenia Safina; Kseniya Komissarova; Liubov Popova; Ludmila Vasilchenko; Maria Nikiforova; Maria Pisareva; Mikhail Bakaev; Nadezhda Kuznetsova; Nikita Yolshin; Oula Mansour; Tamila Musaeva; Veronika Eder; Vladimir Gushchin |
| EPI_ISL_2382733 | NAMRU-6 | Genomics and Discovery, Respiratory Viruses Branch, Division of Viral Diseases, Centers for Disease Control and Prevention | Adam Retchless; Anna Kelleher; Anna Uehara; Brian Lynch; Clinton R. Paden; Dhwani Batra; Haibin Wang; Han Jia Justin Ng; Jasmine Padilla; Jing Zhang; Justin Lee; Krista Queen; Mark Burroughs; Mili Sheth; Morgan Davis; Peter Cook; Rachel Marine; Sarah Nobles; Suxiang Tong; Tara Coalter; Yan Li; Ying Tao |
| EPI_ISL_2550520 | NB-Hôpital Georges L. Dumont | National Microbiology Laboratory (NML) | Anna Majer; Anneliese Landgraff; CanCOGeN's metadata curation team; Darian Hole; Elsie Grudeski; Gary Van Domselaar; Grace Seo; Guillaume Desnoyers; Jennifer Tanner; Kirsten Biggar; Madison Chapel; Morag Graham; Natalie Knox; Nathalie Bastien; Philip Mabon; Public Health Agency of Canada CanCOGeN team; Rhiannon Huzarewich; Richard Garceau; Russell Mandes; Shari Tyson; Timothy Booth; Yan Li |
| EPI_ISL_2710271, EPI_ISL_2726856 | NHLS Charlotte Maxeke Johannesburg Academic Hospital and the University of the Witwatersrand | KRISP, KZN Research Innovation and Sequencing Platform | Bulelani Manene; Florette Treurnicht; Giandhari Jennifer; Kathleen Subramoney; Naidoo Yeshnee; Pillay Sureshnee; San James; Tegally Houriiyah; Tshabuila Derek; Wilkinson Eduan; Yajna Ramphal; de Oliveira Tulio |
| EPI_ISL_912481, EPI_ISL_1818856, EPI_ISL_1818891, EPI_ISL_2162307, EPI_ISL_2162308, EPI_ISL_2162309, EPI_ISL_2162311, EPI_ISL_2162320, EPI_ISL_2162335, EPI_ISL_2162339, EPI_ISL_2162340, EPI_ISL_2162341, EPI_ISL_2162346, EPI_ISL_2162347, EPI_ISL_2162351, EPI_ISL_2162354, EPI_ISL_2162357, EPI_ISL_2162358, EPI_ISL_2162359, EPI_ISL_2162360, EPI_ISL_2162361, EPI_ISL_2162362, EPI_ISL_2162365, EPI_ISL_2162366, EPI_ISL_2162371, EPI_ISL_2162378 | | | |
| see above | NHLS Universitas Academic | UFS Virology | D Goedhals; Emmanuel Ogunbayo; MM Nyaga; MT Mogotsi; P Nthiga; PA Bester; T de Oliveira |
| EPI_ISL_467513, EPI_ISL_518043, EPI_ISL_529765, EPI_ISL_660605, EPI_ISL_660660, EPI_ISL_736971 | NHLS-IALCH | KRISP, KZN Research Innovation and Sequencing Platform | Chimukangara B; ChimukangaraB; Giandhari J; Khan S; Lessells R; Mdlalose K; Pillay S; Tegally H; Wilkinson E; York D; de Oliveira T |
| EPI_ISL_860607 | NHLS-IALCH | KRISP, KZn Research Innovation and Sequencing Platform | Giandhari J; Khan S; Lessells R; Mdlalose K; Pillay S; Tegally H; Wilkinson E; York D; de Oliveira T |
| EPI_ISL_2693039, EPI_ISL_2693050, EPI_ISL_2693075, EPI_ISL_2693084, EPI_ISL_2693093 | NHLS_VIRO | KRISP, KZN Research Innovation and Sequencing Platform | Giandhari Jennifer; Naidoo Yeshnee; Pillay Sureshnee; San James; Tegally Houriiyah; Tshabuila Derek; Wilkinson Eduan; Yajna Ramphal; de Oliveira Tulio |
| EPI_ISL_2494387, EPI_ISL_2494397, EPI_ISL_2494427, EPI_ISL_2494465, EPI_ISL_2494505, EPI_ISL_2494506, EPI_ISL_2494511, EPI_ISL_2494541, EPI_ISL_2494569, EPI_ISL_2494618, EPI_ISL_2494619, EPI_ISL_2494625, EPI_ISL_2727380, EPI_ISL_2727391, EPI_ISL_2727393, EPI_ISL_2727394 | | | |
| see above | NHLS_VIRO | KRISP, KZn Research Innovation and Sequencing Platform | Baillie Vicky; Giandhari Jennifer; Madhi Shabir; Naidoo Yeshnee; Pillay Sureshnee; San James; Tegally Houriiyah; Tshabuila Derek; Wilkinson Eduan; Yajna Ramphal; de Oliveira Tulio; du Plessis Jeanine |
| EPI_ISL_2693824, EPI_ISL_2693825, EPI_ISL_2693828, EPI_ISL_2693832, EPI_ISL_2693835, EPI_ISL_2693860, EPI_ISL_2693863, EPI_ISL_2693864, EPI_ISL_2693865, EPI_ISL_2693899, EPI_ISL_2893253, EPI_ISL_2893257, EPI_ISL_2893259, EPI_ISL_2893260, EPI_ISL_2893268, EPI_ISL_2893284, EPI_ISL_2893296 | | | |
| see above | NHLS_VIRO Eastern Cape | KRISP, KZN Research Innovation and Sequencing Platform | Emmanuel SJ; Giandhari J; Giandhari Jennifer; Khan S; Laguda-Akingba O; Lessells R; Mdlalose K; Naidoo Yeshnee; Pillay S; Pillay Sureshnee; San James; Tegally H; Tegally Houriiyah; Tshabuila Derek; Wilkinson E; Wilkinson Eduan; Yajna Ramphal; York D; de Oliveira T; de Oliveira Tulio |
| EPI_ISL_2727175, EPI_ISL_2727179, EPI_ISL_2727180, EPI_ISL_2727185, EPI_ISL_2727186, EPI_ISL_2727189, EPI_ISL_2727193, EPI_ISL_2727194, EPI_ISL_2727195, EPI_ISL_2727196, EPI_ISL_2727197, EPI_ISL_2727198, EPI_ISL_2727216, EPI_ISL_2727217, EPI_ISL_2955454, EPI_ISL_2955464, EPI_ISL_2955466, EPI_ISL_2955472, EPI_ISL_2955474, EPI_ISL_2955475, EPI_ISL_2955479, EPI_ISL_2955484, EPI_ISL_2955487, EPI_ISL_2955488, EPI_ISL_2955490, EPI_ISL_2955497, EPI_ISL_2955499 | | | |
| see above | NHLS_VIRO Eastern Cape | KRISP, KZn Research Innovation and Sequencing Platform | Emmanuel SJ; Giandhari J; Khan S; Laguda-Akingba O; Lessells R; Mdlalose K; Pillay S; Tegally H; Wilkinson E; York D; de Oliveira T |
| EPI_ISL_766863, EPI_ISL_766864, EPI_ISL_766865, EPI_ISL_766869, EPI_ISL_766873, EPI_ISL_766874, EPI_ISL_766875 | | | |
| see above | NIC Viral Respiratory Unit - Institut Pasteur of Algeria | National Reference Center for Viruses of Respiratory Infections, Institut Pasteur, Paris | Angela Brisebarre; Etienne Simon-Lorière; Fawzi Derrar; Flora Donati; Marion Barbet; Maud Vanpeene; Mélanie Albert; Méline Bizard; Sylvie Behillil; Sylvie van der Werf; Vincent Enouf |
| EPI_ISL_577717 | NIV Influenza | NIV Influenza | NIV; Potdar; Pune; Varsha on behalf of National Influenza Centre |
| EPI_ISL_2161796, EPI_ISL_2484617 | NL-Dr. Leonard A. Miller Centre for Health Services | National Microbiology Laboratory (NML) | Adel Malek; Anna Majer; Anneliese Landgraff; CanCOGeN's metadata curation team; Darian Hole; Elsie Grudeski; Gary Van Domselaar; George Zahariadis; Grace Seo; Jennifer Tanner; Kerri Smith; Kirsten Biggar; Laura Gilbert; Madison Chapel; Morag Graham; Natalie Knox; Nathalie Bastien; Philip Mabon; Public Health Agency of Canada CanCOGeN team; Rhiannon Huzarewich; Robert Needle; Russell Mandes; Shari Tyson; Timothy Booth; Yan Li; Yang Yu |
| EPI_ISL_960304 | NLZOH, Laboratory for Virology | NLZOH, Laboratory for Virology | Cesare Camma (IZSAM); Erik Alm (ECDC); Katarina Prosenc (Laboratory for Virology) |
| EPI_ISL_1579896, EPI_ISL_1628463, EPI_ISL_1654970 | NMVRVI | National Public Health Surveillance Laboratory | Ana Steponkiene; Arnoldas Pautienius; Astra Vitkauskiene; Danas Baksa; Dovydas Gecys; Ingrida Olendraite; Jelena Razmuk; Kamile Tamusauskaite; Laura Pareckaite; Lukas Vasionis; Lukas Zemaitis; Migle Gabrielaite; Svajune Muralyte; Vaiva Lesauskaite |
| EPI_ISL_1016162 | NORTHWELL HEALTH LABORATORIES | Wadsworth Center, New York State Department of Health | Alexis Russel; Daryl M. Lamson; Erasmus Schneider; Erica Lasek-Nesselquist; John Kelly; Jonathan Plitnick; Kirsten St. George; Matthew Shudt; Melissa A Leisner; Navjot Singh |
| EPI_ISL_416742 | NRL for Influenza, Centrum Epidemiology and Microbiology of National Institute of Public Health, Czech Republic | Charite Universitaetsmedizin Berlin, Institute of Virology | Akexander Nagy; Barbara Muehlemann; Christian Drosten; Dusan Trnka; Helena Jirincova; Jaromira Vecerova; Jörn Beheim-Schwarzbach; Julia Schneider; Ludmila Novakova; Talitha Veith; Terry Jones; Victor M Corman |
| EPI_ISL_1272388, EPI_ISL_1272389, EPI_ISL_1272390, EPI_ISL_1272391, EPI_ISL_2162047, EPI_ISL_2162051 | NS-QEII Health Sciences Centre | National Microbiology Laboratory (NML) | Anna Majer; Anneliese Landgraff; CanCOGeN's metadata curation team; Dan Gaston; Darian Hole; Elsie Grudeski; Gary Van Domselaar; Grace Seo; Janice Pettipas; Jason LeBlanc; Jennifer Tanner; Kirsten Biggar; Madison Chapel; Morag Graham; Natalie Knox; Nathalie Bastien; Philip Mabon; Public Health Agency of Canada CanCOGeN team; Rhiannon Huzarewich; Russell Mandes; Shari Tyson; Timothy Booth; Todd Hatchette; Yan Li |
| EPI_ISL_3046227 | NUPIT/UFPE | WallauLab on behalf of Fiocruz COVID-19 Genomic Surveillance Network | Alexandre Freitas da Silva; Cassia Docena; Constância Flávia Junqueira Ayres; Filipe Zimmer Dezordi; Gabriel Luz Wallau; Gustavo Barbosa de Lima; Lais Ceschini Machado; Lilian Caroliny Amorim Silva; Maira Galdino da Rocha Pitta; Marcelo Henrique dos Santos Paiva; Matheus Filgueira Bezerra; Michelly Cristiny Pereira; Rômulo Pessoa e Silva; Sinval Pinto Brandão Filho |
| EPI_ISL_903727 | NV State Public Health Laboratory | Genomics and Discovery, Respiratory Viruses Branch, Division of Viral Diseases, Centers for Disease Control and Prevention | Anna Montmayeur; Anna Uehara; Ben L. Rambo-Martin; Clinton R. Paden; Dhwani Batra; Haibin Wang; Jasmine Padilla; Jing Zhang; Justin Lee; Krista Queen; Lori Rowe; Mark Burroughs; Mili Sheth; Peter W. Cook; Rachel Marine; Sarah Nobles; Suxiang Tong; Yan Li; Ying Tao |

| | | | |
|---|---|---|---|
| EPI_ISL_1226660, EPI_ISL_1226661 | NYC Pandemic Response Lab | Wadsworth Center, New York State Department of Health | Alexis Russel; Daryl M. Lamson; Erasmus Schneider; Erica Lasek-Nesselquist; John Kelly; Jonathan Plitnick; Kirsten St. George; Matthew Shudt; Melissa A Leisner; Navjot Singh |
| EPI_ISL_1922797 | NYU Langone Health | Departments of Pathology and Medicine, New York University School of Medicine | Adriana Heguy; Christian Marier; Dacia Dimartino; Emily Guzman; Gael Westby; Guiqing Wang; Paolo Cotzia; Paul Zappile; Peter Meyn; Sitharam Ramaswami; Yutong Zhang |
| EPI_ISL_1960297, EPI_ISL_2694492 | Nacionalinis maisto ir veterinarijos rizikos vertinimo institutas | National Public Health Surveillance Laboratory | Ana Steponkiene; Danas Baksa; Jelena Razmuk; Lukas Vasionis; Lukas Zemaitis; Migle Gabrielaite; Svajune Muralyte |
| EPI_ISL_2798341, EPI_ISL_2800233, EPI_ISL_2800265, EPI_ISL_2800271 | Nacionālais medicīnas serviss - laboratorija, SIA | Riga East University Hospital, National Microbiology Reference Laboratory; Eurofins Genomics Europe Sequencing GmbH | Arzu Algulieva; Diāna Dušacka; Dārta Pūpola; Ilva Pole; Inita Balta; Jevgenijs Bodrenko; Jūlija Čevere; Nataļja Mikena; Reinis Vangravs; Reinis Zeltmatis; Sergejs Ņikišins; Ģirts Šķenders |
| EPI_ISL_523962, EPI_ISL_523966 | National Agency for Public Health, Republic of Moldova | Charite Universitatsmedizin Berlin, Institute of Virology | Ala Halacu; Barbara Mühlemann; Barbara Mühlemann; Christian Drosten; Elizabeta Jancheska; Golubinka Bosevska; Joern Beheim-Schwarzbach; Jörn Beheim-Schwarzbach; Julia Schneider; Maja Kuzmanovska; Mariana Apostol; Talitha Veith; Terry Jones; Victor M Corman |
| EPI_ISL_2894992, EPI_ISL_2894993, EPI_ISL_2894994, EPI_ISL_2894995, EPI_ISL_2894996 | National Agency for Public Health, Republic of Moldova | Charité Universitätsmedizin Berlin, Institut für Virologie | Ala Halacu; Barbara Mühlemann; Christian Drosten; Julia Schneider; Julia Tesch; Jörn Beheim-Schwarzbach; Mariana Apostol; Talitha Veith; Terry Jones; Tobias Bleicker; Victor M Corman |
| EPI_ISL_1805651, EPI_ISL_1805740, EPI_ISL_1805741 | National Center for Communicable Diseases (NCCD) National Influenza Center | National Center for Communicable Diseases (NCCD) National Influenza Center | Ankhbayar S; Battur L; Bayasgalan N; Darmaa B; Hideka M; Khishigmunkh Ch; Mina N; Naranzul Ts; Nymadawa P; Seiichiro F; Shinji W; Tsogzolmaa G |
| EPI_ISL_1805697 | National Center for Communicable Diseases (NCCD) National Influenza Center | National Centre for Disease Control (NCDC) National Influenza Center | Ankhbayar S; Battur L; Bayasgalan N; Darmaa B; Hideka M; Khishigmunkh Ch; Mina N; Naranzul Ts; Nymadawa P; Seiichiro F; Shinji W; Tsogzolmaa G |
| EPI_ISL_1233442 | National Center for Infectious and Parasitic Diseases (NCIPD) | National Center for Infectious and Parasitic Diseases (NCIPD) | Alexiev et al |
| EPI_ISL_1718293, EPI_ISL_1718294 | National Center of Disease Control and Prevention of the Republic of Armenia | Institute of Molecular Biology NAS RA, Republic of Armenia, Department of Bioengineering, BioinformaticsInstitute and Molecular Biology IBMPh RAU, Republic of Armenia | Andranik Chavushyan; Arsen Arakelyan; Diana Avetyan; Gayane Melik-Pashayan; Gisane Khachatyan; Hovsep Ghazaryan; Maria Nikoghosyan; Nelli Muradyan; Roksana Zakharyan; Shushan Sargrsyan; Siras Hakobyan; Tamara Sirunyan; Zaven Karalyan |
| EPI_ISL_1854607, EPI_ISL_1854610, EPI_ISL_1854624, EPI_ISL_1854625, EPI_ISL_1854638, EPI_ISL_1854647 | National Center of Disease Control and Prevention of the Republic of Armenia | UW Virology Lab | Alexander Greninger; Anahit Hovhannisyan; Andranik Chavushyan; Anna Khazaryan; Arsen Arakelyan; Diana Avetyan; Gisane Khachatyan; Hong Xie; Hovsep Ghazaryan; Keith R. Jerome; Lasata Shrestha; Lyudmila Niazyan; Maria Nikoghosyan; Meei-Li Huang; Michelle Lin; Nelli Muradyan; Pavitra Roychoudhury; Roksana Zakharyan; Shah Mohamed Bakhash; Siras Hakobyan; Tamara Sirunyan |
| EPI_ISL_454572 | National Center of Expertise | National Center for Expertise, Kazakhstan National Center for Biotechnology, Kazakhstan | ; Abdaliyev Askar; Akhmetollayev Ilyas; Amirgazin Asylulan; Aushakhmetova Zabira; Kalendar Ruslan; Lutsay Viktoriya; Rakhmetova Akbota; Ramankulov Yerlan; Shevtsov Alexandr |
| EPI_ISL_454571 | National Center of Expertise | National Center for Expertise, National Center for Biotechnology, Kazakhstan | ; Abdaliyev Askar; Akhmetollayev Ilyas; Amirgazin Asylulan; Aushakhmetova Zabira; Kalendar Ruslan; Lutsay Viktoriya; Rakhmetova Akbota; Ramankulov Yerlan; Shevtsov Alexandr |
| EPI_ISL_1786307, EPI_ISL_2081876, EPI_ISL_2081915, EPI_ISL_2081935, EPI_ISL_2099343, EPI_ISL_2099441, EPI_ISL_2099454, EPI_ISL_2180209, EPI_ISL_2379203, EPI_ISL_2621269, EPI_ISL_2621400, EPI_ISL_2621428, EPI_ISL_2621449, EPI_ISL_2648098, EPI_ISL_2648102, EPI_ISL_2841870, EPI_ISL_2841883, EPI_ISL_2841901, EPI_ISL_2841982, EPI_ISL_2967059, EPI_ISL_2967087, EPI_ISL_2967113, EPI_ISL_2967132, EPI_ISL_2967172 | | | |
| see above | National Center of Infectious and Parasitic Diseases | National Center of Infectious and Parasitic Diseases | Alexiev; Alexiev et al; Dimitrova; Dobrinov; Donchev; Grigorova I.; Grigorova L.; Hristova; Ivanov; Kantardjiev; Korsun; Philipova; Stoitsova; Stoykov; Trifonova |
| EPI_ISL_2332763 | National Centre For Cell Science | National Centre For Cell Science – INSACOG | Ajay Pillai; Dhiraj Paul; INSACOG Consortium team; Manoj Kumar Bhat; Mitali Inamdar; Mohak P Gujare; Shivang P. Bhanushali; Yogesh Shouche |
| EPI_ISL_2459215, EPI_ISL_2459216, EPI_ISL_2459690, EPI_ISL_2459936 | National Centre for Disease Control (NCDC) Biotechnology Division, Delhi | NCDC Delhi, Biotechnology Division INSACOG | Hema Gogia; Hemlata Lall; Kalaiarasan Ponnusamy; Mahesh S Dhar; Manoj K Singh; Meena Datta; Partha Rakshit; Preeti Madan; Priyanka Singh; Radhakrishnan V. S; Robin Marwal; Sandhya Kabra; Sujeet K Singh; Uma Sharma |
| EPI_ISL_1661758 | National Food and Veterinary Risk Assessment Institute | Vilnius University Hospital Santaros Klinikos, Center of Laboratory Medicine | Daniel Naumovas; Dovile Ezerskyte; Gytis Dudas; Ingrida Olendraite; Laimonas Griskevicius; Ligita Raugaite; Mindaugas Stoskus; Monika Katenaite; Rimvydas Norvilas |
| EPI_ISL_1909873 | National Food and Veterinary Risk Assessment Institute (NMVRVI) | National Public Health Surveillance Laboratory | Ana Steponkiene; Danas Baksa; Jelena Razmuk; Lukas Vasionis; Lukas Zemaitis; Migle Gabrielaite; Svajune Muralyte |
| EPI_ISL_528584, EPI_ISL_528585 | National Genomics Core-Center for DNA Fingerprinting and Diagnostics | National Genomics Core- Center for DNA Fingerprinting and Diagnostics (NGC-CDFD)- DBT's PAN-INDIA-1000 Genome consortium | Abhijeeth Singh Thakur; Ashwin Dalal; Bala Pratyusha; Bathula Siddardha; Debashish Mitra; Divya Vashisht; G Shashikanth; Heena Shah; Lavanya Banda; Mohammad Mudassir; Murali Dharan Bashyam; Nalini Raghunathan; R Harinarayanan; Rajeshree Sanyal; Rashna Bhandari; Surya Chodisetty; Vinay Donipadi; Vineesha Oddi |
| EPI_ISL_2492627, EPI_ISL_2492631, EPI_ISL_2492642, EPI_ISL_2492643, EPI_ISL_2492664, EPI_ISL_2492697, EPI_ISL_2492710, EPI_ISL_2493032, EPI_ISL_2493033, EPI_ISL_2493049, EPI_ISL_2493052, EPI_ISL_2493054, EPI_ISL_2493056, EPI_ISL_2493057, EPI_ISL_2686025 | | | |
| see above | National HIV Reference Laboratory, Ministry of Health, Public Health Institute of Malawi | CERI, Centre for Epidemic Response and Innvoation, Stellenbosch University and KRISP, KZN Research Innovation and Sequencing Platform, UKZN. | Auld A; Chilima B; Chiwaula M; Emmanuel SJ; Giandhari J; Kaba M; Kampira E; Kasambara W; Kim L; Lessells R; Maida A; Mvula B; Mwangomba W; Naidoo Y; Panja L; Pillay S; Tegally H; Wadonda N; Wilkinson E; de Oliveira T |
| EPI_ISL_1407121, EPI_ISL_1407184, EPI_ISL_1407198, EPI_ISL_2494782, EPI_ISL_2494844, EPI_ISL_2494924 | National HIV Reference Laboratory, Ministry of Health, Public Health Institute of Malawi | KRISP, KZN Research Innovation and Sequencing Platform | Auld A; Chilima B; Chiwaula M; Emmanuel SJ; Giandhari J; Kaba M; Kampira E; Kasambara W; Kim L; Lessells R; Maida A; Mvula B; Mwangomba W; Naidoo Y; Panja L; Pillay S; Tegally H; Wadonda N; Wilkinson E; de Oliveira T |
| EPI_ISL_560387, EPI_ISL_560388, EPI_ISL_1677703, EPI_ISL_1677707, EPI_ISL_1677716 | National Health Laboratory | Botswana Institute for Technology Research and Innovation | Dineo Emang Tshiamo. Gape Nyepetsi; Dineo Emang Tshiamo. Tefelo Thela; Gape Nyepetsi; Kefentse Arnold Tumedi; Madisa Mine; Maitshwarelo Ignatius Matsheka; Malebogo Kebabonye; Thongbotho Mphoyakgosi |
| EPI_ISL_622942, EPI_ISL_622976, EPI_ISL_622980, EPI_ISL_622984, EPI_ISL_622989, EPI_ISL_622991 | National Health Laboratory Service | National Institute for Communicable Diseases of the National Health Laboratory Service | Allam M; Bhiman JN; Ismail A; Khumalo Z; Kwenda S; Mnyameni F; Mohale T; Mtshali P; Subramoney K |
| EPI_ISL_944143, EPI_ISL_944163, EPI_ISL_1250455, EPI_ISL_1817756, EPI_ISL_1817763, EPI_ISL_1817766, EPI_ISL_1817767, EPI_ISL_1817859, EPI_ISL_2086228, EPI_ISL_2086229, EPI_ISL_2086238, EPI_ISL_2086243, EPI_ISL_2494834, EPI_ISL_2494948 | | | |
| see above | National Health Laboratory Service, South Africa | KRISP, KZN Research Innovation and Sequencing Platform | Emmanuel SJ; Giandhari J; Khan S; Laguda-Akingba O; Lessells R; Maslo C; Mdlalose K; Naidoo Y; Pillay S; Ramphal U; Sitharam L; Tegally H; Wilkinson E; York D; de Oliveira T |
| EPI_ISL_2617196, EPI_ISL_2617201, EPI_ISL_2617210, EPI_ISL_2688538, EPI_ISL_2688543, EPI_ISL_2688549, EPI_ISL_2688550, EPI_ISL_2688553, EPI_ISL_2688563, EPI_ISL_2688564, EPI_ISL_2688573, EPI_ISL_2899689, EPI_ISL_2899695, EPI_ISL_2899725, EPI_ISL_2899740 | | | |
| see above | National Health Laboratory Service, South Africa | KRISP, KZn Research Innovation and Sequencing Platform | Giandhari Jennifer; Naidoo Yeshnee; Pillay Sureshnee; San James; Tegally Houriiyah; Tshabuila Derek; Wilkinson Eduan; Yajna Ramphal; de Oliveira Tulio |
| EPI_ISL_456597, EPI_ISL_456598, EPI_ISL_456601, EPI_ISL_456605, EPI_ISL_456610 | National Health Laboratory, Timor-Leste | Microbiological Diagnostic Unit Public Health Laboratory, The Peter Doherty Institute for Infection and Immunity | Antonia da Costa, E.; Baird, R.; Barreto, I.; Caly, L.; Canisia, D.; Dakh, F.; Dolores de Jesus da Costa, M.; Douglas, N.; Francis, J.; Freeman, K.; Jayanti Pereira Tilman, A.; Marr, I.; Sait, M.; Salles de Sousa, A.; Schultz, M.; Seemann, T.; Sherry, N.; Soares da Silva, E.; Wapling, J.; Ximenes, J. |
| EPI_ISL_2379748 | National Heart Institute | Institute for Medical Research, Infectious Disease Research Centre, National Institutes of Health, Ministry of Health Malaysia | Azizan MA; Kamel K; Mohd Zawawi Z; Ramly N; Robert F; Suppiah J; Thayan R |
| EPI_ISL_2434158 | National Hospital for Tropical Diseases | Oxford University Clinical Research Unit, Hanoi | H.Rogier van Doorn on behalf of the OUCRU COVID-19 research group; Le Van Duyet; Nguyen Thi Hong Thuong; Nguyen Thi Kim Chi; Nguyen Thi Tam; Nguyen Thu Trang; Pham Ngoc Thach; Phan Manh Cuong; Thomas Kesteman; Van Dinh Trang |
| EPI_ISL_2455479, EPI_ISL_2455488, EPI_ISL_2455494, EPI_ISL_2455495, EPI_ISL_2455496, EPI_ISL_2455497, EPI_ISL_2455498, EPI_ISL_2455500 | | | |
| see above | National Hospital for Tropical Diseases | Oxford University Clinical Research Unit, Hanoi, Vietnam | H.Rogier van Doorn on behalf of the OUCRU COVID-19 research group; Le Van Duyet; Nguyen Thi Hong Thuong; Nguyen Thi Kim Chi; Nguyen Thi Tam; Nguyen Thu Trang; Pham Ngoc Thach; Phan Manh Cuong; Thomas Kesteman; Van Dinh Trang |
| EPI_ISL_511898 | National Hospital of Tropical Diseases | Oxford University Clinical Research Unit, Hanoi, Vietnam | H. Rogier van Doorn; Le Nguyen Minh Hoa; Nguyen Thi Hong Thuong; Nguyen Thi Ngoc Diep; Nguyen Thi Tam; Nguyen Thu Trang; Pham Ngoc Thach; Van Dinh Trang; Vu Thi Ngoc Bich; on behalf of the OUCRU COVID-19 research group |
| EPI_ISL_2695785 | National Influenza Center (NIC). Virology department. Institut National d'Hygiène. | National Influenza Center (NIC). Virology department. Institut National d'Hygiène. | Abderrahmane BIMOUHEN; Hassan IHAZMAD; Hicham OUMZIL; Samira BENKERROUM; Zakia REGRAGUI; fatima EL FALAKI and Mohamed RHAJAOUI |
| EPI_ISL_486889 | National Influenza Center, Bahrain | National Influenza Center, Bahrain | AlWasti, H.; Altaif, Z.; Shehab, F.; Zaed, A. |
| EPI_ISL_862077, EPI_ISL_862078, EPI_ISL_1014676, EPI_ISL_1014681, EPI_ISL_1993547, EPI_ISL_1993552, EPI_ISL_1993554, EPI_ISL_1993557 | | | |
| see above | National Influenza Center, Virology Department | National Influenza Center | A Nejati; F Ajaminejad; F Ajaminejad and T Mokhtari Azad; J Yavarian; K Sadeghi; N Ghavvami; N Ghavvami and T Mokhtari Azad; NZ Shafiei Jandaghi; V Salimi |

| | | | |
|---|---|---|---|
| EPI_ISL_2285849, EPI_ISL_2285850, EPI_ISL_2285852, EPI_ISL_2285853, EPI_ISL_2285855, EPI_ISL_2285864, EPI_ISL_2285869, EPI_ISL_2535795, EPI_ISL_2535796, EPI_ISL_2873841, EPI_ISL_2873845, EPI_ISL_2873850, EPI_ISL_2873858, EPI_ISL_2873860, EPI_ISL_2873865, EPI_ISL_2873870, EPI_ISL_2873872 | | | |
| see above | National Influenza Centre | National Influenza Centre | ; Benjamin B. Lindsey; Benjamin H. Foulkes; Dennis Laryea; Ernest Asiedu; Franklin Asiedu-Bekoe; Gordon Awandare; Ivy A. Asante; Joseph Oliver-Commey; Joyce Ngoi; Linda Boatemaa; Lorreta Kwasah; Mathew D. Parker; Michael Marks; Mildred Adusei-Poku; Sharon Hsu; Thushan I de Silva; William K. Ampofo |
| EPI_ISL_476838 | National Influenza Centre for Northern Greece | National Influenza Centre for Northern Greece | Maria Christoforidi |
| EPI_ISL_410301 | National Influenza Centre, National Public Health Laboratory, Kathmandu, Nepal | The University of Hong Kong | Alfonso J. Rodriguez-Morales; Anup Bastola; Basu Dev Pandey; Bibek Kumar Lal; Daniel Chu; Haogao Gu; Leo Poon; Malik Peiris; Ranjit Sah; Runa Jha |
| EPI_ISL_402125 | National Institute for Communicable Disease Control and Prevention (ICDC) Chinese Center for Disease Control and Prevention (China CDC) | National Institute for Communicable Disease Control and Prevention (ICDC) Chinese Center for Disease Control and Prevention (China CDC) | Chen; Dai; F.-H.; Hu, Y.; J.-H.; J.-J.; J.-L. and Zhu; Liu, Y.; Pei; Q.-M.; She; Song; T.-Y.; Tao; Tian; Wang; Wang, W.; Wu, F.; Xu, L.; Y.-L.; Y.-M.; Y.-Y.; Y.-Z.; Yu, B.; Z.-G.; Z.-W.; Zhang; Zhao, S.; Zheng |
| EPI_ISL_450297, EPI_ISL_450299, EPI_ISL_450300, EPI_ISL_450301, EPI_ISL_450495, EPI_ISL_470878, EPI_ISL_490281, EPI_ISL_514417, EPI_ISL_514419, EPI_ISL_515177, EPI_ISL_622930, EPI_ISL_2433725, EPI_ISL_2433832, EPI_ISL_2433834, EPI_ISL_2444005, EPI_ISL_2444020, EPI_ISL_2662723 | | | |
| see above | National Institute for Communicable Diseases of the National Health Laboratory Service | National Institute for Communicable Diseases of the National Health Laboratory Service | Allam M; Amoako DG; Bhiman JN; Everatt J; Ismail A; Khumalo Z; Kwenda S; Mahlangu B; Mnguni A; Mnyameni F; Mohale T; Mtshali P; Ntuli N; Scheepers C; Subramoney K; van Heusden P |
| EPI_ISL_1914886, EPI_ISL_1914931 | National Institute for Communicable Diseases,National Health Laboratory Services, Gauteng, South Africa | National Institute for Communicable Diseases of the National Health Laboratory Service | Amoako DG; Bhiman JN; Ismail A; Mahlangu B; Mohale T; Ntuli N; Scheepers C |
| EPI_ISL_1911458 | National Institute for Food and Veterinary Risk Assessment (NMVRVI) | National Public Health Surveillance Laboratory | Ana Steponkiene; Danas Baksa; Jelena Razmuk; Lukas Vasionis; Lukas Zemaitis; Migle Gabrielaite; Svajune Muralyte |
| EPI_ISL_469254 | National Institute for Viral Disease Control and Prevention, China CDC | Institute of Viral Disease Control and Prevention, China CDC | Chun Huang; Dayan Wang; George Fu Gao; Guizhen Wu; Li Zhao; Lijuan Chen; Peihua Niu , Baoying Huang; Roujian Lu; Wenbo Xu; Wenjie Tan; Wenling Wang; Yubai Bi |
| EPI_ISL_591270, EPI_ISL_591276, EPI_ISL_850947, EPI_ISL_850949 | National Institute for Viral Disease Control and Prevention, China CDC | National Institute for Viral Disease Control and Prevention, China CDC | Cao Chen; Dayan Wang; George F.Gao; Hong Wang; Huilai Ma; Ji Wang; Jingdong Song; Jun Han; Kai Nie; Ruqin Gao; Shiwen Wang; Weimin Zhou; Wenbo Xu; Wenjie Tan; Xiang Zhao; Yang Song; Yanhai Wang; Yao Meng; Yenan Feng; Yong Zhang; Yuchao Wu; Zhaoguo Wang; Zhixiao Chen |
| EPI_ISL_709542 | National Institute of Blood Diseases (NIBD), Molecular Biology Lab | Genomics Lab NIBD | Aneeta Shahni; Arshi Naz; Gul Sufaida; Samina Naz Mukry; Sayed Ali Raza; Shariq Ahmed; Tahir Sultan Shamsi |
| EPI_ISL_2107094, EPI_ISL_2107095, EPI_ISL_2107098, EPI_ISL_2308255, EPI_ISL_2308256, EPI_ISL_2308259 | National Institute of Health (NIH) - Federal Government of Somalia | African Centre of Excellence for Genomics of Infectious Diseases (ACEGID), Redeemer's University | A.T.; Abechi; Ajogbasile; Akano; C.A.; C.T.; Eromon; F.V.; Folarin, O.; Happi; I.B.; J.N.; J.U.; K.O.; Kayode; Nosamiefan, I.; Oguzie; Olawoye; Olumade; Oluniyi; P.E.; P.S.; T.J.; Ugwu; Uwanibe |
| EPI_ISL_833130, EPI_ISL_1915513 | National Institute of Health Research and Development | National Institute of Health Research and Development | A; AA; Adam; Agustiningsih; Arie Ardiansyah Nugraha; F; Fauzul Muna; Febriyani; Febriyanti; HA; HD; Hana Apsari Pawestri; Hariastuti; Hartanti Dian Ikawati; Herman; Herna; Holy Arif Wibowo; IL; Ikawati; Indalao; Irene Lorinda Indalao; J; K; KD; KNA; Kartika Dewi Puspa; Kindi Adam; Kipuw; Krisna Nur Andriana Pangesti; Kurniawati; Muna; Mursinah; N; NI; NK; NL; Natalie Laurencia Kipuw; Nelly Puspandari; Ni Ketut Susilarini; Nike Susanti; Nikmah; Nugraha; Nurika Hariastuti; Pangesti; Pawestri; Puspa; Puspandari; R; Ramadhany; Reni Herman; Ririn Ramadhany; Rizki; Rukminiati; Setiawaty; Soekarso; Subangkit; Susanti; Susilarini; T; Tati Febriyanti; Triyani Soekarso; UA; Ully Alfi Nikmah; V; Vivi Setiawaty.; Wibowo; Y; Yuni Rukminiati |
| EPI_ISL_985395, EPI_ISL_1116455, EPI_ISL_1279947, EPI_ISL_1279951, EPI_ISL_1279953, EPI_ISL_1279966, EPI_ISL_1279972, EPI_ISL_2190103, EPI_ISL_2308251, EPI_ISL_2932503 | | | |
| see above | National Institute of Infectious Diseases-Prof. Dr. Matei Bals Molecular Diagnostics Laboratory | National Institute of Infectious Diseases-Prof. Dr. Matei Bals Molecular Diagnostics Laboratory | Andreea Tudor; Corina Casangiu; Dan Otelea; Leontina Banica; Marius Surleac; Ovidiu Vlaicu; Petre Milu; Robert Hohan; Simona Paraschiv |
| EPI_ISL_943552, EPI_ISL_955172, EPI_ISL_2796523, EPI_ISL_2796527 | National Institute of Laboratory Medicine and Referral Center | Genomic Research Lab, BCSIR | ; A. K. M. Shamsuzzaman; A. K. M. Shamsuzzaman; Abu Sayeed Mohammad Mahmud; Arifa Akram; Asish Kumar Ghosh; Barna Goswami; Eshrar Osman; Iffat Jahan; Mahmuda Yeasmin; Md. Ahasan Habib; Md. Ahashan Habib; Md. Kamrul Islam; Md. Maruf Ahmed Molla; Md. Murshed Hasan Sarkar; Md. Saddam Hossain; Md. Salim Khan; Md. Salim Khan.; Mohammad Mohi Uddin; Mohammad Samir Uzzaman; Shahina Akter; Tanjina Akhtar Banu; Tanjina Akhter Banu; Tasnim Nafisa; Tasnim Nafisa |
| EPI_ISL_2323222, EPI_ISL_2323258, EPI_ISL_2380403 | National Institute of Public Health | Charles University, Faculty of Science, BIOCEV, OMICS Genomics | Blanka Hamplová; Ingrid Poláková; Jana Šmahelová; Jiří Novák; Magdalena Jančářová; Ruth Tachezy; Sebastian Cristian Treitli; Vladimír Hampl; Zoltán Füssy; Štěpánka Hrdá |
| EPI_ISL_2709618, EPI_ISL_2709627 | National Institute of Public Health | Institute of Molecular Genetics CAS | Jan Pačes; Jana Šáchová; Lucie Pfeiferová; Martin Zmuda; Michal Kolář; Miluše Hradilová; Ondřej Moravčík |
| EPI_ISL_2687993 | National Institute of Public Health | National Institute of Infectious Diseases-Prof. Dr. Matei Bals Molecular Diagnostics Laboratory | Corina Casangiu; Dan Otelea; Leontina Banica; Marius Surleac; Petre Milu; Robert Hohan; Simona Paraschiv |
| EPI_ISL_1492241, EPI_ISL_2324674, EPI_ISL_2324821, EPI_ISL_2324940 | National Institute of Public Health | National Institute of Public Health | Alexander Nagy; Dusan Trnka; Helena Jirincova; Jaromira Vecerova; Timotej Suri |
| EPI_ISL_1828716, EPI_ISL_1828718, EPI_ISL_1828724, EPI_ISL_1971075, EPI_ISL_2562079 | National Institute of Public Health | State Veterinary Institute Prague | A; D; H; J; Jirincova; Nagy; Suri; T; Trnka; Vecerova |
| EPI_ISL_2928004, EPI_ISL_2928005, EPI_ISL_2928006, EPI_ISL_2928007, EPI_ISL_2928008, EPI_ISL_2928009, EPI_ISL_2928010, EPI_ISL_2928011, EPI_ISL_2928012 | | | |
| see above | National Institute of Public Health Burundi, WHO Burundi, ASLM Burundi, MRC/UVRI & LSHTM Uganda Research Unit | MRC/UVRI & LSHTM Uganda Research Unit, National Institute of Public Health Burundi, WHO Burundi, ASLM Burundi | Alexis Niyomwungere; Anatole Nkeshimana; Blessing T. Marondera; Butoyi Pascal; Cassien Nduwimana; Dan Lule Bugembe; Eric Kezakarayagwa; Francine Kabatesi; Jean Baptiste Niyibigira; Jerome Ndaruhutse; Jerome Nkurunziza; Joseph Nyandwi; Kouadio Theodore Yao; Matthew Cotten; My V.T. Phan; Xavier Crespin |
| EPI_ISL_1335479, EPI_ISL_1335489 | National Laboratory for Health, Environment and Food, OMM, Celje | CISLD (Clinical Institute of Special Laboratory Diagnostics), University Children's Hospital, University Medical Center Ljubljana | Ana Grom; Barbara Jenko Bizjan; Jernej Kovač; Katarina Kozmos; Marko Pokorn; Maruša Debeljak; Robert Šket; Tadej Battelino; Tine Tesovnik |
| EPI_ISL_1664811, EPI_ISL_2001884 | National Laboratory for Health, Environment and Food, OMM, Celje | NLZOH (National Laboratory for Health, Environment and Food) / CISLD (Clinical Institute of Special Laboratory Diagnostics), University Children's Hospital, University Medical Center Ljubljana | Aleksander Kocuvan; Aleksander Mahnic; Alenka Štorman; Ana Grom; Barbara Jenko Bizjan; Daša Kavka / Jernej Kovač; Kaja Tominc; Katarina Kozmos; Maja Rupnik; Marko Pokorn; Maruša Debeljak; Mateja Borinc; Maša Jarčič; Nika Gobec; Robert Šket; Sandra Janezic; Tadej Battelino; Tine Tesovnik; Tjasa Žohar Čretnik |
| EPI_ISL_2789267, EPI_ISL_2983030 | National Laboratory for Health, Environment and Food, OMM, Koper | NLZOH (National Laboratory for Health, Environment and Food) / CISLD (Clinical Institute of Special Laboratory Diagnostics), University Children's Hospital, University Medical Center Ljubljana | Aleksander Kocuvan; Aleksander Mahnic; Alenka Štorman; Ana Grom; Barbara Jenko Bizjan; Gašper Strugar; Kaja Tominc; Katarina Kozmos; Maja Rupnik; Marko Pokorn; Maruša Debeljak; Maša Jarčič; Mitja Rak / Jernej Kovač; Nika Gobec; Robert Šket; Sandra Janezic; Tadej Battelino; Tina Cvetković; Tine Tesovnik; Tjaša Žohar Čretnik |
| EPI_ISL_2162621 | National Laboratory for Health, Environment and Food, OMM, Kranj | NLZOH (National Laboratory for Health, Environment and Food) / CISLD (Clinical Institute of Special Laboratory Diagnostics), University Children's Hospital, University Medical Center Ljubljana | Aleksander Kocuvan; Aleksander Mahnic; Alenka Štorman; Ana Grom; Barbara Jenko Bizjan; Kaja Tominc; Katarina Kozmos; Maja Rupnik; Marjana Petrevčič / Jernej Kovač; Marko Pokorn; Maruša Debeljak; Mateja Ravnik; Maša Jarčič; Monika Korošec; Nika Gobec; Robert Šket; Sandra Janezic; Tadej Battelino; Tine Tesovnik; Tjasa Žohar Čretnik |
| EPI_ISL_1111786, EPI_ISL_1111889 | National Laboratory for Health, Environment and Food, OMM, Maribor | CISLD (Clinical Institute of Special Laboratory Diagnostics), University Children's Hospital, University Medical Center Ljubljana | Ana Grom; Barbara Jenko Bizjan; Jernej Kovač; Katarina Kozmos; Marko Pokorn; Maruša Debeljak; Robert Šket; Tadej Battelino; Tine Tesovnik |
| EPI_ISL_1664818, EPI_ISL_2162480, EPI_ISL_2162562, EPI_ISL_2162583, EPI_ISL_2463348, EPI_ISL_2463442, EPI_ISL_2789297, EPI_ISL_2982921 | | | |
| see above | National Laboratory for Health, Environment and Food, OMM, Maribor | NLZOH (National Laboratory for Health, Environment and Food) / CISLD (Clinical Institute of Special Laboratory Diagnostics), University Children's Hospital, University Medical Center Ljubljana | Aleksander Kocuvan; Aleksander Mahnic; Alenka Štorman; Ana Grom; Andrej Golle / Jernej Kovač; Barbara Jenko Bizjan; Kaja Tominc; Katarina Kozmos; Maja Rupnik; Marko Pokorn; Maruša Debeljak; Maša Jarčič; Mojca Cimerman; Nika Gobec; Nika Volmajer; Robert Šket; Sandra Janezic; Tadej Battelino; Tine Tesovnik; Tjasa Žohar Čretnik; Tjaša Žohar Čretnik |
| EPI_ISL_2789283 | National Laboratory for Health, Environment and Food, OMM, Novo mesto | NLZOH (National Laboratory for Health, Environment and Food) / CISLD (Clinical Institute of Special Laboratory Diagnostics), University Children's Hospital, University Medical Center Ljubljana | Aleksander Kocuvan; Aleksander Mahnic; Alenka Štorman; Ana Grom; Barbara Jenko Bizjan; David Cvetko; Kaja Tominc; Katarina Kozmos; Maja Rupnik; Marko Pokorn; Maruša Debeljak; Matjaž Retelj / Jernej Kovač; Maša Jarčič; Nika Gobec; Robert Šket; Sandra Janezic; Tadej Battelino; Tatjana Harlander; Tine Tesovnik; Tjaša Žohar Čretnik |
| EPI_ISL_512612, EPI_ISL_512613, EPI_ISL_512636 | National Laboratory for Influenza/Virology reference laboratory, Public Health Center of the Ministry of Health of Ukraine | Respiratory Virus Unit, Microbiology Services Colindale, Public Health England | Dr. Iryna Demchyshyna; PHE Covid Sequencing Team |
| EPI_ISL_647980, EPI_ISL_1191831, EPI_ISL_1191921, EPI_ISL_1192066 | National Microbiology Reference Laboratory | Quadram Institute Bioscience | Agnes Juru; Alexander Goredema; Ana-Victoria Gutierrez; Andrew J. Page; Andrew Tarupiwa; Barbra Murwira; Beuty Makamure; Charles Nyagupe; David Baker; Faustinos T Takawira; Gaetan Thilliez; Gemma Kay; Gibson Mhlanga; Hlanai Gumbo; Isaac Phiri; Justin O'Grady; Kenneth K Maeka; Leonardo de Oliveira Martins; Muchaneta Mugabe; Portia Manangazira; Raiva Simbi; Robert Kingsley; Sekesai Zinyowera; Tapfumanei Mashe; Tatenda Takawira; Thanh Le Viet |
| EPI_ISL_2492582, EPI_ISL_2492707, EPI_ISL_2492718, EPI_ISL_2492732, EPI_ISL_2492775, EPI_ISL_2492860, EPI_ISL_2492916 | | | |

| | | | |
|---|---|---|---|
| see above | National Microbiology Reference Laboratory, Ministry of Health, Harare, Zimbabwe | CERI, Centre for Epidemic Response and Innvoation, Stellenbosch University and KRISP, KZN Research Innovation and Sequencing Platform, UKZN. | Agnes Juru; Air Comodor Dr J. Chimedza; Charles Nyagupe; Dr Raiva Simbi; Emmanuel SJ; Giandhari J; Hlanai Gumbo; Kenneth Maeka; Naidoo Y; Pillay S; Tapfumanei Mashe; Tatenda Takawira; Tegally H; Wilkinson E; de Oliveira T |
| EPI_ISL_1095620 | National Public Health Center, COVID Laboratory | National Public Health Center, National Biosafety Laboratory | Bernadett Pályi; Dániel Déri; Judit Henczkó; Norbert Solymosi; Nóra Magyar; Zoltán Kis |
| EPI_ISL_2479956, EPI_ISL_2479957, EPI_ISL_2479972, EPI_ISL_2479975, EPI_ISL_2479981, EPI_ISL_2479985 | National Public Health Laboartory | CSIR-Institute of Genomics and Integrative Biology | Jyoti Acharya; Lokbandhu Chaudhary; Priya Jha; Runa Jha; Suni Dangol |
| EPI_ISL_2674091, EPI_ISL_2674102 | National Public Health Laboratory | National Public Health Laboratory/CSIR-Institute of Genomics and Integrative Biology | National Public Health Laboratory Team |
| EPI_ISL_845548, EPI_ISL_845549, EPI_ISL_845550, EPI_ISL_845551, EPI_ISL_845563 | National Public Health Laboratory, Cameroon | African Centre of Excellence for Genomics of Infectious Diseases (ACEGID), Redeemer's University | Oluniyi P.E. et al |
| EPI_ISL_2450778, EPI_ISL_2450783, EPI_ISL_2450789, EPI_ISL_2450791 | National Public Health Laboratory, Ministry of Health, Ministry of Health, Republic of South Sudan | South Sudan Ministry of Health, WHO South Sudan, MRC/UVRI & LSHTM Uganda Research Unit | Abe G. Abias; Dan Lule Bugembe; Dennis Kenyi Lodiongo; James Ayei; John Rumunu; Joseph Francis Wamala; Juma John HM; Lul Lojok Deng; Matthew Cotten; My V.T. Phan; Pontiano Kaleebu; Richard Lino Loro Lako; Sudhir Bunga |
| EPI_ISL_428822, EPI_ISL_428830, EPI_ISL_428841, EPI_ISL_435678, EPI_ISL_435686, EPI_ISL_462300, EPI_ISL_493408, EPI_ISL_498594, EPI_ISL_498599, EPI_ISL_516827, EPI_ISL_536427, EPI_ISL_536432, EPI_ISL_548976, EPI_ISL_574541, EPI_ISL_626631, EPI_ISL_626636, EPI_ISL_645124, EPI_ISL_693316, EPI_ISL_693328, EPI_ISL_754076, EPI_ISL_754080, EPI_ISL_1098834, EPI_ISL_1519384, EPI_ISL_1652095, EPI_ISL_1652199, EPI_ISL_1704809, EPI_ISL_1816946, EPI_ISL_1816953, EPI_ISL_1816955, EPI_ISL_1897609, EPI_ISL_1897620, EPI_ISL_2349711, EPI_ISL_2349779, EPI_ISL_2349819, EPI_ISL_2349865, EPI_ISL_2508703, EPI_ISL_2508993, EPI_ISL_2508995, EPI_ISL_2509024, EPI_ISL_2509025, EPI_ISL_2509027, EPI_ISL_2509028, EPI_ISL_2509091, EPI_ISL_2621909, EPI_ISL_2621948, EPI_ISL_2621955, EPI_ISL_2710081, EPI_ISL_2790332, EPI_ISL_2887792, EPI_ISL_2982247, EPI_ISL_2982249, EPI_ISL_3026034 | | | |
| see above | National Public Health Laboratory, National Centre for Infectious Diseases | National Public Health Laboratory, National Centre for Infectious Diseases | Chavatte JM; Chavatte Jean-Marc; Cui L; Cui Lin; Grace Jie Yin Ngan; Katherine Ching; Lin Cui; Lin RTP; Lin Raymond Tzer Pin; Mak TM; Mak Tze Minn; Octavia S; Octavia Sophie; Raymond Tzer Pin Lin; Royce Ang; Sophie Octavia; Tze Minn Mak; Zhenyang Zhou; Zhou Z |
| EPI_ISL_2674074 | National Public Health Laboratory/B.P Koirala Institute of Health Science | National Public Health Laboratory/CSIR-Institute of Genomics and Integrative Biology | National Public Health Laboratory Team /B.P Koirala Institute of Health Science COVID 19 PCR Team |
| EPI_ISL_2674069 | National Public Health Laboratory/Palpa HospitalCOVID-19 Lab | National Public Health Laboratory/CSIR-Institute of Genomics and Integrative Biology | National Public Health Laboratory Team /Palpa HospitalCOVID-19 PCR Team |
| EPI_ISL_2674065 | National Public Health Laboratory/Pokhara Academy of Health Sciences | National Public Health Laboratory/CSIR-Institute of Genomics and Integrative Biology | National Public Health Laboratory Team /Pokhara Academy of Health Sciences COVID 19 PCR Team |
| EPI_ISL_2674086 | National Public Health Laboratory/Provincial Public Health Laboratory -2 | National Public Health Laboratory/CSIR-Institute of Genomics and Integrative Biology | National Public Health Laboratory Team /Provincial Public Health Laboratory -2 Team |
| EPI_ISL_2674070 | National Public Health Laboratory/Provincial Public Health Laboratory -Gandaki | National Public Health Laboratory/CSIR-Institute of Genomics and Integrative Biology | National Public Health Laboratory Team /Provincial Public Health Laboratory -Gandaki Team |
| EPI_ISL_2674108 | National Public Health Laboratory/Provincial Public Health Laboratory -Lumbini | National Public Health Laboratory/CSIR-Institute of Genomics and Integrative Biology | National Public Health Laboratory Team /Provincial Public Health Laboratory -Lumbini Team |
| EPI_ISL_2349378 | National Reference Centre for Retroviruses | Central Public Health Laboratory | Gkikas Magiorkinis et al |
| EPI_ISL_480301, EPI_ISL_480304 | National Reference Laboratory "Influenza and acute respiratory diseases" | NRL-HIV | Ivailo Alexiev; Ivan Ivanov; Ivva Philipova |
| EPI_ISL_1273394, EPI_ISL_1273395, EPI_ISL_1273396, EPI_ISL_1273399, EPI_ISL_1273406, EPI_ISL_1273408 | National Reference Laboratory - Ministry of Health Maseru Lesotho | National Institute for Communicable Diseases of the National Health Laboratory Service | Amoako DG; Banda R; Bhiman JN; Gorova V; Ismail A; Mahlangu B; Mathabo M; Mohale T; Mooko M; Ntuli N; Scheepers C |
| EPI_ISL_737200, EPI_ISL_737201 | National Reference Laboratory, Nigeria Centre for Disease Control. | National Reference Laboratory, Nigeria Centre for Disease Control, Gaduwa,Abuja, Nigeria | Akinpelu Afolabi; Anthony Ahumibe; Catherine Okoi; Celestina Obiekea; Chimaobi Chukwu; Dr Adesuyi Omoare; Dr Chikwe Ihekweazu; Dr Chinwe Ochu; Dr Ndodo Nnaemeka; Dr Sikiru Badaru; Grace Esebanmen; Kingsley Njoku; Nwando Mba; Olusola Akanbi; Shirlee Wohl; oluwaseyi Ajegbe |
| EPI_ISL_435677, EPI_ISL_962877 | National Virology Reference Laboratory | National Public Health Laboratory, National Centre for Infectious Diseases | Chavatte Jean-Marc; Cui Lin; Lin Cui; Lin Raymond Tzer Pin; Mak Tze Minn; Octavia Sophie; Raymond Tzer Pin Lin; Taib Surita; Tze Minn Mak; Zaini Zainun; Zhenyang Zhou |
| EPI_ISL_981252 | National Virus Reference Laboratory | Irish Coronavirus Sequencing Consortium - Helixworks | Conor Crosbie; Nimesh Pinnamaneni; Sachin Chalapati |
| EPI_ISL_500592, EPI_ISL_501261, EPI_ISL_512102, EPI_ISL_525416, EPI_ISL_528457, EPI_ISL_578207, EPI_ISL_671885, EPI_ISL_671920, EPI_ISL_959858, EPI_ISL_1785368, EPI_ISL_1891139, EPI_ISL_1960680, EPI_ISL_1960901, EPI_ISL_1972834, EPI_ISL_2029170, EPI_ISL_2029356, EPI_ISL_2240983, EPI_ISL_2533387, EPI_ISL_2601258, EPI_ISL_2649721, EPI_ISL_2710562, EPI_ISL_2821177, EPI_ISL_2840952, EPI_ISL_2841044, EPI_ISL_2862431, EPI_ISL_2893050, EPI_ISL_2893164, EPI_ISL_2932096, EPI_ISL_3011232, EPI_ISL_3011525, EPI_ISL_3030768 | | | |
| see above | National Virus Reference Laboratory | National Virus Reference Laboratory | Aditi Chaturvedi; Calum Walsh; Charlene Bennet; Charlene Bennett; Cillian F De Gascun; Daniel Hare; Fiona Crispie; Gabriel Gonzalez; Guerrino Macori; Jonathan Dean; Matthew McCabe; Michael Carr; Paul Cotter; Seamus Fanning; Suzie Coughlan; Zoe Yandle |
| EPI_ISL_1627088 | National reference Laboratory, NCDC, Gaduwa, Abuja, Nigeria | National reference Laboratory, NCDC, Gaduwa, Abuja, Nigeria | Akinpelu Afolabi; Anthony Ahumibe; Badaru Sikiru; Catherine Okoi; Celestina Obiekea; Chimaobi Chukwu; Dr Chikwe Ihekweazu; Dr Grace Esebanmen; Dr Ndodo Nnaemeka; Dr Omoare Adesuyi; Kingsley Njoku; Nwando Mba; Olusola Anuoluwapo Akanbi |
| EPI_ISL_2533832, EPI_ISL_2533868, EPI_ISL_2533869, EPI_ISL_2533870, EPI_ISL_2533871, EPI_ISL_2533872, EPI_ISL_2533873, EPI_ISL_2533874, EPI_ISL_2533875, EPI_ISL_2533876, EPI_ISL_2533882, EPI_ISL_2858619, EPI_ISL_2858622, EPI_ISL_2858631, EPI_ISL_2858678, EPI_ISL_2858680, EPI_ISL_2858685, EPI_ISL_2860831, EPI_ISL_2860889 | | | |
| see above | Naval Medical Research Unit No. 3 | Naval Medical Research Center Biological Defense Research Directorate | Andrea E. Luquette; Andrew J. Bennett; Bishwo N. Adhikari; Catherine E. Arnold; Chaselynn M. Watters; Emily K. Stefanov; Francisco Malagon; Kyle A. Long; Logan J. Voegtly; Luis A. Estrella; Michael V. Deschenes; Regina Z. Cer; Stephen M. Eggan; and Kimberly A. Bishop-Lilly |
| EPI_ISL_2754090, EPI_ISL_2967911 | Nebraska Public Health Laboratory | NPHL COVID-19 Response Team | NPHL COVID-19 Response Team |
| EPI_ISL_754068, EPI_ISL_754070 | Nepal Korea Friendship Municipality Hospital | Nepal Health Research Council | Meghnath Dhimal; Pradip Gyanwali |
| EPI_ISL_515391, EPI_ISL_1483251, EPI_ISL_1483264, EPI_ISL_3021067 | Nevada State Public Health Laboratory | Nevada State Public Health Laboratory | Andrew Gorzalski; Chris Laverdure; Cyprian Rossetto; David Jackson; Heather Kerwin; Joel R. Sevinsky; Mark Pandori; Natalie Crawford; Paul Hartley; Richard Tillett; Stephanie Van Hooser; Subhash C. Verma; and Mark Pandori |
| EPI_ISL_2202898 | New Mexico Department of Health Scientific Laboratory Division | Los Alamos National Laboratory Bioscience Division | Alina Deshpande; Cheryl Gleasner; Chien-Chi Lo; Joseph Hicks; Kim McMurry; Michael Edwards; Migun Shakya; Patrick Chain; Twila Kunde |
| EPI_ISL_1660402, EPI_ISL_1660408, EPI_ISL_1660411, EPI_ISL_1697281, EPI_ISL_1756023, EPI_ISL_2107449, EPI_ISL_2107451, EPI_ISL_2107456, EPI_ISL_2107457, EPI_ISL_2107535, EPI_ISL_2811816 | | | |
| see above | New South Wales Health Pathology Royal Prince Alfred Hospital | Microbiology RPAH | Au, J.; Bull, R.; Deveson, I.; Foster, C.; Rawlinson, W.; Ruiz Silva, M.; Van Hal, S. |
| EPI_ISL_1196008 | Nhlangano Health Centre | National Institute for Communicable Diseases of the National Health Laboratory Service | Amoako DG; Bhiman JN; Ismail A; Mahlangu B; Maphalala GP; Mohale T; Ntuli N; Scheepers C |
| EPI_ISL_516934 | Nicolae Testemitanu State University of Medicine and Pharmacy | International Centre for Genetic Engineering and Biotechnology (ICGEB) and ARGO Open Lab Platform for Genome Sequencing | Dal Monego S; Licastro D; Marcello A; Rajasekharan S; Ulinici M |
| EPI_ISL_1073639, EPI_ISL_1073640, EPI_ISL_1381746 | Nigeria Centre For Disease Control | National reference Laboratory, NCDC, Gaduwa, Abuja | Anthony Ahumibe; Catherine Okoi; Celestina Obiekea; Chimaobi Chukwu; Dr Chikwe Ihekweazu; Dr Ndodo Nnaemeka; Dr Omoare Adesuyi; Grace Esebanmen; Naidoo Dhamari; Nwando Mba; Olusola Anuoluwapo Akanbi |
| EPI_ISL_2565890, EPI_ISL_2565894 | Nigeria Centre for Disease Control (NCDC) | Africa Centre for Excellence for Genomics of Infectious Diseases (ACEGID), Redeemer's University | A.T.; Abechi; Ajogbasile; Akano; C.A.; C.T.; Eromon; F.V.; Folarin, O.; Happi; I.B.; J.N.; J.U.; K.O.; Kayode; Nosamiefan, I.; Oguzie; Olawoye; Olumade; Oluniyi; P.E.; P.S.; T.J.; Ugwu; Uwanibe |
| EPI_ISL_1970548 | Nigeria Centre for Disease Control (NCDC) | African Centre for Excellence for Genomics of Infectious Diseases (ACEGID), Redeemer's University | A.T.; Abechi; Ajogbasile; Akano; C.A.; C.T.; Eromon; F.V.; Folarin, O.; Happi; I.B.; J.N.; J.U.; K.O.; Kayode; Oguzie; Olawoye; Olumade; Oluniyi; P.E.; P.S.; T.J.; Ugwu; Uwanibe |
| EPI_ISL_872604, EPI_ISL_872608, EPI_ISL_872610, EPI_ISL_906285, EPI_ISL_906297, EPI_ISL_941276, EPI_ISL_941277, EPI_ISL_941279, EPI_ISL_941285, EPI_ISL_1242022, EPI_ISL_1242023, EPI_ISL_1242025 | | | |
| see above | Nigeria Centre for Disease Control (NCDC) | African Centre for Excellence for Genomics of Infectious Diseases (ACEGID), Redeemer's University | Oluniyi P.E. et al |
| EPI_ISL_455362, EPI_ISL_455412, EPI_ISL_487092, EPI_ISL_487098, EPI_ISL_487101, EPI_ISL_527872, EPI_ISL_527879, EPI_ISL_527881, EPI_ISL_527882, EPI_ISL_527884, EPI_ISL_527887, EPI_ISL_527895, EPI_ISL_527909, EPI_ISL_527914, EPI_ISL_729921, EPI_ISL_729922, EPI_ISL_729924, EPI_ISL_729925, EPI_ISL_729926, EPI_ISL_729929, EPI_ISL_729946, EPI_ISL_729951, EPI_ISL_729962, EPI_ISL_729968, EPI_ISL_729979, EPI_ISL_729982, EPI_ISL_729985, EPI_ISL_730004, EPI_ISL_730016, EPI_ISL_730023, EPI_ISL_730031, EPI_ISL_730043 | | | |
| see above | Nigeria Centre for Disease Control (NCDC) | African Centre for Excellence for Genomics of Infectious Diseases (ACEGID), Redeemer's University, Ede, Osun State, Nigeria | Ajogbasile F.V.; Folarin O.A.; Happi C.T.; Ihekweazu C.; Kayode A.; Oguzie J.; Olawoye I.; Olumade T.; Oluniyi P.E.; Oluniyi P.E. et al; Uwanibe J. |
| EPI_ISL_2240754, | Nigeria Centre for Disease Control (NCDC) | African Centre of Excellence for Genomics of Infectious | A.T.; Abechi; Ajogbasile; Akano; C.A.; C.T.; Eromon; F.V.; Folarin, O.; Happi; I.B.; J.N.; J.U.; K.O.; Kayode; Nosamiefan, I.; Oguzie; Olawoye; Olumade; Oluniyi; P.E.; P.S.; T.J.; Ugwu; Uwanibe |

| | | | |
|---|---|---|---|
| EPI_ISL_2285324 | | Diseases (ACEGID), Redeemer's University | |
| EPI_ISL_1093445, EPI_ISL_1093475 | Nigerian Centre for Disease Control (NCDC) | African Centre of Excellence for Genomics of Infectious Diseases (ACEGID), Redeemer's University | I.B.; Olawoye; et al |
| EPI_ISL_2230650 | Noblis | Noblis | June Kang; Katharine Jennings; Lauren Brinkac; Lee F. Kolakowski; Shane Mitchell; Shiela Diepold; William M. Nelson |
| EPI_ISL_2645140 | NordLab Kokkola | Expert Microbiology, National Institute for Health and Welfare | Carita Savolainen-Kopra; Erika Lindh; Haider al-Hello; Jani Halkilahti; Kirsi Liitsola; Niina Ikonen; Olli Vapalahti; Pekka Ellonen; Phuoc Truong; Päivi Laurila; Ravi Kant; Sari Hannula; Soile Blomqvist; Teemu Smura |
| EPI_ISL_2507749, EPI_ISL_2507872, EPI_ISL_2507873, EPI_ISL_2507875, EPI_ISL_2507876, EPI_ISL_2645062, EPI_ISL_2645166, EPI_ISL_3020668 | | | |
| see above | NordLab Oulu | Expert Microbiology, National Institute for Health and Welfare | Carita Savolainen-Kopra; Erika Lindh; Haider al-Hello; Jani Halkilahti; Kirsi Liitsola; Niina Ikonen; Olli Vapalahti; Pekka Ellonen; Phuoc Truong; Päivi Laurila; Ravi Kant; Sari Hannula; Soile Blomqvist; Teemu Smura |
| EPI_ISL_1599388, EPI_ISL_2333076, EPI_ISL_2509363, EPI_ISL_2725101 | Nordland Hospital - Bodo, Laboratory Department, Molecular Biology Unit | Norwegian Institute of Public Health, Department of Virology | 'Kathrine Stene-Johansen; Atiya R Ali; Debech Nadia; Engebretsen Serina Beate; Garcia Llorente Ignacio; Hilde Elshaug; Hilde Vollan; Jon Bråte; Kamilla Heddeland Instefjord; Karoline Bragstad; Kathrine Stene-Johansen; Line Victoria Moen; Marie Paulsen Madsen; Olav Hungnes; Pedersen Benedikte Nevjen; Rasmus Riis Kopperud |
| EPI_ISL_534243 | Norra Alvsborgs lanssjukhus | The Public Health Agency of Sweden | Anna Risberg; Anna-Malin Linde; Karin Tegmark-Wisell; Maria Lind Karlberg; Mattias Haukland; Mia Brytting; Olov Svartstrom; Oskar Karlsson Lindsjo; Petra Edquist; Reza Advani; Sandra Broddesson |
| EPI_ISL_2931369 | North Lantau Hospital | Hong Kong Department of Health | Alan K.L. Tsang; Edman T.K. Lam; Ken H.L. Ng; Peter C.W. Yip; Rickjason C.W. Chan |
| EPI_ISL_456396, EPI_ISL_548060, EPI_ISL_579406, EPI_ISL_579407 | North Shore Hospital | Institute of Environmental Science and Research (ESR) | Anja Werno; Antje van der Linden; Arlo Upton; Chris Mansell; David Hammer; Dragana Drinkovic; Erasmus Smit; Gary McAuliffe; Hana Sofia Andersson; Hermes Perez; James Ussher; Jill Sherwood; Jing Wang; Joep de Ligt; Josh Freeman; Julia Howard; Juliet Elvy; Lauren Jelly; Mary DeAlmeida; Matt Blakiston; Matt Storey; Matthew Rogers; Max Bloomfield; Michael Addidle; Michelle Balm; Muhammad Faisal; Nikki Freed; Olin Silander; Sally Roberts; Sarah Jefferies; Sharmini Muttaiyah; Susan Morpeth; Susan Taylor; Timothy Blackmore; Vani Sathyendran; Veronica Playle; Virginia Hope; Xiaoyun Ren |
| EPI_ISL_936968 | Northwestern Memorial Hospital | Ozer Lab | Chad J. Achenbach; Egon A. Ozer; Judd F. Hultquist; Lacy M. Simons; Lawrence J. Jennings; Michael G. Ison; Ramon Lorenzo-Redondo |
| EPI_ISL_2788600 | Norwegian Institute of Public Health, Department of Virology | Norwegian Institute of Public Health, Department of Virology | Atiya R Ali; Debech Nadia; Engebretsen Serina Beate; Garcia Llorente Ignacio; Hilde Elshaug; Hilde Vollan; Jon Bråte; Kamilla Heddeland Instefjord; Karoline Bragstad; Kathrine Stene-Johansen; Line Victoria Moen; Marie Paulsen Madsen; Olav Hungnes; Pedersen Benedikte Nevjen; Rasmus Riis Kopperud |
| EPI_ISL_735444, EPI_ISL_735448 | Nucleic Acid Testing - Rwanda National Reference Laboratory | GIGA Medical Genomics | Bouchra Boujemla; Esperence Umumararungu; Jacob Souopgui; Keith Durkin; Léon Mutesa; Maria Artesi; Marie-Pierre Hayette; Patrick Tuyisenge; Robert Rutayisire; Sabin Nsanzimana; Swaibu Gatare; Sébastien Bontems; Vincent Bours; Yvan Butera |
| EPI_ISL_925896, EPI_ISL_925907, EPI_ISL_925910, EPI_ISL_930634, EPI_ISL_960228, EPI_ISL_960237, EPI_ISL_960284, EPI_ISL_960300, EPI_ISL_960301, EPI_ISL_960302, EPI_ISL_1301736, EPI_ISL_1301778, EPI_ISL_1302681, EPI_ISL_2362498, EPI_ISL_2362515, EPI_ISL_2362523, EPI_ISL_2362524, EPI_ISL_2362525, EPI_ISL_2362526, EPI_ISL_2365914, EPI_ISL_2521981, EPI_ISL_2521982, EPI_ISL_2521987, EPI_ISL_2521988, EPI_ISL_2521989, EPI_ISL_2521992, EPI_ISL_2521993, EPI_ISL_2521994, EPI_ISL_2521995, EPI_ISL_2521996, EPI_ISL_2521997, EPI_ISL_2522092, EPI_ISL_2522096, EPI_ISL_2811717, EPI_ISL_2811720, EPI_ISL_2811724, EPI_ISL_2820242, EPI_ISL_2828472, EPI_ISL_2828474, EPI_ISL_2828475, EPI_ISL_2828477, EPI_ISL_2828478, EPI_ISL_2828481, EPI_ISL_2828483, EPI_ISL_2828484, EPI_ISL_2828486, EPI_ISL_2828488, EPI_ISL_2828489, EPI_ISL_2828490, EPI_ISL_2828491, EPI_ISL_2828492, EPI_ISL_2828493, EPI_ISL_2828494, EPI_ISL_2828495, EPI_ISL_2828496, EPI_ISL_2828497, EPI_ISL_2828503, EPI_ISL_2828506, EPI_ISL_2828508, EPI_ISL_2828509, EPI_ISL_2828510, EPI_ISL_2828511, EPI_ISL_2828512, EPI_ISL_2828514, EPI_ISL_2828515, EPI_ISL_2863937, EPI_ISL_2863942, EPI_ISL_3012179, EPI_ISL_3012181, EPI_ISL_3012182, EPI_ISL_3012184, EPI_ISL_3012189, EPI_ISL_3012190, EPI_ISL_3012191, EPI_ISL_3012192, EPI_ISL_3012193, EPI_ISL_3012194, EPI_ISL_3012198, EPI_ISL_3012199, EPI_ISL_3012200, EPI_ISL_3012201, EPI_ISL_3012203, EPI_ISL_3012204, EPI_ISL_3012206, EPI_ISL_3012209, EPI_ISL_3012212, EPI_ISL_3012217, EPI_ISL_3012219, EPI_ISL_3012220, EPI_ISL_3012221, EPI_ISL_3012222, EPI_ISL_3012223, EPI_ISL_3012224, EPI_ISL_3012225, EPI_ISL_3012227, EPI_ISL_3012228, EPI_ISL_3012230, EPI_ISL_3012232, EPI_ISL_3012233, EPI_ISL_3012234, EPI_ISL_3021476, EPI_ISL_3021477, EPI_ISL_3021478 | | | |
| see above | Nucleic Acid Testing, National Reference Laboratory | GIGA Medical Genomics | Bouchra Boujemla; Corinne Fasquelle; Esperence Umumararungu; Jacob Souopgui; Keith Durkin; Léon Mutesa; Maria Artesi; Marie-Pierre Hayette; Nathalie Renotte; Patrick Tuyisenge; Reuben Sindayiheba; Robert Rutayisire; Sabin Nsanzimana; Swaibu Gatare; Sébastien Bontems; Vincent Bours; Yvan Butera |
| EPI_ISL_2779341, EPI_ISL_2779342 | Nyamira County Referral Hopital | USAMRD-A, Basic Science Laboratory | Alan Lemtudo; Beth Mutai; Brian Andika; Carol Kifude; Clement Masakwe; Eric Muthanje; Esther Omuseni; Faith Sigei; Gathii Kimita; George Awinda; John Waitumbi; Josphat Nyataya; Rachel Githii; Rehema Liyai; Stephen Ochola |
| EPI_ISL_2372331, EPI_ISL_2372338, EPI_ISL_2372377, EPI_ISL_2372402 | Nyangabwe Hospital HIV Reference Laboratory | Botswana Harvard HIV Reference Laboratory | Boitumelo Zuze; Botshelo Radibe; Dorcas Maruapula; Joseph Makhema; Keoratile Ntshambiwa; Kgomotso Moruisi; Legodile Kooepile; Madisa Mine; Mosepele Mosepele; Ontlametse T. Bareng; Pamela Smith-Lawrence; Roger Shapiro; Shahin Lockman; Sikhulile Moyo; Simani Gaseitsiwe; Thongbotho Mphoyakgosi; Wonderful T. Choga |
| EPI_ISL_667345 | OHSU Lab Services Molecular Microbiology Lab | Oregon SARS-CoV-2 Genome Sequencing Center | Alec J. Hirsch; Andrew C. Adey; Benjamin N. Bimber; Brendan L. O'Connell; Brian J. O'Roak; Daniel N. Streblow; Donna Hansel; Guang Fan; Ruth V. Nichols; Sally Grindstaff; William B. Messer |
| EPI_ISL_2545564 | OKMI FN Brno | CEITEC MU | Bystry V.; Deissova T.; Dolejska M.; Machackova T.; Pardy F.; Zdrazilova Dubska L. |
| EPI_ISL_1366742, EPI_ISL_2372266 | OUCRU | OUCRU | Guy Thwaites; Huynh Trung Trieu; Lam Minh Yen; Le Manh Hung; Le Nguyen Truc Nhu; Le Thi Thu Huong; Le Van Tan; Nghiem My Ngoc; Ngo Ngoc Quang Minh; Nguyen Thanh Dung; Nguyen Thanh Phong; Nguyen Thanh Truong; Nguyen Thi Han Ny; Nguyen Thi Thu Hong; Nguyen Thi thu Hong; Nguyen To Anh; Nguyen Tri Dung; Nguyen Van Vinh Chau; Tran Nguyen Hoang Tu; Tran Tan Thanh |
| EPI_ISL_2406461, EPI_ISL_2501032 | Oddar Meanchey Rapid Response Team | Virology Unit, Institut Pasteur du Cambodge | Cecile Troupin; Chau Darapheak; Chin Savuth; Erik A Karlsson; Jurre Y Siegers; Kraing Sidonn; Leakhena Pum; Ly Sovann; Veasna Duong; Yi Sengdoeurn |
| EPI_ISL_1138869 | Office of Diseases Prevention and Control Region 4 Saraburi | COVID-19 Network Investigations (CONI) Alliance | Angkana Huang; Anthony R. Jones; Arporn Wangwiwatsin; Bhakbhoom Panthan; Chonticha Klungtong; Duangkamon Loesbanluechai; Ekawat Pasomsub; Elizabeth Batty; Insee Sensorn; Janjira Thaipadungpanit; Jutikul Kaewmalakul; Khajohn Joonlasak; Kingkan Rakmanee; Krittikorn Kumpornsin; Namfon Kotanan; Nathamon Runnachot; Pakjira Rimdusit; Payon Pengyo; Praima Moonmuang; Sataporn Hatsadichart; Sirinapa Singthong; Siriporn Lakesukthom; Siriwan Yaemnimnual; Stefan Fernandez; Suttiruk Changchawai; Thanat Chookajorn; Theerarat Kochakarn; Treewat Watthanachockchai; Wasun Chantratita; Wonvimol Lemprasert; Wudtichai Manasatienkij |
| EPI_ISL_2588639 | Ohio Department of Health Laboratory | Ohio Department of Health Laboratory | Allison Black; Brent Lee; Caitlin McDonnell; Eric Brandt; Erica Leasure; Glen McGillivary; Heather Blankenship; Holmes; Jade Mowery; Jennifer; Kelsey Florek; Keoni Omura; Kirtana Ramadugu; Quanta Brown; Stephanie Mccracken; Tyler Payne; and Tammy Bannerman |
| EPI_ISL_2547342, EPI_ISL_2547351, EPI_ISL_2547353 | Okiep CHC | National Health Laboratory Service/University of Cape Town (NHLS/UCT) | Arash Iranzadeh; Bruna Galvao; Carolyn Williamson; Deelan Doolabh; Diana Hardie; Gert Marais; Innocent Mudau; Lynn Tyers; Marvin Hsiao; Stephen Korsman |
| EPI_ISL_492016 | Oman-NIC | Department of Microbiology and Immunology-SQUH | Abdulla Balkhair; Ahlam Al-Amri; Aisha Al-Amri; Aisha Al-Busaidi; Amina Al Jardani; Fahad Zadjali; Fatma BaAlawi; Hamida AL Barwani; Hanan Al-kindi; Intisar Al-Shukri; Khulood Al-Mammary; Mohammed Al-Tobi; Samiha Al Kharusi; Samira Al-Maruqi; Zeyana AL-Dahmani |
| EPI_ISL_457701, EPI_ISL_457706 | Oman-NIC | Oman-NIC | Abdulla Balkhair; Ahlam Al-Amri; Aisha Al-Amri; Aisha Al-Busaidi; Amina Al Jardani; Fahad Zadjali; Fatma BaAlawi; Hamida AL Barwani; Hanan Al-kindi; Intisar Al-Shukri; Khulood Al-Mammary; Mohammed Al-Tobi; Samiha Al Kharusi; Samira Al-Maruqi; Zeyana AL-Dahmani |
| EPI_ISL_2921171, EPI_ISL_2921174, EPI_ISL_2921181, EPI_ISL_2921182, EPI_ISL_2921186, EPI_ISL_2921187, EPI_ISL_2921188, EPI_ISL_2921200, EPI_ISL_2921201, EPI_ISL_2925607, EPI_ISL_2925610, EPI_ISL_2925612, EPI_ISL_2925622, EPI_ISL_2925623 | | | |
| see above | Oman-NIC | Oman-National Influenza Center | Aisha Al-Busaidi; Amina Al Jardani; Bilal Hussain.; Fahad Al Balushi; Hanan Al Kindi; Intisar Al-Shukri; Intisar Al-Shukri.; Laila Al Balushi; Samiha Al Kharusi |
| EPI_ISL_1795050 | Oman-NIC | Oman-National Influenza Center-Department of Microbiology and Immunology-SQUH | Abdulla Balkhair; Ahlam Al-Amri; Aisha Al-Amri; Aisha Al-Busaidi; Amina Al Jardani; Fahad Zadjali; Fatma BaAlawi; Hamida AL Barwani; Hanan Al-kindi; Intisar Al-Shukri; Khulood Al-Mammary; Mohammed Al-Tobi; Samiha Al Kharusi; Samira Al-Maruqi; Zeyana AL-Dahmani |
| EPI_ISL_1819076 | Oman-NIC | Oman-National Influenza Center-Department of Microbiology and Immunology-SQUH | Abdulla Balkhair; Ahlam Al-Amri; Aisha Al-Amri; Aisha Al-Busaidi; Amina Al Jardani; Fahad Zadjali; Fatma BaAlawi; Hamida AL Barwani; Hanan Al-kindi; Intisar Al-Shukri; Khulood Al-Mammary; Mohammed Al-Tobi; Samiha Al Kharusi; Samira Al-Maruqi; Zeyana AL-Dahmani |
| EPI_ISL_525429, EPI_ISL_1532282, EPI_ISL_2692996 | Oman-National Influenza Center | Biotechnology & OMICs Laboratory | ; Abdul Latif Khan; Adil Al-Wahaibi; Adil Khan; Ahmed Al Harrasi; Ahmed Al-Harrasi; Ahmed Al-Rawahi; Ahmed N Al-Rawahi; Aisha Al-Amri; Amal Al-Maani; Amina Al-Jardani; Amina Al-Jardani.; Bilal Hussain; Hanan Al-Kindi; Intisar Al-Shukri; Sajjad Asaf; Samiha Al-Kharusi; Samira Al-Mahruqi; Samiya Al-Zadjali; Saqib Bilal; Seif Al-Abri |
| EPI_ISL_518845, EPI_ISL_518848 | Oman-National Influenza Center | Biotechnology & OMICs Laboratory, Natural & Medical Sciences Research Center, University of Nizwa | Abdul Latif Khan; Adil Al-Wahaibi; Adil Khan; Ahlam Al-Amri; Ahmed Al-Harrasi; Ahmed Al-Rawahi; Aisha Al-Amri; Aisha Al-Busaidi; Amina Al-Jardani; Hanan Al-Kindi; Intisar Al-Shukri; Sajjad Asaf; Samiha Al-Kharusi; Samira Al-Mahruqi; Seif Al-Abri; Seif Al-Abri. |
| EPI_ISL_2820625 | Omega Diagnostics at Mounes | Omega Diagnostics at Mounes | Cynthia Corley; Latira Haynes-Jacob; Maria Echeverria; Vivek Khare MD |
| EPI_ISL_2405157 | Omics Sciences Laboratory | Omics Sciences Laboratory | Darlyn Amaya; Derly Andrade Molina; Gabriel Morey León; Juan Carlos Fernández Cadena; Rubén Armas González |
| EPI_ISL_569843 | Omsk Research Institute of Natural Focal Infections | WHO National Influenza Centre Russian Federation | Aleksei Vasilenko; Andrey Komissarov; Artem Fadeev; Daria Nashatyreva; Ekaterina Gradoboeva; Ekaterina Savkina; Elena Poleshchuk; Valery Yakimenko |
| EPI_ISL_1739670, EPI_ISL_1739728, EPI_ISL_1739844, EPI_ISL_1739932, EPI_ISL_1739979, EPI_ISL_1740041, EPI_ISL_1740066, EPI_ISL_1740069, EPI_ISL_1740095, EPI_ISL_1740341, EPI_ISL_1740361, EPI_ISL_1740375, EPI_ISL_1740387, EPI_ISL_1740392, EPI_ISL_1740396, EPI_ISL_1740428, EPI_ISL_1740446 | | | |
| see above | Ontario's COVID-19 Genomics Rapid Response Coalition | McMaster University | Ahmed Draia; Allison McGeer; Andrew G. McArthur; Angel Li; Emily Panousis; Hooman Derakhshani; Jalees Nasir; Kuganya Nirmalarajah; Michael Surette; Patryk Aftanas; Samira Mubareka |
| EPI_ISL_1546496, EPI_ISL_1546997, EPI_ISL_1979289 | Originating lab: Wales Specialist Virology Centre Sequencing lab: Pathogen Genomics Unit | Public Health Wales Microbiology Cardiff Wales Specialist Virology Centre | Alec Birchley; Alexander Adams; Amy Gaskin; Angela Marchbank; Bree Gatica-Wilcox; Catherine Moore; Jason Coombes; Joanne Watkins; Joel Southgate; Johnathan Evans; Laura Gifford; Lauren Gilbert; Lee Graham; Malorie Perry; Matthew Bull; Nicole Pacchiarini; Sally Corden; Sara Kumziene-Summerhayes; Sara Rey; Sarah Taylor; Simon Cottrell; Sophie Jones; Tom Connor |
| EPI_ISL_417484, EPI_ISL_2491426 | Oslo University Hospital, Department of Medical Microbiology | Norwegian Institute of Public Health, Department of Virology | Atiya R Ali; Debech Nadia; Engebretsen Serina Beate; Garcia Llorente Ignacio; Hilde Elshaug; Hilde Vollan; Jon Bråte; Kamilla Heddeland Instefjord; Karoline Bragstad; Kathrine Stene-Johansen; Line Victoria Moen; Marie Paulsen Madsen; Olav Hungnes; Pedersen Benedikte Nevjen; Rasmus Riis Kopperud |
| EPI_ISL_1820369, EPI_ISL_1820375, EPI_ISL_2225889, EPI_ISL_2426075, EPI_ISL_2491425, EPI_ISL_2925773 | Ostfold Hospital Trust - Kalnes, Centre for Laboratory Medicine, Section for gene technology and infection serology | Norwegian Institute of Public Health, Department of Virology | Atiya R Ali; Debech Nadia; Engebretsen Serina Beate; Garcia Llorente Ignacio; Hilde Elshaug; Hilde Vollan; Jon Bråte; Kamilla Heddeland Instefjord; Karoline Bragstad; Kathrine Stene-Johansen; Line Victoria Moen; Marie Paulsen Madsen; Olav Hungnes; Pedersen Benedikte Nevjen; Rasmus Riis Kopperud |
| EPI_ISL_3031381, EPI_ISL_3031391, EPI_ISL_3031396, EPI_ISL_3031399 | Othaya Sub-County Hospital | USAMRD-A, Basic Science Laboratory | Alan Lemtudo; Beth Mutai; Brian Andika; Carol Kifude; Clement Masakwe; Eric Muthanje; Esther Omuseni; Faith Sigei; Gathii Kimita; George Awinda; John Waitumbi; Josphat Nyataya; Rachel Githii; Rehema Liyai; Stephen Ochola |
| EPI_ISL_1534378 | Oudtshoorn Hospital wc OUD | NHLS/UCT | Arash Iranzadeh; Bruna Galvao; Carolyn Williamson; Deelan Doolabh; Diana Hardie; Emmanuel SJ; Innocent Mudau; Kruger Marais; Lynn Tyers; Marvin Hsiao; Stephen Korsman; Tegally H; de Oliveira T |
| EPI_ISL_1273214 | Oxford University Clinical Research Unit (OUCRU) | Oxford University Clinical Research Unit (OUCRU) | Guy Thwaites; Huynh Trung Trieu; Lam Minh Yen; Le Manh Hung; Le Nguyen Truc Nhu; Le Van Tan; Nghiem My Ngoc; Ngo Ngoc Quang Minh; Nguyen Thanh Dung; Nguyen Thanh Phong; Nguyen Thanh Truong; Nguyen Thi Thu Hong; Nguyen To Anh; Nguyen Tri Dung; Nguyen Van Vinh Chau; Tran Nguyen Hoang Tu; Tran Tan Thanh; le Thi Thu Huong |
| EPI_ISL_3078887 | Oxford Viromics, NDM, University of Oxford; Oxford University Hospitals; Basingstoke and North Hampshire Hospital | COVID-19 Genomics UK (COG-UK) Consortium | Alex Mobbs; Amy Trebes; Anita Justice; Catrin Moore; Christophe Fraser; David Bonsall; David Buck; Emma Wise; George Macintyre; Jessica Lynch; John Todd; Mariateresa de Cesare; Matilde Mori; Monique Andersson; Nathan Moore; Nick Cortes; Robert Shaw; Stephen Kidd; Tanya Golubchik; Timothy Peto |
| EPI_ISL_2086976 | PATH CARE | National Institute for Communicable Diseases of the National | Amoako DG; Bhiman JN; Ismail A; Mahlangu B; Mohale T; Ntuli N; Scheepers C |

| | | | |
|---|---|---|---|
| | | Health Laboratory Service | |
| EPI_ISL_2695543, EPI_ISL_2709853, EPI_ISL_2709854, EPI_ISL_2709856 | POLOKWANE MANKWENG HOSPITAL | National Institute for Communicable Diseases of the National Health Laboratory Service | Amoako DG; Bhiman JN; Everatt J; Ismail A; Mahlangu B; Mnguni A; Mohale T; Ntuli N; Scheepers C |
| EPI_ISL_2984863 | PORT ELIZABETH LABOARTORY | National Institute for Communicable Diseases of the National Health Laboratory Service | Amoako DG; Bhiman JN; Everatt J; Ismail A; Mahlangu B; Mnguni A; Mohale T; Ntuli N; Scheepers C |
| EPI_ISL_2827902, EPI_ISL_2827903, EPI_ISL_2895717, EPI_ISL_2895720, EPI_ISL_2895726, EPI_ISL_2895727, EPI_ISL_2895735, EPI_ISL_2895741, EPI_ISL_2895744, EPI_ISL_2895748 | | | |
| see above | PORT ELIZABETH LABORATORY | National Institute for Communicable Diseases of the National Health Laboratory Service | Amoako DG; Bhiman JN; Everatt J; Ismail A; Mahlangu B; Mnguni A; Mohale T; Ntuli N; Scheepers C |
| EPI_ISL_2984922, EPI_ISL_2984939 | POTCHEFSTROOM | National Institute for Communicable Diseases of the National Health Laboratory Service | Amoako DG; Bhiman JN; Everatt J; Ismail A; Mahlangu B; Mnguni A; Mohale T; Ntuli N; Scheepers C |
| EPI_ISL_2384025 | PR Public Health Lab | Centers for Disease Control and Prevention Division of Viral Diseases, Pathogen Discovery | Alison Laufer Halpin; Ben L. Rambo-Martin; Clinton R. Paden; Dakota Howard; Darlene Wagner; Dave Wentworth; Dhwani Batra; Jasmine Padilla; Justin Lee; Katie Dillon; Krista Queen; Kristen Knipe; Kristine Lacek; Mark Burroughs; Matthew Schmerer; Mili Sheth; Peter Cook; Sam Shepard; Sarah Nobles; Shoshona Le; Suxiang Tong; Vivien Dugan; Yvette Unoarumhi |
| EPI_ISL_2493252 | PRONTO ATENDIMENTO DE VARGEM GRANDE PAULISTA | Instituto Butantan | Antonio Jorge Martins; Claudia Renata dos Santos Barros; David Schlesinger; Debora Botequio Moretti; Dimas Tadeu Covas; Elaine Cristina Marqueze; Elaine Vieira Santos; Evandra Strazza Rodrigues; Heidge Fukumasu; Jayme Augusto de Souza-Neto; José Salvatore Leister Patané; Luiz Alcantara; Luiz Lehmann Coutinho; Maria Carolina Elias; Maurício Lacerda Nogueira; Rafael dos Santos Bezerra; Raul Machado Neto; Rejane Maria Tommasini Grotto; Ricardo Haddad; Sandra Coccuzzo Sampaio Vessoni; Simone Kashima; Svetoslav Nanev Slavov; Vincent Louis Viala |
| EPI_ISL_2779535 | PSTC | USAMRD-A, Basic Science Laboratory | Alan Lemtudo; Beth Mutai; Brian Andika; Carol Kifude; Clement Masakwe; Eric Muthanje; Esther Omuseni; Faith Sigei; Gathii Kimita; George Awinda; John Waitumbi; Josphat Nyataya; Rachel Githii; Rehema Liyai; Stephen Ochola |
| EPI_ISL_2876986 | Pacaltsdorp Clinic wc PAC | NHLS/UCT | Arash Iranzadeh; Bruna Galvao; Carolyn Williamson; Deelan Doolabh; Diana Hardie; Gert Marais; Innocent Mudau; Lynn Tyers; Marvin Hsiao; Stephen Korsman |
| EPI_ISL_2241616, EPI_ISL_2241618, EPI_ISL_2241619, EPI_ISL_2504093, EPI_ISL_2504094, EPI_ISL_2504099, EPI_ISL_2504100, EPI_ISL_2504101, EPI_ISL_2504102, EPI_ISL_2504103, EPI_ISL_2504104, EPI_ISL_2504105, EPI_ISL_2504106, EPI_ISL_2504107, EPI_ISL_2504108, EPI_ISL_2504109, EPI_ISL_2504110, EPI_ISL_2504112, EPI_ISL_2504114, EPI_ISL_2504115, EPI_ISL_2504116, EPI_ISL_2504117, EPI_ISL_2509333, EPI_ISL_2509334 | | | |
| see above | Palapye Primary Hospital Laboratory | Botswana Harvard HIV Reference Laboratory | Boitumelo Zuze; Botshelo Radibe; David Lawrence; Dorcas Maruapula; Joseph Makhema; Keoratile Ntshambiwa; Kwana Lechiile; Legodile Kooepile; Letsibogo Gaoraelwe; Madisa Mine; Modisa Motswaledi; Mosepele Mosepele; Ontlametse T. Bareng; Roger Shapiro; Shahin Lockman; Sikhulile Moyo; Sikhulile Wonderful T. Choga; Simani Gaseitsiwe; Thela Tefelo; Thongbotho Mphoyakgosi; Wonderful T. Choga |
| EPI_ISL_596509, EPI_ISL_596529, EPI_ISL_596530, EPI_ISL_596532, EPI_ISL_596544, EPI_ISL_596561 | Palestinian Ministry of Health | Molecular Genetics Lab | Damien Richard; Dana Najjar; Francois Balloux; Hisham Darwish; Husam Sallam; Issa Shtayeh; Lucy van Dorp; Mahmoud Ruzayqat; Nouar Qutob; Osama Najjar; Zaidoun Salah |
| EPI_ISL_1098330, EPI_ISL_1173095, EPI_ISL_1543418, EPI_ISL_1543814, EPI_ISL_2633269, EPI_ISL_2634471, EPI_ISL_2634792, EPI_ISL_2716022, EPI_ISL_2923861 | | | |
| see above | Pandemic Response Lab - NYC | Pandemic Response Lab, R&D | Cybill del Castillo; Dylan Law; Haiping Hao; Henry Lee; Isabel Fernandez Escapa; Jon Laurent; Katherine Nelson; Melissa Hopkins; Michael Hammerling; Pradeep Bugga; Shinyoung Clair Kang; Sol Rey; William Ward |
| EPI_ISL_2876996 | Parkwood Clinic wc PRK | NHLS/UCT | Arash Iranzadeh; Bruna Galvao; Carolyn Williamson; Deelan Doolabh; Diana Hardie; Gert Marais; Innocent Mudau; Lynn Tyers; Marvin Hsiao; Stephen Korsman |
| EPI_ISL_2171131 | Pasig City Children's Hospital - Child's Hope | Philippine Genome Center | Alethea R. de Guzman; Anna Ong-Lim; Arianne A. Zamora; Asia Louisa U. Chong; Benedict A. Maralit; Candice Francheska B. Tambaoan; Carlo M. Lapid; Celia Carlos; Devon Ray Pacial; Edsel Maurice Salvaña; El King D. Morado; Elcid Aaron R. Pangilinan; Eva Maria Cutiongco-de la Paz; Francis A. Tablizo; Irish Coleen A. Asin; Jaime C. Montoya; Jan Michael C. Yap; Jo-Hannah S. Llames; John Q. Wong; Joshua Gregor A. Dizon; Juan Antonio R. Magalang; Karol Sophia Agape R. Padilla; Kenneth M. Kim; Kris P. Punayan; Marc Edsel C. Ayes; Marc Jerrone R. Castro; Maria Rosario Singh-Vergeire and Cynthia P. Saloma; Maria Sofia L. Yangzon; Marissa Alejandria; Razel Nikka M. Hao; Rianna Patricia S. Cruz; Sheila Mae M. Araiza |
| EPI_ISL_2035941, EPI_ISL_2035946, EPI_ISL_2035947, EPI_ISL_2035949, EPI_ISL_2035988, EPI_ISL_2036077 | Pasteur Institute - Laboratory of Clinical Virology | Pasteur Institute - Laboratory of Clinical Virology | Anissa Chouikha; Henda Triki; Kais Ghedira; Mariem Gdoura; Sondos Haddad; Wasfi Fares |
| EPI_ISL_2694596, EPI_ISL_2695826 | PathCare, Cape Town | Division of Medical Virology, National Health Laboratory Service (NHLS), Tygerberg Hospital / Stellenbosch University | Bronwyn Kleinhans; Gert van Zyl; Houriiyah Tegally; Jean Maritz; Petra Raimond; San Emmanual James; San Emmanuel James; Susan Engelbrecht; Tania Stander; Tongai Maponga; Tulio de Oliveira; Wolfgang Preiser |
| EPI_ISL_456163, EPI_ISL_456215, EPI_ISL_456301, EPI_ISL_622806, EPI_ISL_755626 | PathLab Bay of Plenty | Institute of Environmental Science and Research (ESR) | Anja Werno; Antje van der Linden; Arlo Upton; Chris Mansell; David Hammer; Dragana Drinkovic; Erasmus Smit; Gary McAuliffe; Hana Sofia Andersson; Hermes Perez; James Ussher; Jill Sherwood; Jing Wang; Joep de Ligt; Josh Freeman; Julia Howard; Juliet Elvy; Lauren Jelly; Mary DeAlmeida; Matt Blakiston; Matt Storey; Matthew Rogers; Max Bloomfield; Michael Addidle; Michelle Balm; Muhammad Faisal; Nikki Freed; Olin Silander; Sally Roberts; Sarah Jefferies; Sharmini Muttaiyah; Susan Morpeth; Susan Taylor; Timothy Blackmore; Vani Sathyendran; Veronica Playle; Virginia Hope; Xiaoyun Ren |
| EPI_ISL_420456, EPI_ISL_470851, EPI_ISL_2713075, EPI_ISL_2758809, EPI_ISL_2932263 | PathWest Laboratory Medicine WA | PathWest Laboratory Medicine WA | Avram Levy; Chisha Sikazwe; David Smith and David Speers; Jurissa Lang; PathWest Laboratory Medicine WA Microbial Surveillance Unit |
| EPI_ISL_596691, EPI_ISL_596741, EPI_ISL_596743, EPI_ISL_596754, EPI_ISL_596770, EPI_ISL_596777, EPI_ISL_596837, EPI_ISL_605854, EPI_ISL_672628, EPI_ISL_672640, EPI_ISL_672646, EPI_ISL_708758, EPI_ISL_708794, EPI_ISL_794672, EPI_ISL_794686, EPI_ISL_794710, EPI_ISL_810967, EPI_ISL_1017678, EPI_ISL_1069391, EPI_ISL_1069392, EPI_ISL_1069393, EPI_ISL_1198802, EPI_ISL_1295933, EPI_ISL_1448427, EPI_ISL_1508992, EPI_ISL_1508993, EPI_ISL_1508994, EPI_ISL_1633676, EPI_ISL_1633681, EPI_ISL_1669117, EPI_ISL_1669120, EPI_ISL_1669123, EPI_ISL_1669125, EPI_ISL_1750965, EPI_ISL_1816917, EPI_ISL_1817558, EPI_ISL_1914666, EPI_ISL_1914667, EPI_ISL_1914668, EPI_ISL_1972908, EPI_ISL_1972909, EPI_ISL_1972911, EPI_ISL_1972912, EPI_ISL_2226229, EPI_ISL_2304306, EPI_ISL_2322898, EPI_ISL_2598297, EPI_ISL_2757655, EPI_ISL_2757656, EPI_ISL_2757657, EPI_ISL_2784344, EPI_ISL_2841828, EPI_ISL_2841829, EPI_ISL_2841830, EPI_ISL_2841831, EPI_ISL_2861270, EPI_ISL_2942965, EPI_ISL_2982303, EPI_ISL_3038899 | | | |
| see above | PathWest Laboratory Medicine WA | PathWest Laboratory Medicine WA Microbial Surveillance Unit | PathWest Laboratory Medicine WA Microbial Surveillance Unit |
| EPI_ISL_2691707 | PathWest Laboratory Medicine WA Microbial Surveillance Unit | PathWest Laboratory Medicine WA Microbial Surveillance Unit | Hospital Ave; Nedlands WA 6009; QEII Medical Centre |
| EPI_ISL_591403, EPI_ISL_591414, EPI_ISL_591426, EPI_ISL_667570, EPI_ISL_667586, EPI_ISL_667621, EPI_ISL_721585, EPI_ISL_779210, EPI_ISL_779237, EPI_ISL_803936, EPI_ISL_825399, EPI_ISL_833371 | | | |
| see above | Pathogen Genomics Center, National Institute of Infectious Diseases | Pathogen Genomics Center, National Institute of Infectious Diseases | Kentaro Itokawa; Makoto Kuroda; Masanori Hashino; Rina Tanaka; Tsuyoshi Sekizuka |
| EPI_ISL_437463, EPI_ISL_437467, EPI_ISL_512962, EPI_ISL_513023, EPI_ISL_513152, EPI_ISL_678037, EPI_ISL_678151, EPI_ISL_678165 | | | |
| see above | Pathogen Genomics Lab King Abdullah University of Science and Technology(KAUST) | Pathogen Genomics Lab King Abdullah University of Science and Technology(KAUST) | Abbas Al Mutair; Abdulaziz Alahmadi; Afrah Alsomali; Amanda; Amanda Ooi; Amit Kumar Subudhi; Anwar Hashem; Arnab Pain; Asim Khogeer; Awad Al-Omari; Fadwa Alofi; Fathia Ben Rached; Jumana Taha; Kahled Algithami; Luke; Luke Esau; Naif Almontashiri; Nashwa Al-khotani; Olga Douvropoulou; Raeece Naeem; Rahul P Salunke; Raushan Nugmanova; Samer Salih; Sara Mfarrej; Sharif Hala |
| EPI_ISL_593656 | Pathology North - Hunter - NSW Health Pathology | NSW Health Pathology - Institute of Clinical Pathology and Medical Research; Westmead Hospital; University of Sydney | CIDM-PH et al. |
| EPI_ISL_407894, EPI_ISL_407896, EPI_ISL_410717 | Pathology Queensland | Public Health Virology Laboratory | Alyssa Pyke; Amanda De Jong; Andrew Van Den Hurk; Ben Huang; Carmel Taylor; David Warrilow; Doris Genge; Elisabeth Gamez; Glen Hewitson; Ian Maxwell Mackay; Inga Sultana; Jamie McMahon; Jean Barcelon; Judy Northill; Mitchell Finger; Natalie Simpson; Neelima Nair; Peter Burtonclay; Peter Moore; Sarah Wheatley; Sean Moody; Sonja Hall-Mendelin; Timothy Gardam; and Frederick Moore. |
| EPI_ISL_490036 | Pathology West - NSW Health Pathology | NSW Health Pathology - Institute of Clinical Pathology and Medical Research; Westmead Hospital; University of Sydney | CIDM-PH et al. |
| EPI_ISL_2876956 | Pelican Park CDC wc PAX | NHLS/UCT | Arash Iranzadeh; Bruna Galvao; Carolyn Williamson; Deelan Doolabh; Diana Hardie; Gert Marais; Innocent Mudau; Lynn Tyers; Marvin Hsiao; Stephen Korsman |
| EPI_ISL_2859337 | Philippine General Hospital (PGH) | Philippine Genome Center | Alethea R. de Guzman; Anna Ong-Lim; Arianne A. Zamora; Benedict A. Maralit; Carlo M. Lapid; Celia Carlos; Devon Ray Pacial; Diomedes A. Carino; Edsel Maurice Salvaña; El King D. Morado; Elcid Aaron R. Pangilinan; Eva Maria Cutiongco-de la Paz; Francis A. Tablizo; Henrietta Marie Rodriguez; Jaime C. Montoya; Jan Michael C. Yap; Jarvin E. Nipales; Jo-Hannah S. Llames; John Q. Wong; Joshua Gregor A. Dizon; Juan Antonio R. Magalang; Karol Sophia Agape R. Padilla; Kenneth M. Kim; Kris P. Punayan; Krisitna Patriz Dela Cruz; Lindsay Claire D.L. Carandang; Ma. Exanil Plantig; Marc Edsel C. Ayes; Maria Rosario Singh-Vergeire and Cynthia P. Saloma; Maria Sofia L. Yangzon; Marielle M Gamboa; Marissa Alejandria; Nina Francesca Bustamante; Razel Nikka M. Hao; Renato Jacinto Q. Mantaring; Rianna Patricia S. Cruz; Sheila Mae M. Araiza; Yvonne Valerie Austria; Zipporah Mariebelle R. Enriquez; Zyrel V. Mollejon |
| EPI_ISL_2155237, EPI_ISL_2859654 | Philippine Red Cross - Port Area | Philippine Genome Center | Alethea R. de Guzman; Anna Ong-Lim; Arianne A. Zamora; Asia Louisa U. Chong; Benedict A. Maralit; Candice Francheska B. Tambaoan; Carlo M. Lapid; Celia Carlos; Devon Ray Pacial; Diomedes A. Carino; Edsel Maurice Salvaña; El King D. Morado; Elcid Aaron R. Pangilinan; Eva Maria Cutiongco-de la Paz; Francis A. Tablizo; Henrietta Marie Rodriguez; Irish Coleen A. Asin; Jaime C. Montoya; Jan Michael C. Yap; Jarvin E. Nipales; Jo-Hannah S. Llames; John Q. Wong; Joshua Gregor A. Dizon; Juan Antonio R. Magalang; Karol Sophia Agape R. Padilla; Kenneth M. Kim; Kris P. Punayan; Krisitna Patriz Dela Cruz; Lindsay Claire D.L. Carandang; Ma. Exanil Plantig; Marc Edsel C. Ayes; Marc Jerrone R. Castro; Maria Rosario Singh-Vergeire and Cynthia P. Saloma; Maria Sofia L. Yangzon; Marielle M Gamboa; Marissa Alejandria; Nina Francesca Bustamante; Razel Nikka M. Hao; Renato Jacinto Q. Mantaring; Rianna Patricia S. Cruz; Sheila Mae M. Araiza; Yvonne Valerie Austria; Zipporah Mariebelle R. Enriquez; Zyrel V. Mollejon |
| EPI_ISL_1164597 | Plateforme de testing Namuroise | Plateforme de testing Namuroise | Degosserie Jonathan; Denis Olivier; Mullier François; Otto Gaetan |
| EPI_ISL_2483324, EPI_ISL_2714810, EPI_ISL_2988339 | Platform BIS UZA/UAntwerpen | Labo Klinische Biologie, UZA | Basil Britto Xavier; Christine Lammens; Herman Goossens; Ines Verbesselt; Jasmine Coppens; Kathleen Holemans; Marie Le Mercier; Veerle Matheeussen |
| EPI_ISL_1120735 | Platform BIS UZA/UAntwerpen | UAntwerp, Laboratory of Medical Microbiology | Basil Britto Xavier; Christine Lammens; Herman Goossens; Jasmine Coppens; Marie Le Mercier; Veerle Matheeussen |
| EPI_ISL_2550759 | Port Dickson Hospital | Institute for Medical Research, Infectious Disease Research Centre, National Institutes of Health, Ministry of Health Malaysia | Azizan MA; Kamel K; Mohd Zawawi Z; Ramly N; Robert F; Suppiah J; Thayan R |
| EPI_ISL_2447801, EPI_ISL_2447809 | Port Elizabeth Laboratory | National Institute for Communicable Diseases of the National Health Laboratory Service | Amoako DG; Bhiman JN; Ismail A; Mahlangu B; Mohale T; Ntuli N; Scheepers C |
| EPI_ISL_712075 | Port Elizabeth Provincial Hospital, National Health Laboratory Services, Eastern Cape, South Africa | National Institute for Communicable Diseases of the National Health Laboratory Service | Allam M; Bhiman JN; Ismail A; Mahlangu B; Mohale T; Ntuli N |
| EPI_ISL_2779427, EPI_ISL_2779428, EPI_ISL_3031388, EPI_ISL_3031389, EPI_ISL_3031390, EPI_ISL_3031419, EPI_ISL_3031420 | | | |

| | | | |
|---|---|---|---|
| see above | Port Health Clinic, Busia border point | USAMRD-A, Basic Science Laboratory | Alan Lemtudo; Beth Mutai; Brian Andika; Carol Kifude; Clement Masakwe; Eric Muthanje; Esther Omuseni; Faith Sigei; Gathii Kimita; George Awinda; John Waitumbi; Josphat Nyataya; Rachel Githii; Rehema Liyai; Stephen Ochola |
| EPI_ISL_2779496, EPI_ISL_2779500, EPI_ISL_2779507 | Port Health Clinic, Malaba border point | USAMRD-A, Basic Science Laboratory | Alan Lemtudo; Beth Mutai; Brian Andika; Carol Kifude; Clement Masakwe; Eric Muthanje; Esther Omuseni; Faith Sigei; Gathii Kimita; George Awinda; John Waitumbi; Josphat Nyataya; Rachel Githii; Rehema Liyai; Stephen Ochola |
| EPI_ISL_529013 | Presidio Ospedaliero "S.Filippo e Nicola"-Avezzano | Istituto Zooprofilattico Sperimentale dell'Abruzzo e Molise "G.Caporale" | Ancora M; Cammà C; Curini V; Di Domenico M; Di Pasquale A; Lorusso A; Mangone I; Marcacci M; Puglia I; Rinaldi A; Savini G. |
| EPI_ISL_1819200 | Presidio di Brindisi Di Summa - Perrino | Istituto Zooprofilattico Sperimentale della Puglia e della Basilicata | Bianco A.; Capozzi L.; Del Sambro L.; Difato L.; Giannico A.; Parisi A.; Ridolfi D.; Santoro A.; Simone D. |
| EPI_ISL_2790410 | Primary Health Care Celinac | Public Health Institute of Republic of Srpska | Branka Culibrk; Dijana Vukajlovic; Milica Celic; Pava Dimitrijevic; Stanka Tomic; Tatjana Markovic; Zeljka Sumic |
| EPI_ISL_2790402, EPI_ISL_2790404, EPI_ISL_2790406 | Primary Health Care Laktasi | Public Health Institute of Republic of Srpska | Branka Culibrk; Dijana Vukajlovic; Milica Celic; Pava Dimitrijevic; Stanka Tomic; Tatjana Markovic; Zeljka Sumic |
| EPI_ISL_2790405 | Primary Health Care Prijedor | Public Health Institute of Republic of Srpska | Branka Culibrk; Dijana Vukajlovic; Milica Celic; Pava Dimitrijevic; Stanka Tomic; Tatjana Markovic; Zeljka Sumic |
| EPI_ISL_417064 | Prince of Wales Hospital | Hong Kong Department of Health | Alan K.L. Tsang; Dominic N.C. Tsang; Edman T.K. Lam; Peter C.W. Yip; Rickjason C.W. Chan |
| EPI_ISL_1402425, EPI_ISL_1402426 | Private clinic of Biogen Med, Tashkent, Uzbekistan | Center of Genomics and bioinformatics, Bioinformatics laboratory | Abdurakhmon N Yusupov; Ibrokhim Y Abdurakhmonov.; Mirzakamol S Ayubov; Mukhammadjon H Mirzakhmedov; Shukhrat E Shermatov; Zabardast T Buriev |
| EPI_ISL_1092071 | Protzer Lab | Protzer Lab, Laboratory for Functional Genome Analysis, Dept. Genomics, Gene Center of the LMU Munich | Alexander Graf; Dieter Hoffmann; Elisabeth Esser; Eva C.Schulte; Helmut Blum; Stefan Krebs; Till Bunse; Ulrike Protzer |
| EPI_ISL_1909922 | Provincial Public Health Reference Laboratory | Provincial Public Health Reference Laboratory | ANDLEEB HANIF; Asim Altaf; Hassan Nawaz; Mariam Javed |
| EPI_ISL_2543563, EPI_ISL_2543615, EPI_ISL_2544508 | Provincial Public Health Reference Laboratory (PPHRL) P&SHD | Provincial Public Health Reference Laboratory (PPHRL) P&SHD | ANDLEEB HANIF |
| EPI_ISL_1180679 | Public Health Authority of the Slovak Republic | Bergthaler laboratory, CeMM Research Center for Molecular Medicine of the Austrian Academy of Sciences | Andreas Bergthaler; Anna Schedl; Bekir Erguner; Benedikt Agerer; Christoph Bock; Fabian Amman; Jan Laine; Lukas Endler; Maelle Le Moing; Martin Senekowitsch; Michael Schuster; Thomas Penz |
| EPI_ISL_1647661, EPI_ISL_1647749, EPI_ISL_1749617, EPI_ISL_1749618, EPI_ISL_2448839, EPI_ISL_2598620, EPI_ISL_2657889, EPI_ISL_2657919, EPI_ISL_2657920, EPI_ISL_2688039 | | | |
| see above | Public Health Authority of the Slovak Republic | Laboratory of Genomics and Bioinformatics, Comenius University Science Park | Anna Gičová; Diana Rusňáková; Jakub Styk; Jaroslav Budiš; Miroslav Böhmer; Tatiana Sedláčková; Tomáš Szemes |
| EPI_ISL_2379751 | Public Health Authority of the Slovak Republic | Public Health Authority of the Slovak Republic | Anna Gičová; Barbora Kotvasová; Elena Tichá; Lucia Ševčíková; Miroslav Böhmer; Pavol Mišenko; Terézia Vrabľová; Tomáš Szemes |
| EPI_ISL_2351129 | Public Health Center 52 Samsennok | Division of Genomic Medicine and Innovation support,Department of Medical Sciences, Ministry of Public Health, Thailand | Archawin Rojanawiwat; Jirapha Pakdee; Natthakul Bunneang; Nuanjun Wichukchinda; Penpitcha Thawong; Pilailuk Akkapaiboon Okada; Pundharika Piboonsiri; Surakameth Mahasirimongkol; Waritta Sawaengdee |
| EPI_ISL_1112327, EPI_ISL_1112329, EPI_ISL_1112753 | Public Health Center of Ukraine | Charité Universitätsmedizin Berlin, Institute of Virology | Barbara Mühlemann; Christian Drosten; Ihor Kuzin; Iryna Demchyshyna; Julia Schneider; Jörn Beheim-Schwarzbach; Liudmyla Chernenko; Roman Rodyna; Talitha Veith; Terry Jones; Victor M Corman |
| EPI_ISL_636973 | Public Health Lab | Public Health Lab | Alwasti; H |
| EPI_ISL_775080, EPI_ISL_775157, EPI_ISL_854650, EPI_ISL_854721, EPI_ISL_875752, EPI_ISL_875764, EPI_ISL_875765, EPI_ISL_875768, EPI_ISL_875771, EPI_ISL_875848, EPI_ISL_875863, EPI_ISL_875864, EPI_ISL_925718, EPI_ISL_925769, EPI_ISL_925781, EPI_ISL_1112618, EPI_ISL_1112619, EPI_ISL_1112631, EPI_ISL_1112803, EPI_ISL_1112836, EPI_ISL_1170498, EPI_ISL_1170552, EPI_ISL_1170615, EPI_ISL_1170627, EPI_ISL_1170659, EPI_ISL_1170706, EPI_ISL_1170769, EPI_ISL_1170787, EPI_ISL_1170801, EPI_ISL_1170838, EPI_ISL_1170842, EPI_ISL_1183470, EPI_ISL_1183583, EPI_ISL_1230694, EPI_ISL_1230736, EPI_ISL_1230755, EPI_ISL_1230800, EPI_ISL_1270899, EPI_ISL_1270994, EPI_ISL_1271039, EPI_ISL_1271347, EPI_ISL_1271359, EPI_ISL_1271393, EPI_ISL_1271396, EPI_ISL_1271455, EPI_ISL_1271458, EPI_ISL_1271532, EPI_ISL_1271545, EPI_ISL_1271550, EPI_ISL_1271649, EPI_ISL_1271754, EPI_ISL_1271774, EPI_ISL_1271827, EPI_ISL_1271914, EPI_ISL_1335164, EPI_ISL_1368590, EPI_ISL_1478357, EPI_ISL_1478379, EPI_ISL_1478747, EPI_ISL_1559221, EPI_ISL_1559226, EPI_ISL_1559230, EPI_ISL_1559294, EPI_ISL_1559308, EPI_ISL_1742848, EPI_ISL_1743549, EPI_ISL_1896723, EPI_ISL_2011880, EPI_ISL_2011881, EPI_ISL_2012922, EPI_ISL_2141305, EPI_ISL_2141308, EPI_ISL_2141368, EPI_ISL_2141369, EPI_ISL_2141377, EPI_ISL_2250400, EPI_ISL_2250575, EPI_ISL_2251679, EPI_ISL_2252430, EPI_ISL_2364408, EPI_ISL_2452463, EPI_ISL_2922328, EPI_ISL_2922543 | | | |
| see above | Public Health Ontario Laboratory | Public Health Ontario Laboratory | Aimin Li; Alireza Eshaghi; Andre Villegas; Ashleigh Sullivan; Christine Frantz; Dean Maxwell; Esha Joshi; Heather Rilkoff; Jared Simpson; Jennifer L Guthrie; Jonathan B Gubbay; Karthikeyan Sivaraman; Lawrence Heisler; Matthew Watson; Michael CY Li; Michael Laszloffy; Nahuel Fittipaldi; Philip Banh; Richard de Borja; Samir N Patel; Sandeep Nagra; Sandra Zittermann; Sarah Teatero; Vanessa G Allen; Yao Chen; Yogi Sundaravadanam |
| EPI_ISL_2860636, EPI_ISL_2860638, EPI_ISL_2860640, EPI_ISL_2860642, EPI_ISL_2860643, EPI_ISL_2860644, EPI_ISL_2860645, EPI_ISL_2860646, EPI_ISL_2860647 | | | |
| see above | Public Health Reference Laboratory | Erasmus Medical Center | Anne van der Linden; Annemiek van der Eijk; Bas Oude Munnink; Corine GeurtsvanKessel; David Nieuwenhuijse; Emmanuelle Munger; Irina Chestakova; Marion Koopmans; Marjan Boter; Omar Elahmer-Abdulla Bashein- Rahma Algheriani-Ahlam Alarif; Reina Sikkema; Richard Molenkamp; on behalf of the Dutch national COVID-19 response team. |
| EPI_ISL_1910858 | Public Health Virology-Forensic and Scientific Services | Public Health Virology-Forensic and Scientific Services | Son Nguyen |
| EPI_ISL_1495211 | Public Health Authority of the Slovak Republic | Bergthaler laboratory, CeMM Research Center for Molecular Medicine of the Austrian Academy of Sciences | Andreas Bergthaler; Anna Schedl; Bekir Erguner; Benedikt Agerer; Christoph Bock; Fabian Amman; Jan Laine; Lukas Endler; Maelle Le Moing; Martin Senekowitsch; Michael Schuster; Petr Triska; Thomas Penz |
| EPI_ISL_2801898 | Pursat Rapid Response Team | Virology Unit, Institut Pasteur du Cambodge | Cecile Troupin; Chau Darapheak; Chin Savuth; Erik A Karlsson; Jurre Y Siegers; Kraing Sidonn; Leakhena Pum; Ly Sovann; Veasna Duong; Yi Sengdoeurn |
| EPI_ISL_1055477, EPI_ISL_1055478, EPI_ISL_1055481, EPI_ISL_1055483, EPI_ISL_1055485, EPI_ISL_1055486, EPI_ISL_1055487, EPI_ISL_1055488, EPI_ISL_1055489, EPI_ISL_1055490, EPI_ISL_1055505, EPI_ISL_1055506, EPI_ISL_1055513, EPI_ISL_1055535, EPI_ISL_1055602, EPI_ISL_1055688 | | | |
| see above | QEII Health Sciences Centre | National Microbiology Laboratory (NML) | Anna Majer; Anneliese Landgraff; CanCOGeN's metadata curation team; Dan Gaston; Darian Hole; Elsie Grudeski; Gary Van Domselaar; Grace Seo; Janice Pettipas; Jason LeBlanc; Jennifer Tanner; Kirsten Biggar; Madison Chapel; Morag Graham; Natalie Knox; Nathalie Bastien; Philip Mabon; Public Health Agency of Canada CanCOGeN team; Rhiannon Huzarewich; Russell Mandes; Shari Tyson; Timothy Booth; Todd Hatchette; Yan Li |
| EPI_ISL_493545, EPI_ISL_493546, EPI_ISL_2519670 | Quadram Institute Bioscience | COVID-19 Genomics UK (COG-UK) Consortium | Alexander J Trotter; Alison E. Mather; Alp Aydin; Ana P. Tedim; Anastasia Kolyva; Andrew Bell; Andrew J. Page; Claire Stuart; Dave J. Baker; Gemma L. Kay; John Wain; Justin O'Grady; Leonardo de Oliveira Martins; Lizzie Meadows; Maria Diaz; Mark Webber; Muhammed Yasir; Nabil-Fareed Alikhan; Ngozi Elumogo; Nicholas M. Thomson; Rachael Stanley; Rachel Gilroy; Reenesh Prakash; Samir Dervisevic; Samuel Bloomfield; Steven Rudder; Thanh Le-Viet |
| EPI_ISL_2860172 | Qualimed Hospital Sta. Rosa | Philippine Genome Center | Alethea R. de Guzman; Anna Ong-Lim; Arianne A. Zamora; Benedict A. Maralit; Carlo M. Lapid; Celia Carlos; Devon Ray Pacial; Diomedes A. Carino; Edsel Maurice Salvaña; El King D. Morado; Elcid Aaron R. Pangilinan; Eva Maria Cutiongco-de la Paz; Francis A. Tablizo; Henrietta Marie Rodriguez; Jaime C. Montoya; Jan Michael C. Yap; Jarvin E. Nipales; Jo-Hannah S. Llames; John Q. Wong; Joshua Gregor A. Dizon; Juan Antonio R. Magalang; Karol Sophia Agape R. Padilla; Kenneth M. Kim; Kris P. Punayan; Krisitna Patriz Dela Cruz; Lindsay Claire D.L. Carandang; Ma. Exanil Plantig; Marc Edsel C. Ayes; Maria Rosario Singh-Vergeire and Cynthia P. Saloma; Maria Sofia L. Yangzon; Marielle M Gamboa; Marissa Alejandria; Nina Francesca Bustamante; Razel Nikka M. Hao; Renato Jacinto Q. Mantaring; Rianna Patricia S. Cruz; Sheila Mae M. Araiza; Yvonne Valerie Austria; Zipporah Mariebelle R. Enriquez; Zyrel V. Mollejon |
| EPI_ISL_820573, EPI_ISL_820578, EPI_ISL_820607 | Queens Medical Centre, Clinical Microbiology Department / DeepSeq Nottingham | COVID-19 Genomics UK (COG-UK) Consortium | Christopher Moore; Fei Sang; Gemma Clark; Hannah Howson-Wells; Johnny Debebe; Jonathan Ball; Joseph Chappell; Manjinder Khakh; Matthew Carlisle; Matthew Loose; Michelle M Lister; Nadine Holmes; Patrick McClure; Theocharis Tsoleridis; Vicki M Fleming; Victoria Wright; Wendy Smith |
| EPI_ISL_593616, EPI_ISL_593617, EPI_ISL_693269, EPI_ISL_693290, EPI_ISL_849686, EPI_ISL_849694, EPI_ISL_849754, EPI_ISL_1465893, EPI_ISL_1516885, EPI_ISL_1516886, EPI_ISL_1516888, EPI_ISL_1563670, EPI_ISL_1563678, EPI_ISL_1633659, EPI_ISL_1754858, EPI_ISL_1910859, EPI_ISL_1938308, EPI_ISL_2137034, EPI_ISL_2137035, EPI_ISL_2137036, EPI_ISL_2274244, EPI_ISL_2274245, EPI_ISL_2274246, EPI_ISL_2274248, EPI_ISL_2283730, EPI_ISL_2385970, EPI_ISL_2385974, EPI_ISL_2507086, EPI_ISL_2507088, EPI_ISL_2507089, EPI_ISL_2507095, EPI_ISL_2603820, EPI_ISL_2661522, EPI_ISL_2661525, EPI_ISL_2675252, EPI_ISL_2675253, EPI_ISL_2727637, EPI_ISL_2811889, EPI_ISL_2811892, EPI_ISL_2811904, EPI_ISL_2842840, EPI_ISL_2842841, EPI_ISL_2842842, EPI_ISL_2842843, EPI_ISL_2896248, EPI_ISL_2896249, EPI_ISL_2956008, EPI_ISL_2993362, EPI_ISL_2993401, EPI_ISL_2993405, EPI_ISL_2993415, EPI_ISL_2993422, EPI_ISL_2993425, EPI_ISL_2993426, EPI_ISL_2993427 | | | |
| see above | Queensland Health Forensic and Scientific Services | Queensland Health Forensic and Scientific Services | Son Nguyen; Son Nguyen et al; Son Nguyen et al. |
| EPI_ISL_530239, EPI_ISL_530279 | Queensland Health Forensic and Scientific Services, Public Health Virology | Public Health Virology Laboratory, Forensic and Scientific Services, Queensland Health | Son Nguyen et al |
| EPI_ISL_1424519 | Queensland Medical Laboratories | Victorian Infectious Diseases Reference Laboratory (VIDRL) and the Melbourne Diagnostic Unit Public Health Laboratory (MDU-PHL) | N.L.; Palou, T.; Seemann, T.; Sherry; Vaccher, S. |
| EPI_ISL_494533, EPI_ISL_571261, EPI_ISL_571975, EPI_ISL_604114 | Quest Diagnostics | Quest Diagnostics | Anderson; Anderson, B.; B.P.; D.F.; Gerasimova, A.; Grover, D.; Hua, M.; K.E.; Kagan; Lacbawan, F.; Liu Y.; Livingston; Owen, R.; R.M.; R.M. and Owen, R.; Rosenthal; S.H.; Shalhout |
| EPI_ISL_1220443, EPI_ISL_1253267, EPI_ISL_1648870, EPI_ISL_1668705, EPI_ISL_1753408, EPI_ISL_1762016, EPI_ISL_1818271, EPI_ISL_1924348, EPI_ISL_1989864, EPI_ISL_2047081, EPI_ISL_2143545, EPI_ISL_2143956, EPI_ISL_2180464, EPI_ISL_2247212, EPI_ISL_2247292, EPI_ISL_2371834, EPI_ISL_2371912, EPI_ISL_2397489, EPI_ISL_2451305, EPI_ISL_2649980, EPI_ISL_2652295, EPI_ISL_2652368, EPI_ISL_2869180, EPI_ISL_2874567, EPI_ISL_2874761, EPI_ISL_2875140, EPI_ISL_2876694, EPI_ISL_2876857, EPI_ISL_2930261 | | | |
| see above | Quest Diagnostics Incorporated | Centers for Disease Control and Prevention Division of Viral Diseases, Pathogen Discovery | A. Gerasimova; A. Perez; Adrian Paskey; B. Anderson; Ben L. Rambo-Martin; Benjamin Rambo-Martin; Christopher Gulvick; Clinton R. Paden; Dakota Howard; Darlene Wagner; Dhwani Batra; Duncan MacCannell; F. Lacbawan; I. A. Shlyakhter; Jason Caravas; K.E. Livingston; Kara Moser; L.E. Bernstein; M. Hua; Matthew Schmerer; P. Tanpaiboon; Peter W. Cook; R. M. Kagan; R. Owen; R. V. Rolando; S. H. Rosenthal; Scott Sammons; Shatavia Morrison; Suxiang Tong; Y. Liu; Yvette Unoarumhi |
| EPI_ISL_1086247, EPI_ISL_1087772, EPI_ISL_1087817 | Quest Diagnostics Incorporated | Respiratory Viruses Branch, Division of Viral Diseases, Centers for Disease Control and Prevention | A. Gerasimova; A. Perez; B. Anderson; Ben L. Rambo-Martin; Clinton R. Paden; Dakota Howard; Dhwani Batra; Duncan MacCannell; F. Lacbawan; I. A. Shlyakhter; K.E. Livingston; L.E. Bernstein; M. Hua; P. Tanpaiboon; Peter W. Cook; R. M. Kagan; R. Owen; R. V. Rolando; S. H. Rosenthal; Suxiang Tong; Y. Liu |
| EPI_ISL_1711855 | RI State Health Laboratories | Centers for Disease Control and Prevention Division of Viral Diseases, Pathogen Discovery | Alison Laufer Halpin; Ben L. Rambo-Martin; Clinton R. Paden; Dakota Howard; Darlene Wagner; Dave Wentworth; Dhwani Batra; Jasmine Padilla; Justin Lee; Katie Dillon; Krista Queen; Kristen Knipe; Kristine Lacek; Mark Burroughs; Matthew Schmerer; Mili Sheth; Peter Cook; Sam Shepard; Sarah Nobles; Shoshona Le; Suxiang Tong; Vivien Dugan; Yvette Unoarumhi |
| EPI_ISL_2531885, EPI_ISL_2531926, EPI_ISL_2531934 | RIIP | National Reference Center for Viruses of Respiratory Infections, Institut Pasteur, Paris | Angela Brisebarre; Camille Capel; Christophe Malabat; Corinne Maufrais; Etienne Simon-Lorière; Frédéric Lemoine; Louise Lefrançois; Marion Barbet; Maud Vanpeene; Méline Bizard; StéPhanie Guyomard-Rabenirina; Sylvie Behillil; Sylvie Van der Werf; Vincent Enouf |
| EPI_ISL_413581 | RIVM | Erasmus Medical Center | Anne van der Linden; Annemiek van der Eijk; Aura Timen; Bas Oude Munnink; Claudia Schapendonk; Corien Swaan; Corine GeurtsvanKessel; David Nieuwenhuijse; Irina Chestakova; Jeroen van Kampen; Jolanda Voermans; Madelief Mollers; Manon Haverkate; Marion Koopmans; Mark Pronk; Mart Stein; Pascal Lexmond; Reina Sikkema; Richard Molenkamp; Sandra Kengne Kamga Mobou; on behalf of the Dutch national COVID-19 response team. |
| EPI_ISL_2086951, EPI_ISL_2273059, EPI_ISL_2273060, EPI_ISL_2273061, EPI_ISL_2273063, EPI_ISL_2273065, EPI_ISL_2273067, EPI_ISL_2273068, EPI_ISL_2273075, EPI_ISL_2273082, EPI_ISL_2273087, EPI_ISL_2442379, EPI_ISL_2444100, EPI_ISL_2444106, EPI_ISL_2454669, EPI_ISL_2454670, EPI_ISL_2454676, EPI_ISL_2454683, EPI_ISL_2454685, EPI_ISL_2454688, EPI_ISL_2454690, EPI_ISL_2454695, EPI_ISL_2582830, EPI_ISL_2582833, EPI_ISL_2695608, EPI_ISL_2695609, EPI_ISL_2695615, EPI_ISL_2695620, EPI_ISL_2695621, EPI_ISL_2695623, EPI_ISL_2695625, EPI_ISL_2695754, EPI_ISL_2695755, EPI_ISL_2695763, EPI_ISL_2695764, EPI_ISL_2695767, EPI_ISL_2695771 | | | |
| see above | ROB FERREIRA LABORATORY | National Institute for Communicable Diseases of the National | Amoako DG; Bhiman JN; Everatt J; Ismail A; Mahlangu B; Mnguni A; Mohale T; Ntuli N; Scheepers C |

| | | | |
|---|---|---|---|
| | | Health Laboratory Service | |
| EPI_ISL_3138836 | RS Jantung Dan Pembuluh Darah Harapan Kita | National Institute of Health Research and Development | Arie Ardiansyah Nugraha; Fajar Nur Sulistiyohadi; Hana Apsari Pawestri; Hartanti Dian Ikawati; Kartika Dewi Puspa; Krisna Pangesti; Nelly Puspandari; Subangkit; Triyani Soekarso; Vivi Setiawaty |
| EPI_ISL_763064 | RS Kasih Ibu, Surakarta | Universitas Sebelas Maret (UNS); Rumah Sakit UNS (RS-UNS); National Institute of Health Research and Development, Indonesian Ministry of Health. | Afif A Ghufron; Arie A Nugraha; Betty Suryawati; Dewi Puspa; Hana A Pawestri; Hartanti Dian Ika; Hartono; Kartika; Maryani; Revi G H Novika; Vivi Setiawaty.; Yulia Sari |
| EPI_ISL_2987618 | RS Mata Cicendo Bandung | National Institute of Health Research and Development | Arie Ardiansyah Nugraha; Fajar Nur Sulistiyohadi; Hana Apsari Pawestri; Hartanti Dian Ikawati; Kartika Dewi Puspa; Krisna Pangesti; Nelly Puspandari; Subangkit; Triyani Soekarso; Vivi Setiawaty |
| EPI_ISL_574611 | RS Premier Bintaro | Eijkman Institute for Molecular Biology, Ministry of Research and Technology/National Agency for Research and Innovation | Amin Soebandrio; David H Muljono; Edison Johar; Frilasita A Yudhaputri; Herawati Sudoyo; Hidayat Trimarsanto; Iskandar A Adnan; Khin Saw Myint; Safarina G Malik; Willy Agustine |
| EPI_ISL_2709370 | RS Pusat Pertamina | Eijkman Institute for Molecular Biology, National Research and Innovation Agency | Amin Soebandrio; Edison Johar; Frilasita A Yudhaputri; Hidayat Trimarsanto; Iskandar Adnan; Khin Saw Myint; Lidwina Priliani; Lydia V. Panggalo; Muhammad Rezki Rasyak; Safarina G Malik; Sukma Oktavianthi; Willy Agustine |
| EPI_ISL_2262254, EPI_ISL_2262265 | RS Umum Kartika Pulomas | National Institute of Health Research and Development | Arie Ardiansyah Nugraha; Hana Apsari Pawestri; Hartanti Dian Ikawati; Kartika Dewi Puspa; Krisna Pangesti; Nelly Puspandari; Subangkit; Triyani Soekarso; Vivi Setiawaty |
| EPI_ISL_906051 | RSA UGM | Genetics Working Group (Pokja Genetik) Faculty of Medicine, Public Health and Nursing Universitas Gadjah Mada (FK-KMK UGM); Disease Investigation Center Wates Ministry of Agriculture Indonesia; Department of Microbiology FK-KMK UGM; Laboratorium Diagnostik Yayasan Tahija World Mosquito Program (WMP) Yogyakarta Center for Tropical Medicine FK-KMK UGM; Integrated Research center FK-KMK UGM; Department of Computer Science and Electronics FMIPA UGM | Afiahayati; Alvin Santoso Kalim; Dwi AA Nugrahaningsih; Dwiki Afandy; Edwin W. Daniwijaya; Eggi Arguni; Endah Supriyati; Gunadi; Hendra Wibawa; Kristy Iskandar; Ludhang P. Rizki; Marcellus; Mohamad S. Hakim; Nungki Anggorowati; Siswanto; Titik Nuryastuti; Tri Wibawa |
| EPI_ISL_454520 | RSE "National Center for Biotechnology" | RSE "National Center for Biotechnology" | Akbota Rakhmetova; Alexandr Shevtsov; Askar Abdaliyev; Asylulan Amirgazin; Ilyas Akhmetollayev; Ruslan Kalendar; Viktoriya Lutsay; Yerlan Ramankulov; Zabira Aushakhmetova |
| EPI_ISL_947247 | RSU Bunda Mulia | Eijkman Institute for Molecular Biology, Ministry of Research and Technology/National Agency for Research and Innovation | Amin Soebandrio; Edison Johar; Frilasita A Yudhaputri; Hidayat Trimarsanto; Iskandar Adnan; Khin Saw Myint; Lydia V. Panggalo; Safarina G Malik; Sukma Oktavianthi; Willy Agustine |
| EPI_ISL_947298 | RSU Medirossa | Eijkman Institute for Molecular Biology, Ministry of Research and Technology/National Agency for Research and Innovation | Amin Soebandrio; Edison Johar; Frilasita A Yudhaputri; Hidayat Trimarsanto; Iskandar Adnan; Khin Saw Myint; Lydia V. Panggalo; Safarina G Malik; Sukma Oktavianthi; Willy Agustine |
| EPI_ISL_560991 | RSUD Dr. Soetomo | Institute of Tropical Disease, Universitas Airlangga | Aldise M Nastri; Gatot Soegiarto; Jezzy R Dewantari; Joni Wahyuhadi; Kazufumi Shimizu; Krisnoadi Rahardjo; Laksmi Wulandari; Maria I Lusida; Mitsuhiro Nishimura; Resti Yudhawati; Retno A Setyoningrum; Rima R Prasetya; Soetjipto; Yasuko Mori; Yohko K Shimizu |
| EPI_ISL_610162, EPI_ISL_1005696 | RSUD Dr. Tjitrowardojo | Genetics Working Group (Pokja Genetik) Faculty of Medicine, Public Health and Nursing Universitas Gadjah Mada (FK-KMK UGM); Disease Investigation Center Wates Ministry of Agriculture Indonesia; Department of Microbiology FK-KMK UGM; Laboratorium Diagnostik Yayasan Tahija World Mosquito Program (WMP) Yogyakarta Center for Tropical Medicine FK-KMK UGM; Integrated Research Center FK-KMK UGM; Department of Computer Science and Electronics FMIPA UGM; Balai Besar Teknik Kesehatan Lingkungan dan Pengendalian Penyakit (BBTKLPP) Yogyakarta | Afiahayati; Alvin Santoso Kalim; Arry Setyowati; Chusni Mubarakh; Dwi AA Nugrahaningsih; Edwin W. Daniwijaya; Eggi Arguni; Endah Supriyati; Gunadi; Havid Setyawan; Hendra Wibawa; Indaryati; Irene; Kristy Iskandar; Ludhang P. Rizki; Marcellus; Mohamad S. Hakim; Nungki Anggorowati; Siswanto; Susan Simanjaya; Titik Nuryastuti; Tri Wibawa; Try Nirmala Sari; Wuryanto |
| EPI_ISL_2854750 | RSUD Dr. W.Z. Johanes kupang | National Institute of Health Research and Development | Arie Ardiansyah Nugraha; Hana Apsari Pawestri; Hartanti Dian Ikawati; Kartika Dewi Puspa; Krisna Pangesti; Nelly Puspandari; Subangkit; Triyani Soekarso; Vivi Setiawaty |
| EPI_ISL_2631472 | RSUD Karawang | Indonesian Institute of Sciences (LIPI)/National Agency of Research and Innovation (BRIN) and National Institute of Health Research and Development (NIHRD) | Andri Wardiana; Herjuno A. Nugroho; Idris; Indriawati; Isa Nuryana; Listiana Oktavia; Puspita Lisdiyanti; Sri Swasthikawati; Subangkit; Sugiyono Saputra; Syam B. Iryanto; Vivi Setiawaty; Yayuk Sri Rahayu; Yuliawati |
| EPI_ISL_632937 | RSUD Saptosari Gunung Kidul | Genetics Working Group (Pokja Genetik) Faculty of Medicine, Public Health and Nursing Universitas Gadjah Mada (FK-KMK UGM); Disease Investigation Center Wates Ministry of Agriculture Indonesia; Department of Microbiology FK-KMK UGM; Laboratorium Diagnostik Yayasan Tahija World Mosquito Program (WMP) Yogyakarta Center for Tropical Medicine FK-KMK UGM; Integrated Research Center FK-KMK UGM; Department of Computer Science and Electronics FMIPA UGM; Balai Besar Teknik Kesehatan Lingkungan dan Pengendalian Penyakit (BBTKLPP) Yogyakarta | Afiahayati; Audric Kenny Tedja; Dwi AA Nugrahaningsih; Edwin W. Daniwijaya; Eggi Arguni; Eko Darmawan; Endah Supriyati; Gunadi; Havid Setyawan; Hendra Wibawa; Indaryati; Irene; Kristy Iskandar; Ludhang P. Rizki; Marcellus; Maria Patricia Inggriani; Mohamad S. Hakim; Nungki Anggorowati; Siswanto; Titik Nuryastuti; Tri Wibawa |
| EPI_ISL_1824615 | RSUD dr. Hasri Ainun Habibie Gorontalo | National Institute of Health Research and Development | Arie Ardiansyah Nugraha; Hana Apsari Pawestri; Hartanti Dian Ikawati; Kartika Dewi Puspa; Krisna Pangesti; Nelly Puspandari; Subangkit; Vivi Setiawaty |
| EPI_ISL_2824996 | RSUP Dr. Hasan Sadikin Bandung | Genomik Solidaritas Indonesia Laboratorium | Alfin Mohammad Abdillah; Anna Christina Brazia; Annisa Muthiah Sukirman; Anuraj Shankar; Ariel Pradipta; Basti Andriyoko; Carissa Sintca Wijaya; Delita Prihatni; Dewi Kartika Turbawati; Dhahlia Agustina Cahyono; Gracia Felias Enos Korompis; Louisa Markus; Meutia Ayuputeri Kumaheri; Normastuti Adhini Tantyo; Reinhart Gabriel; Vania Gavrila Wikasa; Vivi Setyawati |
| EPI_ISL_766032 | RSUP Dr. Sardjito | Eijkman Institute for Molecular Biology, Ministry of Research and Technology/National Agency for Research and Innovation | Amin Soebandrio; David H Muljono; Edison Johar; Frilasita A Yudhaputri; Hidayat Trimarsanto; Khin Saw Myint; Safarina G Malik |
| EPI_ISL_1824606 | RSUP Sanglah | National Institute of Health Research and Development | Arie Ardiansyah Nugraha; Hana Apsari Pawestri; Hartanti Dian Ikawati; I Gede Ketut Sajinadiyasa; I Nengah Tony Rustawan; Kartika Dewi Puspa; Krisna Pangesti; Nelly Puspandari; Subangkit; Vivi Setiawaty |
| EPI_ISL_2827879, EPI_ISL_2827884, EPI_ISL_2827886, EPI_ISL_2895681, EPI_ISL_2895682, EPI_ISL_2895683, EPI_ISL_2895690, EPI_ISL_2895693 | | | |
| see above | RUSTENBURG LABORATORY | National Institute for Communicable Diseases of the National Health Laboratory Service | Amoako DG; Bhiman JN; Everatt J; Ismail A; Mahlangu B; Mnguni A; Mohale T; Ntuli N; Scheepers C |
| EPI_ISL_450508, EPI_ISL_450515 | Rafik Hariri University Hospital | Rafik Hariri University Hospital | Rita Feghali |
| EPI_ISL_2678109 | Ramathibodi Chakri Naruebodindra Hospital | COVID-19 Network Investigations (CONI) Alliance | Angkana Huang; Anthony R. Jones; Arporn Wangwiwatsin; Bhakbhoom Panthan; Chonticha Klungtong; Duangkamon Loesbanluechai; Ekawat Pasomsub; Elizabeth Batty; Insee Sensorn; Janjira Thaipadungpanit; Khajohn Joonlasak; Kingkan Rakmanee; Krittikorn Kumpornsin; Namfon Kotanan; Stefan Fernandez; Thanat Chookajorn; Theerarat Kochakarn; Treewat Watthanachockchai; Wasun Chantratita; Wudtichai Manasatienkij |
| EPI_ISL_429168, EPI_ISL_2104753, EPI_ISL_2678105, EPI_ISL_2802837, EPI_ISL_2802852, EPI_ISL_2802853, EPI_ISL_2928525 | | | |
| see above | Ramathibodi Hospital | COVID-19 Network Investigations (CONI) Alliance | Angkana Huang; Anthony R. Jones; Arporn Wangwiwatsin; Bhakbhoom Panthan; Chonticha Klungtong; Duangkamon Loesbanluechai; Ekawat Pasomsub; Elizabeth Batty; Insee Sensorn; Janjira Thaipadungpanit; Khajohn Joonlasak; Khajohn Joonsalak; Kingkan Rakmanee; Krittikorn Kumpornsin; Namfon Kotanan; Poramate Jiaranai; Stefan Fernandez; Thanat Chookajorn; Theerarat Kochakarn; Treewat Watthanachockchai; Wasun Chantratita; Wudtichai Manasatienkij |
| EPI_ISL_1275880, EPI_ISL_1275902 | Randox Laboratories | Wellcome Sanger Institute for the COVID-19 Genomics UK (COG-UK) Consortium | Cordelia Langford; David K. Jackson; Dominic Kwiatkowski; Ewan Harrison; Ian Johnston; Jeffrey Barrett; John Sillitoe on behalf of the Wellcome Sanger Institute COVID-19 Surveillance Team; Randox Laboratories and Alex Alderton; Roberto Amato; Sonia Goncalves |
| EPI_ISL_2433337 | Rattaphum Hospital | Division of Genomic Medicine and Innovation support,Department of Medical Sciences, Ministry of Public Health, Thailand | Archawin Rojanawiwat; Jirapha Pakdee; Natthakul Bunneang; Nuanjun Wichukchinda; Penpitcha Thawong; Pilailuk Akkapaiboon Okada; Pundharika Piboonsiri; Surakameth Mahasirimongkol; Waritta Sawaengdee |
| EPI_ISL_2157357 | Red - Regional de Vigilancia Genómica del COVID-19 | Laboratory of Respiratory Viruses and Measles, Oswaldo Cruz Institute, FIOCRUZ | Alice Sampaio Rocha; Ana Carolina Mendonca; Anna Carolina Paixao; Claudia Díaz; Eilisa Cavalcante Pereira; Fernando Motta; Luciana Appolinario; Marilda Siqueira on behalf of the Fiocruz COVID-19 Genomic Surveillance Network; Mitzi Castro; Paola Resende; Renata Serrano Lopes; Sandra Paola Paz; Taina Venas |
| EPI_ISL_1415406, EPI_ISL_1415407, EPI_ISL_1415412, EPI_ISL_1415413, EPI_ISL_1415419 | Reference Laboratory of the Ministry of Health | Laboratory of Respiratory Viruses and Measles, Oswaldo Cruz Institute, FIOCRUZ | Alice Sampaio Rocha; Ana Carolina Mendonca; Anna Carolina Paixao; Fernando Motta; Indira Martins; Jessica Edwards; Luciana Appolinario; Marilda Siqueira on behalf of the Fiocruz COVID-19 Genomic Surveillance Network; Paola Resende; Renata Serrano Lopes |
| EPI_ISL_2982838, EPI_ISL_2982850, EPI_ISL_2982851, EPI_ISL_2982858, EPI_ISL_2982860, EPI_ISL_2982861, EPI_ISL_2982863 | | | |
| see above | Reference Laboratory of the Ministry of Health Royal Victoria Gardens | Laboratory of Respiratory Viruses and Measles, Oswaldo Cruz Institute, FIOCRUZ | Agatha Cristinne Prudencio; Alice Sampaio Rocha; Ana Carolina Mendonca; Anna Carolina Paixao; Elisa Cavalcante Pereira; Fernando Motta; Ighor Leonardo Arantes Gomes; Indira Martins; Jessica Edwards; Luciana Appolinario; Marilda Siqueira on behalf of the Fiocruz COVID-19 Genomic Surveillance Network; Paola Resende; Renata Serrano Lopes; Taina Venas |
| EPI_ISL_2276073, EPI_ISL_2276076 | Regional Authority of Public Health Trenčín | Veterinary institute in Zvolen Slovakia | Beňová K.; Bučková L.; Dirbáková Z.; Mojžiš M.; Mokryšová S.; Reizigová L.; Sujová S.; Tinák M. |
| EPI_ISL_768617 | Regional Medical Sciences Center 5 Samut Songkhram | National Institute of Health, Department of Medical Sciences, Ministry of Public Health, Thailand | ; Natchaya Khiadsang; Pakorn Piromtong; Pilailuk Okada; Ratana Tacharoenmuang; Siripaporn Phuygun; Sittiporn Parnmen; Sunthareeya Waicharoen; Thanutsapa Thanadachakul; Warawan Wongboot; sirikanda wimol |
| EPI_ISL_2877034 | Regional Medical Sciences Center 8 Udonthani | National Institute of Health, Department of Medical Sciences, Ministry of Public Health, Thailand | ; Natchaya Khiadsang; Nuttida Thongpramul; Pakorn Piromtong; Pilailuk Okada; Ratana Tacharoenmuang; Siripaporn Phuygun; Sittiporn Parnmen; Sunthareeya Waicharoen; Thanutsapa Thanadachakul; Warawan Wongboot; sirikanda wimol |
| EPI_ISL_708806, EPI_ISL_708807 | Regional medical sciences center 6 chonburi | National Institute of Health, Department of Medical Sciences, Ministry of Public Health, Thailand | Malinee Chittaganpitch; Pakorn Piromtong; Pilailuk Okada; Siripaporn Phuygun; Sittiporn Parnmen; Sunthareeya Waicharoen; Thanutsapa Thanadachakul; Warawan Wongboot |
| EPI_ISL_2544703, EPI_ISL_2544707, | Research Center for Emerging Viral Infections, Chang Gung University, Taiwan | Research Center for Emerging Viral Infections, Chang Gung University, Taiwan | Carol Wang; Chung-Guei Huang; Hsiao-Chen Tu; Hui-Ying Weng; Hung-Yu Shu; Jason Su; Jora Lin; Kuo-Ming Lee; Po-Wei Huang; Pocky Lai; Shih-Feng Tsai; Shin-Ru Shih; Shu-Li Yang; Tsu-Lan Wu; Yu-Nong Gong; Yung-Feng Lin |

| | | | |
|---|---|---|---|
| EPI_ISL_2544708, EPI_ISL_2544710, EPI_ISL_2544711, EPI_ISL_2544713 | | | |
| EPI_ISL_2740439, EPI_ISL_2740490, EPI_ISL_2740549, EPI_ISL_2741203, EPI_ISL_2741308, EPI_ISL_2741766, EPI_ISL_2741814, EPI_ISL_2742068, EPI_ISL_2748953, EPI_ISL_2748955, EPI_ISL_2749048, EPI_ISL_2749111, EPI_ISL_2749225, EPI_ISL_2904339, EPI_ISL_2904411 | | | |
| see above | Respiratory Virus Unit, Microbiology Services Colindale, Public Health England | COVID-19 Genomics UK (COG-UK) Consortium | PHE Covid Sequencing Team |
| EPI_ISL_423103, EPI_ISL_423104, EPI_ISL_423207, EPI_ISL_464172 | Respiratory Virus Unit, Microbiology Services Colindale, Public Health England | Respiratory Virus Unit, Microbiology Services Colindale, Public Health England | Angie Lackenby; Joanna Ellis; Jonathan Hubb; Kirstin Edwards; Leena Bhaw; Maria Zambon; Monica Galiano; Omolola Akinbami; PHE Covid Sequencing Team; Richard Myers; Shahjahan Miah; Steven Platt; Tiina Talts |
| EPI_ISL_747100 | Respiratory Viruses Branch, Centers for Disease Control and Prevention | Respiratory Viruses Branch, Centers for Disease Control and Prevention | C.R.; Cook; Lee, J.; Li, Y.; Marine, R.; Montmayeur, A.; P.W.; Paden; Queen, K.; Sheth, M.; Tao, Y.; Tong, S.; Uehara, A.; Wang, H. |
| EPI_ISL_2603892 | Rhode Island State Health Laboratory | Rhode Island State Health Laboratory | Ewa King; Kristin Carpenter-Azevedo; Richard C. Huard |
| EPI_ISL_2799553, EPI_ISL_2800292 | Riga East University Hospital, National Microbiology Reference Laboratory | Riga East University Hospital, National Microbiology Reference Laboratory; Eurofins Genomics Europe Sequencing GmbH | Arzu Algulieva; Diāna Dušacka; Dārta Pūpola; Ilva Pole; Jevgenijs Bodrenko; Jūlija Čevere; Reinis Vangravs; Reinis Zeltmatis; Sergejs Ņikišins; Ģirts Šķenders |
| EPI_ISL_1239479 | Rob Ferreira Hospital, National Health Laboratory Services, Mpumalanga, South Africa | National Institute for Communicable Diseases of the National Health Laboratory Service | Amoako DG; Bhiman JN; Ismail A; Mahlangu B; Mohale T; Ntuli N; Scheepers C |
| EPI_ISL_2447836, EPI_ISL_2447870 | Rob Ferreira Laboratory | National Institute for Communicable Diseases of the National Health Laboratory Service | Amoako DG; Bhiman JN; Ismail A; Mahlangu B; Mohale T; Ntuli N; Scheepers C |
| EPI_ISL_419833, EPI_ISL_419835 | Royal Darwin Hospital | Victorian Infectious Diseases Reference Laboratory and Microbiological Diagnostic Unit Public Health Laboratory, Doherty Institute | Caly L.; Druce J.; Meumann, E.; Sait, M.; Schultz M.; Seemann T. |
| EPI_ISL_521862, EPI_ISL_779406, EPI_ISL_779408, EPI_ISL_779409, EPI_ISL_779413, EPI_ISL_812423, EPI_ISL_2321158, EPI_ISL_2321160, EPI_ISL_2321161, EPI_ISL_2321163, EPI_ISL_2321166, EPI_ISL_2379269, EPI_ISL_2839562, EPI_ISL_2839563, EPI_ISL_2839567, EPI_ISL_2839568, EPI_ISL_2907544, EPI_ISL_2907545, EPI_ISL_2907546, EPI_ISL_3030412 | | | |
| see above | Royal Darwin Hospital Pathology | MDU-PHL | Caly L.; Druce J.; M.L.; Meumann, E.; N.L.; Sait; Sait, M.; Schultz M.; Seemann T.; Sherry; Sherry, N. |
| EPI_ISL_2250194, EPI_ISL_2250196, EPI_ISL_2250198, EPI_ISL_2250204, EPI_ISL_2250206, EPI_ISL_2250209, EPI_ISL_2250214, EPI_ISL_2250216, EPI_ISL_2250217, EPI_ISL_2250219 | | | |
| see above | Royal Darwin Hospital Pathology | Microbiological Diagnostic Unit Public Health Laboratory (MDU-PHL) | Caly L.; Druce J.; M.L.; Meumann, E.; N.L.; Sait; Seemann T.; Sherry |
| EPI_ISL_426898 | Royal Darwin Hospital Pathology | Microbiological Diagnostic Unit Public Health Laboratory and Victorian Infectious Diseases Reference Laboratory, Doherty Institute | Caly L.; Druce J.; Meumann, E.; Sait, M.; Schultz M.; Seemann T.; Sherry, N. |
| EPI_ISL_430631 | Royal Darwin Hospital Pathology | Microbiological Diagnostic Unit Public Health Laboratory and Victorian Infectious Diseases Reference Laboratory, The Peter Doherty Institute for Infection and Immunity | Caly L.; Druce J.; Meumann, E.; Sait, M.; Schultz M.; Seemann T.; Sherry, N. |
| EPI_ISL_577607 | Royal Hobart Hospital | Royal Hobart Hospital | Cooley L.; van Haeften R. |
| EPI_ISL_522621, EPI_ISL_522739, EPI_ISL_522745 | Royal Hobart Hospital Microbiology Department | MDU-PHL | Cooley L.; M.B.; Sait M.; Schultz; Seemann T.; Sherry N.; van Haeften R. |
| EPI_ISL_516806 | Rumah Sakit PKU Gamping | Genetics Working Group (Pokja Genetik) Faculty of Medicine, Public Health and Nursing Universitas Gadjah Mada (FK-KMK UGM); Disease Investigation Center Wates Ministry of Agriculture Indonesia; Department of Microbiology FK-KMK UGM; Laboratorium Diagnostik Yayasan Tahija World Mosquito Program (WMP) Yogyakarta Center for Tropical Medicine FK-KMK UGM; Integrated Research center FK-KMK UGM; Department of Computer Science and Electronics FMIPA UGM | . Afiahayati; . Marcellus; . Siswanto; Ardorisye Saptaty Fornia; Dwi AA Nugrahaningsih; Edwin W. Daniwijaya; Eggi Arguni; Endah Supriyati; Gunadi; Hendra Wibawa; Kemala Athollah; Ludhang P. Rizki; Mohamad S. Hakim; Titik Nuryastuti; Tri Wibawa |
| EPI_ISL_735445 | Rwanda National Reference Laboratory | GIGA Medical Genomics | Bouchra Boujemla; Esperence Umumararungu; Jacob Souopgui; Keith Durkin; Léon Mutesa; Maria Artesi; Marie-Pierre Hayette; Patrick Tuyisenge; Robert Rutayisire; Sabin Nsanzimana; Swaibu Gatare; Sébastien Bontems; Vincent Bours; Yvan Butera |
| EPI_ISL_1063900, EPI_ISL_1064168 | Rwanda National Reference Laboratory | Rwanda National Reference Laboratory | Enatha Mukantwari; Umuringa Jeanne d'Arc |
| EPI_ISL_451127, EPI_ISL_451144, EPI_ISL_467989, EPI_ISL_468024, EPI_ISL_468039, EPI_ISL_508138, EPI_ISL_510544, EPI_ISL_513911, EPI_ISL_603124, EPI_ISL_603127, EPI_ISL_654795, EPI_ISL_654800, EPI_ISL_755568, EPI_ISL_812358, EPI_ISL_1704675, EPI_ISL_1704796, EPI_ISL_1704798, EPI_ISL_1704799, EPI_ISL_1704804, EPI_ISL_1706366, EPI_ISL_1706367 | | | |
| see above | SA Pathology | SA Pathology | Chuan Kok Lim; Geoff Higgins; Ivan Bastian; Julien Soubrier; Karin Kassahn; Lex Leong; Mark Turra; Song Gao |
| EPI_ISL_1603794 | SARS-CoV-2 Sequencing Castilla y Leon-Spain Consortium | SARS-CoV-2 Sequencing Castilla y Leon-Spain Consortium | Antonio Orduña-Domingo; Carlos Fuster Foz; Carmen Aldea-Mansilla; Carmen Gimeno Crespo; David Abad; Gregoria Megías Lobón; Jose María Eiros Bouza; Laura Sánchez de Prada; M. Isabel Fernandez-Natal; Marta Dominguez-Gil; Marta Hernandez; María Antonia García Castro; Mª Fe Brezmes-Valdivieso; Noelia Arenal Andrés; Silvia Rojo; Sonsoles Garcinuño Pérez |
| EPI_ISL_1046778, EPI_ISL_1198812, EPI_ISL_1927132, EPI_ISL_1927133, EPI_ISL_1927135, EPI_ISL_1927136, EPI_ISL_1927138, EPI_ISL_1927139, EPI_ISL_1927308, EPI_ISL_1927364, EPI_ISL_1927368, EPI_ISL_1927381, EPI_ISL_1927389, EPI_ISL_1927397, EPI_ISL_1927435, EPI_ISL_1927441, EPI_ISL_2131776, EPI_ISL_2131792, EPI_ISL_2447620, EPI_ISL_2447623, EPI_ISL_2447628, EPI_ISL_2447634, EPI_ISL_2447692, EPI_ISL_2621762, EPI_ISL_2674807, EPI_ISL_2828265, EPI_ISL_2828268, EPI_ISL_2930625, EPI_ISL_2933814 | | | |
| see above | SARS-CoV-2 testing team, National Institute of Infectious Diseases | Pathogen Genomics Center, National Institute of Infectious Diseases | Chang-Kweng Lim; Eri Nakayama; Hazuka Y Furihata; Hidemasa Izumiya; Hiromizu Takahashi; Ken Miyazawa; Ken Shimuta; Ken-ichi Lee; Kentaro Itokawa; Koichi Ishikawa; Makoto Kuroda; Masanori Hashino; Masatomo Morita; Masumichi Saito; Minoru Nagi; Naomi Nojiri; Nobuo Koizumi; Nozomu Hanaoka; Rina Tanaka; Sana Uchikoba; Sayuri Seki; Shigeru Kusagawa; Shigeru Tajima; Shouji Yamamoto; Sunao Iyoda; Takashi Sakudoh; Tsuguto Fujimoto; Tsuyoshi Sekizuka |
| EPI_ISL_2324433, EPI_ISL_2567592, EPI_ISL_2828393 | SC (UCO) Igiene e Sanità Pubblica, ASUGI, Trieste | ARGO Laboratorio Genomica ed Epigenomica | Barbone F; Braida C; Breda C; Busetti M; D'Agaro P; Dal Monego S; Degasperi M; Fontana F; Licastro D; Marcello A; Piscianz E; Segat L |
| EPI_ISL_2828450 | SC (UCO) Igiene e Sanità Pubblica, ASUGI, Trieste and SC Microbiologia e Virologia, ASFO, Pordenone | ARGO Laboratorio Genomica ed Epigenomica | Barbone F; Breda C; D'Agaro P; Dal Monego S; De Rosa R; Degasperi M; Licastro D; Marcello A; Segat L |
| EPI_ISL_424841 | SC Dept of Health and Env. Control-Bureau of Laboratories | Pathogen Discovery, Respiratory Viruses Branch, Division of Viral Diseases, Centers for Disease Control and Prevention | Alison S. Laufer Halpin; Anna Uehara; Christopher A. Elkins; Clinton R. Paden; Haibin Wang; Jing Zhang; Krista Queen; Mary S. Keckler; Rachel Marine; Suxiang Tong; Yan Li; Ying Tao |
| EPI_ISL_2482956 | SCM B12 | CNR Virus des Infections Respiratoires - France SUD | Antonin Bal; Bruno Lina; Bruno Simon; Gregory Destras; Gwendolyne Burfin; Hadrien Regue; Laurence Josset; Martine Valette; Quentin Semanas |
| EPI_ISL_2018218 | SD Public Health Laboratory | Centers for Disease Control and Prevention Division of Viral Diseases, Pathogen Discovery | Alison Laufer Halpin; Ben L. Rambo-Martin; Clinton R. Paden; Dakota Howard; Darlene Wagner; Dave Wentworth; Dhwani Batra; Jasmine Padilla; Justin Lee; Katie Dillon; Krista Queen; Kristen Knipe; Kristine Lacek; Mark Burroughs; Matthew Schmerer; Mili Sheth; Peter Cook; Sam Shepard; Sarah Nobles; Shoshona Le; Suxiang Tong; Vivien Dugan; Yvette Unoarumhi |
| EPI_ISL_2934571, EPI_ISL_2966511 | SE Kyiv CDC MHU | The Institute of Molecular Biology and Genetics of NASU | M.Tukalo et al. |
| EPI_ISL_1732275 | SECAO CENTRO DE DIAGNOSTICO SECEDI | Instituto Butantan / Mendelics | Antonio Jorge Martins; Bianca Cechetto Carlos. Mendelics: Bibiana Santos; Claudia Renata dos Santos Barros; David Schlesinger. Hemocentro Ribeirão Preto: Simone Kashima; Debora Botequio Moretti. Centro de Genômica Funcional da ESALQ: Luiz Lehmann Coutinho; Dimas Tadeu Covas; Elaine Cristina Marqueze; Elaine Vieira dos Santos; Elisangela Chicaroni Mattos; Erika Freitas; Evandra Strazza Rodrigues; Felipe Allan da Silva da Costa; Flavia Aburjaile; Guilherme Targino Valente; Heidge Fukumasu. USP-Botucatu: Rejane Maria Tommasini Grotto; Instituto Butantan: Alexander Roberto Precioso; Jayme A. Souza-Neto; Jessika Cristina Chagas Lesbon; José Salvatore Leister Patané; João Paulo Kitajima; Luiz Carlos Junior de Alcantara; Maria Carolina Elias; Marta Giovanetti; Patricia Akemi Assato; Rafael dos Santos Bezerra; Raquel de Lello Rocha Campos Cassano. NGS Soluções Genômicas: Pilar Drummond Sampaio Corrêa Mariani. FZEA-USP Pirassununga: Mirele Daiana Poleti; Raul Machado Neto; Ricardo Augusto Brassaloti; Ricardo Haddad; Rodrigo Tocantins Calado.; Sandra Coccuzzo Sampaio; Svetoslav Nanev Slavov; Vagner Fonseca; Vincent Louis Viala |
| EPI_ISL_2934570, EPI_ISL_2934573, EPI_ISL_2934574, EPI_ISL_2966512 | SECP MHU | The Institute of Molecular Biology and Genetics of NASU | M.Tukalo et al. |
| EPI_ISL_2938096 | SECRETARIA MUNICIPAL DA SAUDE DE GUARATINGUETA LABORATÓRIO 01 | Instituto Butantan | Antonio Jorge Martins; Claudia Renata dos Santos Barros; David Schlesinger; Debora Botequio Moretti; Dimas Tadeu Covas; Elaine Cristina Marqueze; Elaine Vieira Santos; Evandra Strazza Rodrigues; Heidge Fukumasu; Jayme Augusto de Souza-Neto; José Salvatore Leister Patané; Luiz Alcantara; Luiz Lehmann Coutinho; Maria Carolina Elias; Maurício Lacerda Nogueira; Rafael dos Santos Bezerra; Raul Machado Neto; Rejane Maria Tommasini Grotto; Ricardo Haddad; Sandra Coccuzzo Sampaio Vessoni; Simone Kashima; Svetoslav Nanev Slavov; Vincent Louis Viala |
| EPI_ISL_2878465 | SELAS MEDILYS | Department of Virology, Henri Mondor University Hospital, Assistance Publique Hôpitaux de Paris, Université Paris-Est Créteil, INSERM U955 | Alexandre Soulier; Christophe Rodriguez; Elisabeth Trawinski; Guillaume Gricourt; Jean-Michel Pawlotsky; Melissa N'Debi; Slim Fourati; Vanessa Demontant |
| EPI_ISL_2895385, EPI_ISL_2895396, EPI_ISL_2895404, EPI_ISL_2895415 | SESARAM | Instituto Nacional de Saude (INSA) | Borges et al |
| EPI_ISL_2600810 | SIESP CHIETI - DRIVE IN LANCIANO | Istituto Zooprofilattico Sperimentale dell'Abruzzo e Molise "G. Caporale" | Ancora M; Calistri P; Cammà C; Caporale M; Curini V; Delli Compagni E; Di Domenico M; Di Lollo Valeria; Di Pasquale A; Lorusso A; Mangone I; Marcacci M; Puglia I; Rinaldi A; Savini G; Scialabba S |
| EPI_ISL_2483577 | SIESP DIPARTIMENTO DI PREVENZIONE TERAMO | Istituto Zooprofilattico Sperimentale dell'Abruzzo e Molise "G. | Ancora M; Calistri P; Cammà C; Caporale M; Curini V; Delli Compagni E; Di Domenico M; Di Lollo Valeria; Di Pasquale A; Lorusso A; Mangone I; Marcacci M; Puglia I; Rinaldi A; Savini G; Scialabba S |

| | | | |
|---|---|---|---|
| | | Caporale” | |
| EPI_ISL_2828709 | SISP ASL BAT | University of Bari Biomedical Sciences and Human Oncology | Maria Chironna |
| EPI_ISL_2562988, EPI_ISL_2564146, EPI_ISL_2565189, EPI_ISL_2587465, EPI_ISL_2587504, EPI_ISL_2883377, EPI_ISL_2889715, EPI_ISL_2889739 | | | |
| see above | SK-Roy Romanow Provincial Laboratory | National Microbiology Laboratory (NML) | Alanna Senecal; Amanda Lang; Anna Majer; Anneliese Landgraff; CanCOGeN's metadata curation team; Darian Hole; Elsie Grudeski; Gary Van Domselaar; Grace Seo; Jennifer Tanner; Jessica Minion; Kara Loos; Keith MacKenzie; Kirsten Biggar; Madison Chapel; Meredith Faires; Morag Graham; Natalie Knox; Nathalie Bastien; Philip Mabon; Public Health Agency of Canada CanCOGeN team; Rachel DePaulo; Rhiannon Huzarewich; Russell Mandes; Ryan McDonald; Shari Tyson; Timothy Booth; Yan Li |
| EPI_ISL_1660718, EPI_ISL_1708577, EPI_ISL_2139795 | SYNLAB | GIGA Medical Genomics | Bouchra Boujemla; Cécile Meex; Keith Durkin; Maria Artesi; Marie-Pierre Hayette; Nathalie Renotte; Pierrette Melin; Raphaël Boreux; Sébastien Bontems; Vincent Bours |
| EPI_ISL_2642051, EPI_ISL_2642061, EPI_ISL_2642070, EPI_ISL_2642080, EPI_ISL_2642086, EPI_ISL_2642087, EPI_ISL_2642090, EPI_ISL_2642091, EPI_ISL_2642092, EPI_ISL_2642119, EPI_ISL_2642127, EPI_ISL_2642139, EPI_ISL_2642152, EPI_ISL_2642154, EPI_ISL_2642155, EPI_ISL_2642157, EPI_ISL_2642180, EPI_ISL_2642186, EPI_ISL_2642187, EPI_ISL_2642188, EPI_ISL_2642194, EPI_ISL_2642198, EPI_ISL_2642199, EPI_ISL_2642228, EPI_ISL_2642244, EPI_ISL_2642302, EPI_ISL_2642339, EPI_ISL_2642342, EPI_ISL_2642362, EPI_ISL_2642372, EPI_ISL_2642376, EPI_ISL_2642391, EPI_ISL_2642418, EPI_ISL_2642429, EPI_ISL_2642433, EPI_ISL_2642439, EPI_ISL_2642447, EPI_ISL_2642448, EPI_ISL_2642449, EPI_ISL_2642450, EPI_ISL_2642452, EPI_ISL_2642453, EPI_ISL_2642454, EPI_ISL_2642510, EPI_ISL_2642518, EPI_ISL_2642578, EPI_ISL_2642659, EPI_ISL_2642660, EPI_ISL_2642661, EPI_ISL_2642768, EPI_ISL_2642923, EPI_ISL_2642945, EPI_ISL_2642955, EPI_ISL_2642958, EPI_ISL_2642959, EPI_ISL_2642960, EPI_ISL_2642961, EPI_ISL_2642962, EPI_ISL_2642966, EPI_ISL_2642967, EPI_ISL_2642968, EPI_ISL_2643032, EPI_ISL_2643033, EPI_ISL_2643039, EPI_ISL_2643042, EPI_ISL_2643073, EPI_ISL_2643085, EPI_ISL_2643086, EPI_ISL_2643088, EPI_ISL_2643092, EPI_ISL_2643093, EPI_ISL_2643110, EPI_ISL_2643113, EPI_ISL_2643163, EPI_ISL_2643169, EPI_ISL_2643178, EPI_ISL_2643179, EPI_ISL_2643186, EPI_ISL_2643187, EPI_ISL_2643190, EPI_ISL_2643191, EPI_ISL_2643197, EPI_ISL_2643212, EPI_ISL_2643258, EPI_ISL_2643263, EPI_ISL_2643299, EPI_ISL_2643379, EPI_ISL_2643385, EPI_ISL_2643396, EPI_ISL_2643409, EPI_ISL_2643441, EPI_ISL_2643470, EPI_ISL_2643544, EPI_ISL_2643648, EPI_ISL_2643653 | | | |
| see above | SYNLAB Eesti OÜ | Department of Microbiology, Institute of Biomedicine and Translational Medicine, University of Tartu | Aare Abroi; Andrio Lahesaare; Arina Shablinskaja; Dagmar Hoidmets; Ene-Ly Jõgeda; Eveli Kallas; Heiki Niglas; Irja Lutsar; Kai Truusalu; Kaisa Truus; Katrin Kaarna; Kristi Huik; Liidia Dotsenko; Lili Azin Milani; Mari-Anne Härma; Mats Hansen; Meri Pauskar; Olga Sadikova; Paul Naaber; Radko Avi; Taavi Päll; Tuuli Reisberg; Ulvi Gerst Talas |
| EPI_ISL_1145357 | SYNLAB Jena Oncoscreen | Robert Koch Institute | |
| EPI_ISL_1357516 | SYNLAB Labor München Zentrum LMZ | Robert Koch Institute | |
| EPI_ISL_2262015 | SYNLAB MVZ Leinfelden-Echterdingen | Robert Koch Institute | |
| EPI_ISL_2113906, EPI_ISL_2114481 | SYNLAB MVZ Leverkusen | Robert Koch Institute | |
| EPI_ISL_1845492 | SYNLAB MVZ Trier | Robert Koch Institute | |
| EPI_ISL_2389790 | SYNLAB MVZ Weiden | Robert Koch Institute | |
| EPI_ISL_419301, EPI_ISL_419302, EPI_ISL_419303, EPI_ISL_419304 | Saitama Prefectural Institute of Public Health | Pathogen Genomics Center, National Institute of Infectious Diseases | Hajime Kamiya; Kentaro Itokawa; Makoto Kuroda; Masanori Hashino; Michiyo Shinohara; Motoi Suzuki; Rina Tanaka; Tsuyoshi Kishimoto; Tsuyoshi Sekizuka |
| EPI_ISL_684147 | Sakai City Institute of Public Health | Pathogen Genomics Center, National Institute of Infectious Diseases | Kentaro Itokawa; Makoto Kuroda; Masanori Hashino; Rina Tanaka; Tsuyoshi Sekizuka |
| EPI_ISL_2105815, EPI_ISL_2319293, EPI_ISL_2692637, EPI_ISL_2810090, EPI_ISL_2810116, EPI_ISL_2978450, EPI_ISL_2978587 | | | |
| see above | Salud Digna | Instituto Nacional de Medicina Genomica | Abraham Campos-Romero; Cedro-Tanda A; Escobar-Arrazola; Gonzalez-Barrera D; Herrera-Montalvo LA.; Hidalgo-Miranda A; Luna-Ruiz Marco; M; Mendoza-Vargas A; Moreno-Camacho José Luis; Munguia-Garza P; Ramirez-Vega O; Rangel-DeLeon D; Reyes-Grajeda JP; Rodriguez-Gallegos Jorge |
| EPI_ISL_636198, EPI_ISL_667046, EPI_ISL_730115 | San Diego County Public Health Laboratory | Andersen lab at Scripps Research | Brett Austin; Jovan Shephard; SEARCH Alliance San Diego with Tracy Basler |
| EPI_ISL_749155, EPI_ISL_749706, EPI_ISL_750174, EPI_ISL_2754033, EPI_ISL_2754203 | Sanatorio Americano | Institut Pasteur de Montevideo | Ana Carolina Mendonça; Andres Lizasoain; Andrés Lizasoain; Belén González; Camila Simoes; Cecilia Alonso; Cecilia Salazar; Daiana Mir; Emiliano Pereira; Fernando López-Tort; Fernando Motta; Gonzalo Bello; Ighor Arantes; Ignacio Ferrés; Jose Sotelo; Juan Zanetti; Leticia Maya; Letícia Garay Martins; Lucia Bilbao; Luciana Appolinario; Luciana Griffero; Lucía Spangenberg; Lucía Spangenberg; Mailen Arleo; Mariana Brandes; Marilda Mendonça Siqueira; Marilda Tereza Mar da Rosa; María José Benitez-Galeano; Martín Graña; María José Benítez-Galeano; Matías Castells; Matías Victoria; Matías Castells; Matías Salvo; Matías Victoria; Mauricio Méndez; Melissa Duquía; Natalia Rego; Natalia Reyes; Odhille Chappos; Pablo Smircich; Paola Cristina Resende; Pía Techera; Rodney Colina; Tamara Fernandez-Calero; Tamara Fernández-Calero; Tania Possi; Tatiana Schäffer Gregianini; Verónica Noya; Verónica Noya; Yasser Vega |
| EPI_ISL_2547347, EPI_ISL_2547348 | Sanddrift nc Clinic | National Health Laboratory Service/University of Cape Town (NHLS/UCT) | Arash Iranzadeh; Bruna Galvao; Carolyn Williamson; Deelan Doolabh; Diana Hardie; Gert Marais; Innocent Mudau; Lynn Tyers; Marvin Hsiao; Stephen Korsman |
| EPI_ISL_479800 | Sapporo City Institute of Public Health | Pathogen Genomics Center, National Institute of Infectious Diseases | Asami Ohnishi; Hajime Kamiya; Kentaro Itokawa; Makoto Kuroda; Masanori Hashino; Motoi Suzuki; Rina Tanaka; Tsuyoshi Sekizuka |
| EPI_ISL_1971032 | Sapporo City Institute of Public Health | Sapporo City Institute of Public Health | Asami Ohnishi; Kentaro Itokawa; Makoto Kuroda; Masanori Hashino; Masayuki Kikuchi; Rie Shimazaki; Rina Tanaka; Tsuyoshi Sekizuka; Yusuke Oguchi |
| EPI_ISL_1009123 | School of Pharmacy | School of Pharmacy | Ahmed Kandeil; Ghazi Kayali; Mina Nabil Kamel; Mohamed A Ali; Rabeh El-Shesheny; Walid Abi Habib |
| EPI_ISL_661185 | Scientific Veterinary Institute Novi Sad | Veterinary Specialized Institute "Kraljevo", Serbia | Afonso, C.; Banovic Djeri, B.; Jankovic, M.; Jovanovic, T.; Knezevic, A.; Petrovic, T.; Sekler, M.; Tesovic, B.; Vidanovic, D.; Volkening, J. |
| EPI_ISL_408977 | Serology, Virology and OTDS Laboratories (SAViD), NSW Health Pathology Randwick | NSW Health Pathology - Institute of Clinical Pathology and Medical Research; Centre for Infectious Diseases and Microbiology Laboratory Services; Westmead Hospital; University of Sydney | Carter I; Chen SC; Eden J-S; Holmes EC; Kok J and Dwyer DE for the 2019-nCoV Study Group*; Maddocks S; O'Sullivan MV; Rahman H; Rawlinson W; Rockett R; Sintchenko V |
| EPI_ISL_1754133, EPI_ISL_2179780 | Servicio Microbiología Hospital La Paz | Servicio Microbiología Hospital La Paz | Elie Dahdouh; Fernando Lázaro; Jesús Mingorance Cruz; Rubén Cáceres |
| EPI_ISL_856766, EPI_ISL_856774, EPI_ISL_2105574, EPI_ISL_2135170, EPI_ISL_2135699, EPI_ISL_2136027, EPI_ISL_2136086, EPI_ISL_2136141, EPI_ISL_2136143, EPI_ISL_2140067, EPI_ISL_2140071, EPI_ISL_2158708, EPI_ISL_2158745, EPI_ISL_2158754, EPI_ISL_2158755, EPI_ISL_2158761, EPI_ISL_2158762, EPI_ISL_2158766, EPI_ISL_2158773, EPI_ISL_2158774, EPI_ISL_2158779, EPI_ISL_2158815, EPI_ISL_2158817, EPI_ISL_2158830, EPI_ISL_2158850 | | | |
| see above | Servicio Virosis Respiratorias-Departamento Virología-INEI | Instituto Nacional Enfermedades Infecciosas C.G.Malbran | Avaro M.; Baumeister E.; Benedetti E.; Campos J.; Cisterna D.; Dattero ME; Lorenzo F.; Molina V.; Perandones C.; Poklepovich T.; Pontoriero A.; Russo M.; Tuduri E. |
| EPI_ISL_2516849 | Servicio de Microbiología Clínica (Complejo Hospitalario de Navarra, Pamplona) | Centro de Secuenciación NASERTIC | Ana Miqueleiz; Ana Navascués; Carmen Ezpeleta Baquedano |
| EPI_ISL_691645, EPI_ISL_691646, EPI_ISL_691648 | Servicio de Microbiología, Hospital Universitario Son Espases | SeqCOVID-SPAIN consortium/IBV(CSIC) | Antonio Oliver and SeqCOVID-SPAIN consortium; Carla López-Causapé; Jordi Reina |
| EPI_ISL_419687 | Servicio de Microbiología. Consorcio Hospital General Universitario de Valencia | Sequencing and Bioinformatics Service and Molecular Epidemiology Research Group. FISABIO-Public Health | Concepcion Gimeno; Fernando Gonzalez-Candelas; Giuseppe D'Auria; Griselda De Marco; Maria Alma Bracho; Maria Dolores Ocete; Neris Garcia-Gonzalez |
| EPI_ISL_2776001 | Servicio de Microbiología. Hospital General Universitario de Castellón | SeqCOVID-SPAIN consortium/IBV(CSIC) | María Dolores Tirado Balaguer and SeqCOVID-SPAIN consortium; Rosario Moreno Muñoz |
| EPI_ISL_509615 | Servicio de Microbiología. Hospital Universitario Donostia. OSI Donostialdea. Área de Enfermedades Infecciosas, Grupo de Infección Respiratoria y Resistencia Antimicrobiana. Instituto de Investigación Sanitaria Biodonostia | SeqCOVID-SPAIN consortium/IBV(CSIC) | Gustavo Cilla; Jose Maria Marimón and SeqCOVID-SPAIN consortium; Luis Piñeiro; Milagrosa Montes |
| EPI_ISL_452641 | Servicio de Microbiología. Hospital Universitario Donostia. OSI Donostialdea. Área de Enfermedades Infecciosas, Grupo de Infección Respiratoria y Resistencia Antimicrobiana. Instituto de Investigación Sanitaria Biodonostia. | SeqCOVID-SPAIN consortium/IBV(CSIC) | Gustavo Cilla; Jose Maria Marimón and SeqCOVID-SPAIN consortium; Luis Piñeiro; Milagrosa Montes |
| EPI_ISL_710547 | Sestre Milosrdnice University Hospital Center | Ruđer Boškovic Institute; Forensic Science Centre Ivan Vučetić; University of Zagreb Faculty of Science | Ana Livun; Antonela Blažeković; Boris Maček; Danilo Licastro; Dunja Glavaš; Fran Borovečki; Gordana Maravić Vlahoviček; Ivan Šamija; Ivana Ćelap; Josipa Skelin; Kristian Vlahoviček; Kristina Gotovac Jereić; Lidija Cvetko-Krajinović; Lucija Markulin; Maja Kuzman; Marina Korolija; Mario Štefanović; Mirjana Domazet-Lošo; Paula Štancl; Robert Belužić; Rosa Karlić; Sanja Tadinac; Tomislav Domazet-Lošo; Vjekoslav Tomaić; Vladimir Krajinović; Željka Mačak Šafranko |
| EPI_ISL_2545290, EPI_ISL_2834380, EPI_ISL_2834393, EPI_ISL_2834506, EPI_ISL_3019862, EPI_ISL_3022075, EPI_ISL_3022076 | | | |
| see above | Shamir Medical Center (Asaf Harofe) | Shamir Medical Center (Asaf Harofe) | Abu Hamad Ramzia; Adina Bar Chaim; Anna Vishnevsky; Chen Weiner; Netta Zuckerman; Nir Rainy; Patricia Benveniste-Lekovitz; Reut Sorek Abramovich; Yevgeni Yegorov |
| EPI_ISL_582125, EPI_ISL_582126 | Sheikh Khalifa Medical City | Molecular Surveillance lab Sheikh Khalifa Medical City | Amirtharaj Francis; Hala Imambaccus; Hiba Saud; Sahar Almarzooqi; Sajeed Abdul; Stefan Weber |
| EPI_ISL_582611, EPI_ISL_582613, EPI_ISL_582617, EPI_ISL_582646, EPI_ISL_582648, EPI_ISL_582677, EPI_ISL_582679, EPI_ISL_582683 | | | |
| see above | Sheikh Khalifa Medical City | Molecular/Surveillance lab Sheikh Khalifa Medical City | Amirtharaj Francis; Hala Imambaccus; Hiba Saud; Sahar Almarzooqi; Sajeed Abdul; Stefan Weber |
| EPI_ISL_2500849, EPI_ISL_2921027 | Shenzhen Center for Disease Control and Prevention | National Institute for Viral Disease Control and Prevention, China CDC | Can Zhu; Jiancheng Chen; Kai Nie; Long Chen; Peihua Niu; Renli Zhang; Shaoyu Deng; Shaoyu Deng and Yaqing He; Weihua Wu; Xiang Zhao; Xiaoliang Xiao; Xinyi Wei; Yaqing He; Yenan Feng; Ying Sun; Yue Li |
| EPI_ISL_2086220, EPI_ISL_2086224, EPI_ISL_2086226 | ShipMed Inc | KRISP, KZN Research Innovation and Sequencing Platform | Emmanuel SJ; Giandhari J; Lessells R; Naidoo Y; Pillay S; Ramphal U; Tegally H; Wilkinson E; de Oliveira T |
| EPI_ISL_2608422 | Siem Reap Provinical Laboratory | Virology Unit, Institut Pasteur du Cambodge | Cecile Troupin; Chau Darapheak; Chin Savuth; Erik A Karlsson; Jurre Y Siegers; Kraing Sidonn; Leakhena Pum; Ly Sovann; Veasna Duong; Yi Sengdoeurn |
| EPI_ISL_500570, EPI_ISL_610154 | Singapore General Hospital | Department of Microbiology | Chenhao Li; Chenhao Li; Karrie Ko; Karrie Ko; Kenneth Xin Long Chan; Kern Rei Chng; Kern Rei Chng; Kian Sing Chan; Kian Sing Chan; Kun Lee Lim; Kun Lee Lim; Lynette Oon; Lynette Oon; Niranjan Nagarajan; Niranjan Nagarajan; Nurdyana Abdul Rahman; Nurdyana Abdul Rahman; Sui Sin Goh |
| EPI_ISL_2470333 | Sonic - Labor Dr. von Froreich GmbH | Robert Koch Institute | |
| EPI_ISL_1569062 | Sonic - MVZ Medizinisches Labor Bremen GmbH | Robert Koch Institute | |

| | | | |
|---|---|---|---|
| EPI_ISL_2006881, EPI_ISL_2006899, EPI_ISL_2427243, EPI_ISL_2675217, EPI_ISL_3047978 | South Dakota Public Health Laboratory | South Dakota Public Health Laboratory | Chris Carlson; Jacob Garfin and Chris Carlson |
| EPI_ISL_513377, EPI_ISL_593699, EPI_ISL_593726, EPI_ISL_629030, EPI_ISL_740868, EPI_ISL_768602, EPI_ISL_1904462, EPI_ISL_1904464, EPI_ISL_2462416, EPI_ISL_2462422, EPI_ISL_2650459, EPI_ISL_2650461 | | | |
| see above | South Eastern Area Laboratory Services (SEALS) | NSW Health Pathology - Institute of Clinical Pathology and Medical Research; Westmead Hospital; University of Sydney | CIDM-PH et al. |
| EPI_ISL_2928014, EPI_ISL_2928016, EPI_ISL_2928017, EPI_ISL_2928018, EPI_ISL_2928019, EPI_ISL_2928020, EPI_ISL_2928021, EPI_ISL_2928023, EPI_ISL_2928025, EPI_ISL_2928026, EPI_ISL_2928027 | | | |
| see above | South Sudan Ministry of Health, WHO South Sudan, MRC/UVRI & LSHTM Uganda Research Unit | MRC/UVRI & LSHTM Uganda Research Unit, South Sudan Ministry of Health, WHO South Sudan | Abe G. Abias; Dan Lule Bugembe; Dennis Kenyi Lodiongo; James Ayei; John Rumunu; Joseph Francis Wamala; Juma John HM; Lul Lojok Deng; Matthew Cotten; My V.T. Phan; Pontiano Kaleebu; Richard Lino Loro Lako; Sudhir Bunga |
| EPI_ISL_456232, EPI_ISL_456238, EPI_ISL_456297, EPI_ISL_579133 | Southern Community Labs Dunedin | Institute of Environmental Science and Research (ESR) | Anja Werno; Antje van der Linden; Arlo Upton; Chris Mansell; David Hammer; Dragana Drinkovic; Erasmus Smit; Gary McAuliffe; Hana Sofia Andersson; Hermes Perez; James Ussher; Jill Sherwood; Jing Wang; Joep de Ligt; Josh Freeman; Julia Howard; Juliet Elvy; Lauren Jelly; Mary DeAlmeida; Matt Blakiston; Matt Storey; Matthew Rogers; Max Bloomfield; Michael Addidle; Michelle Balm; Muhammad Faisal; Nikki Freed; Olin Silander; Sally Roberts; Sarah Jefferies; Sharmini Muttaiyah; Susan Morpeth; Susan Taylor; Timothy Blackmore; Vani Sathyendran; Veronica Playle; Virginia Hope; Xiaoyun Ren |
| EPI_ISL_849739, EPI_ISL_849743 | Special Operations Medical Research Division, Defence Services Medical Research Centre | Special Operations Medical Research Division, Defence Services Medical Research Centre | Aung; Htun; K.K.; K.Z.; Lwin; Myint, K.; N.M.; Oo; P.K.; Win; Z.W.; Zaw, T. |
| EPI_ISL_513399 | St Vincent's Pathology (SydPath) | NSW Health Pathology - Institute of Clinical Pathology and Medical Research; Westmead Hospital; University of Sydney | CIDM-PH et al. |
| EPI_ISL_596672 | St.Vincent's University Hospital | St.Vincent's University Hospital | Aisling Purcell; Gabriel Gonzalez; Guerrino Macori; Kirsten Schaffer; Lynda Fenelon; Mary Lucey; Niamh Mullane; Suzie Coughlan; Séamus Fanning; Una Sutton-Fitzpatrick |
| EPI_ISL_1495136 | State Institution «Public Health Center of Ministry of Health of Ukraine» | Robert Koch Institute, ZBS1 Highly Pathogenic Viruses, Berlin, Germany | Andreas Nitsche; Annika Brinkmann; Iryna Demchyshyna; Janine Michel; Liudmyla Chernenko; Roman Rodyna; Steven Uddin |
| EPI_ISL_2779638, EPI_ISL_2779639 | State Key Laboratory of Emerging Infectious Diseases, The University of Hong Kong | The University of Hong Kong | Huachen Zhu; Lifeng Li; Xianfan Wu; Yi Guan; Yiu Man Cheung |
| EPI_ISL_2662727 | State Key Laboratory of Pathogen and Biosecurity, Beijing Institute of Microbiology and Epidemiology | State Key Laboratory of Pathogen and Biosecurity, Beijing Institute of Microbiology and Epidemiology | Campbell, A.; Campbell, J.; Gevao; Harding, D.; Lin, L.; P.F.; P.S.; Rogers, J.; Sahr; Wurie, I.; Zhang, J. |
| EPI_ISL_752613, EPI_ISL_753087, EPI_ISL_753170, EPI_ISL_1771478, EPI_ISL_2007889, EPI_ISL_2724554, EPI_ISL_2724734, EPI_ISL_3033249 | | | |
| see above | State Laboratories Division, Hawaii State Department of Health | State Laboratories Division, Hawaii State Department of Health | Ayana Garnet; Drew Kuwazaki; Edward Desmond; Pamela O'Brien; Razvan Sultana; Sabrina Diemert |
| EPI_ISL_2378030, EPI_ISL_2378031, EPI_ISL_2378033, EPI_ISL_2378143, EPI_ISL_2378146, EPI_ISL_2378147, EPI_ISL_2800610 | | | |
| see above | State Testing Facility | Altius Institute for Biomedical Research | Alex Isner; Alex Nguyen; Amanda Gale; Audra Johnson; Clem Green; Daniel Bates; Eric Thorland; Jacob Rodriguez; Jean Robinston; Jemma Nelson; Jessica Kunder; John Stamatoyannopoulos; Joshua Richards; Julia Wald; Kneshay Harper; Lauren Mitchell; Mark Frerker; Matt Hartman; Michael Buckley; Muhammad Halimun; Rebecca Bruders; Sadie Patraw; Sofia Olsson; Tobias Ragoczy |
| EPI_ISL_2685046 | Statens Serum Institut Bioinformatics and Microbial Genomics | Statens Serum Institut Bioinformatics and Microbial Genomics | Danish Covid-19 Genome Consortium |
| EPI_ISL_2375915 | Steinkopf PHC Clinic | NHLS/UCT | Arash Iranzadeh; Bruna Galvao; Carolyn Williamson; Deelan Doolabh; Diana Hardie; Innocent Mudau; Kruger Marais; Lynn Tyers; Marvin Hsiao; Stephen Korsman |
| EPI_ISL_640079, EPI_ISL_2375919 | Stellenbosch Hospital wc STB | NHLS/UCT | Arash Iranzadeh; Bruna Galvao; Carolyn Williamson; Deelan Doolabh; Diana Hardie; Innocent Mudau; Kruger Marais; Lynn Tyers; Marvin Hsiao; Stephen Korsman |
| EPI_ISL_2921612 | Sultan Haji Ahmad Shah Hospital | Institute for Medical Research, Infectious Disease Research Centre, National Institutes of Health, Ministry of Health Malaysia | Anasir MI; Azizan MA; Kamel K; Mohd Zawawi Z; Ramly N; Robert F; Suppiah J; Thayan R |
| EPI_ISL_2931923 | Sultanah Fatimah Specialist Hospital, Muar | Institute for Medical Research, Infectious Disease Research Centre, National Institutes of Health, Ministry of Health Malaysia | Anasir MI; Azizan MA; Kamel K; Mohd Zawawi Z; Ramly N; Robert F; Suppiah J; Thayan R |
| EPI_ISL_2811879 | Sultanah Nora Ismail Hospital, Batu Pahat | Institute for Medical Research, Infectious Disease Research Centre, National Institutes of Health, Ministry of Health Malaysia | Azizan MA; Kamel K; Mohd Zawawi Z; Ramly N; Robert F; Suppiah J; Thayan R |
| EPI_ISL_450747 | Sunnybrook Health Sciences Centre | Department of Laboratory Medicine and Molecular Diagnostics, Sunnybrook Health Sciences Centre | Amogelang R. Raphenya; Andrew G. McArthur; Arinjay Banerjee; Bo Wang; Brian P. Alcock; Finlay Maguire; Hamza Mbareche; Hassaan Maan; Jalees A. Nasir; Julian A. Hiscox; Karen Mossman; Kendrick M. Smith; Muhannad Alruwaili; Natalie C. Knox; Patryk Aftanas; Robert A. Kozak; Samira Mubareka |
| EPI_ISL_2406466 | Svay Rieng Rapid Response Team | Virology Unit, Institut Pasteur du Cambodge | Cecile Troupin; Chau Darapheak; Chin Savuth; Erik A Karlsson; Jurre Y Siegers; Kraing Sidonn; Leakhena Pum; Ly Sovann; Veasna Duong; Yi Sengdoeurn |
| EPI_ISL_1193078, EPI_ISL_1617530, EPI_ISL_1618454, EPI_ISL_1676225, EPI_ISL_1807444, EPI_ISL_1898458, EPI_ISL_1899265, EPI_ISL_1899290, EPI_ISL_1900277, EPI_ISL_2033384, EPI_ISL_2033792, EPI_ISL_2099115, EPI_ISL_2278930, EPI_ISL_2410253, EPI_ISL_2414900, EPI_ISL_2415990, EPI_ISL_2417386, EPI_ISL_2419171, EPI_ISL_2420110, EPI_ISL_2613350, EPI_ISL_2792809, EPI_ISL_2792949, EPI_ISL_2793678, EPI_ISL_2794645, EPI_ISL_2889039, EPI_ISL_2980627 | | | |
| see above | Swedish national genomic surveillance program of SARS-CoV-2 | The Public Health Agency of Sweden | Alma Brolund; Maria Lind Karlberg; Maximilian Riess; Swedish national genomic surveillance program of SARS-CoV-2 |
| EPI_ISL_860745 | Swiss National Reference Centre for Influenza | Swiss National Reference Centre for Influenza | Ana Rita Gonçalves Cabecinhas; Christoph Grünig; Florian Laubscher; Laurent Kaiser; Samuel Cordey |
| EPI_ISL_478718 | Sydney South West Pathology Service (SSWPS) - Concord Repatriation General Hospital - NSW Health Pathology | NSW Health Pathology - Institute of Clinical Pathology and Medical Research; Westmead Hospital; University of Sydney | CIDM-PH et al. |
| EPI_ISL_2811986, EPI_ISL_2812836, EPI_ISL_2812857 | Sydney South West Pathology Service (SSWPS) - Liverpool Hospital - NSW Health Pathology | NSW Health Pathology - Institute of Clinical Pathology and Medical Research; Westmead Hospital; University of Sydney | CIDM-PH et al. |
| EPI_ISL_767863, EPI_ISL_803111, EPI_ISL_2462428 | Sydney South West Pathology Service (SSWPS) - Royal Prince Alfred Hospital - NSW Health Pathology | NSW Health Pathology - Institute of Clinical Pathology and Medical Research; Westmead Hospital; University of Sydney | CIDM-PH et al. |
| EPI_ISL_1319275, EPI_ISL_2107235, EPI_ISL_2107284, EPI_ISL_2546950, EPI_ISL_2547057, EPI_ISL_2788795, EPI_ISL_2788800, EPI_ISL_2788837 | | | |
| see above | Synlab Eesti OÜ | 1. Laboratory of Communicable Diseases (Estonia); 2. Eurofins Genomics Europe Sequencing GmbH | Liidia Dotsenko et al. |
| EPI_ISL_1655749, EPI_ISL_1655750, EPI_ISL_1656735 | Synlab Haut de France | UMR 8199/1283 EGID | Derhourhi Mehdi; Mehdi Derhourhi |
| EPI_ISL_2897132 | Szpital Uniwersytecki w Krakowie | 1. Tricity SARS-CoV-2 sequencing consortium: University of Gdansk, Medical University of Gdansk, Vaxican Ltd., Invicta Ltd. 2. National Institute of Public Health - National Institute of Hygiene, Warsaw, Poland | Celina Cybulska; Karolina Gackowska; Katarzyna Groth; Katarzyna Zacharczuk; Krystyna Bienkowska Szewczyk; Lukasz Rabalski; Maciej Grzybek; Maciej Kosinski; Magdalena Nowakowska; Marcin Lubocki; Małgorzata Sadkowska-Todys; Tomasz Wołkowicz |
| EPI_ISL_2694620, EPI_ISL_2694702, EPI_ISL_2695504, EPI_ISL_2695517 | TAMBO MEMORIAL | National Institute for Communicable Diseases of the National Health Laboratory Service | Amoako DG; Bhiman JN; Everatt J; Ismail A; Mahlangu B; Mnguni A; Mohale T; Ntuli N; Scheepers C |
| EPI_ISL_2662568, EPI_ISL_2662572, EPI_ISL_2662574, EPI_ISL_2709951, EPI_ISL_2709967, EPI_ISL_2709989, EPI_ISL_2827057, EPI_ISL_3024631 | | | |
| see above | TAMBO MEMORIAL LABORATORY | National Institute for Communicable Diseases of the National Health Laboratory Service | Amoako DG; Bhiman JN; Everatt J; Ismail A; Mahlangu B; Mnguni A; Mohale T; Ntuli N; Scheepers C |
| EPI_ISL_914824 | TAMIZAJE COMUNITARIO - PASO CANOAS | Inciensa, Instituto Costarricense de Investigación y Enseñanza en Nutrición y Salud | Adriana Godínez; Claudio Soto-Garita; Estela Cordero; Francisco Duarte; Hebleen Porras; Melany Calderón & Mariel López |
| EPI_ISL_1530791 | TGen North | TGen North | Ashlyn Pfeiffer; Chris French; Darrin Lemmer; Dave Engelthaler; Hayley Yaglom; Heather Centner; Jolene Bowers; The Arizona COVID Genomics Union (ACGU) |
| EPI_ISL_426629, EPI_ISL_436105, EPI_ISL_447252, EPI_ISL_447593, EPI_ISL_457726, EPI_ISL_2693000, EPI_ISL_2693004, EPI_ISL_2693006 | | | |
| see above | TSGH-CP molecular lab | TSGH-CP molecular lab | Cherng-Lih Perng; Chien-Wen Chen; Chih-Kai Chang; Feng-Yee Chang; Hsing-Yi Chung; Hung-Sheng Shang; Jung-Chung Lin; Kuo-Ming Yeh; Kuo-Sheng Hung; Ming-Jr JIAN; Ming-Jr Jian; Sheng-Kang Chiu; Shih-Hung Tsai; Tien-Yao Chang |
| EPI_ISL_2987237, EPI_ISL_2987242, EPI_ISL_2987249, EPI_ISL_2987322, EPI_ISL_2987328, EPI_ISL_2987337, EPI_ISL_2987343, EPI_ISL_2987344 | | | |
| see above | TSHEPONG LABORATORY | National Institute for Communicable Diseases of the National Health Laboratory Service | Amoako DG; Bhiman JN; Everatt J; Ismail A; Mahlangu B; Mnguni A; Mohale T; Ntuli N; Scheepers C |
| EPI_ISL_2827906, EPI_ISL_2827908, EPI_ISL_2827911, EPI_ISL_2984879, EPI_ISL_2984882, EPI_ISL_2984913, EPI_ISL_2984921, EPI_ISL_2984926, EPI_ISL_2984930, EPI_ISL_2984938 | | | |
| see above | TSHEPONG LABORATORY | National Institute for Communicable Diseases of the National Health Laboratory Service | Amoako DG; Bhiman JN; Everatt J; Ismail A; Mahlangu B; Mnguni A; Mohale T; Ntuli N; Scheepers C |
| EPI_ISL_2506492, | TYKS, Kliininen mikrobiologia | Expert Microbiology, National Institute for Health and Welfare | Carita Savolainen-Kopra; Erika Lindh; Haider al-Hello; Jani Halkilahti; Kirsi Liitsola; Niina Ikonen; Olli Vapalahti; Pekka Ellonen; Phuoc Truong; Päivi Laurila; Ravi Kant; Sari Hannula; Soile Blomqvist; Teemu Smura |

| | | | |
|---|---|---|---|
| EPI_ISL_2590709 | | | |
| EPI_ISL_1972539 | Tampin Hospital | Institute for Medical Research, Infectious Disease Research Centre, National Institutes of Health, Ministry of Health Malaysia | Azizan MA; Kamel K; Mohd Zawawi Z; Ramly N; Robert F; Suppiah J; Thayan R |
| EPI_ISL_911750 | Tanjungpura University Hospital | Tanjungpura University Hospital | Ambar Rialita; Andriani; Delima Fajar Liana; Eka Ardiani Putri; Helmi Sastriawan; Mahyarudin; Muhammad Kahtan; Puji Astuti; Rini Andriani; Sofi Siti Shofiyah; Virhan Novianry; Wiwik Windarti |
| EPI_ISL_2611673, EPI_ISL_2651121, EPI_ISL_3019481, EPI_ISL_3019485 | Temporary Specimen Collection Centre at the AsiaWorld-Expo | Hong Kong Department of Health | Alan K.L. Tsang; Dominic N.C. Tsang; Edman T.K. Lam; Ken H.L. Ng; Peter C.W. Yip; Rickjason C.W. Chan |
| EPI_ISL_934972 | Thai Red Cross Emerging Infectious Diseases Health Science Centre, Chulalongkorn Hospital, Faculty of Medicine, Chulalongkorn University | Thai Red Cross Emerging Infectious Diseases Center and Faculty of Medicine, Chulalongkorn University | Apaporn Rodpan; Opass Putcharoen; Rome Buathong; Sininat Petcharat; Sopon Iamsirithaworn; Supaporn Wacharapluesadee; Thiravat Hemachudha; Weenassarin Ampoot; Yutthana Joyjinda |
| EPI_ISL_1490226, EPI_ISL_2230689, EPI_ISL_2230690, EPI_ISL_2230691, EPI_ISL_2230692, EPI_ISL_2230696, EPI_ISL_2230698, EPI_ISL_2230699, EPI_ISL_2478969, EPI_ISL_2500850, EPI_ISL_2544758, EPI_ISL_2545191, EPI_ISL_2545192, EPI_ISL_2545194, EPI_ISL_2545198 | | | |
| see above | The Caribbean Public Health Agency | Carrington Lab, Department of PreClinical Sciences, Faculty of Medical Sciences, The University of the West Indies | Adesh Ramsubhag; Anushka Ramjag; Arianne Brown-Jordan; Avery Hinds; Ayoola Oyinloye; Chinna Chinnadurai; Christine V. F. Carrington; Christopher Oura; Gabriel Escobar; Jaya Jayaraman; Jerome Foster; Karla Georges; Kenneth George; Marsha Ivey; Naresh Nandram; Narine Singh; Nikita S. D. Sahadeo; Nuno Faria; Oliver Pybus; Rahul Naidu; Rajini Haraksingh; Risha Singh; Roshan Parasram; Sarah Hill; Shawn Charles; Stanley Giddings; SueMin Nathaniel; Vernie Ramkissoon |
| EPI_ISL_2631411 | The Caribbean Public Health Agency | Carrington Lab, Department of PreClinical Sciences, Faculty of Medical Sciences, The University of the West Indies, St Augustine Campus | Anushka Ramjag; Arianne Brown-Jordan; Avery Hinds; Christine V. F. Carrington; Christopher Oura; Gabriel Escobar; Karla Georges; Naresh Nandram; Nikita S. D. Sahadeo; Nuno Faria; Oliver Pybus; Risha Singh; Roshan Parasram; Sarah Hill; SueMin Nathaniel; Vernie Ramkissoon |
| EPI_ISL_2557166, EPI_ISL_2565080, EPI_ISL_2621679, EPI_ISL_2621681, EPI_ISL_2621714, EPI_ISL_2621722, EPI_ISL_2626658, EPI_ISL_2626661, EPI_ISL_2626663 | | | |
| see above | The Caribbean Public Health Agency | Carrington Lab, Department of Preclinical Sciences, Faculty of Medical Sciences, The University of the West Indies | Anushka Ramjag; Arianne Brown-Jordan; Avery Hinds; Christine V. F. Carrington; Christopher Oura; Gabriel Escobar; Jacquiline Bisesor-McKenzie; Karla Georges; Kenneth George; Naresh Nandram; Nikita S. D. Sahadeo; Nuno Faria; Oliver Pybus; Risha Singh; Roshan Parasram; Sarah Hill; Sharon Belmar-George; SueMin Nathaniel; Vernie Ramkissoon |
| EPI_ISL_2649790, EPI_ISL_2658366, EPI_ISL_2658367, EPI_ISL_2658376, EPI_ISL_2658405, EPI_ISL_2678129, EPI_ISL_2678143, EPI_ISL_2678145, EPI_ISL_2678146, EPI_ISL_2678161, EPI_ISL_2678163, EPI_ISL_2678165, EPI_ISL_2678166, EPI_ISL_2678168, EPI_ISL_2678169, EPI_ISL_2678171, EPI_ISL_2678172, EPI_ISL_2678180, EPI_ISL_2716565, EPI_ISL_2716569, EPI_ISL_2716570, EPI_ISL_2716574, EPI_ISL_2716576, EPI_ISL_2716577, EPI_ISL_2716580, EPI_ISL_2716583, EPI_ISL_2716594, EPI_ISL_2716595, EPI_ISL_2716602, EPI_ISL_2756569, EPI_ISL_2756571, EPI_ISL_2756578, EPI_ISL_2756579, EPI_ISL_2756580, EPI_ISL_2756581, EPI_ISL_2756582, EPI_ISL_2756585, EPI_ISL_2955588, EPI_ISL_2955590, EPI_ISL_2967979, EPI_ISL_2967980, EPI_ISL_2967981, EPI_ISL_2967982, EPI_ISL_2967983, EPI_ISL_2967984, EPI_ISL_2967985, EPI_ISL_2967986, EPI_ISL_2967991, EPI_ISL_2967992, EPI_ISL_2967993 | | | |
| see above | The Caribbean Public Health Agency | Carrington Lab, Department of Preclinical Sciences, Faculty of Medical Sciences, The University of the West Indies, St Augustine Campus | Anushka Ramjag; Arianne Brown-Jordan; Avery Hinds; Christine V. F. Carrington; Christopher Oura; Gabriel Escobar; Hazel Laws; Karla Georges; Kenneth George; Naresh Nandram; Nikita S. D. Sahadeo; Nuno Faria; Oliver Pybus; Rhonda Sealey-Thomas; Risha Singh; Roshan Parasram; Sarah Hill; Sharon Belmar-George; Shawn Charles; Simone Keizer-Beache; SueMin Nathaniel; Vernie Ramkissoon |
| EPI_ISL_2860169 | The Medical City | Philippine Genome Center | Alethea R. de Guzman; Anna Ong-Lim; Arianne A. Zamora; Benedict A. Maralit; Carlo M. Lapid; Celia Carlos; Devon Ray Pacial; Diomedes A. Carino; Edsel Maurice Salvaña; El King D. Morado; Elcid Aaron R. Pangilinan; Eva Maria Cutiongco-de la Paz; Francis A. Tablizo; Henrietta Marie Rodriguez; Jaime C. Montoya; Jan Michael C. Yap; Jarvin E. Nipales; Jo-Hannah S. Llames; John Q. Wong; Joshua Gregor A. Dizon; Juan Antonio R. Magalang; Karol Sophia Agape R. Padilla; Kenneth M. Kim; Kris P. Punayan; Krisitna Patriz Dela Cruz; Lindsay Claire D.L. Carandang; Ma. Exanil Plantig; Marc Edsel C. Ayes; Maria Rosario Singh-Vergeire and Cynthia P. Saloma; Maria Sofia L. Yangzon; Marielle M Gamboa; Marissa Alejandria; Nina Francesca Bustamante; Razel Nikka M. Hao; Renato Jacinto Q. Mantaring; Rianna Patricia S. Cruz; Sheila Mae M. Araiza; Yvonne Valerie Austria; Zipporah Mariebelle R. Enriquez; Zyrel V. Mollejon |
| EPI_ISL_577632, EPI_ISL_577633, EPI_ISL_792690, EPI_ISL_792699, EPI_ISL_850688 | The National Institute of Public Health | State Veterinary Institute Prague | A; D; H; J; Jirincova; L; Nagy; Novakova; Trnka; Vecerova |
| EPI_ISL_489708 | The National Institute of Public Health | The National Institute of Public Health and State Veterinary Institute Prague | A; D; H; J; Jirincova; L; Nagy; Novakova; Trnka; Vecerova |
| EPI_ISL_424583, EPI_ISL_1586190, EPI_ISL_1586350, EPI_ISL_1586370, EPI_ISL_1586520 | The National University Hospital of Iceland | deCODE genetics | Agnar Helgason; Alma Moller; Arna B Agustsdottir; Arnaldur Gylfason; Asgeir Sigurdsson; Aslaug Jonasdottir; Berglind Eiriksdottir; Bjarni Thorbjornsson; Brynjar O Jensson; Daniel F Gudbjartsson; Droplaug N Magnusdottir; Elisabet E Gardarsdottir; Emil A Thorarensen; Gardar Sveinbjornsson; Gisli Masson; Gudmundur Georgsson; Gudmundur L Norddahl; Gudrun Sigmundsdottir; Hakon Jonsson; Hannes Eggertsson; Hilma Holm; Ingileif Jonsdottir; Jona Saemundsdottir; Kamilla S Josefsdottir; Kari Stefansson; Karl G Kristinsson; Kjartan R Gudmundsson; Kristin E Sveinsdottir; Kristin E Sveinsdottir; Louise le Roux; Maney Sveinsdottir; Olafia S Gretarsdottir; Olafia S Gretarsdottir; Olafur T Magnusson; Pall Melsted; Patrick Sulem; Run Fridriksdottir; Solvi Rognvaldsson; Thora R Gunnarsdottir; Thordur Kristjansson; Thorolfur Gudnason; Unnur Thorsteinsdottir |
| EPI_ISL_648135 | The Public Health Agency of Sweden | The Public Health Agency of Sweden | Anna Risberg; Anna-Malin Linde; Karin Tegmark-Wisell; Maria Lind Karlberg; Mattias Haukland; Mia Brytting; Olov Svartstrom; Oskar Karlsson Lindsjo; Petra Edquist; Reza Advani; Sandra Broddesson |
| EPI_ISL_754229 | The Republican Research and Practical Center for Epidemiology and Microbiology (RRPCEM) | WHO National Influenza Centre Russian Federation | Anatoly Krasko; Andrey Komissarov; Anna Ivanova; Artem Fadeev; Daria Danilenko; Dmitry Bazhenov; Dmitry Lioznov; Elena Gasich; Elena Nabieva; Georgii Bazykin; Kirill Bulda; Ksenia Safina; Kseniya Komissarova |
| EPI_ISL_2631429 | The Trinidad and Tobago Public Health Laboratory | Carrington Lab, Department of PreClinical Sciences, Faculty of Medical Sciences, The University of the West Indies, St Augustine Campus | Anushka Ramjag; Arianne Brown-Jordan; Avery Hinds; Christine V. F. Carrington; Christopher Oura; Gabriel Escobar; Karla Georges; Naresh Nandram; Nikita S. D. Sahadeo; Nuno Faria; Oliver Pybus; Risha Singh; Roshan Parasram; Sarah Hill; SueMin Nathaniel; Vernie Ramkissoon |
| EPI_ISL_2658353 | The Trinidad and Tobago Public Health Laboratory | Carrington Lab, Department of Preclinical Sciences, Faculty of Medical Sciences, The University of the West Indies, St Augustine Campus | Anushka Ramjag; Arianne Brown-Jordan; Avery Hinds; Christine V. F. Carrington; Christopher Oura; Gabriel Escobar; Karla Georges; Naresh Nandram; Nikita S. D. Sahadeo; Nuno Faria; Oliver Pybus; Risha Singh; Roshan Parasram; Sarah Hill; SueMin Nathaniel; Vernie Ramkissoon |
| EPI_ISL_2876985 | Thembalethu CDC wc THC | NHLS/UCT | Arash Iranzadeh; Bruna Galvao; Carolyn Williamson; Deelan Doolabh; Diana Hardie; Gert Marais; Innocent Mudau; Lynn Tyers; Marvin Hsiao; Stephen Korsman |
| EPI_ISL_729274, EPI_ISL_729275, EPI_ISL_755695, EPI_ISL_755883, EPI_ISL_792087, EPI_ISL_961405 | Toronto Invasive Bacterial Diseases Network | McMaster University | Ahmed Draia; Allison McGeer; Andrew G. McArthur; Angel Li; Emily Panousis; Hooman Derakhshani; Jalees Nasir; Kuganya Nirmalarajah; Michael Surette; Patryk Aftanas; Samira Mubareka |
| EPI_ISL_853223, EPI_ISL_853236, EPI_ISL_853247, EPI_ISL_853567, EPI_ISL_853586 | Toronto Invasive Bacterial Diseases Network | Ontario Institute for Cancer Research | Allison McGeer; Angel Li; Bernard Lam; Ilinca Lungu; Jared T. Simpson; Kuganya Nirmalarajah; Lawrence E. Heisler; Lubaina Kothari; Michael Laszloffy; Patryk Aftanas; Paul Krzyzanowski; Richard de Borja; Samira Mubareka |
| EPI_ISL_2140718 | Touwsranten Clinic wc TST | NHLS/UCT | Arash Iranzadeh; Bruna Galvao; Carolyn Williamson; Deelan Doolabh; Diana Hardie; Innocent Mudau; Kruger Marais; Lynn Tyers; Marvin Hsiao; Stephen Korsman |
| EPI_ISL_2779295 | Trans Nzoia hospital | USAMRD-A, Basic Science Laboratory | Alan Lemtudo; Beth Mutai; Brian Andika; Carol Kifude; Clement Masakwe; Eric Muthanje; Esther Omuseni; Faith Sigei; Gathii Kimita; George Awinda; John Waitumbi; Josphat Nyataya; Rachel Githii; Rehema Liyai; Stephen Ochola |
| EPI_ISL_610265 | TriCore Reference Laboratories | Center for Global Health, University of New Mexico Health Sciences Center | Darrell Dinwiddie; Daryl Domman; Joseph Hicks; Kurt Schwalm; Michael Edwards; Twila Kunde |
| EPI_ISL_717692, EPI_ISL_717697, EPI_ISL_756362, EPI_ISL_2230685, EPI_ISL_2230686 | Trinidad Public Health Laboratory | Carrington Lab, Department of PreClinical Sciences, Faculty of Medical Sciences, The University of the West Indies | Adesh Ramsubhag; Arianne Brown-Jordan; Avery Hinds; Chinna Chinnadurai; Christine V. F. Carrington; Christopher Oura; Gabriel Escobar; Jaya Jayaraman; Jerome Foster; Karla Georges; Marsha Ivey; Naresh Nandram; Nikita S. D. Sahadeo; Nuno Faria; Oliver Pybus; Rahul Naidu; Rajini Haraksingh; Risha Singh; Roshan Parasram; Sarah Hill; Stanley Giddings; SueMin Nathaniel; Vernie Ramkissoon |
| EPI_ISL_2547369 | Tshwaragano Hospital | National Health Laboratory Service/University of Cape Town (NHLS/UCT) | Arash Iranzadeh; Bruna Galvao; Carolyn Williamson; Deelan Doolabh; Diana Hardie; Gert Marais; Innocent Mudau; Lynn Tyers; Marvin Hsiao; Stephen Korsman |
| EPI_ISL_2802121 | Tsineng Clinic | NHLS/UCT | Arash Iranzadeh; Bruna Galvao; Carolyn Williamson; Deelan Doolabh; Diana Hardie; Gert Marais; Innocent Mudau; Lynn Tyers; Marvin Hsiao; Stephen Korsman |
| EPI_ISL_1225161 | Turkish Society of Internal Medicine | Gen Era Diagnostics Corporation Life Science Research and Molecular Diagnostics | Serhat Ünal |
| EPI_ISL_2625892 | UAB Diagnostikos laboratorija | Institute of Biotechnology, Life Sciences Center, Vilnius University | Emilija Vasiliunaite__ Milda Norkiene__ Danguole Ziogiene__ Albertas Timinskas__ Alma Gedvilaite |
| EPI_ISL_2334699 | UAB Diagnostikos laboratorija | National Public Health Surveillance Laboratory | Ana Steponkiene; Danas Baksa; Jelena Razmuk; Lukas Vasionis; Lukas Zemaitis; Migle Gabrielaite; Svajune Muralyte |
| EPI_ISL_2428034, EPI_ISL_2694419, EPI_ISL_2694447 | UAB InMedica | National Public Health Surveillance Laboratory | Ana Steponkiene; Danas Baksa; Jelena Razmuk; Lukas Vasionis; Lukas Zemaitis; Migle Gabrielaite; Svajune Muralyte |
| EPI_ISL_1581485, EPI_ISL_2427828, EPI_ISL_2427854 | UAB InMedica | Vilnius University Hospital Santaros Klinikos, Center of Laboratory Medicine | Daniel Naumovas; Dovile Ezerskyte; Gytis Dudas; Ingrida Olendraite; Laimonas Griskevicius; Ligita Raugaite; Mindaugas Stoskus; Monika Katenaite; Rimvydas Norvilas |
| EPI_ISL_2340539 | UAB Medicina practica laboratorija | Lithuanian University of Health Sciences | Arnoldas Pautienius; Dovydas Gecys; Gediminas Alzbutas; Kamile Tamusauskaite; Lukas Zemaitis; Vaiva Lesauskaite |
| EPI_ISL_2082397, EPI_ISL_2694049 | UAB Medicina practica laboratorija | National Public Health Surveillance Laboratory | Ana Steponkiene; Danas Baksa; Jelena Razmuk; Lukas Vasionis; Lukas Zemaitis; Migle Gabrielaite; Svajune Muralyte |
| EPI_ISL_2758730 | UEL | IPEC Guarapuava | NAPI-Genômica (Novos Arranjo de Pesquisa e Inovação em Genômica): Ademar Dantas da Cunha Júnior Adriano Ferrasa Adriano Mondini Aldo Przybysz Alessandra Lourenço Cecchini Armani Alex Sandro Jorge Alexandra Ivo de Medeiros Alexandre Maller Aline Cristina Batista Rodrigues Johann Ana Lucia Ferreira Ana Marisa Fusco Almeida Anderson Joel Martino Andrade André Luís Laforga Vanzela Andrea Duarte Doetzer Andrea Name Colado Simao Andressa Pereira de Souza Anelisa Ramão Angelica Beate Winter Boldt Anna Hermínia Castro Gomes de Amorim Anna Silvia Penteado Setti da Rocha Antonio Camilo da Silva Filho Antonio Stabelini Neto Arthur Hirata Bertachi Barbara Mendes Paz Chao Betty Cristiane Kuhn Bruno Ambrozio Galindo Bruno Ribeiro Cruz Camilla Reginatto De Pierri Carla Fredrichsen Moya Araujo Carla Fredrichsen Moya Araujo Carlos Alberto Oliveira de Biagi Junior Carlos Augusto Nassar Carlos Eduardo Buss Carlos Gilberto Carlotti Junior Carlos Henrique Schneider Carolina Panis Carolina Weigert |

| | | | |
|---|---|---|---|
| | | | Galvão Caroline de Jesus Coelho Donha Caroline Guisantes de Salvo Toni Caryna Eurich Mazur Catiuscie Cabreira da Silva Tortorella Celso F. D. Doliveira Cesar Luiz Boguszewski Christiane Pienna Soares Chung Man Chin Claudia Moro Cleverson Busso Cristiane Cominetti Daiane Priscila Simão-Silva Dalila Luciola Zanette Daniel de Paula Daniel de Paula Daniel Rech Daniela Fiori Gradia Daniela Pretti da Cunha Tirapelli Daniela Viganó Zanoti Jeronymo Daniele Ukan Danielle Malheiros Ferreira Danielle Venturini Deborah Catharine de Assis Leite Deivid Calebe de Souza Dennis Armando Bertolini Edenir Inez Pamero Edna Maria Vissoci Reiche Edson Roberto Arpini Miguel Eduardo José de Almeida Araújo Eliana Carolina Vespero Eliandro Reis Tavares Elza Kimura Grimshaw Emanuel Maltempi de Souza Emanuele Cristina Gustani Buss Émerson Carraro Emiliana Cristina Melo ENILze Maria de Souza Fonseca Ribeiro Enilze Maria de Souza Fonseca Ribeiro Erika Izumi Erika Seki Kioshima Cotica Evani Marques Pereira Fabio Negretti Fábio Rodrigues Ferreira Seiva Felipe Dunin dos Santos Felipe Tuon Fernanda Andreia Rosa Fernanda Cestaro Prado Cortez Fernanda Ivanski Fernanda Maris Peria Flavia Regina Oliveira de Barros Franciele Aní Caovilla Follador Franciele Mara Lucca Zanardo Bohm Francinete Ramos Campos Fulviana Silva Nishiyama GABRIEL RIBEIRO CORDEIRO Gabriela Datsch Bennemann Gisele Santos de Oliveira Glaucio Valdameri Glauco Akelinghton Freire Vitiello Glauco Vieira Miranda Glaura Scantamburlo ALves Fernandes Guilherme Ferreira Silveira Gustavo Bianchini Porfírio Gustavo Lenci Marques Hélito Volpato Hildebrando Masshiro Nagai Huei Diana Lee Ilce Mara de Syllos Cólus Iris Rabinovich Israel Gomy Jackson Kawakami Jacques Duílio Brancher Jaime Luís Lopes Rocha Jaqueline Carvalho de Oliveira Jean Henrique da Silva Rodrigues Jean Leandro dos Santos Jeane Eliete Laguila Visentainer João Paulo Bianchi Ximenez Joaquim Manoel da Silva Jociani Ascari Joel Donazzolo Jorge Luis Maria Ruiz Jose Knopfholz José Luis da Conceição Silva José Sebastião dos Santos Joseane Carla Schabarum Juliana Cheleski Wiggers Juliana Mara Serpeloni Juliana Morini Küpper Cardoso Perseguini Karen Brajão de Oliveira Karin Braun Prado Karine Aparecida de Lima Katiany Rizzieri Caleffi Ferracioli Katiuscia de Oliveira Francisco Gabriel Kelvinson Fernandes Viana Larissa Beatriz Cossalter Larissa Danielle Bahls Pinto Laurival Antonio Vilas Boas Léia Carolina Lucio Líbero Mezzadri Neto Ligia Carla Faccin Galhardi Lirane Elize Defante Ferreto Luciana Furlaneto Maia Luciana Oliveira de Fariña Luciana Reis Azevedo Alanis Luciane Regina Cavalli Lucy Megumi Yamauchi Lioni Luis Paulo Gomes Mascarenhas Luis Paulo Gomes Mascarenhas Luís Paulo Mascarenhas Lupe Furtado Alle Lyvia Regina Biagi Silva Bertachi Mara Antonia Ramos Costa Mara L. Cordeiro Marcela Maria Birolim Marcelo Ricardo Vicari Marcia Edilaine Lopes Consolaro Marcia Holsbach Beltrame Marcia Regina Eches Perugini Marcos Abdo Arbex Marcos Pileggi MARCOS TADEU GRZELCZAK Marcus Peikriszwili Tartaruga Maria Angelica Ehara Watanabe Maria Antonia Ramos Costa Maria Claudia Gross Maria José Soares Mendes Giannini Maria Leandra Terencio Maria Lúcia Bonfleur Maria Luiza Guimarães de Oliveira Maria Luiza Petzl-Erler Mariana Abe Vicente Cavagnari Marina Kimiko Kadowaki Marise Fonseca dos Santos Marla Karine Amarante Maurício Turkiewicz Mauro Antonio Alves Castro Michel Rodrigo Zambrano Passarini Michele Potrich Michelle Orane Schemberger Milena Massumi Kozonoe Mônica Degraf Cavallin Monica Tereza Suldofski Mucio Luiz de Assis Cirino Nadia Graciele Krohn Najeh Maissar Khalil Nédia de Castilhos Ghisi Neide Tomimura Costa Neiva Leite Neyva Maria Lopes Romeiro Patrícia Amâncio da Rosa Patricia Dayane Carvalho Schaker Patricia Oehlmeyer Nassar Patrícia Savio de Araújo-Souza Patricia Silva Lucio Paulo Henrique Couto Souza Paulo Roberto Donadio Percy Nohama Quirino Alves de Lima Neto Rafael Deminice Rafael dos Santos Bezerra Raquel Alves dos Santos Renan Manozzo Galante Renata Ernlund Freitas de Macedo Rita de Cássia Garcia Simão Roberta Losi Guembarovski Roberto H. Herai Roberto Rosati Rodrigo Ferreira Rodrigo Rodrigues Matiello Rogério Neri Shinsato Rogério Pincela Mateus Rosane Aparecida Ribeiro Rosilene Fressatti Cardoso Rosilene Fressatti Cardoso Sandra Mara Guse Scós Venske Selene Elifio Esposito Sérgio Ossamu Ioshii Silvana Giuliatti Silvia Mara de Souza Halick Silvio Henrique Maia de Almeida Simone Neumann Wendt Spencer Luiz Marques Payão Stefan Wolanski Negrão Stephane Janaina de Moura Escobar Sueli Fumie Yamada Ogatta SUELI PERCIO QUINAIA Taciane Finatto Tatiana Mayumi Veiga Iriyoda Tayza Katelline Danilau Ostroski Tony Alexander Hild Valeria Valente Vanessa Nascimento Kozak Vanessa Santos Sotomaior Victor Breno Pedrosa Victoria Zeghbi Cochenski Borba Vivian Rotuno Moure Valdameri Wander Rogerio Pavanelli Weber Cláudio Francisco Nunes da Silva Willian Augusto de Melo Yohandra Reyes Torres |
| EPI_ISL_2812356, EPI_ISL_2812359, EPI_ISL_2812362, EPI_ISL_2812363, EPI_ISL_2812364, EPI_ISL_2812367, EPI_ISL_2812368, EPI_ISL_2812369, EPI_ISL_2812373, EPI_ISL_2812374, EPI_ISL_2812386, EPI_ISL_2812387, EPI_ISL_2812391, EPI_ISL_2812396, EPI_ISL_2812402, EPI_ISL_2812407, EPI_ISL_2812411, EPI_ISL_2812412, EPI_ISL_2812419, EPI_ISL_2812425, EPI_ISL_2812426, EPI_ISL_2812429, EPI_ISL_2841638 | | | |
| see above | UFS Virology | KRISP, KZn Research Innovation and Sequencing Platform | Giandhari Jennifer; Naidoo Yeshnee; Pillay Sureshnee; San James; Tegally Houriiyah; Tshabuila Derek; Wilkinson Eduan; Yajna Ramphal; de Oliveira Tulio |
| EPI_ISL_628760 | UHAS COVID-19 Lab | UHAS COVID-19 Lab | John O. Gyapong and the UHAS COVID-19 Lab Team; Jones Gyamfi; Kwabena O. Duedu; Reuben Ayivor-Djanie |
| EPI_ISL_747486, EPI_ISL_747487 | ULSS 5 Polesana | Istituto Zooprofilattico Sperimentale delle Venezie | Adelaide Milani; Alessia Schivo; Alice Fusaro; Ambra Pastori; Annalisa Salviato; Antonia Ricci; Bianca Zecchin; Calogero Terregino; Erika Giorgia Quaranta; Isabella Monne |
| EPI_ISL_1664743 | UMMC-Health | WHO National Influenza Centre Russian Federation | Andrey Komissarov; Anna Ivanova; Artem Fadeev; Daria Danilenko; Dmitry Lioznov; Elena Nabieva; Georgii Bazykin; Ksenia Safina; Kseniya Komissarova; Maria Pisareva; Maria Timofeeva; Mikhail Bakaev; Tamila Musaeva; Tatiana Platonova; Veronika Eder |
| EPI_ISL_2092861 | UNC Charlotte COVID-19 Testing Lab | UNC Charlotte Environmental Monitoring Laboratory | Angelica Martins; Cynthia Gibas; Jannatul Ferdous; Jessica Schlueter; Kevin Lambirth; Visva Barua |
| EPI_ISL_2645959, EPI_ISL_2989027, EPI_ISL_2989037 | UNILABS | Instituto Nacional de Saude (INSA) | Borges et al |
| EPI_ISL_2796010 | UNILABS | Instituto Nacional de Saude (INSA) and Instituto Gulbenkian de Ciencia (IGC) | Borges et al |
| EPI_ISL_853342 | UPMC Clinical Microbiology Laboratory | Microbial Genome Sequencing Center; Microbial Genomic Epidemiology Laboratory | Chinelo Ezeonwuku; Dan Snyder; Jane W. Marsh; Kady D. Waggle; Lee H. Harrison; Marissa P. Griffith; Mustapha M. Mustapha; Stephanie L. Mitchell; Vatsala R. Srinivasa; Vaughn S. Cooper |
| EPI_ISL_1137240, EPI_ISL_1732367 | UPMC Clinical Microbiology Laboratory | Microbial Genome Sequencing Center; Microbial Genomic Epidemiology Laboratory, University of Pittsburgh | Daniel J. Snyder; Jane W. Marsh; Kady D. Waggle; Lee H. Harrison; Marissa P. Griffith; Stephanie L. Mitchell; Vatsala R. Srinivasa; Vaughn S. Cooper |
| EPI_ISL_983464, EPI_ISL_1227749, EPI_ISL_1227750, EPI_ISL_2278770 | URMC LABS | Wadsworth Center, New York State Department of Health | Alexis Russel; Alexis Russell; Catharine Prussing; Daryl M. Lamson; Erasmus Schneider; Erica Lasek-Nesselquist; John Kelly; Jonathan Plitnick; Kirsten St. George; Matthew Shudt; Melissa A Leisner; Navjot Singh |
| EPI_ISL_2383566, EPI_ISL_2661466 | US Air Force School of Aerospace Medicine | US Air Force School of Aerospace Medicine | Amanda Javorina; Anthony Fries; Carol Garrett; Clarise Starr; Cole Anderson; Elizabeth Macias; Fritz Castillo; Jennifer Meyer; Sarah Purves; William Gruner |
| EPI_ISL_436045 | US VI Department of Health | Pathogen Discovery, Respiratory Viruses Branch, Division of Viral Diseases, Centers for Disease Control and Prevention | Anna Uehara; Bettina Bankamp; Clinton R. Paden; Haibin Wang; Jing Zhang; Krista Queen; Suxiang Tong; Yan Li; Ying Tao; Zachary Weiner |
| EPI_ISL_903891 | UT-Unified State Labs: Public Health Utah DOH | Genomics and Discovery, Respiratory Viruses Branch, Division of Viral Diseases, Centers for Disease Control and Prevention | Anna Montmayeur; Anna Uehara; Ben L. Rambo-Martin; Clinton R. Paden; Dhwani Batra; Haibin Wang; Jasmine Padilla; Jing Zhang; Justin Lee; Krista Queen; Lori Rowe; Mark Burroughs; Mili Sheth; Peter W. Cook; Rachel Marine; Sarah Nobles; Suxiang Tong; Yan Li; Ying Tao |
| EPI_ISL_424229, EPI_ISL_1324125, EPI_ISL_1621123, EPI_ISL_1785784, EPI_ISL_2194129, EPI_ISL_2773932 | UW Virology Lab | UW Virology Lab | Alexander Greninger; Hong Xie; Keith Jerome; Keith R Jerome; Lasata Shrestha; Margaret Mills; Meei-Li Huang; Michelle Lin; Nathan Breit; Noah Baker; Noah R. Baker; Patrick Mathias; Pavitra Roychoudhury; Ricardo Perez; Robert J. Livingston; Saraswathi Sathees; Sean Ellis; Shah Mohamed Bakhash; Tien V. Nguyen |
| EPI_ISL_734804, EPI_ISL_734899, EPI_ISL_735130 | UZ Leuven, National Reference Laboratory for Coronaviruses, Laboratory Medicine, Leuven, Belgium | KU Leuven, Rega Institute, Clinical and Epidemiological Virology | Bert Vanmechelen; Joan Marti-Carerras; Piet Maes; Tony Wawina-Bokalanga |
| EPI_ISL_737958, EPI_ISL_738036 | Uganda Central Public Health Lab and Uganda Virus Research Institute | MRC/UVRI & LSHTM Uganda Research Unit | Dan Lule Bugembe; Matthew Cotten; My V.T. Phan; Pontiano Kaleebu et al. |
| EPI_ISL_451189 | Uganda Virus Research Institute | MRC/UVRI & LSHTM Uganda Research Unit | Beatrice Dhaala; Dan Lule Bugembe; Deogratius Ssemwanga; Henry Kyobe; Henry Mwebesa; Jane Aceng; John Kayiwa; Jonas Lexow; Julius Lutwama; Matthew Cotten; My V.T Phan; Phionah Tushabe; Pontiano Kaleebu; Stephen Balinandi |
| EPI_ISL_2828019 | Unidad de Investigación Molecular (UNIMOL) | Instituto Nacional de Salud- Dirección de Investigación en Salud Pública | Carlos Franco-Muñoz; Carmen Osorio; Diana Malo; Diego A. Álvarez-Díaz; Diego Andrés Prada; Gerardo Santamaría; Hector Alejandro Ruiz-Moreno; Jhonnatan Reales-González; Jorge Rivera; Juan Camilo Martinez; Julian Naizaque; Katherine Laiton-Donato; Lisseth Pardo; Magdalena Wiesner; Marcela Mercado-Reyes; Maria T. Herrera-Sepúlveda; Marta Lopez Blanco; Martha Lucia Ospina Martinez; Paola Rojas; Sergio Gomez; Sheryll Corchuelo; Ángela Alarcon Cruz |
| EPI_ISL_1640127 | Uniklinikum Carl Gustav Carus an der TU Dresden; Institut fÃ¼r Virologie | Robert Koch Institute | |
| EPI_ISL_549175, EPI_ISL_1317690, EPI_ISL_1490459, EPI_ISL_2509362, EPI_ISL_2509368, EPI_ISL_2509369, EPI_ISL_3031028 | | | |
| see above | Unilabs Laboratory Medicine | Norwegian Institute of Public Health, Department of Virology | Atiya R Ali; Debech Nadia; Engebretsen Serina Beate; Garcia Llorente Ignacio; Hilde Elshaug; Hilde Synnøve Vollan; Hilde Vollan; Jon Bråte; Kamilla Heddeland Instefjord; Karoline Bragstad; Kathrine Stene-Johansen; Line Victoria Moen; Marie Paulsen Madsen; Olav Hungnes; Pedersen Benedikte Nevjen; Rasmus Riis Kopperud |
| EPI_ISL_1299863, EPI_ISL_1299864 | Unit of lab surveillance of viral emerging diseases, National Lab of Influenza | Respiratory Virus Unit, National Infection Service, Public Health England | Iris Hasibra; PHE Covid Sequencing Team; Prof Albana Fico; Prof Silvia Bino |
| EPI_ISL_812548, EPI_ISL_831801 | United States Air Force School of Aerospace Medicine | United States Air Force School of Aerospace Medicine | Amanda Javorina; Anthony Fries; Clarise Starr; Elizabeth Macias; Jennifer Meyer; Sarah Purves; William Gruner |
| EPI_ISL_569963, EPI_ISL_570008, EPI_ISL_2823743 | Unity Health Toronto | Ontario Institute for Cancer Research | Bernard Lam; Felicia Vincelli; Ilinca Lungu; Jared T. Simpson; Jeremy Johns; Karel Boissinot; Larissa M. Matukas; Le Luu; Mark Downing; Paul Krzyzanowski; Philip Zuzarte; Ramzi Fattouh; Richard de Borja; Samira Mubareka; TIBDN; Trina Otterman; Wai Sum Siu; Yan Chen; Zhi Cui |
| EPI_ISL_888804 | Univeristy of New Mexico Hospital | Center for Global Health, University of New Mexico Health Sciences Center | Darrell Dinwiddie; Daryl Domman; Jon Femling; Justin Bacca; Kurt Schwalm |
| EPI_ISL_3024078 | Universidad Autonoma de Yucatan | New York Genome Center | Amy Baldwin; Andre Corvelo; Aviles-Gomez E; Ayora-Talavera G; Chan-Gasca M; Dayna M. Oschwald; Flores-Quintal F; Granja-Perez P; Lopez-Coral L.; Michael Zody; Samantha Fennessey; Tom Maniatis; Villanueva-Jorge S; Yam-Pool E |
| EPI_ISL_697796 | Universidad Regional Amazonica IKIAM | Institute of Microbiology, Universidad San Francisco de Quito | Andrea Carrera; Belén Prado-Vivar; Bernardo Gutiérrez; Carolina Proaño-Bolaños; Fabian Aguilar; Gabriel Trueba; Giovanna Moran; Juan José Guadalupe; Katherine Apunte; Marcelo Ortiz; Michelle Grunauer; Monica Becerra-Wong; Nina Espinoza de los Monteros; Patricio Rojas-Silva; Paúl Cárdenas; Sonia Sislema; Sully Márquez; Verónica Barragán; Yeimy Rojas |
| EPI_ISL_1656146 | Universidad Tecnica Particular de Loja | Institute of Microbiology, Universidad San Francisco de Quito | Belén Prado-Vivar; Bernardo Gutiérrez; David Zuñiga; Fernando Serrano; Gabriel Trueba; Juan José Guadalupe; Katherine Ojeda; Luis Flores; Melissa Ortega; Michelle Grunauer; Monica Becerra-Wong; Paola Dalgo; Patricio Rojas-Silva; Paúl Cárdenas; Raiza Briceño; Sully Márquez; Verónica Barragán |
| EPI_ISL_2123875 | Universitatsklinikum Koln; Institut fur Virologie | Robert Koch Institute | |
| EPI_ISL_812967 | University Clinical Research Center, University of Sciences | University Clinical Research Center, University of Sciences | A.A.; Bane, S.; Dao, S.; Diakite, M.; Diarra, B.; Doumbia, S.; Guindo, I.; Iknane; Kone, A. |
| EPI_ISL_2895002 | University College Hospital, Ibadan | African Centre of Excellence for Genomics of Infectious Diseases (ACEGID), Redeemer's University, Nigeria | A.T.; Abechi; Ajogbasile; Akano; C.A.; C.T.; Eromon; F.V.; Folarin, O.; Fowotade, A.; Happi; I.B.; J.N.; J.U.; K.O.; Kayode; Nosamiefan, I.; Oguzie; Olawoye; Olumade; Oluniyi; P.E.; P.S.; T.J.; Ugwu; Uwanibe |
| EPI_ISL_741244, EPI_ISL_865865 | University College London, Great Ormond Street Hospital for Children NHS Foundation Trust, Imperial College Healthcare NHS Trust | COVID-19 Genomics UK (COG-UK) Consortium | Alison Holmes; Charlotte Williams; Helena Tutill; Jacqueline Findlay; James Price; Judith Breuer; Julianne Brown; Kathryn Harris; Leysa Forrest; Mark Kristiansen; Paola Niola; Paola Resende Silva; Patricia Dyal; Paul Randell; Rachel Williams; Samuel Weeks; Sergi Castellano; Sunando Roy; Tony Brooks; Yasmin Panchbhaya |
| EPI_ISL_982819, EPI_ISL_982938, EPI_ISL_982943, EPI_ISL_982954, EPI_ISL_982981, EPI_ISL_983011, EPI_ISL_983187 | | | |
| see above | University Health Network/Mount Sinai Hospital Department of Microbiology | Ontario Institute for Cancer Research | Aimee Paterson; Allison McGeer; Angel Liu; Bernard Lam; Cassandra Bergwerff; Ilinca Lungu; Jared T. Simpson; Javier Hernandez; Jeff Wrana; Jessica Bourke; Kin Chan; Kuganya Nirmalarajah; Laurence Pelletier; Lawrence E. Heisler; Lubaina Kothari; Marc Mazzulli; Marie-Ming Aynaud; Michael Laszloffy; Patryk Aftanas; Paul Krzyzanowski; Richard de Borja; Samira Mubareka; Seda Barutcu; Tony Mazzulli |
| EPI_ISL_710555, EPI_ISL_710559, EPI_ISL_710570 | University Hospital Dubrava | Ruđer Boškovic Institute; Forensic Science Centre Ivan Vučetić; University of Zagreb Faculty of Science | Ana Livun; Antonela Blažeković; Boris Maček; Danilo Licastro; Dunja Glavaš; Fran Borovečki; Fuad Ćosović; Gordana Maravić Vlahoviček; Ivan Šamija; Ivana Ćelap; Jasna Kašman; Josipa Skelin; Katarina Marija Tupek; Kristian Vlahoviček; Kristina Gotovac Jerešić; Lidija Cvetko-Krajinović; Lucija Basić; Lucija Markulin; Maja Kuzman; Marina Korolija; Mario Štefanović; Mirjana Domazet-Lošo; Paula Štancl; Petra Vrabec; Robert Beluzić; Rosa Karlić; Sanja Tadinac; Senčica Pejša; Tomislav Domazet-Lošo; Valentina Đumlijan-Combaj; Vjekoslav Tomaić; Vladimir Krajinović; Željka Maček Šafranko |

| | | | |
|---|---|---|---|
| EPI_ISL_2790388 | University Hospital Foca | Public Health Institute of Republic of Srpska | Branka Culibrk; Dijana Vukajlovic; Milica Celic; Pava Dimitrijevic; Stanka Tomic; Tatjana Markovic; Zeljka Sumic |
| EPI_ISL_681761 | University Hospital Limerick | Irish Coronavirus Sequencing Consortium - Teagasc Moorepark | Amy Fitzpatrick; Calum Walsh; Carolyn Meaney; Elaine Lawton; Fiona Crispie; John Kenny; Patrick Stapleton; Paul Cotter |
| EPI_ISL_1254573, EPI_ISL_2357760 | University Hospital of Northern Norway, Department for Microbiology and Infectious Disease Control | Norwegian Institute of Public Health, Department of Virology | Atiya R Ali; Debech Nadia; Engebretsen Serina Beate; Garcia Llorente Ignacio; Hilde Elshaug; Hilde Vollan; Jon Bråte; Kamilla Heddeland Instefjord; Karoline Bragstad; Kathrine Stene-Johansen; Line Victoria Moen; Marie Paulsen Madsen; Olav Hungnes; Pedersen Benedikte Nevjen; Rasmus Riis Kopperud |
| EPI_ISL_429207 | University Hospitals of Geneva Laboratory of Virology | University Hospitals of Geneva Laboratory of Virology | Laubscher F. |
| EPI_ISL_897794 | University Hospitals of Geneva, Laboratory of Virology | HUG, Laboratory of Virology and the Health2030 Genome Center | Ana Rita Goncalves; Deborah Penet; Emmanouil Dermitzakis; Henri Pegeot; Ioannis Xenarios; Keith Harshman; Laurent Kaiser; Lorenzo Cerutti; Melyssa Elies; Samuel Cordey |
| EPI_ISL_2384717 | University Medical Center Hamburg Eppendorf | Heinrich Pette Institute, Leibniz Institute for Experimental Virology | Adam Grundhoff; Alexis Robitaille; Johannes Knobloch; Martin Aepfelbacher; Nicole Fischer; Thomas Günther |
| EPI_ISL_2931002 | University of Arkansas for Medical Sciences | Center for Global Health, University of New Mexico Health Sciences Center | Catherine Kirkpatrick; Darrell Dinwiddie; Daryl Domman; David Ussery; Jennifer Laudadio; Joshua L. Kennedy; Kurt Schwalm; Susan Harley; Uday Chalwadi; Valerie Morley |
| EPI_ISL_1072985 | University of Balamand | Microbial Genomics Lab, Lebanese American University, Byblos | Mira El Chaar; Sima Tokajian; Youssef Bassim |
| EPI_ISL_2100503 | University of Health Sciences | Quadram Institute Bioscience | Alam Khan; Alexander J Trotter; Alison E. Mather; Almina Shafiq; Alp Aydin; Ana P. Tedim; Anastasia Kolyva; Andrew Bell; Andrew J. Page; Claire Stuart; Dave J. Baker; Gemma L. Kay; Javed Akram; John Wain; Justin O'Grady; Leonardo de Oliveira Martins; Lizzie Meadows; Maria Diaz; Mark Webber; Muhammad Bilal Sarwar; Muhammad Roman; Muhammed Yasir; Nabil-Fareed Alikhan; Nadeem Afzal; Ngozi Elumogo; Nicholas M. Thomson; Rachael Stanley; Rachel Gilroy; Reenesh Prakash; Samir Dervisevic; Samuel Bloomfield; Shah Jahan; Sidra-tul-muntaha; Steven Rudder; Thanh Le-Viet |
| EPI_ISL_2864611 | University of Health and Allied Sciences (UHAS) COVID-19 Testing and Research Centre | University of Health and Allied Sciences (UHAS) COVID-19 Testing and Research Centre | John O. Gyapong and the UHAS COVID-19 Lab Team; Jones Gyamfi; Kwabena O. Duedu; Reuben Ayivor-Djanie |
| EPI_ISL_2500011, EPI_ISL_2728657 | University of Liège COVID-19 testing center | GIGA Medical Genomics | Bouchra Boujemla; Claire Gourzonès; Cécile Meex; Fabrice Bureau; Keith Durkin; Laurent Gillet; Maria Artesi; Marie-Pierre Hayette; Nadine Cambisano; Nathalie Renotte; Olivier Ek; Sébastien Bontems; Vincent Bours; Wouter Coppieters |
| EPI_ISL_2242506, EPI_ISL_2243223 | University of Maiduguri Teaching Hospital | International Centre for Genetic Engineering and Biotechnology (ICGEB) and ARGO Open Lab for Genome Sequencing | Alessandro Marcello; Bamidele S Oderinde; Danilo Licastro; Emanuele Orsini; Galadima Gadzama; Marycelin M Baba; Monilade Akinola; Simeone Dal Monego; Zara Wuduri |
| EPI_ISL_1118703 | University of Michigan Clinical Microbiology Laboratory | Lauring Lab, University of Michigan, Department of Microbiology and Immunology | Valesano |
| EPI_ISL_955142, EPI_ISL_955144, EPI_ISL_955148, EPI_ISL_955170, EPI_ISL_1016690, EPI_ISL_1016822 | University of Sarajevo, Veterinary Faculty, Laboratory for Molecular Diagnostic and Research Laboratory | University of Sarajevo, Veterinary Faculty, Laboratory for Molecular Diagnostic and Research Laboratory | Alic-Seho A.; Alić-Šeho A.; Goletic S.; Goletic T.; Goletić T.; Goletić Š.; Hodzic A.; Hodžić A.; Jazic A.; Jažić A.; Nicevic M.; Nicević M.; Sabic E.; Softic A.; Softić A.; Terzic I.; Terzić I.; Terzić I. Jažić A.; Šabić E. |
| EPI_ISL_476078, EPI_ISL_677765 | University of Szeged, Institute of Clinical Microbiology | National Laboratory of Virology, Szentágothai Research Centre | Balázs Somogyi; Brigitta; Brigitta Zana; Endre Gábor Tóth; Ferenc Jakab; Gabriella Terhes; Gábor Kemenesi; Terhes Gabriella |
| EPI_ISL_485391, EPI_ISL_485395 | University of Ulsan College of Medicine and Asan Medical Center | University of Ulsan College of Medicine and Asan Medical Center | Heungsup Sung; Jaewoong Lee; Jina Lee; Jiwon Jung; Jongsik Chun; Kihyun Lee; Kuenyoul Park; Kyu-Hwa Hur; Mauricio Chalita; Mi-Na Kim; Seok-Hwan Yoon; Sung-Han Kim; and Hae Kyung Lee |
| EPI_ISL_509950, EPI_ISL_1822080 | University of Wisconsin-Madison AIDS Vaccine Research Laboratories | University of Wisconsin-Madison AIDS Vaccine Research Laboratories | Gage Moreno; Katarina Braun; et al. AIDS Vaccine Research Laboratories |
| EPI_ISL_977291, EPI_ISL_977342, EPI_ISL_977350, EPI_ISL_977357, EPI_ISL_977367, EPI_ISL_977373, EPI_ISL_977421, EPI_ISL_977422, EPI_ISL_2803362, EPI_ISL_2803437, EPI_ISL_2803449, EPI_ISL_2803512, EPI_ISL_2803529, EPI_ISL_2803584, EPI_ISL_2803629, EPI_ISL_2803630, EPI_ISL_2803631, EPI_ISL_2803632, EPI_ISL_2803635, EPI_ISL_2803636, EPI_ISL_2803638, EPI_ISL_2803639, EPI_ISL_2803645, EPI_ISL_2803646, EPI_ISL_2803648, EPI_ISL_2803666, EPI_ISL_2803668, EPI_ISL_2803673, EPI_ISL_2803674, EPI_ISL_2803685, EPI_ISL_2803687, EPI_ISL_2803693, EPI_ISL_2803694, EPI_ISL_2803698, EPI_ISL_2803701, EPI_ISL_2803702, EPI_ISL_2803709, EPI_ISL_2803714, EPI_ISL_2803716, EPI_ISL_2803718, EPI_ISL_2803729, EPI_ISL_2803730, EPI_ISL_2803732 | | | |
| see above | University of Zambia, School of Veterinary Medicine | UNZAVET and PATH | Daniel Bridges; Mulenga Mwenda-Chimfwembe; Ngonda Saasa; ZNPHI and ZGSC |
| EPI_ISL_1721045 | UniversitÃ¤tsklinikum des Saarlandes - Institut fÃ¼r Virologie | Robert Koch Institute | |
| EPI_ISL_2382023, EPI_ISL_2382028, EPI_ISL_2382031, EPI_ISL_2382032 | Universität Innsbruck, Institut für Mikrobiologie | Bergthaler laboratory, CeMM Research Center for Molecular Medicine of the Austrian Academy of Sciences | Andreas Bergthaler; Anna Schedl; Bekir Erguner; Benedikt Agerer; Christoph Bock; Fabian Amman; Jan Laine; Lukas Endler; Maelle Le Moing; Martin Senekowitsch; Michael Schuster; Petr Triska; Thomas Penz |
| EPI_ISL_1286241 | Universitätsklinikum Heidelberg | Robert Koch Institute | |
| EPI_ISL_628939, EPI_ISL_1313869, EPI_ISL_2279888, EPI_ISL_2290332, EPI_ISL_2290425, EPI_ISL_2679594, EPI_ISL_2856552, EPI_ISL_2857105 | | | |
| see above | Utah Public Health Laboratory | Utah Public Health Laboratory | Erin L. Young; Erin Young; Kelly F. Oakeson; Kelly Oakeson; Olinto Linares-Perdomo; Tara Gallagher |
| EPI_ISL_2382667 | VERMAAK | National Institute for Communicable Diseases of the National Health Laboratory Service | Amoako DG; Bhiman JN; Ismail A; Mahlangu B; Mohale T; Ntuli N; Scheepers C |
| EPI_ISL_2662532, EPI_ISL_2662536 | VERMAAK LABORATORY | National Institute for Communicable Diseases of the National Health Laboratory Service | Amoako DG; Bhiman JN; Everatt J; Ismail A; Mahlangu B; Mnguni A; Mohale T; Ntuli N; Scheepers C |
| EPI_ISL_1094245 | VT Dept. of Health Laboratory | Respiratory Viruses Branch, Division of Viral Diseases, Centers for Disease Control and Prevention | Anna Montmayeur; Anna Uehara; Ben L. Rambo-Martin; Clinton R. Paden; Dhwani Batra; Haibin Wang; Jasmine Padilla; Jing Zhang; Justin Lee; Krista Queen; Lori Rowe; Mark Burroughs; Mili Sheth; Peter W. Cook; Rachel Marine; Sarah Nobles; Suxiang Tong; Yan Li; Ying Tao |
| EPI_ISL_2494898 | Vaccines and Infectious Diseases Analytics Research Unit (VIDA) | KRISP, KZN Research Innovation and Sequencing Platform | Baillie Vicky; Giandhari Jennifer; Madhi Shabir; Naidoo Yeshnee; Pillay Sureshnee; San James; Tegally Houriiyah; Wilkinson Eduan; de Oliveira Tulio; du Plessis Jeanine |
| EPI_ISL_2360774, EPI_ISL_2360878, EPI_ISL_2494391, EPI_ISL_2494501, EPI_ISL_2494557, EPI_ISL_2494585 | Vaccines and Infectious Diseases Analytics Research Unit (VIDA) | KRISP, KZn Research Innovation and Sequencing Platform | Baillie Vicky; Giandhari Jennifer; Madhi Shabir; Naidoo Yeshnee; Pillay Sureshnee; San James; Tegally Houriiyah; Tshabuila Derek; Wilkinson Eduan; de Oliveira Tulio; du Plessis Jeanine |
| EPI_ISL_2521377, EPI_ISL_3016436 | Valais Hospital, Central Institute | Valais Hospital, Central Institute | Alexis Dumoulin; Deborah Penet; Emmanouil Dermitzakis; Henri Pegeot; Ioannis Xenarios; Keith Harshman; Lorenzo Cerutti; Melyssa Elies |
| EPI_ISL_1373898 | Vanderbilt University Medical Center | Pathogen Discovery, Respiratory Viruses Branch, Division of Viral Diseases, Centers for Disease Control and Prevention | Anna Kelleher; Anna Uehara; Brian Lynch; Clinton R. Paden; Haibin Wang; Han Jia Justin Ng; Jing Zhang; Krista Queen; Peter Cook; Suxiang Tong; Yan Li; Ying Tao |
| EPI_ISL_640096 | Vanguard CHC wc VGC | NHLS/UCT | Arash Iranzadeh; Bruna Galvao; Carolyn Williamson; Deelan Doolabh; Diana Hardie; Innocent Mudau; Kruger Marais; Lynn Tyers; Marvin Hsiao; Stephen Korsman |
| EPI_ISL_2582736 | Vermaak & Partners Laboratory | National Institute for Communicable Diseases of the National Health Laboratory Service | Amoako DG; Bhiman JN; Everatt J; Ismail A; Mahlangu B; Mnguni A; Mohale T; Ntuli N; Scheepers C |
| EPI_ISL_2001620 | Vestfold Hospital, Toensberg Department of Microbiology | Norwegian Institute of Public Health, Department of Virology | Atiya R Ali; Debech Nadia; Engebretsen Serina Beate; Garcia Llorente Ignacio; Hilde Elshaug; Hilde Vollan; Jon Bråte; Kamilla Heddeland Instefjord; Karoline Bragstad; Kathrine Stene-Johansen; Marie Paulsen Madsen; Olav Hungnes; Pedersen Benedikte Nevjen; Rasmus Riis Kopperud |
| EPI_ISL_678490 | Veterinary Specialized Institute "Sabac", Serbia | Veterinary Specialized Institute "Kraljevo", Serbia | Afonso, C.; Banovic Djeri, B.; Jankovic, M.; Jovanovic, T.; Knezevic, A.; Mrkovacki, S.; Petrovic, T.; Sekler, M.; Tesovic, B.; Vidanovic, D.; Volkening, J. |
| EPI_ISL_833517 | Veterinary Specialized Instute "Nis" | Veterinary Specialized Institute "Kraljevo", Serbia | Afonso, C.; Banovic Djeri, B.; Jankovic, M.; Jovanovic, T.; Knezevic, A.; Manic, M.; Petrovic, M.; Petrovic, T.; Sekler, M.; Tesovic, B.; Vidanovic, D.; Volkening, J. |
| EPI_ISL_2227287 | Veterinary institute in Zvolen, Pod Drahami 918, 960 86 Zvolen Slovakia | Veterinary institute in Zvolen, Pod Drahami 918, 960 86 Zvolen Slovakia | Dirbáková Z.; Mojžiš M.; Mokryšová S.; Sujová S.; Tinák M. |
| EPI_ISL_1040814, EPI_ISL_2140693, EPI_ISL_2375993 | Victoria Hospital wc VHW | NHLS/UCT | Arash Iranzadeh; Bruna Galvao; Carolyn Williamson; Deelan Doolabh; Diana Hardie; Innocent Mudau; Kruger Marais; Lynn Tyers; Marvin Hsiao; Stephen Korsman |
| EPI_ISL_426857, EPI_ISL_456455, EPI_ISL_456565 | Victorian Infectious Diseases Reference Laboratory (VIDRL) | Microbiological Diagnostic Unit Public Health Laboratory and Victorian Infectious Diseases Reference Laboratory, Doherty Institute | Caly L.; Druce J.; Sait, M.; Schultz M.; Seemann T.; Sherry, N. |
| EPI_ISL_521971, EPI_ISL_640784, EPI_ISL_779621, EPI_ISL_779622, EPI_ISL_877578, EPI_ISL_1913202, EPI_ISL_1913204, EPI_ISL_1913205, EPI_ISL_1913206, EPI_ISL_1913211, EPI_ISL_2920928, EPI_ISL_2920932, EPI_ISL_2920946, EPI_ISL_2920948, EPI_ISL_2920952, EPI_ISL_2920955, EPI_ISL_2920960, EPI_ISL_2920963, EPI_ISL_2920974, EPI_ISL_2920989, EPI_ISL_2920990, EPI_ISL_2920998, EPI_ISL_2978349, EPI_ISL_3030415 | | | |
| see above | Victorian Infectious Diseases Reference Laboratory (VIDRL) | VIDRL and MDU-PHL | Caly L.; Druce J.; M.L.; N.L.; Sait; Sait, M.; Schultz M.; Schultz M.B.; Seemann T.; Sherry; Sherry, N. |
| EPI_ISL_416410, EPI_ISL_416411, EPI_ISL_419733, EPI_ISL_419735 | Victorian Infectious Diseases Reference Laboratory (VIDRL) | Victorian Infectious Diseases Reference Laboratory and Microbiological Diagnostic Unit Public Health Laboratory, Doherty Institute | Caly L.; Druce J.; Sait, M.; Schultz M.; Seemann T.; Sherry, N.; Taiaroa, G. |
| EPI_ISL_2694573 | Viesoji istaiga Respublikine Siauliu ligonine | National Public Health Surveillance Laboratory | Ana Steponkiene; Danas Baksa; Jelena Razmuk; Lukas Vasionis; Lukas Zemaitis; Migle Gabrielaite; Svajune Muralyte |

| | | | |
|---|---|---|---|
| EPI_ISL_2339638, EPI_ISL_2510724 | Viesoji istaiga Vilniaus universiteto ligonine Santaros klinikos | Vilnius University Hospital Santaros Klinikos, Center of Laboratory Medicine | Daniel Naumovas; Dovile Ezerskyte; Gytis Dudas; Ingrida Olendraite; Laimonas Griskevicius; Ligita Raugaite; Mindaugas Stoskus; Monika Katenaite; Rimvydas Norvilas |
| EPI_ISL_2779343 | Vihiga County referral hospital | USAMRD-A, Basic Science Laboratory | Alan Lemtudo; Beth Mutai; Brian Andika; Carol Kifude; Clement Masakwe; Eric Muthanje; Esther Omuseni; Faith Sigei; Gathii Kimita; George Awinda; John Waitumbi; Josphat Nyataya; Rachel Githii; Rehema Liyai; Stephen Ochola |
| EPI_ISL_2802125 | Villiersdorp Clinic wc VDP | NHLS/UCT | Arash Iranzadeh; Bruna Galvao; Carolyn Williamson; Deelan Doolabh; Diana Hardie; Gert Marais; Innocent Mudau; Lynn Tyers; Marvin Hsiao; Stephen Korsman |
| EPI_ISL_904938, EPI_ISL_904952, EPI_ISL_904953 | Vilnius University Hospital Santaros Klinikos, Vilnius University | Institute of Biotechnology, Life Sciences Center, Vilnius University | Albertas Timinskas; Alma Gedvilaite; Aurelija Zvirbliene; Daniel Naumovas; Emilija Vasiliunaite; Laimonas Griskevicius; Milda Norkiene |
| EPI_ISL_560398, EPI_ISL_560405 | Vilnius University Hospital Santaros Klinikos, Vilnius University | Institute of Biotechnology, Life Sciences Center, Vilnius University and Thermo Fisher Scientific | Albertas Timinskas; Alma Gedvilaite; Aurelija Zvirbliene; Daniel Naumovas; Justinas Slikas; Laimonas Griskevicius; Ligita Jancioriene; Mindaugas Paulauskas |
| EPI_ISL_933832, EPI_ISL_933912, EPI_ISL_934070, EPI_ISL_1005262, EPI_ISL_1005284, EPI_ISL_1005354 | Vilnius university hospital Santaros Klinikos, Center of Laboratory Medicine | Vilnius university hospital Santaros Klinikos, Center of Laboratory Medicine | Daniel Naumovas; Dovile Ezerskyte; Gytis Dudas; Ingrida Olendraite; Justinas Slikas; Laimonas Griskevicius; Rimvydas Norvilas |
| EPI_ISL_1658231, EPI_ISL_1914247, EPI_ISL_1914248 | Viollier AG | Department of Biosystems Science and Engineering, ETH Zurich | Andrea Patrignani; Andreia Cabral de Gouvea; Catharine Aquino; Chaoran Chen; Christian Beisel; Christiane Beckmann; Christoph Noppen; David Dreifuss; Doris Popovic; Elodie Burcklen; Griffin White; Ina Nissen; Ivan Topolsky; Jay Tracy; Katharina Jahn; Lara Fuhrmann; Laura Neff; Lennart Opitz; Maria Domenica Moccia; Maurice Redondo; Mirjam Feldkamp; Natascha Santacroce; Niko Beerenwinkel; Noemie Santamaria de Souza; Olivier Kobel; Philipp Jablonski; Ralph Schlapbach; Rebecca Denes; Sarah Nadeau; Simon Gruter; Sophie Seidel; Tanja Stadler; Timothy Sykes |
| EPI_ISL_451864, EPI_ISL_486506, EPI_ISL_500907, EPI_ISL_693877, EPI_ISL_721718, EPI_ISL_721726, EPI_ISL_737488, EPI_ISL_737507, EPI_ISL_737516, EPI_ISL_737673, EPI_ISL_737925, EPI_ISL_737926, EPI_ISL_767993, EPI_ISL_899061, EPI_ISL_899109, EPI_ISL_899362, EPI_ISL_1001507, EPI_ISL_1002753, EPI_ISL_1002759, EPI_ISL_1002868, EPI_ISL_1003152, EPI_ISL_1003836, EPI_ISL_1004279, EPI_ISL_1059306, EPI_ISL_1119265, EPI_ISL_1119341, EPI_ISL_1119430, EPI_ISL_1119431, EPI_ISL_1130149, EPI_ISL_1130743, EPI_ISL_1130884, EPI_ISL_1195147, EPI_ISL_1195162, EPI_ISL_1195173, EPI_ISL_1259963, EPI_ISL_1260061, EPI_ISL_1260151, EPI_ISL_1260423, EPI_ISL_1406785, EPI_ISL_1407469, EPI_ISL_1408298, EPI_ISL_1408489, EPI_ISL_1408490, EPI_ISL_1408791, EPI_ISL_1496311, EPI_ISL_1597912, EPI_ISL_1750693, EPI_ISL_1750789, EPI_ISL_2152242, EPI_ISL_2211587, EPI_ISL_2307509, EPI_ISL_2308029, EPI_ISL_2375082, EPI_ISL_2375120, EPI_ISL_2662389, EPI_ISL_2820594, EPI_ISL_2820604, EPI_ISL_2938399, EPI_ISL_3007533 | | | |
| see above | Viollier AG | Department of Biosystems Science and Engineering, ETH Zürich | Andrea Patrignani; Andrea Patrizia Salzmann; Andreia Cabral de Gouvea; Catharine Aquino; Chaoran Chen; Christian Beisel; Christiane Beckmann; Christoph Noppen; Daniel Ehrsam; David Dreifuss; Deborah Penet; Doris Popovic; Elodie Burcklen; Emmanouil Dermitzakis; Griffin White; Henri Pegeot; Henriette Kurth; Ina Nissen; Ioannis Xenarios; Isabel Stürmer; Ivan Topolsky; Jay Tracy; Katharina Jahn; Keith Harshman; Kim Philipp Jablonski; Lara Fuhrmann; Laura Neff; Lennart Opitz; Lorenzo Cerutti; Maria Domenica Moccia; Maurice Redondo; Mirjam Feldkamp; Natascha Santacroce; Niko Beerenwinkel; Noemie Santamaria de Souza; Olivier Kobel; Pedro Ferreira; Philipp Jablonski; Ralph Schlapbach; Rebecca Denes; Sarah Nadeau; Simon Grüter; Sophie Seidel; Susana Posada-Céspedes; Tanja Stadler; Timothy Sykes; Tobias Schär |
| EPI_ISL_417946, EPI_ISL_420841, EPI_ISL_513597, EPI_ISL_514125, EPI_ISL_527492, EPI_ISL_527532, EPI_ISL_527543, EPI_ISL_527547, EPI_ISL_591086, EPI_ISL_965122, EPI_ISL_965123, EPI_ISL_965124, EPI_ISL_1623757, EPI_ISL_1785567, EPI_ISL_2135837, EPI_ISL_2135838, EPI_ISL_2135840, EPI_ISL_2135841, EPI_ISL_2135843, EPI_ISL_2135844, EPI_ISL_2135845, EPI_ISL_2135846, EPI_ISL_2966635, EPI_ISL_2966637, EPI_ISL_2966638, EPI_ISL_2966639, EPI_ISL_2966640, EPI_ISL_2966644, EPI_ISL_2966646, EPI_ISL_2966650, EPI_ISL_2966653, EPI_ISL_2968537, EPI_ISL_2968539, EPI_ISL_2968551, EPI_ISL_2968556, EPI_ISL_2968557, EPI_ISL_2968689, EPI_ISL_2968690, EPI_ISL_2968691, EPI_ISL_2968701, EPI_ISL_2968706, EPI_ISL_2968711, EPI_ISL_2968714, EPI_ISL_2968715, EPI_ISL_2968718, EPI_ISL_2968721, EPI_ISL_2968723, EPI_ISL_2968726, EPI_ISL_2968728, EPI_ISL_2968730, EPI_ISL_2982483, EPI_ISL_2982501, EPI_ISL_2982504, EPI_ISL_2982568, EPI_ISL_2982622 | | | |
| see above | Viral Respiratory Lab, National Institute for Biomedical Research (INRB) | Pathogen Sequencing Lab, National Institute for Biomedical Research (INRB) | Allison Black; Amuri Aziza; Andrew Rambaut; Catherine Pratt; Eddy Kinganda-Lusamaki; Edith Nkwembe; Emmanuel Lokilo Lofiko; Francisca Muyembe Mawete; Ian Goodfellow; James Hadfield; Jean Claude Makangara; Jean-Claude Makangara Cigolo; Jean-Jacques Muyembe Tamfum; Josh Quick; Kristian Andersen; Matthias Pauthner; Michael Wiley; Nick Loman; Placide Mbala-Kingebeni; Steve Ahuka-Mundeke; Trevor Bedford |
| EPI_ISL_528386 | Viral vaccines, VSVRI- Veterinary serum and vaccine research institute | Viral vaccines, VSVRI- Veterinary serum and vaccine research institute | A.A.; M.A.; Saad; Saleh |
| EPI_ISL_2344484 | Virginia Division of Consolidated Laboratory Services | Virginia Division of Consolidated Laboratory Services | Virginia DCLS |
| EPI_ISL_2694588 | Virology Centre, School of public health, Tehran University of Medical Sciences, Tehran, Iran | Genetics Research Center, University of Social Welfare and Rehabilitation Sciences | Hossein Najmabadi.; Jila Yavarian; Khadijeh Jalalvand; Kimia Kahrizi; Marzieh Mohseni; Talat Mokhtari-Azad; Zohreh Fattahi |
| EPI_ISL_2499903, EPI_ISL_2499907, EPI_ISL_2499909, EPI_ISL_2499913, EPI_ISL_2499916, EPI_ISL_2499917, EPI_ISL_2549198, EPI_ISL_2549199, EPI_ISL_2657327, EPI_ISL_2657329, EPI_ISL_2657330, EPI_ISL_2657332, EPI_ISL_2657333, EPI_ISL_2657334, EPI_ISL_2657335, EPI_ISL_2657336, EPI_ISL_2657337, EPI_ISL_2657338, EPI_ISL_2657339, EPI_ISL_2657341, EPI_ISL_2657343, EPI_ISL_2657344, EPI_ISL_2657345, EPI_ISL_2657346, EPI_ISL_2657347, EPI_ISL_2657352, EPI_ISL_2657353, EPI_ISL_2657354, EPI_ISL_2657355, EPI_ISL_2657356 | | | |
| see above | Virology Department, Central Health Laboratory | The Francis Crick Institute | Bahadoor BS; Crawford M; Daniels RS; Goldstone R; Harvey R; Manraj SS; Nicod J; Patel H; Ramuth M; Sonoo J |
| EPI_ISL_1104285 | Virology Department, Royal Infirmary of Edinburgh, NHS Lothian / School of Biological Sciences, University of Edinburgh | COVID-19 Genomics UK (COG-UK) Consortium | Colquhoun R; Cotton S; Dewar R; Hill V; Jackson B; McCrone JT; McHugh M; O'Toole Á; Rambaut A; Rooke S; Scher E; Templeton K; Yu X |
| EPI_ISL_473793 | Virology Department, Royal Infirmary of Edinburgh, NHS Lothian / School of Biological Sciences, University of Edinburgh / Institute of Genetics and Molecular Medicine, University of Edinburgh | COVID-19 Genomics UK (COG-UK) Consortium | Balcaza C; Colquhoun R; Dewar R; Gallagher M; Hill V; Jackson B; McCrone JT; McHugh M; O'Toole Á; Rambaut A; Rooke S; Scher E; Templeton K; Williams TC; Yu X |
| EPI_ISL_1191603, EPI_ISL_1191604, EPI_ISL_1191605, EPI_ISL_1827649, EPI_ISL_1827650, EPI_ISL_1827667, EPI_ISL_1827674, EPI_ISL_1827677, EPI_ISL_1827690 | | | |
| see above | Virology Department, Victoria Hospital, Plaine-Wilhems, Mauritius | National Institute for Communicable Diseases of the National Health Laboratory Service | Allam M; Amoako DG; Baboo SB; Bhiman JN; Ismail A; Mahlangu B; Manraj SS; Mohale T; Ntuli N; Ramuth M; Scheepers C; Sonoo J |
| EPI_ISL_636982 | Virology Lab, National Institute for Biomedical Research (INRB) | Project group Epidemology of Highly Pathogenic Microorganisms, Robert Koch-Institute | Eddy Kinganda-Lusamaki; Essia Belarbi; Fabian Leendertz; Gabriel Mbunsu; Grit Schubert; Jasmin Schlotterbeck; Jean-Jacques Muyembe Tamfum; Sheila Makiala; Steve Ahuka-Mundeke |
| EPI_ISL_2955336 | Virology Laboratories | Erasmus Medical Center | Abd Moniem Ain shoka; Amel Nagiub; Anne van der Linden; Annemiek van der Eijk; Bas Oude Munnink; Corine GeurtsvanKessel; Dalia Ramadan; David Nieuwenhuijse; Emmanuelle Munger; Galal Mahmoud; Irina Chestakova; Marion Koopmans; Marjan Boter; Mohamed Hassany; Mohamed K. Khalifa; Nancy El Guindy; Ramy Galal; Reina Sikkema; Richard Molenkamp; Salma Sayed; Shymaa A. Showky; Wael H. Roshdy; on behalf of the Dutch national COVID-19 response team. |
| EPI_ISL_2960150 | Virology Laboratories | Erasmus Medical Center Department of Virology | Abd Moniem Ain shoka; Amel Nagiub; Anne van der Linden; Annemiek van der Eijk; Bas Oude Munnink; Corine GeurtsvanKessel; Dalia Ramadan; David Nieuwenhuijse; Emmanuelle Munger; Galal Mahmoud; Irina Chestakova; Marion Koopmans; Marjan Boter; Mohamed Hassany; Mohamed K. Khalifa; Nancy El Guindy; Ramy Galal; Reina Sikkema; Richard Molenkamp; Salma Sayed; Shymaa A. Showky; Wael H. Roshdy; on behalf of the Dutch national COVID-19 response team. |
| EPI_ISL_1711654, EPI_ISL_1711656 | Virology Laboratory of São Vicente | Institut Pasteur de Dakar | Dia Ndongo; Diagne Moussa Moïse; Diallo Amadou; Diop Mamadou; Faye Ousmane; Loucoubar Cheikh; Nadia Rodrigues; Ndiaye Ndack; Sall Amadou Alpha; Sanders Kevin; Sankhe Safietou; da Luz Lima Mendonça Maria |
| EPI_ISL_2361901 | Virology Laboratory, International Centre for Diarrhoeal Disease Research, Bangladesh (ICDDR,B) | Virology Laboratory, International Centre for Diarrhoeal Disease Research, Bangladesh (ICDDR,B) | Md. Mahfuzur Rahman; Mohammad Enayet Hossain; Mohammed Ziaur Rahman; Mojnu Miah; Mustafizur Rahman; Rashedul Hasan |
| EPI_ISL_1855056 | Virology Laboratory, Scientific Department, Army Medical Center | Virology Laboratory, Scientific Department, Army Medical Center | Anella Monte; Anna Anselmo; Antonella Fortunato; Filippo Molinari; Florigio Lista; Francesco Giordani; Giancarlo Petralito; Giandomenico Cerreto; Riccardo De Sanctis; Silvia Fillo; Vanessa Vera Fain |
| EPI_ISL_508863, EPI_ISL_625456, EPI_ISL_677634, EPI_ISL_677636, EPI_ISL_1660225, EPI_ISL_1660226, EPI_ISL_1660228, EPI_ISL_1660235, EPI_ISL_1660241, EPI_ISL_1660245, EPI_ISL_1660246, EPI_ISL_1660250, EPI_ISL_1660258, EPI_ISL_1660261, EPI_ISL_1660271, EPI_ISL_1660279, EPI_ISL_1660281, EPI_ISL_1660289, EPI_ISL_1660297, EPI_ISL_1660298, EPI_ISL_1660301, EPI_ISL_1660305, EPI_ISL_1660310, EPI_ISL_1660312, EPI_ISL_1660314, EPI_ISL_1660317, EPI_ISL_1660318, EPI_ISL_1660322, EPI_ISL_1660329, EPI_ISL_1660330 | | | |
| see above | Virology Unit, Institut Pasteur de Madagascar | Virology Unit, Institut Pasteur de Madagascar | Angela Brisebarre; Camille Capel; Cara Brook; Cara E. Brook; Christian Ranaivoson; Christophe Malabat; Corinne Maufrais; Cristina M. Tato; Emmanuelle Permal; Etienne Simon-Lorière; Frédéric Lemoine; Helisoa Razafimanjato; Jean-Michel Heraud; Joseph L. DeRisi; Louise Lefrançois; Marion Barbet; Maud Vanpeene; Michelle Tan; Méline Bizard; Norosoa Razanajatovo; Philippe Dussart; Soa Fy Andriamandimby; Sylvie Behillil; Sylvie van der Werf; Tsiry Randriambolamanantsoa; Vida Ahyong; Vincent Enouf; Vololoniaina Raharinosy |
| EPI_ISL_918365, EPI_ISL_918370, EPI_ISL_1534532, EPI_ISL_1706599, EPI_ISL_2501048, EPI_ISL_2693949, EPI_ISL_2801925, EPI_ISL_3031525, EPI_ISL_3031530, EPI_ISL_3031544, EPI_ISL_3045601 | | | |
| see above | Virology Unit, Institut Pasteur du Cambodge | Virology Unit, Institut Pasteur du Cambodge | Cecile Troupin; Chau Darapheak; Chin Savuth; Erik A Karlsson; Etienne Simon-Loriere; Jurre Y Siegers; Kraing Sidonn; Leakhena Pum; Ly Sovann; Sokhoun Yann; Sophoannadedh Rath; Teyputita Ou; Veasna Duong; Yi Sengdoeurn |
| EPI_ISL_411902 | Virology Unit, Institut Pasteur du Cambodge. | Virology Unit, Institut Pasteur du Cambodge (Sequencing done by: Jessica E Manning/Jennifer A Bohl at Malaria and Vector Research Research Laboratory, National Institute of Allergy and Infectious Diseases and Vida Ahyong from Chan-Zuckerberg Biohub) | Erik A Karlsson; Jennifer A Bohl; Jessica E Manning.; Philippe Dussart; Veasna Duong; Vida Ahyong |
| EPI_ISL_416458 | Virology laboratory Ministry of Health Kuwait sequenced at Dasman Diabetes Institute | Dasman Diabetes Institute | Ebaa alOzairi; Fahd Al-Mulla; Motasem Melhem; Qais Al-Duwairi; Rasheeba iqbal; Sara Alqabandi; Sumi John |
| EPI_ISL_708021 | Virology, Universitätsklinikum des Saarlandes | Epigenetics, Saarland University | Jörn Walter; Kathrin Kattler; Markus Vogelgesang; Sascha Tierling; Sigrun Smola; Stefan Lohse |
| EPI_ISL_3050736 | Virus Ecology, NIH | Virus Ecology, NIH | Banakis, S.; Bushmaker, C.; Bushmaker, T.; Chung, M.; Ghedin, E.; Holbrook; M.G.; Munster; Roder, A.; V.J.; Van Doremalen, N.; Wang, W. |
| EPI_ISL_2507964 | Vita Laboratoriot Oy | Expert Microbiology, National Institute for Health and Welfare | Carita Savolainen-Kopra; Erika Lindh; Haider al-Hello; Jani Halkilahti; Kirsi Liitsola; Niina Ikonen; Olli Vapalahti; Pekka Ellonen; Phuoc Truong; Päivi Laurila; Ravi Kant; Sari Hannula; Soile Blomqvist; Teemu Smura |
| EPI_ISL_2547367 | Vredendal North Clinic wc VDN | National Health Laboratory Service/University of Cape Town (NHLS/UCT) | Arash Iranzadeh; Bruna Galvao; Carolyn Williamson; Deelan Doolabh; Diana Hardie; Gert Marais; Innocent Mudau; Lynn Tyers; Marvin Hsiao; Stephen Korsman |
| EPI_ISL_2836853, EPI_ISL_2836857, EPI_ISL_2836861, EPI_ISL_2836864, EPI_ISL_2836868, EPI_ISL_2836869, EPI_ISL_2836871, EPI_ISL_2836872, EPI_ISL_2836875, EPI_ISL_2836880, EPI_ISL_2836883, EPI_ISL_2836885, EPI_ISL_2836886, EPI_ISL_2836887, EPI_ISL_2836890, EPI_ISL_2836897, EPI_ISL_2836898, EPI_ISL_2836899, EPI_ISL_2836901, EPI_ISL_2836905, EPI_ISL_2836907, EPI_ISL_2836908, EPI_ISL_2836909, EPI_ISL_2836910, EPI_ISL_2836911, EPI_ISL_2836912, EPI_ISL_2836913, EPI_ISL_2836918, EPI_ISL_2836919, EPI_ISL_2836920, EPI_ISL_2836921, EPI_ISL_2836922, EPI_ISL_2836924, EPI_ISL_2836925, EPI_ISL_2836930, EPI_ISL_2836932, EPI_ISL_2836933, EPI_ISL_2836934, EPI_ISL_2836936, EPI_ISL_2836938, EPI_ISL_2836942, EPI_ISL_2836944, EPI_ISL_2836945, EPI_ISL_2836946, EPI_ISL_2836947, EPI_ISL_2836952, EPI_ISL_2836953, EPI_ISL_2836954, EPI_ISL_2836955, EPI_ISL_2836956, EPI_ISL_2836957, EPI_ISL_2836958, EPI_ISL_2836961, EPI_ISL_2836962, EPI_ISL_2836963, EPI_ISL_2836965, EPI_ISL_2836966, EPI_ISL_2836968, EPI_ISL_2967997, EPI_ISL_2968002, EPI_ISL_2968004, EPI_ISL_2968009, EPI_ISL_2968010, EPI_ISL_2968011, EPI_ISL_2968012, EPI_ISL_2968015, EPI_ISL_2968016, EPI_ISL_2968021, EPI_ISL_2968023, EPI_ISL_2968025, EPI_ISL_2968030, EPI_ISL_2968031, EPI_ISL_2968034 | | | |
| see above | WACCBIP, University of Ghana, Accra, Ghana | WACCBIP, University of Ghana, Volta Road, Legon-Accra, Ghana | Bright K. Yemi; Collins M. Morang'a; Deborah N. A. Mettle; Dominic S. Y. Amuzu; Dominic S.Y. Amuzu; Evelyn B. Quansah; Evelyn Y. Bonney; Frederick Tei-Maya; Israel Osei-Wusu; Ivy A. Asante; Joe K. Mutungi; John K. Odoom; Joseph H.K. Bonney; Joyce M. Ngoi; Lawrence Ofori-Boadu; Lucas N. Amenga-Etego and Gordon A. Awandare; Mildred Adusei-Poku; Nicaise T. Ndam; Oliver Commey; Peter K Quashie; Philip M. Soglo; Samirah Saiid; Samuel Armoo; Vincent Appiah; Violette V. M'cormack; William K. Ampofo; Yaw Bediako |
| EPI_ISL_733021, EPI_ISL_1372304, EPI_ISL_1797437, EPI_ISL_2385079, EPI_ISL_2523488, EPI_ISL_2816149 | WHO National Influenza Centre Russian Federation | WHO National Influenza Centre Russian Federation | Andrey Komissarov; Anna Ivanova; Artem Fadeev; Daria Danilenko; Dmitry Bazhenov; Dmitry Lioznov; Elena Nabieva; Georgii Bazykin; Kirill Varchenko; Ksenia Safina; Kseniya Komissarova; Maria Pisareva; Maria Timofeeva; Mikhail Bakaev; Nikita Yolshin; Oula Mansour; Oula Masour; Tamila Musaeva; Veronika Eder |
| EPI_ISL_860802, | WHO/Minsk | Charité Universitätsmedizin Berlin, Institut für Virologie | Barbara Mühlemann; Christian Drosten; Julia Schneider; Julia Tesch; Jörn Beheim-Schwarzbach; Shmialiova Natallia; Sivets Natallia; Talitha Veith; Terry Jones; Tobias Bleicker; Victor M Corman |

| | | | |
|---|---|---|---|
| EPI_ISL_860805, EPI_ISL_860806, EPI_ISL_860809 | | | |
| EPI_ISL_1627990 | WSSE w Krakowie | 1. National Institute of Public Health - National Institute of Hygiene; 2. Eurofins Genomics Europe Sequencing GmbH | ECDC COVID-19 WGS support team; Eurofins Genomics Europe Sequencing Team; Gierczyński Rafał; Sadkowska-Todys Małgorzata; Wołkowicz Tomasz; Zacharczuk Katarzyna |
| EPI_ISL_2365408, EPI_ISL_2365412 | WWF Bayanga field laboratory | Robert Koch Institute | F. H. Leendertz; F. S. Niatou-Singa; M. Ulrich; S. Calvignac-Spencer; T. B. Tombolomako; T. Fuh-Neba; U. Vickos |
| EPI_ISL_2365414, EPI_ISL_2365415, EPI_ISL_2365416, EPI_ISL_2365417 | WWF Bayanga field laboratory | WWF Bayanga field laboratory | F. H. Leendertz; F. S. Niatou-Singa; M. Ulrich; S. Calvignac-Spencer; T. B. Tombolomako; T. Fuh-Neba; U. Vickos |
| EPI_ISL_2987085 | WY Public Health Laboratory | Centers for Disease Control and Prevention Division of Viral Diseases, Pathogen Discovery | Alison Laufer Halpin; Ben L. Rambo-Martin; Clinton R. Paden; Dakota Howard; Darlene Wagner; Dave Wentworth; Dhwani Batra; Jasmine Padilla; Justin Lee; Katie Dillon; Krista Queen; Kristen Knipe; Kristine Lacek; Mark Burroughs; Matthew Schmerer; Mili Sheth; Peter Cook; Sam Shepard; Sarah Nobles; Shoshona Le; Suxiang Tong; Vivien Dugan; Yvette Unoarumhi |
| EPI_ISL_754579 | Wadsworth Center, New York State Department.of Health | Wadsworth Center, New York State Department.of Health | Alexis Russel; Daryl M. Lamson; Erasmus Schneider; Erica Lasek-Nesselquist; John Kelly; Jonathan Plitnick; Kirsten St. George; Matthew Shudt; Melissa A Leisner; Navjot Singh; Sara Griesemer |
| EPI_ISL_456190, EPI_ISL_536790, EPI_ISL_548064, EPI_ISL_548101 | Waikato Hospital | Institute of Environmental Science and Research (ESR) | Anja Werno; Antje van der Linden; Arlo Upton; Chris Mansell; David Hammer; Dragana Drinkovic; Erasmus Smit; Gary McAuliffe; Hana Sofia Andersson; Hermes Perez; James Ussher; Jill Sherwood; Jing Wang; Joep de Ligt; Josh Freeman; Julia Howard; Juliet Elvy; Lauren Jelly; Mary DeAlmeida; Matt Blakiston; Matt Storey; Matthew Rogers; Max Bloomfield; Michael Addidle; Michelle Balm; Muhammad Faisal; Nikki Freed; Olin Silander; Sally Roberts; Sarah Jefferies; Sharmini Muttaiyah; Susan Morpeth; Susan Taylor; Timothy Blackmore; Vani Sathyendran; Veronica Playle; Virginia Hope; Xiaoyun Ren |
| EPI_ISL_1833203 | Wales Specialist Virology Centre Sequencing lab: Pathogen Genomics Unit | Public Health Wales Microbiology Cardiff Wales Specialist Virology Centre | Alec Birchley; Alexander Adams; Amy Gaskin; Angela Marchbank; Bree Gatica-Wilcox; Catherine Moore; Jason Coombes; Joanne Watkins; Joel Southgate; Johnathan Evans; Laura Gifford; Lauren Gilbert; Lee Graham; Malorie Perry; Matthew Bull; Nicole Pacchiarini; Sally Corden; Sara Kumziene-Summerhayes; Sara Rey; Sarah Taylor; Simon Cottrell; Sophie Jones; Tom Connor |
| EPI_ISL_417160 | Washington State Department of Health | Seattle Flu Study | Chu etl al |
| EPI_ISL_416538 | Wellington Hospital | Institute of Environmental Science and Research (ESR) | New Zealand; Newtown; Riddiford Street; Wellington 6021; Wellington Hospital; Wellington SCL |
| EPI_ISL_456161, EPI_ISL_456185, EPI_ISL_456320 | Wellington SCL | Institute of Environmental Science and Research (ESR) | Anja Werno; Antje van der Linden; Arlo Upton; Chris Mansell; David Hammer; Dragana Drinkovic; Erasmus Smit; Gary McAuliffe; Hana Sofia Andersson; James Ussher; Jill Sherwood; Joep de Ligt; Josh Freeman; Julia Howard; Juliet Elvy; Lauren Jelly; Mary DeAlmeida; Matt Blakiston; Matt Storey; Matthew Rogers; Max Bloomfield; Michael Addidle; Michelle Balm; Sally Roberts; Sarah Jefferies; Sharmini Muttaiyah; Susan Morpeth; Susan Taylor; Timothy Blackmore; Vani Sathyendran; Veronica Playle; Virginia Hope; Xiaoyun Ren |
| EPI_ISL_1016878, EPI_ISL_1250695, EPI_ISL_2964938, EPI_ISL_2964939 | Wellington SCL (WN) | Institute of Environmental Science and Research (ESR) | Anja Werno; Antje van der Linden; Arlo Upton; Chris Mansell; David Hammer; Dragana Drinkovic; Erasmus Smit; Gary McAuliffe; Hana Sofia Andersson; Hermes Perez; James Ussher; Jill Sherwood; Jing Wang; Joep de Ligt; Josh Freeman; Julia Howard; Juliet Elvy; Lauren Jelly; Mary DeAlmeida; Matt Blakiston; Matt Storey; Matthew Rogers; Max Bloomfield; Michael Addidle; Michelle Balm; Muhammad Faisal; Nikki Freed; Olin Silander; Olivia Stroeven; Rachel Boyle; Sally Roberts; SallyAnn Harbison; Sarah Jefferies; Sharmini Muttaiyah; Susan Morpeth; Susan Taylor; Timothy Blackmore; Vani Sathyendran; Veronica Playle; Virginia Hope; Xiaoyun Ren |
| EPI_ISL_2422499, EPI_ISL_2422500, EPI_ISL_2422505, EPI_ISL_2422515, EPI_ISL_2422517, EPI_ISL_2422520, EPI_ISL_2422521, EPI_ISL_2422526, EPI_ISL_2422530, EPI_ISL_2422549, EPI_ISL_2422571, EPI_ISL_2422595, EPI_ISL_2422614, EPI_ISL_2422624 | | | |
| see above | West African Centre for Cell Biology of Infectious Pathogen, University of Ghana, Legon | WACCBIP, University of Ghana, Volta Road, Legon, Accra | Collins M. Morang'a; Dominic S. Y. Amuzu; Dominic S.Y. Amuzu; Edward Danso Fenteng; Emmanuel Kudjo; Evelyn B. Quansah; Evelyn Y Bonney; Frederick Tei-Maya; Ivy A Asante; Joe K. Mutungi; John K. Odoom; Joseph HK Bonney; Joyce M. Ngoi; Lawrence Ofori-Boadu; Lucas N. Amenga-Etego and Gordon A Awandare; Lucas N. Amenga-Etego and Gordon A. Awandare; Mildred Adusei-Poku; Nicaise T. Ndam; Patrick Tetteh Ababio; Peter K Quashie; Peter K. Quashie; Philip M Soglo; Philip M. Soglo; Samirah Saiid; Theophilus Odoom; Vincent Appiah; Violet MCormack; Violette M’cormack; William K. Ampofo; Yaw Bediako |
| EPI_ISL_1255107, EPI_ISL_1255110, EPI_ISL_1255150, EPI_ISL_1255152, EPI_ISL_1255164, EPI_ISL_1255170, EPI_ISL_1255176, EPI_ISL_1255186, EPI_ISL_1255188, EPI_ISL_1255199, EPI_ISL_1255229, EPI_ISL_1255231, EPI_ISL_1255244, EPI_ISL_1255250 | | | |
| see above | West African Centre for Cell Biology of Infectious Pathogens (WACCBIP), University of Ghana, Accra, Ghana | West African Centre for Cell Biology of Infectious Pathogens (WACCBIP), University of Ghana, Volta Road, Legon-Accra, Ghana | ; Abdoulaye B Diallo; Abdul-Karim Abass; Aisha Mohammed; Benjamin Demah Nuertey; Collins M. Morang'a; Dam Kenneth Mibut; Dominic S.Y. Amuzu; Emmanuella Amoako4; Evelyn B. Quansah; Frederick Kumi-Ansah; Frederick Tei-Maya; Gordon A Awandare; Joyce M. Ngoi; Kesego Tapela; Lucas N. Amenga-Etego; Nelson Kibinge; Oliver D Boakye5; Peter K Quashie; Philip M. Soglo; Samirah Saiid; Samuel Kaba Akoriyea; Theophilus Odoom; Vanessa Magnussen; Vincent Appiah; Yaw Bediako |
| EPI_ISL_451397 | West China Hospital of Sichuan University | State Key Laboratory of Biotherapy of Sichuan University | Baowen Du; Binwu Ying; Chao Tang; Chuan Chen; Hancheng Wei; Jia Geng; Jing-wen Lin; Lu Chen; Mingxia Yu; Minjin Wang; Weimin Li; Yongzhao Zhou |
| EPI_ISL_2382409 | Wisma Atlet | National Institute of Health Research and Development | Arie Ardiansyah Nugraha; Hana Apsari Pawestri; Hartanti Dian Ikawati; Kartika Dewi Puspa; Krisna Pangesti; Nelly Puspandari; Subangkit; Triyani Soekarso; Vivi Setiawaty |
| EPI_ISL_1300721, EPI_ISL_1493351, EPI_ISL_1731710, EPI_ISL_2500299, EPI_ISL_2692941, EPI_ISL_2692972 | Wojewodzka Stacja Sanitarno-Epidemiologiczna w Olsztynie, Laboratorium Badan Epidemiologiczno-Klinicznych | Wojewodzka Stacja Sanitarno-Epidemiologiczna w Olsztynie, Laboratorium Badan Epidemiologiczno-Klinicznych | Aleksandra Kobiatko; Barbara Dolinska; Emilia Tarabasz; Ewa Liszewska; Marta Lukian; Monika Czerminska; Patryk Bielecki; Paulina Rozycka; Sylwia Krzetowska; Tomasz Jakubczak |
| EPI_ISL_3009173 | Wojewódzki Szpital Specjalistyczny Laboratorium Mikrobiologiczne | 1. Academic Center for Pathomorphological and Genetic-Molecular Diagnostics ltd, Bialystok, Poland 2. National Institute of Public Health - National Institute of Hygiene, Warsaw, Poland | Anetta Sulewska; Jacek Nikliński; Janusz Dzięcioł; Joanna Kiśluk; Katarzyna Zacharczuk; Konrad Raczkowski; Magdalena Nowakowska; Małgorzata Sadkowska-Todys; Piotr Karabowicz; Piotr Majewski; Przemysław Biecek. Joanna Reszeć; Radosław Charkiewicz; Tomasz Wołkowicz |
| EPI_ISL_454905, EPI_ISL_454939, EPI_ISL_454982 | Wuhan Chain Medical Labs (CMLabs) | State Key Laboratory of Biotherapy of Sichuan University | Baowen Du; Binwu Ying; Chao Tang; Chuan Chen; Hancheng Wei; Jia Geng; Jing-wen Lin; Lu Chen; Mingxia Yu; Minjin Wang; Weimin Li; Yongzhao Zhou |
| EPI_ISL_462931 | Wyoming Public Health Laboratory | Center for Global Health, University of New Mexico Health Sciences Center | Cari Sloma; Darrell Dinwiddie; Daryl Domman; Kurt Schwalm; Noah Hull; Rob Christensen; Wanda Manley |
| EPI_ISL_1532099, EPI_ISL_2404707, EPI_ISL_2444758, EPI_ISL_2716283, EPI_ISL_2775587 | Wyoming Public Health Laboratory | Wyoming Public Health Laboratory | Ashley Norberg; Brian Dominguez; Brittany Oher; Cari Sloma; Channing Weber; Chayse Rowley; Elliot Thomasson; Jim Mildenberger; Lynette Gumbleton; Marley Goetz; Noah Hull; Robert Petit; Sam Britz; Taylor Fearing; Wanda Manley; and Rob Christensen |
| EPI_ISL_1595852 | Yunan Center for Disease Control and Prevention | National Institute for Viral Disease Control and Prevention, China CDC | Jienan Zhou; Meiling Zhang; Senquan Jia; Xiang Zhao; Xiaoqing Fu; Yenan Feng; Yuchao Wu; Zhixiao Chen |
| EPI_ISL_2727271 | ZARV/NHLS, Department Medical Virology, University of Pretoria | KRISP, KZN Research Innovation and Sequencing Platform | Adriano Mendes; Amy Strydom; Emmanuel SJ; Giandhari J; Lessells R; Micheala Davids; Naidoo Y; Pillay S; Ramphal U; Sim Mayaphi and Marietjie Venter; Tegally H; Wilkinson E; de Oliveira T |
| EPI_ISL_2727350, EPI_ISL_2899747 | ZARV/NHLS, Department Medical Virology, University of Pretoria | KRISP, KZn Research Innovation and Sequencing Platform | Adriano Mendes; Amy Strydom; Emmanuel SJ; Giandhari J; Lessells R; Micheala Davids; Naidoo Y; Pillay S; Ramphal U; Sim Mayaphi and Marietjie Venter; Tegally H; Wilkinson E; de Oliveira T |
| EPI_ISL_3039570 | ZOL | Jessa | Berden et al. on behalf of the Jessa_cmdLab |
| EPI_ISL_666636, EPI_ISL_718079, EPI_ISL_718096 | ZOTZ KLIMAS MVZ Düsseldorf-Centrum GbR ÜBAG für Labormedizin, Genetik, Zytologie, Pathologie | Center of Medical Microbiology, Virology, and Hospital Hygiene, University of Duesseldorf | Alexander Dilthey; Andreas Walker; Ashley-Jane Duplessis; Daniel Strelow; Jessica Nicolai; Jörg Timm; Katrin Hoffmann; Klaus Pfeffer; Lisanna Hülse; Malte Kohns Vasconcelos; Marek Korencak; Maximilian Damagnez; Nadine Lübke; Patrick Finzer; Rainer Zotz; Tobias Wienemann; Torsten Houwaart |
| EPI_ISL_2377264 | Zakład Laboratoryjnej Diagnostyki Klinicznej USK Białystok | 1. National Institute of Public Health - National Institute of Hygiene; 2. Eurofins Genomics Europe Sequencing GmbH | ECDC COVID-19 WGS support team; Eurofins Genomics Europe Sequencing Team; Gierczyński Rafał; Sadkowska-Todys Małgorzata; Wołkowicz Tomasz; Zacharczuk Katarzyna |
| EPI_ISL_2674012, EPI_ISL_2674013 | Zavod za javno zdravstvo Šibensko- Kninske Županije | Hrvatski zavod za javno zdravstvo | Irena Tabain; Ivana Ferenčak |
| EPI_ISL_1911196, EPI_ISL_1911197, EPI_ISL_1911250 | Zhejiang Provincial Center for Disease Control and Prevention Zhoushan Center for Disease Prevention and Contol | Zhejiang Province Center of Disease Control and prevention | Bing WU; Hongling Wang; Yanjun Zhang |
| EPI_ISL_2716621 | aboratoire National de Référence pour les Fièvres Hémorragiques Virales, Centre Muraz | Centre Muraz | Ange Badjo; Arsène Zongo; Essia Belarbi; Fabian Leendertz; Grit Schubert; Jasmin Schlotterbeck; Saïdou Ouedraogo; Soumeya Ouangraoua; Thérèse Kagone |
| EPI_ISL_1727293, EPI_ISL_1727369 | amedes MVZ Hannover | Robert Koch Institute | |
| EPI_ISL_1281348 | amedes MVZ Jena | Robert Koch Institute | |
| EPI_ISL_1141025 | amedes MVZ für Laboratoriumsmedizin Rhein/Ruhr | Robert Koch Institute | |
| EPI_ISL_828071, EPI_ISL_828410, EPI_ISL_829200 | deCODE genetics | deCODE genetics | Agnar Helgason; Alma Moller; Arna B Agustsdottir; Arnaldur Gylfason; Asgeir Sigurdsson; Aslaug Jonasdottir; Berglind Eiriksdottir; Bjarni Thorbjornsson; Brynjar O Jensson; Daniel F Gudbjartsson; Droplaug N Magnusdottir; Elisabet E Gardarsdottir; Emil A Thorarensen; Gardar Sveinbjornsson; Gisli Masson; Gudmundur Georgsson; Gudmundur L Norddahl; Gudrun Sigmundsdottir; Hakon Jonsson; Hannes Eggertsson; Hilma Holm; Ingileif Jonsdottir; Jona Saemundsdottir; Kamilla S Josefsdottir; Kari Stefansson; Karl G Kristinsson; Kjartan R Gudmundsson; Kristin E Sveinsdottir; Louise le Roux; Maney Sveinsdottir; Olafia S Gretarsdottir; Olafur T Magnusson; Pall Melsted; Patrick Sulem; Run Fridriksdottir; Solvi Rognvaldsson; Thora R Gunnarsdottir; Thordur Kristjansson; Thorolfur Gudnason; Unnur Thorsteinsdottir |
| EPI_ISL_732911 | genXone SA, Molecular Diagnostics Laboratory / NZOZ | genXone SA, Research & Development Laboratory | Grzegorz Nowicki; Jakub Grabowski; Maciej Sykulski; Michał Kaszuba; Monika Mańkowska-Woźniak; Natalia Drwęska-Matelska; Łukasz Krych |
| EPI_ISL_458286 | unknown | Bundeswehr Institute of Microbiology | Antwerpen; Bestehorn-Willmann; Eckstein, S.; Handrick, S.; M.C.; M.H.; M.S.; Naija, H.; R. and Ben Moussa, M.; Rehn, A.; Stoecker, K.; Walter; Woelfel |
| EPI_ISL_463005, EPI_ISL_463006 | unknown | Clinical virology | Fares, W.; Triki, H. |
| EPI_ISL_487270 | unknown | Communicable Disease Laboratory, Public Health Directorate | AlTaif, Z.; AlWasti, H.; Shehab, F.; Zaed, A. |
| EPI_ISL_437612, | unknown | Faculty of Medicine | Buathong, R.; Bunprakob, S.; Ghai, S.; Joyjinda, Y.; Mungaomklang, A.; Petcharat, S.; Plipat; Prasithsirikul, W.; Rodpan, A.; Sirichan, N.; T. and Hemachudha, T.; Wacharapluesadee, S. |

| | | | |
|---|---|---|---|
| EPI_ISL_437616, EPI_ISL_437618, EPI_ISL_437622 | | | |
| EPI_ISL_507012 | unknown | Infectious Diseases Research, King Abdullah International Medical Research Center (KAIMRC) | Alghoribi; M.F. |
| EPI_ISL_476559 | unknown | Laboratoire Sciences et Technologies de la Santé (STS) Institut Supérieur des Sciences de la Santé Université Hassan 1er, Settat, Morocco | Abderraouf Hilali; Amal Souiri; Hajar Lemriss; Jawad Bouzid; Mustapha Mouallif; Narjis Amar; Sanaâ Lemriss; Saâd EL Kabbaj; Touria Essayagh |
| EPI_ISL_483060 | unknown | Microbiology, Canterbury Health Laboratories | Anderson, T.; Dilcher, M. |
