## Supplementary figures and images for "SARS-CoV-2 Genetic diversity and lineage dynamics of in Egypt"

### Figure S1

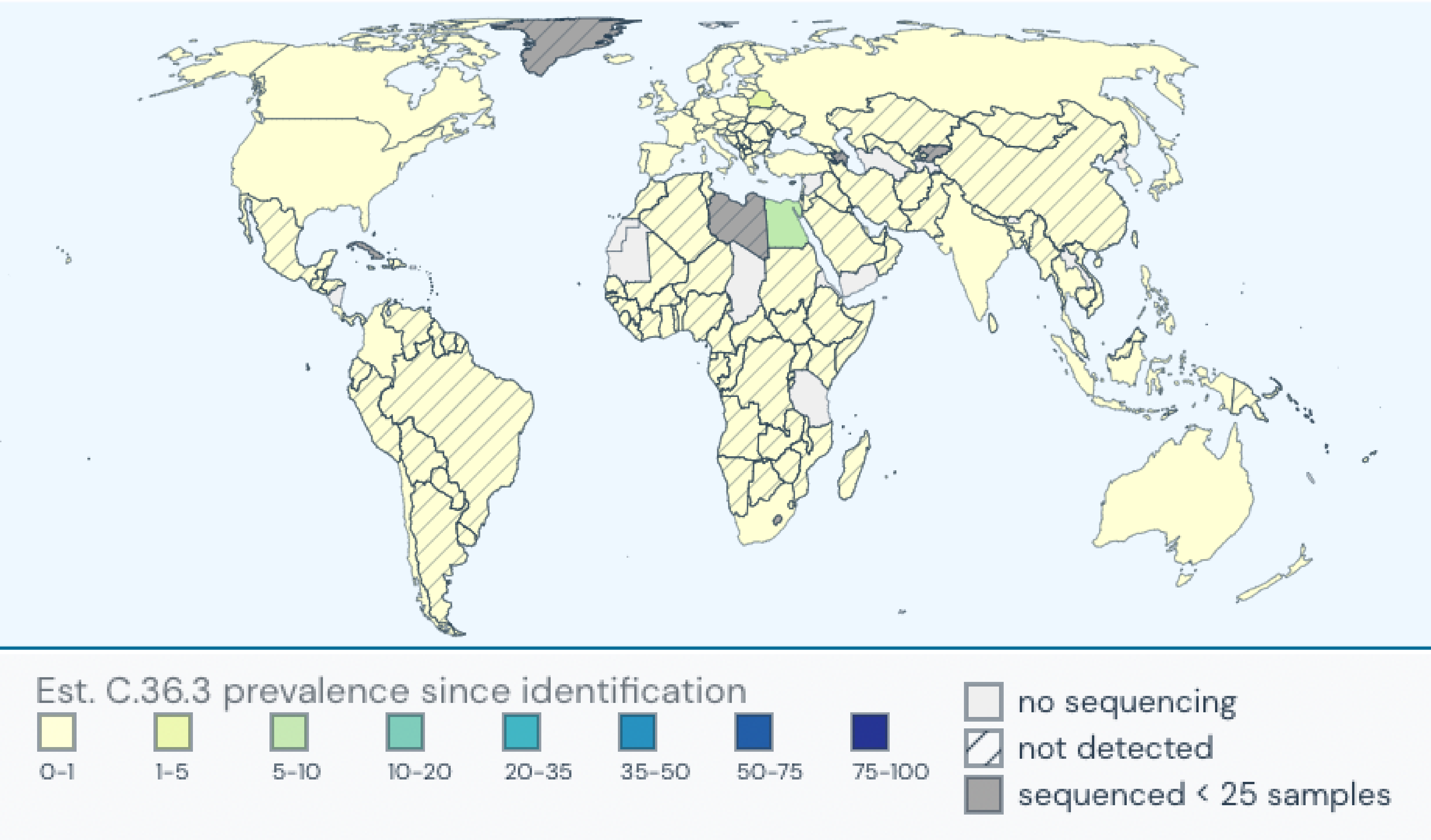

### Figure S2

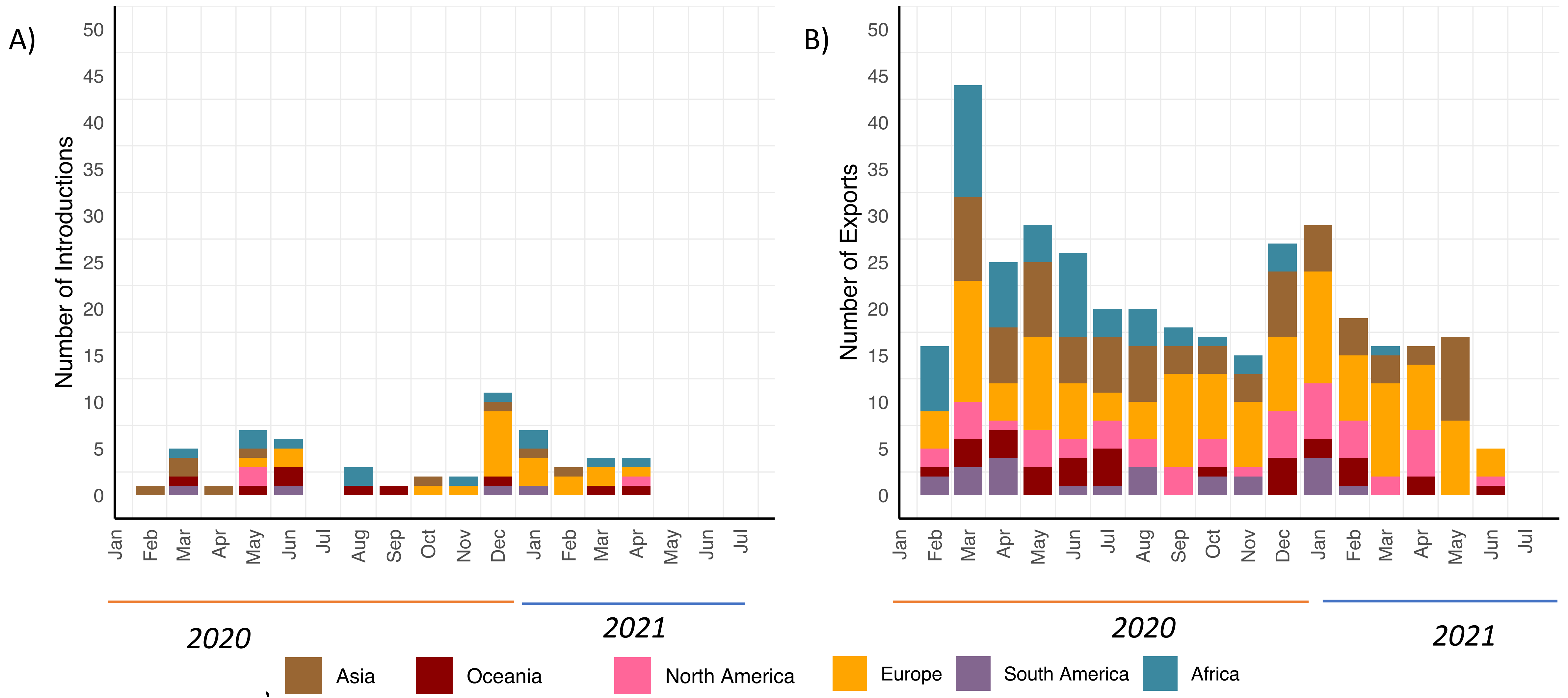
